# Colchicine for COVID-19 in adults in the community (PRINCIPLE): a randomised, controlled, adaptive platform trial

**DOI:** 10.1101/2021.09.20.21263828

**Authors:** PRINCIPLE Trial Collaborative Group, Jienchi Dorward, Ly-Mee Yu, Gail Hayward, Benjamin R Saville, Oghenekome Gbinigie, Oliver Van Hecke, Emma Ogburn, Philip H Evans, Nicholas PB Thomas, Mahendra G Patel, Duncan Richards, Nicholas Berry, Michelle A Detry, Christina Saunders, Mark Fitzgerald, Victoria Harris, Milensu Shanyinde, Simon de Lusignan, Monique I Andersson, Christopher C Butler, FD Richard Hobbs

**Affiliations:** Nuffield Department of Primary Care Health Sciences, University of Oxford, Oxford, UK; Centre for the AIDS Programme of Research in South Africa (CAPRISA), University of KwaZulu–Natal, Durban, South Africa; Berry Consultants, Texas, USA; Department of Biostatistics, Vanderbilt University School of Medicine, Tennessee, USA; College of Medicine and Health, University of Exeter, Exeter, UK; National Institute for Health Research (NIHR) Clinical Research Network, National Institute for Health Research, London, UK; Royal College of General Practitioners, London, UK; Oxford Clinical Trials Research Unit, Botnar Research Centre, University of Oxford, UK; Nuffield Department of Clinical Medicine, University of Oxford, Oxford, UK

**Author notes:** Writing committee listed below on behalf of the PRINCIPLE Trial Collaborative Group. PRINCIPLE trial collaborators are listed in the appendix. Drs Dorward and Yu and contributed equally to this article. Professors Hobbs and Butler contributed equally to this article. **Corresponding authors:** Professors Richard Hobbs and Chris Butler, Address: Nuffield Department of Primary Care Health Sciences, University of Oxford, Gibson Building 1st Floor, Radcliffe Observatory Quarter, Woodstock Road, Oxford, OX2 6GG.

## Abstract

**Objectives:** Colchicine has been proposed as a COVID-19 treatment, but its effect on time to recovery is unknown. We aimed to determine whether colchicine is effective at reducing time to recovery and COVID-19 related hospitalisations/deaths among people in the community.

**Design:** Prospective, multicentre, open-label, multi-arm, adaptive Platform Randomised Trial of Treatments in the Community for Epidemic and Pandemic Illnesses (PRINCIPLE).

**Setting:** National trial run remotely from a central trial site and at multiple primary care centres across the United Kingdom.

**Participants:** Adults aged ≥65, or ≥18 years with comorbidities or shortness of breath, and unwell ≤14 days with suspected COVID-19 in the community.

**Interventions:** Participants were randomised to usual care, usual care plus colchicine (500µg daily for 14 days), or usual care plus other interventions.

**Main outcome measures:** The co-primary endpoints were time to first self-reported recovery, and hospitalisation/death related to COVID-19, within 28 days, analysed using Bayesian models. The hypothesis for the time to recovery endpoint is evaluated first, and if superiority is declared on time to recovery, the hypothesis for the second co-primary endpoint of hospitalisation/death is then evaluated. To determine futility, we pre-specified a clinically meaningful benefit in time to first reported recovery as a hazard ratio of 1.2 or larger (equating to approximately 1.5 days benefit in the colchicine arm, assuming 9 days recovery in the usual care arm).

**Results:** The trial opened on April 2, 2020, with randomisation to colchicine starting on March 04, 2021 and stopping on May 26, 2021, because the pre-specified time to recovery futility criterion was met. The primary analysis model included 2755 SARS-CoV-2 positive participants, randomised to colchicine (n=156), usual care (n=1145), and other treatments (n=1454). Time to first self-reported recovery was similar in the colchicine group compared with usual care with an estimated hazard ratio of 0.919 [95% credible interval 0.72 to 1.16] and an estimated increase of 1.14 days [−1.86 to 5.21] in median time to self-reported recovery for colchicine versus usual care. The probability of meaningful benefit in time to recovery was very low at 1.8%. Results were similar in comparisons with concurrent controls. COVID-19 related hospitalisations/deaths were similar in the colchicine group versus usual care, with an estimated odds ratio of 0.76 [0.28 to 1.89] and an estimated difference of −0.4% [−2.7% to 2.4]. One serious adverse event occurred in the colchicine group and one in usual care.

**Conclusions:** Colchicine did not improve time to recovery in people at higher risk of complications with COVID-19 in the community.

**Trial registration:** ISRCTN86534580.

## Introduction

Colchicine is a widely used for the treatment and prophylaxis of gout. Colchicine inhibits cellular transport and mitosis by binding to tubulin and preventing its polymerisation as part of the cytoskeleton transport system.^1^ Although the precise mechanism is unclear, colchicine has an inhibitory action on the NLRP3 inflammasome.^1^ Inflammasomes are activated in COVID-19 and that the degree of activation is correlated with disease severity.^2^ Colchicine is therefore an attractive candidate to test the role of the inflammasome in COVID-19.^3^

Several observational studies and one small randomised controlled trial suggested that colchicine may be an effective treatment for patients hospitalised with COVID-19.^4–7^ However, the large RECOVERY trial of hospitalised COVID-19 patients clearly demonstrated that colchicine did not improve the primary outcome of 28 day mortality, or any secondary outcomes, when compared with usual care.^8^ Though the actions of colchicine may be more relevant earlier in disease to prevent the progression from inflammatory activation to a hyperinflammatory state,^3^ evidence for effectiveness of colchicine in the community is lacking. The COLCORONA randomised controlled trial, among 4488 people aged ≥40 years with suspected COVID-19 in the community, was stopped early for administrative reasons and did not reach the pre-specified superiority criterion for a reduction in the primary outcome of COVID-19 related hospitalisation/death.^9^ However, in a pre-specified secondary analysis among severe acute respiratory distress syndrome coronavirus 2 (SARS-CoV-2) polymerase chain reaction (PCR) positive participants, there was a marginally significant reduction in COVID-19 related hospitalisations and death compared to placebo (4.6% vs 6.0%; OR 0.75, 0.57–0.99; p=0.042). Pulmonary emboli and gastrointestinal adverse events were significantly higher in the colchicine arm, and the trial did not measure time to recovery. While encouraging, these data are generally considered insufficient to support a recommendation and several key bodies have called for more information.^10–12^

We aimed to determine whether colchicine speeds recovery and reduces COVID-19 related hospital admission or death in people in the community.

## Methods

### Trial design

We assessed the effectiveness of colchicine in the UK national, multi-centre, primary care, open-label, multi-arm, prospective adaptive Platform Randomised trial of Treatments in the Community for Pandemic and Epidemic Illnesses (PRINCIPLE), which opened on April 2, 2020, and is ongoing. The protocol is available in the appendix (pp 4-80) and at the trial website, www.principletrial.org. A “platform trial” allows multiple treatments for the same disease to be tested simultaneously. A master protocol defines prospective decision criteria for dropping interventions for futility, declaring interventions superior, or adding new interventions.^13^ This allows interventions with little evidence of meaningful benefit to be rapidly dropped for futility and replaced by new interventions, thereby directing resources towards identifying community-based treatments for COVID-19. Interventions evaluated in PRINCIPLE include hydroxychloroquine, azithromycin,^14^ doxycycline,^15^ inhaled budesonide,^16^ favipiravir, ivermectin and, reported here, colchicine.

The UK Medicines and Healthcare products Regulatory Agency and the South Central-Berkshire Research Ethics Committee (Ref: 20/SC/0158) approved the trial protocol. Online consent was obtained from all participants. The authors vouch for the accuracy and completeness of the data and for fidelity to the protocol. An independent Trial Steering Committee and Data Monitoring and Safety Committee provided trial oversight.

### Participants

From the beginning of the trial, people in the community were eligible if they were aged ≥65 years, or 50-65 years with comorbidities (appendix p 8), and had ongoing symptoms from polymerase chain reaction (PCR) confirmed or suspected COVID-19 (in accordance with the UK National Health Service definition of high temperature and/or new, continuous cough and/or change in sense of smell/taste),^17,18^ which had started within the previous 14 days. When the colchicine arm opened, eligibility criteria were expanded to allow enrolment of people aged 18-65 years with comorbidities or shortness of breath.^19^ Comorbidities required for eligibility were: heart disease; hypertension; asthma or lung disease; diabetes; hepatic impairment; stroke or neurological problems; weakened immune system (e.g. chemotherapy); and self-reported obesity or body mass index ≥35 kg/m^2^. People were ineligible to be randomised to colchicine if they were already taking colchicine or if colchicine was contraindicated according to the British National Formulary. Initially, eligible people were recruited, screened and enrolled through participating general medical practices, but from May 17, 2020, people across the UK could enrol online or telephonically. After patients completed a baseline and screening questionnaire, a clinician or trained research nurse confirmed eligibility using the patient’s primary care medical record, accessed remotely where necessary, before randomisation. We implemented several community outreach strategies aiming to increase recruitment of those from ethnically diverse communities and socioeconomically deprived backgrounds, who have been disproportionally affected by COVID-19.^20^

### Randomisation and masking

Eligible, consenting participants were randomised using a secure, in-house, web-based randomisation system (Sortition version 2.3). Randomisation was stratified by age (< 65 years /≥ 65 years), and presence of comorbidity (yes/no) and probabilities were determined using response adaptive randomisation via regular interim analyses, which allows allocation of more participants to interventions with better observed time to recovery outcomes (appendix pp 140-142). However, between March 31, 2021 and April 08, 2021, only the colchicine and usual care arms were active, with 1:1 allocation between each. The trial team was blinded to randomisation probabilities.

### Trial procedures

Participants were followed up through an online, daily symptom diary for 28 days after randomisation, supplemented with telephone calls to non-responders on days 7, 14 and 28. The diary includes questions about illness recovery (ascertained by answering the question, “Do you feel recovered today? (i.e. symptoms associated with illness are no longer a problem) Yes/No”), overall illness severity (a rating of how well they are feeling on a scale of 1-10 [1 being the worst and 10 being the best]), individual symptom severity on a four-point scale (0 = no problem to 3 = major problem), and healthcare service utilisation. Participants could nominate a trial partner to help provide follow up data. We obtained consent to ascertain healthcare use outcome data from general practice and hospital records. We aimed to provide a self-swab for SARS-CoV-2 confirmatory PCR testing, but capacity issues early in the pandemic meant testing was unavailable for some participants.

### Trial interventions

Participants received usual care plus colchicine 500µg daily for 14 days, or usual care alone. Colchicine was either prescribed or issued directly by the participant’s general medical practitioner, or issued centrally by the study team and delivered to the participant by urgent courier. Usual care in the UK National Health Service for suspected COVID-19 in the community is largely focused on managing symptoms with antipyretics,^21^ although previous results from PRINCIPLE^16^ led to the introduction of inhaled budesonide on an off-label, case-by-case basis for people aged ≥65 years or 50-65 with co-morbidities.^22^

### Primary outcomes

The trial commenced with the primary outcome of COVID-19 related hospitalisation or death within 28 days. However, hospitalisation rates in the UK^23^ were lower than initially expected^24^. Therefore, the Trial Management Group and Trial Steering Committee recommended amending the primary outcome to also include illness duration,^25,26^ which is an important outcome for patients and has substantial economic and social impacts. This received ethical approval on September 16, 2020, and was implemented before performing any interim analyses. Thus, the trial has two co-primary endpoints measured within 28 days of randomisation: 1) time to first reported recovery defined as the first instance that a participant reports feeling recovered; and 2) hospitalisation or death related to COVID-19. Decisions about COVID-19 relatedness were made after independent review of available data by two clinicians blinded to treatment allocation and study identifiers.

### Secondary outcomes

Secondary outcomes (defined in section 3.3. of the Master Statistical Analysis Plan, appendix pp 102-109) include a binary outcome of early, sustained recovery (recovered by day 14 and remains recovered until day 28), time to sustained recovery (date participant first reports recovery and subsequently remains well until 28 days), daily rating from 1-10 of how well participants feel, time to initial alleviation of symptoms (date symptoms first reported as minor or none), time to sustained alleviation of symptoms (date symptoms first reported as minor or none and subsequently remain minor or none until 28 days), time to initial reduction of severity of symptoms (among people with symptom at baseline, date symptom severity reported at least one scale lower), worsening of symptoms (worsening symptom by one grade from mild to moderate/severe, or from moderate to severe, and excluding individuals reporting symptom severity as major at baseline), contacts with healthcare services, hospital assessment without admission, duration of hospital admission, oxygen administration, Intensive Care Unit admission, mechanical ventilation, WHO ordinal scale of clinical progression, adherence to study treatment, WHO-5 Well-Being Index,^27^ serious adverse events, all cause death or urgent, non-elective hospitalisation and reports of new household infections. All time to event analyses used date of randomisation as baseline. We included secondary outcomes that capture sustained recovery due to the often recurrent and relapsing nature of COVID-19 symptoms.

### Statistical analysis

Sample size calculation and statistical analysis are detailed in the Adaptive Design Report (appendix pp 130-230) and the Master Statistical Analysis Plan (appendix pp 81-129). In the Adaptive Design Report we justify sample sizes by simulating the operating characteristics of the adaptive design in multiple scenarios, which explicitly account for response adaptive randomisation, early stopping for futility/success and multiple interventions. In brief, for the primary outcome analyses, assuming a median time to recovery of nine days in the usual care group, approximately 400 participants per group would provide 90% power to detect a 2-day difference in median recovery time. Assuming 5% hospitalisation in the usual care group, approximately 1500 participants per group would provide 90% power to detect a 50% reduction in the relative risk of hospitalisation/death.

The first co-primary outcome, time to first self-reported recovery, was analysed using a Bayesian piecewise exponential model. The second co-primary outcome, hospitalisation/death, was analysed using a Bayesian logistic regression model. Both models were regressed on treatment group and stratification covariates (age < 65 years /≥ 65 years and comorbidity yes/no). These primary outcomes were evaluated using a “gate-keeping” strategy to preserve the overall Type I error without additional adjustments for multiple hypotheses. The hypothesis for the time-to-first-recovery endpoint was evaluated first, and if the null hypothesis was rejected, the hypothesis for the second co-primary endpoint of hospitalisation/death was evaluated. In the context of multiple interim analyses, the master protocol specifies that each null hypothesis is rejected if the Bayesian posterior probability of superiority exceeded 0.99 for the time to recovery endpoint and 0.975 (via gate-keeping) for the hospitalisation/death endpoint. For the purposes of defining futility rules, we pre-specified a clinically meaningful hazard ratio for time to first reported recovery as 1.2 or larger (equating to approximately 1.5 days difference in median time to recovery, assuming 9 days recovery in the usual care arm), and a clinically meaningful odds ratio as 0.80 or smaller for hospitalisations/deaths (equating to approximately a 1% decrease in the hospitalisation rate, assuming a rate of 5% in the usual care arm). If there is insufficient evidence of a clinically meaningful benefit in time to recovery, futility is declared and randomisation to that intervention is stopped, meaning other interventions can be evaluated more rapidly in the trial. For each primary outcome endpoint (time to recovery and hospitalisation/death), a model-based estimate of absolute benefit (days and percent, respectively) was obtained by applying the model-based estimate of treatment benefit (hazard ratio or odds ratio, respectively) to a bootstrap sample of the concurrent and eligible usual care population.

At the beginning of the trial, due to initial difficulties with community SARS-CoV-2 PCR testing in the UK, participants with suspected COVID-19 were included in the primary analysis population, irrespective of confirmatory testing. When testing became more accessible, the Trial Steering Committee recommended restricting the primary analysis population to those with confirmed COVID-19. This change was included in protocol version 7.1 on February 22, 2021 and approved on March 15, 2021, before any interim colchicine results were disclosed to the Trial Management Group. Therefore, the pre-specified primary analysis population includes all eligible SARS-CoV-2 positive participants randomised to colchicine, usual care, and other interventions, from the start of the platform trial until the colchicine arm was closed, on May 26, 2021. This population includes participants randomised to usual care before the colchicine group opened, who may differ from concurrently randomised participants because of changes in the inclusion/exclusion criteria (e.g. participants aged ≥18 years with comorbidity or shortness of breath became eligible when the colchicine group opened), and changes over time in the predominant variant and amount of circulating SARS-CoV-2 or usual care, including increasing availability of vaccinations. Therefore, the primary analysis models include parameters to adjust for potential temporal drift in the trial population, by estimating the primary endpoint in the usual care group across time via Bayesian hierarchical modelling.

We also conducted a key pre-specified sensitivity analysis of the primary outcomes using the concurrent randomised population; defined as all SARS-CoV-2 positive participants randomised during the time period when the colchicine arm was active. To determine the applicability of our results to situations where PCR testing may not be readily available, we also conducted secondary analyses of time to recovery and COVID-19 related hospitalisation/death among the overall study population, irrespective of SARS-CoV-2 status.

Analyses of all secondary outcomes, and pre-specified sub-group analyses, were conducted using SARS-CoV-2 positive participants eligible for colchicine, and concurrently randomised to colchicine or usual care; the concurrently randomised and eligible SARS-CoV-2 positive population. Secondary time-to-event outcomes were analysed using Cox proportional hazard models, and binary outcomes were analysed using logistic regression, adjusting for comorbidity, age, duration of illness and vaccination status. Due to the high proportion contributing to the analysis of primary outcomes (95%), we did not explore the potential impact of missing data. All model assumptions were evaluated. Analyses were conducted using R (version 4.0.3) and Stata (version 16.1).

### Patient and public involvement

We convened a patient and public involvement group of five women and two men who input into the patient-facing materials, choice of trial outcomes, and delivery plans. They were very supportive of the option of a trial partner to assist participants, and of our work to evaluate community treatments for COVID-19. Two public contributors serve on the Trial Steering Committee and continue to input on patient facing material, trial design and dissemination. Feedback from a UK wide survey of 291 PRINCIPLE participants showed that 90% found the study information prepared them for participation and 94% felt they were treated well by the study team. We made improvements in our communications about study procedures following this detailed feedback. PPI support has been forthcoming from a diverse range of ethnic minority communities for widespread promotion of the study, for example in community and faith meetings, and broadcast and print media.^20^

## Results

### Population

The first participant was randomised into PRINCIPLE on April 2, 2020. Enrolment into the colchicine group started on March 04, 2021. On May 26, 2021, the Trial Steering Committee advised the Trial Management Group to stop randomisation to colchicine because the pre-specified futility criterion had been met on time to recovery. Those participants taking colchicine at the time randomisation was stopped were advised that we had identified evidence for futility. All participants were followed up for the full 28 days.

4997 participants had been randomised of whom 212 were allocated to colchicine, 2081 to usual care alone, and 2704 to other treatments (Figure 1); 4221 of 4880 (86.5%) eligible participants had a SARS-CoV-2 test result available, of which 2900 (68.7%) tested positive. The Bayesian primary analysis model includes data from 2755 of 2900 (95.0%) SARS-CoV-2 positive participants who provided follow up data and were randomised to colchicine (n = 156), usual care alone (n = 1145), and other treatment groups (n = 1454). To protect the integrity of the platform trial and other interventions, we only provide descriptive summaries of participants randomised to colchicine and usual care. The average age (range) of participants was 61 (18 to 100) years, 1260 (91.2%) were White and 1165 (84.4%) had comorbidities. At randomisation, median time from symptom onset was 6 (interquartile range 4 to 9) days. Baseline characteristics were similar between the comparison groups (Tables 1 and S1). Data regarding inhaled corticosteroid was not consistently recorded early in the trial, but in the concurrent randomisation analysis population, 13/174 (7%) of the colchicine arm and 14/140 (10%) of the usual care arm reported taking inhaled corticosteroids at randomisation or during follow-up.

**Table 1.**
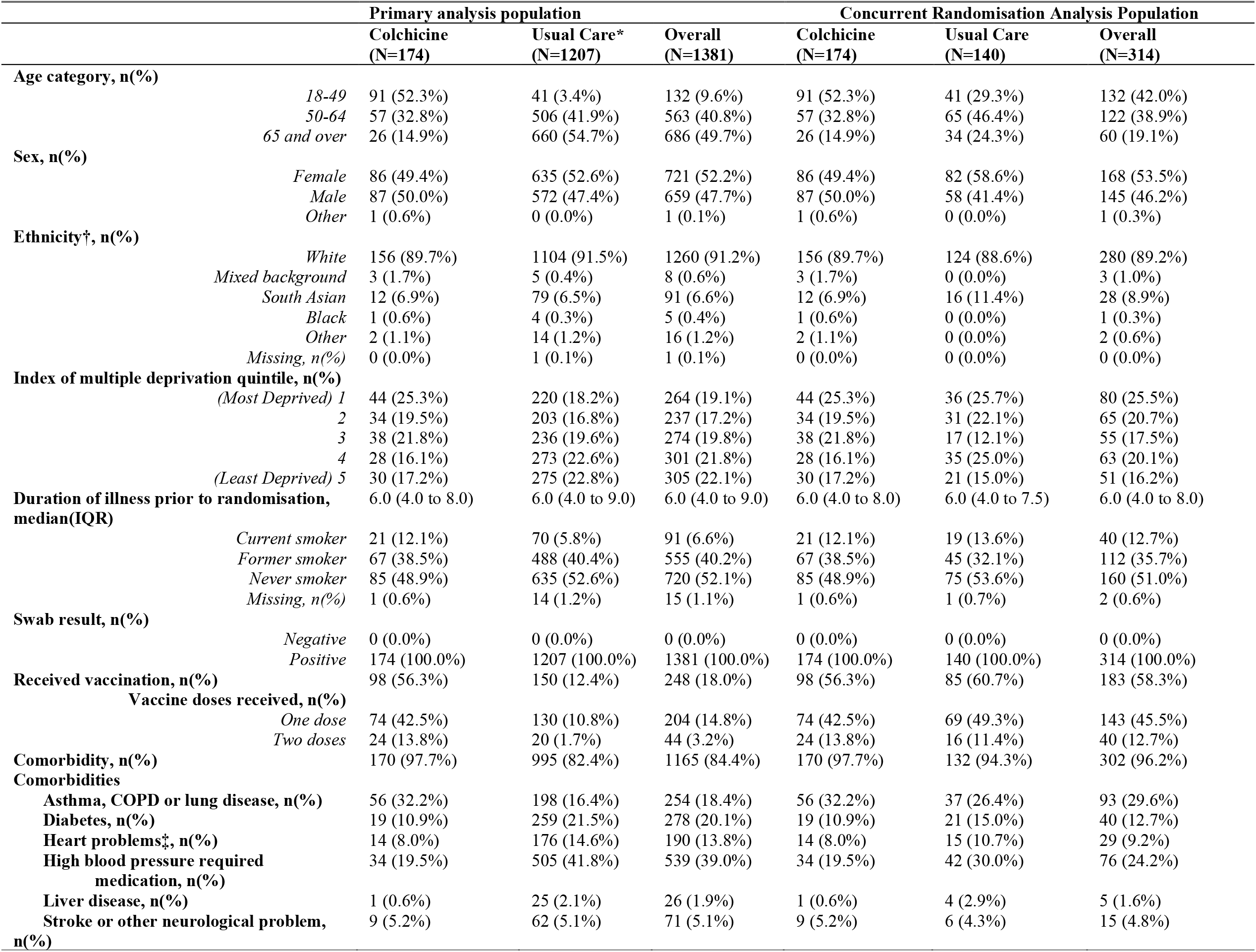
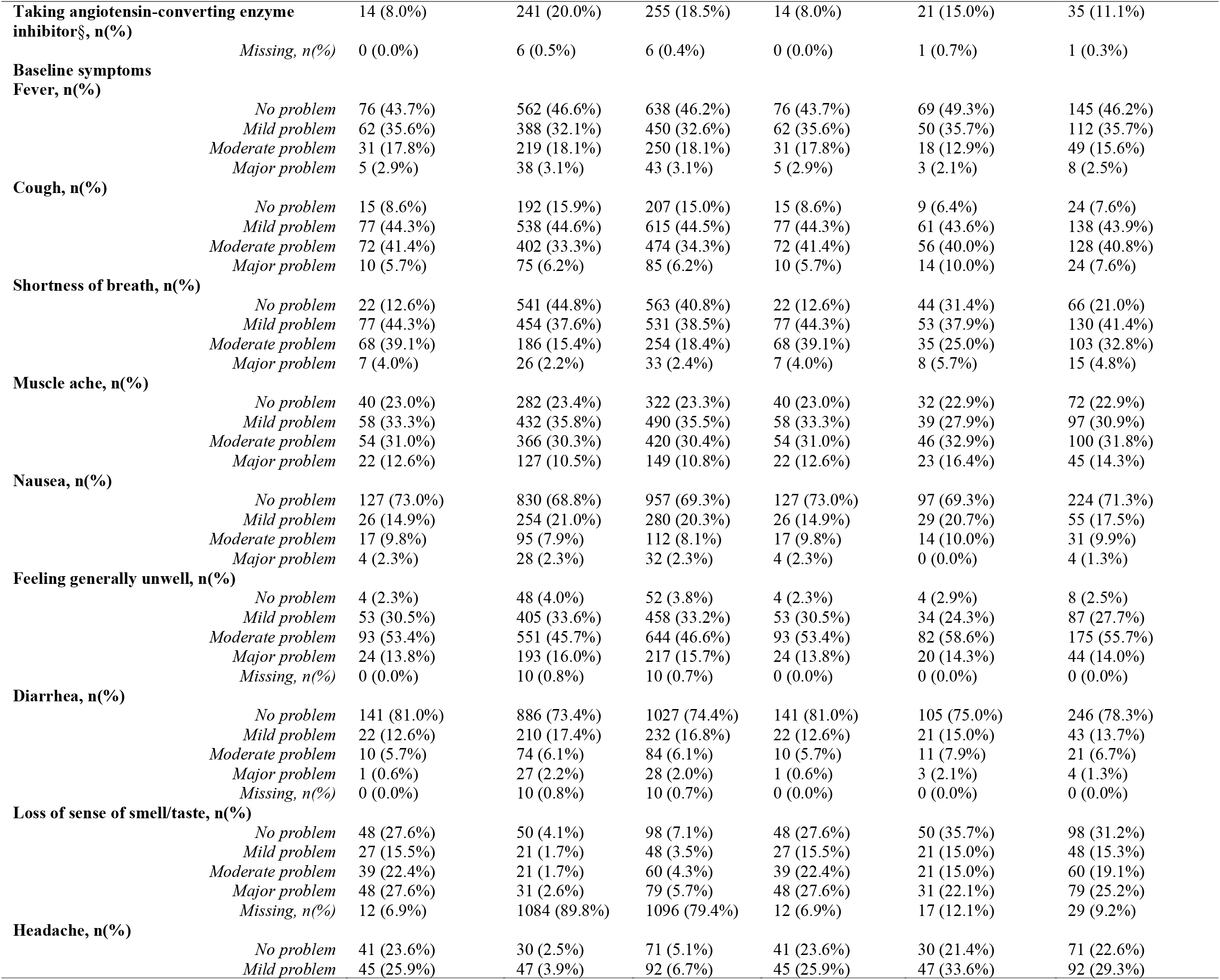
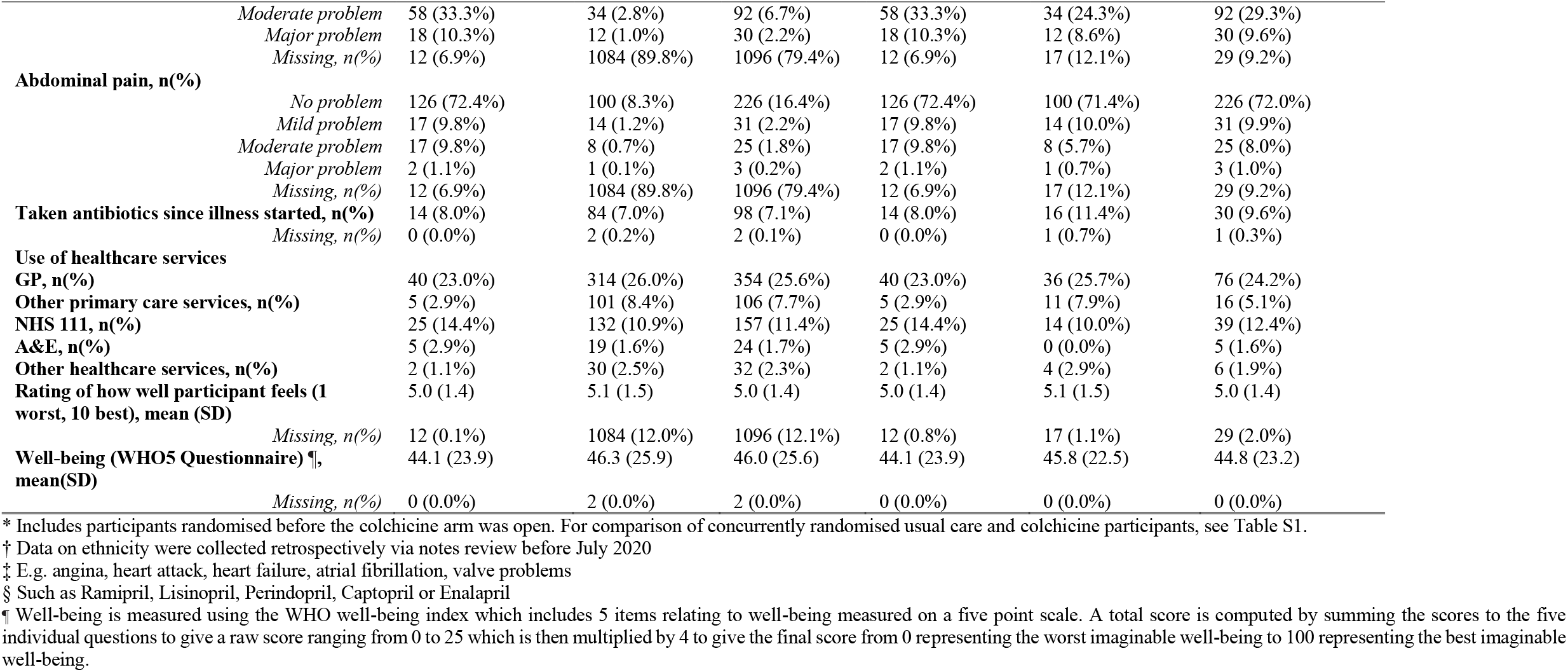
Baseline characteristics of SARS-CoV-2 positive participants by treatment group.

**Figure 1.**
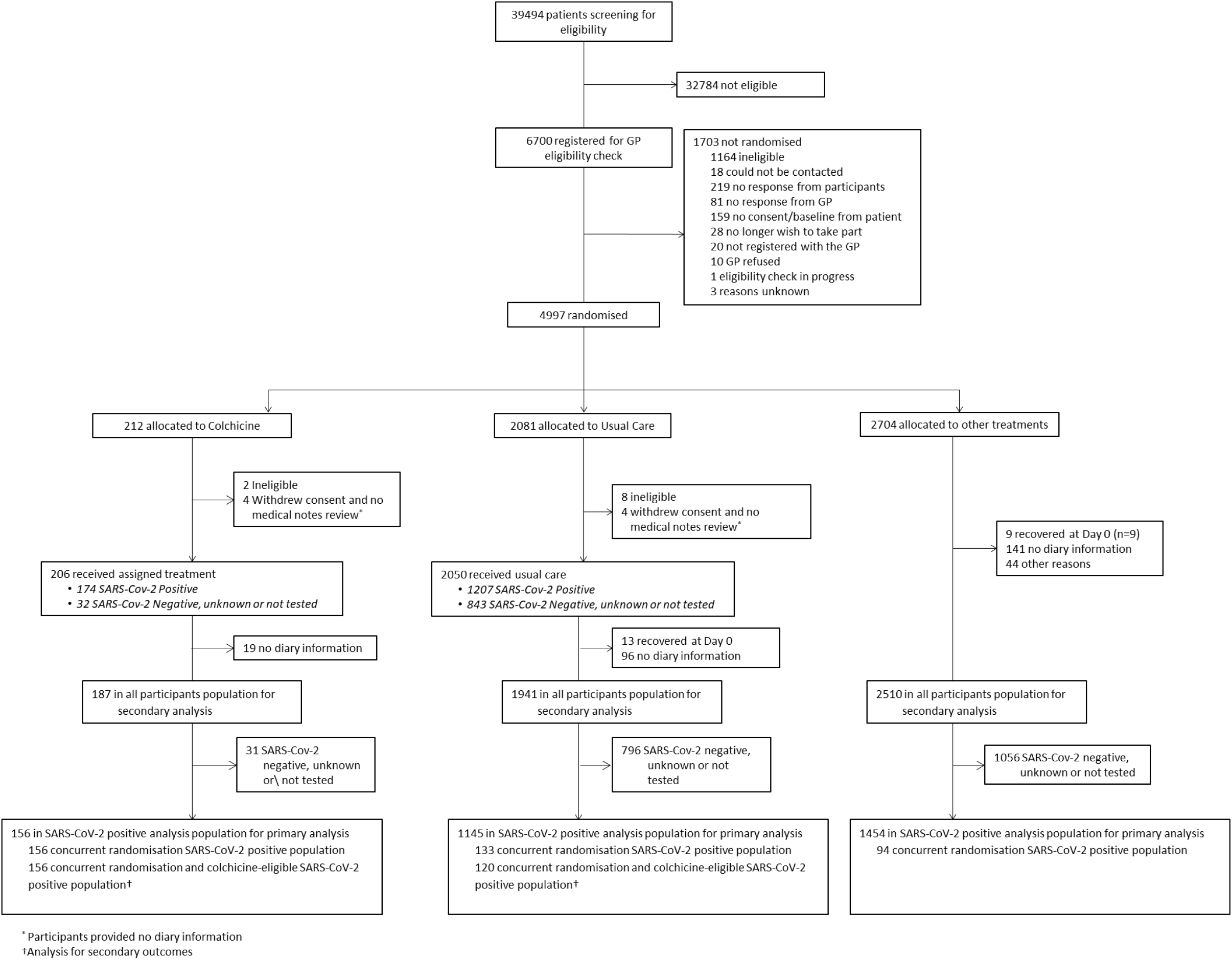
Participant flow diagram.

Of 184 participants randomised to colchicine who provided medication use information, 138 (75%) reported taking colchicine for at least seven days.

### Primary Outcomes

In the SARS-CoV-2 positive primary analysis population, the observed median time to first recovery was 15 days in the colchicine group compared to 14 in the usual care group (Figure 2a). In the concurrent randomisation analysis population, (excluding participants randomised to usual care before the colchicine arm opened) the observed median time to first recovery was 15 in the colchicine group and 14 in the usual care group. Based on the Bayesian primary analysis model which adjusts for temporal drift, there was no evidence of a benefit in time-to-first-recovery in the colchicine group versus usual care (hazard ratio 0.92, 95% Bayesian credible interval [0.72 to 1.16]. Based on a bootstrap estimated median time to recovery of 13 days in the concurrent and eligible usual care SARS-CoV-2 positive population, the model-based estimated hazards ratio corresponds to an estimated 1.14 (−1.86 to 5.21) additional days in median time to first reported recovery for colchicine relative to usual care. The probability that time to recovery was shorter in the colchicine group versus usual care (i.e. probability of superiority) was 0.241, which did not meet the pre-specified superiority threshold of 0.99. The probability of meaningful effect (pre-specified as a hazard ratio ≥1.2 for the purpose of evaluating futility) was 0.018 (Table 2). The low probability of a clinically meaningful treatment effect was consistent in the concurrent randomisation and overall study population (Table 2).

**Table 2:**
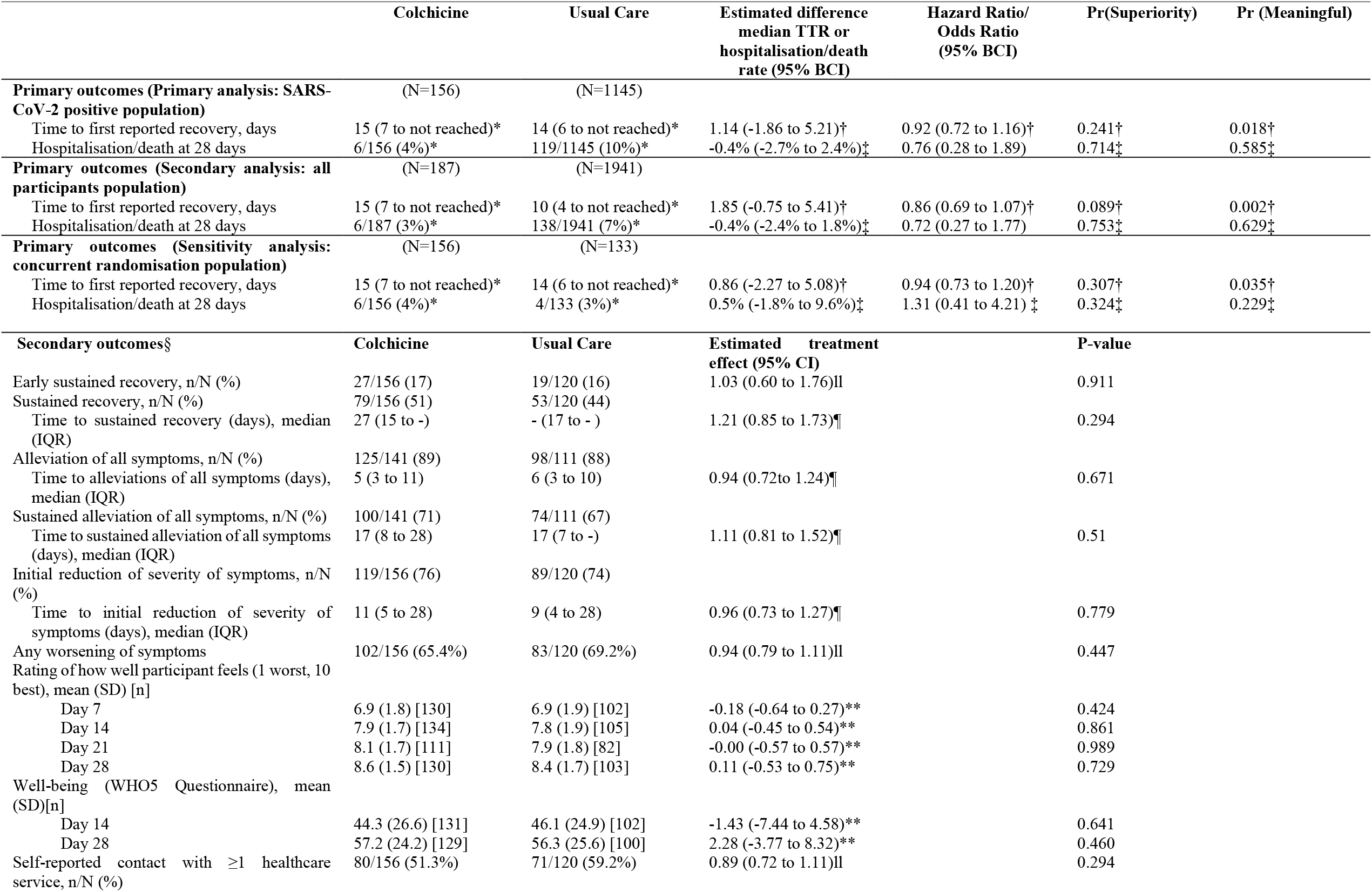
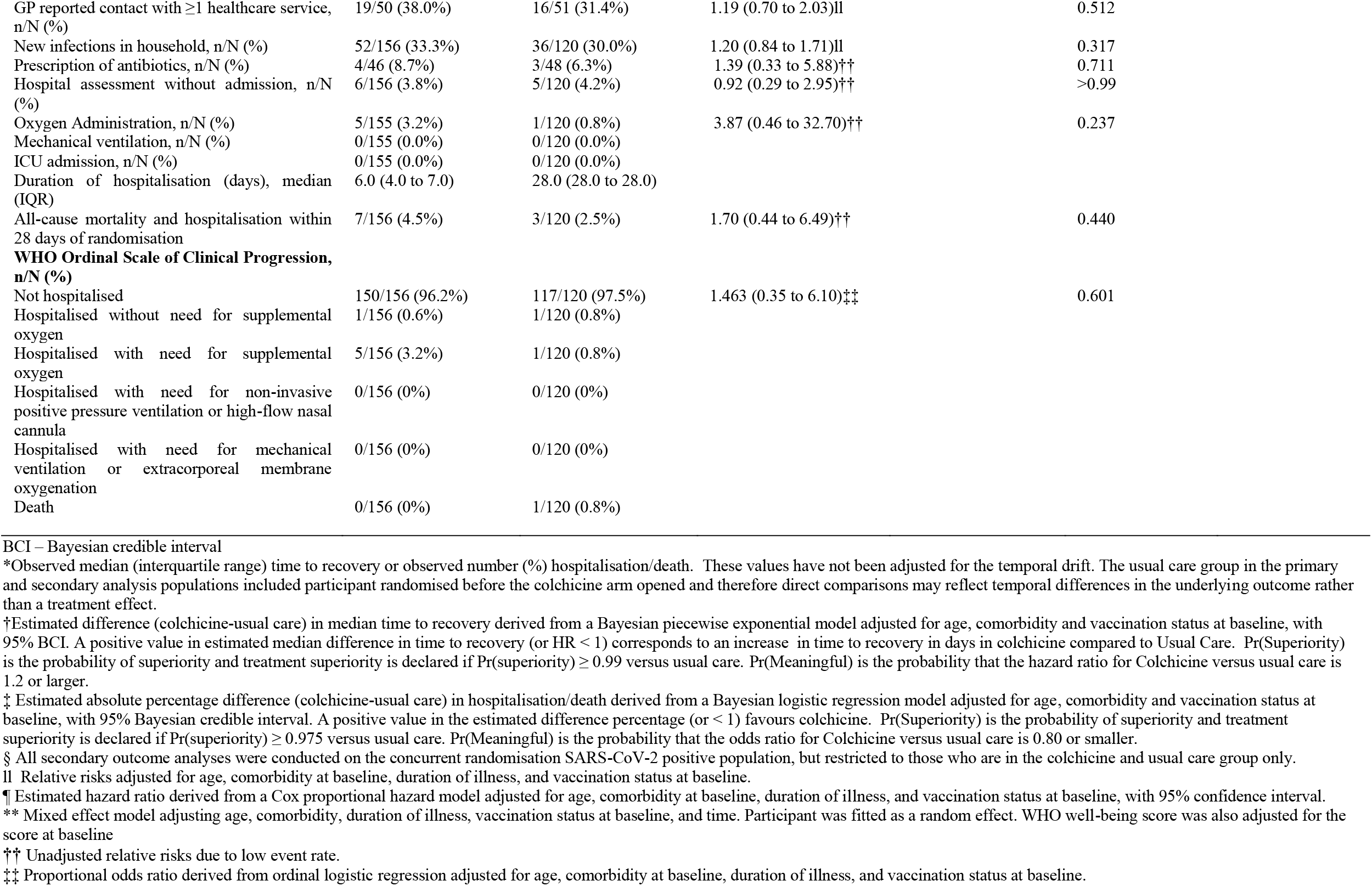
Primary and Secondary Outcomes.

|  | Colchicine | Usual Care | Estimated difference<br>median TTR or<br>hospitalisation/death<br>rate (95% BCI) | Hazard Ratio/<br>Odds Ratio<br>(95% BCI) | Pr(Superiority) | Pr (Meaningful) |
| --- | --- | --- | --- | --- | --- | --- |
| **Primary outcomes (Primary analysis: SARS-CoV-2 positive population)** | (N=156) | (N=1145) |  |  |  |  |
| Time to first reported recovery, days | 15 (7 to not reached)* | 14 (6 to not reached)* | 1.14 (-1.86 to 5.21)† | 0.92 (0.72 to 1.16)† | 0.241† | 0.018† |
| Hospitalisation/death at 28 days | 6/156 (4%)* | 119/1145 (10%)* | -0.4% (-2.7% to 2.4%)‡ | 0.76 (0.28 to 1.89) | 0.714‡ | 0.585‡ |
| **Primary outcomes (Secondary analysis: all participants population)** | (N=187) | (N=1941) |  |  |  |  |
| Time to first reported recovery, days | 15 (7 to not reached)* | 10 (4 to not reached)* | 1.85 (-0.75 to 5.41)† | 0.86 (0.69 to 1.07)† | 0.089† | 0.002† |
| Hospitalisation/death at 28 days | 6/187 (3%)* | 138/1941 (7%)* | -0.4% (-2.4% to 1.8%)‡ | 0.72 (0.27 to 1.77) | 0.753‡ | 0.629‡ |
| **Primary outcomes (Sensitivity analysis: concurrent randomisation population)** | (N=156) | (N=133) |  |  |  |  |
| Time to first reported recovery, days | 15 (7 to not reached)* | 14 (6 to not reached)* | 0.86 (-2.27 to 5.08)† | 0.94 (0.73 to 1.20)† | 0.307† | 0.035† |
| Hospitalisation/death at 28 days | 6/156 (4%)* | 4/133 (3%)* | 0.5% (-1.8% to 9.6%)‡ | 1.31 (0.41 to 4.21) ‡ | 0.324‡ | 0.229‡ |
| **Secondary outcomes§** | **Colchicine** | **Usual Care** | **Estimated treatment effect (95% CI)** |  | **P-value** |  |
| Early sustained recovery, n/N (%) | 27/156 (17) | 19/120 (16) | 1.03 (0.60 to 1.76)II |  | 0.911 |  |
| Sustained recovery, n/N (%) | 79/156 (51) | 53/120 (44) |  |  |  |  |
| Time to sustained recovery (days), median (IQR) | 27 (15 to -) | - (17 to -) | 1.21 (0.85 to 1.73)¶ |  | 0.294 |  |
| Alleviation of all symptoms, n/N (%) | 125/141 (89) | 98/111 (88) |  |  |  |  |
| Time to alleviations of all symptoms (days), median (IQR) | 5 (3 to 11) | 6 (3 to 10) | 0.94 (0.72to 1.24)¶ |  | 0.671 |  |
| Sustained alleviation of all symptoms, n/N (%) | 100/141 (71) | 74/111 (67) |  |  |  |  |
| Time to sustained alleviation of all symptoms (days), median (IQR) | 17 (8 to 28) | 17 (7 to -) | 1.11 (0.81 to 1.52)¶ |  | 0.51 |  |
| Initial reduction of severity of symptoms, n/N (%) | 119/156 (76) | 89/120 (74) |  |  |  |  |
| Time to initial reduction of severity of symptoms (days), median (IQR) | 11 (5 to 28) | 9 (4 to 28) | 0.96 (0.73 to 1.27)¶ |  | 0.779 |  |
| Any worsening of symptoms | 102/156 (65.4%) | 83/120 (69.2%) | 0.94 (0.79 to 1.11)II |  | 0.447 |  |
| Rating of how well participant feels (1 worst, 10 best), mean (SD) [n] |  |  |  |  |  |  |
| Day 7 | 6.9 (1.8) [130] | 6.9 (1.9) [102] | -0.18 (-0.64 to 0.27)** |  | 0.424 |  |
| Day 14 | 7.9 (1.7) [134] | 7.8 (1.9) [105] | 0.04 (-0.45 to 0.54)** |  | 0.861 |  |
| Day 21 | 8.1 (1.7) [111] | 7.9 (1.8) [82] | -0.00 (-0.57 to 0.57)** |  | 0.989 |  |
| Day 28 | 8.6 (1.5) [130] | 8.4 (1.7) [103] | 0.11 (-0.53 to 0.75)** |  | 0.729 |  |
| Well-being (WHO5 Questionnaire), mean (SD)[n] |  |  |  |  |  |  |
| Day 14 | 44.3 (26.6) [131] | 46.1 (24.9) [102] | -1.43 (-7.44 to 4.58)** |  | 0.641 |  |
| Day 28 | 57.2 (24.2) [129] | 56.3 (25.6) [100] | 2.28 (-3.77 to 8.32)** |  | 0.460 |  |
| Self-reported contact with ≥1 healthcare service, n/N (%) | 80/156 (51.3%) | 71/120 (59.2%) | 0.89 (0.72 to 1.11)II |  | 0.294 |  |

|  |  |  |  |  |
| --- | --- | --- | --- | --- |
| GP reported contact with $\geq 1$ healthcare service, n/N (%) | 19/50 (38.0%) | 16/51 (31.4%) | 1.19 (0.70 to 2.03)<sup>II</sup> | 0.512 |
| New infections in household, n/N (%) | 52/156 (33.3%) | 36/120 (30.0%) | 1.20 (0.84 to 1.71)<sup>II</sup> | 0.317 |
| Prescription of antibiotics, n/N (%) | 4/46 (8.7%) | 3/48 (6.3%) | 1.39 (0.33 to 5.88)<sup>††</sup> | 0.711 |
| Hospital assessment without admission, n/N (%) | 6/156 (3.8%) | 5/120 (4.2%) | 0.92 (0.29 to 2.95)<sup>††</sup> | >0.99 |
| Oxygen Administration, n/N (%) | 5/155 (3.2%) | 1/120 (0.8%) | 3.87 (0.46 to 32.70)<sup>††</sup> | 0.237 |
| Mechanical ventilation, n/N (%) | 0/155 (0.0%) | 0/120 (0.0%) |  |  |
| ICU admission, n/N (%) | 0/155 (0.0%) | 0/120 (0.0%) |  |  |
| Duration of hospitalisation (days), median (IQR) | 6.0 (4.0 to 7.0) | 28.0 (28.0 to 28.0) |  |  |
| All-cause mortality and hospitalisation within 28 days of randomisation | 7/156 (4.5%) | 3/120 (2.5%) | 1.70 (0.44 to 6.49)<sup>††</sup> | 0.440 |
| **WHO Ordinal Scale of Clinical Progression, n/N (%)** |  |  |  |  |
| Not hospitalised | 150/156 (96.2%) | 117/120 (97.5%) | 1.463 (0.35 to 6.10)<sup>‡‡</sup> | 0.601 |
| Hospitalised without need for supplemental oxygen | 1/156 (0.6%) | 1/120 (0.8%) |  |  |
| Hospitalised with need for supplemental oxygen | 5/156 (3.2%) | 1/120 (0.8%) |  |  |
| Hospitalised with need for non-invasive positive pressure ventilation or high-flow nasal cannula | 0/156 (0%) | 0/120 (0%) |  |  |
| Hospitalised with need for mechanical ventilation or extracorporeal membrane oxygenation | 0/156 (0%) | 0/120 (0%) |  |  |
| Death | 0/156 (0%) | 1/120 (0.8%) |  |  |

BCI – Bayesian credible interval
\*Observed median (interquartile range) time to recovery or observed number (%) hospitalisation/death. These values have not been adjusted for the temporal drift. The usual care group in the primary and secondary analysis populations included participant randomised before the colchicine arm opened and therefore direct comparisons may reflect temporal differences in the underlying outcome rather than a treatment effect.
†Estimated difference (colchicine-usual care) in median time to recovery derived from a Bayesian piecewise exponential model adjusted for age, comorbidity and vaccination status at baseline, with 95% BCI. A positive value in estimated median difference in time to recovery (or HR < 1) corresponds to an increase in time to recovery in days in colchicine compared to Usual Care. Pr(Superiority) is the probability of superiority and treatment superiority is declared if Pr(superiority) $\geq 0.99$ versus usual care. Pr(Meaningful) is the probability that the hazard ratio for Colchicine versus usual care is 1.2 or larger.
‡ Estimated absolute percentage difference (colchicine-usual care) in hospitalisation/death derived from a Bayesian logistic regression model adjusted for age, comorbidity and vaccination status at baseline, with 95% Bayesian credible interval. A positive value in the estimated difference percentage (or < 1) favours colchicine. Pr(Superiority) is the probability of superiority and treatment superiority is declared if Pr(superiority) $\geq 0.975$ versus usual care. Pr(Meaningful) is the probability that the odds ratio for Colchicine versus usual care is 0.80 or smaller.
§ All secondary outcome analyses were conducted on the concurrent randomisation SARS-CoV-2 positive population, but restricted to those who are in the colchicine and usual care group only.
II Relative risks adjusted for age, comorbidity at baseline, duration of illness, and vaccination status at baseline.
¶ Estimated hazard ratio derived from a Cox proportional hazard model adjusted for age, comorbidity at baseline, duration of illness, and vaccination status at baseline, with 95% confidence interval.
\*\* Mixed effect model adjusting age, comorbidity, duration of illness, vaccination status at baseline, and time. Participant was fitted as a random effect. WHO well-being score was also adjusted for the score at baseline
†† Unadjusted relative risks due to low event rate.

**Figure 2.**
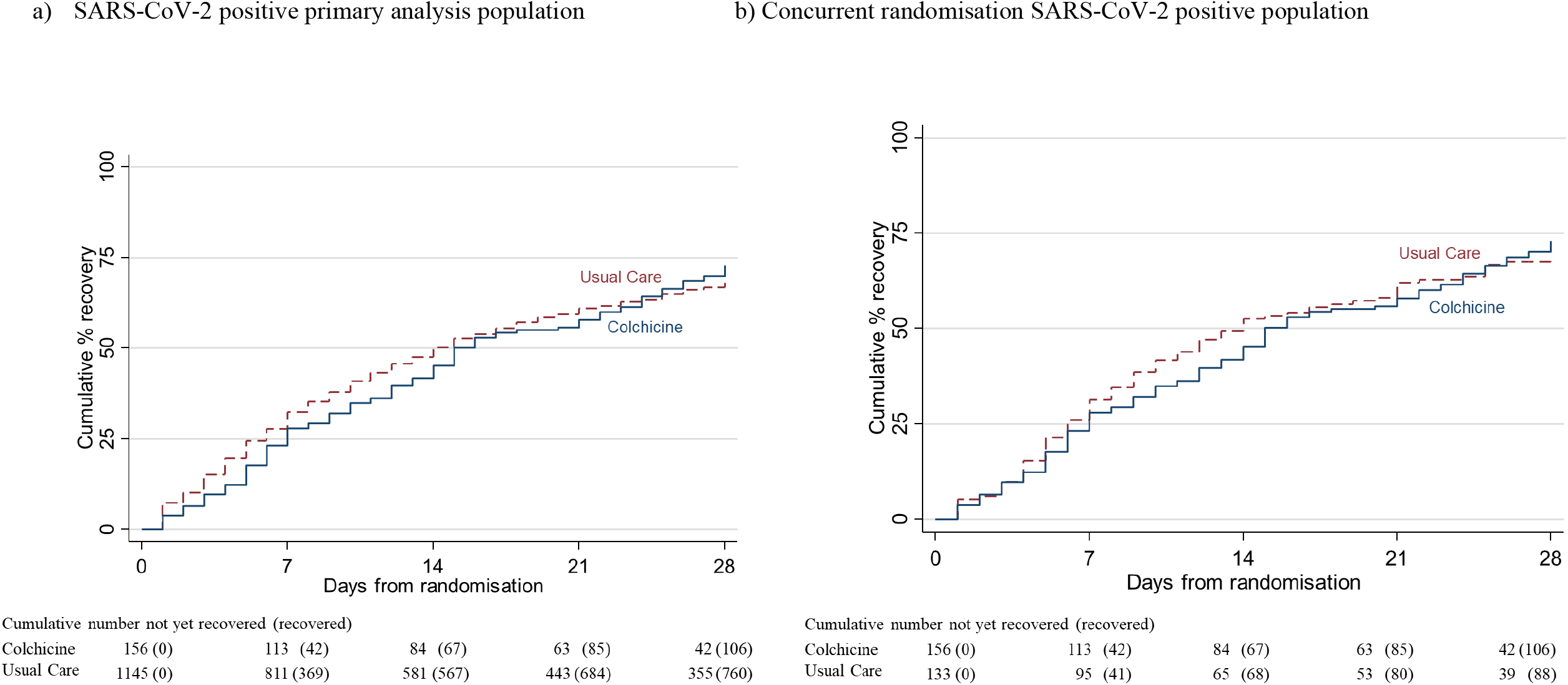
Time to first reported recovery.

In the SARS-CoV-2 positive primary analysis population, there were 6/156 (3.8%) COVID-19 related hospitalisations/deaths in the colchicine group (six hospitalisations, no deaths), and 119/1145 (10.4%) in the usual care group (116 hospitalisations, of whom nine died, and three deaths without hospitalisation). The high levels of hospitalisations/deaths in the usual care group in the primary analysis population were driven by the high event rate before the colchicine arm opened. In the usual care group in concurrent randomisation analysis population, which excluded participants randomised to usual care before the colchicine arm opened, there were 4/133 (3.0%) COVID-19 related hospitalisations/deaths in the usual care group (three hospitalisations, and one death without hospitalisation). In the Bayesian primary analysis model, which takes into account the temporal change in event rates, COVID-19 related hospitalisation/deaths in the colchicine group compared to usual care were similar, with an estimated odds ratio of 0.76 (95% credible interval 0.28 to 1.89). Based on a bootstrap estimated hospitalization rate of 2.5% in the concurrent and eligible usual care population, the model-based estimated odds ratio corresponds to an estimated difference in the hospitalisation rate of −0.4% [−2.7% to 2.4%]) (Table 2). The probability that COVID-19 related hospitalisations/deaths were lower in the colchicine arm versus usual care (i.e. probability of superiority) was 0.714 and because superiority was not reached on time to recovery, the hospitalisation/death outcome was not formally analysed for significance due to the gate-keeping hypothesis structure. The probability that there was a meaningful reduction in COVID-19 related hospitalisations/deaths (predefined as an odds ratio of 0.80 or smaller) was 0.585. The point estimates of the model-based effects on hospitalisations were not consistent across the primary analysis population (odds ratio = 0.76 [95% BCI: 0.28,1.89)]), the concurrent randomisation population (odds ratio = 1.31 [95% BCI: 0.41,4.21]), and the overall study population (odds ratio = 0.72 [95% BCI: 0.27,1.77]). This is due to the very small number of events observed in the concurrent analysis population and corresponds to wide credible intervals. The general conclusion is consistent across these populations, in that there is lack of evidence of a clinically meaningful treatment effect on hospitalisations/deaths (Table 2).

### Secondary outcomes

Analyses of secondary outcomes, using the concurrent randomisation and eligible SARS-CoV-2 positive population, are presented in Table 2, Figures S1, S2, S3b and S3c. There was no clear evidence of benefit for any of the secondary outcomes.

In the pre-specified subgroup analyses, there was no strong statistical evidence that symptom duration prior to randomisation, baseline illness severity score, inhaled corticosteroid use, age or comorbidity modified the effect of colchicine on time to first reported recovery (Figure 3), although numbers were small. In post-hoc sub-group analyses, there was no evidence that colchicine effects differed by vaccination status (Figure 3), although numbers were small. Event rates were too low to conduct sub-group analyses of the hospitalisation/death outcome. Regarding serious adverse events, there was one hospitalisation unrelated to COVID-19 in the colchicine group and one in usual care.

**Figure 3.**
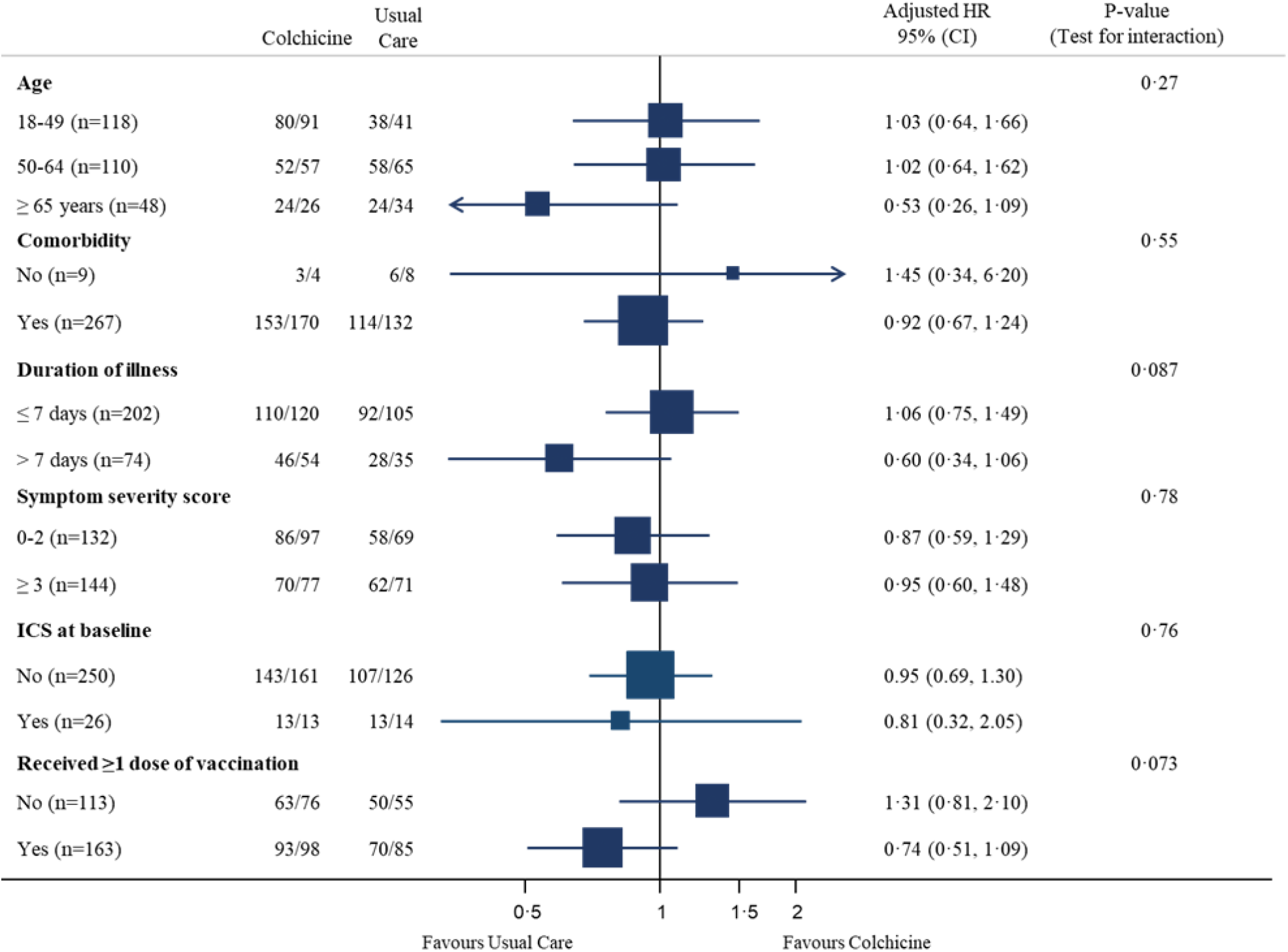
Forest plot of subgroup analysis of time to first reported recovery (concurrent randomisation and eligible SARS-CoV-2 positive population)

## Discussion

### Principal findings

This analysis from a platform, randomised trial involving people in the community with COVID-19 found that colchicine did not meaningfully improve time to first recovery compared with usual care alone. There was also no evidence of a difference in secondary measures of wellbeing, sustained recovery, symptoms or healthcare service use. In line with the protocol, due to futility reaching the futility criterion on time to recovery, the colchicine arm was stopped prior to collecting substantial data on hospitalisations/deaths, meaning the estimates of effect on hospitalisation had wide confidence intervals. Overall, these findings do not support the use of colchicine as treatment for the symptoms of COVID-19 in the community.

### Strengths and weaknesses

Strengths include the pragmatic design of the PRINCIPLE trial which allowed for efficient evaluation of the effectiveness of colchicine as an early, standalone intervention as it might be used in the community. We focused on patients at increased risk of complications and used routine electronic health records to confirm hospitalisation/death, and obtained primary outcome data on over 95% of participants. Although our primary analysis was restricted to SARS-CoV-2 positive patients, we conducted secondary analyses of the co-primary outcomes among patients with suspected COVID-19 but without PCR confirmed SARS-CoV-2 infection, as limited SARS-CoV-2 testing may necessitate early empirical treatment in low resource settings. Furthermore, variation in PCR testing sensitivity, particularly if self-administered, means some participants will have had false negative tests.^28^ Time to recovery estimates were similar in the SARS-CoV-2 positive population, all participants irrespective of SARS-CoV-2 status, as well as the concurrent randomisation SARS-CoV-2 positive population (the latter populations are most analogous to those in traditional two arm trials).

Although the sample size of the colchicine group in PRINCIPLE was relatively small, the Bayesian primary analysis model leverages previous enrolments in the usual care arm to increase the precision of estimates, which allow us to declare futility with high precision due to very low probability of a meaningful benefit of colchicine on time to recovery. This allowed the colchicine arm to be dropped and for trial resources to be rapidly redirected to other interventions, with the ultimate aim of identifying effective treatments for COVID-19 in the community, attesting to the efficiency of the trial design in keeping with the aim of rapid generation of evidence for use within the pandemic itself. We used a pragmatic, open label design, similar to other large COVID-19 platform trials,^29,30^ to evaluate the addition of colchicine to usual care, rather than to assess benefit of colchicine compared to a placebo. If a positive placebo effect influenced our self-reported time to recovery outcome, it would likely be masking even greater negative effect of colchicine. We used this outcome as it was of greatest interest to our patient and public contributors and is best ascertained by direct patient report, rather than by the use of surrogate measures. We hypothesised that a treatment that does not reduce recovery time is also unlikely to reduce COVID-19 related hospitalisations/death. However, it is possible for a treatment to reduce the likelihood of severe disease without reducing duration of the illness.

### Strengths and weaknesses in relation to other studies

PRINCIPLE is the first randomised trial to evaluate the effect of colchicine on time to recovery for people with COVID-19 in the community. Our finding of no benefit contrasts with some of the findings of the COLCORONA trial, which was terminated early at approximately 75% of targeted recruitment and which found a possible reduction in hospitalisations/deaths with colchicine, although time to recovery was not measured.^9^ The primary endpoint of COVID-19 related hospitalisation or death occurred in 104/2235 (4.7%) in the colchicine group and 131/2253 (5.8%) in the placebo group (odds ratio 0.79; 95.1% confidence interval (CI) 0.61 to 1.03; P=0.08). In the 4159 SARS-CoV-2 positive patients, the primary endpoint was lower in the colchicine arm compared to placebo (4.6% versus 6.0%; odds ratio 0.75; 95% CI, 0.57 to 0.99; P=0.04). Diarrhoea (13.7% versus 7.3%), gastrointestinal adverse events (23.9% versus 14.8%) and pulmonary emboli (0.5% versus 0.1%) were higher in the colchicine group compared to placebo. Participants in the COLCORONA trial were slightly older than PRINCIPLE (median age in colchicine arm 53 vs 48 years). Taking into account the pharmacokinetic variability and narrow therapeutic index of colchicine, we selected a moderate dose with no loading dose in PRINCIPLE. The loading dose used in the COLCORONA study may mean intracellular concentrations rose faster and may explain the difference in clinical effect, including the higher proportion of gastrointestinal adverse events and diarrhoea seen in the COLCORONA colchicine arm. We did not find evidence that colchicine led to a worsening of any symptoms, although there was a trend towards a longer duration of diarrhoea symptoms. In the RECOVERY trial among 11,340 people hospitalised with COVID-19, there was no evidence that colchicine improved the primary outcome of 28-day mortality.^8^

### Implications and future research

While the COLCORONA trial found limited evidence that colchicine may reduce COVID-19 related hospitalisation/death,^9^ most treatment guidelines have not recommended colchicine’s use in the community, potentially because of its increased gastrointestinal side effect profile and the observed higher rates of pulmonary embolism. Our results support this position; we found no evidence that colchicine shortens time to recovery among patients with COVID-19 in the community. Therefore, with the evidence available to date, colchicine should neither be used for treatment of COVID-19 in community nor hospital settings. We are planning analyses of the effect of colchicine on longer term symptoms related to “long COVID”.

## Conclusion

Colchicine does not improve time to recovery for adults with COVID-19 in the community, and there was no evidence for a benefit in COVID-19 related hospitalisations/deaths but estimates were imprecise due to the few hospitalisations.

## Summary box

### What is already known on this topic

- Colchicine has been proposed as treatment for COVID-19 due to its anti-inflammatory properties, but evidence to support its use is inconclusive, and its effect on time to recovery in the community has not been evaluated.
- The RECOVERY trial found no benefit with colchicine use among people hospitalised with COVID-19, while the COLCORONA trial found some evidence of a 1.1% and 1.4% absolute reduction in hospitalisations/deaths among adults with suspected or confirmed COVID-19 in the community respectively.

### What this study adds

- In this national, platform adaptive randomised controlled trial, we found evidence of no meaningful benefit with colchicine on time to recovery, and because the threshold for futility on time to recovery was met, randomisation to colchicine was stopped before collecting substantial data on hospitalisations and death, leading to imprecise estimates for that outcome.
- Our findings add to the evidence currently available and suggest that colchicine should not be recommended for treating symptoms of COVID-19.

## Supporting information

CONSORT checklist

## Data Availability

Data can be shared with qualifying researchers who submit a proposal with a valuable research question as assessed by a committee formed from the TMG including senior statistical and clinical representation. A contract should be signed.

## Ethics statements

The study conformed to the ethical guidelines of the 1975 Declaration of Helsinki and was approved by the UK Medicines and Healthcare products Regulatory Agency and the South Central-Berkshire Research Ethics Committee (Ref: 20/SC/0158). Online consent was obtained from all participants.

## Writing group contributions

FRDH and CCB had full access to all of the data in the study, take responsibility for the conduct of the study, decided to publish the paper attests that all writing group members meet authorship criteria and that no others meeting the criteria have been omitted. BS, NB, L-MY, CCB, FDRH, GH, MS, OVH, OG, JD, DR contributed to trial design. SdeL, PHE, NT and MPG helped plan the trial and ongoing recruitment. EO, NT, SdeL, were responsible for acquisition of data. CCB, L-MY, JD, BS, MB, FDRH, GH, OVH and OG drafted the manuscript. BS, NB, L-MY, MAD, MF, CS, VH, and MS contributed to statistical analysis. All members of the PRINCIPLE writing group critically revised the manuscript. The members of the PRINCIPLE Collaborative Group and their roles in the conduct of the trial are listed at the end of the manuscript.

## Acknowledgments

We thank the patients who participated in this study. We also thank the many health, and social care professionals and who contributed. The PRINCIPLE trial platform is led from the Primary Care and Vaccines Collaborative Clinical Trials Unit at the University of Oxford’s Nuffield Department of Primary Care Health Sciences. PRINCIPLE is supported by a large network of care homes, pharmacies, NHS 111 Hubs, hospitals, and 1,401 GP practices across England, Wales, Scotland, and Northern Ireland. The trial is integrated with the Oxford-Royal College of General Practitioners (RCGP) Research and Surveillance Centre (RSC) ORCHID digital platform. PRINCIPLE has been supported by the NIHR and its Clinical Research Network, NHS DigiTrials, Public Health England, Health and Care Research Wales, NHS Research Scotland, the Health and Social Care Board in Northern Ireland, and the Therapeutics Task Force.

CCB acknowledges part support as Senior Investigator of the National Institute of Health Research, the NIHR Community Healthcare Medtech and In-Vitro Diagnostics Co-operative (MIC), and the NIHR Health Protection Research Unit on Health Care Associated Infections and Antimicrobial Resistance. FDRH acknowledges his part support as an NIHR Senior Investigator, as Director, NIHR Applied Research Collaboration (ARC) Oxford Thames Valley, as Theme Lead, NIHR Oxford Biomedical Research Centre (BRC, UHT), and the NIHR Community Healthcare Medtech and In-Vitro Diagnostics Co-operative (MIC). JD and OG are funded by the Wellcome Trust PhD Programme for Primary Care Clinicians (216421/Z/19/Z and 203921/Z/16/Z respectively). GH is funded by an NIHR Advanced Fellowship and by the NIHR Community Healthcare Medtech and In-Vitro Diagnostics Co-operative (MIC). For the purpose of Open Access, the author has applied a CC BY public copyright license to any Author Accepted Manuscript version arising from this submission.

