## Supplementary material for "Colchicine for COVID-19 in adults in the community (PRINCIPLE): a randomised, controlled, adaptive platform trial"

### Supplementary Appendix Content

### PRINCIPLE COLLABORATIVE GROUP

#### PRINCIPLE Trial Management Group (TMG)

Julie Allen[1], Monique Andersson[2], Nick Berry[3*], Emily Bongard[1], Aleksandra Borek[1], Christopher C Butler[1] (Chair), Simon de Lusignan[1], Jienchi Dorward[1,4], Philip H Evans[5,6], Filipa Ferreira[1], Oghenekome Gbinigie[1], Jenna Grabey[1], Gail Hayward[1], FD Richard Hobbs[1], , David Judge[1], Mona Koshkouei[1], Emma Ogburn[1], Mahendra G. Patel[1], Dan Richards-Doran[1], Heather Rutter[1], Benjamin R. Saville[2,7], Hannah Swayze[1], Nicholas PB Thomas[5,8], Manasa Tripathy[1], Sarah Tonkin-Crine[1,9], Jared Robinson[1], Sharon Tonner[1], Oliver Van Hecke[1], Ly-Mee Yu[1]
* TMG membership ended when NB became unblinded and joined the SAC

##### Trial Management Group affiliations

1. Nuffield Department of Primary Care Health Sciences, University of Oxford, Oxford, UK
2. Department of Microbiology, Oxford University Hospitals NHS Trust, Oxford, UK
3. Berry Consultants, Texas, USA,
4. Centre for the AIDS Programme of Research in South Africa (CAPRISA), University of KwaZulu–Natal, Durban, South Africa.
5. National Institute for Health Research, Clinical Research Network
6. College of Medicine and Health, University of Exeter
7. Department of Biostatistics, Vanderbilt University School of Medicine, Tennessee, USA
8. Royal College of General Practitioners, London, UK
9. National Institute for Health Protection Research Unit in Healthcare Associated Infections and Antimicrobial Resistance, Oxford, UK

##### Statistical Analysis Committee

Nick Berry, Michelle A. Detry (Chair), Christina Saunders, Mark Fitzgerald

##### Trial Steering Committee

Carol Green, Phil Hannaford, Paul Little (Chair), Tim Mustill, Matthew Sydes

##### Data Monitoring and Safety Committee

Deborah Ashby (Chair), Nick Francis, Simon Gates, Gordon Taylor, Patrick White

##### Principle Trial Coordinating Office

*Co-Chief Investigators:* CC Butler, FDR Hobbs

*Trial management:* E Ogburn (coordinator), H Swayze, E Bongard, J Allen, S Tonner, R Edeson, J Brooks, R Edwards, N Maeder, S Barrett, S Brann, A Maloney, K Dempster, J de Henau, J Robinson, N Begum, Micheal McKenna, Clare Bateman, Rebecca Lowe

*Clinical Team:* H Rutter (Coordinator), K Madronal, B Mundy, B Ianson, I Noel, B Thompson, O Gbinigie, J Dorward, G Hayward, O van Hecke, N Jones, H van der Westhuizen, K Kotze

*Data and Programming Team:* D Judge, J Grabey, L Castello, D Watt, R Zhao

*Statistics:* B Saville, L-M Yu, N Berry, M Detry, V Harris, C Saunders, M Fitzgerald, J Mollison, M Shanyinde

*Oxford-Royal College of General Practitioner's Research and Surveillance Centre:* S de Lusignan (coordinator) M Tripathy, F Ferreira

*National Institute for Health Research Clinical Research Network Coordinating Centre:* PH Evans, NPB Thomas, H Collins, K Priddis, L Owen, K Hannaby, B Drew

### APPENDIX 2

**Trial Title:** **P**latform **R**andomised tr**I**al of treatme**N**ts in the **C**ommunity for ep**I**demic and **P**andemic i**L**ln**E**sses

**Internal Reference Number / Short title:** PRINCIPLE
**Ethics Ref:** 20/SC/0158
**IRAS Project ID:** 281958
**EudraCT Number**: 2020-001209-22
**Date and Version No**: 12th April 2021 version 8.1

| | |
|---|---|
| **Chief Investigator and trial leader:** | Professor Chris Butler, Department of Primary Care Health Sciences University of Oxford |
| **Co-Principal Investigator and Co-trial lead:** | Prof Richard Hobbs, Department of Primary Care Health Sciences University of Oxford |
| **Co-Principal Investigators:** | Prof Simon de Lusignan, RCGP Research Surveillance Centre, University of Oxford<br>Prof Gail Hayward, Department of Primary Care Health Sciences University of Oxford |
| **Investigators:** | Dr Ly-Mee Yu, Primary Care Clinical Trials Unit, Department of Primary Care Health Sciences, University of Oxford<br>Dr Emma Ogburn, Primary Care Clinical Trials Unit, Department of Primary Care Health Sciences, University of Oxford<br>Dr Oliver Van Hecke, Department of Primary Care Health Sciences, University of Oxford<br>Ms Julie Allen, Primary Care Clinical Trials Unit, Department of Primary Care Health Sciences, University of Oxford<br>Dr Emily Bongard, Primary Care Clinical Trials Unit, Department of Primary Care Health Sciences, University of Oxford<br>Dr Hannah Swayze, Primary Care Clinical Trials Unit, Department of Primary Care Health Sciences, University of Oxford<br>Ben Saville, PhD, Berry Consultants, Texas, USA, & Department of Biostatistics, Vanderbilt University School of Medicine, Tennessee, USA<br>Prof Martin Llewellyn, Professor in Infectious Diseases, Medical Research Building, Room 1.08, BSMS, University of Sussex<br>Dr Oghenekome Gbinigie, Department of Primary Care Health Sciences University of Oxford<br>Dr Jienchi Dorward, Department of Primary Care Health Sciences University of Oxford |

© Copyright: The University of Oxford and Oxford University Hospitals NHS Foundation Trust 2019

Dr Monique Andersson, Oxford University Hospital NHS Trust

Dr Susan Hopkins, Incident Director for COVID-19, Public Health England

Dr Sarah Tonkin Crine, Department of Primary Care Health Sciences, University of Oxford

Dr Aleksandra Borek, Department of Primary Care Health Sciences, University of Oxford

Prof. Mahendra G Patel, Honorary Visiting Professor of Pharmacy University of Bradford , Honorary Senior Lecturer Academic Unit Primary Care Medical School University of Sheffield

**Sponsor:** University of Oxford
Joint Research Office
1st floor, Boundary Brook House
Churchill Drive,
Headington
Oxford
OX3 7GB

**Funder:** UKRI, DHSC, NIHR

No potential conflict of interest

**Confidentiality Statement**
This document contains confidential information that must not be disclosed to anyone other than the Sponsor, the Investigator Team, HRA, host organisation, and members of the Research Ethics Committee and Regulatory Authorities unless authorised to do so.

See *supplementary material B* for **Key Trial Contacts**.

© Copyright: The University of Oxford and Oxford University Hospitals NHS Foundation Trust 2019

### Platform Randomised trIal of treatmeNts in the Community for epIdemic and Pandemic iLlnEsses (PRINCIPLE): Overview

**Background:** There is an urgent need to identify effective treatments for SARS-CoV-2 infection that helps people recover quicker and reduces the need for hospital admission. We have established an open, adaptive, platform trial to evaluate treatments suitable for use in the community for treating COVID-like-illness that might help people recover sooner and prevent hospitalisation.

**Eligibility and randomisation:** This protocol describes a randomised trial for people in the community at higher risk of an adverse outcome from possible or confirmed SARS-CoV-2 infection, defined in accordance with the United Kingdom's National Health Service syndromic case definition (https://www.nhs.uk/conditions/coronavirus-covid-19/symptoms/). Participants are randomised to receive either usual care or a trial treatment in addition to usual care (see Intervention Specific Appendices). Participants can take part in the study if they are eligible to be randomised to at least one intervention arm, as well as the Usual Care arm.

**Platform trial:** A "platform trial" is a trial in which multiple treatments for the same disease are tested simultaneously. New interventions can be added or replace existing ones during the course of the trial in accordance with pre-specified criteria.

**Response adaptive randomisation:** The initial randomisation ratio is fixed 1:1 for a comparison between two trial arms, but the trial has the capability for these proportions to be altered according to participants' responses to interventions. Pre-specified decision criteria allow for dropping a treatment for futility, declaring a treatment superior, or adding a new treatment to be tested. If at any point a treatment is deemed superior to the usual care arm, the superior treatment may replace the usual care arm as the new standard of care. In the context of the COVID-19 pandemic, the trial may continue as long as the pandemic persists.

**Outcomes:** The trial has co-primary endpoints: 1) Time taken to self-reported recovery from randomisation; and 2) hospitalisation and/or death. The main objective of the trial is to assess the effectiveness of the interventions in reducing time to recovery and in reducing the incidence of hospitalisation and/or death.

Key secondary outcomes include: Hospital assessment without admission; Oxygen administration; Intensive Care Unit admission; Mechanical ventilation (components of the WHO Clinical Progression Ordinal Scale); Duration of hospital admission; Duration of severe symptoms; Sustained recovery; Contacts with the health services; Consumption of antibiotics; Effects in those with a positive test for COVID-19 infection; WHO Well-being Index; daily rating of how well participant feels; Safety.

See *supplementary material C* for details of objectives and outcome measures.

**Efficient study design:** All enrolment (screening, informed consent, eligibility review and baseline data) and follow-up procedures (daily diary, data capture of hospitalisations and deaths) can be performed and captured online on the trial website or by telephone with a member of the trial

© Copyright: The University of Oxford and Oxford University Hospitals NHS Foundation Trust 2019

team. Randomisation is online and automatic following eligibility confirmation. Participant packs and medications are sent from the central study team directly to the participant.

**Data to be recorded:** We will capture demographic features including ethnicity and care home residency at baseline. In the online daily diary (completed for 28 days)/ during telephone calls, participants or their Study Partners will rate the severity of symptoms including how well they are feeling, record contacts with the health services (including hospital admission), record medication use, and new infections in the household. The WHO-5 Wellbeing Index, a five-question instrument, will assess wellbeing at baseline and on days 14 and 28. Follow-up beyond 28 days after randomisation will be by accessing electronic medical records and by participant questionnaire for information relevant to the longer term consequences of COVID-19.

**Numbers to be randomised:** The trial will continue until either superiority or futility is claimed for each intervention. We estimate that approximately 400 participants per arm (800 participants total if only a single intervention vs. Usual Care) will be required to provide 90% power for detecting an approximate difference of 2 days in median recovery time in the primary analysis population. We estimate that approximately 1500 participants per arm (3000 participants total if only a single intervention vs. usual care) will be required to provide 90% power for detecting a 50% reduction in the relative risk of hospitalisation and/or death in the primary analysis population.

**To enquire about the trial, contact the PRINCIPLE Trial Team:**

PRINCIPLE Trial
Nuffield Department of Primary Care Health Sciences
Radcliffe Primary Care
Radcliffe Observatory Quarter, Woodstock Road
Oxford
OX2 6GG

Website: www.principletrial.org

© Copyright: The University of Oxford and Oxford University Hospitals NHS Foundation Trust 2019

© Copyright: The University of Oxford and Oxford University Hospitals NHS Foundation Trust 2019

5.2.6 Virtual Trial Simulations ... 24
5.2.7 Procedure for Accounting for Missing, Unused, and Spurious Data ... 25
5.3 Primary Analysis Population ... 25
5.4 Procedures for Reporting Unplanned Deviation(s) from the Master Statistical Analysis Plan 25
5.5 Qualitative sub-study analysis ... 25
6 DATA MANAGEMENT ... 25
6.1 Source Data ... 25
6.2 Access to Data ... 26
6.3 Data Recording and Record Keeping ... 26
7 QUALITY ASSURANCE PROCEDURES ... 27
7.1 Risk assessment and Monitoring ... 27
7.2 Trial committees ... 27
8 PROTOCOL DEVIATIONS ... 27
9 SERIOUS BREACHES ... 28
10 ETHICAL AND REGULATORY CONSIDERATIONS ... 28
10.1 Declaration of Helsinki ... 28
10.2 Guidelines for Good Clinical Practice ... 28
10.3 Approvals ... 28
10.4 Other Ethical Considerations ... 28
10.5 Reporting ... 29
10.6 Transparency in Research ... 29
10.7 Participant Confidentiality ... 29
10.8 Expenses and Benefits ... 29
11 FINANCE AND INSURANCE ... 29
11.1 Funding ... 30
11.2 Insurance ... 30
11.3 Contractual arrangements ... 30
12 PUBLICATION POLICY ... 30
13 DEVELOPMENT OF A NEW PRODUCT/ PROCESS OR THE GENERATION OF INTELLECTUAL PROPERTY ... 30
14 ARCHIVING ... 30
15 REFERENCES ... 31
16 APPENDIX A: SCHEDULE OF PROCEDURES ... 36
17 APPENDIX B: AMENDMENT HISTORY ... 39
18 APPENDIX C: USUAL CARE ARM ... 42
1. Background and rationale ... 42
2. Changes to outcome measures ... 42

© Copyright: The University of Oxford and Oxford University Hospitals NHS Foundation Trust 2019

3. Detail of intervention ........ 42
a. Investigational Medicinal Product (IMP) description ........ 42
b. Storage of IMP ........ 42
4. Safety reporting ........ 42
19 APPENDIX D: USUAL CARE PLUS HYDROXYCHLOROQUINE ARM (DISCONTINUED) ........ 43
1. Background and rationale ........ 43
a. Evidence for potential Hydroxychloroquine benefits in COVID-19 ........ 43
2. Eligibility criteria specifically related to hydroxychloroquine ........ 47
3. Outcome measures related to hydroxychloroquine ........ 48
4. Detail of intervention ........ 48
a. Investigational Medicinal Product (IMP) description ........ 48
b. Storage of IMP ........ 48
c. SmPC precautions and concomitant medication ........ 49
*i.* Precautions ........ 49
ii. Concomitant medication ........ 49
iii. Pregnancy and Breastfeeding ........ 50
5. Safety reporting ........ 50
20 APPENDIX E: USUAL CARE PLUS AZITHROMYCIN ARM (DISCONTINUED) ........ 51
1. Background and rationale ........ 51
a. Evidence for potential Azithromycin benefits in COVID-19 ........ 51
b. Importance of treating CAP or CAP risk in the elderly or immuno-compromised ........ 51
2 Changes to outcome measures ........ 52
3 Eligibility criteria specifically related to azithromycin ........ 52
4 Detail of intervention ........ 52
a. Investigational Medicinal Product (IMP) description ........ 53
b. Storage of IMP ........ 53
c. SmPC precautions and concomitant medication ........ 53
i. Precautions ........ 53
ii. Concomitant medications ........ 53
iii. Fertility, pregnancy and lactation ........ 57
5 Safety reporting ........ 57
21 APPENDIX F: USUAL CARE PLUS DOXYCYCLINE ARM (DISCONTINUED) ........ 58
1. Background and rationale ........ 58
a. Evidence for potential doxycycline benefits in COVID-19 ........ 58
b. Importance of treating CAP or CAP risk in the elderly or immuno-compromised ........ 58
2. Changes to outcome measures ........ 59
3. Eligibility criteria specifically related to doxycycline ........ 59

© Copyright: The University of Oxford and Oxford University Hospitals NHS Foundation Trust 2019

4. Detail of intervention ........ 59
a. Investigational Medicinal Product (IMP) description ........ 59
b. Storage of IMP ........ 60
c. SmPC precautions and concomitant medication ........ 60
i. Precautions ........ 60
ii. Concomitant medications ........ 60
5. Safety reporting ........ 60
22 APPENDIX G: USUAL CARE PLUS INHALED CORTICOSTEROID (ICS) ARM ........ 61
1. Background and rationale ........ 61
a. Evidence for potential benefits of inhaled corticosteroids in COVID-19 illness ........ 61
2. Changes to outcome measures ........ 62
3. Eligibility criteria specifically related to ICS ........ 62
4. Detail of intervention ........ 62
a. Investigational Medicinal Product (IMP) description ........ 62
b. Storage of IMP ........ 62
c. SmPC precautions and concomitant medication ........ 63
i. SmPC Precautions ........ 63
ii. Concomitant medications ........ 63
5. Safety reporting ........ 63
23 APPENDIX H: USUAL CARE PLUS COLCHICINE ........ 64
1. Background and rationale ........ 64
a. Evidence for potential benefits of colchicine in COVID-19 illness ........ 64
*b.* Potential mechanism of action ........ 65
2. Changes to outcome measures ........ 65
3. Eligibility criteria specifically related to colchicine ........ 66
4. Detail of intervention ........ 66
a. Investigational Medicinal Product (IMP) description ........ 67
b. Storage of IMP ........ 68
i. SmPC Precautions ........ 68
ii. Concomitant medications ........ 68
iii. Fertility, pregnancy and lactation ........ 69
5. Safety reporting ........ 69
Drug Accountability ........ 70
24 APPENDIX I: USUAL CARE PLUS FAVIPIRAVIR ........ 71
1. Background and rationale ........ 71
a. Evidence for potential benefits of favipiravir in COVID-19 illness ........ 71
b. Potential mechanism of efficacy ........ 72

© Copyright: The University of Oxford and Oxford University Hospitals NHS Foundation Trust 2019

2. Changes to outcome measures ..... 72
3. Eligibility criteria specifically related to favipiravir ..... 72
4. Detail of intervention ..... 73
a. Investigational Medicinal Product (IMP) description ..... 73
b. Storage of IMP ..... 73
c. Precautions, concomitant medications, pregnancy and lactation ..... 73
i. Precautions ..... 74
ii. Concomitant medications ..... 74
iii. Pregnancy and lactation ..... 74
5. Safety reporting ..... 74
a. Side-effects ..... 74
b. Reference Safety Information (RSI) ..... 75
c. Drug Accountability ..... 75
d. Risk/Benefit Assessment ..... 75
i. Risks ..... 75
ii. Benefits ..... 76
25. APPENDIX J: USUAL CARE PLUS IVERMECTIN ..... 77
1. Background and rationale ..... 77
a. Evidence for potential benefits of ivermectin in COVID-19 illness ..... 77
*b.* Potential mechanism of action ..... 79
2. Changes to outcome measures ..... 79
3. Eligibility criteria specifically related to Ivermectin ..... 79
4. Detail of intervention ..... 80
a. Investigational Medicinal Product (IMP) description ..... 80
b. Storage of IMP ..... 81
c. Precautions, concomitant medications, pregnancy and lactation ..... 81
i. Precautions ..... 81
ii. Concomitant medications ..... 81
iii. Pregnancy and lactation ..... 82
5. Safety reporting ..... 82
b. Reference Safety Information (RSI) ..... 83
c. Drug Accountability ..... 83
26. Supplementary Material ..... 85
A. Abbreviations ..... 85
B. Key Trial Contacts ..... 86
C. Objectives and Outcome Measures ..... 88
D. Adverse Events ..... 89

© Copyright: The University of Oxford and Oxford University Hospitals NHS Foundation Trust 2019

Definitions ........................................................................................................ 89
E. Data Recording and Record Keeping ..................................................................... 91
F. Qualitative Sub-study ........................................................................................... 91

© Copyright: The University of Oxford and Oxford University Hospitals NHS Foundation Trust 2019

#### 1. BACKGROUND and RATIONALE

We urgently need to know whether potential treatments for COVID-19-like-illness that are suitable for use in the community might help affected individuals recover more quickly and reduce the risk of hospitalisation and/or death.(1) PRINCIPLE is a platform trial designed to efficiently evaluate potential treatments for people with COVID-19-like-illness in the community, and who may be at higher risk of poorer outcomes. Eligible participants are those who meet the United Kingdom's National Health Service syndromic case definition (https://www.nhs.uk/conditions/coronavirus-covid-19/symptoms/), who are being managed in the community, and who are aged 65 and over, or 18 to 64 and experiencing shortness of breath as part of COVID-19 illness, or aged 18-64 with certain comorbidities (2-6)

The platform trial has the flexibility to allow additional interventions to be added in, or to replace existing interventions according to pre-specified criteria. If at any point a treatment is deemed superior to the usual care arm, the superior treatment may replace the usual care arm as the new standard of care. All approved intervention arms are outlined in Intervention Specific Appendices (ISAs).

The trial has co-primary endpoints: 1) Time taken to self-reported recovery from randomisation; and 2) Hospitalisation and/or death. The main objective of the trial is to assess the effectiveness of the respective interventions in reducing time to recovery and in reducing the incidence of hospitalisation and/or death.

The primary analysis will include all participants as specified in the master statistical analysis plan and the adaptive design report. Clinical data, and information from swab and blood tests, where available, will be used to classify participants according to aetiology.

#### 2. TRIAL DESIGN AND PROCEDURES

PRINCIPLE is an open, prospective, individually randomised, platform, response adaptive, controlled clinical trial in community care.

##### 2.1 Participant Identification

###### 2.1.1 Trial Participants

The trial aims to include symptomatic participants with confirmed, or possible COVID-19 who meet the current NHS case definition for possible COVID-19, and who are well enough to remain in the community. This definition can be found here: https://www.nhs.uk/conditions/coronavirus-covid-19/symptoms/. Participants must be aged 65 and over, **OR** aged 18 to 64 and experiencing shortness of breath as part of COVID-19 illness, **OR** aged 18-64 with certain comorbidities.

Participants experiencing shortness of breath have a greater risk of severe and critical disease outcomes with COVID-19 (5).

The study is for people who have ongoing symptoms.

© Copyright: The University of Oxford and Oxford University Hospitals NHS Foundation Trust 2019

##### 2.1.2 Inclusion Criteria

Inclusion requires the following:

1. Participant or their legal representative, is willing and able to give informed consent for participation in the study, and is willing to comply with all trial procedures

2. Suspected COVID-19 using the NHS syndromic definition, OR symptoms consistent with COVID-19* and with a positive test for SARS-CoV-2 infection within the past 14 days

3. Symptoms must have started within the past 14 days and be ongoing

**AND**

4. Participant is aged 65 or over **OR**

Participant is aged 18- 64, and is experiencing shortness of breath as part of COVID-19 illness **OR**

Participant is aged 18-64 and has any of the following underlying health conditions

a) Known weakened immune system due to a serious illness or medication (e.g. chemotherapy);
b) Known heart disease and/or a diagnosis of high blood pressure
c) Known chronic lung disease (e.g. asthma)
d) Known diabetes
e) Known mild hepatic impairment;
f) Known stroke or neurological problem;
g) Self-report obesity or body mass index ≥35 $kg/m^2$

*These symptoms may include, but are not limited to, shortness of breath, general feeling of being unwell, muscle pain, diarrhoea and vomiting.

##### 2.1.3 Exclusion Criteria

- Patient currently admitted in hospital
- Almost recovered (generally much improved and symptoms now mild or almost absent)
- Judgement of the recruiting clinician deems ineligible.
- Previous randomisation to an arm of the PRINCIPLE trial

Additional exclusions specific to each intervention arm are listed in the ISAs. For participation, participants must be eligible to be randomised to at least one intervention arm as well as the Usual Care arm.

#### 2.2 Trial procedures

##### 2.2.1 Recruitment

Recruitment is possible through a variety of mechanisms:

© Copyright: The University of Oxford and Oxford University Hospitals NHS Foundation Trust 2019

##### 2.2.2 Face to face

Attending clinicians including video consultations, including research nurses or other health care professionals, at general medical practices, paramedic services, hospital emergency departments, clinical care hubs, Hospital at Home facilities, care of the elderly services, pharmacies, social care services, residential and nursing homes, or any health and social care facility, can facilitate recruitment into the trial. They can do this by discussing the study with potentially eligible participants, guiding them through informed consent procedures, collection of baseline data, completion of screening questions, collecting information for eligibility assessment, and randomising the participant. If required and appropriate, licensed prescribers may prescribe the medication appropriate to the group to which the participant is randomised. Alternatively, health care professionals may revert to the PC-CTU to complete the activity, including eligibility confirmation and issue of study medication and materials.

##### 2.2.3 Remote recruitment

i) All Health, health related, and Social Care professionals will be able to give information verbally or via a trial text, email, poster, social media post, adverts, media release, leaflet or letter, to potential study participants and their study partners. They may also direct patients to the online study information and the study website.

ii) Potential participants may present directly to the study team via the website or by the study telephone contact. The study team can provide information about joining the trial and guide them through the consent and enrolment process.

iii) A General Practice may be contacted by a potential participant or the practice may contact patients, by text (or by letter), who may match the trial eligibility criteria, through running searches of their database. They will then direct patients to the trial enrolment website or seek verbal consent to be contacted by the trial team.

iv) NHS Digital will provide the PRINCIPLE trial with a daily list of contact details from the COVID-19 testing Pillar 2 data, for patients receiving a positive test result for SARS-Co-V2 infection, via a secure transfer system. NHS Digital will apply an age filter to ensure only the details of those within the age range of the trial are passed on to PRINCIPLE. The trial team will make a limited number (maximum of 3) of attempts to telephone, text or email these patients to provide them with information about the trial, to invite them to consider taking part, and to enrol them if they provide full informed consent and are deemed eligible at screening.

Patient details will be provided in accordance with section 251 under the General Notice under the Health Service Control of Patient Information Regulations 2002, which applies only in England and Wales, providing patient information without consent for COVID-19 public health, surveillance and research purposes. The notice provides a temporary legal basis to avoid a breach of confidentiality for COVID-19 purposes.

v) Join Dementia Research (JDR) - We will also be using JDR as a recruitment tool. This is an on-line self-registration service that enables volunteers with memory problems or dementia, carers of those with memory problems or dementia and healthy volunteers to register their interest in taking part in research.

© Copyright: The University of Oxford and Oxford University Hospitals NHS Foundation Trust 2019

For all recruitment models:

- Study Partner: at screening the potential participant will be asked to provide contact details for a Study Partner, to assist in completing trial procedures and to provide information on their behalf where necessary, but this is not a requirement for trial participation. However, it is strongly encouraged that participants who may be frailer and/or lack capacity to consent make use of a study partner to facilitate their participation. In addition to family member or friend, the study partner may also be a carer or other suitable person.
- Participants may be asked if they wish to receive information about other studies that do not conflict with the main PRINCIPLE trial. Those who do not screen as eligible for PRINCIPLE may be given information about other approved trials they could potentially participate in.

#### 2.3 Screening

An online screening, eligibility and consent procedure is used. If online access is not possible, a member of the trial team collects this information during a telephone call. A trial free-phone number enables participants to contact the trial team for further information and study participation support. Participants are screened after they have read the PIS by completing an online eligibility questionnaire.

#### 2.4 Informed Consent

If participants meet the screening criteria, they will be asked to provide informed consent and a screening trial ID number will be assigned to them. Remote, paperless online/telephone consent is required, and appropriate during the pandemic. Participants will be able to download their consent form, or it may be printed by the central study team and delivered to participants with their study materials if they so prefer.

Written and summary versions of the PIS and ICF will be presented to participants detailing no less than: the exact nature of the trial; the known side-effects and risks involved in taking part. It will be clear that the participant is free to withdraw from the study at any time. A summary, pictorial PIS is available which can be read by those feeling very unwell, lack capacity or have low reading comprehension skills. Adequate time will be given to the participant to consider the information and to ask any questions about the trial before deciding whether to participate. After consent, the participant will enter online baseline information, including their address, contact details and those of a Study Partner.

Population groups such as care home residents have been amongst those hardest hit by the pandemic and therefore stand to benefit the most from any effective treatments. However, some care home residents lack capacity to consent to research themselves. If the recruiting clinician deems a care home resident lacks capacity to consent then a personal or professional legal representative (England and Wales only) will be asked to provide consent for those lacking capacity to consent for themselves. A personal legal representative is defined as a person not connected with the conduct of the trial who is suitable to act as the legal representative by virtue of their relationship with the adult. A professional legal representative may be a doctor responsible for the medical treatment of the adult if they are independent of the study, or a

© Copyright: The University of Oxford and Oxford University Hospitals NHS Foundation Trust 2019

person nominated by the healthcare provider. In all instances, a personal legal representative will be sought first and a professional legal representative sought only if a personal legal representative cannot be identified. A professional legal representative will be sought in order not to deny access to research to older adults who may not have personal legal representatives. Only residents of care homes who lack capacity to consent will be recruited, adults who lack capacity to consent will not be recruited from the wider community. Legal Guardians and recruiting clinicians will not endeavour to obtain consent for or recruit into the trial residents who, in addition to their lack of capacity, have a quality of life which can reasonably be considered as not acceptable to the potential participant.

The legal representative will be provided with information about the trial and made aware of the following:

- They are being asked to give consent on behalf of the incapacitated adult,
- They are free to decide whether they wish to make this decision or not, and
- They are being asked to consider what the adult would want, and to set aside their own personal views when making this decision.

#### 2.5 Eligibility Assessment

Eligibility of those who have provided appropriate consent can be checked at study sites or centrally by a medically qualified clinician or a research nurse, who is suitably trained and experienced and has been delegated this responsibility, and who has appropriate access to the participant's summary care record or relevant medical information. If a participant's summary care record cannot be accessed centrally, the clinician/delegate will contact the participant's primary care medical practice for information relevant to confirming eligibility. Participants will not be randomised to an arm if an exclusion criterion to that arm applies to them, but will need to have no exclusions relevant to at least one intervention and the usual care arm.

#### 2.6 Randomisation

Participants will be randomised using a fully validated and compliant web-based randomisation system called Sortition. Once deemed eligible, the clinician or a member of the trial team will randomise the participant, to one of the arms they are eligible for (at least two arms, usual care and at least one intervention), automatically by Sortition. Full details of response adaptive randomisation are described in section 5.2.2.

The participant, legal representative if applicable, trial team and participant's GP will be notified electronically of the treatment allocation. If the participant does not have an email address, they will be notified by telephone. The research team may also send the GP or Care Home an email or letter via secure systems, containing personally identifiable data and treatment allocation.

#### 2.7 Blinding and code-breaking

© Copyright: The University of Oxford and Oxford University Hospitals NHS Foundation Trust 2019

PRINCIPLE is an open-label trial. The participant, legal representative if applicable, and the recruiting clinician will know the participant's allocation. Therefore, no unblinding or code breaking is required. However, those managing the data will be blind to participant allocation.

The trial team and recruiting clinicians will be blinded to emerging results and the response adaptive randomisation allocation ratio. During the course of the trial, only the unblinding statisticians and the independent members of the Data Monitoring and Safety Committee will have access to the unblinded interim results.

#### 2.8 Baseline Assessments

Once randomised, study medication (if so randomised), and a participant pack will be sent to participants, from their general practice, study team, Public Health England (PHE) or other approved central service (or collected from a general practice or pharmacy). Participants may be offered a swab test as part of standard care. Where possible, and availability of sampling kits allows, one sample will be taken as close to study entry as possible to assess COVID-19 status and other viral aetiologies. While the aim is to have a swab result for all patients, where swabs are unavailable, patients may still participate and be included in the primary intention to treat analysis only.

#### 2.9 Subsequent Visits

There is no requirement for participants to have a face-to-face visit as part of trial participation. Those participants randomised to an unlicensed medication will receive a call from the study team within one day of randomisation, to reaffirm consent, to explain when to call the 24 hour safety phone line, what to do in an emergency, and to answer any other trial questions. This information is also included in the participant information sheet and the participation pack that they will receive. All subsequent measurements consist of self-completed questionnaires online or through telephone calls, and primary care and/or hospital record searches. We will ascertain relevant data from primary care and/or hospital medical records about length of hospital stay, oxygen therapy, and ICU admission and ventilation, if applicable.

Participants will be sent a link to their online diary, which they will be asked to complete for 28 days. They will be asked to rate the severity of symptoms, record contacts with the health services (including hospital admission), record medication use and new infections in the household. It is becoming increasingly apparent the COVID-19 infection may have a considerable negative impact on well-being (7) and so the five questions of WHO-5, validated for measuring wellbeing over time, will be presented at baseline and on days 14 and 28. We will not ask for WHO-5 questions to be completed for participants who lack capacity. We will capture ethnicity and care home residency at baseline and day 28 (if missed at baseline).

All participants receive a call from the trial team on day 2/3 to reaffirm consent, to confirm that they have received a participant pack, and trial medication (if randomised to a trial medication), and to explain that they should complete the daily diary for 28 days even if they feel better or their swab result is negative. The trial team calls participants/study partners on days 7, 14 and 28 if they do not have internet access or have *not* completed their diary for at least 2 consecutive days prior to the call. No more than six contact attempts will be made at each of these follow-up points. For those on unlicensed medication, if the participant/study partner does not answer the

© Copyright: The University of Oxford and Oxford University Hospitals NHS Foundation Trust 2019

calls and hasn't completed online diaries, their GP will be contacted to allow us to monitor any potential side-effects associated with the medication.

We will seek consent from participants to contact them on a monthly basis for up to 12 months after enrolment (via email, text message or phone call) to collect information about any ongoing symptoms, hospitalisations and well-being. We will re-consent those already enrolled in the trial.

In addition to the swab being undertaken as part of the national RCGP RSC surveillance programme with PHE, trial participants will also be asked to consent to the trial team accessing a blood sample result. The study team will obtain the result from RCGP RSC/PHE.

The RCGP RSC will report to the central trial office at regular intervals about healthcare contacts in the participant's clinical records, as they are able to download this information centrally. This will be used as confirmation and a back-up for information obtained directly from study participants and other data sources outlined above. If obtaining data is not possible using this route, the GP surgery will be contacted to request a limited notes review. Participant records will be accessed up to twelve months following enrolment to ascertain follow up data from enrolment to day 28. Data will be collected as close to real time as possible; RCGP RSC, EMIS and NHS Digital and other sources of routinely collected data will be utilised if required To investigate the impact of trial interventions on the longer-term effects of COVID-19, we will use these data collection methods to follow-up participants, for up to 10 years.

##### 2.10 Qualitative Sub-study

A qualitative sub-study will be nested within the trial, to capture data to understand how patients conceptualise their illness and how they respond to taking medication(s) as part of the trial. Once participants have completed the trial, we will interview their respective clinicians to explore their views of taking part in trials during a pandemic. Healthcare professionals will also be asked about their experiences of taking part in the trial. See *supplementary material F* for further details. Participants who lack capacity will not be invited to participate in the qualitative sub study.

##### 2.11 Early Discontinuation/Withdrawal of Participants

Each participant, or their legal representative on the participant's behalf, has the right to withdraw from the study at any time. For those that lack capacity, expression of dissent in any form will be taken as an indication they do not wish to be included and they will be withdrawn. In addition, the Investigator may discontinue a participant from the study at any time if the Investigator considers it necessary for any reason including:

- Ineligibility (either arising during the study or retrospectively having been overlooked at screening)
- Withdrawal of consent

The reason for withdrawal will be recorded on the CRF. Data that has already been collected about the participant will be kept and used.

##### 2.12 Definition of End of Trial

© Copyright: The University of Oxford and Oxford University Hospitals NHS Foundation Trust 2019

Last data capture of last participant, when: no further suitable interventions are available and/or COVID-19 is no longer prevalent. March 2022 has been decided as the formal end date at this stage, but this date may need to be amended depending on circumstances prevailing at the time.

### 3 TRIAL INTERVENTIONS

IMP information can be found in the relevant ISAs.

In general, re-packaging and issuing of medication can be completed by: the patient's registered GP surgery or treatment and assessment facility; an accredited licensed central facility; an online, community or hospital pharmacy, and The Primary Care Clinical Trials Unit (as approved by the MHRA). Distribution of trial packs to participants will be tracked via courier or call/text message. Clinicians can prescribe trial medications that can be issued in the community and pharmacies can issue medication to the patient by community pharmacy services 'on-line pharmacy' services, NHS volunteers, or it can be collected from the pharmacy by the participant or someone on their behalf.

To record presence of symptoms and severity, as well as adherence to trial treatment, participants will receive a daily email asking them to complete an online diary where they will record their symptoms and medicines use. If incomplete, the trial team will contact the participant and/or their Study Partner to obtain the data. Section 2.9 explains the additional oversight of those participants receiving unlicensed medication. A risk-adapted approach will be used for drug accountability. Accountability logs will be kept by PC-CTU when they ship drug.

### 4 SAFETY REPORTING

All symptoms, medication side-effects and SAEs will be collected from participant daily diaries, calls to participants/Study Partners, medical records, notes reviews and RCGP data downloads. SAE information will be analysed as part of the interim and whole trial analysis and will be reviewed at each Data Safety & Monitoring Committee meeting.

#### 4.1 Procedures for Reporting Adverse Events and Serious Adverse Events

The severity of events and symptoms will be assessed by participants in daily diaries on the following scale: no problem/mild problem/moderate problem/major problem.

| | Participant reported symptom rating |
|---|---|
| No problem | Symptom not experienced |
| Mild problem | Short-lived or mild symptoms; medication may be required. No limitation to usual activity |
| Moderate problem | Moderate limitation in usual activity. Medication may be required. |
| Major problem | Considerable limitation in activity. Medication or medical attention required. |

i. **AE reporting**

© Copyright: The University of Oxford and Oxford University Hospitals NHS Foundation Trust 2019

AEs will be monitored daily for participants allocated to either hydroxychloroquine or treatments not licensed in the UK, whilst they are taking the drug, to allow successful safety monitoring of these less familiar treatments. Participants will be free to withdraw from taking the treatment if they perceive they have an intolerable AE.

For each treatment not licensed in the UK, the following AEs from the start of medication until the specified follow-up period, will be assessed by a clinician for causality and severity (definitions below): i) pre-defined AEs detailed in the ISA that are rated by the participant as 'moderate' and ii) other reported 'major' AEs.

Participants will also be provided with a Trial Wallet Emergency Card detailing potential side-effects and a 24-hour contact telephone line, manned by a clinical team, enabling them to report any moderate or major AEs they experience whilst taking the drug. The clinician will contact the participant directly within 24 hrs of becoming aware of a major AE reported in their daily diary or on the Freephone number, to advise the participant on the appropriate clinical care, as well as notifying the participant's GP about the event. In the event of a medical emergency, trial participants will be instructed to show this card to the clinician they see.

##### ii. AE Severity Assessment (for assessing clinician):

| | Clinical assessment of severity |
|---|---|
| **GRADE 1 (Mild)** | Short-lived or mild symptoms; medication may be required. No limitation to usual activity |
| **GRADE 2 (Moderate)** | Moderate limitation in usual activity. Medication may be required. |
| **GRADE 3 (Severe)** | Considerable limitation in activity. Medication or medical attention required. |

##### iii. SAEs

Hospitalisation and/or death due to confirmed or possible SARS-Cov-2 infection is a primary outcome, we will collect this data using a risk-adapted approach and will not report such SAEs. SAEs other than hospitalisation or death due to COVID-19 must be reported for all treatments.

SAEs must be reported by the person who has discovered the SAE or nominated delegate within 24 hours of becoming aware of the event. Some SAEs occurring within the 28 day follow-up period, may be identified retrospectively from data extracts, these will be reported within 24 hours of becoming aware of the event. The sponsor or delegate will ensure it is reviewed by the CI or other delegated personnel for relatedness and expectedness as soon as possible taking into account the reporting time for a potential SUSAR according to the relevant competent authority. If the event has not resolved, at the 28 day time point the SAE will be reviewed again to see if resolution has occurred. If the event is considered 'resolved' or 'resolving' no further follow up is required. If not, the event must be followed up until such a time point.

© Copyright: The University of Oxford and Oxford University Hospitals NHS Foundation Trust 2019

All SAEs that have not resolved by the end of the study or those that are identified retrospectively, or that have not resolved upon discontinuation of the participant's participation in the study, must be followed until any of the following occurs:

• The event resolves

• The event stabilises

• The event returns to “baseline”, if a “baseline” value/status is available

• The event can be attributed to agents other than the study intervention or to factors unrelated to study conduct

• It becomes unlikely that any additional information can be obtained (participant or health care practitioner refusal to provide additional information, lost to follow-up after demonstration of due diligence with follow-up efforts)

*See Supplementary Material D for definitions of adverse events*

##### 4.1.1. Other events exempt from immediate reporting as SAEs

Hospitalisations will be defined as at least a one night admission to hospital. Hospitalisation for a pre-existing condition, including elective procedures planned prior to study entry, which has not worsened, does not constitute an SAE, and standard supportive care for the disease under study are not SAEs and do not require SAE reporting.

##### 4.1.2. Procedure for immediate reporting of Serious Adverse Events

- Trial team will complete an SAE report form for all reportable SAEs.
- GP practice/trial team/RCGP will provide additional, missing or follow up information in a timely fashion.
- If necessary the participant may be contacted to provide additional, missing or follow up information as required.

The CI or delegate will review the SAE once reported, collect as much information and report to the Sponsor within the timeframe according to the PC-CTU SOPs.

##### 4.1.3 Expectedness and Causality

For SAEs that require reporting, expectedness of SARs will be determined according to the relevant RSI section of the Summary of Product Characteristics/IB. The RSI will be the current Sponsor and MHRA approved version at the time of the event occurrence.

#### Assessment of Causality

The relationship of each serious adverse event to the trial medication must be determined by a medically qualified individual according to the following definitions:

- **Unrelated** – where an event is not considered to be related to the IMP

© Copyright: The University of Oxford and Oxford University Hospitals NHS Foundation Trust 2019

- **Possibly** – although a relationship to the IMP cannot be completely ruled out, the nature of the event, the underlying disease, concomitant medication or temporal relationship make other explanations possible.
- **Probably** – the temporal relationship and absence of a more likely explanation suggest the event could be related to the IMP.
- **Definitely** – the known effects of the IMP, its therapeutic class or based on challenge testing suggest that the IMP is the most likely cause.

All AEs/SAEs labelled possibly, probably or definitely will be considered as related to the IMP.

##### 4.2 SUSAR Reporting

All SUSARs will be reported by the sponsor delegate to the relevant Competent Authority and to the REC and other parties as applicable. For fatal and life-threatening SUSARS, this will be done no later than seven calendar days after the Sponsor or delegate is first aware of the reaction. Any additional relevant information will be reported within eight calendar days of the initial report. All other SUSARs will be reported within 15 calendar days.

Principal Investigators will be informed of all SUSARs for the relevant IMP for all studies with the same Sponsor, whether or not the event occurred in the current trial.

##### 4.3 Development Safety Update Reports

The DSUR will be developed and submitted annually on the anniversary date that the trial receives Clinical Trial Authorisation +60 days. Due to the nature of this trial and the importance of sharing the science of COVID-19 and the drug, internationally, we expect to produce reports to the UK Government and regulatory agency more frequently upon request.

#### 5 STATISTICS

##### 5.1 Master Statistical Analysis Plan (M-SAP)

Details of the statistical design and methods will be described in a Master Statistical Analysis Plan (M-SAP), in which an appendix to the M-SAP titled "Adaptive Design Report" (ADR) provides complete specifications for the primary analysis and pre-specified adaptive algorithm. In addition, the M-SAP will be accompanied by arm-specific appendices to describe any planned deviations from the M-SAP. A broad overview of the design and primary analyses is provided below.

##### 5.2 Open Adaptive Platform Trial

PRINCIPLE is an open, adaptive, platform trial to evaluate emerging treatments for symptomatic COVID-19-like illness. A "platform trial" is a trial in which multiple treatments for the same disease are tested simultaneously. The backbone of the trial is an adaptive clinical trial design. Pre-specified decision criteria allow for dropping a treatment for futility, declaring a treatment superior, or adding a new treatment to be tested. If at any point a treatment is deemed superior to the Usual Care arm, the superior treatment may replace the Usual Care arm as the new

© Copyright: The University of Oxford and Oxford University Hospitals NHS Foundation Trust 2019

standard of care. Because the process of dropping and adding treatments may be on-going for an indefinite period of time, platform trials may be better conceived of as a process rather than a singular clinical trial. In the context of the COVID-19 pandemic, the trial may continue as long as the pandemic persists.

The PRINCIPLE trial will begin as a two arm, 1:1 randomised trial but will have the capability to add additional interventions over time. The evaluation of any new interventions will be governed by this master protocol and M-SAP (including adaptive algorithm and decision criteria), with any planned deviations from the master protocol and M-SAP to be specified in arm-specific appendices. The inclusion of any new interventions will require additional arm-specific appendices to the master protocol and M-SAP.

##### 5.2.1 Co-Primary Endpoints & Analyses

There are two co-primary endpoints. The first co-primary endpoint is time to recovery from possible COVID-19 infection within 28 days from randomisation, where time to recovery is defined as the first instance that a participant reports feeling recovered. The second co-primary endpoint is hospital admission or death related to possible or confirmed COVID-19 within 28 days from randomisation. Unless otherwise specified in the Intervention Specific Appendices (ISA), the co-primary outcomes will be evaluated using a “gate-keeping” strategy. For a given treatment, the hypothesis for the time to recovery endpoint will be evaluated first, and if the recovery null hypothesis is rejected, the hypothesis for the second co-primary endpoint of hospitalisation/death will be evaluated. This gate-keeping strategy preserves the overall Type I error of the primary endpoints without additional adjustments for multiple hypotheses. In addition, the gate-keeping structure reflects the clinical belief that an intervention is unlikely to demonstrate benefit on the hospitalisation/death endpoint without first demonstrating benefit on the time to recovery endpoint.

The primary outcome of time to recovery is defined as the first instance that a participant reports feeling recovered. The corresponding primary analysis for this outcome is a Bayesian piecewise exponential model, with time to recovery regressed on treatment and stratification covariates (age, comorbidity). Let $\theta_j$ denote the log hazards ratio comparing the hazards of recovery for participants in treatment group *j* versus participants in the Usual Care arm. A corresponding Bayesian posterior distribution will be derived for the estimated log hazards ratio. The first co-primary analysis for intervention *j* will test the following hypothesis:

$$H_{10}: \theta_j \leq 0$$
$$H_{11}: \theta_j > 0$$

If the Bayesian posterior probability of superiority (a log hazards ratio greater than 0 corresponding to quicker recovery) for a treatment versus Usual Care is sufficiently large (e.g. ≥ 0.99), the null hypothesis will be rejected and the intervention will be deemed superior to Usual Care with respect to time to recovery. The exact threshold of the superiority decision criterion (e.g. 0.99) will be determined *a priori* via simulation to control the one-sided Type I error of the study at approximately 0.025, and will be specified in the Adaptive Design Report (Appendix to the M-SAP). The Adaptive Design Report will also specify appropriate methodology for the primary analysis when the Usual Care arm is replaced by a superior treatment, and for when the comparison of a treatment versus Usual Care includes non-concurrent randomisations.

© Copyright: The University of Oxford and Oxford University Hospitals NHS Foundation Trust 2019

The second co-primary endpoint is hospital admission or death due to possible SARS-CoV-2 infection. The corresponding analysis will be a Bayesian generalised linear model of hospitalisation/death regressed on treatment and stratification covariates (age, comorbidity). Let $\delta_j$ denote the log odds ratio comparing the odds of hospitalisation/death for persons in treatment group *j* versus persons in the Usual Care arm. A corresponding Bayesian posterior distribution will be derived for the estimated log odds ratio. If the first co-primary endpoint hypothesis (for time to recovery) is rejected for intervention *j*, the second co-primary hypothesis for intervention *j* be tested:

$$H_{20}: \delta_j \leq 0$$
$$H_{21}: \delta_j > 0$$

If the Bayesian posterior probability of superiority on hospitalisation/death for a treatment versus Usual Care is sufficiently large (e.g. $\geq 0.99$), the null hypothesis will be rejected and the intervention will be deemed superior to Usual Care with respect to hospitalisation/death. The exact threshold of the superiority decision criterion (e.g. 0.99) will be determined *a priori* via simulation to control the one-sided Type I error of the study at approximately 0.025, and will be specified in the M-SAP.

##### 5.2.2 Adaptive Design

The pre-specified design will allow adaptations to the trial based on the observed co-primary endpoint data. These adaptations include the declaration of success or futility of an intervention at an interim analysis, the addition or removal of treatment arms, and changes in the randomisation probabilities. Adaptations will occur at a given interim analysis if pre-specified conditions are satisfied. The adaptive algorithm will be documented in the Adaptive Design Report, including pre-specified criteria for decisions regarding futility or effectiveness of interventions and/or replacing interventions in the trial.

##### 5.2.3 Interim Analyses

Precise timing of the first interim analysis and frequency of subsequent interim analyses will be specified in the Adaptive Design Report, based on both simulations and logistical considerations. At each interim analysis, all enrolled intervention arms will be evaluated for success and futility on both co-primary endpoints using the Bayesian primary analyses. These interim analyses will maintain the gate-keeping sequential order by first evaluating the hypothesis for time to recovery, and if the recovery endpoint null hypothesis is rejected, subsequently evaluating the hypothesis for hospitalisation and/or death. If the Bayesian posterior probability of superiority of a given intervention versus Usual Care is sufficiently large for a given endpoint (e.g. $\geq 0.99$) within the gate-keeping structure, superiority will be declared versus Usual Care with respect to that endpoint.

If the Bayesian posterior probability of a clinically meaningful treatment effect is sufficiently small (e.g. $< 0.01$) for the first co-primary endpoint (time to recovery), the intervention arm may be dropped from the study for futility. If there are no other intervention arms available, the trial may be suspended; otherwise accrual continues to the remaining treatment arms. The exact futility thresholds will be pre-specified in the Adaptive Design Report and determined via simulation.

© Copyright: The University of Oxford and Oxford University Hospitals NHS Foundation Trust 2019

##### 5.2.4 Allocation & Response Adaptive Randomisation

Initially, randomisation will be fixed 1:1 for a comparison between two trial arms, with stratification by age (less than 65, greater than or equal to 65), and comorbidity (yes/no). If a second experimental intervention arm is added to the study, randomisation allocation will be modified and the additional intervention will be included in the interim analyses (with evaluation for success and futility) as detailed in the Adaptive Design Report. If there are at least 3 arms (2 intervention arms plus Usual Care) in the study, each interim analysis may incorporate modified randomisation probabilities via response adaptive randomisation (RAR). Full details for implementing RAR will be provided in the Adaptive Design Report; the general idea is to allocate more participants to the intervention arms that have the best observed outcomes. Except for the CTU programmer, the rest of the trial team are blinded to the RAR ratios.

##### 5.2.5 Sample Size Justification

Given the open perpetual trial structure, the trial does not have a finite ending based on sample size. Rather, the trial will continue until either superiority or futility is claimed for each intervention, or until the pandemic expires in the population. We estimate that approximately 400 participants per arm (800 participants total if only a single intervention vs. Usual Care) will be required to provide 90% power for detecting a hazard ratio of 1.3 (approximate difference of 2 days in median recovery time). This calculation is based on the assumption of an exponential distribution for time to recovery with a median of 9 days in the Usual Care arm, with some adjustments for missing data and multiple interim analyses. On average, we expect fewer participants to be required when there is a large treatment benefit or complete lack of benefit. For example, if the true hazard ratio is 1.5 (3 day benefit in median time to recovery), on average only 150 subjects per arm are required to provide sufficient power. The primary advantage of the adaptive design is the ability to adapt the sample size to the observed data, thus addressing the primary hypothesis as quickly and as efficiently as possible.

We estimate that approximately 1500 participants per arm (3000 participants total if only a single intervention vs. usual care) will be required to provide 90% power for detecting a 50% reduction in the relative risk of hospitalisation and/or death. This calculation is based on the assumption of an underlying 5% combined hospitalisation and/or death rate in the Usual Care arm, with an intervention lowering the hospitalisation and/or death rate to 2.5%, with some adjustments for the multiple interim analyses. We expect fewer participants to be required to detect a 50% reduction if the event rate in the Usual Care arm is greater than 5%.

##### 5.2.6 Virtual Trial Simulations

Because of the adaptive platform trial structure, there exists no simple formula(s) to calculate power and Type I error (false positive rate). Hence, virtual trial simulations will be used to fully characterize and quantify the power and Type I error of the design. These simulations will be conducted prior to the first interim analysis (with results described in the Adaptive Design Report), and will be used to optimize the adaptive decision criterion and RAR parameters. The simulations will include a comprehensive evaluation of trial performance across a wide range of assumptions (e.g. underlying distribution of outcome in Usual Care arm, treatment effect, accrual rates, etc.). This will include summaries regarding the number of subjects required to make a success or

© Copyright: The University of Oxford and Oxford University Hospitals NHS Foundation Trust 2019

futility conclusions for each intervention. Complete details of the simulations will be provided in the Adaptive Design Report.

###### 5.2.7 Procedure for Accounting for Missing, Unused, and Spurious Data.

Full details of handling missing data will be specified in the M-SAP.

##### 5.3 Primary Analysis Population

The primary analysis population is defined as all randomised participants with a COVID-19 positive test, according to the groups they were randomly allocated to as specified in the M-SAP. All other analysis populations will be defined in the M-SAP.

##### 5.4 Procedures for Reporting Unplanned Deviation(s) from the Master Statistical Analysis Plan

Analyses will be carried out in accordance with the M-SAP and corresponding appendices. Any additional analysis that is not specified in the M-SAP/appendices or any unplanned deviation(s) from the M-SAP/appendices will be specified in the Statistical Report. Reasons for these changes will be documented and authorised by the Chief Investigator.

##### 5.5 Qualitative sub-study analysis

Audio-recordings of interviews will be transcribed verbatim and transcripts analysed using thematic analysis. Patient and HCP interviews transcripts will be analysed separately but findings will be compared and triangulated if deemed appropriate. Thematic analysis allows the research team to take a pragmatic approach to data collection, remaining grounded in the data but ensuring that the analysis answers the research objectives. NVivo software will be used to assist with the organisation and coding of data. Codes will be compared with one another to create categories, grouping similar codes together. A thematic framework will be developed to code all data and represent key themes for both sets of interviews.

#### 6 DATA MANAGEMENT

The data management aspects of the study are summarised here with details fully described in the Data Management Plan.

##### 6.1 Source Data

Source documents are where data are first recorded. These include, but are not limited to, hospital records (from which medical history and previous and concurrent medication may be summarised into the CRF), clinical and office charts, laboratory and pharmacy records, diaries, microfiches, radiographs, and correspondence.

If a participant fails to complete data online and after six attempts at contacting the participant/Study Partner, the RCGP RSC may be utilised to obtain missing data. Data collected will include participant identifiable information and will be accessed at the University of Oxford

© Copyright: The University of Oxford and Oxford University Hospitals NHS Foundation Trust 2019

according to PC-CTU Information Governance policies and GDPR. Data will only be held for the duration it is required, this will be reviewed annually.

CRF entries will be considered source data if the CRF is the site of the original recording (e.g. there is no other written or electronic record of data). All documents will be stored safely in confidential conditions. On all study-specific documents, other than the signed consent, the participant will be referred to by the study participant number/code, not by name.

##### 6.2 Access to Data

Direct access will be granted to authorised representatives from the Sponsor and host institution for monitoring and/or audit of the study to ensure compliance with regulations.

##### 6.3 Data Recording and Record Keeping

In accordance with the principles of Good Clinical Practice and the recommendations and guidelines issued by regulatory agencies, the design, conduct and analysis of this trial is focussed on issues that might have a material impact on the wellbeing and safety of study participants (in the community with suspected or confirmed SARS-CoV-2 infection) and the reliability of the results that would inform the care for future patients.

The critical factors that influence the ability to deliver these quality objectives are:

- to minimise the burden on thousands of busy clinicians working in an overstretched primary care setting and undertaking research during a major epidemic
- to ensure that suitable patients have access to the trial medication
- to provide information on the study to patients and clinicians in a timely and readily digestible fashion but without impacting adversely on other aspects of the trial or the patient's care
- to collect comprehensive information on the mortality and disease status

In assessing any risks to patient safety and well-being, a key principle is that of proportionality. Risks associated with participation in the trial must be considered in the context of usual care. At present, there are no proven treatments for COVID-19 for clinicians in the community to prescribe safely with a sound evidence base.

Although data entry should be mindful of the desire to maintain integrity and audit trails, in the circumstances of this epidemic, the priority is on the timely entry of data that is sufficient to support reliable analysis and interpretation about treatment effects.

The Investigators will maintain appropriate medical and research records for this trial, in compliance with the requirements of the Medicines for Human Use (Clinical Trial) Regulations 2004, ICH E6 GCP and regulatory and institutional requirements for the protection of confidentiality of volunteers. The Chief Investigator, Principal Investigator, Co-Investigators, clinical team, including Clinical Research Nurses, and other authorised members of the trial team will have access to records. The Investigators will permit authorised representatives of the sponsor, and regulatory agencies to examine (and when required by applicable law, to copy) clinical records for the purposes of quality assurance reviews, audits and evaluation of the study safety and progress. The software used for the trial is described in *supplementary material E*.

© Copyright: The University of Oxford and Oxford University Hospitals NHS Foundation Trust 2019

### 7 QUALITY ASSURANCE PROCEDURES

The study will be conducted in accordance with the current approved protocol, GCP, relevant regulations and PC-CTU Standard Operating Procedures. All PIs, coordinating centre staff and site staff will receive training in trial procedures according to GCP where required. Regular monitoring will be performed according to GCP using a risk-based approach. Data will be evaluated for compliance with the protocol and accuracy in relation to source documents where possible.

The PC-CTU Trial Management Group will be responsible for the monitoring of all aspects of the trial's conduct and progress and will ensure that the protocol is adhered to and that appropriate action is taken to safeguard participants and the quality of the trial itself. The TMG will be comprised of individuals responsible for the trial's day to day management and will meet regularly throughout the course of the trial.

#### 7.1 Risk assessment and Monitoring

A risk assessment and monitoring plan will be prepared before the study opens and will be reviewed as necessary over the course of the study to reflect significant changes to the protocol or outcomes of monitoring activities. Monitoring will be performed by the PC-CTU Quality Assurance Manager or delegate. The level of monitoring required will be informed by the risk assessment.

#### 7.2 Trial committees

The responsibilities of each group are as follows:

- Data Monitoring and Safety Committee (DMSC) - to review the data at each interim analysis as described in the Statistical Analysis section, as the updates to the randomisation scheme occur in order to ensure that the process is working correctly and to review and monitor the accruing data to ensure the rights, safety and wellbeing of the trial participants. Composition, and roles and responsibilities of the DMSC are detailed in the DMSC charter.
- Trial Steering Committee (TSC) - the Trial Steering Committee ensure the rights, safety and wellbeing of the trial participants. They will make recommendations about how the study is operating, any ethical or safety issues and any data being produced from other relevant studies that might impact the trial. Composition, and roles and responsibilities of the TSC are detailed in the TSC charter.
- Trial Management Group (TMG) - is responsible for the day-to-day running of the trial, including monitoring all aspects of the trial and ensuring that the protocol is being adhered to. It will include Co-Investigators and will meet weekly in the first instance. . Composition, and roles and responsibilities of the TMG are detailed in the TMG charter.
- A core project team (PT) from within the TMG will meet weekly or as required for operational decision making (met daily at the start of the trial).

### 8 PROTOCOL DEVIATIONS

A study related deviation is a departure from the ethically approved study protocol or other study document or process (e.g. consent process or administration of study intervention) or from Good

© Copyright: The University of Oxford and Oxford University Hospitals NHS Foundation Trust 2019

Clinical Practice (GCP) or any applicable regulatory requirements. Any deviations from the protocol will be documented in a protocol deviation form and filed in the study master file.

A PC-CTU SOP is in place describing the procedure for identifying non-compliances, escalation to the central team and assessment of whether a non-compliance /deviation may be a potential Serious Breach.

### 9 SERIOUS BREACHES

A "serious breach" is a breach of the protocol or of the conditions or principles of Good Clinical Practice which is likely to affect to a significant degree –

(a) the safety or physical or mental integrity of the trial subjects; or
(b) the scientific value of the research.

In the event that a serious breach is suspected the Sponsor must be contacted within one working day. In collaboration with the CI, the serious breach will be reviewed by the Sponsor and, if appropriate, the Sponsor will report it to the approving REC committee and the relevant NHS host organisation within seven calendar days.

### 10 ETHICAL AND REGULATORY CONSIDERATIONS

#### 10.1 Declaration of Helsinki

The Investigator will ensure that this study is conducted in accordance with the principles of the Declaration of Helsinki.

#### 10.2 Guidelines for Good Clinical Practice

The Investigator will ensure that this trial is conducted in accordance with relevant regulations and with Good Clinical Practice.

#### 10.3 Approvals

Following Sponsor approval, the protocol, informed consent form, participant information sheets and any proposed informing material will be submitted to an appropriate Research Ethics Committee (REC), regulatory authorities, and host institution(s) for written approval. The PI and coordinating centres for each country will ensure and confirm correct regulatory approvals are gained prior to recruitment.

The Investigator will submit and, where necessary, obtain approval from the above parties for all substantial amendments to the original approved documents.

#### 10.4 Other Ethical Considerations

If a particular arm is deemed futile and dropped, no further participants will be randomised to this arm and anyone who is currently on this arm will be informed it has been dropped.
Once a particular intervention has been declared superior and effective, that will become the comparator arm (i.e. standard care).

© Copyright: The University of Oxford and Oxford University Hospitals NHS Foundation Trust 2019

Some participant's, due to their co-morbidities, will be exempt from prescription charges. Medication is most often sent from PC-CTU and so prescription charges will not apply. All participants will receive a £20 voucher to cover any prescriptions and other expenses they may incur as a consequence of study participation.

Participants who lack capacity to consent for themselves will only be recruited after consultation with their legal representative. Any sign of dissent in any form from the participant who lacks consent will be taken as an indication they do not wish to be involved and they will be withdrawn. Only residents of care homes who lack capacity to consent will be recruited, adults who lack capacity to consent will not be recruited from the wider community.

##### 10.5 Reporting

The CI shall submit once a year throughout the clinical trial, or on request, an Annual Progress Report to the REC, HRA (where required), host organisation, funder (where required) and Sponsor. In addition, an End of Trial notification and final report will be submitted to the MHRA, the REC, host organisation and Sponsor.

##### 10.6 Transparency in Research

Prior to the recruitment of the first participant, the trial will have been registered on a publicly accessible database. Results will be uploaded to the European Clinical Trial (EudraCT) Database within 12 months of the end of trial declaration by the CI or their delegate. Where the trial has been registered on multiple public platforms, the trial information will be kept up to date during the trial, and the CI or their delegate will upload results to all those public registries within 12 months of the end of the trial declaration.

##### 10.7 Participant Confidentiality

The study will comply with the General Data Protection Regulation (GDPR) and Data Protection Act 2018, which require data to be de-identified as soon as it is practical to do so. The processing of the personal data of participants will be minimised by making use of a unique participant study number only on all study documents and any electronic database(s). All documents will be stored securely and only accessible by study staff and authorised personnel. The study staff will safeguard the privacy of participants' personal data.

##### 10.8 Expenses and Benefits

All participants will be reimbursed with a £20 voucher, to cover the payment of a prescription, should they incur this as a result of study participation, and a token of recognition of giving their time and contribution to the study. A proportion of people with the co-morbidities outlined and in the over 50 age-range , are not required to pay for prescriptions. Furthermore, medication is most often sent from the PC-CTU, and so prescription charges will not apply. Participants who complete a telephone interview as part of the qualitative sub-study will be reimbursed with a (second) £20 voucher for their time to participate.

#### 11 FINANCE AND INSURANCE

© Copyright: The University of Oxford and Oxford University Hospitals NHS Foundation Trust 2019

##### 11.1 Funding

The study is funded by the UKRI/NIHR via an MRC call.
The Department of Health & Social Care have provided the following drugs free of charge for trial Use: Hydroxychloroquine, Favipiravir.

##### 11.2 Insurance

The University has a specialist insurance policy in place, which would operate in the event of any participant suffering harm as a result of their involvement in the research (Newline Underwriting Management Ltd, at Lloyd's of London). NHS indemnity operates in respect of the clinical treatment that is provided.

##### 11.3 Contractual arrangements

Appropriate contractual arrangements will be put in place with all third parties.

#### 12 PUBLICATION POLICY

The Investigators (those listed on the protocol and others to be decided at publication) will be involved in reviewing drafts of the manuscripts, abstracts, press releases and any other publications arising from the study. Authors will acknowledge the study funders. Authorship will be determined in accordance with the ICMJE guidelines and other contributors will be acknowledged.

#### 13 DEVELOPMENT OF A NEW PRODUCT/ PROCESS OR THE GENERATION OF INTELLECTUAL PROPERTY

Ownership of IP generated by employees of the University vests in the University. The University will ensure appropriate arrangements are in place as regards any new IP arising from the trial.

#### 14 ARCHIVING

Archiving will be done according to the UOXF PC-CTU SOP and study specific working instructions.

© Copyright: The University of Oxford and Oxford University Hospitals NHS Foundation Trust 2019

© Copyright: The University of Oxford and Oxford University Hospitals NHS Foundation Trust 2019

21. Chen J, Liu D, Liu L, Liu P, Xu Q, Xia L, et al. [A pilot study of hydroxychloroquine in treatment of patients with moderate COVID-19]. Zhejiang Da Xue Xue Bao Yi Xue Ban. 2020;49(2):215-9.
22. Gautret P, Lagier JC, Parola P, Hoang VT, Meddeb L, Mailhe M, et al. Hydroxychloroquine and azithromycin as a treatment of COVID-19: results of an open-label non-randomized clinical trial. Int J Antimicrob Agents. 2020:105949.
23. Retallack H, Di Lullo E, Arias C, Knopp KA, Laurie MT, Sandoval-Espinosa C, et al. Zika virus cell tropism in the developing human brain and inhibition by azithromycin. Proc Natl Acad Sci U S A. 2016;113(50):14408-13.
24. E Bosseboeuf MA, T Nhan, JJ de Pina, JM Rolain, D Raoult, et al. **Azithromycin inhibits the replication of Zika virus.** J Antivirals Antiretrovirals. 2018;10:6-11.
25. Madrid PB, Panchal RG, Warren TK, Shurtleff AC, Endsley AN, Green CE, et al. Evaluation of Ebola Virus Inhibitors for Drug Repurposing. ACS Infect Dis. 2015;1(7):317-26.
26. Bacharier LB, Guilbert TW, Mauger DT, Boehmer S, Beigelman A, Fitzpatrick AM, et al. Early Administration of Azithromycin and Prevention of Severe Lower Respiratory Tract Illnesses in Preschool Children With a History of Such Illnesses: A Randomized Clinical Trial. JAMA. 2015;314(19):2034-44.
27. Villarino AV, Kanno Y, O'Shea JJ. Mechanisms and consequences of Jak-STAT signaling in the immune system. Nat Immunol. 2017;18(4):374-84.
28. Hodge S, Hodge G, Holmes M, Jersmann H, Reynolds PN. Increased CD8 T-cell granzyme B in COPD is suppressed by treatment with low-dose azithromycin. Respirology. 2015;20(1):95-100.
29. Hodge S, Hodge G, Jersmann H, Matthews G, Ahern J, Holmes M, et al. Azithromycin improves macrophage phagocytic function and expression of mannose receptor in chronic obstructive pulmonary disease. Am J Respir Crit Care Med. 2008;178(2):139-48.
30. Hodge S, Reynolds PN. Low-dose azithromycin improves phagocytosis of bacteria by both alveolar and monocyte-derived macrophages in chronic obstructive pulmonary disease subjects. Respirology. 2012;17(5):802-7.
31. Baines KJ, Wright TK, Gibson PG, Powell H, Hansbro PM, Simpson JL. Azithromycin treatment modifies airway and blood gene expression networks in neutrophilic COPD. ERJ Open Res. 2018;4(4).
32. Mandell LA, Wunderink RG, Anzueto A, Bartlett JG, Campbell GD, Dean NC, et al. Infectious Diseases Society of America/American Thoracic Society consensus guidelines on the management of community-acquired pneumonia in adults. Clin Infect Dis. 2007;44 Suppl 2:S27-72.
33. Kawai S, Ochi M, Nakagawa T, Goto H. Antimicrobial therapy in community-acquired pneumonia among emergency patients in a university hospital in Japan. J Infect Chemother. 2004;10(6):352-8.
34. Gleason PP, Meehan TP, Fine JM, Galusha DH, Fine MJ. Associations between initial antimicrobial therapy and medical outcomes for hospitalized elderly patients with pneumonia. Arch Intern Med. 1999;159(21):2562-72.
35. Wu C, Liu Y, Yang Y, Zhang P, Zhong W, Wang Y, et al. Analysis of therapeutic targets for SARS-CoV-2 and discovery of potential drugs by computational methods. Acta Pharm Sin B. 2020.
36. Kim HS, Luo L, Pflugfelder SC, Li DQ. Doxycycline inhibits TGF-beta1-induced MMP-9 via Smad and MAPK pathways in human corneal epithelial cells. Invest Ophthalmol Vis Sci. 2005;46(3):840-8.
37. Clemens DL, Duryee MJ, Sarmiento C, Chiou A, McGowan JD, Hunter CD, et al. Novel Antioxidant Properties of Doxycycline. Int J Mol Sci. 2018;19(12).
38. Hsu AT, Barrett CD, DeBusk GM, Ellson CD, Gautam S, Talmor DS, et al. Kinetics and Role of Plasma Matrix Metalloproteinase-9 Expression in Acute Lung Injury and the Acute Respiratory Distress Syndrome. Shock. 2015;44(2):128-36.
39. NICE. COVID-19 rapid guideline: managing suspected or confirmed pneumonia in adults in the community. NICE; 2020.
40. Nicolau DV, Bafadhel M. Inhaled corticosteroids in virus pandemics: a treatment for COVID-19? Lancet Respir Med. 2020.
41. BTS/SIGN. BTS/SIGN British Guideline on the Management of Asthma. British Thoracic Society; 2019.
42. GINA. Global Strategy for Asthma Management and Prevention (2019 update). 2019.

© Copyright: The University of Oxford and Oxford University Hospitals NHS Foundation Trust 2019

43. Bloom CI, Elkin SL, Quint JK. Changes in COPD inhaler prescriptions in the United Kingdom, 2000 to 2016. Int J Chron Obstruct Pulmon Dis. 2019;14:279-87.
44. Artigas A, Camprubi-Rimblas M, Tantinya N, Bringue J, Guillamat-Prats R, Matthay MA. Inhalation therapies in acute respiratory distress syndrome. Ann Transl Med. 2017;5(14):293.
45. Ortiz-Diaz E, Li G, Kor D, Gajic O, Akca O, Adesanya A, et al. Preadmission Use of Inhaled Corticosteroids Is Associated With a Reduced Risk of Direct Acute Lung Injury/Acute Respiratory Distress Syndrome. CHEST. 2011;140(4):912A.
46. Festic E, Carr GE, Cartin-Ceba R, Hinds RF, Banner-Goodspeed V, Bansal V, et al. Randomized Clinical Trial of a Combination of an Inhaled Corticosteroid and Beta Agonist in Patients at Risk of Developing the Acute Respiratory Distress Syndrome. Crit Care Med. 2017;45(5):798-805.
47. Yamaya M, Nishimura H, Deng X, Sugawara M, Watanabe O, Nomura K, et al. Inhibitory effects of glycopyrronium, formoterol, and budesonide on coronavirus HCoV-229E replication and cytokine production by primary cultures of human nasal and tracheal epithelial cells. Respir Investig. 2020;58(3):155-68.
48. Merad M, Martin JC. Pathological inflammation in patients with COVID-19: a key role for monocytes and macrophages. Nature Reviews Immunology. 2020;20(6):355-62.
49. Barnes PJ. Inhaled Corticosteroids. Pharmaceuticals (Basel). 2010;3(3):514-40.
50. Peters MC, Sajuthi S, Deford P, Christenson S, Rios CL, Montgomery MT, et al. COVID-19 Related Genes in Sputum Cells in Asthma: Relationship to Demographic Features and Corticosteroids. Am J Respir Crit Care Med. 2020.
51. Inhaled corticosteroids downregulate the SARS-CoV-2 receptor ACE2 in COPD through suppression of type I interferon - preliminary report. 13 June 2020 (pre-print).
52. Lan J, Ge J, Yu J, Shan S, Zhou H, Fan S, et al. Structure of the SARS-CoV-2 spike receptor-binding domain bound to the ACE2 receptor. Nature. 2020;581(7807):215-20.
53. Hoffmann M, Kleine-Weber H, Schroeder S, Krüger N, Herrler T, Erichsen S, et al. SARS-CoV-2 Cell Entry Depends on ACE2 and TMPRSS2 and Is Blocked by a Clinically Proven Protease Inhibitor. Cell. 2020;181(2):271-80.e8.
54. Matsuyama S, Kawase M, Nao N, Shirato K, Ujike M, Kamitani W, et al. The inhaled corticosteroid ciclesonide blocks coronavirus RNA replication by targeting viral NSP15. bioRxiv. 2020:2020.03.11.987016.
55. Yamaya M, Nishimura H, Deng X, Sugawara M, Watanabe O, Nomura K, et al. Inhibitory effects of glycopyrronium, formoterol, and budesonide on coronavirus HCoV-229E replication and cytokine production by primary cultures of human nasal and tracheal epithelial cells. Respir Investig. 2020.
56. Reyes AZ, Hu KA, Teperman J, Wampler Muskardin TL, Tardif J-C, Shah B, et al. Anti-inflammatory therapy for COVID-19 infection: the case for colchicine. Annals of the Rheumatic Diseases. 2020:annrheumdis-2020-219174.
57. Rodriguez-Nava G, Trelles-Garcia DP, Yanez-Bello MA, Chung CW, Trelles-Garcia VP, Friedman HJ. Atorvastatin associated with decreased hazard for death in COVID-19 patients admitted to an ICU: a retrospective cohort study. Crit Care. 2020;24(1):429.
58. Deftereos SG, Giannopoulos G, Vrachatis DA, Siasos GD, Giotaki SG, Gargalianos P, et al. Effect of Colchicine vs Standard Care on Cardiac and Inflammatory Biomarkers and Clinical Outcomes in Patients Hospitalized With Coronavirus Disease 2019: The GRECCO-19 Randomized Clinical Trial. JAMA Netw Open. 2020;3(6):e2013136.
59. Scarsi M, Piantoni S, Colombo E, Airo P, Richini D, Miclini M, et al. Association between treatment with colchicine and improved survival in a single-centre cohort of adult hospitalised patients with COVID-19 pneumonia and acute respiratory distress syndrome. Ann Rheum Dis. 2020;79(10):1286-9.
60. Lopes MIea. Beneficial effects of colchicine for moderate to severe COVID-19: an interim analysis of a randomized, double-blinded, placebo controlled clinical trial. medRxiv preprint. 2020.
61. Chiu L, Chow R, Chiu N, Lo C-H, Aggarwal R, Lee J, et al. Colchicine use in patients with COVID-19: a systematic review and meta-analysis. medRxiv. 2021:2021.02.02.21250960.
62. Tardif J-C, Bouabdallaoui N, L'Allier PL, Gaudet D, Shah B, Pillinger MH, et al. Efficacy of Colchicine in Non-Hospitalized Patients with COVID-19. medRxiv. 2021:2021.01.26.21250494.

© Copyright: The University of Oxford and Oxford University Hospitals NHS Foundation Trust 2019

63. Piantoni S, Colombo E, Airò P, Tincani A, Brucato A, Franceschini F, et al. The rationale for the use of colchicine in COVID-19: comments on the letter by Cumhur Cure M et al. Clin Rheumatol. 2020;39(8):2489-90.
64. Piantoni S, Colombo E, Furloni R, Andreoli L, Brucato A, Imazio M, et al. Anti-inflammatory action of colchicine in hospitalised patients with COVID-19. Response to: 'Colchicine treatment in community healthcare setting to prevent severe COVID-19' by Della-Torre <em>et al</em>. Annals of the Rheumatic Diseases. 2020:annrheumdis-2020-218806.
65. Piantoni S, Patroni A, Toniati P, Furloni R, Franceschini F, Andreoli L, et al. Why not to use colchicine in COVID-19? An oldanti-inflammatory drug for a novel auto-inflammatory disease. Rheumatology. 2020;59(7):1769-70.
66. Prieto-Pena D, Dasgupta B. Biologic agents and small-molecule inhibitors in systemic autoimmune conditions: an update. Pol Arch Intern Med. 2020.
67. Dalbeth N, Lauterio TJ, Wolfe HR. Mechanism of action of colchicine in the treatment of gout. Clin Ther. 2014;36(10):1465-79.
68. Rodrigues TS, de Sá KSG, Ishimoto AY, Becerra A, Oliveira S, Almeida L, et al. Inflammasomes are activated in response to SARS-CoV-2 infection and are associated with COVID-19 severity in patients. J Exp Med. 2021;218(3).
69. Stewart S, Yang KCK, Atkins K, Dalbeth N, Robinson PC. Adverse events during oral colchicine use: a systematic review and meta-analysis of randomised controlled trials. Arthritis Research & Therapy. 2020;22(1):28.
70. Levy M, Spino M, Read SE. Colchicine: a state-of-the-art review. Pharmacotherapy. 1991;11(3):196-211.
71. Chen C, Zhang Y, Huang J, Yin P, Cheng Z, Wu J, et al. Favipiravir versus Arbidol for COVID-19: A Randomized Clinical Trial. medRxiv. 2020:2020.03.17.20037432.
72. Udwadia ZF, Singh P, Barkate H, Patil S, Rangwala S, Pendse A, et al. Efficacy and Safety of Favipiravir, an Oral RNA-Dependent RNA Polymerase Inhibitor, in Mild-to-Moderate COVID-19: A Randomized, Comparative, Open-Label, Multicenter, Phase 3 Clinical Trial. Int J Infect Dis. 2020.
73. Ivashchenko AA, Dmitriev KA, Vostokova NV, Azarova VN, Blinow AA, Egorova AN, et al. AVIFAVIR for Treatment of Patients With Moderate Coronavirus Disease 2019 (COVID-19): Interim Results of a Phase II/III Multicenter Randomized Clinical Trial. Clinical Infectious Diseases. 2020.
74. Khamis F, Al Naabi H, Al Lawati A, Ambusaidi Z, Al Sharji M, Al Barwani U, et al. Randomized controlled open label trial on the use of favipiravir combined with inhaled interferon beta-1b in hospitalized patients with moderate to severe COVID-19 pneumonia. International Journal of Infectious Diseases. 2021;102:538-43.
75. Cai Q, Yang M, Liu D, Chen J, Shu D, Xia J, et al. Experimental Treatment with Favipiravir for COVID-19: An Open-Label Control Study. Engineering. 2020;6(10):1192-8.
76. Pilkington V, Pepperrell T, Hill A. A review of the safety of favipiravir – a potential treatment in the COVID-19 pandemic? Journal of Virus Eradication. 2020;6(2):45-51.
77. Sanders JM, Monogue ML, Jodlowski TZ, Cutrell JB. Pharmacologic Treatments for Coronavirus Disease 2019 (COVID-19): A Review. JAMA. 2020;323(18):1824-36.
78. Coomes EA, Haghbayan H. Favipiravir, an antiviral for COVID-19? Journal of Antimicrobial Chemotherapy. 2020;75(7):2013-4.
79. Arshad U, Pertinez H, Box H, Tatham L, Rajoli RK, Curley P, et al. Prioritisation of potential anti-SARS-CoV-2 drug repurposing opportunities based on ability to achieve adequate plasma and target site concentrations derived from their established human pharmacokinetics. medRxiv. 2020:2020.04.16.20068379.
80. Kaptein SJF, Jacobs S, Langendries L, Seldeslachts L, ter Horst S, Liesenborghs L, et al. Favipiravir at high doses has potent antiviral activity in SARS-CoV-2–infected hamsters, whereas hydroxychloroquine lacks activity. Proceedings of the National Academy of Sciences. 2020;117(43):26955-65.
81. Driouich J-S, Cochin M, Lingas G, Moureau G, Touret F, Petit PR, et al. Favipiravir antiviral efficacy against SARS-CoV-2 in a hamster model. bioRxiv. 2020:2020.07.07.191775.

© Copyright: The University of Oxford and Oxford University Hospitals NHS Foundation Trust 2019

82. Cevik M, Tate M, Lloyd O, Maraolo AE, Schafers J, Ho A. SARS-CoV-2, SARS-CoV, and MERS-CoV viral load dynamics, duration of viral shedding, and infectiousness: a systematic review and meta-analysis. The Lancet Microbe.

83. CDC. https://www.cdc.gov.tw/File/Get/ht8jUiB_MI-aKnlwstwzvw.

84. Sissoko D, Laouenan C, Folkesson E, M'Lebing AB, Beavogui AH, Baize S, et al. Experimental Treatment with Favipiravir for Ebola Virus Disease (the JIKI Trial): A Historically Controlled, Single-Arm Proof-of-Concept Trial in Guinea. PLoS Med. 2016;13(3):e1001967.

© Copyright: The University of Oxford and Oxford University Hospitals NHS Foundation Trust 2019

#### 16 APPENDIX A: SCHEDULE OF PROCEDURES

| **Procedures** | **Participant contacts** | | | | | | | |
|---|---|---|---|---|---|---|---|---|
| | **Visit timing Day 0** | **Day 0** | **Day 0** | **Day 0** | **Daily Day 1-28 incl** | **Day 28-12 months (monthly contact)** | **Day 29-12mths** | **Up to 10 years** |
| | **Screening completed by participant online/phone** | **Eligibility completed by participant online/phone** | **Baseline completed by participant online/phone** | **Eligibility completed by Clinician online/phone** | **Symptom Diaries completed by participant online/phone** | **Contacted by study team if consent provided** | **Retrospective data collection by study team** | **By data extraction from clinical records** |
| Informed consent | X | X | X | X | X | | | |
| Demographics | X | X | X | | | | X | |
| Medical history | X | X | X | X | | | X | |
| Swab as part of the RCGP RSC/PHE national surveillance programme | When available, preferably by self-swabbing at study entry | | | | | | | |
| Concomitant medications | | X | | | | | X | |
| Eligibility assessment | X | X | | | | | | |

© Copyright: The University of Oxford and Oxford University Hospitals NHS Foundation Trust 2019

| | | | | | | | | |
|---|---|---|---|---|---|---|---|---|
| Randomisation | | | | X | | | | |
| Dispensing of trial drugs | | | | X | X | | | |
| Questionnaire | | | | | X | X | | |
| WHO 5 Well Being Index | X | | | | Day 14 and day 28 | X | | |
| Telephone interview (for subset of patient participants) | | | | | X | | | |
| Compliance | | | | | X | | | |
| Adverse event assessments | | | | | X* | | X | |
| Optional SARS-CoV-2 blood test as part of the RCGP RSC/PHE national surveillance programme | | | | | | | X | |
| Evidence of sequalae and health care utilisation | | | | | | X | | X |

© Copyright: The University of Oxford and Oxford University Hospitals NHS Foundation Trust 2019

* Patient reported AEs will be assessed by a clinician for certain IMPs, as specified in the Intervention Specific Appendices. These treatments include HCQ (no longer an active treatment arm) and drugs that are not licensed for use in the UK.

© Copyright: The University of Oxford and Oxford University Hospitals NHS Foundation Trust 2019

#### 17 APPENDIX B: AMENDMENT HISTORY

| Amendment No. | Protocol Version No. | Date issued | Author(s) of changes | Details of Changes made |
|---|---|---|---|---|
| 1 (SA1) | 1.1 | | Emma Ogburn; Chris Butler; Gail Hayward | Inclusion criteria: change 'known heart disease' to 'Known heart disease and/or hypertension';<br>Exclusion criteria: exclude patients taking the following drugs: penicillamine, amiodarone, ciclosporin, chloroquine.<br>Update section 9.6 to include vision changes and lowering of blood sugar.<br>Update change in Funder and update Investigator list to reflect UKRI funder bid. |
| 2 (SA2) | 2.0 | | Emma Ogburn; Chris Butler; Gail Hayward, Hannah Swayze | Inclusion of TSC; central facility to distribute patient packs; addition of third arm; update of secondary outcomes to include WHO wellbeing questions; qualitative sub study; sign posting to other RCGP RSC study; eligibility confirmation by research nurse. |
| 3 (SA3) | 2.1 | | Hannah Swayze; Chris Butler; Emma Ogburn; Gail Hayward | Trial rationale; secondary outcomes to include blood test; 14 days of covid-19 symptoms; call to participant at day 2; poster |
| *4 (SA4)* | *2.1* | | *No changes to the protocol* | |
| 5 (SA5) | 3.0 | | Hannah Swayze; Chris Butler; Emma Ogburn; Gail Hayward | Updated Azithromycin information; broadening of inclusion criteria; first interim analysis; primary analysis details; care home materials; administrative and typographical updates; study partner letter; recruitment via social media, care homes and pharmacies; GPs prescribe trial medication; eligibility to at least one intervention arm as well as the |

© Copyright: The University of Oxford and Oxford University Hospitals NHS Foundation Trust 2019

| | | | | |
|---|---|---|---|---|
| | | | | Usual Care arm; ICF may be sent to participants. |
| 6 (SA6) | 4.0 | | Chris Butler; Emma Ogburn; Gail Hayward; Ben Saville; Ly-Mee Yu; Hannah Swayze | Updating inclusion criteria; updating the rationale and evidence for safety of hydroxychloroquine; inclusion of a new arm, doxycycline; AE reporting for hydroxychloroquine arm; typographical clarifications. |
| 7 (NS1) | 4.0 | | *No changes to the protocol* | |
| 8 (SA7) | 5.0 | | Chris Butler; Emma Ogburn; Ben Saville; Ly-Mee Yu; Hannah Swayze | Including a second primary outcome, time to recovery, change to sample size estimation, new eligibility criteria: obesity, formatting changes, blood test process. |
| 9 (SA8) | 5.0 | | *No changes to the protocol* | |
| 10 (SA9) | 5.0 | | *No changes to the protocol* | |
| 11 (NS2) | 5.0 | | *No changes to the protocol* | |
| 12 (SA10) | 6.0 | | Chris Butler; Emma Ogburn; Hannah Swayze | Addition of inhaled corticosteroid treatment arm, enrolment to additional trials, long-term follow-up, access to NHS Digital Pillar 2 test data, removal of investigators, additional trial contact with participants for up to 12 months, changes to objectives/outcomes/ time-points, removal of sampling from study |
| 13 (NS3) | 6.1 | | Sharon Tonner | Removal of patient already taking a treatment arm medication as an exclusion |
| 14 (NS4) | 6.1 | | *No changes to the protocol* | |
| 15 (SA11) | 6.2 | | Sharon Tonner, Hannah Swayze | Inclusion of patients who lack capacity to consent, discontinuation of azithromycin arm |
| 16 (SA12) | 7.0 | | Chris Butler; Emma Ogburn; Hannah Swayze; | Addition of colchicine treatment arm. Data management proportional approach. Discontinuation of doxycycline |

© Copyright: The University of Oxford and Oxford University Hospitals NHS Foundation Trust 2019

| | | | | |
|---|---|---|---|---|
| | | | Emily Bongard; Julie Allen; Jienchi Dorward; Oliver Van Hecke | arm. Stylistic and typographical updates. Funding arrangements. Inclusion criteria - to include those aged 18 or over. Rephrasing secondary outcome: daily rating of how well participant feels. Statistical analysis. |
| 17 (SA13) | 7.1 | | Chris Butler; Emma Ogburn; Hannah Swayze; Emily Bongard; Ly-Mee Yu; Jienchi Dorward | Addition of safety monitoring procedures for drugs that are not licensed in the UK. Addition of safety as a secondary endpoint. Addition of favipiravir treatment arm. Stylistic and typographical updates. GP Contact for safety monitoring. In-kind contributions. Using JDR as a recruitment tool. The primary analysis population defined as those with a COVID-19 positive test. Drug accountability for favipiravir. |
| 18 (SA14) | 8.0 | | Chris Butler; Emma Ogburn; Hannah Swayze; Emily Bongard; Jienchi Dorward; Ly-Mee Yu | Addition of Ivermectin treatment arm and safety reporting for this arm. Trial name change. Updated safety reporting for the colchicine arm.<br><br>Rejected by the MHRA |
| 18 (SA15) | 8.1 | | Chris Butler; Emma Ogburn; Hannah Swayze; Emily Bongard; Jienchi Dorward; Ly-Mee Yu | Addition of Ivermectin treatment arm and safety reporting for this arm. Trial name change. Updated safety reporting for the colchicine arm. SAE reporting clarification. Discontinuation of Budesonide arm. |

Lists details of all protocol amendments whenever a new version of the protocol is produced.

Protocol amendments must be submitted to the Sponsor for approval prior to submission to the REC committee, HRA (where required) or MHRA.

© Copyright: The University of Oxford and Oxford University Hospitals NHS Foundation Trust 2019

### 18 APPENDIX C: USUAL CARE ARM

#### 1. Background and rationale

COVID-19 disproportionately affects people with comorbidities, more severe illness, and who are older. The disease causes considerable morbidity and mortality in this population group in particular, and is having a devastating effect on people's health, and society in the UK and internationally.(2, 3, 6, 8) So far, there are no specific treatments for COVID-19 that have been proven in rigorous clinical trials to be effective and that can be used in the community. Clinicians managing possible COVID-19 in the community will make clinical judgements about best treatment based on the clinical situation, but care is usually supportive to begin with, unless patients deteriorate and require hospital admission https://www.nice.org.uk/guidance/ng163). The National Institute for Health and Care Excellence does not recommend the immediate use of antibiotics unless there are signs of pneumonia (https://www.nice.org.uk/guidance/ng163).

This Usual Care arm will follow current NHS care provision, and provides a control against which the effect of new interventions that are added to usual care can be assessed. If a new trial intervention plus usual care is found to be superior to usual care alone, then the usual care alone arm will be dropped, and the intervention that is found to be most effective will become the standard of care within the trial.

#### 2. Changes to outcome measures

None

#### 3. Detail of intervention

Participants randomised to the usual care arm will receive usual clinical care as per NHS care delivery practice.

##### a. Investigational Medicinal Product (IMP) description

Not applicable

##### b. Storage of IMP

Not applicable

#### 4. Safety reporting

Mechanisms for safety reporting are outlined in the trial protocol.

© Copyright: The University of Oxford and Oxford University Hospitals NHS Foundation Trust 2019

### 19 APPENDIX D: USUAL CARE PLUS HYDROXYCHLOROQUINE ARM (DISCONTINUED)

#### 1. Background and rationale

##### a. Evidence for potential Hydroxychloroquine benefits in COVID-19

A candidate intervention for COVID-19, a drug called hydroxychloroquine, has become available following early evaluation in some studies in China.(9, 10) Hydroxychloroquine is a hydroxylated version of the drug chloroquine.(10, 11) Both agents are commonly in use as anti-malarials, and are used in a variety of auto-immune diseases. They have received significant recent interest as potential modifiers of disease activity in COVID-19. (10, 12, 13) Hydroxychloroquine is already available within the NHS on prescription for other indications, and has a generally benign safety profile.(14) Chloroquine is available to buy in the UK over the counter in some formulations and is used as antimalarial prophylaxis and treatment.

Chloroquine is known to block virus infection by increasing endosomal pH required for virus/cell fusion, as well as interfering with the glycosylation of cellular receptors of SARS-CoV-2.(5) Besides its antiviral activity, chloroquine has an immune-modulating activity, which may synergistically enhance its antiviral effect *in vivo*.(11) Chloroquine is widely distributed in the whole body, including lungs, after oral administration.(10) The $EC_{90}$ value of chloroquine against the 2019-nCoV in Vero E6 cells was 6.90 μM in one study (9) which can be clinically achievable as demonstrated in the plasma of rheumatoid arthritis patients who received 500 mg administration.(14)

Hydroxychloroquine has been found to be effective against intracellular micro-organisms including malaria and intracellular bacteria *Coxiella burnetii* and *Tropheryma Whipplei*.(11) Both chloroquine and hydroxychloroquine have been shown to have *in vitro* antiviral activity against SARS coronavirus in a number of studies.(11) Most recently activity against SARSCOV2 was shown to be greater for hydroxychloroquine than chloroquine (15).

**Key publications that have relevance to the safety and rationale for use of hydroxychloroquine in the PRINCIPLE Trial:**

**1.** ***The Mahévas study*** was an observational study that assessed whether hydroxychloroquine reduced the need for transfer to ICU in patients already sick enough to be hospitalised.(16) It focussed on sicker patients with hypoxic pneumonia, some requiring ITU care. It did not find a difference in transfers to ICU. So the question and population in the Mahevas study are very different compared to PRINCIPLE. Most importantly, unlike PRINCIPLE, the Mahevas study is ***not a randomised clinical tri***al. Numbers were relatively small (n=181), and it is at high risk of bias due to the observational design.

***Regarding safety***, those receiving hydroxychloroquine were prescribed 600mg per day, whereas the dose in the PRINCIPLE trial is 400mg per day; 18% of those who received hydroxychloroquine in the Mahévas study were also on azithromycin (which can be arrhythmogenic), and this combination is not possible in PRINCIPLE because of the additive risk. Moreover, PRINCIPLE excludes several other drug combinations that could be arrhythmogenic. In the Mahevas study, eight patients (10%) who were taking hydroxychloroquine experienced electrocardiographic changes that required discontinuation of hydroxychloroquine. Critically, those in the control

© Copyright: The University of Oxford and Oxford University Hospitals NHS Foundation Trust 2019

group did not have ECGs done, so we don't know if there was indeed a difference between groups, and we cannot therefore attribute the ECG changes to hydroxychloroquine. COVID-19 itself, or drug interactions, may well have been underlying reasons. The authors state, "Although hydroxychloroquine is considered safe in the context of systemic lupus erythematosus, these adverse events might be explained by the use of high dose hydroxychloroquine in patients older than 75 years with renal impairment and frequent drug interactions. We cannot rule out the possibility that these cardiac effects attributed to hydroxychloroquine were caused by COVID-19, especially given electrocardiograms were unavailable during follow-up in the control group."

**2. *The Tang study*** was a hospital-based, randomised study and included 150 patients; randomisation was done using sealed envelopes.(17) The trial found no difference in the proportion of patients with two sequential negative swab results.

***Regarding safety***,75 participants received hydroxychloroquine 1200 mg daily for 3 days and then 800 mg for either 2 or 3 weeks. Again, the dose used in this study was much higher that the dose being used in PRINCIPLE (initially three times, and subsequently twice as high as PRINCIPLE). However, 63% and 64% of patients in the hydroxychloroquine and control groups respectively also received other antiviral agents. In PRINCIPLE, we are not evaluating the combination of antiviral agents and hydroxychloroquine. Importantly, this study did not find evidence of cardiac arrhythmias associated with hydroxychloroquine use. The authors sate, "Events of cardiac arrhythmia, such as prolonged QT interval were not observed in our trial, possibly because of the relatively mild to moderate disease of patients investigated or the short term period of follow-up."

**3. *The Mehra study*** published in the Lancet on 22.05.2020 reported an association between hydroxychloroquine use and cardiac events and mortality amongst patients hospitalised with COVID-19.(18) The *observational* study design is inherently susceptible to bias, the study data integrity has been queried given the homogeneity of the baseline characteristics, the adequacy of the adjustment for confounders cannot be assessed from the published methods, and the registries used are in a different patient population compared to PRINCIPLE. Patients were much sicker and more advanced in the illness than in PRINCIPLE. The authors themselves state that "*Randomised clinical trials will be required before any conclusion can be reached regarding benefit or harm of these agents (hydroxychloroquine and chloroquine) in COVID-19 patients*." The authors also state "These data do not apply to the use of any treatment regimen used in the ambulatory, out-of-hospital setting." This study has proved hugely controversial on social media, with a number of methodological and data integrity concerns already raised, for example:

1. There were inadequate adjustments for known and measured confounders (disease severity, temporal effects, site effects, dose used).
2. The authors have not adhered to standard practices in the machine learning and statistics community. They have not released their code or data. There is no data/code sharing and availability statement in the paper. The Lancet was among the many signatories on the Wellcome statement on data sharing for COVID 19 studies.
3. There was no ethics review.
4. There was no mention of the countries or hospitals that contributed to the data source, no acknowledgments to their contributions. A request to the authors for information on the contributing centres was denied.
5. Data from Australia are not compatible with government reports (too many cases for

© Copyright: The University of Oxford and Oxford University Hospitals NHS Foundation Trust 2019

just five hospitals, more in-hospital deaths than had occurred in the entire country during the study period). Surgisphere (the data company) have since claimed this was an error of classification.

6. Data from Africa indicate over 40% of all COVID-19 cases and deaths in the continent occurred in *Surgisphere*-associated hospitals which had sophisticated electronic patient data recording, and patient monitoring able to detect and record “non-sustained [at least 6 secs] or sustained ventricular tachycardia or ventricular fibrillation”. This seems unlikely.
7. Unusually small reported variances in baseline variables, interventions and outcomes between continents
8. Mean daily doses of hydroxychloroquine that are 100 mg higher than FDA recommendations, whilst 66% of the data are from North American hospitals.
9. Implausible ratios of chloroquine to hydroxychloroquine use in some continents.
10. The tight 95% confidence intervals reported for the hazard ratios are unlikely. For instance, for the Australian data this would need about double the numbers of recorded deaths that were reported in the paper.

This paper has now been retracted, and the data cannot be verified.

**4. *The Geleris study*** was an observational study of 1,376 consecutive COVID-19 patients at a New York hospital to determine whether hydroxychloroquine use was associated with intubation or death, as a primary composite outcome.(19) 811 (58.9%) of these patients received hydroxychloroquine. The authors excluded patients who were intubated, died, or who were transferred to another facility within 24 hours after presentation to the emergency department from the analyses. A propensity score matching model (C-statistic of 0.81) was used to ensure that groups were similar at baseline.

***Regarding safety***, multivariable adjusted analyses with inverse probability weighting revealed no significant association between treatment with hydroxychloroquine and intubation or death (HR 1.04 (95% CI 0.82 – 1.32)). Whilst the patient population in this study is different to that of PRINCIPLE, it is interesting that the findings contrast with those of a recent Lancet study published by Mehra *et al*. One possible reason for the difference is that patients receiving interventions like hydroxychloroquine in the study by Mehra *et al* were sicker than those in the study’s control group. This may have arisen through use of crude measures to account for baseline disease severity (qSOFA score and SpO2 < 94%) in their propensity score matching model, and may also explain the big differences seen in patients requiring mechanical ventilation between controls (7.7%) and those in intervention groups (20-21.6%).

**5. *Boulware*** and colleagues conducted a Covid-19 postexposure prophylaxis, placebo controlled randomised trial of hydroxychloroquine in 821 asymptomatic patients; 11.8% of those taking hydroxychloroquine vs 14.3 of those taking placebo experienced a new illness compatible with COVID-19 (absolute difference -2.4%) but this difference was not statistically significant, indicating no evidence of benefit from the hydroxychloroquine. (20)

***Regarding safety***, while side effects were more common with hydroxychloroquine than with placebo (40.1% vs. 16.8%), no serious adverse reactions were reported.

© Copyright: The University of Oxford and Oxford University Hospitals NHS Foundation Trust 2019

**Earlier studies of hydroxychloroquine for COVID-19**

**1. *Chen*** and colleagues conducted a *randomised* controlled trial to test the effectiveness of hydroxychloroquine in 30 adult patients who tested positive for COVID-19 in China.(21) Patients in the treatment group received 400mg of hydroxychloroquine for 5 days, while the control group received usual care. The result of a nasopharyngeal swab on Day 7 was used as the primary outcome. The intention- to- treat analysis revealed that the treatment group did not differ from the control group in the number of patients testing negative for COVID-19 on Day 7 (13 versus 14 patients), nor the duration of illness (all P>0.05).

***Regarding safety*,** the authors report three adverse events in the control group (one patient with abnormal liver function and anaemia, and one patient with abnormal renal function), and four adverse events in the treatment group (two patients with diarrhoea, one with lethargy, and one patient with abnormal liver function tests), which the authors argue were not linked to treatment with HCQ. One patient in the treatment group deteriorated significantly and thus HCQ was stopped on Day 4 of the treatment. This study was under-powered according to their own calculations.

***2. Gautret*** and colleagues presented the results of an open- label, non-randomised trial with 36 patients diagnosed with COVID-19 in French hospitals.(22) Six participants were asymptomatic, 22 had upper respiratory tract infection symptoms, and eight had lower respiratory tract infection symptoms. The twenty patients in the treatment group received HCQ 200mg three times a day for 10 days. Patients declining to take part in the study and not meeting the inclusion criteria were assigned to the control group and received usual care. Six of the patients in the treatment group additionally received azithromycin to prevent bacterial superinfection. The primary outcome was SARS- CoV-2 carriage at Day 6 on nasopharyngeal swabs. Patients treated with hydroxychloroquine were significantly more likely to test negative for SARS- CoV-2 on Day 6 compared with controls (70% versus 12.5% virologically cured, p<0.001). All patients treated with hydroxychloroquine and azithromycin tested negative on Day 6.

***Regarding safety*,** the authors did not report any safety data, stating that this would follow in a subsequent publication. Aside from a lack of adverse event reporting, there are many problems with the study methodology including the non-randomized design, under-powered sample size, lack of intention-to-treat analysis, and absence of medium to long-term follow-up data.

***3. Chen*** and colleagues conducted a *randomised* clinical trial of adult patients admitted to hospital with confirmed COVID-19.(7) Sixty two patients were randomly assigned to usual care (n=31) or hydroxychloroquine (200 mg BD) for five days in addition to usual care (n=31). The authors report that there were 'significant differences' in time to clinical recovery (TTCR) between the two groups, with TTCR defined as the return of body temperature and cough relief, maintained for more than 72 hours. They also report that all four patients who 'progressed to severe disease' were in the control group. The reporting of empirical data by the authors is limited and unclear. They did not include a power calculation, but presumably this study was under-powered to detect differences between groups. No medium to long-term follow-up data is presented.

***Regarding safety,*** the authors report that two mild adverse events occurred (a rash and a headache), both of which were in patients receiving hydroxychloroquine. No patients receiving usual care experienced adverse events.

© Copyright: The University of Oxford and Oxford University Hospitals NHS Foundation Trust 2019

**In summary**
The large scale hospital based Recovery trial has recently announced that they found no benefit from hydroxychloroquine (as yet unpublished). No safety concerns have been reported by the Principle Trial. A post exposure prophylaxis study found no benefit from hydroxychloroquine, but also found no safety concerns. These studies address a different research question and focus on different patient populations in comparison to the Principle Trial. Evidence about early treatment of COPVID-19 in the community is urgently needed: the potential application of the findings of the PRINCIPLE Trial of community treatment is considerable, and the 'reach' of the study is now nation-wide. Our study population are patients in the community and our trial question is about early treatment. Outcome data from studies with sicker hospitalised patients may not apply to our study population

A key, controversial observational study (Mehra et al) reported that those taking hydroxychloroquine had worse outcomes and suffered more cardiac events than those not taking hydroxychloroquine. However, major doubts have been expressed about the data integrity of this study and insufficient detail in the paper to judge the adequacy of the methods employed to adjust for the inevitable confounders in an observational study. Hydroxychloroquine is not a licensed drug for treating COVID-19. Patients doing well are therefore less likely to be prescribed this drug. When a patient is causing their clinical team more concern or their condition is deteriorating, the chances of them being prescribed hydroxychloroquine will be greater. Adjustment for potential confounders has been inadequate in the observational studies. Critically, these studies cannot adjust for the clinician's sense of how the patient is faring over time. The Mehra study has been retracted and can't be relied upon.

The deficiencies and differences in all of these studies highlight the need for well-conducted, adequately powered randomised clinical trials, to provide definitive evidence of the safety and effectiveness of hydroxychloroquine for the early community treatment COVID-19 illness. PRINCIPLE will assess whether hydroxychloroquine is safe and effective if given earlier in the course of illness and in patients with milder symptoms not requiring hospital admission.

#### 2. Eligibility criteria specifically related to hydroxychloroquine

Inclusion criteria:

Exclusion criteria:

- Pregnancy;
- Breastfeeding;
- Known severe hepatic impairment;
- Known severe renal impairment;
- Known porphyria;
- Type 1 diabetes or insulin dependent Type 2 Diabetes mellitus ;
- Known G6PD deficiency;
- Known myasthenia gravis;
- Known severe psoriasis;
- Known severe neurological disorders (especially those with a history of epilepsy—may lower seizure threshold)
- Previous adverse reaction to, or currently taking, hydroxychloroquine or chloroquine

© Copyright: The University of Oxford and Oxford University Hospitals NHS Foundation Trust 2019

Patients currently taking the following drugs: penicillamine, amiodarone, ciclosporin, digoxin: the following antimicrobials; azithromycin, clarithromycin, erythromycin, ciprofloxacin, levofloxacin, moxifloxacin, ketoconazole, itraconazole, or mefloquine: the following antidepressants; amitriptyline, citalopram, desipramine, escitalopram, imipramine, doxepin, fluoxetine, wellbutrin, venlafaxine; the following antipsychotics or mood stabilizers; haloperidol, droperidol, lithium, quetiapine, thioridazine, ziprasidone: methadone: sumatriptan, zolmitriptan

- Known congenital or documented QT prolongation
- Known retinal disease

#### 3. Outcome measures related to hydroxychloroquine

There are no outcome measures related specifically to this usual care plus hydroxychloroquine arm

#### 4. Detail of intervention

Participants randomised to the usual care plus hydroxychloroquine arm will receive usual clinical care as per NHS guidelines, plus a course of oral hydroxychloroquine 200mg twice daily for seven days.

##### a. Investigational Medicinal Product (IMP) description

Hydroxychloroquine sulphate 200 milligram (mg) tablets. The tablets are for oral administration. One tablet (200mg) hydroxychloroquine to be taken twice daily for 7 days by mouth (14 tablets in total).
Special instructions: Each dose should be taken with a meal or glass of milk. Antacids may reduce absorption of hydroxychloroquine so it is advised that a 4-hour interval be observed between taking hydroxychloroquine and an antacid.
This is the standard therapeutic dose for its normal indication in adults which is for the treatment of rheumatoid arthritis, discoid and systemic lupus erythematosus, and dermatological conditions caused or aggravated by sunlight.

The Marketing Authorisation holder is Zentiva Pharma UK Limited Guildford Surrey GU1 4YS United Kingdom. Marketing authorisation number is PL 17780/0748.

##### b. Storage of IMP

: Stored at room temperature in locked cupboards in restricted access rooms in the Nuffield Department of Primary Care Health Sciences; in locked cupboards in restricted access rooms in GP Practices; in Pharmacies.

For hydroxychloroquine, GP practices can order a supply of trial medication from Public Health England using the existing ImmForm process. GPs will be provided with an envelope by the trial team which will be labelled appropriately for trial medication, and they will add the patient's details to this label. This pack, containing instructions on using the medication will be provided to the patient or their representative.

© Copyright: The University of Oxford and Oxford University Hospitals NHS Foundation Trust 2019

##### c. SmPC precautions and concomitant medication

Hydroxychloroquine: Hydroxychloroquine will be used for short-term use (7 days) in this trial. The SmPC and precautions listed below focus on longer term chronic use.

###### *i.* Precautions

Hydroxychloroquine might lower blood sugar levels in some people. If participants develop these symptoms, they will be advised in the Patient Information documents to eat something sweet and seek clinical advice if the symptoms continue.

Hydroxychloroquine occasionally causes blurred vision, which typically resolves once the medication is stopped. Participants will be advised via the Participant Information documents that if they develop any problems with vision, they should stop taking the medication immediately, seek clinical advice, and not drive or operate any heavy machinery.

###### ii. Concomitant medication

Hydroxychloroquine sulfate has been reported to increase plasma digoxin levels. Serum digoxin levels should be closely monitored in participants receiving concomitant treatment.

Hydroxychloroquine sulfate may also be subject to several of the known interactions of chloroquine even though specific reports have not appeared. These include: potentiation of its direct blocking action at the neuromuscular junction by aminoglycoside antibiotics; inhibition of its metabolism by cimetidine which may increase plasma concentration of the antimalarial; antagonism of effect of neostigmine and pyridostigmine; reduction of the antibody response to primary immunisation with intradermal human diploid-cell rabies vaccine.

As with chloroquine, antacids may reduce absorption of hydroxychloroquine so it is advised that a four hour interval be observed between hydroxychloroquine and antacid dosaging.

As hydroxychloroquine may enhance the effects of a hypoglycaemic treatment, a decrease in doses of insulin or antidiabetic drugs may be required.

Halofantrine prolongs the QT interval and should not be administered with other drugs that have the potential to induce cardiac arrhythmias, including hydroxychloroquine. Also, there may be an increased risk of inducing ventricular arrhythmias if hydroxychloroquine is used concomitantly with other arrhythmogenic drugs, such as amiodarone and moxifloxacin.

An increased plasma ciclosporin level was reported when ciclosporin and hydroxychloroquine were co-administered.

Hydroxychloroquine can lower the convulsive threshold. Co-administration of hydroxychloroquine with other antimalarials known to lower the convulsion threshold (e.g. mefloquine) may increase the risk of convulsions. Also, the activity of anti-epileptic drugs might be impaired if co-administered with hydroxychloroquine. In a single-dose interaction study, chloroquine has been reported to reduce the bioavailability of praziquantel. It is not known if there is a similar effect when hydroxychloroquine and praziquantel are co-administered. Per extrapolation, due to the similarities in structure and pharmacokinetic parameters between hydroxychloroquine and chloroquine, a similar effect may be expected for hydroxychloroquine.

© Copyright: The University of Oxford and Oxford University Hospitals NHS Foundation Trust 2019

There is a theoretical risk of inhibition of intra-cellular α-galactosidase activity when hydroxychloroquine is co-administered with agalsidase.

##### iii. Pregnancy and Breastfeeding

A moderate amount of data on pregnant women (between 300 – 1000 pregnancy outcomes), including prospective studies in long-term use with large exposure, have not observed a significant increased risk of congenital malformations or poor pregnancy outcomes. Hydroxychloroquine crosses the placenta. Only limited non-clinical data are available for hydroxychloroquine, data on chloroquine have shown developmental toxicity at high supratherapeutic doses and a potential risk of genotoxicity in some test systems. Therefore, hydroxychloroquine sulfate should be avoided in pregnancy except when, in the judgement of the physician, the individual potential benefits outweigh the potential hazards. Careful consideration should be given to using hydroxychloroquine during lactation, since it has been shown to be excreted in small amounts in human breast milk, and it is known that infants are extremely sensitive to the toxic effects of 4-aminoquinolines.
Pregnancy and breastfeeding are exclusion criteria for the hydroxychloroquine arm of the PRINCIPLE trial.

#### 5. Safety reporting

Hydroxychloroquine: has a well-documented safety profile and is a commonly used medication in a primary care setting (see above).

Common symptoms of hydroxychloroquine include abdominal pain; appetite decreased; diarrhoea; emotional lability; headache; nausea; skin reactions; vision disorders; and vomiting.
Mechanisms for safety reporting are outlined in the trial protocol.

We will call all participants randomised to hydroxychloroquine on day 7 to ask about cardiovascular AEs. Our team of clinicians will review any AEs relating to cardiovascular symptoms from the day 7 call, and assess whether these may be related to hydroxychloroquine. If AEs are thought to be related and it's deemed necessary by the assessing clinician, the participant's GP will be contacted to arrange a face-to-face visit for further clinical evaluation.

© Copyright: The University of Oxford and Oxford University Hospitals NHS Foundation Trust 2019

### 20 APPENDIX E: USUAL CARE PLUS AZITHROMYCIN ARM (DISCONTINUED)

#### 1. Background and rationale

##### a. Evidence for potential Azithromycin benefits in COVID-19

Atypical macrolides, especially Azithromycin, have activities that may be beneficial in the treatment of COVID-19 patients, and especially those in the at-risk or age range of the PRINCIPLE trial.

Firstly, Azithromycin appears to have some anti-viral mechanisms. In COVID-19, Azithromycin appears to inhibit viral replication and therefore reduces shedding. In the small open observational trial of Gautret *et al* the addition of azithromycin to hydroxychloroquine (HCQ) (at 200 tds for 10 days) in 6 of the 14 HCQ subjects of the total 36 COVID-19 patients in the study significantly reduced viral shedding at 3 days to 15% (one subject) versus 70% in the HCQ arm and 95% in the indirect control arm, with no shedding at 6 days in the combination arm versus 50% and 90% respectively.(22) Azithromycin was also used in some Chinese observational and interventional studies.

Azithromycin has also been shown to be active ***in vitro*** against Zika and Ebola viruses,(23-25) and to prevent severe respiratory tract infections when administrated to patients suffering viral infection.(26) Inhibition of viral infections by azithromycin may be linked to its suppressive effect on the production of viral interferon.(27) Longer term administration of low dose azithromycin in COPD has been shown to suppress proinflammatory cytokine production, potentiate macrophage phagocytosis and anti-inflammatory cytokine expression.(28-30) Azithromycin use is also associated with a decrease in the expression of human HLA (human leukocyte antigen) complex molecules in the respiratory tract, including HLA-A, HLA-B, HLA-DPA1, HLA-DRA, HLA-DRB4.(31)

##### b. Importance of treating CAP or CAP risk in the elderly or immuno-compromised

An important secondary pathway to severe illness and death with COVID-19 may be secondary infection and sepsis in the immune-compromised state, especially secondary community or hospital acquired pneumonia. Older people are more susceptible to pneumonia because of comorbidities, a weakened immune system and are therefore more likely to die.(32) The onset of pneumonia in the elderly can often be rapid, and for severe pneumonia, the prognosis is poor: as many as one in five will die.(32) Severe pneumonia is more prevalent the older you are and in those with more serious underlying diseases.(33) The leading cause of death is respiratory insufficiency. Death has been shown to increase in those not responding to initial antimicrobials, and consequently, the initial selection of the agent is important.

Common causative organisms in the elderly admitted to the hospital with pneumonia include *Haemophilus influenza, Staphylococcus aureus, Streptococcus pneumoniae,* and *Mycoplasma pneumoniae*. In severe pneumonia, *S. aureus, Klebsiella pneumoniae*, and *Pseudomonas aeruginosa* have been identified as common causative organisms. Older patients often have polymicrobial infections, which may be a factor in non-responders. Assessment of 12,945 US

© Copyright: The University of Oxford and Oxford University Hospitals NHS Foundation Trust 2019

Medicare inpatients over 65 with pneumonia found that initial treatment with a second-generation cephalosporin plus macrolide ([HR, 0.71; 95% CI, 0.52-0.96), a non-pseudomonas third-generation cephalosporin plus a macrolide (HR, 0.74; 0.60-0.92), or a fluoroquinolone alone (HR, 0.64; 0.43-0.94) was associated with lower 30-day mortality.(34)

For CAP management NICE guidance currently recommends Amoxycillin 500mg tds combined with Clarithromycin 500mg bd for 5 days or, in penicillin sensitive, Clarithromycin 500mg bd for 5 days or Doxycycline 200mg stat then 100mg daily for the next 4 days. They also recommend starting therapy within 4 hours. The identification of the early stages of pneumonia in older patients can prove challenging since traditional symptoms and signs, including fever, may be lacking.

Azithromycin will have at least as broad a spectrum of action as clarithromycin in terms of bacterial infections and the additional potential anti-viral activity which has not been observed for other macrolides like Clarithromycin. It will also cover atypical organisms.

#### 2 Changes to outcome measures

The addition of this usual care plus azithromycin arm will not require any changes to outcome measures

#### 3 Eligibility criteria specifically related to azithromycin

Inclusion criteria: No changes

Exclusion criteria:

- Pregnancy
- Breastfeeding
- Known severe hepatic impairment;
- Known severe renal impairment;
- Known myasthenia gravis;
- Previous adverse reaction to, or currently taking, azithromycin or other macrolides or ketolides
- Patients taking the following drugs: hydroxychloroquine or chloroquine, sotalol, amiodarone, ciclosporin, digoxin, bromocriptine, cabergoline, ergotamine, ergometrine, methysergide or any ergot derivatives.
- Already taking antibiotics for an acute condition
- Known congenital or documented QT prolongation
- Known allergy to soya or peanut due to the risk of hypersensitivity reactions

#### 4 Detail of intervention

Participants randomised to the usual care plus azithromycin arm will receive usual clinical care as per NHS guidelines, plus a course of oral azithromycin 500mg daily for three days. We will use the IMP distribution methods described in the protocol to deliver IMP to participants.

© Copyright: The University of Oxford and Oxford University Hospitals NHS Foundation Trust 2019

##### a. Investigational Medicinal Product (IMP) description

Azithromycin 250mg capsules. Participants in this arm will take 500 mg (two capsules) once daily for 3 days. The capsules are for oral administration.

Special instructions:
Azithromycin must be taken at least 1 hour before or 2 hours after antacids as this affects overall bioavailability. Azithromycin must be taken at least 1 hour before or 2 hours after food.

The marketing authorisation holder is: Teva UK Limited, Brampton Road, Hampden Park, Eastbourne, East Sussex, BN22 9AG, UK.
Marketing authorisation number: PL 00289/1570

##### b. Storage of IMP

Azithromycin: Stored at room temperature in locked cupboards in restricted access rooms in the Nuffield Department of Primary Care Health Sciences; in locked cupboards in restricted access rooms in GP Practices; in Pharmacies.

##### c. SmPC precautions and concomitant medication

###### i. Precautions

Azithromycin is a commonly prescribed antibiotic with an established safety profile. The SmPC advises caution using azithromycin in the following conditions:
Elderly people with proarrhythmic conditions due to the risk of developing cardiac arrhythmia and torsades de pointes including patients with congenital or documented QT prolongation; receiving treatment with other active substances known to prolong QT interval such as anti-arrhythmics (e.g. amiodarone and sotalol), cisapride, and fluoroquinolones such as moxifloxacin and levofloxacin; known hypokalaemia and hypomagnesaemia; significant hepatic or renal impairment; patients with neurological or psychiatric disorders; myasthenia gravis. Azithromycin as other with the use of nearly all antibacterial agents, alters the normal flora of the colon leading to overgrowth of Clostridium difficile which can lead to Clostridium difficile associated diarrhoea.

###### ii. Concomitant medications

Effects of other medicinal products on azithromycin:

*Antacids*

In a pharmacokinetic study investigating the effects of simultaneous administration of antacids and azithromycin, no effect on overall bioavailability was seen, although the peak serum concentrations were reduced by approximately 25%. In patients receiving both azithromycin and antacids, the medicinal products should not be taken simultaneously. Azithromycin must be taken at least 1 hour before or 2 hours after the antacids.

Co-administration of azithromycin prolonged-release granules for oral suspension with a single 20 ml dose of co-magaldrox (aluminium hydroxide and magnesium hydroxide) did not affect the rate and extent of azithromycin absorption.

Co-administration of a 600 mg single dose of azithromycin and 400 mg efavirenz daily for 7 days did not result in any clinically significant pharmacokinetic interactions.

© Copyright: The University of Oxford and Oxford University Hospitals NHS Foundation Trust 2019

*Fluconazole*

Co-administration of a single dose of 1200 mg azithromycin did not alter the pharmacokinetics of a single dose of 800 mg fluconazole. Total exposure and half-life of azithromycin were unchanged by the coadministration of fluconazole, however, a clinically insignificant decrease in $C_{max}$ (18%) of azithromycin was observed.

*Nelfinavir*

Co-administration of azithromycin (1200 mg) and nelfinavir at steady state (750 mg three times daily) resulted in increased azithromycin concentrations. No clinically significant adverse effects were observed and no dose adjustment is required.

*Rifabutin*

Coadministration of azithromycin and rifabutin did not affect the serum concentrations of either medicinal product.

Neutropenia was observed in subjects receiving concomitant treatment of azithromycin and rifabutin. Although neutropenia has been associated with the use of rifabutin, a causal relationship to combination with azithromycin has not been established.

*Terfenadine*

Pharmacokinetic studies have reported no evidence of an interaction between azithromycin and terfenadine. There have been rare cases reported where the possibility of such an interaction could not be entirely excluded; however, there was no specific evidence that such an interaction had occurred.

*Cimetidine*

In a pharmacokinetic study investigating the effects of a single dose of cimetidine, given 2 hours before azithromycin, on the pharmacokinetics of azithromycin, no alteration of azithromycin pharmacokinetics was seen.

Effect of azithromycin on other medicinal products:
*Ergotamine derivatives*

Due to the theoretical possibility of ergotism, the concurrent use of azithromycin with ergot derivatives is not recommended.

*Digoxin and colchicine (P-gp substrates)*

Concomitant administration of macrolide antibiotics, including azithromycin, with P-glycoprotein substrates such as digoxin and colchicine, has been reported to result in increased serum levels of the P-glycoprotein substrate. Therefore, if azithromycin and P-gp substrates such as digoxin are administered concomitantly, the possibility of elevated serum concentrations of the substrate should be considered.

*Coumarin-Type Oral Anticoagulants*

In a pharmacokinetic interaction study, azithromycin did not alter the anticoagulant effect of a single 15-mg dose of warfarin administered to healthy volunteers. There have been reports received in the post-marketing period of potentiated anticoagulation subsequent to co-administration of azithromycin and coumarin-type oral anticoagulants. Although a causal relationship has not been established, consideration should be given to the frequency of

© Copyright: The University of Oxford and Oxford University Hospitals NHS Foundation Trust 2019

monitoring prothrombin time when azithromycin is used in patients receiving coumarin-type oral anticoagulants.

*Cyclosporin*

In a pharmacokinetic study with healthy volunteers that were administered a 500 mg/day oral dose of azithromycin for 3 days and were then administered a single 10 mg/kg oral dose of cyclosporin, the resulting cyclosporin $C_{max}$ and $AUC_{0-5}$ were found to be significantly elevated. Consequently, caution should be exercised before considering concurrent administration of these drugs. If coadministration of these drugs is necessary, cyclosporin levels should be monitored and the dose adjusted accordingly.

*Theophylline*

There is no evidence of a clinically significant pharmacokinetic interaction when azithromycin and theophylline are co-administered to healthy volunteers. As interactions of other macrolides with theophylline have been reported, alertness to signs that indicate a rise in theophylline levels is advised.

*Trimethoprim/sulfamethoxazole*

Coadministration of trimethoprim/sulfamethoxazole DS (160 mg/800 mg) for 7 days with azithromycin 1200 mg on Day 7 had no significant effect on peak concentrations total exposure or urinary excretion of either trimethoprim or sulfamethoxazole. Azithromycin serum concentrations were similar to those seen in other studies.

*Zidovudine*

Single 1000 mg doses and multiple 1200 mg or 600 mg doses of azithromycin had little effect on the plasma pharmacokinetics or urinary excretion of zidovudine or its glucuronide metabolite. However, administration of azithromycin increased the concentrations of phosphorylated zidovudine, the clinically active metabolite, in peripheral blood mononuclear cells. The clinical significance of this finding is unclear, but it may be of benefit to patients.

Azithromycin does not interact significantly with the hepatic cytochrome P450 system. It is not believed to undergo the pharmacokinetic drug interactions as seen with erythromycin and other macrolides. Hepatic cytochrome P450 induction or inactivation via cytochrome-metabolite complex does not occur with azithromycin.

*Astemizole, alfentanil*

There are no known data on interactions with astemizole or alfentanil. Caution is advised in the co-administration of these medicines with azithromycin because of the known enhancing effect of these medicines when used concurrently with the macrolid antibiotic erythromycin.

*Atorvastatin*

Coadministration of atorvastatin (10 mg daily) and azithromycin (500 mg daily) did not alter the plasma concentrations of atorvastatin (based on a HMG CoA-reductase inhibition assay).

However, post-marketing cases of rhabdomyolysis in patients receiving azithromycin with statins have been reported.

*Carbamazepine*

© Copyright: The University of Oxford and Oxford University Hospitals NHS Foundation Trust 2019

In a pharmacokinetic interaction study in healthy volunteers, no significant effect was observed on the plasma levels of carbamazepine or its active metabolite in patients receiving concomitant azithromycin.

*Cisapride*

Cisapride is metabolized in the liver by the enzyme CYP 3A4. Because macrolides inhibit this enzyme, concomitant administration of cisapride may cause the increase of QT interval prolongation, ventricular arrhythmias and torsades de pointes.

*Cetirizine*

In healthy volunteers, coadministration of a 5-day regimen of azithromycin with cetirizine 20 mg at steady-state resulted in no pharmacokinetic interaction and no significant changes in the QT interval.

*Didanosins (Dideoxyinosine)*

Coadministration of 1200 mg/day azithromycin with 400 mg/day didanosine in 6 HIV-positive subjects did not appear to affect the steady-state pharmacokinetics of didanosine as compared with placebo.

*Efavirenz*

Coadministration of a 600 mg single dose of azithromycin and 400 mg efavirenz daily for 7 days did not result in any clinically significant pharmacokinetic interactions.

*Indinavir*

Coadministration of a single dose of 1200 mg azithromycin had no statistically significant effect on the pharmacokinetics of indinavir administered as 800 mg three times daily for 5 days.

*Methylprednisolone*

In a pharmacokinetic interaction study in healthy volunteers, azithromycin had no significant effect on the pharmacokinetics of methylprednisolone.

*Midazolam*

In healthy volunteers, coadministration of azithromycin 500 mg/day for 3 days did not cause clinically significant changes in the pharmacokinetics and pharmacodynamics of a single 15 mg dose of midazolam.

*Sildenafil*

In normal healthy male volunteers, there was no evidence of an effect of azithromycin (500 mg daily for 3 days) on the AUC and $C_{max}$ of sildenafil or its major circulating metabolite.

*Triazolam*

In 14 healthy volunteers, coadministration of azithromycin 500 mg on Day 1 and 250 mg on Day 2 with 0.125 mg triazolam on Day 2 had no significant effect on any of the pharmacokinetic variables for triazolam compared to triazolam and placebo.

© Copyright: The University of Oxford and Oxford University Hospitals NHS Foundation Trust 2019

##### iii. Fertility, pregnancy and lactation

*Pregnancy*

There are no adequate data from the use of azithromycin in pregnant women. In reproduction toxicity studies in animals azithromycin was shown to pass the placenta, but no teratogenic effects were observed. The safety of azithromycin has not been confirmed with regard to the use of the active substance during pregnancy. Therefore azithromycin should only be used during pregnancy if the benefit outweighs the risk.

#### 5 Safety reporting

Mechanisms for safety reporting are outlined in the protocol. In brief, we will collect symptoms and side effects of azithromycin from symptom diaries and participant telephone calls.

Common symptoms of azithromycin include diarrhoea, abdominal pain, nausea and flatulence. It may also cause headache, dizziness, insomnia, altered taste, pins and needles, changes in vision or hearing, rash, itching, joint pains or fatigue.

© Copyright: The University of Oxford and Oxford University Hospitals NHS Foundation Trust 2019

### 21 APPENDIX F: USUAL CARE PLUS DOXYCYCLINE ARM (DISCONTINUED)

#### 1. Background and rationale

##### a. Evidence for potential doxycycline benefits in COVID-19

Doxycycline may be beneficial in the treatment of COVID-19 patients, and especially those in the at-risk or age range of the PRINCIPLE trial.
The rationale for testing doxycycline is based on three reasons:
Firstly, doxycycline may have direct antiviral activity against SARS-CoV-2 based on computer modelling. Analysing all the proteins encoded by SARS-CoV-2 genes and then predicting potential targets by performing target-based virtual ligand screening, doxycycline ranked in the group of compounds with the highest binding affinity to 3CLpro (3-chymotrypsin-like protease). 3CLpro is the main protease in SARS- CoV-2 which is critical in the life-cycle of the virus (35).

Secondly, doxycycline has known anti-inflammatory effects in various human diseases by inhibiting mitogen-activated protein kinase (MAPK) and SMAD pathways (36), as well as potent antioxidant properties(37). Doxycycline reduces the hyperinflammation associated with severe COVID-19 by antagonising metalloproteinases such as MMP9 that are linked with lung injury, including SARS and ARDS(38).

Lastly, from extensive experience in other infectious diseases, doxycycline has broad antimicrobial activity and is efficacious against a broad spectrum of bacteria including atypical bacteria and other pathogens including intracellular plasmodia, chlamydia, rickettsia, and RNA viruses like Dengue fever and chikungunya.

##### b. Importance of treating CAP or CAP risk in the elderly or immuno-compromised

An important secondary pathway to severe illness and death with COVID-19 may be secondary infection and sepsis in the immune-compromised state, especially secondary community or hospital acquired pneumonia. Older people are more susceptible to pneumonia because of comorbidities, a weakened immune system and are therefore more likely to die.(32) The onset of pneumonia in the elderly can often be rapid, and for severe pneumonia, the prognosis is poor: as many as one in five will die.(32) Severe pneumonia is more prevalent the older you are and in those with more serious underlying diseases.(33) The leading cause of death is respiratory insufficiency. Death has been shown to increase in those not responding to initial antimicrobials, and consequently, the initial selection of the agent is important. Common causative organisms in the elderly admitted to the hospital with pneumonia include *Haemophilus influenza, Staphylococcus aureus, Streptococcus pneumoniae* and less commonly, atypical organisms, such as *Mycoplasma pneumoniae* and *Klebsiella pneumoniae*. All these organisms fall under doxycycline's antimicrobial spectrum.

We are aware that currently NICE, in their COVID-19 rapid guideline, advocates that clinicians offer oral doxycycline for treatment of suspected pneumonia in people who can or wish to be

© Copyright: The University of Oxford and Oxford University Hospitals NHS Foundation Trust 2019

treated in the community if: the likely cause is bacterial or; it is unclear whether the cause is bacterial or viral and symptoms are more concerning or; they are at high risk of complications (older or frail patients, pre-existing comorbidity or have a history of severe illness following previous lung infection).(39) Doxycycline will have at least as broad a spectrum of action as azithromycin in terms of bacterial infections with the potential anti-viral and anti-inflammatory effects.

Doxycycline for acute cough and community acquired pneumonia is recommended in the British National Formulary at a dose of Doxycycline 200mg stat then 100mg daily for the next 4 days. However, its use in COVID-19 is not proven and therefore important to address in this trial. Given the potential anti-inflammatory properties of doxycycline, we will use a slightly extended 7 day course.

#### 2. Changes to outcome measures

The addition of this usual care plus doxycycline arm will not require any changes to outcome measures

#### 3. Eligibility criteria specifically related to doxycycline

Inclusion criteria: No changes
Exclusion criteria:

- Pregnancy
- Breastfeeding
- Myasthenia gravis
- Systemic lupus erythematosus
- Previous adverse reaction to, or currently taking, doxycycline or other tetracyclines
- Sucrose intolerance (i.e. rare hereditary problems of fructose intolerance, glucose galactose malabsorption or sucrose-isomaltase insufficiency)
- Already taking antibiotics for an acute condition
- Patients taking the following drugs: ciclosporin, retinoids (acitretin, alitretinoin, isotretinoin, tretinoin), methotrexate, ergotamine, methoxyflurane, lithium.

#### 4. Detail of intervention

Participants randomised to the usual care plus doxycycline arm will receive usual clinical care as per NHS guidelines, plus a course of oral doxycycline for 7 days. We will use the IMP distribution methods described in the protocol to deliver IMP to participants.

##### a. Investigational Medicinal Product (IMP) description

Doxycycline 100mg capsules. Participants in this arm will take 200mg on the first day (as a single dose or in divided doses with a twelve hour interval) followed by 100mg a day for 6 days (7 day course in total). The capsules are for oral administration.

© Copyright: The University of Oxford and Oxford University Hospitals NHS Foundation Trust 2019

Special instructions:
Capsules should be swallowed whole with plenty of fluid, while sitting or standing. Capsules should be taken during meals, well before going to bed. Due to the risk of photosensitivity, patients should be advised to avoid exposure to sunlight or sun lamps.

The marketing authorisation holder is:

Accord-UK Ltd (Trading style: Accord), Whiddon Valley, Barnstaple, Devon, EX32 8NS
Marketing authorisation number: PL 0142/0407

##### b. Storage of IMP

Doxycycline: Stored at room temperature in locked cupboards in restricted access rooms in the Nuffield Department of Primary Care Health Sciences; in locked cupboards in restricted access rooms in GP Practices; in Pharmacies.

##### c. SmPC precautions and concomitant medication

###### i. Precautions

Doxycycline is a commonly prescribed antibiotic with an established safety profile. The SmPC states that in elderly patients "doxycycline may be prescribed in the usual dose with no special precautions. No dosage adjustment is necessary in the presence of renal impairment".

###### ii. Concomitant medications

*Warfarin*
There have been reports of prolonged prothrombin time in patients taking warfarin and doxycycline. Tetracyclines depress plasma prothrombin activity and reduced dosage of concomitant anti-coagulants may be necessary

#### 5. Safety reporting

Mechanisms for safety reporting are outlined in the protocol. In brief, we will collect symptoms and side effects from symptom diaries and participant telephone calls.

Common side effects of doxycycline include: Angioedema; diarrhoea; headache; Henoch-Schönlein purpura; hypersensitivity; nausea/vomiting; pericarditis; skin and photosensitivity reaction; dyspnoea; hypotension; peripheral oedema; tachycardia.

© Copyright: The University of Oxford and Oxford University Hospitals NHS Foundation Trust 2019

### 22 APPENDIX G: USUAL CARE PLUS INHALED CORTICOSTEROID (ICS) ARM (DISCONTINUED)

#### 1. Background and rationale

##### a. Evidence for potential benefits of inhaled corticosteroids in COVID-19 illness

Inhaled corticosteroids (ICS) are a commonly prescribed class of medication throughout the world. They are reasonably cheap and have been used widely for the last 60 years. The inhaled action and type2 pneumocyte target of COVID make ICS a potential therapeutic agent in COVID-19(40). They have been shown to be very effective in improving asthma and COPD care over the long term, where the recommendation is that most, if not all, patients with asthma should be prescribed an inhaled corticosteroid(41),(42) and up to 90% of patients with COPD in the UK are prescribed ICS(43). The rationale of ICS is to reduce the inflammatory process that underlies exacerbations, which can be triggered by viruses in asthma and COPD. Systemic corticosteroids have been found to be effective at reducing mortality amongst hospitalised patients with COVID-19 [46, 47], but it is not known whether pre-hospital treatment with ICS is also beneficial.

Further evidence is as described below:

**Evidence from the ARDS literature**
ICS in patients at risk of acute respiratory distress syndrome (ARDS) have been shown to improve physiology and reduce inflammatory markers(44). In patients admitted to hospital at risk of ARDS or acute lung injury, there was an almost 50% reduction of ARDS in patients that were using ICS pre-admission, even controlling for covariates such as age, gender and chronic respiratory disease(45). Moreover, this ICS effect can also be seen to improve pulmonary physiology(46).

**Potential mechanism of efficacy**
Recently published *in vitro* data suggest a role for ICS inhibition of coronavirus replication in infected epithelial cells(47), whilst there is an indication that there is accelerated hyperinflammation at the onset of SARS-CoV-2 infection(48), which potentially can be modified by anti-inflammatory therapy. This suggests a plausible mechanism for ICS efficacy against COVID-19 in which ICS has a dual role: firstly, toning down the inflammatory “runaway train” (ARDS-like) response affecting a minority of COVID-19 patients; and secondly, inhibiting viral replication. It has long been known that the ICS effect on epithelial cells is as a direct consequence of gene transcription(49), and investigation of gene expression of ACE2 and TMPRSS2 in the sputum of asthmatic patients has very recently demonstrated lower expression of these key receptors in the presence of ICS(50). Furthermore, ICS attenuates expression of the ACE2 receptor in human and murine in vitro and in vivo models(51). This is of relevance as the SARS-CoV-2 mechanism of action is upon direct action of the ACE2 receptor, a receptor highly expressed on epithelial cells in the oral mucosa and type 2 alveolar cells and the serine protease TMPRSS2 for SARS-CoV-2 spike protein priming(52, 53). Furthermore, there is experimental evidence that inhaled corticosteroids inhibit coronavirus replication in vitro(54, 55). SARS-CoV-2 binds to cells via the angiotensin converting enzyme 2 (ACE2) receptor. ACE2 is highly expressed on epithelial cells in the oral mucosa and type 2 alveolar epithelial cells. The use of inhaled corticosteroids as a therapy suggests it would target the cells of interest. Furthermore, the primary action of the inhaled steroids is on the type 2 pneumocytes where viral replication is going to be at its most, where we know that ACE2 receptor expression is high.

© Copyright: The University of Oxford and Oxford University Hospitals NHS Foundation Trust 2019

#### 2. Changes to outcome measures

The addition of this arm will not require any changes to outcome measures.

#### 3. Eligibility criteria specifically related to ICS

Inclusion criteria:

Age criteria: Patients aged ≥65 years, or Patients aged 50-64 years and meeting at least one of the following criteria:

- Known weakened immune system due to a serious illness or medication (e.g. chemotherapy);
- Known heart disease and/or a diagnosis of high blood pressure;
- Known asthma or lung disease;
- Known diabetes;
- Known mild hepatic impairment;
- Known stroke or neurological problem;
- Self-report obesity or body mass index ≥35 kg/m$^2$

Exclusion criteria:

- A known allergy to inhaled corticosteroids
- Any known contraindication to inhaled corticosteroids (as per SmPC, *patients with rare hereditary problems of galactose intolerance, the Lapp lactase deficiency or glucose-galactose malabsorption should not take this medicine. Lactose, the excipient in the product, contains small amounts of milk proteins and can therefore cause allergic reactions*).
- Patient currently prescribed inhaled or systemic corticosteroids
- Unable to administer inhaler

#### 4. Detail of intervention

Participants randomised to the usual care plus ICS arm will receive usual clinical care as per NHS guidelines, plus inhaled corticosteroids for 14 days. We will use the IMP distribution methods described in the protocol to deliver IMP to participants.

##### a. Investigational Medicinal Product (IMP) description

The IMP is the inhaled corticosteroid budesonide (dose 400mcg, Pulmicort turbohaler®). Inhaled budesonide comes in a polyethylene container consisting of a white cover screwed onto a brown bottom plate. Inside this is the inhaler with its main parts: a mouthpiece, a dosing mechanism and a substance store. The device will have 50 actuations of 400mcg/actuation. This product has marketing authorisation in the UK (PL 17901/0164) and is manufactured by AstraZeneca UK Ltd, 600 Capability Green, Luton, LU1 3LU, UK. This IMP will be taken as 2 puffs twice a day for 14 days.

##### b. Storage of IMP

© Copyright: The University of Oxford and Oxford University Hospitals NHS Foundation Trust 2019

Stored at room temperature in locked cupboards in restricted access rooms in the Nuffield Department of Primary Care Health Sciences; in locked cupboards in restricted access rooms in GP practices; in Pharmacies

##### c. SmPC precautions and concomitant medication

###### i. SmPC Precautions

Budesonide is a commonly prescribed inhaled steroid with an established safety profile.

###### ii. Concomitant medications

Largely, there is no restriction to concomitant medications using inhaled budesonide. The SmPC states that concomitant treatment with ketoconazole, HIV protease inhibitors or other potent CYP3A inhibitors may increase systemic budesonide levels, but that this is of little clinical significance for a short term treatment of 2 weeks, which is the duration of IMP use in the trial.

#### 5. Safety reporting

Mechanisms for safety reporting are outlined in the protocol. In brief, we will collect symptoms and side effects from symptom diaries and participant telephone calls.

Common and/or potential side effects from IMP include:

- Cough immediately after inhaling
- Mouth and throat pain
- Hoarse voice
- Oral candidiasis (thrush)

These are all reversible upon ceasing IMP.

© Copyright: The University of Oxford and Oxford University Hospitals NHS Foundation Trust 2019

### 23 APPENDIX H: USUAL CARE PLUS COLCHICINE

#### 1. Background and rationale

##### a. Evidence for potential benefits of colchicine in COVID-19 illness

Colchicine is licenced and widely used in the UK for the treatment of acute gout and has been investigated as a possible treatment for COVID-19. Reyes and colleagues (56) have summarised existing clinical evidence for colchicine for COVID-19 thus:

"A retrospective single-centre study of 87 ICU patients with COVID-19 demonstrated a lower risk of death in patients on colchicine (adjusted HR 0.41, 95% CI 0.17 to 0.98).(57) The Greek Effects of Colchicine in COVID-19 (GRECCO-19) trial was the first prospective open-label randomised trial evaluating colchicine versus usual care in early hospitalised patients. This study of 105 patients found a significant reduction in the primary clinical outcome of a two-point deterioration on WHO disease severity scale.(58) An Italian study compared 122 hospitalised patients who received colchicine plus standard-of-care (lopinavir/ritonavir, dexamethasone or hydroxychloroquine) with 140 hospitalised patients receiving standard-of-care alone. Colchicine had a significant mortality benefit versus controls (84% vs 64% survival).(59) A third prospective study randomised 38 hospitalised COVID-19 patients to colchicine or placebo in a double-blinded manner.(60) Patients receiving colchicine had less need for supplemental oxygen at day 7 (6% vs 39%) and were more likely to be discharged at day 10 (94% vs 83%). Colchicine subjects also had greater reduction of CRP, and no increase in serious adverse events."

More recently, a systematic review and meta-analysis (in preprint) supports the notion that colchicine lowers the risk of mortality (HR of 0.25, 95% CI [0.09, 0.66], six studies, n=5,033) However, the summary point estimate from the three included RCTs showed a signal towards mortality benefit that was not statistically significant among patients receiving colchicine versus placebo (OR 0.49, 95% CI [0.20, 1.24]).(61)

The COLCORONA randomised clinical trial has now reported in a pre-print.(62) It randomised 4488 patients to treatment with colchicine (0.5 mg twice daily for 3 days and once daily thereafter) or placebo for 28 days. The primary endpoint occurred in 4.7% of the patients in the colchicine group and 5.8% of those in the placebo group (odds ratio, 0.79; 95.1% confidence interval (CI), 0.61 to 1.03; P=0.08). Among the 4159 patients with PCR-confirmed COVID-19, the primary endpoint occurred in 4.6% and 6.0% of patients in the colchicine and placebo groups, respectively (odds ratio, 0.75; 95% CI, 0.57 to 0.99; P=0.04). In these patients with PCR-confirmed COVID-19, the odds ratios were 0.75 (95% CI, 0.57 to 0.99) for hospitalization due to COVID-19, 0.50 (95% CI, 0.23 to 1.07) for mechanical ventilation, and 0.56 (95% CI, 0.19 to 1.66) for death. Serious adverse events were reported in 4.9% and 6.3% in the colchicine and placebo groups (P=0.05); pneumonia occurred in 2.9% and 4.1% of patients (P=0.02). Diarrhoea was reported in 13.7% and 7.3% in the colchicine and placebo groups (P<0.0001).

This large-scale study of early treatment in those 40 years and over with symptoms of more severe illness or comorbidity suggests that colchicine treatment early on in the illness reduces the need for hospitalisation or COVID-19. However, the study did not assess impact on recovery, so we

© Copyright: The University of Oxford and Oxford University Hospitals NHS Foundation Trust 2019

don't know from this study if colchicine reduced symptom burden. This is important as those receiving colchicine, predictably, experienced more gastrointestinal side-effects. The study did not recruit to target and some of the findings are not statistically significant. However, these findings need replication before this drug can be considered for routine use for COVID-19.

Another Phase 3 trial in Canadian pre-hospital and hospital settings is investigating colchicine paired with aspirin or interferon beta (ACTCOVID). The trial is still in progress (www2.phri.ca/ACT-COVID-19/).

##### *b.* Potential mechanism of action

Colchicine is a broad-spectrum anti-inflammatory agent.(63-66) Colchicine inhibits cellular transport and mitosis by binding to tubulin and preventing its polymerisation as part of the cytoskeleton transport system.(67) Several of the biological therapies that have been studied and/or used in the setting of severe COVID-19 target some of the same pathways as colchicine, including IL-1β (ie, anakinra) and IL-6 (ie, tocilizumab and sarilumab). Colchicine differs from these agents in having pleotropic mechanisms of action, being less potent on any single target, and being an oral agent. Potential benefits of colchicine compared to these biological therapies when used in the midst of cytokine storm, are that colchicine is not immunosuppressive, is not known to increase risk of infection, and is inexpensive.

There is evidence that the inflammasome is activated in COVID-19 and that the degree of activation is correlated with disease severity.(68) Inflammasomes are key components of effective host immune responses to pathogens. Excessive inflammasome activation (specifically NLRP3 inflammasome) is implicated in chronic inflammatory diseases such as inflammatory bowel disease, rheumatoid arthritis and gout, and with ARDS and ALI (acute lung injury) pathology following respiratory viral infections. Additionally, colchicine may have relevance to COVID-19 associated inflammatory pathology that include: inhibition of neutrophil chemotaxis in response to cytokines, inhibition of NFkB activation (a protein complex that controls transcription of DNA, cytokine production and cell survival) or expression, inhibition of neutrophil adhesion to endothelium, inhibition of neutrophil respiratory burst and reactive oxygen species generation, reduced TNF receptor expression on macrophages and endothelial cells and increased TGFβ expression(67) Of note though is that many of these latter actions of colchicine occur at much lower concentrations that are required for NLRP3 inflammasome activation in response to MSU crystals. Symptoms such as fever, joint and muscle ache, and headache may be ameliorated by a general anti-inflammatory action.

Therefore, when used early in the course of COVID-19, colchicine may prevent the progression from inflammatory activation to a hyperinflammatory state.  The potential benefits of colchicine may therefore be maximised when used in the community, where earlier treatment could alleviate symptom burden, and prevent disease progression, hospitalisation and adverse outcomes.

#### 2. Changes to outcome measures

The addition of the usual care plus colchicine arm will not require any changes to outcome measures.

© Copyright: The University of Oxford and Oxford University Hospitals NHS Foundation Trust 2019

##### 3. Eligibility criteria specifically related to colchicine

Inclusion criteria: No changes required

Exclusion criteria:

- Hypersensitivity to the active substance or to any of the excipients listed in section (Lactose, Pregelatinised Maize Starch, Stearic Acid, Purified Talc, Purified Water, Ethanol 96%)
- Known or suspected pregnancy
- Breastfeeding
- Women of childbearing potential (premenopausal female that is anatomically and physiologically capable of becoming pregnant*) and not prepared to use highly effective contraception for the 28 day duration of follow up in the study**
- Known blood dyscrasias
- Known severe renal impairment or requiring dialysis
- Known severe hepatic impairment
- Currently taking any of the following drugs: colchicine, clarithromycin, erythromycin, ketoconazole, itraconazole, voriconazole, HIV protease inhibitors (e.g. ritonavir, atazanavir), cobicistat, verapamil, diltiazem, cyclosporin, quinidine, disulfiram, grapefruit juice
- Inflammatory bowel disease or chronic diarrhoea

* As recorded by the participant on the screening form and confirmed on Day 3 telephone call

***Highly effective methods have typical-use failure rates of less than 1% and include male or female sterilisation and long-acting reversible contraceptive (LARC) methods (intrauterine devices and implants). Women using another method of contraception, such as a combined hormonal method, progestogen only pill or injection, are eligible **if** they are willing to use an additional barrier method (e.g. male condom) for the 28 day duration of follow-up in the trial.*

The Patient Information Sheet Appendix, which the participant must read prior to providing informed consent, will clearly state the exclusion criteria listed above and the participant will be asked if they meet any of these exclusion criteria at the screening stage of the trial. The assessing clinician will then review the participant's responses against their medical record to confirm eligibility.

##### 4. Detail of intervention

Participants randomised to the usual care plus colchicine arm will receive usual clinical care as per NHS guidelines, plus 500 micrograms of colchicine to be taken each day for 14 days. We will use the IMP distribution methods described in the protocol to deliver IMP to participants.

Pharmacokinetic modelling shows that for a given dose of colchicine plasma exposure is greater in older (especially female) people than younger people; the target patient population for this study will be predominantly older subjects. Colchicine is also subject to accumulation in leucocytes- a target cell in this COVID19. Taking these factors into account and given the relatively narrow therapeutic index of colchicine, the dosing regimen will be 500 microgrammes daily for

© Copyright: The University of Oxford and Oxford University Hospitals NHS Foundation Trust 2019

14 days. A loading dose such as that used in the COLCORONA study and the use of a higher daily dose increases the risk of dose related adverse drug reactions (ADRs). Given the outpatient setting for this study with limited opportunity for laboratory and clinical monitoring, the proposed dose is expected to achieve a balance between clinically meaningful exposure in target cells while minimising the risk of ADRs. Some other studies have treated for longer than 14 days (7) but the natural history of COVID19 is that in most cases the course of disease for that individual has been established within 2 weeks.

It is acknowledged that the total dose of colchicine administered in this study (7 mg) is modestly greater than that recommended for treatment of acute gout (6 mg) but the proposed regimen has been designed to minimise the risk of ADRs.

Note: The British National Formulary advises a maximum total of 6mg per treatment course for acute gout (1mg less than the total for this study). However, the treatment course for gout is given over up to three days at 500mcg 2-4 times per day initially. In the PRINCIPLE Trial, the treatment will be spread over two weeks at a lower daily dose.

We propose a shorter duration and no loading dose compared to the COLCORONA study (62), given the incidence of side-effects found in that study, and that by two weeks, most patients with COVID-19 have either recovered or been hospitalised. Therefore, the window of opportunity for a positive benefit is mainly over two weeks, and a shorter duration without a loading dose will minimize risk of side-effects, while offering potential benefit. There are no dose-findings studies for colchicine in COVID-19. Our proposed dosing regime is based on expert pharmacological opinion and an appraisal of side-effects balanced against potential benefit in a large-scale community study without face-to-face recruitment and monitoring.

**a. Investigational Medicinal Product (IMP) description**

Colchicine 500 microgram (µg) tablets. The tablets are for oral administration. One tablet to be taken daily by mouth for 14 days (14 tablets in total).

Special instructions: Tablets should be swallowed whole with a glass of water.

**Manufacturer:**
The Marketing Authorisation holder is:
Accord-UK Ltd
(Trading style: Accord)
Whiddon Valley
Barnstaple
Devon
EX32 8NS
Marketing authorisation number is: PL 0142/0918

**Labelling and QP release:**
Vertical Pharma Resources Ltd (trading as IPS Pharma), 41 Central Avenue, West Molesey, KT8 2QZ, UK
Authorisation number: WDA(H) 32879

© Copyright: The University of Oxford and Oxford University Hospitals NHS Foundation Trust 2019

**b. Storage of IMP**

Colchicine: This medicine does not need any special storage conditions, but we will ask participants to store the medication at room temperature. The medication will be stored in locked cupboards in restricted access rooms in the Nuffield Department of Primary Care Health Sciences; in locked cupboards in restricted access rooms in GP Practices; in Pharmacies.

**c. SmPC Precautions, concomitant medications, pregnancy and lactation**

**i. SmPC Precautions**

Colchicine is a commonly prescribed drug in UK primary care and has a well-described safety profile due to its regulatory assessments for the authorisation in gout. Typical treatment doses for acute gout are 500 micrograms 2–4 times a day until symptoms relieved, maximum 6 mg per course.

Colchicine is teratogenic in animal studies and contraindicated in patients with severe renal (including patients undergoing haemodialysis) or severe hepatic impairments. Colchicine may cause severe bone marrow depression (agranulocytosis, aplastic anaemia, thrombocytopenia) and blood cell dyscrasia.

Colchicine is potentially toxic with a narrow therapeutic window.
Symptoms of acute overdosage may be delayed (3 hours on average): nausea, vomiting, abdominal pain, haemorrhagic gastroenteritis, volume depletion, electrolyte abnormalities, leucocytosis, hypotension in severe cases. The second phase with life threatening complications develops 24 to 72 hours after drug administration: multisystem organ dysfunction, acute renal failure, confusion, coma, ascending peripheral motor and sensory neuropathy, myocardial depression, pancytopenia, dysrhythmias, respiratory failure, consumption coagulopathy.

Patients at particular risk of toxicity are those with renal or hepatic impairment, gastrointestinal or cardiac disease, and patients at extremes of age. However, colchicine has a good safety profile when used according to the established therapeutic guidelines, and toxicity is rare if the recommended doses are not exceeded.

In PRINCIPLE, we exclude patients with known severe renal and known liver impairment, and have used a cautious dosing regimen, with no loading dose and low daily dose, to minimise risk in participants with other less severe co-morbidities. Our dosing schedule is also shorter in duration than the 30 days used in the large scale, remotely managed COLCORONA trial.(62) In addition, we will mitigate the risk of toxicity by asking each participant taking colchicine the number of tablets remaining via their diary entry to ensure drug accountability.

**ii. Concomitant medications**

Colchicine is a substrate for both CYP3A4 and the transport protein P-gp. In the presence of CYP3A4 or P-gp inhibitors, the concentrations of colchicine in the blood increase. Toxicity, including fatal cases, have been reported during concurrent use of CYP3A4 or P-gp inhibitors such

© Copyright: The University of Oxford and Oxford University Hospitals NHS Foundation Trust 2019

as macrolides (clarithromycin and erythromycin), ciclosporin, ketoconazole, itraconazole, voriconazole, HIV protease inhibitors, calcium channel blockers (**verapami**l and **diltiazem**) and disulfiram.
Colchicine is contraindicated in patients with renal or hepatic impairment who are taking a P-gp inhibitor (e.g. ciclosporin, verapamil or quinidine) or a strong CYP3A4 inhibitor (e.g. ritonavir, atazanavir, indinavir, clarithromycin, telithromycin, itraconazole or ketaconazole).
Therefore, patients using the above medications will not be eligible for enrolment into the colchicine arm of PRINCIPLE.

**iii. Fertility, pregnancy and lactation**

Pregnancy and breast-feeding are exclusions for the colchicine arm.

Women of childbearing potential (premenopausal female that is anatomically and physiologically capable of becoming pregnant) and not prepared to use highly effective contraception for the 28 day duration of follow up in the study are excluded from the trial.

Pregnancy in the study will be reported as an AE. Follow-up information regarding the outcome of the pregnancy and any postnatal sequelae in the infant will be required.

#### 5. Safety reporting

Mechanisms for safety reporting are outlined in the protocol. In brief, we will collect symptoms and side effects from symptom diaries and participant telephone calls.

<u>Common and/or potential side effects that may be associated with colchicine include:</u>

Abdominal pain, diarrhoea*, nausea, vomiting.

A meta-analysis of 35 randomised trials of colchicine versus placebo found that the most common and significant adverse effect was diarrhoea. The only other adverse effect that occurred at a greater frequency than placebo was a set of pooled gastrointestinal symptoms including nausea, vomiting, diarrhoea, abdominal pain, loss of appetite, and bloating.(69, 70)
<u>Rare side effects that may be associated with colchicine:</u>

Agranulocytosis; alopecia; bone marrow disorders; gastrointestinal haemorrhage; kidney injury; liver injury; menstrual cycle irregularities; myopathy; nerve disorders; rash; sperm abnormalities; thrombocytopenia

** Side effects that may be associated with COVID-19*

Events rated as a 'major problem' (except loss of smell and/or taste) will be assessed by a clinician for potential reporting as an SAE, for the 14 days of IMP treatmentWe will report SAEs as defined in the main protocol for hospitalisation and/or death.

Pregnancy in the study will be reported as an AE (see pregnancy and breast-feeding section).

© Copyright: The University of Oxford and Oxford University Hospitals NHS Foundation Trust 2019

**Drug Accountability**

We will telephone all participants on Day 3 after randomisation to confirm that they have received their medication. For those receiving a trial treatment, 3 attempts are made to contact the participant to confirm receipt of the medication.

If we are unable to contact patients in the colchicine group, we will confirm and log IMP receipt by checking the patient's daily diary, where they are asked on a daily basis whether they have taken their trial treatment and how many tablets they have left. We can also check via the DHL portal, whether the participant pack containing the medication has been received by the participant, for additional confirmation. IMP receipt will be logged on the central IMP log.

© Copyright: The University of Oxford and Oxford University Hospitals NHS Foundation Trust 2019

### 24 APPENDIX I: USUAL CARE PLUS FAVIPIRAVIR

#### 1. Background and rationale

##### a. Evidence for potential benefits of favipiravir in COVID-19 illness

Small clinical trials assessing favipiravir for treatment of COVID-19 have been published, but results are inconclusive.

A randomised controlled trial in China of 240 adults hospitalised with moderate to severe COVID-19 compared favipiravir (1600mg BD day 1; 600mg BD days 2-7) with umifenovir (200mg TDS).(71) In the per-protocol analysis, there was no difference in the primary outcome of clinical recovery by day 7 between the two arms. In a post-hoc analysis restricted to patients with moderate COVID-19, 70/98 (71.43%) had recovered by day 7 in the favipiravir arm, vs 62/111 (55.86%) in the umifenovir arm (p = 0.0199). In other post-hoc analyses, time to cessation of fever (p < 0.0001) and cough (p < 0.0001) was shorter in the favipiravir arm. Adverse events were similar between the two arms, apart from hyperuricaemia which occurred in 13.8% of participants who received favipiravir vs 2.5% among those receiving umifenovir (p < 0.001).(71)

A trial in India of 150 patients with mild to moderate COVID-19 compared favipiravir (1800 mg BD day 1; 800 mg BD days 2-14) with usual care.(72) The primary outcome of time to viral clearance was not significantly different between the two arms (favipiravir 5 days [95% CI 4-7] vs usual care 7 days [95% CI 5-8, P=0.129), but there was a significant decrease in the secondary outcome of time to clinical cure in the favipiravir arm (favipiravir 3 days [95% CI 3- 4] vs usual care 5 days [95% CI 4-6], P=0.030). Adverse events were more common in the favipiravir arm (36% vs 8%); these were mainly hyperuricaemia (16.4%) and abnormal liver function tests (6.8%); 76.9% were mild and 23.1% were moderate.

A trial in Russia among 60 adults hospitalised with moderate COVID-19 compared usual care (n=20) vs lower dose favipiravir (1600 mg BD day 1; 600 mg BID days 2–14) (n=20) vs higher dose favipiravir (1800 mg BD day 1; 800 mg BD days 2–14) (n=20).(73) Viral clearance was higher in the favipiravir arms vs usual care by day 5 (25/40 [62.5%] vs 6/20 [30.0%], p = 0.018), although this difference was non-significant by day 10 (37/40 [92.5%] vs 16/20 [80.0%], p = 0.155). Temperature normalisation was quicker in the favipiravir arms (2 days [IQR 1–3] vs 4 days [IQR 1–8], p = .007). 17.5% of patients in the favipiravir arm experienced adverse drug reactions including gastrointestinal disturbances and raised liver function tests. Two patients discontinued the drug early.

A trial in Oman randomised 89 adults hospitalised with moderate to severe COVID-19 to receive favipiravir (1600 mg OD day 1; 600 mg BD days 2-10) combined with inhaled interferon beta-1b, versus hydroxychloroquine. There was no difference between the two groups with regard to the primary outcomes of inflammatory markers at discharge, length of hospital stay, transfer to ICU, or mortality. Adverse events were not reported.(74)

A smaller, non-randomised study among adults hospitalised with COVID-19 in China compared 35 patients treated with favipiravir (1600 mg BD day 1; 600 mg BD days 2-14) with 45 historical controls who were treated with lopinavir/ritonavir (400mg/100mg BD). All patients also

© Copyright: The University of Oxford and Oxford University Hospitals NHS Foundation Trust 2019

received aerosolised IFN-α. Viral clearance was quicker in the favipiravir cohort (median 4 days, IQR 2.5-9) compared to the lopinavir/ritonavir cohort (11 days, IQR 8-13, p <0.001). Improvements in CT chest imaging at 14 days were higher in the favipiravir group (91.4% vs 62.2%, p = 0.004).(75)

These mixed findings highlight the need for a large randomised controlled trial of favipiravir to treat COVID-19 in the community.

##### b. Potential mechanism of efficacy

Favipiravir is an oral antiviral that is licensed in Japan for use against novel and re-emerging influenzae, and has been used in clinical trials for Ebola (76).(76) Like remdesivir, it is a nucleoside analogue which selectively inhibits viral RNA polymerase, and has been shown to have in vitro activity against a range of RNA viruses (77, 78), including SARS-CoV-2.(9) Favipiravir was one of seven antiviral agents reported to achieve plasma concentrations at least double the reported concentrations required to inhibit 90% of SARS-CoV-2 replication in vitro.(79) In animal models, high dose favipiravir was found to reduce viral titres and lung pathology in SARS-CoV-2 infected hamsters.(80, 81) Given that upper respiratory tract SARS-CoV-2 viral loads peak in the first 3-5 days of illness,(82) antiviral treatments for COVID-19 may be of particular value early in the course of the disease.

#### 2. Changes to outcome measures

The addition of the *usual care plus favipiravir arm* will require the introduction of the secondary outcome measure, safety.

#### 3. Eligibility criteria specifically related to favipiravir

Inclusion criteria: No changes

Exclusion criteria:

- Aged <50 years
- Known or suspected pregnancy
- Breastfeeding
- Women of childbearing potential (premenopausal female that is anatomically and physiologically capable of becoming pregnant*), or male with a partner of childbearing potential, not willing to use highly effective contraceptive** for 28 day duration of the trial.
- Known allergy to favipiravir
- Currently taking favipiravir
- Known history of gout
- Known severe liver disease

* As recorded by the participant on the screening form and confirmed on Day 1 by a call between clinician and participant

© Copyright: The University of Oxford and Oxford University Hospitals NHS Foundation Trust 2019

** Highly effective methods have typical-use failure rates of less than 1% and include male or female sterilisation and long-acting reversible contraceptive (LARC) methods (intrauterine devices and implants) OR If a couple are using another method of contraception, such as a combined hormonal method, progestogen only pill or injection, they are only eligible if they are willing to use an additional barrier method (e.g. male condom) for the 28 day duration of follow-up in the trial.

Note: a barrier method on its own is not sufficient.

The Favipiravir Patient Information Sheet Appendix, which the participant must read prior to providing informed consent, will clearly state the exclusion criteria listed above and the participant will be asked if they meet any of these exclusion criteria at the screening stage of the trial including whether they have any known history of gout or known severe liver disease. The assessing clinician will then review the participant's responses against their medical record to confirm eligibility.

#### 4. Detail of intervention

Participants randomised to the usual care plus favipiravir arm will receive usual clinical care as per NHS guidelines, plus one batch of favipiravir for five days. We will use the IMP distribution methods described in the protocol to deliver IMP to participants.

##### a. Investigational Medicinal Product (IMP) description

Favipiravir 200 milligram (mg) tablets. The tablets are for oral administration. Nine tablets (1800mg) favipiravir to be taken twice a day on day one, and then four tablets (800mg) twice daily for four days (50 tablets in total).

This product is not licensed for use in the UK.

**Manufacturer:**
It is manufactured by FujifilmToyama Chemical Company Ltd., TOYAMA CHEMICAL CO., LTD. 2-5, Nishishinjuku 3-chome, Shinjuku-ku, Tokyo 160-0023, Japan.
Manufacturirng Licence No. NHI Number 87625
MA Approval No from outside the EEA: 22600AMX0053000 Japan

**Importer, Labelling and QP release:**
IPS Pharma, 41 Central Ave, East Molesey, West Molesey KT8 2QZ (32879)

##### b. Storage of IMP

All study medication is to be kept in a dry area, stored at 1° to 30°C (59° to 86°F) and shielded from direct light in locked cupboards in restricted access rooms in the Nuffield Department of Primary Care Health Sciences; in locked cupboards in restricted access rooms in GP practices; in Pharmacies. The IMP is stable at 1-30°C.

##### c. Precautions, concomitant medications, pregnancy and lactation

© Copyright: The University of Oxford and Oxford University Hospitals NHS Foundation Trust 2019

**i. Precautions**

Favipiravir has been used in over 30 clinical trials and has a favourable safety profile.(76) A review of six phase 2 and 3 controlled trials, including 4299 participants and 175 person-years-of-follow-up, found no statistically significant differences in overall proportion of AEs, SAEs, discontinuations due to AEs or LFT elevations between favipiravir and placebo or other treatment arms.(76) However, there was evidence of mild to moderate, asymptomatic, uric acid elevations among patients receiving favipiravir, although these generally returned to normal by 21 days. Overall, follow up times were short (5-21 days) and participants were generally young.

Patients with gout or a history of gout, and patients with hyperuricaemia (blood uric acid level may increase, and symptoms may be aggravated) are excluded.

The participant must avoid excessive exposure to sunlight or artificial ultraviolet light.

**ii. Concomitant medications**

Restrictions to paracetamol use (limiting daily use in adults to no more than 3000 mg/day) have been incorporated into all clinical study protocols. Participants will be advised they can't consume more than 6 paracetamol tablets in 24 hours.

**iii. Pregnancy and lactation**

Evidence from animal models suggests that favipiravir has teratogenic potential, and there are no human studies of its use among pregnant or lactating women. The Japanese drug safety bureau advise that women of child-bearing potential should use effective contraception for up to 7 days after the end of treatment (https://www.cdc.gov.tw/File/Get/ht8jUiB_MI-aKnlwstwzvw). Male patients should use the most effective form of contraception (see eligibility criteria for examples) for up to seven days after the end of treatment if they have a female partner of child-bearing potential. Men should also avoid intercourse with pregnant women.

As the effect of favipiravir on pregnancy is unknown, pregnancy in the study will be reported as an AE. Follow-up information regarding the outcome of the pregnancy and any postnatal sequelae in the infant will be required.

#### 5. Safety reporting

##### a. Side-effects

Mechanisms for safety reporting are outlined in the protocol. In brief, we will collect symptoms and side effects from symptom diaries and participant telephone calls.

Common and/or potential side effects from IMP include:

- Diarrhoea*[1]
- Nausea[1]
- Headache*[1]

© Copyright: The University of Oxford and Oxford University Hospitals NHS Foundation Trust 2019

- Urinary Tract Infections[1]
- Vomiting[1]
- Raised liver enzymes
- Elevated uric acid concentrations

*side-effects also seen with COVID-19

These are all reversible upon ceasing IMP.

Certain pre-defined moderate (defined above[1]) AEs experienced in the 5 days of favipiravir drug administration, will be reviewed daily and will be assessed by a clinician until resolution.

A systematic review suggests that liver changes do not differ between favipiravir and placebo/other treatments (76). To mitigate the risk of elevated uric acid concentrations, we exclude people with gout. Evidence shows mild to moderate, asymptomatic, uric acid elevations, which return to normal after stopping the medication (76).

As the effect of favipiravir on pregnancy is unknown, pregnancy in the study will be reported as an AE (see pregnancy and breast-feeding section).

We will report SAEs as defined in the main protocol for hospitalisation and/or death.

##### b. Reference Safety Information (RSI)

See section 7.13 of the Investigator Brochure. No serious adverse reactions are considered expected for the purpose of expedited reporting of suspected unexpected serious adverse reactions (SUSAR).

##### c. Drug Accountability

We telephone all participants on Day 3 after randomisation to confirm that they have received their medication and read the instructions on the medication card. For those receiving a trial treatment, 3 attempts are made to contact the participant to confirm receipt of the medication.

If we are unable to contact patients in the favipiravir group, we will confirm and log IMP receipt by checking the patient's daily diary, where they are asked on a daily basis whether they have taken their trial treatment, the number of tablets taken and the number of tablets remaining. We can also check via the courier portal, whether the participant pack containing the medication has been received by the participant, for additional confirmation. IMP receipt will be logged on the central IMP log.

If a participant decides that they no longer wish to take their medication, we will provide a pre-paid envelope so that they can return the medication to the trial team, via courier.

##### d. Risk/Benefit Assessment

The UK COVID-19 Therapeutics Advisory Panel recommends including favipiravir into the PRINCIPLE platform with an 1800mg loading dose, followed by an 800mg BD maintenance dose, based on a review of efficacy and safety data.

###### i. Risks

© Copyright: The University of Oxford and Oxford University Hospitals NHS Foundation Trust 2019

Phase I studies and pharmacokinetic studies of favipiravir have indicated increased blood levels of uric acid and elevation of liver aminotransferases in some individuals (76). In addition, animal studies have indicated potential teratogenicity and the drug is distributed in sperm (83). Taking this evidence into account and to ensure patient safety we will exclude known pregnancy, breastfeeding, severe liver disease and known history of gout, and require participants to use adequate contraception for the duration of the treatment and 28 days of follow-up.

**ii. Benefits**

The benefits of favipiravir have been shown in several phase I-III studies to significantly alleviate influenza symptoms with a good safety profile (83). From the studies detailed below, the frequency of adverse events was 386/1472 (26.2%) with favipiravir versus 227/894 (25.4%) with placebo and there were no differences in the incidence of any specific adverse event between groups.

A Phase I/II study in type A or B influenza using favipiravir doses 1800mg bd for 1 day followed by 800mg bd for 4 days (101 patients) or 2400mg/600mg/600mg for 1 day followed by 600mg tds for 4 days (82 patients) or placebo (88 patients).

Two Phase III studies in type A or B influenza using favipiravir doses 1800mg bd for 1 day followed by 800mg bd for 4 days (301 and 526 patients respectively) or placebo (322 and 169 patients respectively).

A global phase III study in type A or B influenza using favipiravir 1200mg/400mg for 1 day followed by 400mg bd for 4 days or oseltamivir 75mg bd for 5 days (377 and 380 patients respectively).

A phase II study in type A or B influenza using favipiravir doses 1000mg bd for 1 day followed by 400mg bd for 4 days (88 patients) or 1200mg bd for 1 day followed by 800mg bd for 4 days (121 patients) or placebo (124 patients).

Favipiravir was also used for Ebola virus disease, especially in the JIKI trial in Guinea (84). Doses (2400mg/2400mg/1200mg on Day 1 followed by 1200mg bd) were significantly higher than proposed for the current study. No drug related grade 3 or 4 clinical events were observed. In 41 of the 48 patients who survived, biochemical abnormalities of renal and liver function rapidly improved on treatment; 7 of 48 patients saw transient rises in one marker but all subsequently normalised despite continuing favipiravir. Biochemical abnormalities in patients who died were attributed to severe viral infection.

In London institutions such as Royal Free Hospital and Great Ormond Street Hospital (GOSH), favipiravir has been used as post-exposure prophylaxis for Ebola virus, for chronic norovirus infection, refractory influenza infection, astrovirus, respiratory syncytial virus and seasonal coronavirus. Occasional asymptomatic elevation of liver aminotransferase levels were observed, but no serious adverse events related to the drug. This includes in patients with immunodeficiencies who have received several months-worth of treatment (FLARE Trial).

Favipiravir has been used clinically in several viral infections and has shown beneficial treatment effects in both influenza and COVID-19 studies, as well as having a good safety profile.

© Copyright: The University of Oxford and Oxford University Hospitals NHS Foundation Trust 2019

#### 25. APPENDIX J: USUAL CARE PLUS IVERMECTIN

##### 1. Background and rationale

Ivermectin is an anti-parasitic drug that is licensed in the United Kingdom (UK) for topical use in the treatment of rosacea [70]. Oral ivermectin is approved by the US FDA and is on the World Health Organisation Essential Medicines List [71]. It is widely used globally to treat strongyloidiasis, onchocerciasis, lymphatic filariasis and hyperkeratotic scabies, with over 2.7 billion treatments given as part of mass drug administration programmes for these conditions.[72] Although it remains unlicensed for these indications in the UK, [70] there have been calls for this to change, in particular for treatment of scabies [73].

Ivermectin is well tolerated in humans and has a good safety profile. A single dose of 200µg/kg is used to treat onchocerchiasis, and doses of 300-400 µg/kg are recommended for ***Wuchereria bancrofti*** (which causes lymphatic filiarisis) when administered annually in mass drug administration programmes [74]. Higher doses of 800 µg/kg have been assessed as safe treatments for onchocerciasis. [75-80] while a recent dose ranging study of ivermectin 300 µg/kg dose for three days, as part of combination treatment for malaria, was assessed as safe [81] and is being taken forward in larger trials.[82]

##### a. Evidence for potential benefits of ivermectin in COVID-19 illness

Ivermectin has in vitro activity against SARS-CoV-2, and emerging clinical evidence from multiple small trials suggest it may effectively reduce SARS-CoV-2 viral loads, reduce time to recovery, and reduce mortality among people with COVID-19. However, data from large, robust clinical trials is lacking.

*In vitro studies*

*In vitro* studies suggest that ivermectin has anti-viral activity against Dengue, HIV-1 and Influenza A [86, 87]. More latterly, Caly and colleagues found that ivermectin inhibited SARS-CoV-2 replication *in vitro* [83]. There was a 93% reduction in viral RNA production when cells infected with SARS-CoV-2 at a multiplicity of infection of 0.1 were treated with 5 µM ivermectin, compared with untreated cells [83]. The ivermectin $IC_{50}$ was estimated at 2.5µM, (equivalent to 2190ng/ml), which is significantly higher than plasma concentrations ($C_{max}$ approx 40ng/ml) achieved with doses of 150-200µg/kg,[88]. However, in animal studies, ivermectin concentrations in the lung have been up to three times higher than in plasma, [89, 90] and doses of up to 2000 µg/kg have been well tolerated.[75] While concentrations that are inhibitory against SARS-CoV02 *in vitro* may be difficult to attain *in vivo*, the relationship between *in vitro* inhibitory effects and clinical efficacy is not clear, and emerging *in vivo* evidence is promising.

*In vivo studies*

A number of RCTs have assessed the use of oral ivermectin as a treatment for COVID-19, with some evidence of benefit. The majority of these trials were conducted in hospitals and had small numbers of participants [91-95].

Ahmed and colleagues report the findings of a pilot randomised, double blind trial in which 72 hospitalized patients with mild COVID-19 within 7 days of hospital admission were treated with oral ivermectin alone (12 mg once a day for five days), ivermectin (12mg on day one only) and doxycycline, or placebo [96]. The primary outcome of mean time to SARS-CoV-2 clearance on PCR was quickest in the five day ivermectin alone arm (9.7 days, 95% CI 7.8–11.8, p = 0.02), 11.5 days

© Copyright: The University of Oxford and Oxford University Hospitals NHS Foundation Trust 2019

(95% CI 9.8–13.2, p = 0.27) in the one day ivermectin plus doxycycline arm, and 12.7 days (95% CI 11.3–14.2) in the placebo control arm, suggesting a potential dose response effect for ivermectin. There were no severe adverse drug events in the pilot study.

Chaccour and colleagues report the findings of a pilot randomised double-blind trial of 24 patients presenting to an emergency department within COVID-19 within 72 hours of symptom onset, in a pre-print [97]. Patients were randomly assigned to receive a single dose of ivermectin (400 mcg/kg) or placebo. There was no significant difference in the primary endpoint of proportion of PCR positive patients at day seven (p = 1.0), however, patients receiving ivermectin reported fewer patient-days of symptoms compared with placebo controls (171 compared with 255 patient-days, respectively. No p value reported). Fifteen adverse events were reported within the 28 day follow-up period – seven in the ivermectin group (including dizziness and blurred vision) and eight in the placebo group. There were no serious adverse events.

A pilot randomised controlled trial of 45 adults with COVID-19 in Argentina compared high dose ivermectin (600 μg/kg) for five days versus standard care controls (no ivermectin). [98] In a pre-print, the investigators report no difference in viral load reduction at 5 days between the two groups, but in a stratified analysis, those with median ivermectin concentrations > 160ng/ml had a greater reduction in 5 day viral load (72% IQR 59 – 77) versus untreated controls (42% IQR 31 – 73) (p=0·004).[98] There were 5 adverse events in the standard care arm compared to 17 in the high dose ivermectin arm (most common was rash reported in 3 participants, all mild). One serious adverse event in the ivermectin arm was reported; hyponatraemia that was thought to be related to COVID-19. There was no association between higher ivermectin plasma concentrations and adverse events.

A rolling meta-analysis by Andrew Hill and colleagues at the University of Liverpool has identified 18 ongoing or completed randomised clinical trials (n=2,167) with another 37 clinical trials registered (total 7,491 patients).[99] Included trials were mostly conducted in low- and middle-income countries and found benefit in terms of viral clearance, reduction in inflammatory markers, time to recovery, hospitalisation and mortality in patients with COVID-19. In a meta-analysis (n = 1143, 603 for ivermectin, 540 for controls), the risk of all-cause mortality was significantly lower in patients receiving ivermectin (risk ratio 0.25, 95% CI 0.11-0.58). Furthermore, there was a suggestion of a dose response effect, with findings consistently favouring a longer treatment duration (of up to 5 days) over a single dose regimen, and a doses of ≥400 μg/kg body weight compared to 200 μg body weight (personal communication and reference [99]).

A meta-analysis of safety data from anthelmintic clinical trials showed no difference in the severity of the adverse events between standard (up to 400 μg/kg) and higher doses of ivermectin regardless of treatment duration [80].

Ivermectin is licensed for use at a single daily dose of 400 μg/kg in France and the Netherlands. The daily dose in this study is lower (300 μg/kg), but will be for over three days. This dosing regimen is not currently licensed anywhere, but is being used in studies of treatment for malaria. Higher doses of 400 μg/kg for 5 days have been used in studies of COVID-19 with no adverse safety signals.

About 3.7 billion doses of ivermectin have been given in mass drug administration programs over 30 years. At 10-100 times higher than current doses, ivermectin has been found to be teratogenic in animals. A review of 893 pregnant women inadvertently given ivermectin found no neonatal

© Copyright: The University of Oxford and Oxford University Hospitals NHS Foundation Trust 2019

deaths, maternal morbidity, pre-term births, or low birthweight. There was low certainty of evidence about impact on congenital anomalies or spontaneous abortions and stillbirths.[100]

##### *b.* Potential mechanism of action

Caly *et al* propose that ivermectin may inhibit binding of viral proteins to Importin alpha/Importin beta 1, preventing the transport of these viral proteins from the host cell cytoplasm to the host cell nucleus [83]. When in the cell nucleus, these viral proteins normally suppress the host cell's anti-viral response. Treatment with ivermectin may therefore enhance the ability of host cells to respond to SARS-CoV-2 infection [83]. In a molecular modelling study, Sen Gupta *et al* found that Ivermectin appears to bind to Importin alpha, Helicase, and RNA dependent RNA polymerase (RdRp). The ivermectin-RdRp complex appeared to have the highest stability, suggesting that ivermectin may inhibit SARS-CoV-2 activity through RdRp inhibition.[84] In vitro and animal studies have demonstrated that ivermectin significantly reduces the production of pro-inflammatory cytokines such as TNF-alpha, Interleukin – 1beta and Interleukin 6 [85]. It is therefore possible that ivermectin could reduce the cytokine storm and ensuing cellular damage that occurs in some individuals with COVID-19.

#### 2. Changes to outcome measures

The 'safety' secondary outcome measure, will apply to ivermectin.

#### 3. Eligibility criteria specifically related to Ivermectin

Inclusion criteria: No changes

Exclusion criteria:

- A known allergy to Ivermectin or any of its excipients
- Known or suspected pregnancy
- Breastfeeding
- Women of childbearing potential (premenopausal female that is anatomically and physiologically capable of becoming pregnant*) and not prepared to use highly effective contraception for the 28 day duration of follow up in the study**
- Ever having travelled to countries that are endemic for *Loa loa* (Angola, Cameroon, Central African Republic, Chad, Democratic Republic of Congo, Ethiopia, Equatorial, Guinea, Gabon, Republic of Congo, Nigeria and Sudan)
- Known bleeding disorder
- Known severe liver disease
- Currently taking the following drugs:
    - quinidine, amiodarone, diltiazem, spironolactone, verapamil, clarithromycin, erythromycin, itraconazole, ketoconazole, cyclosporine, tacrolimus, sirolimus, indinavir, ritonavir, cobicistat, warfarin
- Consumption of grapefruit juice

* As recorded by the participant on the screening form and confirmed on Day 3 telephone call

**Highly effective methods have typical-use failure rates of less than 1% and include male or female sterilisation and long-acting reversible contraceptive (LARC) methods (intrauterine

© Copyright: The University of Oxford and Oxford University Hospitals NHS Foundation Trust 2019

devices and implants). Women using another method of contraception, such as a combined hormonal method, progestogen only pill or injection, are eligible **if** they are willing to use an additional barrier method (e.g. male condom) for the 28 day duration of follow-up in the trial.

Note: a barrier method on its own is not sufficient.

The Ivermectin Patient Information Sheet Appendix, which the participant must read prior to providing informed consent will clearly state the exclusion criteria listed above and the participant will be asked if they meet any of these criteria at the screening stage of the trial including whether they have any known bleeding disorders or known severe liver disease. The assessing clinician will then review the participant's responses against their medical record to confirm eligibility.

#### 4. Detail of intervention

Participants randomised to the usual care plus ivermectin arm will receive usual clinical care as per NHS guidelines, plus Ivermectin to be taken each day for 3 days. We will use the IMP distribution methods described in the protocol to deliver IMP to participants.

##### a. Investigational Medicinal Product (IMP) description

Ivermectin tablets (3mg) are round, white (or off-white) tablets with a 5.5 mm diameter and a 2.1 mm thickness (IB). The tablets are for oral administration. Tablets to be taken as one dose each day for 3 days at 300 μg/kg body weight (weight range below) up to maximum of 30 mg tablets can be taken with water but no food should be taken two hours before or after administration, as the influence of food on absorption is not known.

**Table: Ivermectin dosing by weight to target 300-400 μg/kg**

| Participant weight | Ivermectin dose | Number of 3 mg ivermectin tablets per day |
|---|---|---|
| 45-64 kg | 18 mg | 6 |
| 65-84 kg | 24 mg | 8 |
| ≥84 kg | 30 mg | 10 |

This product is not licensed for use in the UK.

**Manufacturer:**
It is manufactured by
Edenbridge Pharmaceuticals, LLC
169 Lackawanna Avenue, Suite 110
Parsippany, NJ 07054
ANDA 204154.
The drug product is on the WHO Prequalification List (WHO Reference Number NT009)

**Importer, Labelling and QP release:**
Torbay Pharmaceuticals, Torbay and South Devon NHS Foundation Trust

© Copyright: The University of Oxford and Oxford University Hospitals NHS Foundation Trust 2019

Wilkins Drive | Paignton | TQ4 7FG
MIA(IMP) 13079

The IMP is available in aluminium blisters of 20 tablets (Edenbridge Pharmaceuticals http://www.edenbridgepharma.com/pharmacists-ivermectin.php).

Participants will be asked to enter their body weight at screening, and if an invalid weight is entered they will be deemed ineligible. The participant's weight will also be confirmed during the Day 1 follow-up call.

**Dosing Risks**

There are no concerns with the proposed trial dose for PRINCIPLE. Please see Tables 11 and 12 of the Investigator's Brochure for the safety data.

**b. Storage of IMP**

Ivermectin: This medicine does not need any special storage conditions, but we will ask participants to store the medication at room temperature. The medication will be stored in locked cupboards in restricted access rooms in the Nuffield Department of Primary Care Health Sciences; in locked cupboards in restricted access rooms in GP Practices; in Pharmacies.

**c. Precautions, concomitant medications, pregnancy and lactation**

**i. Precautions**

Ivermectin has a good safety profile, which is reflected in its widespread use in non-clinical settings as part of mass drug administration programmes [58][100], often provided in the community by lay health workers. Most side effects are related to parasitic treatment effects and are mild and transient. More serious parasite load related effects are highly unlikely in UK primary care settings. A non-significant increase in prothrombin time has been reported among patients taking ivermectin in one study, with no related increase in bleeding related adverse events. There have been rare post-marketing reports of increased International Normalized Ratio (INR) when warfarin is co-administered with ivermectin. There have been some reports of elevated liver enzymes with ivermectin use. As ivermectin is extensively metabolized by the liver, caution should be exercised if ivermectin is administered to patients with impaired hepatic function.

Rarely, severe and potentially fatal cases of encephalopathy have been described following administration of ivermectin for treatment of *Loa loa*, particularly in patients heavily infected with *Loa Loa*. People who have visited a *Loa loa* endemic country will be excluded from the ivermectin arm.

**ii. Concomitant medications**

Ivermectin is a substrate of the P-glycoprotein (P-gp), thus, its penetration into the CNS maybe increased by P-gp inhibitors. Common P-gp inhibitors include: Amiodarone, Ketoconazole/Itraconazole, Clarithromycin/Erythromycin, Ciclosporin, Verapamil, Diltiazem, Quinidine, Protease inhibitors (indinavir, ritonavir or those containing cobicistat), Sirolimus/Tacrolimus, grapefruit juice [101]. Those taking these drugs will be ineligible.

Ivermectin is primarily metabolised by CYP3A4, thus, plasma levels may be increased by potent CYP3A4 inhibitors including clarithromycin, erythromycin, diltiazem, itraconazole, ketoconazole, ritonavir, verapamil and grapefruit juice [85]. However, there is data supporting the safety of

© Copyright: The University of Oxford and Oxford University Hospitals NHS Foundation Trust 2019

ivermectin at higher doses than proposed in this study,[80] and these drugs already constitute exclusion criteria due to being P-gp inhibitors.

| **Class** | **Drug** | **Rationale** |
|---|---|---|
| Antiarrhythmic/ Antihypertensive | Quinidine | May increase ivermectin exposure by inhibiting its metabolism and excretion or competing with CYPs or the P-gp. |
| | Amiodarone | |
| | Diltiazem | |
| | Spironolactone | |
| | Verapamil | |
| Antibiotic- Macrolides | Clarithromycin | |
| | Erythromycin | |
| Antifungal agents | Itraconazole | |
| | Ketoconazole | |
| Immunosuppressants | Cyclosporine | |
| | Tacrolimus | |
| | Sirolimus | |
| Anti-HIV therapy | Indinavir | |
| | Ritonavir | |
| | Cobicistat | |
| Foods / vitamins | Grapefruit juice | |
| Anticoagulant | Warfarin | Theoretical risk of bleeding due to potential increase in prothrombin time |

**iii. Pregnancy and lactation**

Ivermectin does not appear to be selectively fetotoxic to the developing fetus, as it is only teratogenic in animal studies at doses high enough to cause toxicity in the mother (significantly greater than the dose proposed for PRINCIPLE). However, as there are no adequate and well-controlled studies in pregnant women, ivermectin should not be used during pregnancy. Pregnancy (known or suspected) and breast-feeding are exclusions for the ivermectin arm of the trial. During the day 1 call, the clinician will confirm this exclusion criteria with the participant.

As the effect of ivermectin on pregnancy is unknown, pregnancy in the study will be reported as an AE. Follow-up information regarding the outcome of the pregnancy and any postnatal sequelae in the infant will be required.

#### 5. Safety reporting

##### a. Side-effects

© Copyright: The University of Oxford and Oxford University Hospitals NHS Foundation Trust 2019

Mechanisms for safety reporting are outlined in the protocol. In brief, we will collect symptoms and side effects from symptom diaries and participant telephone calls.

Common and/or potential side effects from IMP include the following, as outlined in section 6.11 Clinical Trial Adverse Events of the IB:

- Visual disturbances[1]: such as blurred vision, difficulty reading, tunnel vision, floaters, black spots, abnormal colours or shapes.
- CNS: dizziness, tremor, somnolence, vertigo, difficulty focussing, confusion[1], seizures[1], headache*
- Gastrointestinal: diarrhoea*, nausea, vomiting, abdominal pain, lack of appetite*
- Skin: rashes, which are normally mild and resolve after stopping the medication
- General: fatigue*, general malaise*, muscle pain*

* Side effects that may be associated with COVID-19

Please see section 5.4 Reference Safety Information, for the full list of common and potential side-effects.

Rare side effects that may be associated with ivermectin:

- Stevens-johnson
- Toxic epidermal necrolysis
- Blood in urine, as there is a very rare risk of bleeding
- Prolonged activated partial thromboplastic time (aPTT)

Many observed side effects are likely related to the parasite load of infections that are rare in UK primary care. Most side effects are mild and short-lived.

Certain pre-defined moderate AEs (defined above[1]) experienced in the 14 days from initial drug administration, will be reviewed daily and will be assessed by a clinician until resolution.

As the effect of ivermectin on pregnancy is unknown, pregnancy in the study will be reported as an AE (see pregnancy and breast-feeding section).

We will report SAEs as defined in the main protocol for hospitalisation and/or death.

##### b. Reference Safety Information (RSI)

RSI for Ivermectin can be found in section 5.4 (Reference Safety Information) of the Investigator's Brochure.

##### c. Drug Accountability

We telephone all participants on Day 3 after randomisation to confirm that they have received their medication and read the instructions on the medication card. For those receiving a trial treatment, 3 attempts are made to contact the participant to confirm receipt of the medication.

If we are unable to contact patients in the ivermectin group, we will confirm and log IMP receipt by checking the patient's daily diary, where they are asked on a daily basis whether they have taken their trial treatment. We can also check via the courier portal, whether the participant pack

© Copyright: The University of Oxford and Oxford University Hospitals NHS Foundation Trust 2019

containing the medication has been received by the participant, for additional confirmation. IMP receipt will be logged on the central IMP log.

Participants will be asked to return any unused medication to the trial team in a pre-paid envelope, via courier. Returned medication will be counted to monitor adherence/real dose and documented on the central drug accountability log. Any discrepancy will be followed up by the clinical team until resolution.

© Copyright: The University of Oxford and Oxford University Hospitals NHS Foundation Trust 2019

### 26. Supplementary Material

#### A. Abbreviations

| | |
|---|---|
| AE | Adverse event |
| AR | Adverse reaction |
| CI | Chief Investigator |
| CRF | Case Report Form |
| CT | Clinical Trials |
| CTA | Clinical Trials Authorisation |
| CTRG | Clinical Trials and Research Governance |
| DMSC | Data Monitoring Committee / Data Monitoring and Safety Committee |
| DSUR | Development Safety Update Report |
| GCP | Good Clinical Practice |
| GP | General Practitioner |
| HRA | Health Research Authority |
| HCP | Healthcare professional |
| IB | Investigators Brochure |
| ICF | Informed Consent Form |
| ICH | International Conference on Harmonisation |
| IMP | Investigational Medicinal Product |
| ISA | Intervention Specific Appendix |
| MHRA | Medicines and Healthcare products Regulatory Agency |
| NHS | National Health Service |
| NIHR | National Institute of Health Research |
| RES | Research Ethics Service |
| PHE | Public Health England |
| PI | Principal Investigator |
| PIL | Participant/ Patient Information Leaflet |
| R&D | NHS Trust Research and Development Department |
| RCGP RSC | Royal College of General Practitioners Research Surveillance Centre |
| REC | Research Ethics Committee |
| RSI | Reference Safety Information |
| SAE | Serious Adverse Event |
| SAR | Serious Adverse Reaction |
| SDV | Source Data Verification |
| SMPC | Summary of Medicinal Product Characteristics |
| SOP | Standard Operating Procedure |
| TSC | Trial Steering Committee |
| SUSAR | Suspected Unexpected Serious Adverse Reactions |
| TMF | Trial Master File |

© Copyright: The University of Oxford and Oxford University Hospitals NHS Foundation Trust 2019

#### B. Key Trial Contacts

| | |
|---|---|
| **Chief Investigator** | Professor Chris Butler<br>Nuffield Department of Primary Care Health Sciences<br>Gibson Building<br>Radcliffe Observatory Quarter<br>Woodstock Road<br>Oxford<br>OX2 6GG<br> |
| **Sponsor** | Joint Research Office<br>1st floor, Boundary Brook House<br>Churchill Drive,<br>Headington<br>Oxford OX3 7GB<br><br>Tel: +44 (0)1865572224<br>Fax: +44 (0)1865572228 |
| **Funder(s)** | UKRI/NIHR |
| **Clinical Trials Unit** | Primary Care Clinical Trials Unit,<br>Nuffield Department of Primary Care Health Sciences<br>Radcliffe Observatory Quarter<br>Woodstock Road<br>Oxford<br>OX2 6GG<br><br>01865 289296 |
| **Statistician** | Dr Ben Saville,<br>Berry Consultants,<br>Austin, Texas, USA,<br>And<br>Department of Biostatistics,<br>Vanderbilt University School of Medicine,<br>Nashville, Tennessee,<br>USA.<br><br>Dr Ly-Mee Yu<br>Primary Care Clinical Trials Unit,<br>Nuffield Department of Primary Care Health Sciences<br>Radcliffe Observatory Quarter<br>Woodstock Road<br>Oxford<br>OX2 6GG |
| **Committees** | **DMSC Chair:**<br>Prof. Deborah Ashby<br>Chair in Medical Statistics and Clinical Trials<br>Director of the School of Public Health<br>Imperial College London Faculty of Medicine, School of Public Health,<br>153 Medical School<br>St Mary's Campus<br>Imperial College London<br> |

© Copyright: The University of Oxford and Oxford University Hospitals NHS Foundation Trust 2019

| | |
|---|---|
| | (0)20 7594 8704<br><br>**DMSC Members:** Prof Simon Gates<br>Cancer Research Clinical Trials Unit (CRCTU)<br>Institute of Cancer and Genomic Sciences<br>University of Birmingham<br>Edgbaston<br>Birmingham<br>B15 2TT<br><br><br>Prof Gordon Taylor<br>College House,<br>University of Exeter, St Luke's Campus,<br>Heavitree Road,<br>Exeter, EX1 2LU, UK<br><br><br>Prof Nick Francis<br>Primary Care and Population Science,<br>University of Southampton, Southampton, UK.<br><br><br>Dr Patrick White<br>7th Floor<br>Capital House<br>Guy's<br>United Kingdom<br> |
| | **TSC Chair**<br>Prof Paul Little,<br>Primary Care and Population Science,<br>University of Southampton, Southampton, UK.<br><br><br>**TSC Members**<br>Prof Philip Hannaford,<br>NHS Professor of Primary Care<br>University of Aberdeen, Aberdeen, UK<br><br><br>Prof Matt Sydes,<br>Professor of Clinical Trials & Methodology,<br>MRC Clinical Trials Unit, University College of London, London, UK<br><br><br>*PPI representatives*<br>Ms Carol Green<br>Mr Tim Mustill |

© Copyright: The University of Oxford and Oxford University Hospitals NHS Foundation Trust 2019

#### C. Objectives and Outcome Measures

| | Objectives | Outcome Measures | Timepoint (s) |
|---|---|---|---|
| Primary | To assess the effectiveness of trial treatments in reducing<br>1) Time to recovery, for patients<br>2) Hospitalisation and/or death. | 1) Time to self-reported recovery, defined as the first instance that a participant reports feeling recovered from possible COVID-19, and<br>2) Hospitalisation and/or death | Within 28 days of randomisation<br>Patient report, Study Partner report, medical records, Daily online symptom scores |
| Secondary | To explore whether trial treatment will affect<br>1) Participant reported illness severity, reported by daily rating of how well participant feels.<br>2) Duration of severe symptoms and symptom recurrence<br>3) Contacts with the health services<br>4) Consumption of antibiotics<br>5) Hospital assessment without admission<br>6) Oxygen administration<br>7) Intensive Care Unit admission<br>8) Mechanical ventilation<br>9) Duration of hospital admission<br>10) Negative effects on well being<br>11) New infections in household<br>12) To determine if effects are specific to those with a | 1-3. Participant reports daily and monthly (after 28 days) symptoms.<br>4. Contacts with health services reported by patients and/or captured by reports of patients' medical records if the practice is a member of the RCGP RSC network<br>5. Bi-weekly reports from participants' primary care medical records<br>6-10. Patient report/carer report/medical record in primary and secondary care<br>11. WHO-5 Well Being Index<br>12. Reports of new infections in the household (from daily questionnaire)<br>13. Swab test results will indicate an "Intention to Treat Infected" group within the overall cohort for sub analysis. Blood test results on recovery (optional) for | Daily online symptom scores.<br>Telephone call or text on days 2, 7, 14 and 28 and once a month for 12 months if data is not obtained through the online diary.<br><br>GP notes review if available through Oxford RCGP RSC network; otherwise, other sources of routinely collected data after 28 days. Medical notes review for up to 10 years.<br><br>HES/ONS/EMIS/Medical record data linkage after 28 days if patients have been assessed in hospital<br><br>Swab result from medical records, the supporting laboratory and/or convalescent blood test result for evidence of historic COVID-19 |

© Copyright: The University of Oxford and Oxford University Hospitals NHS Foundation Trust 2019

| | | | |
|---|---|---|---|
| | positive test for SARS-CoV-2<br><br>13) To investigate the safety of treatments that are not licensed in the UK | evidence of historic COVID-19<br><br>Evaluation of overall safety of drugs by the monitoring of adverse events (AEs as defined in the ISAs) | WHO 5 Well Being Index at baseline, day 14, and day 28 and monthly for up to 12 months, either via online diary or telephone<br><br>For the duration of the treatment course and a defined period after the treatment finishes (see ISAs) |
| Qualitative sub-study | 1. To explore patients' experiences of consulting, being tested and taking (trial) medication for possible COVID-19.<br>2. To explore healthcare professionals' views of taking part in research during pandemics. | 1. Telephone interviews with patients.<br>2. Telephone interviews with healthcare professionals. | 1. After 28 days.<br>2. Once practice has completed recruitment. |
| Intervention(s) | All trial interventions are detailed in the Appendices. Further interventions may be added or replaced during the course of the trial, subject to suitable interventions becoming available and all necessary approvals being obtained. | | |
| Comparator | In the first instance, this will be a two-arm trial, with the intervention arm being usual care plus a trial drug and the comparator being usual care. There will be no placebo control in this study. Additional arms may be added as the trial progresses. These will be detailed in the Appendices. If an intervention arm is shown to be superior, then this will become the new standard of care. However, the primary analysis of subsequent interventions will correspond to the comparison versus the original Usual Care arm. | | |

##### D. Adverse Events

#### Definitions

© Copyright: The University of Oxford and Oxford University Hospitals NHS Foundation Trust 2019

| | |
|---|---|
| Adverse Event (AE) | Any untoward medical occurrence in a participant to whom a medicinal product has been administered, including occurrences which are not necessarily caused by or related to that product. |
| Adverse Reaction (AR) | An untoward and unintended response in a participant to an investigational medicinal product which is related to any dose administered to that participant.<br><br>The phrase "response to an investigational medicinal product" means that a causal relationship between a trial medication and an AE is at least a reasonable possibility, i.e. the relationship cannot be ruled out.<br><br>All cases judged by either the reporting medically qualified professional or the Sponsor as having a reasonable suspected causal relationship to the trial medication qualify as adverse reactions. |
| Serious Adverse Event (SAE) | A serious adverse event is any untoward medical occurrence that:<br>• results in death<br>• is life-threatening<br>• requires inpatient hospitalisation or prolongation of existing hospitalisation<br>• results in persistent or significant disability/incapacity<br>• consists of a congenital anomaly or birth defect*.<br><br>Other 'important medical events' may also be considered a serious adverse event when, based upon appropriate medical judgement, the event may jeopardise the participant and may require medical or surgical intervention to prevent one of the outcomes listed above.<br><br>NOTE: The term "life-threatening" in the definition of "serious" refers to an event in which the participant was at risk of death at the time of the event; it does not refer to an event which hypothetically might have caused death if it were more severe.<br><br>*NOTE: Pregnancy is not, in itself an SAE. In the event that a participant or his/her partner becomes pregnant whilst taking part in a clinical trial or during a stage where the foetus could have been exposed to the medicinal product (in the case of the active substance or one of its metabolites having a long half-life) the pregnancy should be followed up by the investigator until delivery for congenital abnormality or birth defect, at which point it would fall within the definition of "serious". |
| Serious Adverse Reaction (SAR) | An adverse event that is both serious and, in the opinion of the reporting Investigator, believed with reasonable probability to be due to one of the trial treatments, based on the information provided. |

© Copyright: The University of Oxford and Oxford University Hospitals NHS Foundation Trust 2019

| | |
|---|---|
| Suspected Unexpected Serious Adverse Reaction (SUSAR) | A serious adverse reaction, the nature and severity of which is not consistent with the Reference Safety Information for the medicinal product in question set out:<br>• in the case of a product with a marketing authorisation, in the approved summary of product characteristics (SmPC) for that product<br>• in the case of any other investigational medicinal product, in the approved investigator's brochure (IB) relating to the trial in question. |

NB: To avoid confusion or misunderstanding the difference between the terms "serious" and "severe", the following note of clarification is provided: "Severe" is often used to describe intensity of a specific event, which may be of relatively minor medical significance. "Seriousness"

#### E. Data Recording and Record Keeping

The data will be entered into the CRFs in an electronic format by the participant, Trial Partner or trial team (using OpenClinica™ database via Sentry). OpenClinica™ is stored on a secure server – data will be entered in a web browser and then transferred to the OpenClinica Database by encrypted (Https) transfer. OpenClinica™ meets FDA part 11B standards. This includes safety data, laboratory data and outcome data. Safety data will also be collected through electronic diaries which are stored on a secure server.

Sentry is an online secure data entry system developed in-house at PC-CTU and hosted at Oxford. It is designed to collect sensitive data, such as participant and Trial Partner contact details, and securely retain them separate form a trial's clinical data. Sentry can also act as a central participant portal to manage online eligibility, eConsent and ePRO - acting as an intermediary between the participant and the clinical databases. Sentry is accessed via a secure HTTPS connection and all stored sensitive data is encrypted at rest to AES-256 standards. Participant and Trial Partner data will be kept and stored securely for as long as it's required by the study and reviewed on annual basis.

#### F. Qualitative Sub-study

With consent, participants will be contacted for a telephone interview within three months after they complete their day 28 follow up. The researcher will provide study information over the telephone and the Interview Patient PIS, and ICF will be available on the study website and emailed to participants if requested.

Once a practice has completed patient recruitment and one of their patients has been interviewed, we may ask 1-2 healthcare professionals who would be willing to share their experiences of taking part in the trial. Healthcare professionals will include clinicians and non-clinicians with the main criteria for inclusion in interviews being that HCP participants should have carried out trial activities in their practice. Potential HCP participants will be contacted in person

© Copyright: The University of Oxford and Oxford University Hospitals NHS Foundation Trust 2019

or by email by the practice contact. They will be provided with the Interview HCP Invitation Email, Interview HCP PIS and Interview HCP ICF by email.

Patients recruited to both the intervention and usual care arms will be purposively sampled across the recruiting period with approximately 15-20 patients in each arm (30-40 interviews in total). We will seek to obtain maximum variation in age and symptom severity (as reported in daily diary at baseline). When the research team receives responses from HCPs, they will collect basic demographics to purposively select participants based on practice location, practice size, practice patient recruitment and job role. We aim to complete 20-25 interviews with HCPs.

All participants will only be required to take part in a single interview. Patient participant interviews will follow a semi-structured topic guide (Interview Patient Topic Guide) and ask about reasons for consulting and illness perceptions prior to the consultation, experiences of the consultation, the COVID-19 testing process (if applicable, and result if the participant has been notified) and medication adherence. The topic guide will be informed by the Common Sense Model which describes how people perceive and cope with symptoms of illness.
HCP interviews will follow the Interview HCP Topic Guide and will ask about experiences of carrying out trial activities, recruiting patients and the work required to set up a clinical trial during a pandemic.

Interviews with patient participants are expected to last approximately 30-45 minutes and interviews with HCPs are expected to last 15-30 minutes.

Data Collection:
Each interview will be audio-recorded with the participant's permission. Recordings will allow verbatim transcription of interviews. Transcription will be completed by an independent transcription company. Once transcribed and transcripts are checked, audio-recordings will be deleted. Transcripts will be labelled with a unique participant number and will omit any identifiable data either identifying the participant or their general practice.

© Copyright: The University of Oxford and Oxford University Hospitals NHS Foundation Trust 2019

Primary Care
Clinical Trials Unit

PRINCIPLE
Platform Randomised Trial of Treatments in the Community for Epidemic and Pandemic Illnesses

### APPENDIX 3

### MASTER STATISTICAL ANALYSIS PLAN

**P**latform **R**andomised tr**I**al of treatme**N**ts in the **C**ommunity for ep**I**demic and **P**andemic i**L**ln**E**sses Internal Reference Number / Short title: PRINCIPLE

**Ethics Ref:** 20/SC/0158

IRAS Project ID: 281958

**EudraCT Number**: 2020-001209-22

Based on protocol version 8.1 12th April 2021

Version 4.0 Date: 2nd July 2021

Version History

| Version: | Version Date: | Changes: |
|---|---|---|
| 0.1 | 25th March 2020 | MS initial draft |
| 0.2 | 31st March 2020 | Updated post JM initial comments |
| 0.3 | 12 May 2020 | JM/UG writing SAP |
| 0.4 | 10 Aug<br>4th August 2020 | Updated to protocol version 4.0<br>NW updating SAP |
| 0.5 | 11th August 2020 | Merging of 2 0.4 versions |
| 0.6 | 1st October 2020 | Updates following comments from JG, ST and CB<br>Update primary outcomes and sample size<br>Remove references to RCGP data as this is stored in the notes review |
| 0,7 | 15th October 2020 | Updates following meetings with JG, ST, LMY<br>Remove much of text from introduction/DMC sections and refer to protocol/DMC charter<br>Update analysis of binary outcomes to logistic regression |
| 0.8 | 29th October 2020 | Updates following review from LMY<br>Health service use split into participant reported and GP reported health service use<br>Addition of sustained recovery outcome<br>Addition of appendix III – analysis of HYDROXYCHLOROQUINE arm |
| 0.9 | 30th October 2020 | Further update to definition of sustained recovery. Also changed the definition of duration of severe symptoms. |
| 0.10 | 3rd November 2020 | Update following comments from CB |
| 0.11 | 4th November 2020 | Incorporated Jienchi Dorward & Ben Saville's comments |

| | | |
|---|---|---|
| 0.12 | 6th November 2020 | Incorporated Richard Hobbs' comments and made changes following meeting with LMY, RH, CB, BS, JD:<br><br>Updated definition of sustained recovery<br>Change 'consumption' to 'prescription' for antibiotics<br>Addition of WHO ordinal scale |
| 0.13 | 18th November 2020 | Amending typos<br><br>Addition of appendices detailing analysis of azithromycin and doxycycline<br><br>Addition of subgroup analyses |
| 0.14 | 24th November 2020 | Update to protocol v6.0<br>New outcome: new infections in the household<br>Addition of appendix detailing analysis of inhaled corticosteroid arm<br><br>Update subgroup analysis section<br><br>Accept changes from BS and CB from previous version |
| 0.15 | 26th November 2020 | Updates following comments from Philip Hannaford – update to ICS dose and subgroup analyses to include deaths and deaths/hospitalisation combined |
| 1.0 | 1st December 2020 | Change to version 1.0 for signing off |
| 1.1 | 20th January 2021 | Remove viral shedding outcome<br>Update timing of swab results<br>Clarification of censoring for time to event outcomes<br>Addition of moderation analyses |

| | | |
|---|---|---|
| 1.2 | 9th February 2021 | Updated objectives to protocol version 6.3 30.12.2020.<br>Inclusion of Addendum 1.1 and clarification of analysis populations.<br>Updated derivation of: hospitalisation primary outcome (hierarchy of data sources); duration of hospital admission;<br>Definition of last contact date for time to event outcomes<br>Schedule of procedures and flow diagram of trial participants updated<br>Updated timing of swab results<br>Updated handling missing data |
| 2.0 | 10th February 2021 | Signatures |
| 2.1 | 26th February 2021 | Updated to reflect protocol V7.0, addition of colchicine, target population, Appendices. Addition of new secondary outcome |
| 2.2 | 16th March 2021 | Updated to reflect protocol V7.1. Addition of safety as a secondary endpoint. Addition of favipiravir treatment arm. The primary analysis population defined as those with a COVID-19 positive test. |
| 3.0 | 16th March 2021 | Change to version 3.0 for signing |
| 3.1 | 30th March 2021 | Addition of sensitivity analysis specific to Budesonide. Clarification of analysis population for sensitivity analysis. Definition of Vaccination status and addition as covariate, update to definition of sustanined recovery (secondary outcome). Moved derivation of COVID-19 test status to section 2.2.4 |

| | | |
|---|---|---|
| 3.2 | 2nd July 2021 | Updated to protocol version 8.1. Addition of Ivermectin arm<br>Updated definition of vaccination status in section 2.2.5<br>Daily diary added as a source for derivation of COVID-19 status<br>Sources of data for derivation of mortality updated, section 3.2.2.1<br>All cause death or non-elective/urgent hospitalisation within 28 days of randomisation added as secondary outcome<br>Section 3.3.2 Duration of symptoms: derivation of time to event outcomes updated<br>Section 9. Moderation analyses added and intervention specific subgroups specified<br>Section 4.6.4 clarification of population for safety analysis.<br>Updates to intervention specific appendices for colchicine, favipirivir and ivermectin<br>Appendix II Participant flow diagram updated<br>Section 6.2 Removal of secondary analysis of the secondary outcomes using all participants analysis population<br>Section 3.3.2.5 addition of secondary outcome, worsening of symptoms following randomisation. |
| 4.0 | 2nd July 2021 | Signatures |

### 1 INTRODUCTION

#### 1.1 PREFACE

Chief Investigator: Professor Chris Butler

Co-study lead: Professor Richard Hobbs

Senior Trial Manager(s): Dr Hannah Swayze, Dr Emma Ogburn, Dr Emily Bongard, Julie Allen

Trial Manager: Jared Robinson

Data manager: Jenna Grabey

Lead Trial Statistician(s): Dr Ly-Mee Yu and Dr Ben Saville

There are two teams of unblinded statisticians involved in the PRINCIPLE trial. The unblinded statisticians in Oxford Primary Care Clinical Trials Unit (PC-CTU) are responsible for data management, derivation of outcomes, transfer of data to the Statistical Analysis Committee (SAC) of Berry Consultants and analysis of the secondary and safety outcomes. The unblinded SAC is responsible for the interim analysis and to provide a summary of results for the Data Monitoring Safety Monitoring Committee (DMSC) members in an interim analysis report. The SAC will be responsible for the co-primary analyses and any related sensitivity and subgroup analyses requiring a similar Bayesian framework.

This version of the Statistical Analysis Plan was written based on protocol version 7.1 22nd February 2021, and may be updated in the light of further amendments to the study protocol.

#### 1.2 PURPOSE AND SCOPE OF THE PLAN

This Master Statistical Analysis Plan (M-SAP) will detail the statistical design and methods of the PRINCIPLE trial. It will include an appendix titled "Adaptive Design Report" (ADR), which will provide complete specifications for the primary analyses and pre-specified adaptive algorithm. In addition, the M-SAP will be accompanied by arm-specific appendices to describe any planned deviations from the M-SAP. Plans for the analysis of qualitative outcomes is beyond the scope of this statistical analysis plan, and therefore will not be covered in this M-SAP.

Analyses-related decisions may need to be made based on the observed data, such as a review of the distribution of outcome data. These decisions will be made prior to the proposed statistical analyses.

The plan draws on statistical guidance ICH Harmonised Tripartite Guideline: Statistical Principles for Clinical Trials and PSI Guidelines for the Standard Operating Procedures (SOP) for Good Statistical Practice in Clinical Research, the CONSORT statement for operating trials and PC-CTU statistical SOPs.

Analyses will be carried out in accordance with the M-SAP and corresponding appendices. Any additional analysis that is not specified in the M-SAP/appendices or any unplanned deviation(s) from the M-SAP/appendices will be specified in the Statistical Report. Reasons for these changes will be documented and authorised by the Chief Investigator.

Due to the nature of the design of this trial, results for specific treatments will be analysed while the trial is ongoing. This will be done with prior agreement from the Trial Steering Committee (TSC) and DMSC, and the trial team will remain blind to these analyses until such time as the TSC, informed by data and advice from the DMSC, advise that findings should be declared.

#### 1.3 Trial overview

PRINCIPLE is an open, adaptive, platform trial to evaluate emerging treatments for COVID-like-illness. A "platform trial" is a trial in which multiple treatments for the same disease are tested simultaneously. The backbone of the trial is an adaptive clinical trial design. Pre-specified decision criteria allow for dropping a treatment for futility, declaring a treatment superior, or adding a new treatment to be tested. If at any point a treatment is deemed superior to the Usual Care arm, the superior treatment may replace the Usual Care arm as the new standard of care within the trial. However, the primary analysis of subsequent interventions will correspond to the comparison versus the original Usual Care arm. Because the process of dropping and adding treatments may be on-going for an indefinite period of time, platform trials may be better conceived of as a process rather than a single clinical trial. In the context of the COVID-19 pandemic, the trial may continue as long as the pandemic persists.

The PRINCIPLE trial began as a two arm, 1:1 randomised trial but with the capability to add additional interventions over time. The evaluation of any new interventions is governed by the master protocol and M-SAP (including adaptive algorithm and decision criteria), with any planned deviations from the master protocol and M-SAP to be specified in arm-specific appendices. The inclusion of any new interventions will require additional arm-specific appendices to the master protocol and M-SAP.

#### 1.4 Objectives

The primary and secondary objectives as well as time points to evaluate these outcome measures as stated in the protocol.

| | Objectives | Outcome Measures | Timepoint (s) |
|---|---|---|---|
| Primary | To assess the effectiveness of trial treatments in reducing<br><br>1) Time to recovery, for patients | 1) Time to self-reported recovery, defined as the first instance that a participant reports feeling recovered from possible COVID-19, and | Within 28 days of randomisation<br><br>Patient report, Study Partner report, daily online symptom scores |
| | 2) Hospitalisation and/or death. | 2) Hospitalisation and/or death | Within 28 days of randomisation<br><br>Patient report, Study Partner report, medical records |
| Secondary | To explore whether trial treatment will affect<br><br>1) participant-reported illness severity reported by daily rating of how well participant feels<br>2) Duration of severe symptoms and symptom recurrence | Participant reports of daily and monthly (after 28 days) symptoms. | Daily online symptom scores.<br><br>Telephone call or text on days 2, 7, 14 and 28 and once a month for 12 months if data is not obtained through the online diary |
| | 3) Contacts with the health services | Contacts with health services reported by patients and/or captured by reports in patients' medical records if the practice is a member of the RCGP RSC network | GP notes review if available through Oxford RCGP RSC network; otherwise, other sources of routinely collected data after 28 days. Medical notes review for up to 10 years. |
| | 4) Prescription of antibiotics | Bi-weekly reports from participants' primary care medical records | |

| | Objectives | Outcome Measures | Timepoint (s) |
|---|---|---|---|
| | 5) Hospital assessment without admission<br>6) Oxygen administration<br>7) Intensive Care Unit admission<br>8) Mechanical ventilation<br>9) Duration of hospital admission | Patient report/carer report/medical record in primary and secondary care | HES/ONS/EMIS/Medical record data linkage after 28 days if patients have been assessed in hospital |
| | 10) Negative effects on well being | WHO-5 Well Being Index | WHO 5 Well Being Indat baseline, day 14, and day 28 and monthly for up to 12 months, either via online diary or telephone |
| | 11) New infections in household | Reports of new infections in the household (from daily questionnaire) | Within 28 days of randomisation |
| | 12) To determine if effects are specific to those with a positive test for SARS-CoV-2 | Swab test results will indicate an "Intention to Treat Infected" group within the overall cohort for sub analysis. Blood test results on recovery (optional) for evidence of historic COVID-19 | Swab result from medical records, the supporting laboratory and/or convalescent blood test for evidence of historic COVID-19 |
| | 13) To investigate the safety of treatments that are not licensed in the UK | Evaluation of overall safety of drugs by the monitoring of adverse events (AEs as defined in the Intervention specific Appendices (ISAs)) | For the duration of the treatment course and a defined period after the treatment finishes. |
| Qualitative sub-study<br><br>(not covered in this M-SAP) | 1. To explore patients' experiences of consulting, being tested and taking (trial) medication for suspected COVID-19.<br><br>2. To explore healthcare professionals' views of | 1. Telephone interviews with patients.<br><br>2. Telephone interviews with healthcare professionals. | 1. After 28 days. |

| | Objectives | Outcome Measures | Timepoint (s) |
|---|---|---|---|
| | taking part in research during pandemics. | | 2. Once practice has completed recruitment. |
| Intervention(s) | All trial interventions are detailed in the Appendices of the protocol. Further interventions may be added or replaced during the course of the trial, subject to suitable interventions becoming available and all necessary approvals being obtained. | | |
| Comparator | PRINCIPLE began as a two-arm trial, with the intervention arm being Usual Care without the addition of a trial drug. There will be no placebo control in this study. Additional arms may be added as the trial progresses. These will be detailed in the Appendices of the protocol. If an intervention arm is shown to be superior, then this will become the new standard of care. However, the primary analysis of subsequent interventions will correspond to the comparison versus the original Usual Care arm. | | |

### 2 TRIAL DESIGN

PRINCIPLE is an open, adaptive, platform trial to evaluate emerging treatments of COVID-like-illness. A "platform trial" is a trial in which multiple treatments for the same disease are tested simultaneously. The backbone of the trial is an adaptive clinical trial design. Pre-specified decision criteria allow for dropping a treatment for futility, declaring a treatment superior, or adding a new treatment to be tested. If at any point a treatment is deemed superior to the control arm, the superior treatment will replace the control arm as the new standard of care, and all subsequent treatments may be compared to the new standard of care within the trial. Because the process of dropping and adding treatments may be on-going for an indefinite period of time, platform trials may be better conceived of as a process rather than a single clinical trial. In the context of the COVID-19 pandemic, the trial may continue as long as the pandemic persists, and while there is a need to evaluate treatments for acute respiratory tract infections in the community.

The PRINCIPLE trial began as a 1:1 randomised trial of standard care versus standard care plus hydroxychloroquine but with the capability to add additional interventions over time. The evaluation of any new interventions is governed by the master protocol, including adaptive and decision criteria. In addition, the inclusion of any new interventions will require amendments and/or supplements to the protocol and M-SAP.

#### 2.1 ADAPTIVE DESIGN

The pre-specified design will allow adaptations to the trial based on the observed data. These adaptations include the declaration of success or futility of an intervention at an interim analysis, the addition or removal of treatment arms, and changes in the randomisation probabilities. Adaptations will occur at a given interim analysis if pre-specified conditions are satisfied. The adaptive algorithm will be documented in the Adaptive Design Report, including pre-specified criteria for decisions regarding futility or effectiveness of interventions and/or replacing interventions in the trial.

#### 2.2 DEFINITIONS

##### 2.2.1 FOLLOW-UP PERIOD

Patients will be followed from date of randomisation to 28 days by daily symptom diary and/or telephone, and review of their medical records. Participant records will be accessed up to 3 months following randomisation to ascertain outcome data up to 28 days from randomisation. Participants will be followed up on a monthly basis for up to 12 months after enrolment (via email, text message or phone call) to collect information about any ongoing symptoms, hospitalisations and well-being.

##### 2.2.2 LAST CONTACT DATE

In the derivation of time to event outcomes, the last contact date will be defined as the maximum of;

- Last diary entry
- Last call CRF completed
- Date of hospital admission
- Date of hospital discharge

##### 2.2.3 Duration of illness prior to randomisation

Duration of illness prior to randomisation will be derived using the date the participant reported starting to feel unwell (Screening CRF) and date of randomisation. If date patient started to feel unwell (screening CRF) is missing, the date the screening form was completed will be used instead.

##### 2.2.4 Derivation of COVID-19 test status

Participants will be categorised as being infected or not, based on any swab result obtained 14 days or less prior to randomisation and up to and including 7 days following randomisation. Swab result data is collected in the screening CRF, baseline CRF, daily diary, Lab results CRF and notes review CRF.

Any swab result obtained 14 days or less prior to randomisation and up to and including 7 days following randomisation will be used to classify participants into three groups. A positive test result at any time will put them into group 1, regardless of other negative results within the window specified;

1. COVID-19 positive test result
2. COVID-19 Negative test result
3. No swab result available

##### 2.2.5 Vaccination status

Vaccination status will be categorized as “Yes" vs. “other", where “yes" indicates at least one COVID-19 vaccination dose was given two weeks (or more) prior to randomisation and “other" indicates either no vaccination at least 2 weeks prior to randomisation or no vaccine information is available.

#### 2.3 Outcomes

##### 2.3.1 Primary outcome

There are two co-primary outcomes as listed below:

- Time to recovery from suspected COVID-19 infection within 28 days from randomisation, where recovery is defined as the first instance that a participant reports feeling recovered.
- Hospital admission or death related to suspected or confirmed COVID-19 within 28 days from randomisation

##### 2.3.2 Secondary outcomes

###### 2.3.2.1 Patient reported illness severity

Participants are asked to rate how well they are feeling overall each day on a scale of 1-10 (1 being the worst and 10 being the best). This is captured on the patients’ daily diaries and the Call CRF.

###### 2.3.2.2 Duration of severe symptoms and symptom recurrence

Participants are asked to rate their symptoms of fever, cough, shortness of breath, muscle ache, and nausea/vomiting on a four point scale from 0=no problem, 1=mild problem, 2=moderate problem and 3=major problem.

This is captured on patients' daily diaries and Call CRF and is only collected for participants who responded "No" to "Do you feel recovered today?"

The outcomes to be analysed will be time to alleviation of symptoms, time to initial reduction in severe symptoms, time to sustained recovery and time to sustained alleviation of symptoms.

###### 2.3.2.3 Contacts with health services between randomisation and day 28 of follow-up (FU).

This is captured on patients' daily diaries, telephone call CRFs, and medical notes review

###### 2.3.2.4 prescription of antibiotics between randomisation and day 28 of FU

Notes review at 28 days will record information regarding prescription of antibiotics. Hospital assessment without admission overnight

This is captured on patients' daily diaries, telephone call CRFs, notes review and hospitalisation and death CRF.

###### 2.3.2.5 Oxygen Administration

This is captured in patients' daily diaries, telephone call CRF, Notes review and hospitalisation and death CRF.

###### 2.3.2.6 Intensive care unit admission

This is captured in patients' daily diaries, telephone call CRF, Notes review, and hospitalisation and death CRF.

###### 2.3.2.7 Mechanical ventilation

This is captured in patients' daily diaries, telephone call CRF, Notes review and hospitalisation and death CRF.

###### 2.3.2.8 Duration of hospital admission

Patient report/carer report/medical record in primary care and hospital care in relation to duration of hospital admission between date of randomisation and day 28 of follow-up.

This is captured in patients' daily diaries, Call CRF, Notes review and hospitalisation and death CRF.

###### 2.3.2.9 Negative effects on well-being (WHO-5)

The WHO-5 well-being index is collected from daily diaries or telephone call at baseline, day 14 and day 28, and monthly for up to 12 months.

###### 2.3.2.10 New infections in the Household

This is collected in the daily diary and telephone call CRF through the question "has anybody else in your house become unwell today with a respiratory illness?"

###### 2.3.2.11 Safety of treatments not licensed in the UK

Medication side-effects and SAEs will be collected from participant daily diaries, calls to participants/study partners, medical records, notes reviews and RCGP data downloads.

###### 2.3.2.12 All cause death or non-elective/urgent hospitalisation within 28 days of randomisation

#### 2.4 Target population

The trial aims to include symptomatic participants with confirmed, or possible COVID-19 who meet the current NHS case definition for possible COVID-19, and who are well enough to remain in the community. This definition can be found here:

https://www.nhs.uk/conditions/coronavirus-covid-19/symptoms/

Participants must be aged 65 and over, OR aged 18 to 64 and experiencing shortness of breath as part of COVID-19 illness, OR aged 18-64 with certain comorbidities.

Participants experiencing shortness of breath have a greater risk of severe and critical disease outcomes with COVID-19.

The study is for people who have ongoing symptoms.

See protocol for detailed inclusion and exclusion criteria.

#### 2.5 Treatments

Based on version 7.1 of the protocol the main randomisation will be between the following treatment arms (although not all treatments may be available at any one time and not all participants are eligible for all treatments). However, the trial design accommodates treatments being added and dropped as appropriate.

- Usual care
- Usual care plus hydroxychloroquine, 200mg twice daily for 3 days (discontinued)
- Usual care plus azithromycin, 500mg once daily for 3 days (discontinued)
- Usual care plus doxycycline, 200mg on day 1 followed by 100mg daily for 6 days (discontinued)
- Usual care plus the inhaled corticosteroid, budesonide, 400mcg daily (as two puffs bd) for 14 days (discontinued)
- Usual care plus colchicine, 500 microgram (µg) once daily for 14 days Usual care plus favipiravir 1800mg twice a day on day one, and then 800mg twice a day for four days (discontinued)

- Usual care plus favipiravir, 1800mg twice a day on day 1 and then 800mg twice daily for four days
- Usual care plus ivermectin, one dose each day for 3 days at 300µg/kg body weight

Subsequent reference to a treatment group refers to treatment plus usual care, and subsequent reference to usual care group refers to the usual care without a study drug.

#### 2.6 Sample size

Given the open perpetual trial structure, the trial does not have a prespecified end based on sample size. Rather, the trial will continue until either superiority or futility is claimed for each intervention, or until the pandemic expires in the population. We estimate that approximately 400 participants per arm (800 participants total if only a single intervention vs. Usual Care) will be required to provide 90% power for detecting a hazard ratio of 1.3 in the primary population (approximate difference of 2 days in median recovery time). This calculation is based on the assumption of an exponential distribution for time to recovery with a median of 9 days in the Usual Care arm, with some adjustments for missing data and multiple interim analyses. On average, we expect fewer participants to be required when there is a large treatment benefit or complete lack of benefit. For example, if the true hazard ratio is 1.5 (3 day benefit in median time to recovery), on average only 150 subjects per arm are required to provide sufficient power. The primary advantage of the adaptive design is the ability to adapt the sample size to the observed data, thus addressing the primary hypothesis as quickly and as efficiently as possible.

We estimate that approximately 1500 participants per arm (3000 participants total if only a single intervention vs. usual care) will be required to provide 90% power for detecting a 50% reduction in the relative risk of hospitalisation and/or death in the primary analysis population. This calculation is based on the assumption of an underlying 5% combined hospitalisation and/or death rate in the Usual Care arm, with an intervention lowering the hospitalisation and/or death rate to 2.5%, with some adjustments for the multiple interim analyses. We expect fewer participants to be required to detect a 50% reduction if the event rate in the Usual Care arm is greater than 5%.

Virtual trial simulations

Because of the adaptive platform trial structure, there exists no simple formula(s) to calculate power and Type I error (false positive rate). Hence, virtual trial simulations will be used to fully characterize and quantify the power and Type I error of the design. These simulations will be conducted prior to the first interim analysis (with results described in the Adaptive Design Report), and will be used to optimize the adaptive decision criterion and RAR parameters. The simulations will include a comprehensive evaluation of trial performance across a wide range of assumptions (e.g. underlying distribution of outcome in Usual Care arm, treatment effect, accrual rates, etc.). This will include summaries regarding the number of subjects required to make a success or futility conclusions for each intervention. Complete details of the simulations will be provided in the Adaptive Design Report.

#### 2.7 Randomisation and blinding in the analysis stage

Participants will be randomised using a fully validated and compliant web-based randomisation system called Sortition. Once deemed eligible, the clinician or a member of the trial team will randomise the participant. The randomisation process will take only a few moments via the online system and will not delay trial participation. Participants will be randomised to the arm/arms they are eligible for (at least two arms, usual care and at least one intervention), automatically by Sortition.

Initially, randomisation will be fixed 1:1 for a comparison between two trial arms, with stratification by age (less than 65, greater than or equal to 65), and comorbidity (yes/no). If a second intervention arm is added to the study, randomisation allocation will be modified and the additional intervention will be included in the interim analyses (with evaluation for success and futility) as detailed in the Adaptive Design Report. If there are at least 3 arms (2) treatment and Usual Care) in the study, each interim analysis may incorporate modified randomisation probabilities via response adaptive randomisation (RAR). Full details for implementing RAR will be provided in the Adaptive Design Report; the general idea is to allocate more participants to the intervention arms that have the best observed outcomes. Except for the CTU programmer, the rest of the trial team are blinded to RAR ratios.

PRINCIPLE is an open-label trial. The participant, a person guiding the participant through the randomisation process, and the participants' primary care clinician will know the participant's allocation. Therefore, no unblinding or code breaking is required in the event of a relevant emergency. However, those managing the data will be blind to participant allocation.

The trial team and recruiting clinicians will be blinded to emerging results. During the course of the trial, only the unblinding statisticians and independent members of the Data Monitoring and Safety Committee will have access to the unblinded interim results.

### 3 Data management and Derivation of outcomes

#### 3.1 Sources of Data

Data is collected from multiple sources. For the derivation of outcomes, should data be collected from more than one source, we will specify which source should be utilised. For example, if data for an outcome are obtained from both daily diary and telephone calls, the daily diary data will be utilised first.

(i) Death and hospitalisation CRF
(ii) Online daily diaries for 28 days, and then monthly up to 12 months
(iii) Telephone call CRF at day 7, 14 and 28, and then monthly up to 12 months
(iv) Lab results CRF
(v) Notes review CRF
(vi) When available, the Secondary Uses Services data, which is a collection of healthcare data in England provided by NHS Digital, and other sources of hospital data.

Data management for derivation of primary and secondary outcomes will be carried out by PC-CTU statisticians, and primary outcomes (and any data that are relevant to the analysis of the primary outcomes) data transferred to SAC. The PC-CTU unblinded statistician will ensure that data transfer to the SAC for each treatment comparison includes only participants that meet the definition of the analysis population as defined in Section 4.6.

#### 3.2 Primary outcomes

##### 3.2.1 Time to recovery

The first primary outcome is time to recovery from suspected COVID-19 infection within 28 days from randomisation, where recovery is defined as the first instance that a participant reports feeling recovered.

Time (in days) taken to self-reported recovery will be computed as time to reported “Yes” to the question “Do you feel recovered today (I.e. symptoms associated with illness are no longer a problem)?”. The variable is recorded as (**WELLYN**=1). This will be calculated as date from randomisation to date (VISDAT_P) of participant self-report of recovery if using the patient diary. If the call CRF is being used then the date to feeling fully recovered is ‘**WELLDAT**’. Where patient recall of date of feeling recovered (as recorded on the call CRF) is prior to date of randomisation, the date of feeling recovered will be assumed to be day 0.

If a participant has more than one date for when they reported feeling recovered (i.e. relapse or more than one data source completed), the date of first reported recovery will be taken.

If the participant is in hospital on the date of feeling recovered then this will still be counted as a recovery (hospitalisation recorded in any 1 or more of the following sources: notes review; death and hospitalisation CRF; daily diaries; call CRF).

Participants who withdraw or are lost to follow-up will be censored at their last contact date (definition 2.2.2).

Participants who die will be censored at 28 days.

##### 3.2.2 Hospital admission or death

The second primary outcome is hospital admission or mortality related to suspected or confirmed COVID-19 measured within 28 days from randomisation.

###### 3.2.2.1 Derivation of mortality

Mortality related to suspected COVID-19 within 28 days of randomisation

Data collected via the death and hospitalisation CRF, call CRF, medical notes review, electronic health record data such as HES, mortaility data, SUS, and GP prescription data provided by NHS Digital, and data extracted from RSC, will be used to derive the primary outcome. If the following is true from any source then the patient will be recorded as having death related to suspected COVID-19 within 28 days of randomisation.

**DDYN** =1 AND date of death (**DDDAT**) ≤28 days from randomisation AND COVID-19 contributed to death [**DD_CV19=1**].

If death is recorded as "Yes" but the COVID-19 related variable is missing, then the outcome will be cross checked with the SAE data. Any death not related to suspected COVID-19 should be recorded as an SAE.

###### 3.2.2.2 Derivation of hospital admission

Hospital admission within 28 days will be defined as an overnight stay in hospital and likely to be related to COVID-19. This can be recorded from the daily diaries, calls at days 7, 14 and 28, notes review CRF, death and hospitalisation CRF and data from NHS digital. The data sources will be considered in the following hierarchy; (1) Patient reported (daily diaries, calls, hospitalisation CRF), (2) Notes review, (3) data from NHS digital. The NHS digital data will only be used if no data can be obtained from sources 1 and 2. Data from NHS digital will be adjudicated by two clinicians blind to randomised group as to whether the hospital admission is likely to be related to COVID-19. Hospital admissions on the day of randomisation will be assumed to have occurred post randomisation and will be included in the analysis.

###### 3.2.2.2.1 Death and hospitalisation CRF

Participant attended hospital [**HOSP_HOOCCUR = 1**] AND attendance likely related to COVID-19 [**CVYN** = 1] AND [number of days in hospital > 1 or not yet discharged [**HOSP_HOENYN=0**] AND date they were admitted to hospital (**HOSP_HOSTDAT**) is ≤28 days from date of randomisation.

###### 3.2.2.2.2 Daily diary data

Have you attended hospital in the last 24 hours [**HOSP_HOOCCUR** = 1] AND attendance likely related to COVID-19 [**CVYN** = 1] AND admitted overnight [**HOSP_NIGHTYN** = 1] AND first date of overnight hospital stay (**HOSP_HOSTDAT**) is ≤28 days from date of randomisation.

###### 3.2.2.2.3 Calls at 7, 14, 28 days

Admitted to hospital [**HOSP_HOOCCUR** = 1] AND was the admission related to suspected COVID-19 infection [**CVYN** = 1] AND admitted overnight [**HOSP_NIGHTYN** = 1] AND first date of overnight hospital stay (**HOSP_HOSTDAT**) is ≤28 days from date of randomisation.

###### 3.2.2.2.4 Notes review

Participant attended hospital [**HOSP_HOOCCUR** = 1] AND attendance likely related to COVID-19 [**CVYN** = 1] AND admitted overnight [**HOSP_NIGHTYN**=1] AND date they went to hospital is ≤28 days from date of randomisation.

If either hospital admission (as derived above) OR mortality (as derived above) occurs then this primary outcome has occurred (Primary outcome = 1).

Early versions of online daily diaries and call CRF did not include a qualification for suspected COVID-19 infection so these will be cross checked with the SAE data. Any hospitalisation or death not thought to be related to suspected COVID-19 should be recorded as an SAE

#### 3.3 Secondary outcomes

##### 3.3.1 Patient reported illness severity

Participants are asked to rate how well they are feeling each day on a scale of 1-10 (1 being the worst and 10 being the best). This is captured on the participants' daily diaries and the Call CRF.

Day 7, 14 and 28 data will be obtained from the call CRF if not available in the daily diary.

Four variables will be derived; illness severity on day 7, illness severity on day 14, illness severity on day 21 and illness severity on day 28. If day 7, 14 or 28 is not available from the daily diaries, then data from the call CRF will be used. If no data available from diaries or calls on days 7, 14, 21 and 28 illness severity will be regarded as missing for that time point.

##### 3.3.2 Duration of severe symptoms

Participants are asked to rate their symptoms of fever, cough, shortness of breath, muscle ache, nausea/vomiting, diarrhoea, generally feeling unwell, loss of smell/taste (added 26th February 2021), headache (added 26th February 2021) and abdominal pain (added 26th February 2021) on a four point scale from 0=no problem, 1=mild problem, 2=moderate problem and 3=major problem. From the start of the trial until 22nd March 2021 participants were asked to rate their symptoms whilst they did NOT feel recovered. From 23rd March 2021 participants were asked to rate symptoms irrespective of recovery.

'Severe' symptoms will be defined as a score of 3 (major) on the four point rating scale.

For time to event outcomes for overall symptoms, the following symptoms will be considered; fever, cough, shortness of breath, muscle ache, nausea/vomiting, diarrhoea, generally unwell, loss of smell/taste, headache and abdominal pain.

Participants can have a missing symptom severity rating for the following reasons

- Participant feels recovered so was not required to complete symptom data (before 23rd March 2021)
- Symptom was added to daily diary CRF at a later date than when patient completed the diary

If a participant has reported they have recovered AND not rated individual symptoms, they will be assumed to have a rating of 0 for symptom severity for each symptom not rated.

For each participant, only symptoms that were collected at baseline will be used when assessing the overall and individual time to event symptom outcomes.

###### 3.3.2.1 Time to alleviation of symptoms

Time to alleviation of symptoms will be defined as the time from randomisation to symptoms being rated as mild or none. For those who have call data only, the time to alleviation will be defined as the day the call was made. Patients who report none/mild symptoms at baseline will be censored at time 0. Participants who withdraw or are lost to follow-up will be censored at their last contact date (definition 2.2.2).

Time to alleviation of overall symptoms will be the time for all symptoms specified in section 3.3.2 to be rated as none or mild.

###### 3.3.2.2 Time to initial reduction of severity of symptoms

Time to initial reduction of severity of symptoms will be defined as time to reduction in severity of each individual symptoms to at least one grade lower. For those who have call data only, the time that the symptoms reduced to at least one grade lower will be defined as the day the call was made. This will be calculated for each symptom and also for overall symptoms. Participants who withdraw or are lost to follow-up will be censored at their last contact date (definition 2.2.2). Participants who die will be censored at 28 days. Participants with symptoms rated as none at baseline will be censored at time 0.

###### 3.3.2.3 Time to Sustained Recovery

Time to sustained recovery will be defined as the time to first reported recovery on the question 'Do you feel recovered today?' with subsequent responses of 'yes' to this question until day 28. For those who have call data only, the time to sustained recovery will be defined as the date at which they felt recovered or if this information is missing, the date the call was made. Subsequent calls must report feeling recovered. Participants who withdraw or are lost to follow-up will be censored at their last contact date (definition 2.2.2). Participants who die will be censored at 28 days. Where information from call data is not consistent with diary data, the diary data will take precedence. Where call data at days 7, 14 and 28 days report inconsistent dates of recovery, the date furthest from randomisation will be used as the date for sustained delivery (assuming recovery sustained).

In addition a binary variable shall be derived as sustained recovery or otherwise.

Sustained recovery will be derived as being recovered within the first 14 days and reports feeling recovered for the next 14 days. Where the participant has only call data, they require to have reported feeling recovered at both 14 days and 28 days. Where participants have got incomplete diary data for days 14-28 and no call data at 28 days, they are classified as otherwise.

Only participants who have reached day 28 follow-up will be included in the analysis of time to sustained recovery.

###### 3.3.2.4 Time to Sustained symptoms alleviation

This will follow the same principle as time to initial reduction in severity of symptoms but there must be no subsequent symptom severity recorded as moderate or major. This will be calculated for individual symptoms separately and also for overall symptoms. Time to sustained allevation for overall symptoms will be the time for all symptoms specified in section 3.3.2 to be rated as mild or none with no subsequent symptom severity coded as moderate or major.

###### 3.3.2.5 worsening of symptoms after randomisation

Participants who report symptoms worsening over the period of 28 days follow up. Participants will be categorised with respect to each symptom into two categories (i) worsening symptoms and (ii) no change or improved during the 28 day period from randomisation.

##### 3.3.3 Contacts with health services between randomisation and day 28 of follow-up.

This will be split into participant reported health service use and health service use from GP records.

###### 3.3.3.1 Participant reported health service use

Sources of data for this outcome are participant daily diaries and telephone call CRF (day 7, day 14 and day 28).

This will be presented as 2 outcomes:

1. A binary outcome indicating whether the participant has had any contact with health services during 28 days of follow-up. This will be 'yes' if any of the following are recorded as yes in the daily diary or call CRF:

    - GP (**GP_HOOCCUR**)
    - Other primary care services (**PCS_HOOCCUR**)
    - NHS 111 and other central advice resources (**NHS_HOOCCUR**)
    - A&E (**AE_HOOCCUR**)
    - Hospital (**HOSP_HOOCCUR**)
    - Other (**OTH_HOOCCUR, OTH_HOOCCUR_DEF**– free text)

2. A continuous variable of the number of health service contacts whilst alive during 28 days of FU.

    The number of health service contacts from the diary data will be the total number of times the participant has responded 'yes' to any of the following:

    **GP_HOOCCUR, PCS_HOOCCUR, NHS_OOCCUR, AE_OOCUR, OTH_OOCCUR**

    If the diary data is missing then the call CRF data will be utilised. The number of health service contacts from the call CRF will be the total of the following for each of the 7, 14 and 28 day calls:

    **GP_HOOCCURNUM, PCS_HOOCCURNUM, NHS_HOOCCURNUM, AE_HOOCCURNUM, OTH_HOOCCURNUM**

###### 3.3.3.2 Health Service Use From GP Records

Data for this outcome will come from the GP notes review after 28 days.

This will be presented as 2 outcomes:

1. A binary outcome indicating whether the participant has had any contact with health services during 28 days of FU. This will be 'yes' if any of the following are recorded as yes in the notes review:
    - GP (**GP_HOOCCUR**)
    - Other primary care services (**PCS_HOOCCUR**)
    - NHS 111 and other national resources (**NHS_HOOCCUR**)
    - A&E (**AE_HOOCCUR**)
    - Hospital (**HOSP_HOOCCUR**)
    - Other (**OTH_HOOCCUR, OTH_HOOCCUR_DEF**– free text)

2. A continuous variable of the number of health service contacts whilst alive during 28 days of FU. This will be derived by totalling the following for those with a notes review:

    **GP_NUM, PCS_NUM, NHS_NUM, AE_NUM, OTH_NUM**

##### 3.3.4 Prescribing of antibiotics between randomisation and day 28 of FU

Notes review at 28 days will record information regarding prescription of antibiotics.

- Antibiotic prescribed (Yes = 1, No=1) (**ATBYN**)

This outcome will consider prescription of antibiotics whilst alive to account for truncation by death.

###### 3.3.4.1 Derivation of outcome

Antibiotic prescribed = Yes, IF prescribed antibiotic [**ATBYN**=1] AND start date of antibiotic ≤28 days from data of randomisation. If case note review is available for a participant but no information is provided regarding antibiotics, it will be assumed that NO antibiotic was prescribed. If no notes review is available for a participant this will be recorded as missing with respect to antibiotic prescribing.

##### 3.3.5 Hospital assessment without admission

This is captured on patients' daily diaries, Call CRF, notes review and the death and hospitalisation CRF. All sources will be considered and if any indicates a hospital assessment without admission it will be coded as such. If there are discrepancies between the sources of data the diary data will be assumed to be correct.

###### 3.3.5.1 Derivation

Notes review, patient diary and call CRF:

Hospital assessment without admission = YES if on any day the participant reports going to hospital [**HOSP_HOOCUR** = Yes] AND on that date they were not admitted overnight [**HOSP_NIGHTYN** = No]

Death and hospitalisation CRF:

Hospital assessment without admission = YES if the patient has been admitted to hospital since they joined the trial (**HOSP_HOOCCUR**=1) and Date of admission (**HOSP_HOSTDAT**) = date of discharge (**HOSP_HOENDAT_DD**)

##### 3.3.6 Oxygen administration

This is captured in patients' daily diaries, Call CRF, Notes review and death and hospitalisation CRF.

This outcome will reflect oxygen administration whilst alive to account for truncation due to death

###### 3.3.6.1 Derivation

Oxygen administration = yes if patient reports receiving oxygen whilst in hospital [**OXY_HOOCCUR**=Yes] AND date of admission to hospital is ≤28 days from date of randomisation.

If oxygen use is reported via any data source then it will be counted as yes.

##### 3.3.7 Intensive care unit admission

This is captured in patients' daily diaries, Call CRF, Notes review and the death and hospitalisation CRF. It will be calculated in 2 ways:

1. To reflect ICU admission whilst alive to account for truncation due to death prior to ICU admission. This definition of the outcome would give an estimate of the requirement for ICU between the randomised groups from a healthcare resource use perspective. This is derived as:

   Intensive care unit admission = yes if report of participant staying in ICU [ICUYN=Yes or ICU_HOOCCUR=Yes] AND date of admission to ICU is ≤28 days from date of randomisation. If intensive care unit admission is reported via any data source then it will be counted as an ICU admission.

2. To consider this outcome from a patient benefit perspective the outcome will be defined as a composite by assigning a "poor" outcome (i.e. ICU admission) to participants who die before requiring ICU admission. This is derived as:

   Intensive care unit admission = yes if report of participant staying in ICU [**ICUYN**=Yes or **ICU_HOOCCUR**=Yes] AND date of admission to ICU is ≤28 days from date of randomisation) OR participant has died.

If an ICU admission is recorded on any data source then it will be coded as an ICU admission.

##### 3.3.8 Mechanical ventilation.

a. This outcome will reflect mechanical ventilation whilst alive to account for truncation due to death. This definition of the outcome would give an estimate of the requirement for ventilation between the randomised groups from a healthcare resource use perspective.
b. To consider this outcome from a patient benefit perspective the outcome will be defined as a composite by assigning a “poor” outcome to participants who die before requiring ventilation.

This is captured in patients’ daily diaries, Call CRF, Notes review and death and hospitalisation CRF and captured from the question related to receiving mechanical ventilation.

###### 3.3.8.1 Derivation

(a) Mechanical ventilation = yes if report of participant on mechanical ventilation [**MV_HOOCCUR**=Yes] AND date of admission to hospital is ≤28 days from date of randomisation.
(b) Mechanical ventilation = yes if participant dies OR (participant on mechanical ventilation AND date of admission to hospital is ≤28 days from date of randomisation [**MV_HOOCCUR**=Yes])

If mechanical ventilation is recorded on any data source then it will be coded as mechanical ventilation=YES.

##### 3.3.9 Duration of hospital admission

This is captured in patient diaries, Call CRF, notes review and death and hospitalisation CRF.

The duration of hospital admission is calculated in the following ways from the different data sources:

Notes review: **HOSP_DUR**

Death and hospitalisation CRF: Difference between date of admission and date of discharge

Participant diaries: **HOSP_DUR**

Call CRF: **HOSP_NONIGHTS**

If a participant has been admitted with suspected COVID-19 more than once in the FU period of 28 days, the duration of hospital stay will be the sum of all admissions during FU, truncated at day 28.

All sources of data will be considered and if any indicates a hospital stay it will be coded as such. If there are discrepancies between the sources of data the diary data will be assumed to be correct. Only participants with a hospitalisation likely to be related to COVID-19 will be included in the analysis.

Participants admitted with likely COVID-19 and who die whilst in hospital will not be included in the estimate of mean duration and the number in each arm who die whilst in hospital will be reported in a footnote.

##### 3.3.10 Negative effect on well-being (WHO-5)

Well-being is measured using the WHO well-being index which includes 5 items relating to well-being measured on a five point scale (scale of 5 =all of the time, 4=most of the time, 3=more than half the time, 2=less than half the time, 1=some of the time, 0=at no time). A total score is computed by summing the scores to the five individual questions to give a raw score ranging from 0 to 25 which is then multiplied by 4 to give the final score from 0 representing the worst imaginable well-being to 100 representing the best imaginable well-being. Negative effect on well-being is collected at baseline, 14 days and 28 days via the daily diary and call CRF.

From a patient benefit perspective participants who die before the measured time point will be given a score of 0 for that time point and included in the analysis.

##### 3.3.11 WHO Ordinal Scale of Clinical Progression

There are various versions of this scale. We will use a score based on a number of factors including hospitalisation, use of oxygen, ventilation and death. It ranges from 1 (not hospitalised) to 6 (dead) and is defined as follows:

1 = Not hospitalised

2 = Hospitalised without need for supplemental oxygen

3 = Hospitalised with need for supplemental oxygen

4 = Hospitalised with need for non-invasive positive pressure ventilation or high-flow nasal cannula

5 = Hospitalised with need for mechanical ventilation or extracorporeal membrane oxygenation

6 = Death

This outcome will be derived at days 7, 14 and 28.

##### 3.3.12 New infections in the household

This will be coded as ‘yes’ if the answer to the question “has anybody else in your house become unwell today with a respiratory illness?” is yes in the daily diary and/or the telephone call CRF at any time during the 28 days after randomisation.

##### 3.3.13 Safety of treatments not licensed in the UK

For each treatment not licensed in the UK, the following AEs from the start of medication until the specified follow-up period, will be assessed by a clinician for causality and severity (See protocol for definitions): i) pre-defined AEs detailed in the intervention specific appendices (ISA) that are rated by the participant as ‘moderate’ and ii) other reported ‘major’ AEs. Severity will be rated by the clinician as Grade 1 (mild), grade 2 (moderate) and grade 3 (severe).Derivation of COVID-19 positive test result population

##### 3.3.14 All cause death or non-elective/urgent hospitalisation within 28 days of randomisation

The derivation will follow that detailed in sections 3.2.2.1 and 3.2.2.2 with no requirement for cause of death or reason for admission to be related to suspected COVID-19. Hospitalisations will be limited to non-elective/urgent admissions.

### 4 Analysis – General considerations

The following sections detail the final analysis for any treatment comparison within 28 days of randomisation. Methods for the primary analyses, including interim analyses, are specified in the Adaptive Design Report.

#### 4.1 Participant throughput

The flow of participants through the trial will be reported following CONSORT and will include number of participants randomly assigned, receiving allocated treatment, followed up, withdrawn and analysed for primary outcome. Protocol deviations and information regarding screening information and number of ineligible participants randomised will be reported.

#### 4.2 Descriptive statistics

Baseline characteristics of participants will be summarised both overall and by randomised group, including stratification factors and important prognostic, demographic and clinical characteristics. Binary and unordered categorical variables will be summarised using number, number missing and proportions. Continuous variables that are approximately normally distributed will be summarised using number, number missing, mean and standard deviation. Continuous variables that are not normally distributed or ordered categorical variables will be summarised using number, number missing, median and interquartile range.

There will be no tests of statistical significance nor confidence intervals for differences between randomised groups with respect to any baseline variable.

#### 4.3 Characteristics of participants

Characteristics of participants to be described include age, sex (male/female/other), presence of comorbidities (Asthma COPD or other lung disease, Diabetes, Heart problems, High blood pressure, Liver disease, Stroke or other neurological problem), duration of symptoms prior to randomisation, symptoms (fever, cough shortness of breath, muscle ache and nausea/vomiting, other) rated as no problem, mild, moderate or major, medications, use of antibiotics , contact with health care services (GP, other primary care services, NHS 111, A&E, Hospital and other), test results for SARS-CoV-2 infection (not tested, missing, positive, negative), care home residency and ethnicity (collected at 28 days in addition to baseline in case these data were missed at baseline). Stratification factors will be described.

Tables of baseline characteristics will include only participants in the analysis population (as per section 4.6 i.e. excluded participants post randomisation will not be included). The number of randomised participants excluded from the analysis will be reported.

#### 4.4 DESCRIPTION OF AVAILABLE DATA

The number of participants with available data for primary and secondary outcomes for the final analysis will be reported by treatment group.

Details describing methods for dealing with missing data with respect to the primary outcome will be described within the Adaptive Design Report (ADR).

#### 4.5 COMPLIANCE WITH MEDICATION

Participants are asked in the online daily diaries to record whether they have taken their medication and if not, the reason why. The call CRF records the number of days they took the trial medication. For participants in randomised groups receiving medication, compliance with medication will be reported. The number of days that the allocated medication was taken will be reported along with withdrawals from treatment.

#### 4.6 DEFINITION OF POPULATION FOR ANALYSIS

The analysis population will include all participants as defined by the protocol eligibility criteria.
As per ICH E9 guidance the following participants will be excluded from the analysis population;
(a) Participants randomised but subsequently found to be not eligible for randomization
(b) Participants previously randomised to an arm in the PRINCIPLE trial (subsequent randomisations will be excluded)
In addition, the following participants will be excluded;
(c) Participants who withdraw consent for data linkage and notes review and for whom no outcome data has been collected.

##### 4.6.1 PRIMARY ANALYSIS POPULATION

The primary analysis population is defined as all randomised participants with a COVID-19 positive test for whom data were available with participants analysed according to the groups they were randomly allocated to, regardless of deviation from protocol.. In the setting of an adaptive platform trial with sequential overlap of treatment arms, the primary analysis for each intervention may be based on a distinct population. For each intervention that stops randomisation (due to futility) or becomes standard of care (due to superiority) based on interim analysis criteria, the date of the implementation of change in randomisation per interim decision will determine the intervention primary analysis population. For example, if randomisation is stopped for an intervention due to interim futility criteria, the final primary analysis includes all trial participants enrolled up to the date when randomisation was stopped to that intervention. If the randomisation is stopped for an intervention for other reasons (e.g. external reasons not related to the trial design), the primary analysis population will be specified in the intervention specific Appendix. Generally speaking, the primary analysis for a given intervention will be based on complete 28-day follow-up of these participants, but the duration of follow-up may be impacted by public disclosure of interim results as determined by the Trial Management Group.

###### 4.6.2 Concurrent Randomised and Eligible Analysis population

Analysis of the secondary outcomes for a given intervention will be based on participants who were randomised to the usual care arm during the same time frame when the intervention was actively randomised and who were eligible for randomisation to the intervention (i.e. Concurrent Randomised and Eligible Analysis Population).

###### 4.6.3 Concurrent Randomisation Analysis Population

Sensitivity analyses of the primary analysis for a given intervention will be based on participants with a positive COVID-19 test result randomised during the same time frame when the intervention was actively being randomised, i.e. a concurrent randomisation analysis population.

###### 4.6.4 Safety Analysis Population

Safety analysis will be conducted on the as treated population (i.e. the treatment that participants have received). Participants randomised to receive an intervention, but whom do not receive , or do take at least one dose of the intervention or for whom there is no information will belong to the usual care (non-treatment arm) arm for the safety analysis.  Safety analysis will be conducted using the concurrent randomisation population, irrespective of COVID-19 swab status.

##### 4.7 Comparative analysis

For all outcomes the primary analysis will be performed on the primary analysis population at 28 days after randomisation.

Each treatment arm will be compared with the usual care arm.  If a treatment is deemed superior to usual care on both co-primary endpoints and replaces the usual care arm as the new standard of care, subsequent treatments will be compared with the original usual care arm.

There will be no formal adjustment for multiple comparisons.

##### 4.8 Pooling of investigational sites

Data from all sites will be combined and analysed collectively. A sensitivity analysis of the primary outcomes may be carried out adjusting for geographical clinical research network (CRN) if deemed necessary.

##### 4.9 Data Monitoring Committee And Interim Analyses

Details of the data monitoring committee and interim analyses can be found in the interim analysis report and the DMC charter.

### 5 PRIMARY ANALYSIS

#### 5.1 PRIMARY OUTCOMES

Refer to ADR for details.

##### 5.1.1 SUMMARY OF BAYESIAN ANALYSES TO BE CONDUCTED BY STATISTICAL ANALYSIS COMMITTEE

Below is a summary of the pre-specified Bayesian analyses that may be requested from the Statistical Analysis Committee (SAC) for interventions that stop randomisation (due to futility) or become standard of care (due to superiority) based on interim analysis criteria:

B1. Bayesian interim analysis that satisfies the interim decision criteria

a. Details in ADR
b. Subgroup estimates from Bayesian model corresponding to categories in ADR)

B2. Bayesian primary analysis on “final data”

a. Details in ADR and M-SAP
b. Include subgroup estimates

B3. Sensitivity analysis: Repeat Bayesian primary analysis on concurrent randomisation analysis population

a. Details in ADR
b. Include subgroup estimates

B4. Secondary analysis: Repeat Bayesian primary analysis on overall population (i.e. regardless of COVID-19 test status). These models will be identical to the primary analysis models, but will include an additional baseline covariate and corresponding parameters for COVID-19 status (negative, positive, unknown).

a. Details in M-SAP
b. Includes subgroup estimates

The specific pre-specified Bayesian analyses required for each intervention will be specified in the Appendix.

### 6 SECONDARY ANALYSIS

#### 6.1 PRIMARY OUTCOME ON OVERALL POPULATION

The primary outcomes will be analysed using the same method as detailed in the adaptive design report, but using the overall population with additional baseline covariate for COVID-19 test status (see B4 above).

#### 6.2 SECONDARY OUTCOMES

For all secondary outcomes, the analysis will compare each treatment arm with the usual care arm, in a pairwise comparison. For each analysis the Concurrent Randomisation and Eligible Analysis Population will be used. The analysis will be conducted on the Concurrent Randomisation and Eligible Analysis population with a COVID-19 positive test result. Regression models (appropriate for each endpoint) will include randomised group (treatment/usual care) and stratification factors (age (continuous), comorbidity (Yes/no)). They will also include duration of illness at randomisation and vaccination status (as far as possible). Should it be necessary to compare more than one intervention with control at the same time, a covariate indicating which arms of the trial the participant was eligible to be randomised to will also be included. For binary outcomes with a low event rate, results will be reported descriptively by treatment group and a Chi-square test or Fisher's exact test may be used instead of the analysis detailed below. For continuous outcomes where the data are skewed, alternative non-parametric methods will be considered.

##### 6.2.1 PATIENT REPORTED ILLNESS SEVERITY

A linear mixed model will be used to analyse this outcome. The illness severity at each time point (7, 14, 21 and 28 days) will be included as the response variable, along with randomised group, age, presence of comorbidity, eligibility for treatment arm, duration of illness prior to randomisation vaccination status and time as fixed effects. Participant will be included as a random effect. Mean scores at each time point by randomised group will be described graphically.

##### 6.2.2 DURATION OF SEVERE SYMPTOMS

###### 6.2.2.1 TIME TO ALLEVIATION OF SYMPTOMS

Time to alleviation of symptoms will be compared between each treatment arm with the usual care arm using Cox proportional hazards model, adjusting for randomised group, age and presence of comorbidity at baseline. The model will also be adjusted for duration of illness (days) prior to randomisation and vaccination status. This will be calculated as the date of randomisation minus the start date of symptoms as reported on the screening CRF. If this date is unavailable then the date the patient registered with Principle will be used. The adjusted hazard ratio and 95% CI will be estimated from the model. A Kaplan Meier plot will also be presented. If the assumption of proportionality is not met, then an alternative survival model such as restricted mean survival method will be used.

Separate analyses will be carried out for overall and for each symptom separately.

###### 6.2.2.2 TIME TO INITIAL REDUCTION OF SEVERITY OF SYMPTOMS

This will be analysed using the method described for time to alleviation of symptoms.

##### 6.2.3 TIME TO SUSTAINED RECOVERY

This will be analysed using the method described for time to alleviation of symptoms.

##### 6.2.4 Time to Sustained symptoms alleviation

This will be analysed using the method described for time to alleviation of symptoms.

##### 6.2.5 Contacts with health care services

The number and percentage of participants with at least one contact with health services will be presented for treatment and usual care groups. This outcome will be analysed using a logistic regression model. Randomised group, age, presence of comorbidity, duration of illness and vaccination status will be included as covariates. The adjusted odds ratios will be reported for each pairwise comparison along with their associated 95% confidence interval and P-value. In addition, the adjusted absolute risk differences will be reported.

The number of contacts with health care services over 28 days will be analysed using a Poisson model. Randomised group, age, presence of comorbidity, duration of illness and vaccination status will be included as covariates. Adjusted incidence ratios will be presented with their 95% confidence intervals and related P value.

##### 6.2.6 Prescribing of antibiotics

The count and percentage of participants with an antibiotic prescription will be presented for each treatment group. This outcome will be analysed using a logistic regression model. Randomised group, age, presence of comorbidity, duration of illness and vaccination status will be included as covariates. The adjusted odds ratios will be reported for each pairwise comparison along with their associated 95% confidence interval and P-value. In addition, the adjusted absolute risk differences will be reported.

##### 6.2.7 Hospital assessment without admission

The count and percentage of participants with hospital assessment without admission will be presented for each treatment group. This outcome will be analysed using a logistic regression model. Randomised group, age, presence of comorbidity, duration of illness and vaccination status will be included as covariates. The adjusted odds ratios will be reported for each pairwise comparison along with their associated 95% confidence interval and P-value. In addition, the adjusted absolute risk differences will be reported.

##### 6.2.8 Oxygen administration

The count and percentage of participants with oxygen administration will be presented for each treatment group. This outcome will be analysed using a logistic regression model. Randomised group, age, presence of comorbidity, duration of illness and vaccination status will be included as covariates. The adjusted odds ratios will be reported for each pairwise comparison along with their associated 95% confidence interval and P-value. In addition, the adjusted absolute risk differences will be reported.

##### 6.2.9 Intensive care unit admission

The count and percentage of participants with intensive care unit administration will be presented for each treatment group. This outcome will be analysed using a logistic regression model. Randomised group, age, presence of comorbidity, duration of illness and vaccination status will be included as covariates. The adjusted odds ratios will be reported for each pairwise comparison along with their associated 95% confidence interval and P-value. In addition, the adjusted absolute risk differences will be reported.

##### 6.2.10 Mechanical ventilation

The count and percentage of participants with mechanical ventilation will be presented for each treatment group. This outcome will be analysed using a logistic regression model. Randomised group, age, presence of comorbidity, duration of illness and vaccination status will be included as covariates. The adjusted odds ratios will be reported for each pairwise comparison along with their associated 95% confidence interval and P-value. In addition, the adjusted absolute risk differences will be reported.

##### 6.2.11 Duration of hospital admission

The mean duration of hospital admission will only be considered for those with a hospital admission and will be compared between each treatment arm with the usual care arm using a multiple linear regression model. The model will include outcome as the response variable, randomised group, age, presence of comorbidity, duration of illness and vaccination status as covariates. The mean (SD) duration will be presented in each group and the adjusted difference in means and 95% CI for each pairwise treatment arm comparison with the usual care group.

##### 6.2.12 Well-being

The distribution of the WHO well-being index will be considered. Assuming there are not a large number of deaths or hospitalisations, a linear mixed effect model will be fitted to the data. Baseline well-being score will be fitted as a covariate in the model. Fixed effects will include randomised group, age, presence of comorbidity, duration of illness, vaccination status, time and a time x randomised group interaction. The mean (SD) well-being score at 14 and 28 days will be reported for each group and the adjusted difference in means (95% CI) for each pairwise treatment comparison with the usual care group will be presented.

##### 6.2.13 WHO Ordinal Scale of Clinical Progression

If the data are available to calculate this scale then the following analysis will be carried out. The number and percentage of participants on each level of the scale will be presented by treatment group at days 7, 14 and 28. The outcome will be analysed using an ordinal logistic regression model, including the following covariates: randomised group; age; presence of comorbidity, duration of illness and vaccination status.

##### 6.2.14 New infections in the Household

The count and percentage of participants with a new infection in the household will be presented for each treatment group. This outcome will be analysed using a logistic regression model. Randomised group, age, presence of comorbidity, duration of illness and vaccination status will be included as covariates. The adjusted odds ratios will be reported for each pairwise comparison along with their associated 95% confidence interval and P-value. In addition, the adjusted absolute risk differences will be reported.

###### 6.2.15 Safety of treatments not licensed in the UK

For treatment arms that are not licensed in the UK, the number and percentage of participants reporting AEs will be reported by grade of severity.

###### 6.2.16 All cause mortality and hospitalisation within 28 days of randomisation

The count and percentage of participants with all cause mortality or hospital admission will be presented for each treatment group. This outcome will be analysed using a logistic regression model. Randomised group, age, presence of comorbidity, duration of illness and vaccination status will be included as covariates. The adjusted odds ratios will be reported for each pairwise comparison along with their associated 95% confidence interval and P-value. In addition, the adjusted absolute risk differences will be reported.

##### 6.3 Handling missing data

Participants who withdraw or are lost to FU will be included in the primary analysis and censored at last contact date. Participants with complete missing data (e.g. no diaries or calls) for a given endpoint will not contribute data to the respective primary analysis. The proportion of participants contributing no data to the respective primary analysis is expected to be low. Various imputation strategies will be considered when the proportion of participants contributing no data exceeds 15%.

##### 6.4 Multiple comparisons and multiplicity

There will be no adjustment for multiplicity in the analysis of secondary outcomes.

##### 6.5 Model assumptions

For the analysis of the secondary outcomes, model diagnostics will be checked.

#### 7 Sensitivity analysis of co-primary outcomes using concurrent randomised analysis population

Upon conclusion of the study or intervention (and at interims as needed), a stand-alone sensitivity analysis of each co-primary analysis will be conducted for each completed intervention, in which each intervention is compared to Usual Care using the concurrent randomisation analysis population. These analyses will take the same form as the primary analysis models but may require modified priors/parameters for the temporal adjustment (carried out by the SAC and detailed in ADR and section 5.1 M-SAP). For some interventions (e.g. hydroxychloroquine), temporal adjustment may not be necessary for this sensitivity analysis.

#### 8 Subgroup estimates

Model-based estimated treatment group differences in median time to recovery will be provided for each of the covariate subgroups with 95% Bayesian credible intervals from the first co-primary analysis model. Similarly, model-based estimated differences in hospitalization rates will be provided for each of the covariate subgroups with 95% Bayesian credible intervals from the second co-primary analysis model (carried out by the SAC refer to section 5.1 M-SAP).

### 9 MODERATION ANALYSIS

Moderation analyses of the time to recovery and death or hospitalisation outcomes will be carried out. Time to recovery will be analysed using the model specified for the analysis of time to alleviation of symptoms (section 6.2.2.1) and death or hospitalisation will be carried out using the same model used for other secondary binary outcomes (logistic regression model with randomised group, age and presence of comorbidity, duration of illness and vaccination status included as covariates). In addition the models will include an interaction between treatment group and the subgroup of interest. The P-values for the interaction effects will be presented and forest plots presented to show the effects in each subgroup and overall. Unless specified in the intervention specific appendices, the following analyses will be undertaken.

- Age group (<65/≥65 years)
- Presence of comorbidity at baseline (yes/no)
- Duration of symptoms prior to randomisation (this will be assessed as both a continuous outcome and using a cut off of ≤7 days vs. >7 days)
- Severity of symptoms at baseline (at least one symptom rated major vs no symptoms rated major)
  Using an inhaled corticosteroid steroid (ICS) at randomisation or during 28 days of FU (Yes v No). Participant report will be taken as the primary source and only if missing will prescription of ICS in the two months prior to randomisation or during follow up from notes review or NHS Digital be considered.

### 10 SAFETY ANALYSIS

#### 10.1 ADVERSE EVENTS

Number and severity of serious adverse events (SAE) will be summarised across treatment arms using numbers and proportions.

Frequencies and percentages of adverse events (AE) will be reported. Details in the intervention specific appendices.

### 11 VALIDATION

The analysis of the primary outcome will be validated by a second statistician from the SAC. The final analysis of the secondary and safety outcomes will be validated by a Senior Trial Statistician or suitably qualified delegate from Oxford PC-CTU.

### 12 CHANGES TO THE PROTOCOL OR PREVIOUS VERSIONS OF SAP

The current protocol (V7.1) has an outcome of 'consumption of antibiotics'. Information regarding antibiotics has only been collected in the notes review and therefore relates to prescriptions of antibiotics rather than patient reported consumption.

Protocol version 7.1 states that Clinical data, and information from swab and blood tests, where available, will be used to classify participants according to aetiology. Blood test results have not been used to define the infected population for the intention to treat infected analyses.

### 13 APPENDICES

#### 13.1 APPENDIX I. SCHEDULE OF PROCEDURES

| **Procedures** | **Participant contacts** | | | | | | | |
|---|---|---|---|---|---|---|---|---|
| | **Visit timing**<br>**Day 0** | **Day 0** | **Day 0** | **Day 0** | **Daily Day 1-28 incl** | **Day 28-12 months (monthly contact)** | **Day 29-12mths** | **Up to 10 years** |
| | **Screening completed by participant online/phone** | **Eligibility completed by participant online/phone** | **Baseline completed by participant online/phone** | **Eligibility completed by Clinician online/phone** | **Symptom Diaries completed by participant online/phone** | **Contacted by study team if consent provided** | **Retrospective data collection by study team** | **By data extraction from clinical records** |
| Informed consent | X | X | X | X | X | | | |
| Demographics | X | X | X | | | | X | |
| Medical history | X | X | X | X | | | X | |

| | | | | | | | |
|---|---|---|---|---|---|---|---|
| Swab as part of the RCGP RSC/PHE national surveillance programme | When available, preferably by self-swabbing at study entry | | | | | | |
| Concomitant medications | | X | | | | | X |
| Eligibility assessment | X | X | | | | | |
| Randomisation | | | | X | | | |
| Dispensing of trial drugs | | | | X | X | | |
| Questionnaire | | | | | X | X | |
| WHO 5 Well Being Index | X | | | | Day 14 and day 28 | X | |
| Telephone interview (for subset of | | | | | X | | |

| | | | | | | | | |
|---|---|---|---|---|---|---|---|---|
| patient participants) | | | | | | | | |
| Compliance | | | | | X | | | |
| Adverse event assessments | | | | | X* | | X | |
| Optional SARS-<br><br>CoV-2 blood test as part of the RCGP RSC/PHE national surveillance programme | | | | | | | X | |
| Evidence of sequalae and health care utilisation | | | | | | X | | X |

#### 13.2 APPENDIX II. FLOW DIAGRAM OF TRIAL PARTICIPANTS

Figure 1 Participant flow diagram

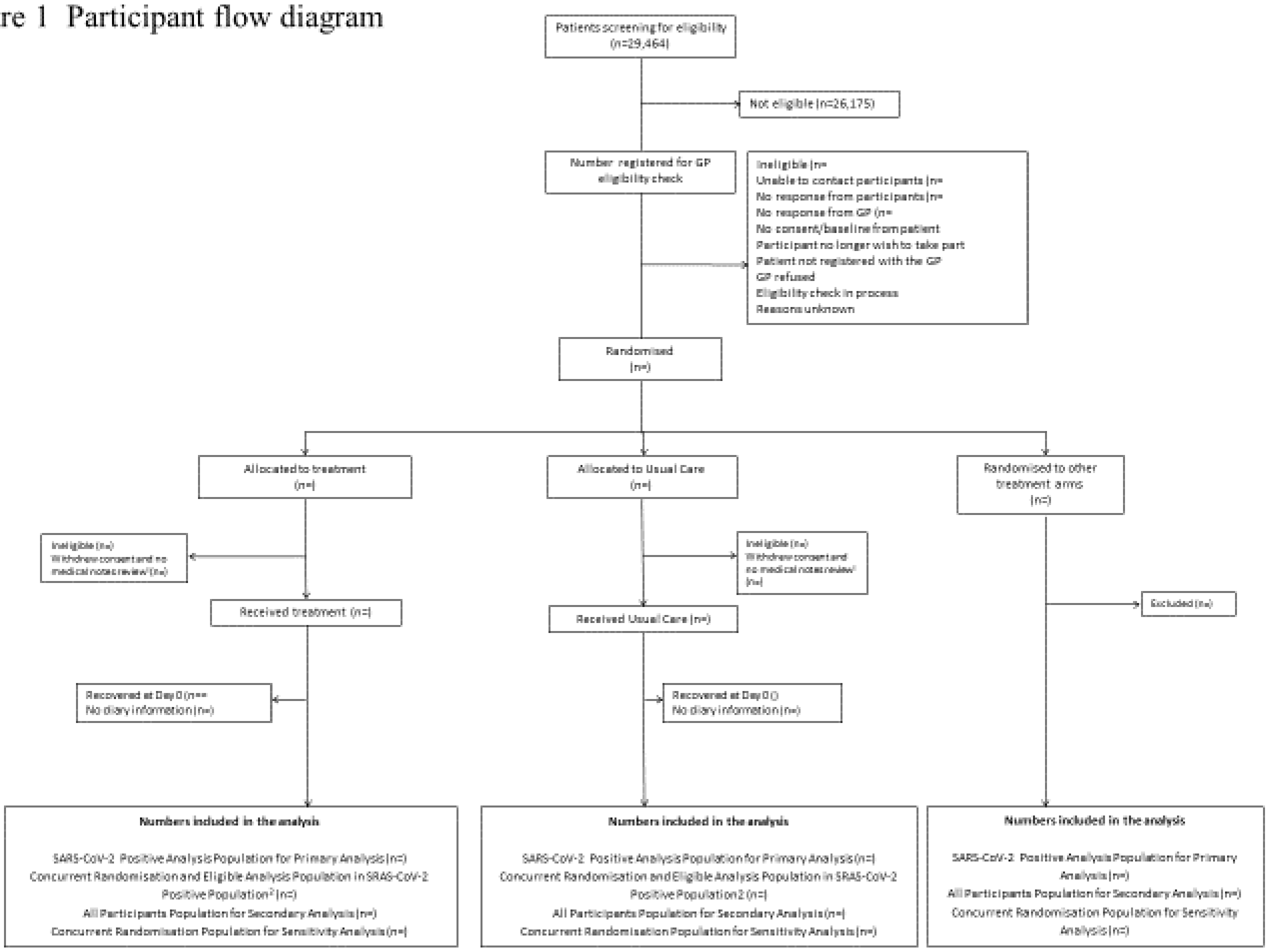

[1] Participants provided no diary information.
[2] Secondary outcomes only

#### 13.3 APPENDIX III: Plans for analysis of Hydroxychloroquine

##### Population of Analysis

The primary analysis population for the co-primary outcomes is defined as all randomised participants for whom data were available with participants analysed according to the groups they were randomly allocated to, at the time of the first trial interim analysis regardless of deviation from protocol and irrespective of their COVID-19 status. Although, participants randomised to other treatment arms were included in this interim analysis, only results of the hydroxychloroquine intervention will be reported.

Sensitivity and secondary analyses will be based on all participants randomised up to the point when hydroxychloroquine stopped recruitment.

##### Descriptive statistics

Baseline characteristics of participants will be summarised both overall and by randomised group, for all participants up to the point when randomisation to hydroxychloroquine was stopped. Data will be summarised as described in the M-SAP.

##### Primary Outcome Analysis

Analysis of the co-primary outcomes, i.e. time-to-recovery from randomisation and hospitalisation or death at 28 days from randomisation, are detailed in the Adaptive Design Report. Because Hydroxychloroquine was stopped for external reasons not related to the trial design, the primary analysis for Hydroxychloroquine is based on all participants enrolled up to 30th November 2020 . This date coincides with the date that randomisation was stopped to Azithromycin per interim futility criteria, and is meant to optimise precision of the Hydroxychloroquine analysis. Data from participants allocated to the hydroxychloroquine arm, and all control participants' data available at the time of the first interim analysis will be used, so there will be more control participants in the primary analysis than participants allocated to the hydroxychloroquine arm contributing to the primary analyses. However, due to the general lack of availability of swab tests during the Hydroxychloroquine testing phase, secondary analyses on the SARS-CoV-2 positive population will not be performed.

| Analysis | Date last participant randomised to be included in the analysis | Duration Follow-up | Date of data transfer/lock |
|---|---|---|---|
| **HCQ** | 30th Nov 2020 | 28 days | 11th Jan 2021 |

##### Specific sensitivity and secondary analysis of Primary Outcome

Bayesian analysis B2 and B3 (Section 5.1 M-SAP) will be requested.

##### Concurrent Randomisation analysis population

| | Date first participant randomised | Date last participant randomised | Duration Follow-up | Date of data transfer/lock |
|---|---|---|---|---|
| HCQ | 2nd April 2020 | 22nd May 2020 | 28 days | 11th Jan 2021 |

#### SECONDARY OUTCOME ANALYSIS

Secondary outcomes will be analysed as described in the M-SAP. All secondary outcomes will be analysed as detailed in the M-SAP.

##### Analysis of Safety Data

All serious adverse events will be reported as described in the M-SAP.

#### 13.4 APPENDIX IV: PLANS FOR ANALYSIS OF AZITHROMYCIN

##### POPULATION OF ANALYSIS

The primary analysis population for the co-primary outcomes is defined as all randomised participants for whom data were available with participants analysed according to the groups they were randomly allocated to, at the time of the interim analysis regardless of deviation from protocol and irrespective of their COVID-19 status. Although, participants randomised to other treatment arms were included in this interim analysis, only results of the azithromycin intervention will be reported.

Sensitivity and secondary analyses will be based on all participants randomised between the first and last dates a patient was randomised to receive azithromycin. Only concurrent controls eligible for the azithromycin arm will be included.

##### DESCRIPTIVE STATISTICS

Baseline characteristics of participants will be summarised both overall and by randomised group, for all participants included in the primary analysis population. Data will be summarised as described in the M-SAP.

##### PRIMARY OUTCOME ANALYSIS

Analysis of the co-primary outcomes, i.e. time-to-recovery from randomisation and hospitalisation or death at 28 days from randomisation, are detailed in the Adaptive Design Report. All relevant data available at the time of the interim will be used in the primary analysis.

| **Analysis** | **Date last participant randomised to be included in the analysis** | **Duration Follow-up** | **Date of data transfer/lock** |
|---|---|---|---|
| **Azithromycin** | 30th Nov 2020 | 28 days | 11th Jan 2021 |

##### SPECIFIC SENSITIVITY AND SECONDARY ANALYSIS OF PRIMARY OUTCOME

Bayesian analysis B1, B2, B3, B4 (Section 5.1 M-SAP) will be requested.

##### CONCURRENT RANDOMISATION ANALYSIS POPULATION

| | **Date first participant randomised** | **Date last participant randomised** | **Duration Follow-up** | **Date of data transfer/lock** |
|---|---|---|---|---|
| **Azithromycin** | 23rd May 2020 | 30th Nov 2020 | 28 days | 11th Jan 2021 |

##### SECONDARY OUTCOME ANALYSIS

Secondary outcomes will be analysed as described in the M-SAP. All secondary outcomes will be analysed as detailed in the M-SAP.

##### ANALYSIS OF SAFETY DATA

All serious adverse events will be reported as described in the M-SAP.

#### 13.5 APPENDIX VI: PLANS FOR ANALYSIS OF DOXYCYCLINE

##### POPULATION OF ANALYSIS

The primary analysis population for the co-primary outcomes is defined as all randomised participants for whom data were available with participants analysed according to the groups they were randomly allocated to, at the time of the interim analysis regardless of deviation from protocol and irrespective of their COVID-19 status. Although, participants randomised to other treatment arms were included in this interim analysis, only results of the doxycycline intervention will be reported.

Sensitivity and secondary analyses will be based on all participants randomised between the first and last dates a patient was randomised to receive doxycycline. Only concurrent controls eligible for the doxycycline arm will be included.

##### DESCRIPTIVE STATISTICS

Baseline characteristics of participants will be summarised both overall and by randomised group, for all participants included in the primary analysis population. Data will be summarised as described in the M-SAP.

##### PRIMARY OUTCOME ANALYSIS

Analysis of the co-primary outcomes, i.e. time-to-recovery from randomisation and hospitalisation or death at 28 days from randomisation, are detailed in the Adaptive Design Report. All relevant data available at the time of the interim will be used in the primary analysis.

| Analysis | Date last participant randomised to be included in the analysis | Duration Follow-up | Date of data transfer/lock |
|---|---|---|---|
| **Doxycycline** | 14th Dec 2020 | 28 days | 14th Jan 2021 |

##### SPECIFIC SENSITIVITY AND SECONDARY ANALYSIS OF PRIMARY OUTCOME

Bayesian analysis B1, B2, B3, B4 (Section 5.1 M-SAP) will be requested.

##### CONCURRENT RANDOMISATION ANALYSIS POPULATION

| Analysis | Date first participant randomised | Date last participant randomised | Duration Follow-up | Date of data transfer/lock |
|---|---|---|---|---|
| **Doxycycline** | 24th July 2020 | 14th Dec 2020 | 28 days | 14th Jan 2021 |

##### SECONDARY OUTCOME ANALYSIS

Secondary outcomes will be analysed as described in the M-SAP. All secondary outcomes will be analysed as detailed in the M-SAP.

##### ANALYSIS OF SAFETY DATA

All serious adverse events will be reported as described in the M-SAP.

#### 13.6 APPENDIX VI: PLANS FOR ANALYSIS OF INHALED CORTICOSTEROID BUDESONIDE

##### POPULATION OF ANALYSIS

The primary analysis population for the co-primary outcomes is defined as all randomised participants with a positive COVID-19 test result for whom data were available with participants analysed according to the groups they were randomly allocated to, at the time of the interim analysis regardless of deviation from protocol. Although, participants randomised to other treatment arms were included in this interim analysis, only results of the inhaled corticosteroid intervention will be reported.

Sensitivity and secondary analyses will be based on all participants randomised between the first and last dates a patient was randomised to receive inhaled corticosteroid. Only concurrent controls eligible for the inhaled corticosteroid arm will be included.

##### DESCRIPTIVE STATISTICS

Baseline characteristics of participants will be summarised both overall and by randomised group, for all participants included in the primary analysis population. Data will be summarised as described in the M-SAP.

##### PRIMARY OUTCOME ANALYSIS

Analysis of the co-primary outcomes, i.e. time-to-recovery from randomisation and hospitalisation or death at 28 days from randomisation, are detailed in the Adaptive Design Report. All relevant data available at the time of the interim will be used in the primary analysis.

##### SPECIFIC SENSITIVITY AND SECONDARY ANALYSIS OF PRIMARY OUTCOME

Bayesian analysis B1, B2, B3, B4(Section 5.1 M-SAP) will be requested.
Sensitivity analysis of co-primary outcomes using the concurrent randomised and eligible analysis population with a positive COVID-19 test result. Time to recovery will be analysed using Cox proportional hazards model, adjusting for randomised group, age, presence of comorbidity at baseline, duration of illness prior to randomisation and vaccination status. Death or hospitalisation will be carried out using a logistic regression model with randomised group, age and presence of comorbidity, duration of illness and vaccination status included as covariates. In addition, these models will be repeated on the concurrent randomised analysis population with a positive COVID-19 test result with additional adjustment for asthma/COPD at baseline as a surrogate for taking inhaled corticosteroid at baseline.

##### SECONDARY OUTCOME ANALYSIS

Secondary outcomes will be analysed as described in the M-SAP. All secondary outcomes will be analysed as detailed in the M-SAP.

##### ANALYSIS OF SAFETY DATA

All serious adverse events will be reported as described in the M-SAP.

#### 13.7 Appendix VII: plans for analysis of colchicine

##### Population of Analysis

The primary analysis population for the co-primary outcomes is defined as all randomised participants with a positive COVID-19 test result for whom data were available with participants analysed according to the groups they were randomly allocated to, at the time of the interim analysis regardless of deviation from protocol. Although, participants randomised to other treatment arms were included in this interim analysis, only results of the colchicine intervention will be reported.

##### Descriptive statistics

Baseline characteristics of participants will be summarised both overall and by randomised group, for all participants included in the primary analysis population. Data will be summarised as described in the M-SAP.

##### Primary Outcome Analysis

Analysis of the co-primary outcomes, i.e. time-to-recovery from randomisation and hospitalisation or death at 28 days from randomisation, are detailed in the Adaptive Design Report. All relevant data available at the time of the interim will be used in the primary analysis.

##### Specific sensitivity and secondary analysis of Primary Outcome

Bayesian analysis B1, B2, B3, B4 (Section 5.1 M-SAP) will be requested.

| **Date randomisation opened to colchicine** | **Date randomisation closed to colchicine** | **Duration Follow-up** | **Date of data transfer/lock** |
|---|---|---|---|
| 4th March 2021 | 26th May 2021 | 28 days | 5th July 2021 |

##### Secondary Outcome Analysis

Secondary outcomes will be analysed as described in the M-SAP. All secondary outcomes will be analysed as detailed in the M-SAP.

##### Analysis of Safety Data

All serious adverse events will be reported as described in the M-SAP. AEs are monitored daily whilst the participant is taking the drug. Events rated as a 'major problem' (except loss of smell and/or taste) will be assessed by a clinician for potential reporting as an SAE, for the 14 days of IMP treatment. Pregnancy in the study will be reported as an AE.

Duration of IMP treatment: 14 days
Duration of AE reporting: 14 days

##### Moderation analysis

The following moderation analyses or modification to subgroups will be carried out in addition to those specified in section 9.0

- Age (<50, 50-64, ≥65)
- Shortness of breath at baseline (no problem vs mild/moderate/major)

#### 13.8 APPENDIX VIII: PLANS FOR ANALYSIS OF FAVIPIRAVIR

##### Population of Analysis

The primary analysis population for the co-primary outcomes is defined as all randomised patients with a positive COVID-19 test result for whom data were available with participants analysed according to the groups they were randomly allocated to, at the time of the interim analysis regardless of deviation from protocol. Although, participants randomised to other treatment arms were included in this interim analysis, only results of the favipiravir intervention will be reported.

##### Descriptive statistics

Baseline characteristics of participants will be summarised both overall and by randomised group, for all participants included in the primary analysis population. Data will be summarised as described in the M-SAP.

##### Primary Outcome Analysis

Analysis of the co-primary outcomes, i.e. time-to-recovery from randomisation and hospitalisation or death at 28 days from randomisation, are detailed in the Adaptive Design Report. All relevant data available at the time of the interim will be used in the primary analysis.

##### Specific sensitivity and secondary analysis of Primary Outcome

Bayesian analysis B1, B2, B3, B4 (Section 5.1 M-SAP) will be requested.

| **Date randomisation opened to faviprivir** | **Date randomisation closed to favipirivir** | **Duration Follow-up** | **Date of data transfer/lock** |
|---|---|---|---|
| 8th April 2021 | | 28 days | |

##### Secondary Outcome Analysis

Secondary outcomes will be analysed as described in the M-SAP. All secondary outcomes will be analysed as detailed in the M-SAP. AEs will be reported by grade of severity.

##### Analysis of Safety Data

All serious adverse events will be reported as described in the M-SAP. AEs are monitored daily whilst the participant is taking the drug. Pre-defined AEs rated by the participant as 'moderate' and any symptoms reported as 'major' will be reported and assessed by a clinican for severity and causality.

Duration of treatment: 5 days
Duration of AE reporting: 5 days
Pre-defined AEs : Diarrhoea, nusea, headache, urinary tract infections, vomiting

#### 13.9 APPENDIX IX: PLANS FOR ANALYSIS OF IVERMECTIN

##### Population of Analysis

The primary analysis population for the co-primary outcomes is defined as all randomised patients with a positive COVID-19 test result for whom data were available with participants analysed according to the groups they were randomly allocated to, at the time of the interim analysis regardless of deviation from protocol. Although, participants randomised to other treatment arms were included in this interim analysis, only results of the Ivermectin intervention will be reported.

##### Descriptive statistics

Baseline characteristics of participants will be summarised both overall and by randomised group, for all participants included in the primary analysis population. Data will be summarised as described in the M-SAP.

##### Primary Outcome Analysis

Analysis of the co-primary outcomes, i.e. time-to-recovery from randomisation and hospitalisation or death at 28 days from randomisation, are detailed in the Adaptive Design Report. All relevant data available at the time of the interim will be used in the primary analysis.

##### Specific sensitivity and secondary analysis of Primary Outcome

Bayesian analysis B1, B2, B3, B4 (Section 5.1 M-SAP) will be requested.

| **Date randomisation opened to ivermectin** | **Date randomisation closed to ivermectin** | **Duration Follow-up** | **Date of data transfer/lock** |
|---|---|---|---|
| 23rd June 2021 | | 28 days | |

##### Secondary Outcome Analysis

Secondary outcomes will be analysed as described in the M-SAP. All secondary outcomes will be analysed as detailed in the M-SAP. AEs will be reported by grade of severity.

##### Analysis of Safety Data

All serious adverse events will be reported as described in the M-SAP. AEs are monitored daily whilst the participant is taking the drug. Pre-defined AEs (visual disturbances, confusion, seizures) rated by the participant as ‘moderate’ or ‘major’ and any symptom reported as ‘major’ will be reported and assessed by a clinican for severity and causality. Pregnancy will be reported as an AE.

Duration of treatment: 3 days
Duration of AE reporting: 14 days from initial drug administration
Pre defined AEs: visual disturbances, confusion, seizures.

##### Moderation analyses

The following moderation analyses or modification to subgroups will be carried out in addition to those specified in section 9.0

- Age (<50, 50-64, ≥65)
- Shortness of breath at baseline (no problem vs mild/moderate/major)

### APPENDIX 4

### PRINCIPLE
### Adaptive Design Report
### Version 4.1

Ben Saville, Ph.D.
Nick Berry, Ph.D.

May 15, 2021

### 1 Trial Overview

PRINCIPLE is an open, adaptive, platform trial to evaluate emerging treatments of persons with infection or suspected infection of the novel COVID-19 virus, with a focus on adults with existing comorbidities and/or older ages. The purpose of this document is to define the primary analysis and adaptive design of the PRINCIPLE trial. Complete details of all planned analyses are described in the Master Statistical Analysis Plan (M-SAP).

A “platform trial” is a trial in which multiple treatments for the same disease are tested simultaneously. The backbone of the trial is an adaptive clinical trial design. Pre-specified decision criteria allow for dropping a treatment for futility, declaring a treatment superior, or adding a new treatment to be tested. Because the process of dropping and adding treatments may be on-going for an indefinite period of time, platform trials may be better conceived of as a process rather than a singular clinical trial. In the context of the COVID-19 pandemic, the trial may continue as long as the pandemic persists.

The PRINCIPLE trial will begin as a 1:1 randomized trial of Hydroxychloroquine versus Usual Care and will have the capability to add additional interventions over time. The evaluation of any new interventions will be governed by the master protocol, including adaptive and decision criteria. In addition, the inclusion of any new interventions will require supplementary appendices to the protocol and M-SAP.

#### 1.1 Version Control

Multiple versions of this document (and trial design) have existed over time. Below we summarize key versions:

- Version 1.0, 09 May 2020:
  - Initial draft design report, with primary endpoint of hospitalization
- Version 2.0, 07 July 2020:
  - Second draft of design report, with primary endpoint changed to time to recovery
- Version 3.0, 09 September 2020:
  - First version of completed design report, with co-primary endpoints of time to recovery and hospitalization
- Version 3.1, 22 September 2020:
  - Minor changes to text; simulations and design unchanged
- Version 3.2, 23 October 2020:
  - Changes to futility rules
  - Update to accrual rates used in simulations (simulations incomplete)
- Version 3.3, 30 October 2020:
  - Updated simulations to reflect modifications to design implemented in version 3.2
- Version 3.4, 06 November 2020:
  - Minor changes to text (regarding interim communication channel); simulations and design unchanged
- Version 3.5, 08 March 2021:
  - Changes to the hospitalization analysis (including temporal adjustment)
  - Modified model covariates (COVID swab result)
  - Modified futility rules

- Version 4.0, 23 March 2021:
  - Changed primary analysis population to those with COVID-19 positive swab test
  - Clarified that adaptations (including RAR) are based on the primary analysis population (i.e. COVID-19 positive)
  - Changes to covariates used in both co-primary analyses (e.g. remove COVID swab)
  - Changed time intervals to 4 weeks (instead of 8 weeks) in the temporal adjustment of the hospitalization endpoint
- Version 4.1, 15 May 2021:
  - Clarified that the minimum numbers of patients for interims are based on the primary analysis population (Section 4.1)
  - Clarified that the timing of interim analyses are to be based on the primary analysis population (Section 4.1)
  - Removed text throughout the document indicating a superior arm will replace Usual Care
  - Modified action if superiority is achieved on recovery or on both endpoints (Section 4.1.1)
  - Clarified lack of futility rule on hospitalisation endpoint, and fixed a typo in the probability of a meaningful effect on hospitalisation (Section 4.1.2)
  - Modified RAR details if an intervention demonstrates superiority (Section 4.3.1)
  - Removed mentions of "Standard of Care" and replaced with Usual Care (e.g., Section 4.3.4)
  - Modified language describing accuracy of simulations (Section 5)
  - Added Sections 3.1.2 and 3.2.2 to describe the estimation of treatment benefit on the scales of time to recovery and hospitalization rate
  - Modified definition of vaccination covariate (Section 2.1)

### 2 Co-primary Hypotheses

The trial has two co-primary endpoints. The first co-primary endpoint is time to recovery from suspected COVID-19 infection within 28 days from randomization, where time to recovery is defined as the first instance that a participant reports feeling recovered. The second co-primary endpoint is hospital admission or death related to suspected COVID-19 infection within 28 days from randomization. The SAP defines three groups based on COVID-19 test status: 1) COVID-19 Positive; 2) COVID-19 Negative; and 3) No swab result available. The primary analysis population includes only individuals who are COVID-19 positive. A secondary analysis will replicate the primary analyses on the overall population.

Unless otherwise specified in the intervention-specific appendices, the co-primary outcomes will be analyzed using a "gate-keeping" strategy. For a given treatment, time to recovery will be analysed first, and if the first null hypothesis is rejected, the second co-primary hypothesis regarding hospitalization/death will be subsequently evaluated. This gate-keeping strategy preserves the overall Type I error of the primary endpoints without additional adjustments for multiple hypotheses. In addition, the gate-keeping structure reflects the clinical belief that an intervention is unlikely to demonstrate benefit on the hospitalization/death endpoint without first demonstrating benefit on the time to recovery endpoint.

#### 2.1 Time to Recovery

The first primary analysis is a Bayesian piecewise exponential of time to recovery regressed on intervention and baseline covariates (age, comorbidity, vaccination status) with temporal adjustment. For the purposes of modeling, age will be categorized as 1) at least 18 but less than 30 years old;

2) at least 30 but less than 50 years old; 3) at least 50 but less than 65 years old; or 4) at least 65 years old; comorbidity will indicate the presence or absence of comorbidities at baseline; and vaccination status will be categorized as "yes" vs. "other", where "yes" indicates at least one COVID-19 vaccination dose was given at least 2 weeks prior to randomization and "other" indicates either no vaccination at least 2 weeks prior to randomization or no vaccine information is available. Covariate categories may be collapsed for the purposes of modeling if insufficient sample size within a given category.

Let $\theta_j$ for $j > 0$ denote the log hazard ratio for time to recovery for persons on intervention j versus the Usual Care arm ($j = 0$), where $\theta_j > 0$ corresponds to faster recovery. Based on a Bayesian posterior distribution of $\theta_j$, the primary analysis for intervention $j$ will test the following hypothesis:

$$\begin{aligned} H_0 &: \theta_j \leq 0 \\ H_1 &: \theta_j > 0 \end{aligned} \tag{1}$$

If the Bayesian posterior probability of beneficial treatment effect ($\psi_{1j}$ in equation 2) is greater than or equal to 0.99,

$$\psi_{1j} = \Pr(\theta_j > 0) \tag{2}$$

the null hypothesis will be rejected and the intervention will be deemed superior to Usual Care in the primary analysis population. The decision criteria controls the one-sided Type I error of each intervention at approximately 0.025.

#### 2.2 Hospital Admission or Death

The second co-primary analysis is a Bayesian generalized linear model of the primary outcome regressed on treatment and baseline covariates (age, comorbidity, vaccination status) with temporal adjustment. For the purposes of modeling, covariate categories will be the same as the time to recovery analysis, where covariate categories may be collapsed if insufficient sample size within a given category.

Let $p_j$ denotes the probability of hospitalization/death for persons in treatment group j, where j=0 denotes the Usual Care arm and $j = 1$ denotes an intervention arm. A Bayesian posterior distribution will be derived for the estimated difference in probability of hospitalization/death between treatment groups. Let $\delta_j$ denote the log odds ratio of hospitalization/death comparing intervention $j$ to Usual Care. The primary analysis for intervention $j$ will test the following hypothesis:

$$\begin{aligned} H_0 &: \delta_j \geq 0 \\ H_1 &: \delta_j < 0 \end{aligned} \tag{3}$$

If the Bayesian posterior probability of beneficial treatment effect $\psi_{2j}$ is greater than or equal to 0.975, with $\psi_{2j}$ given in (4),

$$\psi_{2j} = \Pr(\delta_j < 0) \tag{4}$$

the null hypothesis will be rejected and the intervention will be deemed superior to Usual Care with respect to Hospitalization/Death in the primary analysis population. Note the decision criterion is lower than the recovery endpoint decision criterion (due to the gate-keeping structure), and controls the one-sided Type I error of each intervention at approximately 0.025 for plausible scenarios.

### 3 Co-primary Analyses

#### 3.1 Time to Recovery Analysis Model

Let $T_{ij}$ be the progression event time (in days) for time to recovery for the $i$th subject in arm $j$, where an event represents a positive outcome for the subject. We model the event times as piecewise exponential:

$$T_{ij} \sim PE(\lambda_{ij1}, \lambda_{ij2}, \lambda_{ij3}, \lambda_{ij4}), \tag{5}$$

where $\mathbf{\Lambda} = (\lambda_{ij1}, \lambda_{ij2}, \lambda_{ij3}, \lambda_{ij4})$ represents the set of hazard rates (events per day per subject) within each time segment $s$ corresponding to subject $i$. The hazard rate for subject $i$ on treatment $j$ within time segment $s$ is given by Equation (6),

$$\lambda_{ijs} = \exp(\gamma_s + \theta_j + \boldsymbol{x}_i' \boldsymbol{\beta} + \eta_{t(i)}), \tag{6}$$

where $\gamma_s$ is the log hazard rate corresponding to time segment $s$, $\theta_j$ is the log hazard ratio for treatment relative to Usual Care, $\boldsymbol{x}_i$ is a vector of baseline covariates (age, comorbidity, vaccination status), $\boldsymbol{\beta}$ the corresponding vector of covariate parameters, and $\eta_{(i)}$ is a time drift parameter described in Section (3.1.1).

The log hazard rate corresponding to time segment $s$ is given by equation (7),

$$\gamma_s = \alpha_1 + \alpha_2 I(s = 2) + \alpha_3 I(s = 3) + \alpha_4 I(s = 4) \tag{7}$$

where I() is an indicator function equal to 1 if the condition is satisfied and 0 otherwise, $\alpha_1$ is the log hazard rate corresponding to time segment one (0-7 days), $\alpha_2$ is the increment in log hazards for time segment two (8-14 days), $\alpha_3$ is the increment in log hazards for time segment three (15-21 days), and $\alpha_4$ is the increment in log hazards for time segment four (> 21 days). The prior distributions corresponding to the time segment parameters are given by equation (8),

$$\begin{aligned} \alpha_1 &\sim N(-2.3, 0.3^2) \\ \alpha_s &\sim N(0, 0.3^2) \text{ for } s > 1 \end{aligned} \tag{8}$$

where $\alpha_1$ is centered at a hazard rate of 0.10, with increments in the log hazards centered at 0 for subsequent time segments, all with weakly informative variance.

For a time to recovery endpoint, treatment hazard ratios greater than one indicate treatment benefit. The log hazard ratio for treatment has the weak informative prior

$$\theta_j \sim N(0, 0.3^2), \tag{9}$$

and is assumed to be constant over time. The weak informative prior for the log hazard ratio places the prior mass of the HR between 0.5 and 2.0, which in line with clinical expectations for potential therapies, and also will be quickly overwhelmed with accruing data. A similar prior distribution is used for the covariate parameters given by equation (10),

$$\beta_k \sim N(0, 0.3^2) \tag{10}$$

where the covariate parameters are also assumed to be constant over time. Subjects with values of time to recovery equal to 0 days will be excluded from the analysis. Subjects with values for time to recovery greater than 28 days will be censored at 28 days. Subjects with incomplete diary data will be censored at the last date of entry. However, subjects who are censored due to death before first recovery will be censored at 28 days.

##### 3.1.1 Adjustment for Temporal Changes

In the COVID-19 pandemic setting, there is a risk of temporal changes in the time to recovery, for example due to changes in virus prevalence, strain severity, or clinical care (perhaps due to results released from this trial). Hence treatment arm comparisons based on non-concurrently randomized participants (e.g. due to staggered entry) can provide misleading results if temporal changes are not appropriately accounted for. Because the primary analysis is based on the comparison of the intervention versus a potentially changing Usual Care (see exception for combination arms in Section 4.3.5), adjustment for potential temporal changes is essential to the primary analysis. The temporal adjustment is made possible due to the overlapping enrollment across the respective treatment arms.

Bayesian methods are used to smooth the estimates across time intervals over the course of the trial. More specifically, we define 2-week time intervals from the start of the trial, and count backwards from the interval at the time of the data cut ($t = 1$) to the start of the trial, i.e. "walking backwards" in time. Let $\eta_{t(i)}$ denote the time offset parameter corresponding to the time of randomization for subject $i$. We set $\eta_1 = \eta_2 = 0$ corresponding to $t = 1$ and $t = 2$, i.e. for subjects randomized in the most recent two intervals. For every previous 2-week interval, the time parameter is modeled with the following Bayesian second order normal dynamic linear model (NDLM):

$$\eta_t \sim N(2\eta_{t-1} - \eta_{t-2}, \tau_\eta^2), \text{ for } t \geq 3 \tag{11}$$

The drift parameter $\tau_\eta^2$ specifies the degree of smoothing over the time intervals, for which the following hyperprior distribution is used,

$$\tau_\eta^2 \sim \text{InvGamma}(a = 2, b = 0.0125) \tag{12}$$

and the inverse gamma (InvGamma) distribution is given by:

$$p(x) = \frac{b^a e^{-b/x}}{x^{a+1}\Gamma(a)} \tag{13}$$

This hyperprior distribution for $\tau_\eta^2$ is equivalent to 4 intervals of data with $\tau_\eta$ centered at 0.079, i.e. a prior that time interval effects have small changes from one interval to the next. The individual time interval effects will be heavily shaped by the data from patients within the intervals. If there are insufficient numbers of patients within a given interval, time intervals will be collapsed as necessary to ensure stable model estimates.

Although the temporal adjustment in the primary analysis model is pre-specified, the unblinded statistical analysis committee may make adjustments to the temporal components as needed (e.g. change in prior distributions or length of intervals) to provide stable estimates of model parameters.

##### 3.1.2 Estimated Benefit on Median Time to Recovery

The primary analysis model provides an estimated hazard ratio that quantifies the benefit for each intervention relative to Usual Care. To better understand the clinical meaning of the estimated hazard ratio, the model-based estimated hazard ratio will be used to estimate an average benefit for each intervention versus Usual Care on median time to recovery. This is done for each intervention by taking a bootstrap sample with replacement of sample size $N_j$ from Usual Care participants among concurrent and eligible randomizations, and estimating the median time to recovery for the bootstrap sample (e.g. via Kaplan-Meier methods). This process is repeated $K$ times to create a total of $K$ estimates of median time to recovery for Usual Care (concurrent and eligible for given intervention), where $K$ is the number of MCMC draws used to fit the Bayesian primary analysis model. For each MCMC sample $k$ ($k = 1, \ldots, K$) of the Bayesian posterior of the hazard ratio, we apply the sampled hazard ratio to the estimated median time to recovery in Usual Care for bootstrap $k$ (e.g. under the assumption of exponential time to recovery), which produces a distribution for treatment benefit on median time to recovery for each intervention versus Usual Care. We summarize the estimated benefit with the median of this distribution, along with the corresponding 95% credible interval from the 2.5% and 97.5% quantiles.

#### 3.2 Hospitalization/Death Analysis Model

We define the second co-primary Bayesian analysis model for hospital admission or death related to suspected COVID-19. Among participants who have the opportunity to complete 28 days of follow-up, let $Y_{ij}$ be the primary outcome measured at 28 days for subject $i$ on arm $j$, with $j = 0$ denoting the Usual Care (control) and $j$ denotes an intervention arm. We model the primary outcome as

$$\begin{aligned} \text{logit}(p_{ij}) &= \gamma_0 + \delta_j z_{ij} + \boldsymbol{x}'_{ij}\boldsymbol{\beta} + \eta_{t(i)} \\ Y_{ij} &\sim \text{Bernoulli}(p_{ij}) \end{aligned} \tag{14}$$

where $p_{ij}$ is the probability of hospital admission/death for patient $i$ on arm $j$; $\gamma_0$ is the log odds of hospitalization for the Usual Care arm (at reference levels of covariates); $z_j$ is an indicator variable equal to 1 if subject $i$ is randomized to intervention $j$ (for $j > 0$) and 0 otherwise; $\delta_j$ is the log odds ratio of hospitalization/death comparing intervention $j$ to Usual Care; $\boldsymbol{x}_{ij}$ is a vector of baseline covariates (age, comorbidity, vaccination status) specific to subject $i$ and arm $j$; $\boldsymbol{\beta}$ is a vector of corresponding covariate log odds ratios; and $\eta_{(i)}$ is a time drift parameter described in Section (3.2.1). Note we use similar notation for some parameters from the first primary endpoint $(\boldsymbol{\beta}, \eta)$, but this is for notational convenience only, as these parameters do not represent the same population parameters across endpoints. The priors are given by:

$$\begin{aligned} \gamma_0 &\sim N(0, 2^2) \\ \delta_j &\sim N(0, 2^2) \text{ for } j \geq 1 \\ \beta_k &\sim N(0, 2^2) \text{ for } k = 1, 2, \ldots \end{aligned} \tag{15}$$

which are non-informative (disperse) prior distributions on the logit scale, and are expected to be overwhelmed with increasing number of events. If the number of events are insufficient to provide stable model estimates, the primary analysis model may be modified by removing or collapsing the baseline covariates in the model.

##### 3.2.1 Adjustment for Temporal Changes

Similar to the time to recovery endpoint, Bayesian methods are used for the hospitalization endpoint to smooth the estimates across time intervals over the course of the trial. More specifically, we define 4-week time intervals from the start of the trial, and count backwards from the interval at the time of the data cut ($t = 1$) to the start of the trial, i.e. “walking backwards” in time. Let $\eta_{t(i)}$ denote the time offset parameter corresponding to the time of randomization for subject $i$. We set $\eta_1 = 0$ for subjects randomized in the most recent interval. For every previous 4-week interval, the time parameter is modeled with the following Bayesian second order normal dynamic linear model (NDLM):

$$\begin{aligned} \eta_2 &\sim N(0, \tau_\eta^2) \\ \eta_t &\sim N(2\eta_{t-1} - \eta_{t-2}, \tau_\eta^2), \text{ for } t \geq 3 \end{aligned} \tag{16}$$

The drift parameter $\tau_\eta^2$ specifies the degree of smoothing over the time intervals, for which the following hyperprior distribution is used,

$$\tau_\eta^2 \sim \text{InvGamma}(a = 2, b = 0.1) \tag{17}$$

This hyperprior distribution for $\tau_\eta^2$ is equivalent to 4 intervals of data with $\tau_\eta$ centered at 0.22, i.e. a prior that time interval effects have small changes from one interval to the next, after accounting for changes in baseline covariates. The individual time interval effects will be heavily shaped by the data from patients within the intervals.

Although the temporal adjustment in the primary analysis model is pre-specified, the unblinded statistical analysis committee may make adjustments to the temporal components as needed (e.g. change in prior distributions or length of intervals) to provide stable estimates of model parameters.

For example, if there are insufficient numbers of hospitalizations within a given interval, time intervals may be collapsed as necessary to ensure stable model estimates. At the beginning of the trial, the proportion of COVID positive participants and hospitalization rates are both relatively low. Hence, the SAC may choose to combine the last two 4-week intervals (measured backwards from time of analysis) into a single 16-week time interval. Alternatively, the SAC may choose to maintain 4-week intervals, but modify the prior distributions for the last two intervals to induce higher correlation between the adjacent 4-week intervals. The goal is to provide a reasonable model of the temporal drift with respect to hospitalization/death rates.

###### 3.2.2 Estimated Benefit on Probability of Hospitalization

The primary analysis model provides an estimated odds ratio that quantifies the benefit for each intervention relative to Usual Care. To better understand the clinical meaning of the estimated odds ratio, the model-based estimated odds ratio will be used to estimate an average benefit for each intervention versus Usual Care on the probability of hospitalization. This is done for each intervention $j$ by taking a bootstrap sample with replacement of sample size $N_j$ from Usual Care participants among concurrent and eligible randomizations, and calculating the proportion of participants hospitalized in the bootstrap sample. This process is repeated $K$ times to create a total of $K$ estimated proportions for Usual Care (concurrent and eligible for a given intervention), where $K$ is the number of MCMC draws used to fit the Bayesian primary analysis model. For each MCMC sample $k$ ($k = 1, \ldots, K$) of the Bayesian posterior of the odds ratio, we apply the sampled odds ratio to the estimated hospitalization proportion in Usual Care for bootstrap $k$ (via the logit), which produces a distribution for treatment benefit on the probability of hospitalization for each intervention versus Usual Care. We summarize the estimated benefit with the median of this distribution, along with the corresponding 95% credible interval from the 2.5% and 97.5% quantiles.

##### 3.3 Sensitivity Analyses

Upon conclusion of the study or intervention (and at interims as needed), the following sensitivity analyses are planned with respect to the primary outcome

- A sensitivity analysis will be conducted for each completed intervention, in which each intervention is compared to Usual Care using only concurrent randomizations in the primary analysis population. These analyses will take the same form as the primary analysis models but may require modified priors/parameters for the temporal adjustment. This analysis will include participants randomized to all regimens that are being concurrently randomized. For some interventions (e.g. hydroxychloroquine), temporal adjustment may not be necessary for the sensitivity analysis.
- Analyses evaluating the impact of missing data will be conducted as outlined in the M-SAP.

##### 3.4 Secondary Analyses

Upon completion of each intervention (e.g. futility), the following secondary analyses may be conducted (details provided in M-SAP):

- The co-primary outcomes analyses will be replicated for the overall population, i.e. comparing time to recovery and hospitalization/death rates between treatment groups, among individuals regardless of test status. These models will be identical to the primary analysis models, but will include an additional baseline covariate and corresponding parameters for COVID-19 status (negative, positive, unknown).
- Other secondary analyses listed in the M-SAP

Each of the hypotheses for the overall population will be conducted at the same Bayesian posterior thresholds (0.99 for time to recovery and 0.975 for hospitalization) as the co-primary outcomes analyses using the gate-keeping structure. In other words, we first evaluate time to recovery in the overall population, and if significant we will evaluate the hospitalization/death endpoint in the overall population. Because other interventions may still be accruing information and using Usual Care subjects in their respective analyses, caution should be used in reporting and/or publishing results of an intervention, and will be limited to the analyses pre-specified in the M-SAP. Further guidance is provided in the PRINCIPLE Data Sharing/Access Policy.

### 4 Adaptive Design

The pre-specified design will allow adaptations to the trial based on the observed data. These adaptations include the declaration of superiority or futility of an arm at an interim analysis, the addition or removal of treatment arms, and changes in the randomization probabilities. All adaptations are based on the primary analysis population, and will occur at a given interim analysis if pre-specified conditions are satisfied.

#### 4.1 Interim Analyses for Superiority & Futility

The timing of the first interim analysis will be determined by the trial management group (TMG) and statistical analysis committee (SAC), and will include logistical and operational considerations. Subsequent interim analyses will be conducted on a monthly basis provided the accrual rate is is approximately 25 or less participants per week in the primary analysis population. If accrual increases to approximately 50 participants per week, interims may be conducted every 2 weeks. If accrual increases above approximately 100 participants per week, interims may be conducted weekly. The TMG and SAC will monitor accrual rates and will have flexibility to adapt the frequency of the interim analysis to the observed accrual rate, with the goal of interim analyses occurring approximately every 100 completed observations in the primary analysis population. At each interim analysis, all enrolled intervention arms will be evaluated for superiority using the Bayesian interim primary analysis, provided that the intervention arm has at least 100 randomized participants with the opportunity to complete 28-days of follow-up for the primary endpoint in the primary analysis population. In addition, at each interim analysis all enrolled intervention arms will be evaluated for futility using the Bayesian interim primary analysis, provided that the intervention arm has at least 75 randomized participants with the opportunity to complete 28-days of follow-up for the primary endpoint in the primary analysis population.

##### 4.1.1 Interim Superiority

At an interim analysis, if the Bayesian posterior probability of superiority of a given intervention over Usual Care is greater than or equal to 0.99 for the recovery endpoint, and greater than or equal to 0.975 for the hospitalization endpoint, superiority versus Usual Care will be declared on both endpoints, and randomisation will be stopped for that intervention. The primary analyses may be updated after observing full 28-day follow-up on all participants randomized to the given intervention, but the superiority conclusion will not change, regardless of any updated posterior probabilities.

At an interim analysis, if the Bayesian posterior probability of superiority exceeds the pre-specified threshold for the first co-primary endpoint (time to recovery) but not the second (hospitalization/death), superiority versus Usual Care will be declared on time to recovery, and randomisation will be stopped for that intervention. The second co-primary endpoint will be evaluated again for superiority after complete 28-days of follow-up on on all participants randomized to the given intervention. The primary analysis for time to recovery may be updated after observing full 28-day follow-up on all participants randomized to the given intervention, but the superiority conclusion will not change, regardless of any updated posterior probabilities.

In either of the above scenarios, if the NHS decides to adopt the superior intervention into national recommended treatment guidelines, the trial will continue randomising to the Usual Care, with the understanding that Usual Care arm may change with any modified national guidelines. Additional information may be collected corresponding to these changes on the Usual Care of trial participants. The primary analysis of each intervention arm will continue to be a comparison versus the Usual Care arm.

###### 4.1.2 Interim Futility

We define a futility rule based on the the Bayesian posterior probability of the hazard ratio from model (6). Let $\omega_{1j}$ be the model-based Bayesian posterior probability of a clinically meaningful treatment effect of the recovery endpoint, defined as the probability that the hazard ratio $e^{\theta}_{j}$ is at least 1.2 (equation 18).

$$\omega_{1j} = \Pr(e^{\theta_j} \geq 1.2) \tag{18}$$

A hazard ratio of 1.2 corresponds to a 1.5 day benefit on median time to recovery, assuming an exponential distribution with median time to recovery of 9 days on the Usual Care Arm. If the probability of a meaningful effect on recovery, $\omega_{1j}$, is less than or equal to 0.01, intervention $j$ will be dropped from the study for futility. If there are no other intervention arms available, the trial will be suspended; otherwise accrual continues to the remaining treatment arms.

In addition, a Bayesian posterior probability of a clinically meaningful treatment effect will be calculated for the second co-primary endpoint (hospitalization) using model (14). This probability, $\omega_{2j}$, is defined as the probability that the odds ratio of hospitalization/death for intervention $j$ relative to Usual Care is smaller than or equal to 0.80 (equation 19).

$$\omega_{2j} = \Pr(e^{\delta_j} \leq 0.80) \tag{19}$$

An odds ratio of 0.80 corresponds to an absolute decrease in the probability of hospitalization/death of 0.01 (i.e. 1 percentage point decrease), assuming a hospitalization rate of 0.05 in the Usual Care arm. There is no futility rule associated with the quantity $\omega_{2j}$; rather it is a simple summary to help describe the treatment effect for the hospitalisation endpoint.

The futility rule for time to recovery is non-binding, meaning that the SAC and DMSC may choose to override a futility trigger if they mutually agree it is in the best interests of the trial. For example, suppose there is only a single active intervention and the futility threshold is met, despite there being some evidence of a small treatment benefit. Rather than suspend the trial due to lack of active interventions, it may be desirable to continue randomization to the remaining intervention despite the futility threshold being met, with the TSC or TMG being informed that they should consider adding additional interventions as soon as possible.

###### 4.1.3 Maximum Sample Size per Arm

There is no cap on sample size per arm; hence interventions will remain active in the trial unless futility criteria are met or accrual is halted for budgetary or other considerations.

##### 4.2 Additional Treatment Arms

At any point in the trial (per recommendations of the Trial Steering Committee), the trial management group may elect to add an additional intervention to the trial. The evaluation of any new interventions will be governed by the master protocol, including adaptive and decision criteria. In addition, the inclusion of any new interventions will require supplementary appendices to the protocol and M-SAP. This includes updated simulations describing the revised operating characteristics of the trial with the additional arm.

#### 4.3 Allocation

At the start of the trial, randomization was fixed 1:1 to hydroxychloroquine (HCQ) versus Usual Care, with stratification by age (less than 65, greater than or equal to 65), and comorbidity (yes/no). Randomization was suspended to HCQ on May 23, 2020 per the Medicine and Health Care Products Regulatory Agency (MHRA), at which time the trial began randomizing 1:1 to Azithromycin versus Usual Care. Protocol Amendment 6 included the addition of the intervention Doxycycline, which is began enrolling around the end of July 2020. This adaptive design report was finalized prior to conducting the first interim analysis, which is expected to accomplish the following tasks:

1. Evaluate primary hypotheses for Hydroxychloroquine vs. Usual Care
   - Due to the sudden halt in accrual to hydroxychloroquine, there is only a single analysis evaluating hydroxychloroquine benefit versus Usual Care. Hence the posterior probability thresholds for hydroxychloroquine versus Usual Care equal 0.975 for BOTH of the co-primary endpoints. In addition, the analysis of hydroxychloroquine will include all subjects in the Usual Care arm up until the time of the interim analysis.
2. Evaluate early superiority/futility for all active interventions with a sufficient number of participants with the opportunity to complete 28 days of follow-up (per Section 4.1)
3. Determine subsequent randomization probabilities of active interventions

Subsequent interim analyses will evaluate superiority/futility of all active interventions with a sufficient number of participants with the opportunity to complete 28 days of follow-up (per Section 4.1), as well as determine subsequent randomization probabilities for all active interventions. An active intervention is defined as an intervention currently randomizing participants. Note Hydroxychloroquine will remain an inactive treatment regardless of the first interim results.

When there is more than one active intervention in the trial, the allocation probability to Usual Care will be set to $1/Z$, where $Z$ is the total number of active arms in the trial (e.g., $Z = 3$ for Usual Care and two active interventions).

##### 4.3.1 Response Adaptive Randomization

Prior to the first interim analysis, randomization will be set to $1/Z$ for each of the intervention arms. Response adaptive randomization (RAR) will be activated at the time of the first interim analysis if there at least two active interventions in the trial. When RAR is activated, the Usual Care arm will continue to receive a fixed allocation of $1/Z$. The remaining $(Z-1)/Z$ allocation probability will be divided among the intervention arms based on interim RAR probabilities. The purpose of RAR is to allocate more participants to the intervention arms with the best observed outcomes (relative to Usual Care).

For example, if there are 3 active arms (2 active intervention arms and Usual Care), the Usual Care allocation will be fixed at 1/3, and the remaining 2/3 allocation will be split among the two intervention arms via response adaptive randomization. The RAR probabilities ($q_j$) for the intervention arms will be proportional to the Bayesian posterior probability that a given intervention is superior to Usual Care ($\psi_{1j}$) with respect to the first co-primary endpoint (time to recovery) in the primary analysis population; see equation (2). The calculation of $\psi_{1j}$ is based on the MCMC posterior samples from the Bayesian interim primary analysis. The randomization probabilities $q_j$ of the intervention arms at a given interim are normalized so that they sum to $(Z-1)/Z$.

##### 4.3.2 Adding Interventions

When a new intervention is added, the RAR allocation for the new arm will be bounded between $1/(2Z)$ and $1/2$ for the first 4 weeks, after which this condition is removed. The remaining allocation will be given to the Usual Care (fixed $1/Z$) and the other interventions, proportional to probability of superiority on time to recovery versus Usual Care ($\psi_{1j}$).

###### 4.3.3 Comparing Interventions

Although the primary analysis compares each intervention versus Usual Care, the primary analysis model also enables pairwise comparisons between interventions. These pairwise comparisons will be conducted at each interim analysis. If an intervention is superior to another intervention per criterion of Bayesian posterior probability of superiority $\geq 0.99$ for recovery and $\geq 0.975$ for hospitalization, the inferior intervention will be dropped from the trial, regardless of how the intervention compares to Usual Care. Upon completion of enrollment for two interventions, the same criteria will be evaluated, i.e. a Bayesian posterior probability $\geq 0.99$ and $\geq 0.975$ for recovery and hospitalization, respectively, for one intervention versus another intervention will indicate pairwise superiority for each co-primary endpoint, with an identical gate-keeping sequential order of hypotheses for a given pairwise comparison.

###### 4.3.4 Usual Care

Usual Care will refer to the current state of national medical practice, in which physicians may choose and prescribe treatments as deemed appropriate. This may change during the course of the trial, and will be modeled in the primary analysis models using parameters for temporal drift.

###### 4.3.5 Combination Interventions

A combination intervention is defined as a combination of one of the existing interventions plus another treatment. The primary analysis for the combination arm will be as specified in Section (3), except when the stand-alone component of the combination has been determined to be superior to Usual Care for a given primary endpoint. In such settings, the primary analysis of the combination arm will be versus the standalone component arm. For example, if azithromycin is superior to Usual Care on the first co-primary endpoint (recovery) only, and a new combination enters the trial as a combination of azithromycin plus intervention X, the primary analysis of the combination will be to evaluate superiority versus azithromycin for the first co-primary endpoint, and superiority versus Usual Care for the second co-primary endpoint. If azithromycin is superior to Usual Care on both co-primary endpoints, the primary analysis of the combination will evaluate superiority versus azithromycin for both co-primary endpoints. Note the combination arm will use the same gate-keeping strategy for evaluating the two co-primary endpoints regardless of the comparison group.

###### 4.3.6 Arm Suspension

If the randomization probability $q_j < 0.05$ for any intervention $j$ at a given interim, the intervention $j$ is suspended from the trial, which is done by setting the randomization probability $q_j$ equal to 0 until the next interim analysis. If the next interim analysis no longer meets this threshold for a previously suspended dose, randomization to the intervention may then resume according to the algorithm. If at any point in the trial there only exist two active arms, randomization will be fixed at 1:1.

###### 4.3.7 Eligibility Exclusions by Intervention

A participant is eligible for randomization provided the participant meets the eligibility criteria for at least two arms, one of which includes Usual Care. If such a participant is ineligible for one of the interventions, the randomization probabilities will be re-normalized (i.e. probabilities sum to 1.0) among the remaining arms prior to randomization. Eligibility status will be recorded for each participant as (eligible, ineligible, or unknown) for each of the treatment arms. If the number of participants who are known to be eligible for one active intervention but ineligible for another active intervention becomes substantial, the primary analysis model may be modified to to include eligibility status as a covariate.

##### 4.4 Sample Size Justification

Given the open perpetual trial structure, the trial does not have a finite ending based on sample size. Rather, the trial will continue until either superiority or futility is claimed for an intervention, or until the pandemic expires in the population. We estimate that approximately 400 participants per arm (800 participants total if only a single intervention vs. usual care) will be required to provide 90% power for detecting a difference of 2 days in median recovery time in the primary analysis population. This calculation is based on the assumption of an exponential distribution for time to recovery with a median of 9 days in the Usual Care group, with some adjustments for missing data and multiple interim analyses. Alternative assumptions are explored in the virtual trial simulations

On average, we expect fewer participants to be required when there is a large treatment benefit or complete lack of benefit. For example, if the true benefit is a 3 day benefit in median time to recovery (6 days intervention vs. 9 days Usual Care), on average only 155 subjects per arm are required to provide sufficient power. The primary advantage of the adaptive design is the ability to adapt the sample size to the observed data, thus addressing the primary hypothesis as quickly and as efficiently as possible.

In addition, we estimate that approximately 1500 participants per arm (3000 participants total if only a single intervention vs. usual care) will be required to provide 90% power for detecting a 50% reduction in the relative risk of hospitalization/death in the primary analysis population. This calculation is based on the assumption of an underlying 5% combined hospitalization/death rate in the Usual Care arm, with an intervention lowering the hospitalization/death rate to 2.5%, with some adjustments for the multiple interim analyses. We expect fewer participants to be required to detect a 50% reduction if the event rate in the Usual Care arm is greater than 5%.

For a more accurate sample size justification (via simulation) of the adaptive platform trial design, we refer to Section 5.

#### 5 Simulated Operating Characteristics

Because of the adaptive platform trial structure, there exists no simple formula(s) to calculate power and Type I error (false positive rate). Hence, virtual trial simulations are used to fully characterize and quantify the power and Type I error of the design. The simulations include a comprehensive evaluation of trial performance across a wide range of assumptions (e.g. underlying distribution of outcome in control arm, treatment effect, accrual rates, etc.). This includes summaries regarding the number of subjects required to make a superiority or futility conclusions for each intervention.

Simulations below were conducted in October 2020 based on the available knowledge and trial design at the time, which is detailed in the Adaptive Design Report version 3.3 (30 October 2020). Although some modifications have been made to the trial design (e.g. futility rules, primary analysis population of COVID-19 positive participants, replacing Usual Care with superior treatment) to address a dynamic pandemic situation, the simulations can be viewed as an approximation to the performance of the adaptive design for evaluating future interventions in the primary analysis population.

##### 5.1 Simulation Scenarios

The time to recovery is simulated using an exponential distribution with rate parameter $\lambda$. Based on available knowledge, we expect $\lambda_0 = \log(2)/9$ for the Usual Care arm (corresponding to a median of 9 days), but also consider lower and higher values of $\lambda$ corresponding to a median of 7 and 11 days. We explore different scenarios for the hazard ratio reduction proportion hospitalized relative to control; these are given by HR=(1, 1.10, 1.20, 1.30). We also explore different scenarios for the treatment effects on hospitalization rates, where the aggregate hospitalization rate is assumed to be 0.025. This is done by imposing treatment effect sizes in absolute percentage decrease relative to the Usual Care arm equal to the null (0), small (1%), medium (2%), or large (3%) treatment effects while enforcing that the average hospitalization rate for the simulated arms (including Usual Care) equals 0.025.

At the time of running these simulations, the trial had been enrolling for approximately 6 months. Hence, our simulations include the observed accrual over the first 6 months (20-70 participants per week), followed by expected accrual thereafter based on a Poisson process. Given the uncertainty in accrual due to the state of the COVID pandemic, we consider maximum accrual rates of 30 ("expected") and 75 ("fast") subjects per week.

Given the large number of potential scenarios based on the factors listed above, simulations are conducted on a base set of scenarios with a median time to recovery of 9 days, hazard ratios of 1.0, 1.1, 1.2, and 1.3 for time to recovery, and hospitalization treatment effects of null (0%), small (1%), medium(2%), or large(3%). We then explore additional scenarios for a small subset of the base scenarios, where we 1) cross the null hospitalization scenarios with various treatment effects for the recovery endpoint; 2) cross different SOC median time to recovery rates with various treatment effects for the recovery endpoint; and 3) simulate faster accrual of 75 participants per week with bi-weekly interim analyses.

#### 5.2 Operating Characteristics Base Scenarios (Three Interventions)

This section explores the simulated trial operating characteristics (OCs) for the first three interventions (Hydroxychloroquine, azithromycin, doxycycline) versus Usual Care using a base set of scenarios. Given simulated virtual trials, the adaptive algorithm is applied at each interim update to conduct a virtual trial exactly as described. Various summaries are captured, including the average sample size, average trial duration (in weeks), the probability of stopping accrual for early futility or superiority, and the total probability of success, where success is defined as claiming superiority versus Usual Care. Note "power" is calculated as the proportion of simulated trials that claim superiority on the primary endpoint for scenarios with a treatment benefit for a given endpoint, and Type I error is calculated as the proportion of simulated trials that claim superiority on the primary endpoint for scenarios with no treatment benefit for a given endpoint. This is done for all combinations of accrual, treatment effect, and the assumed median time to recovery in the Usual Care arm.

### Principle OCs

Berry Consultants

10/26/2020

These operating characteristics are based on 1000 simulations per scenario across a variety of assumed treatment effects for experimental arms in the trial. The true number of patients randomized per day from April 1, 2020 to October 19, 2020. After this period is over, the simulations assume a 50 patient per week accrual rate. In absence of a definitive rule, these simulations conduct an interim every 28 days. In addition, the true day of enrollment stopping of HCQ is used as well as the days for opening enrollment to the Azithromycin and Doxycycline arms. As such, up to at least October 19, 2020 (day 199) of the trial, the enrollment rates to each arm in these simulations are expected to align closely with the true enrollment rates to the arms in the trial.

| Median TTR | Hazard Ratio | | | Power for TTR | | | | Hospitalization Rate Benefit (%) | | | Power for TTR & Hosp | | |
|---|---|---|---|---|---|---|---|---|---|---|---|---|---|
| SOC | HCQ | Azith | Doxy | SOC | HCQ | Azith | Doxy | HCQ | Azith | Doxy | HCQ | Azith | Doxy |
| 9 | 1.0 | 1.0 | 1.0 | 0.952 | 0.017 | 0.031 | 0.022 | 0 | 0 | 0 | 0.000 | 0.001 | 0.000 |
| 9 | 1.0 | 1.0 | 1.1 | 0.623 | 0.016 | 0.022 | 0.370 | 0 | 0 | 1 | 0.000 | 0.000 | 0.065 |
| 9 | 1.0 | 1.0 | 1.2 | 0.117 | 0.014 | 0.018 | 0.883 | 0 | 0 | 2 | 0.000 | 0.000 | 0.496 |
| 9 | 1.0 | 1.0 | 1.3 | 0.011 | 0.013 | 0.012 | 0.989 | 0 | 0 | 3 | 0.000 | 0.000 | 0.952 |
| 9 | 1.0 | 1.1 | 1.0 | 0.592 | 0.014 | 0.401 | 0.030 | 0 | 1 | 0 | 0.000 | 0.079 | 0.000 |
| 9 | 1.0 | 1.1 | 1.2 | 0.108 | 0.013 | 0.383 | 0.878 | 0 | 1 | 2 | 0.000 | 0.070 | 0.459 |
| 9 | 1.0 | 1.1 | 1.3 | 0.013 | 0.014 | 0.238 | 0.986 | 0 | 1 | 3 | 0.000 | 0.032 | 0.883 |
| 9 | 1.0 | 1.2 | 1.0 | 0.096 | 0.011 | 0.904 | 0.016 | 0 | 2 | 0 | 0.000 | 0.510 | 0.000 |
| 9 | 1.0 | 1.2 | 1.1 | 0.085 | 0.012 | 0.900 | 0.326 | 0 | 2 | 1 | 0.000 | 0.487 | 0.052 |
| 9 | 1.0 | 1.2 | 1.2 | 0.037 | 0.015 | 0.879 | 0.865 | 0 | 2 | 2 | 0.000 | 0.422 | 0.409 |
| 9 | 1.0 | 1.2 | 1.3 | 0.007 | 0.012 | 0.798 | 0.986 | 0 | 2 | 3 | 0.000 | 0.342 | 0.816 |
| 9 | 1.0 | 1.3 | 1.0 | 0.013 | 0.015 | 0.987 | 0.012 | 0 | 3 | 0 | 0.000 | 0.923 | 0.002 |
| 9 | 1.0 | 1.3 | 1.1 | 0.015 | 0.015 | 0.985 | 0.183 | 0 | 3 | 1 | 0.000 | 0.867 | 0.022 |
| 9 | 1.0 | 1.3 | 1.2 | 0.003 | 0.015 | 0.987 | 0.741 | 0 | 3 | 2 | 0.000 | 0.820 | 0.325 |
| 9 | 1.0 | 1.3 | 1.3 | 0.000 | 0.015 | 0.978 | 0.972 | 0 | 3 | 3 | 0.000 | 0.755 | 0.760 |
| 9 | 1.1 | 1.1 | 1.1 | 0.451 | 0.180 | 0.412 | 0.357 | 1 | 1 | 1 | 0.012 | 0.078 | 0.068 |
| 9 | 1.2 | 1.2 | 1.2 | 0.042 | 0.540 | 0.883 | 0.848 | 2 | 2 | 2 | 0.117 | 0.378 | 0.349 |
| 9 | 1.3 | 1.3 | 1.3 | 0.003 | 0.839 | 0.970 | 0.947 | 3 | 3 | 3 | 0.406 | 0.636 | 0.633 |

Decisions on Time to Recovery Benefit By Time

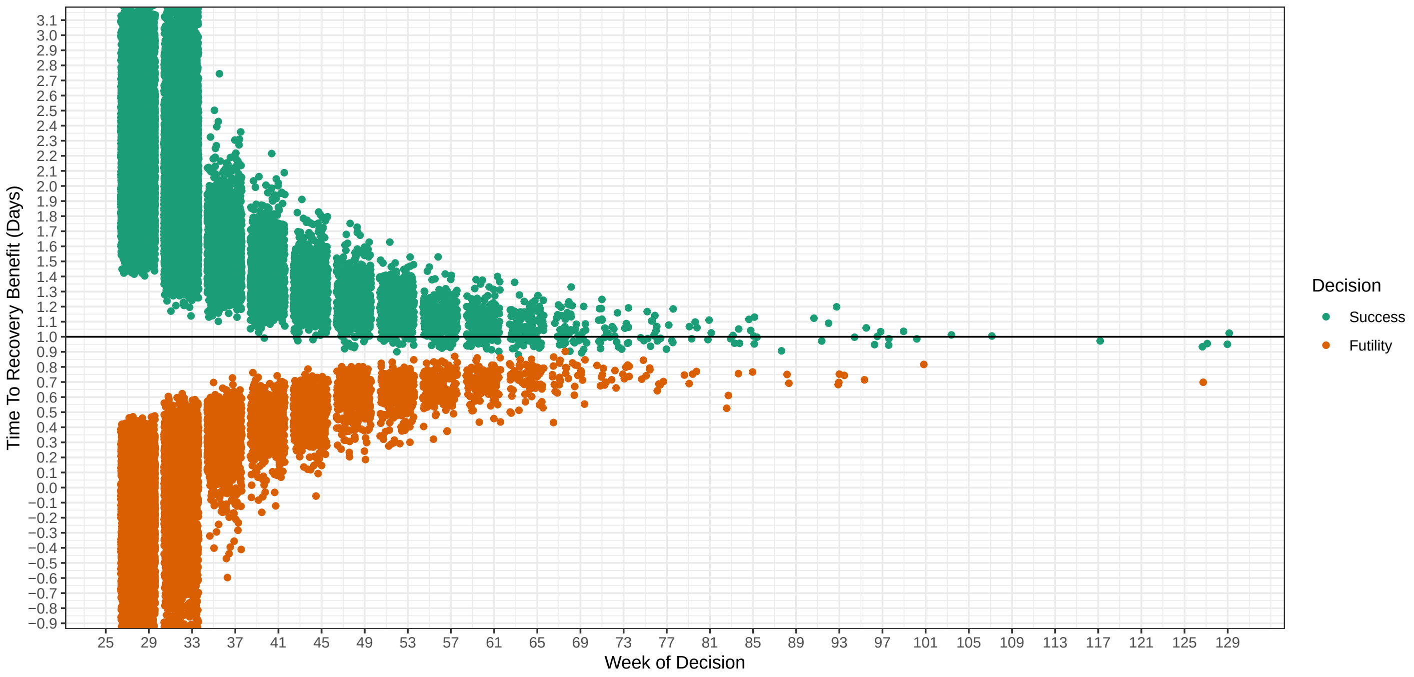

### Hazard Ratios: HCQ: 1 Azith: 1 Doxy: 1

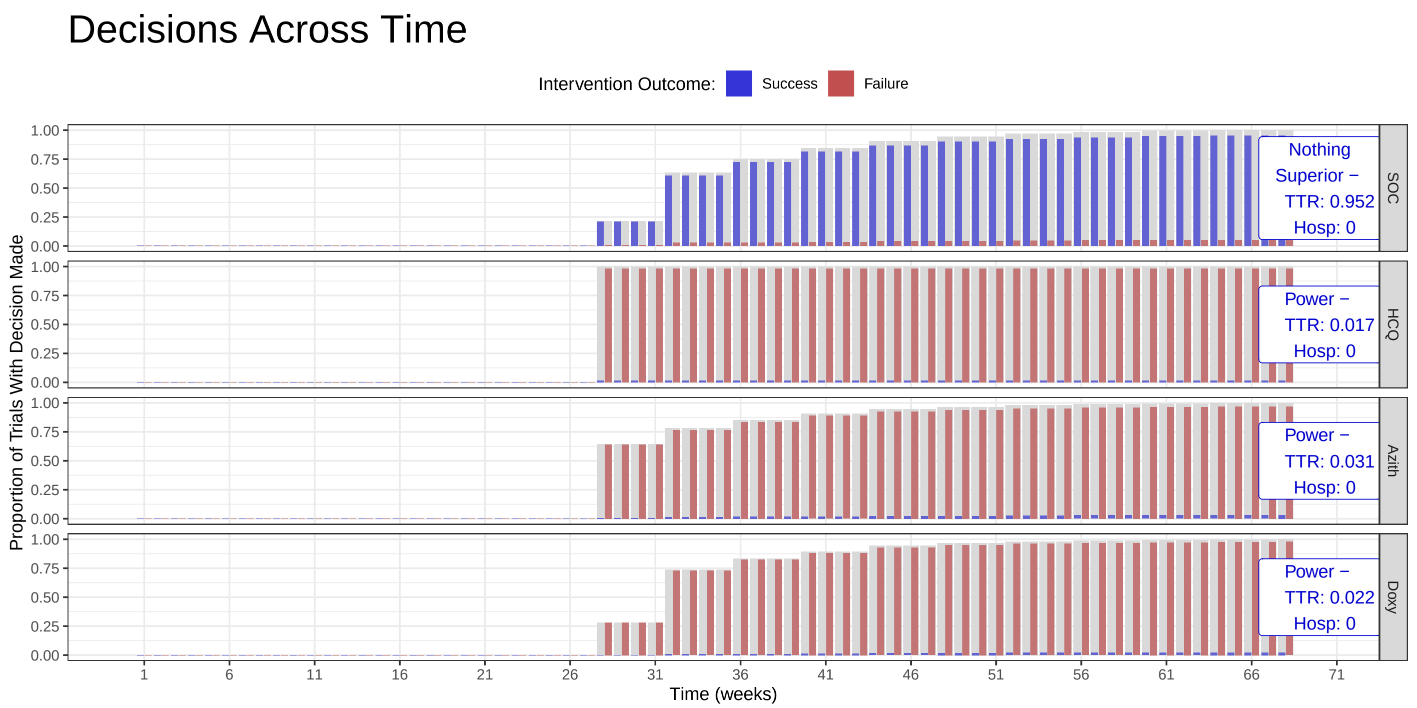

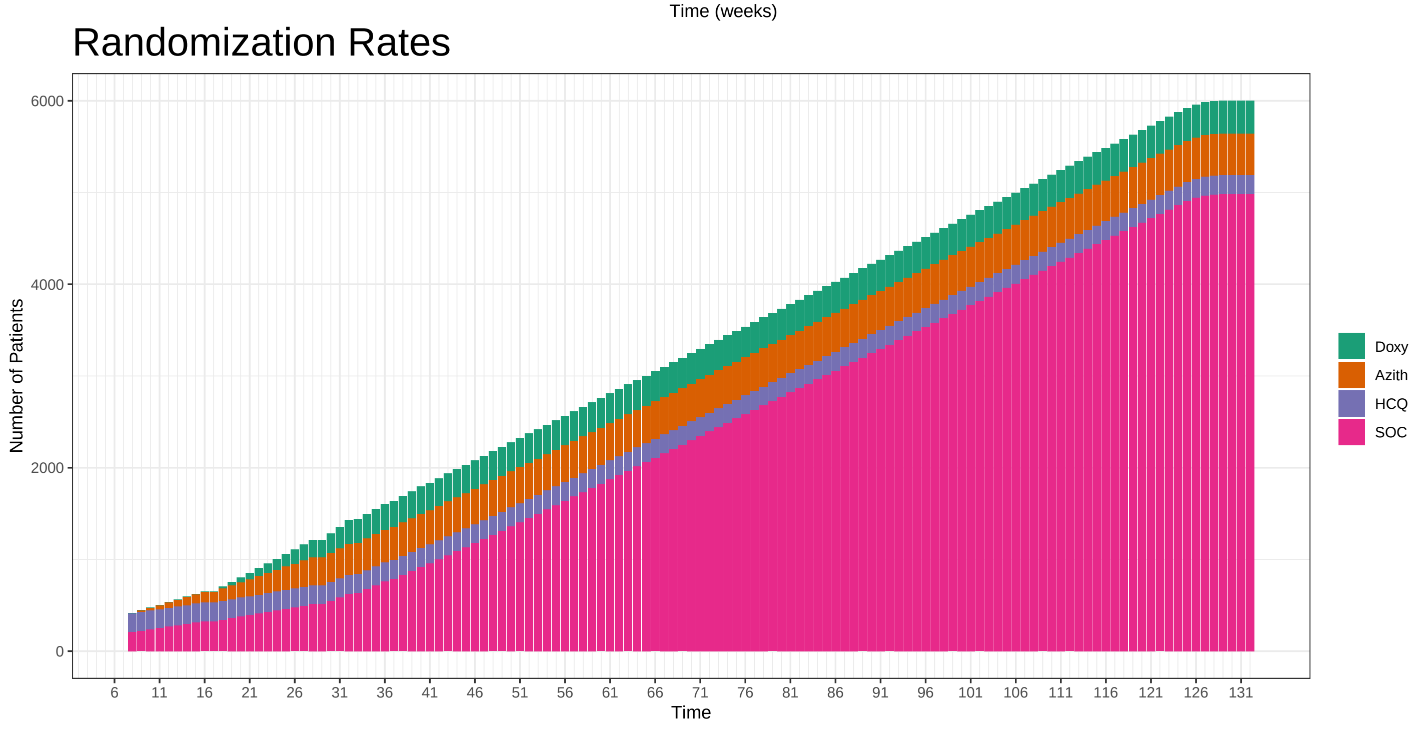

### Hazard Ratios: HCQ: 1 Azith: 1 Doxy: 1.1

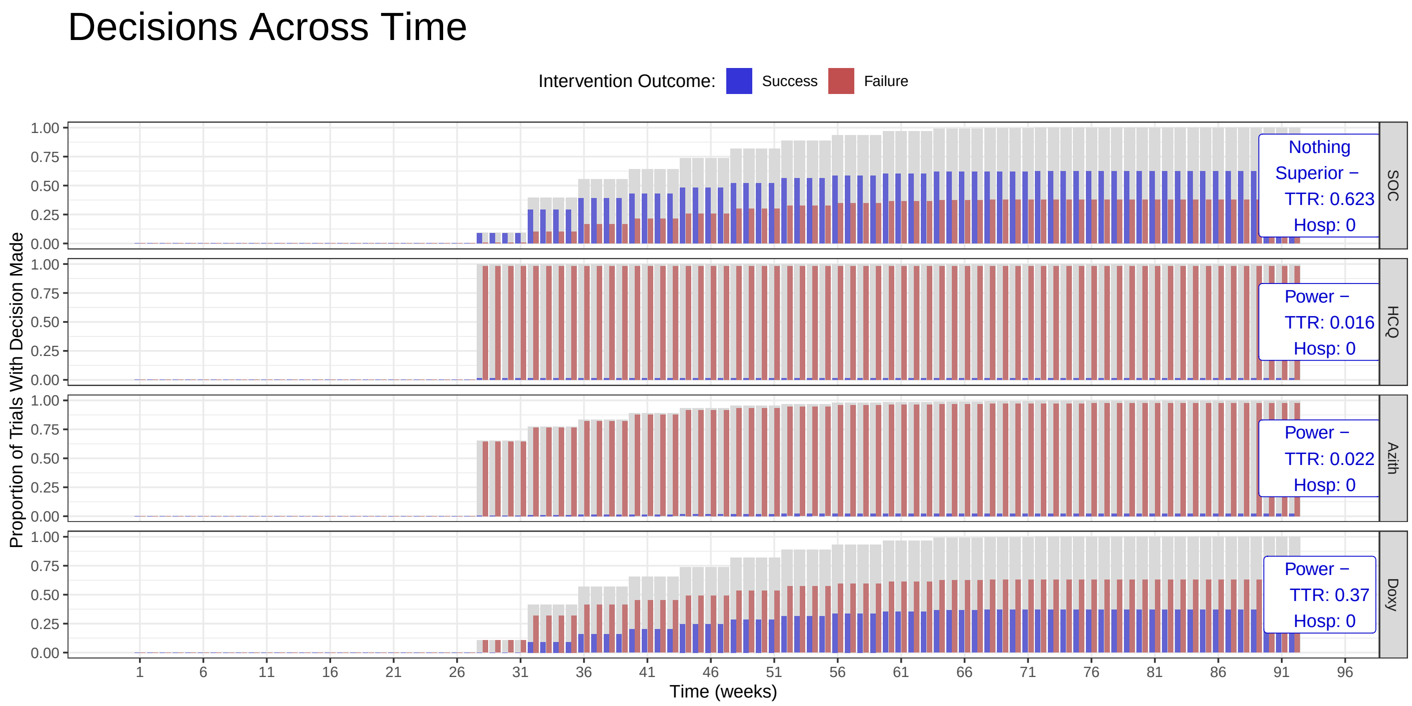

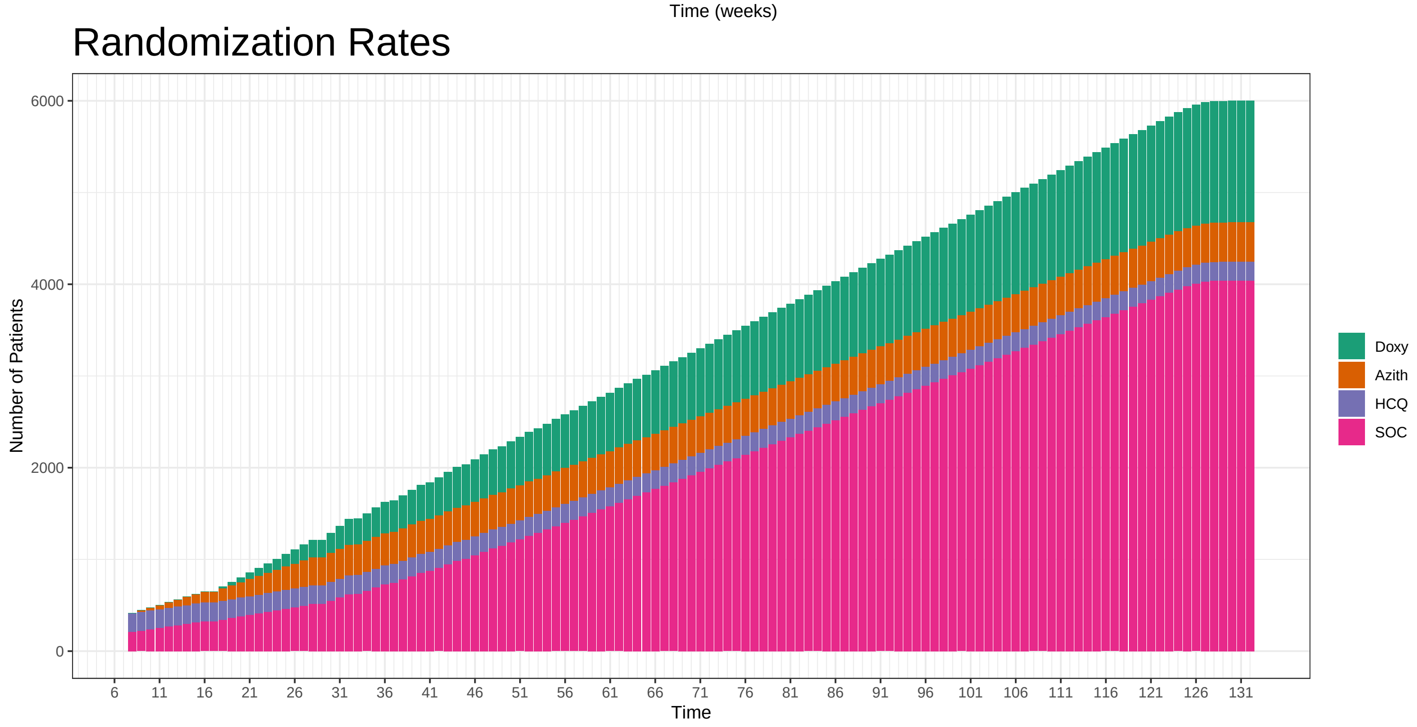

#### Hazard Ratios: HCQ: 1 Azith: 1 Doxy: 1.2

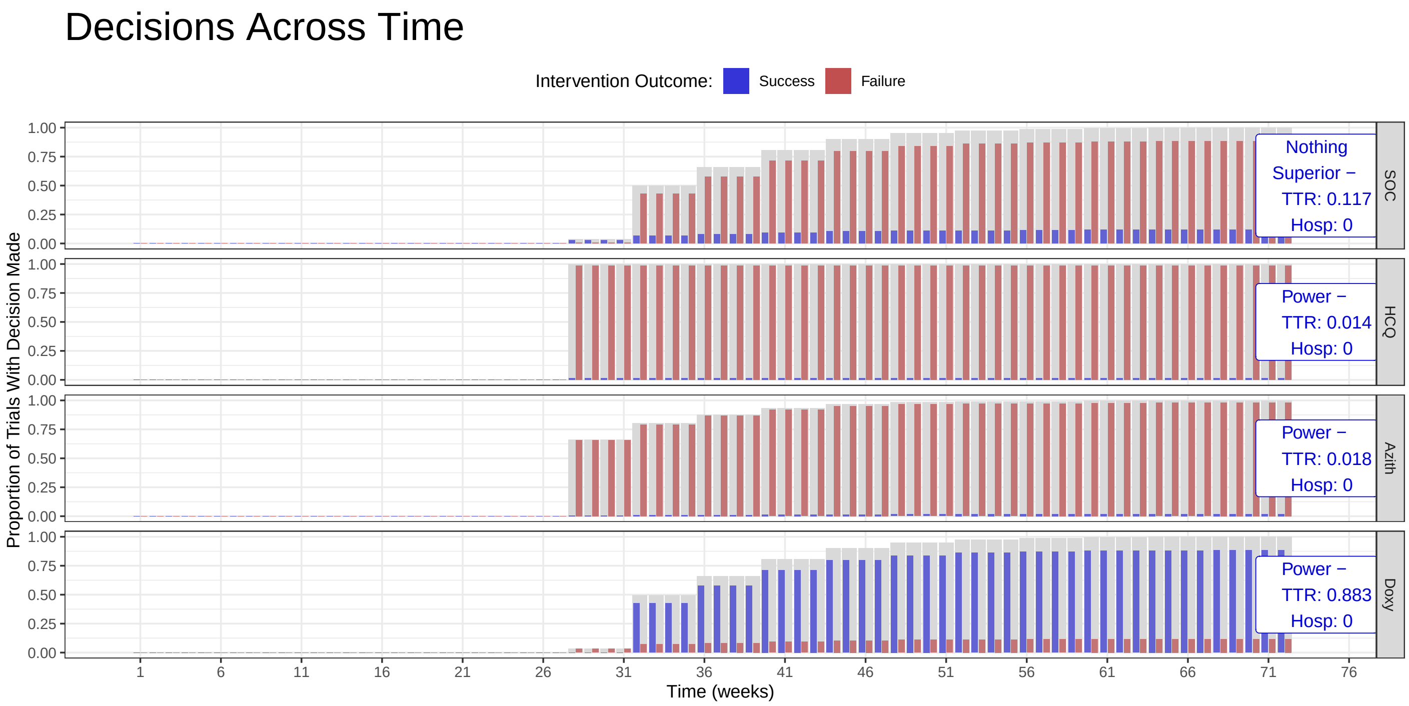

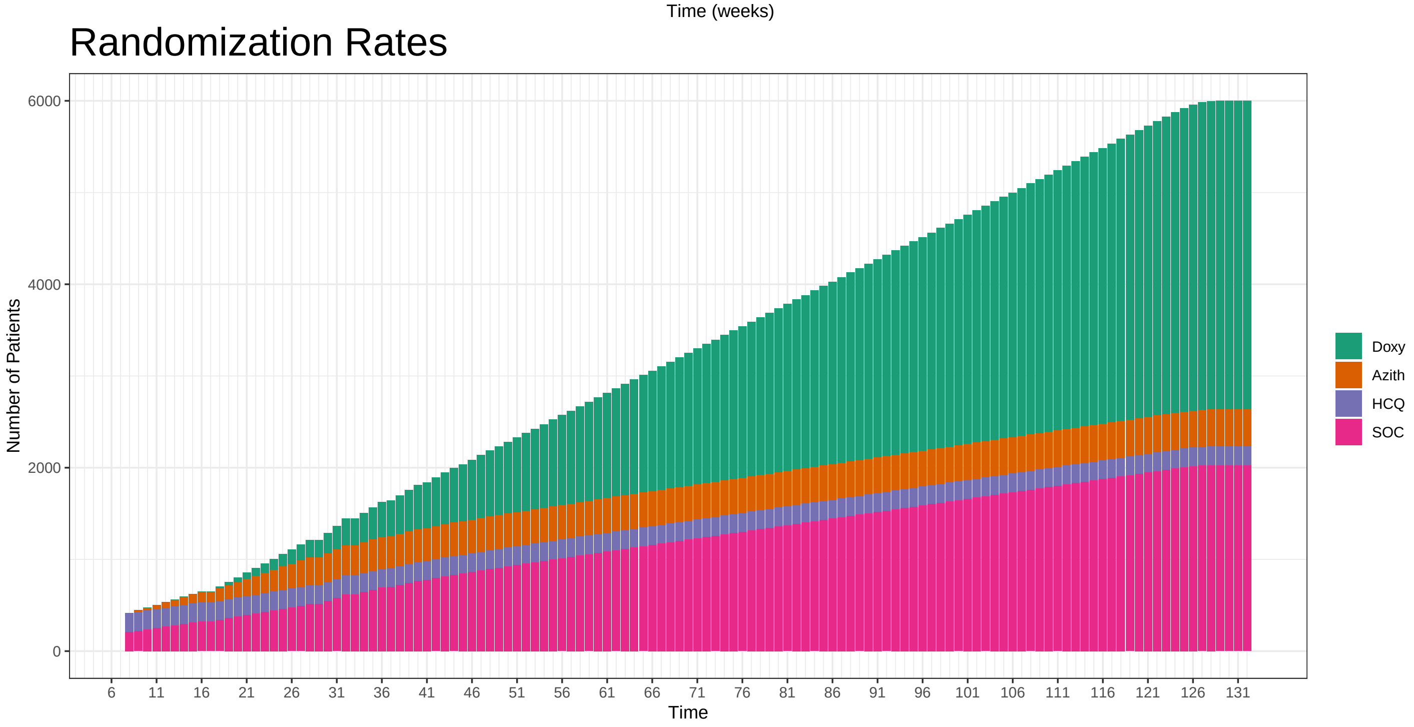

### Hazard Ratios: HCQ: 1 Azith: 1 Doxy: 1.3

#### Decisions Across Time

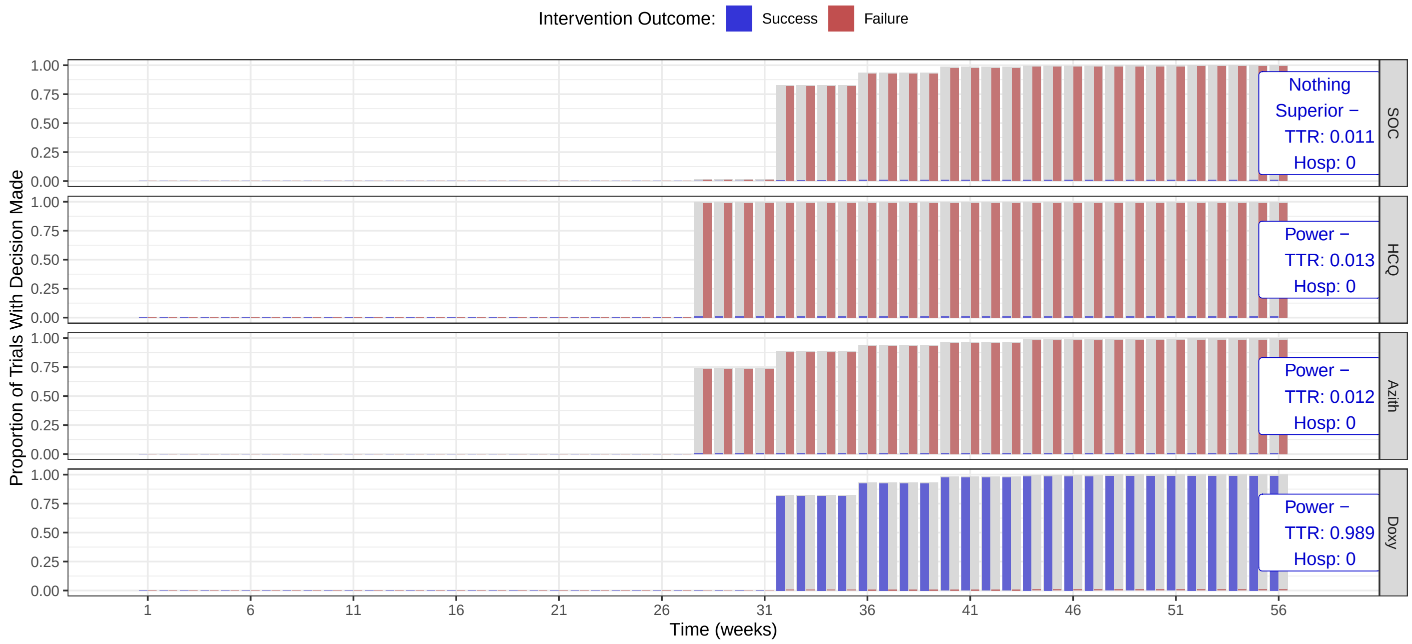

#### Randomization Rates

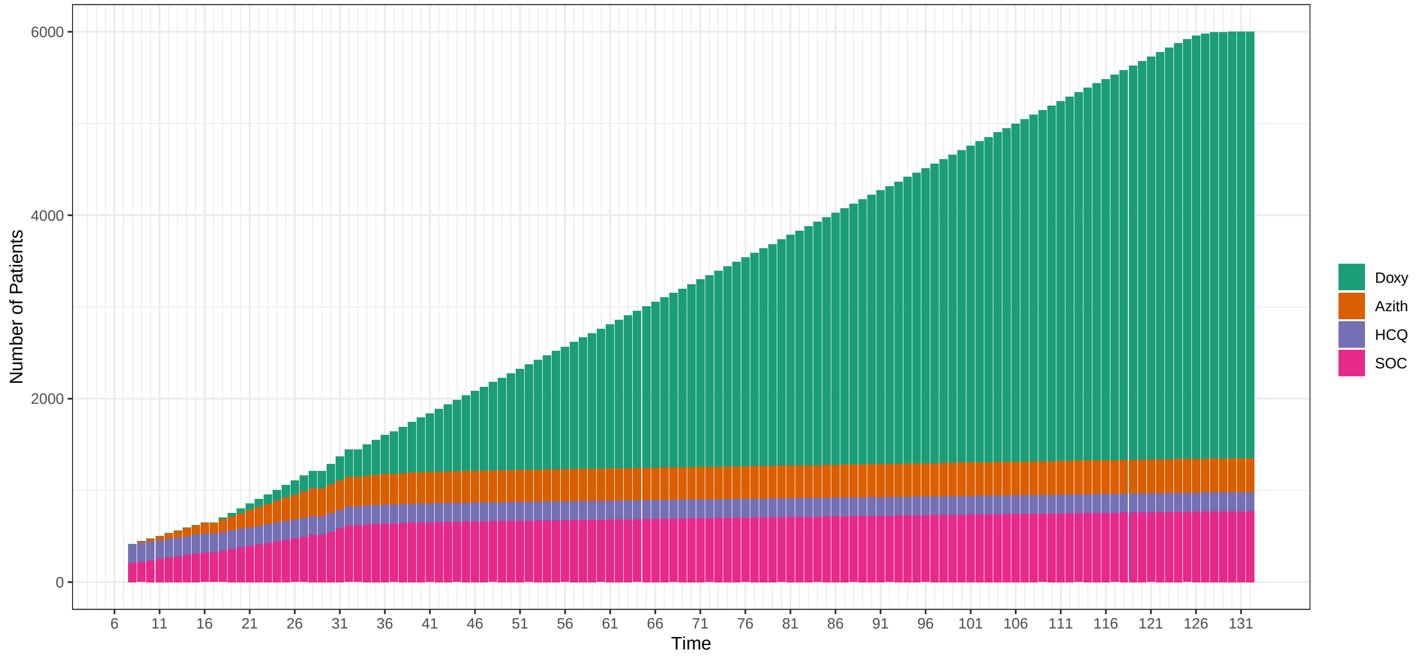

### Hazard Ratios: HCQ: 1 Azith: 1.1 Doxy: 1

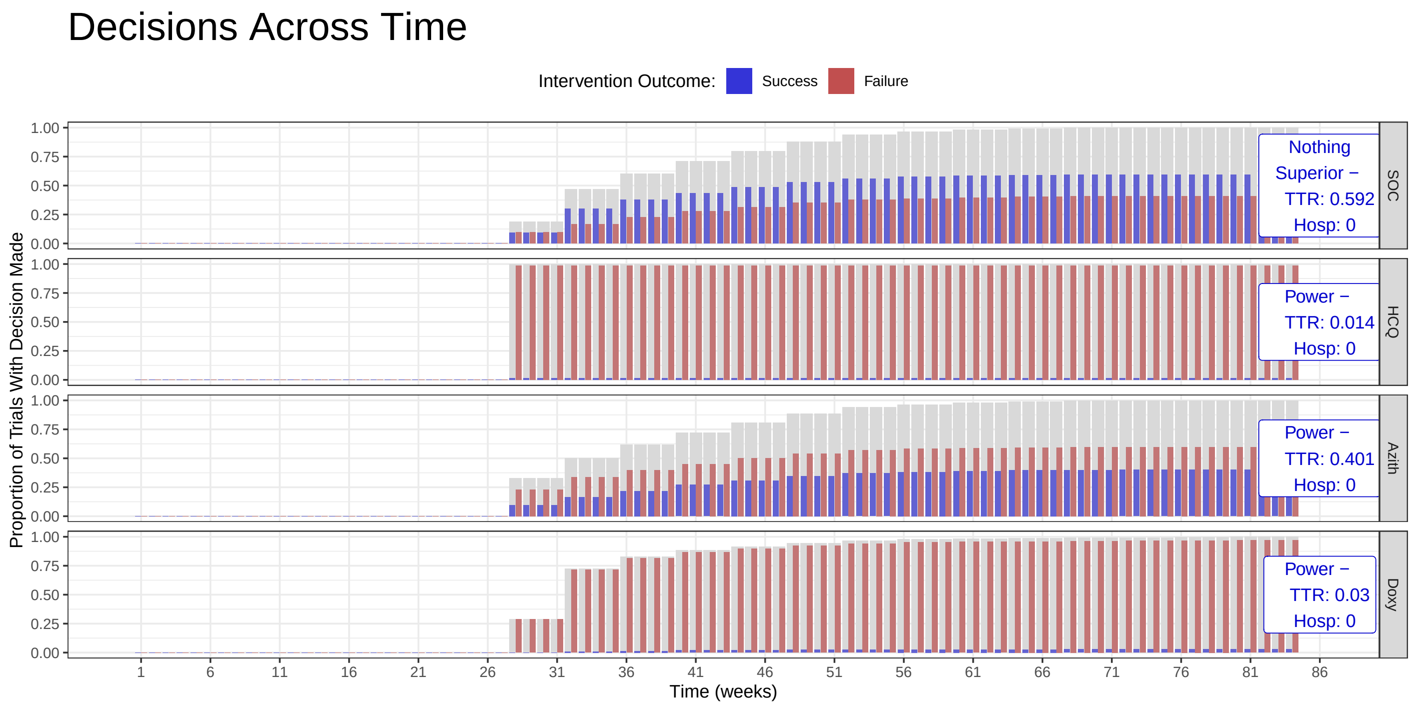

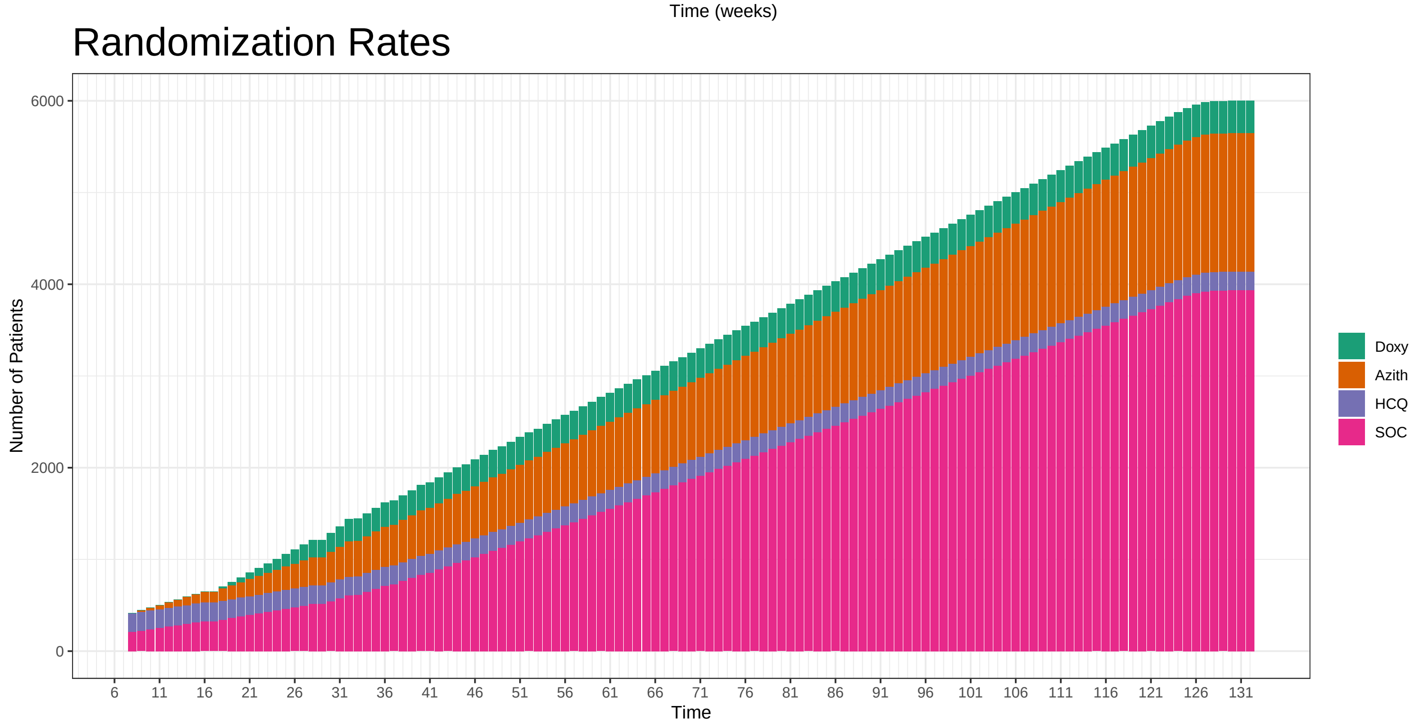

### Hazard Ratios: HCQ: 1 Azith: 1.1 Doxy: 1.2

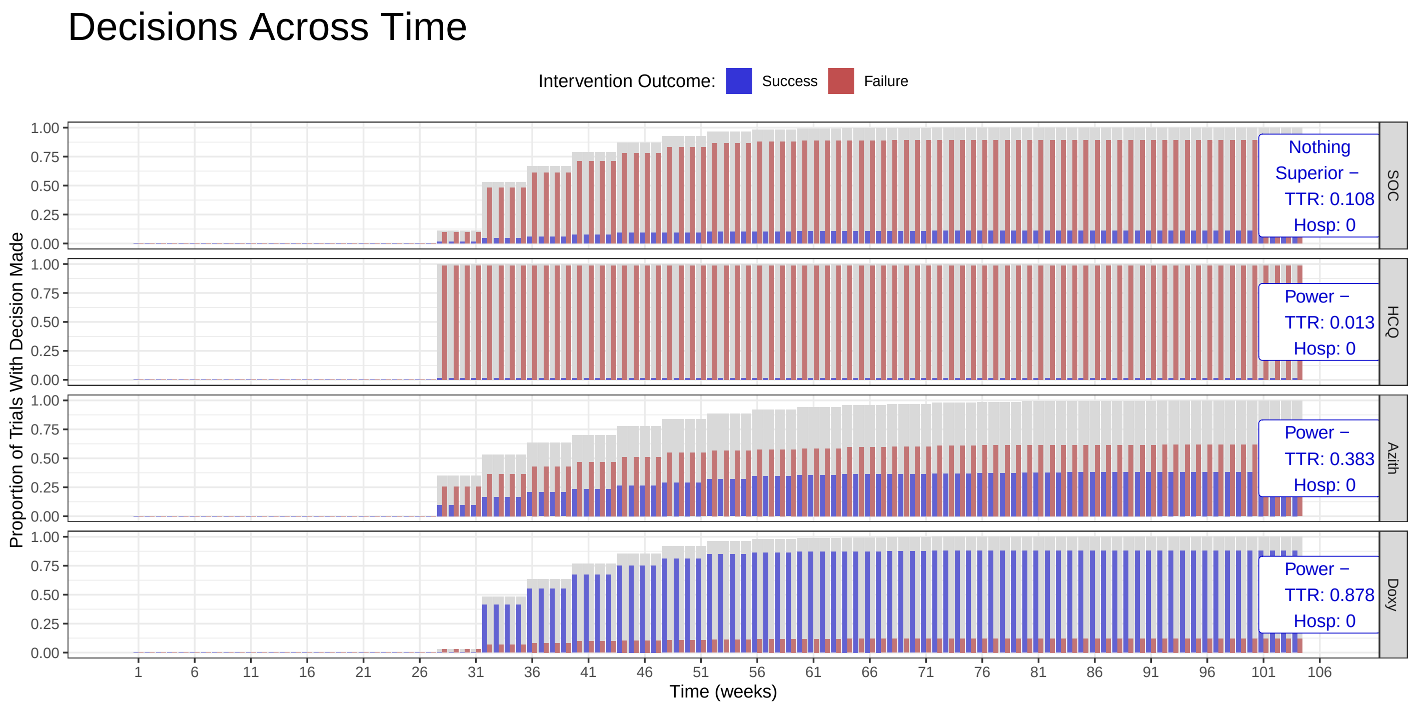

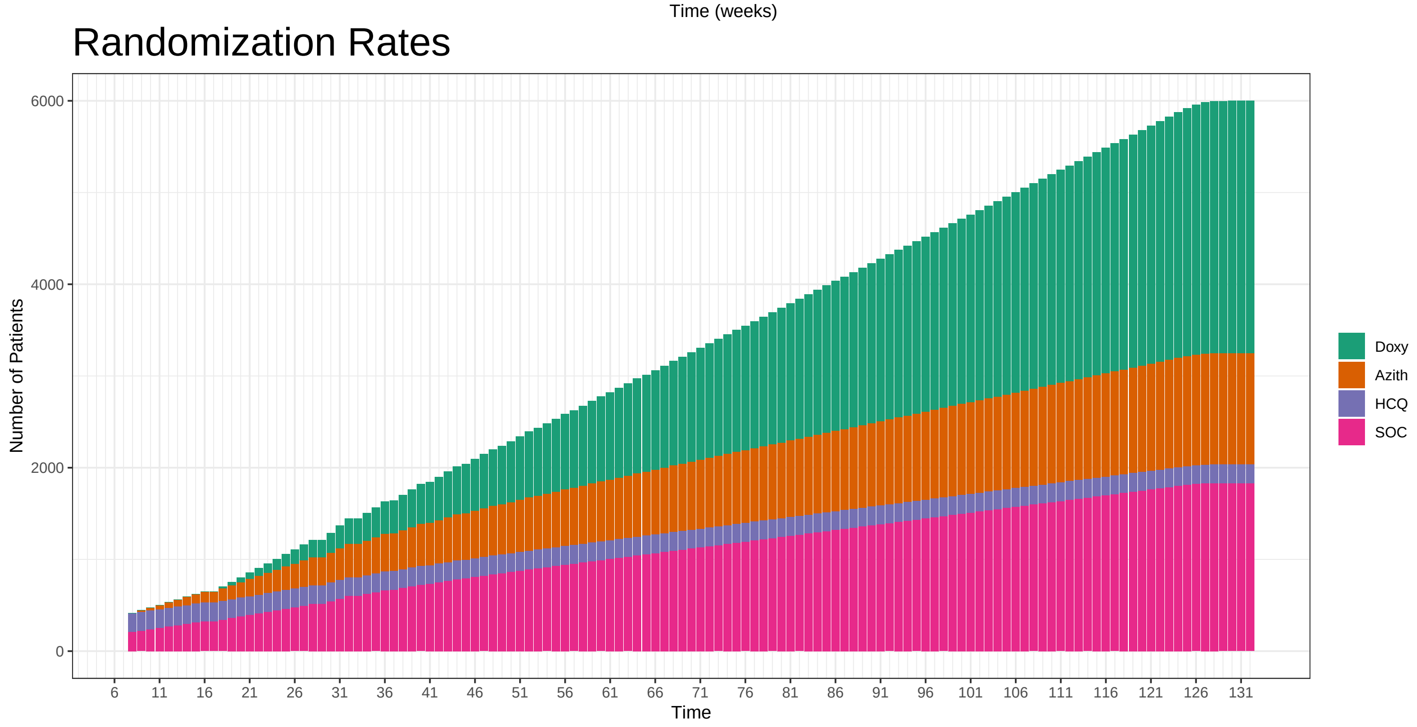

### Hazard Ratios: HCQ: 1 Azith: 1.1 Doxy: 1.3

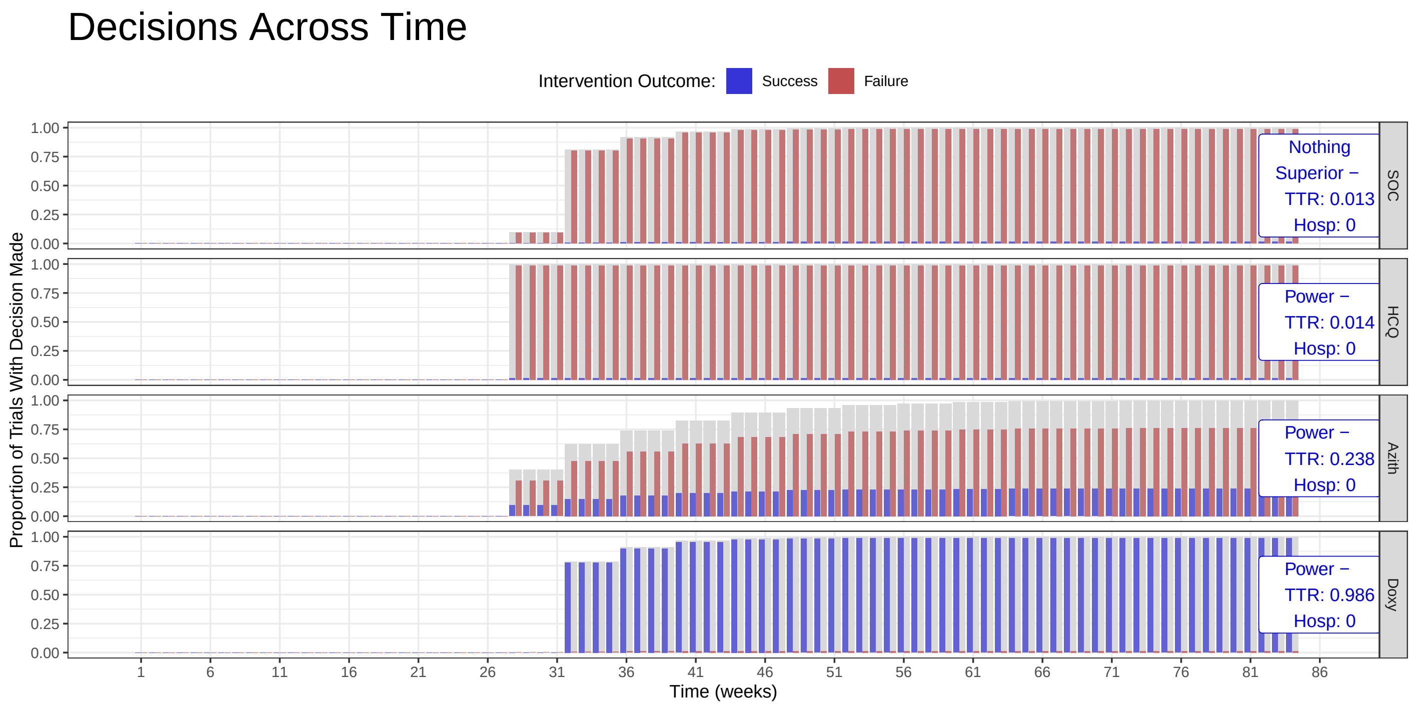

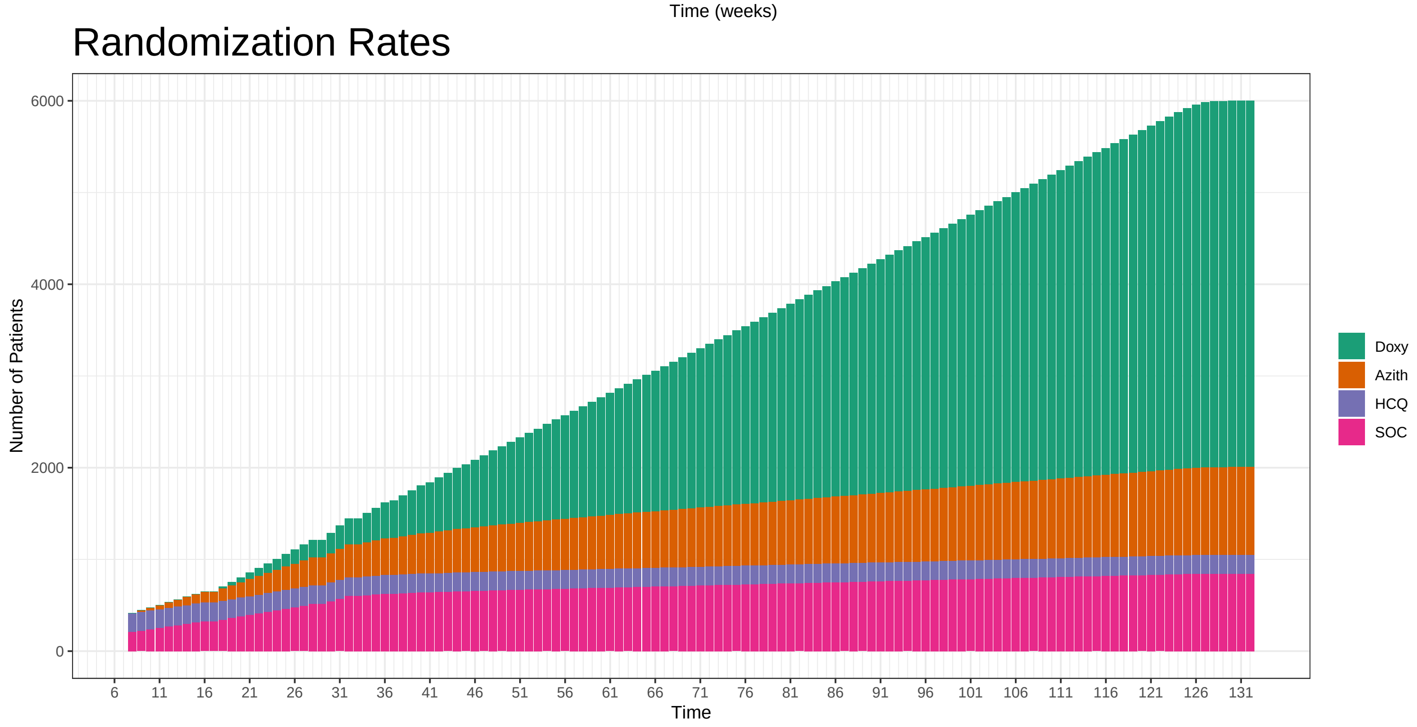

### Hazard Ratios: HCQ: 1 Azith: 1.2 Doxy: 1

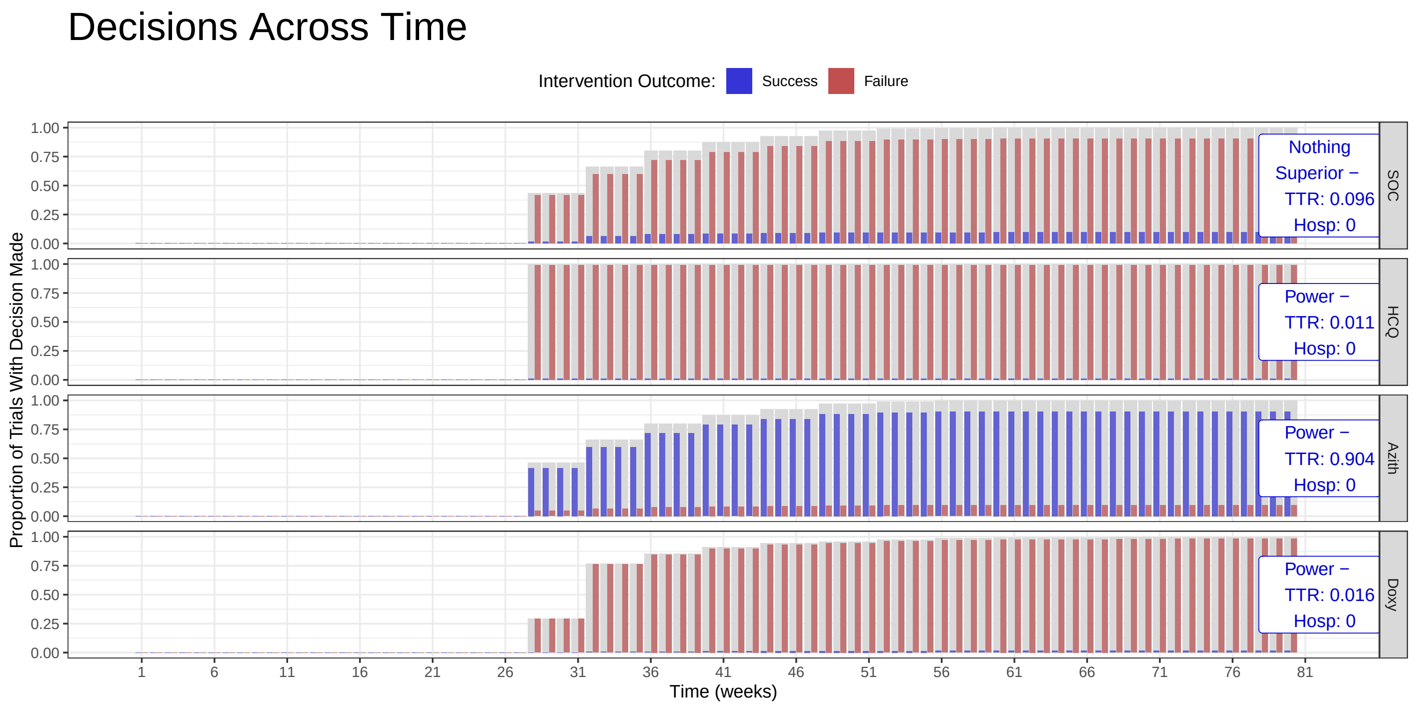

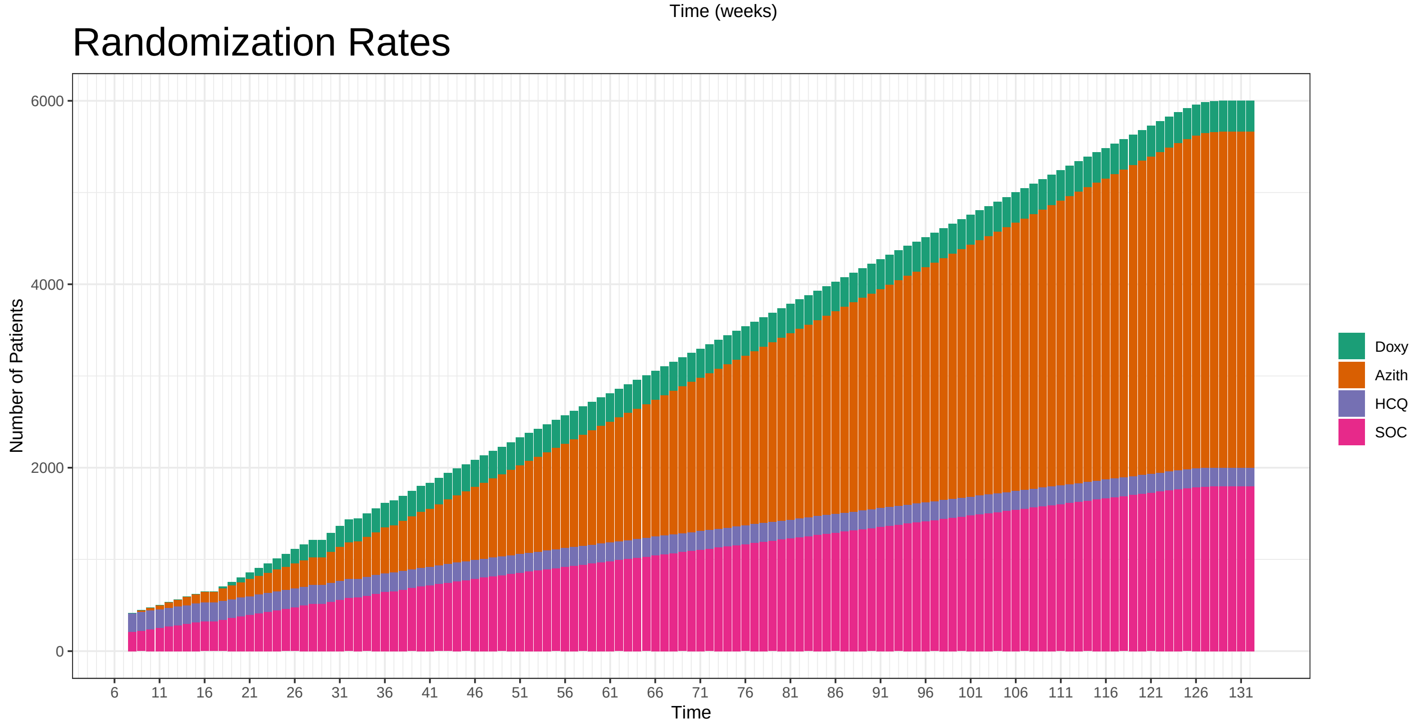

### Hazard Ratios: HCQ: 1 Azith: 1.2 Doxy: 1.1

#### Decisions Across Time

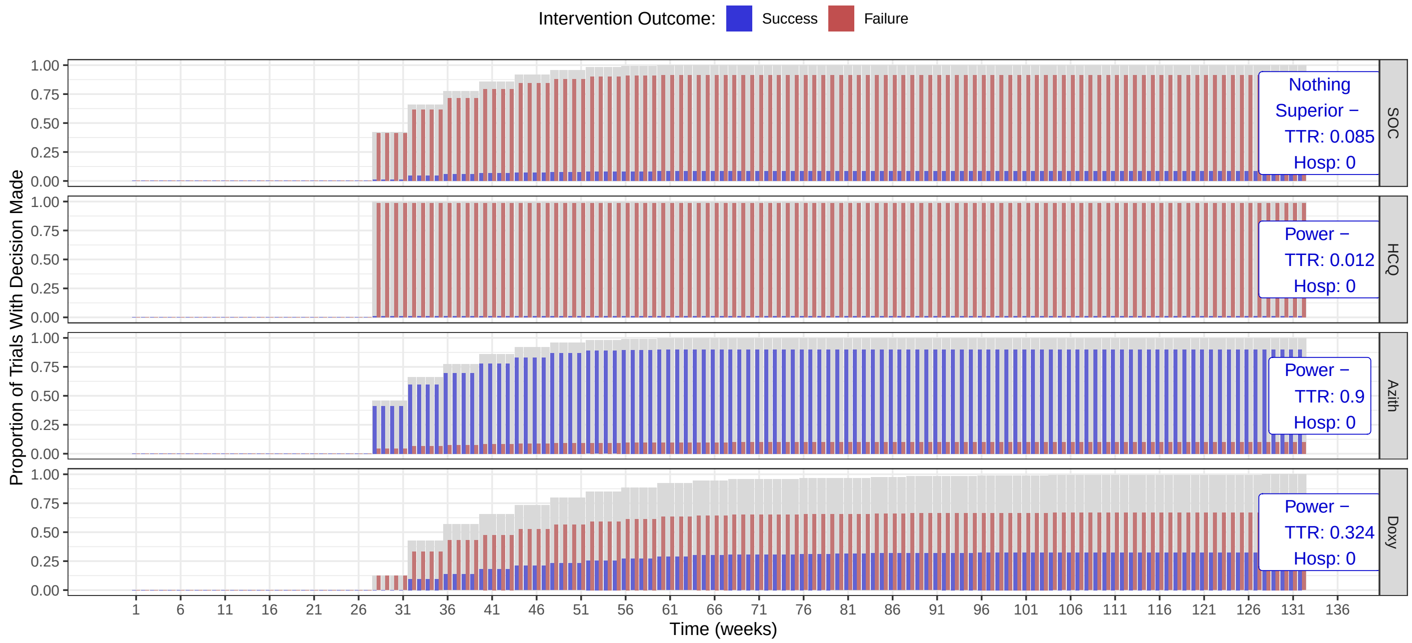

#### Randomization Rates

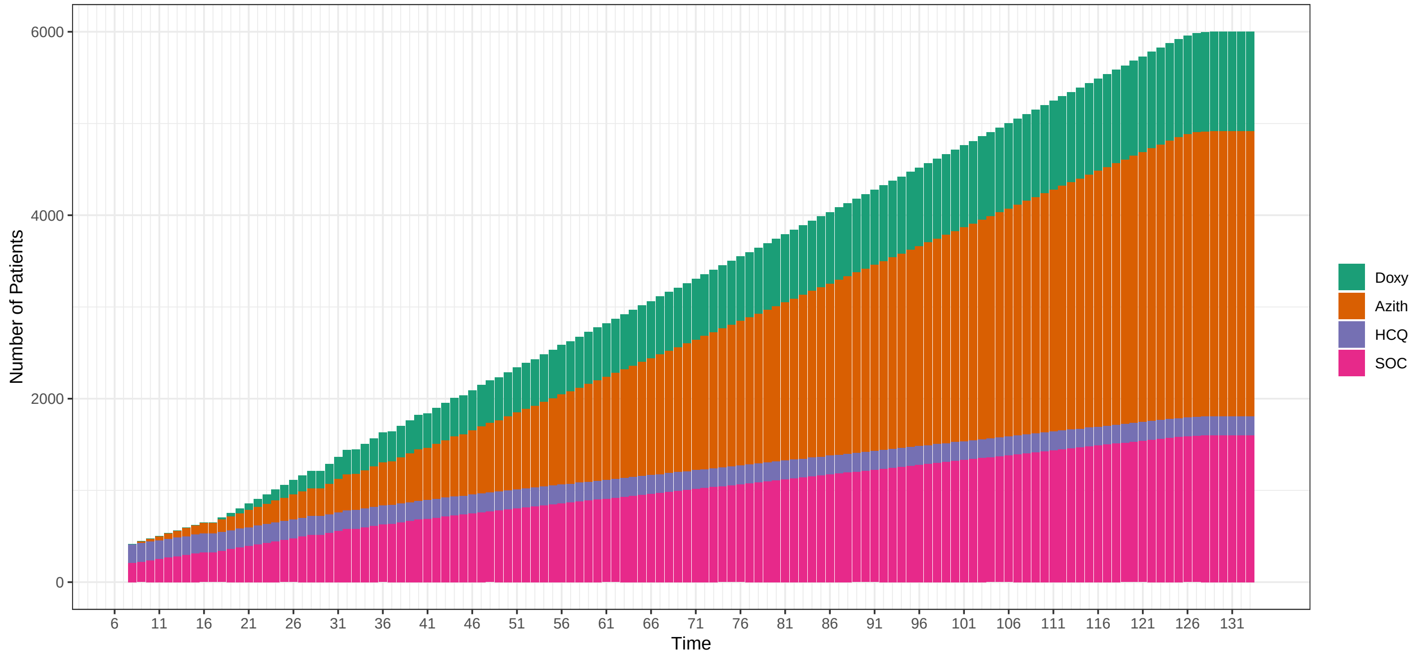

**Hazard Ratios: HCQ: 1 Azith: 1.2 Doxy: 1.2**

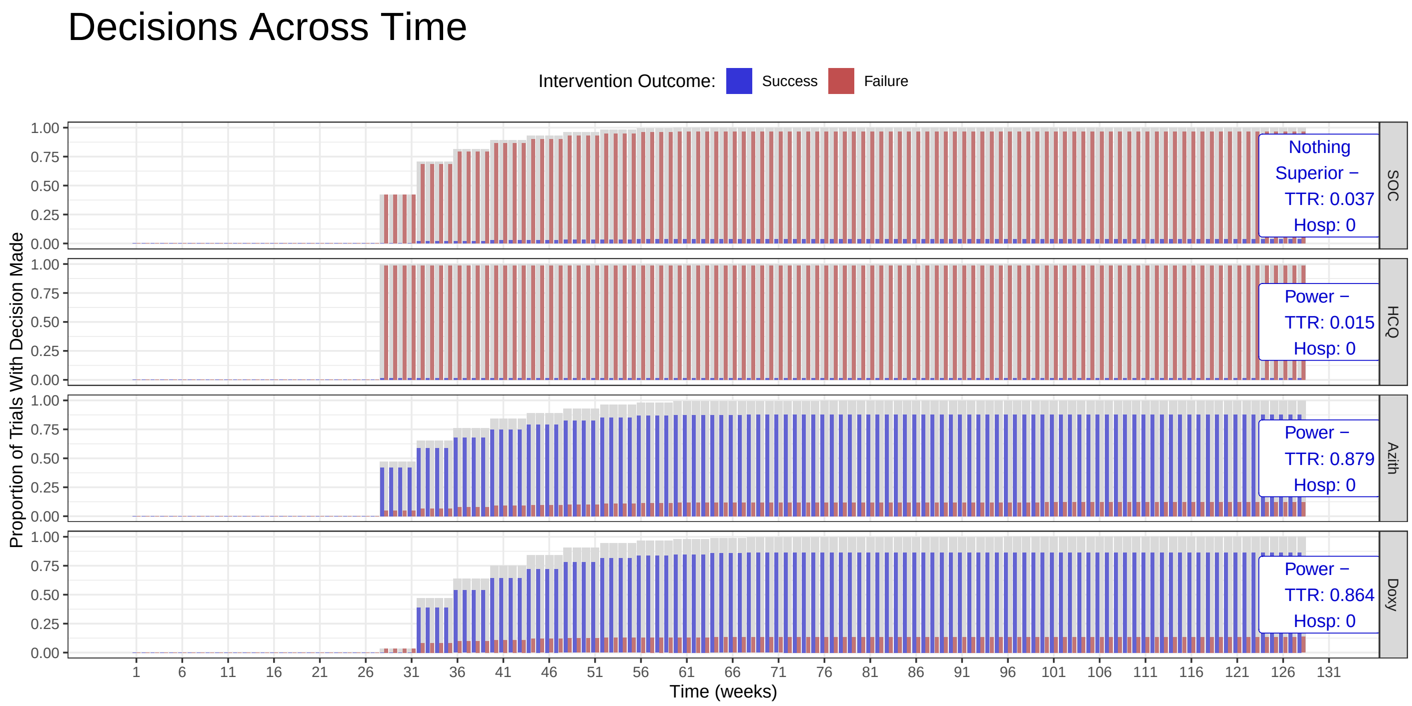

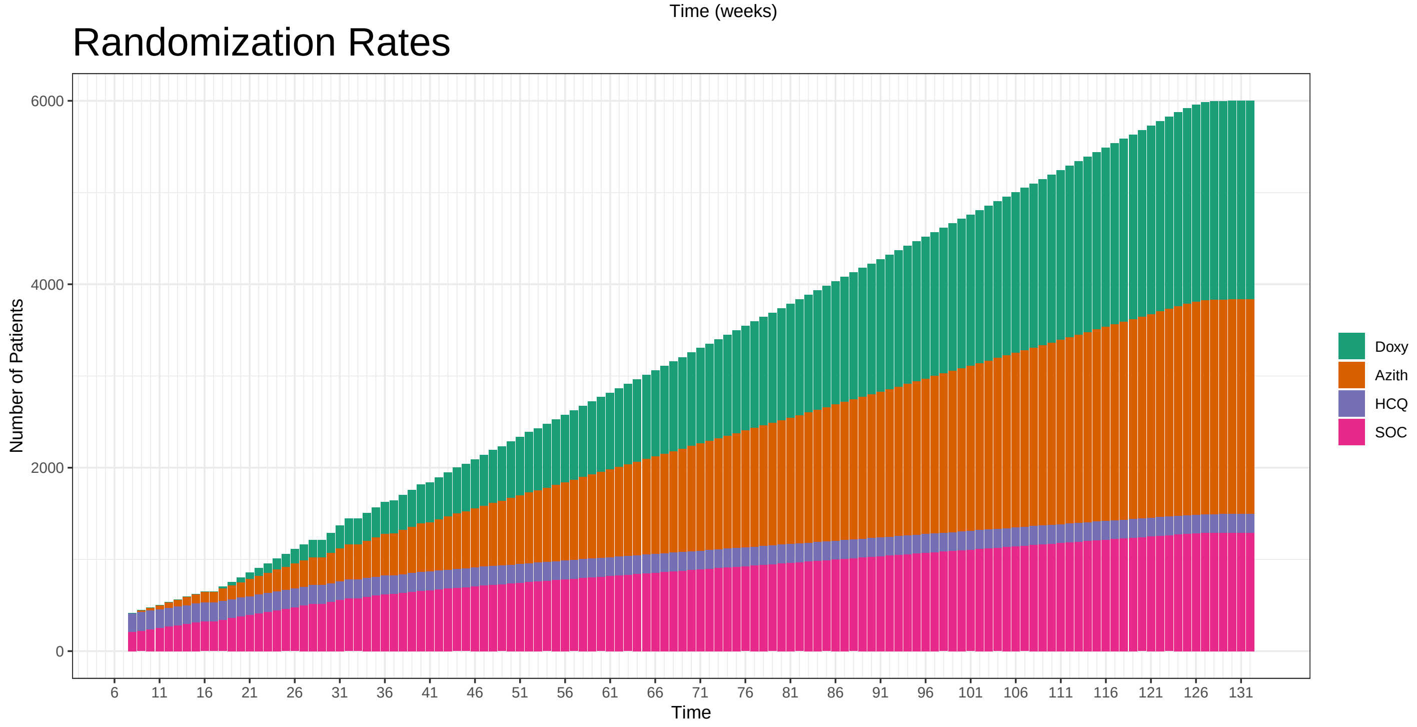

### Hazard Ratios: HCQ: 1 Azith: 1.2 Doxy: 1.3

#### Decisions Across Time

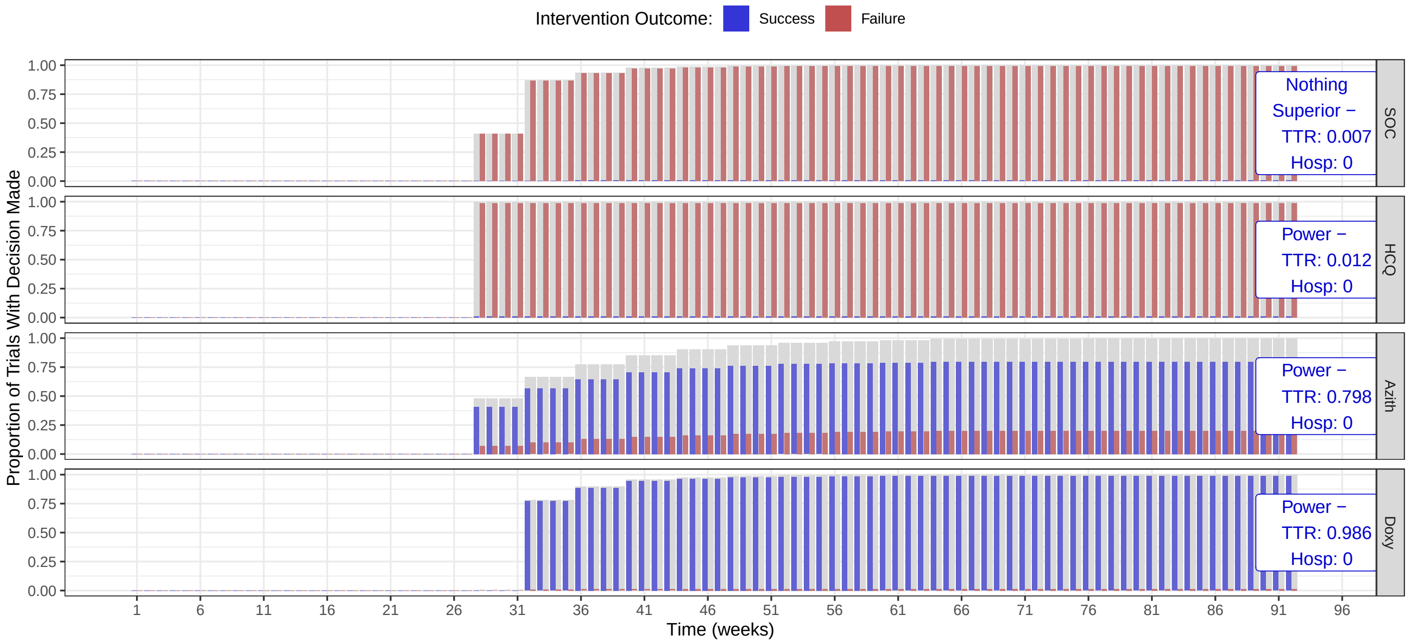

#### Randomization Rates

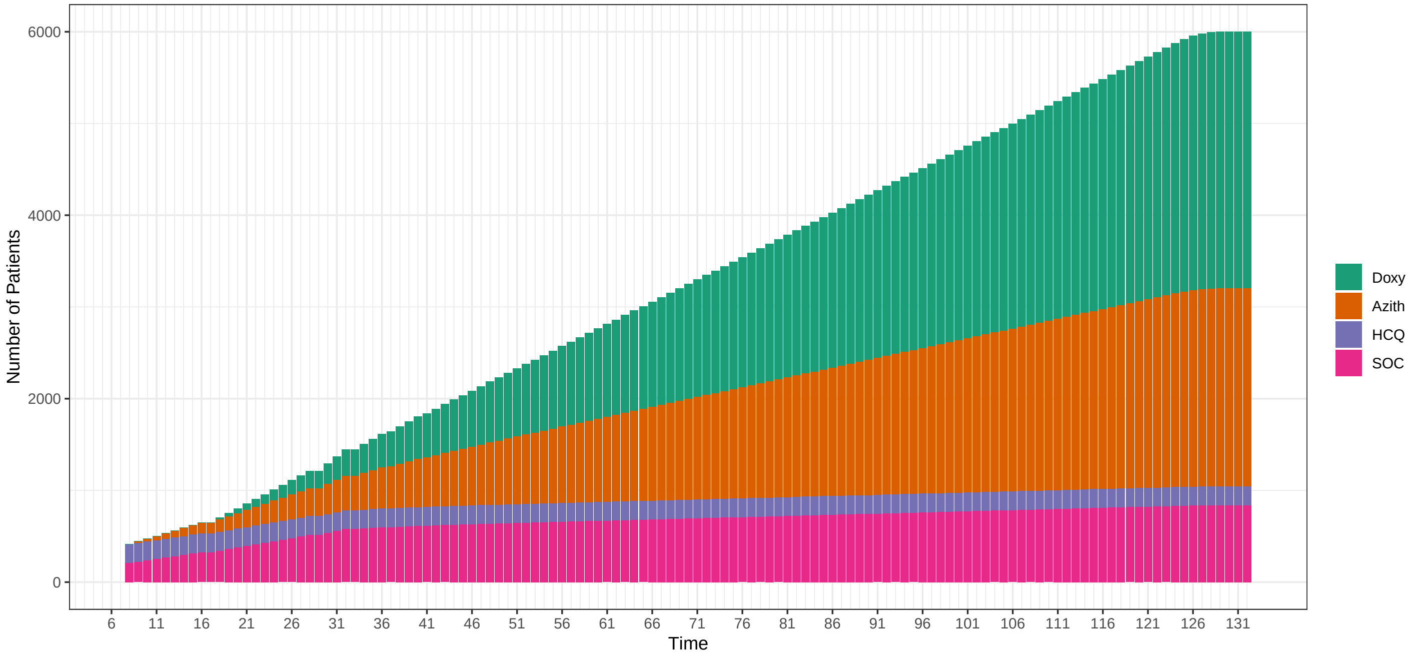

#### Hazard Ratios: HCQ: 1 Azith: 1.3 Doxy: 1

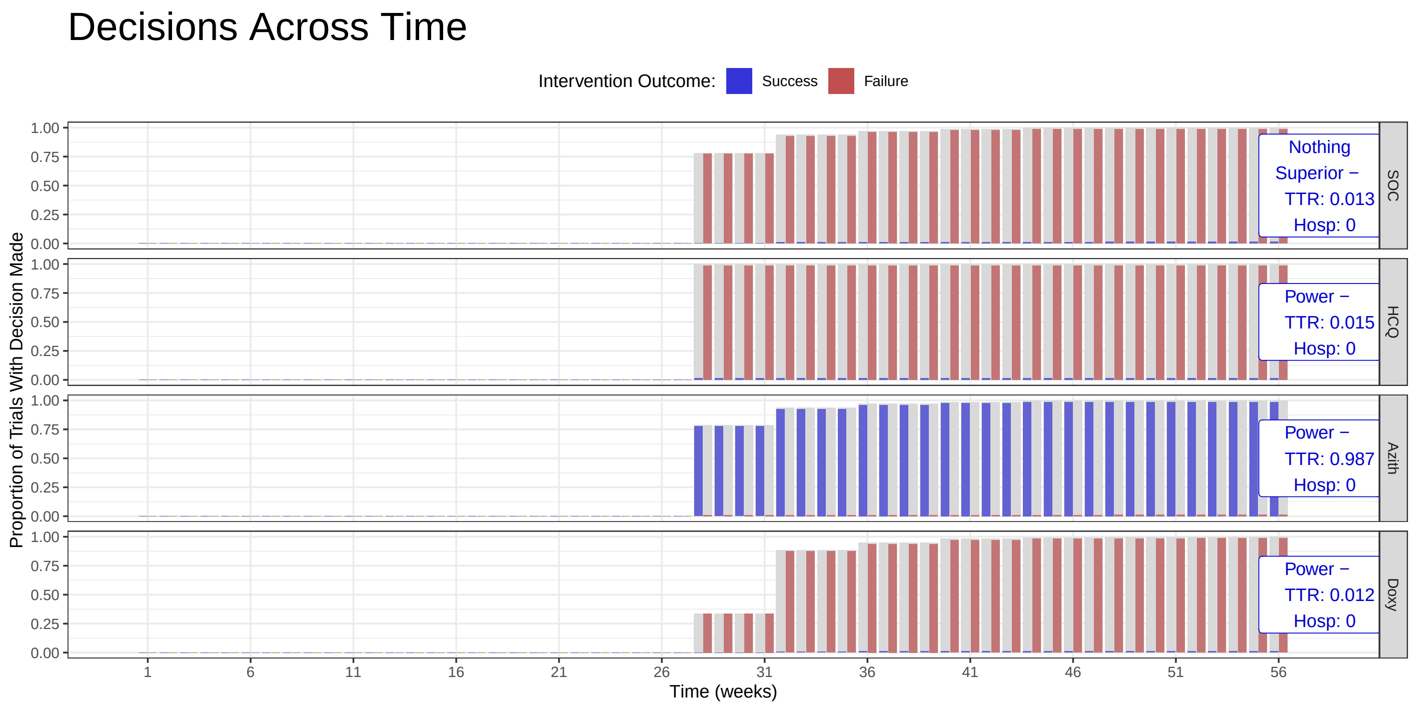

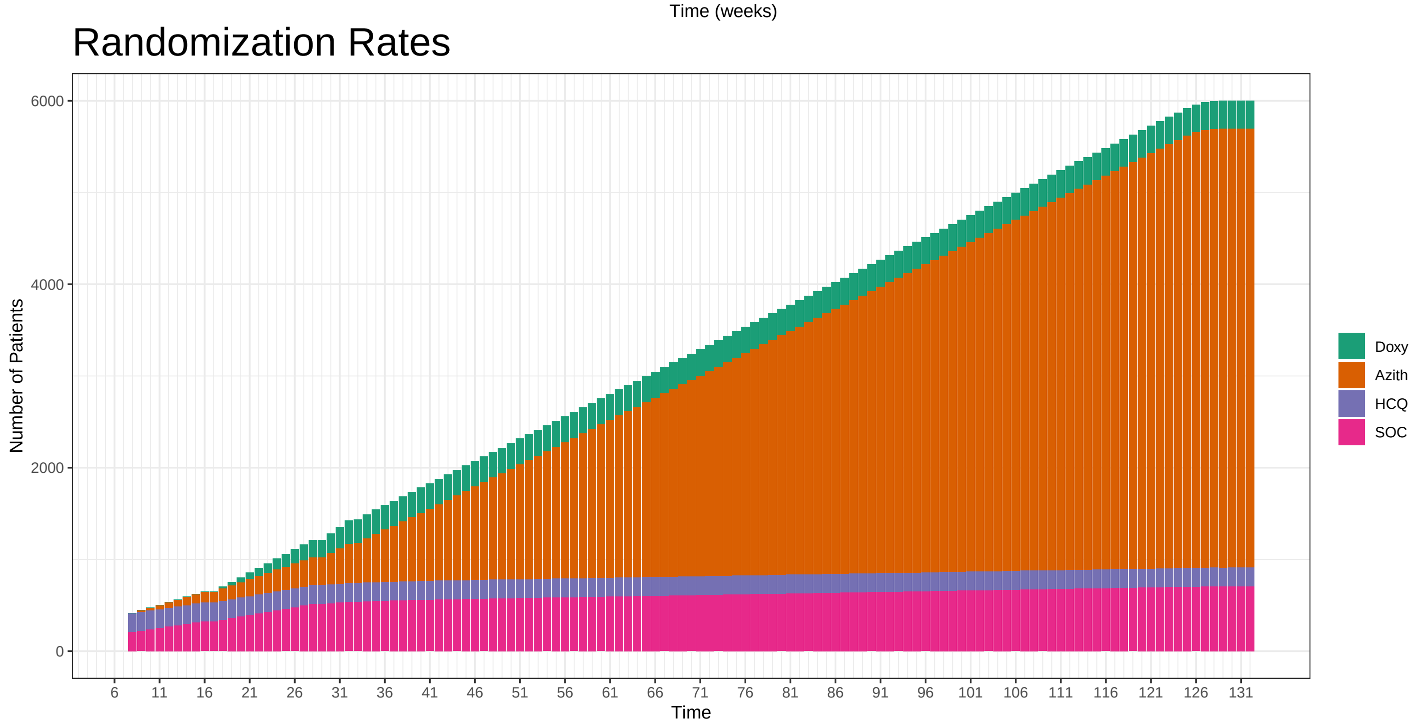

### Hazard Ratios: HCQ: 1 Azith: 1.3 Doxy: 1.1

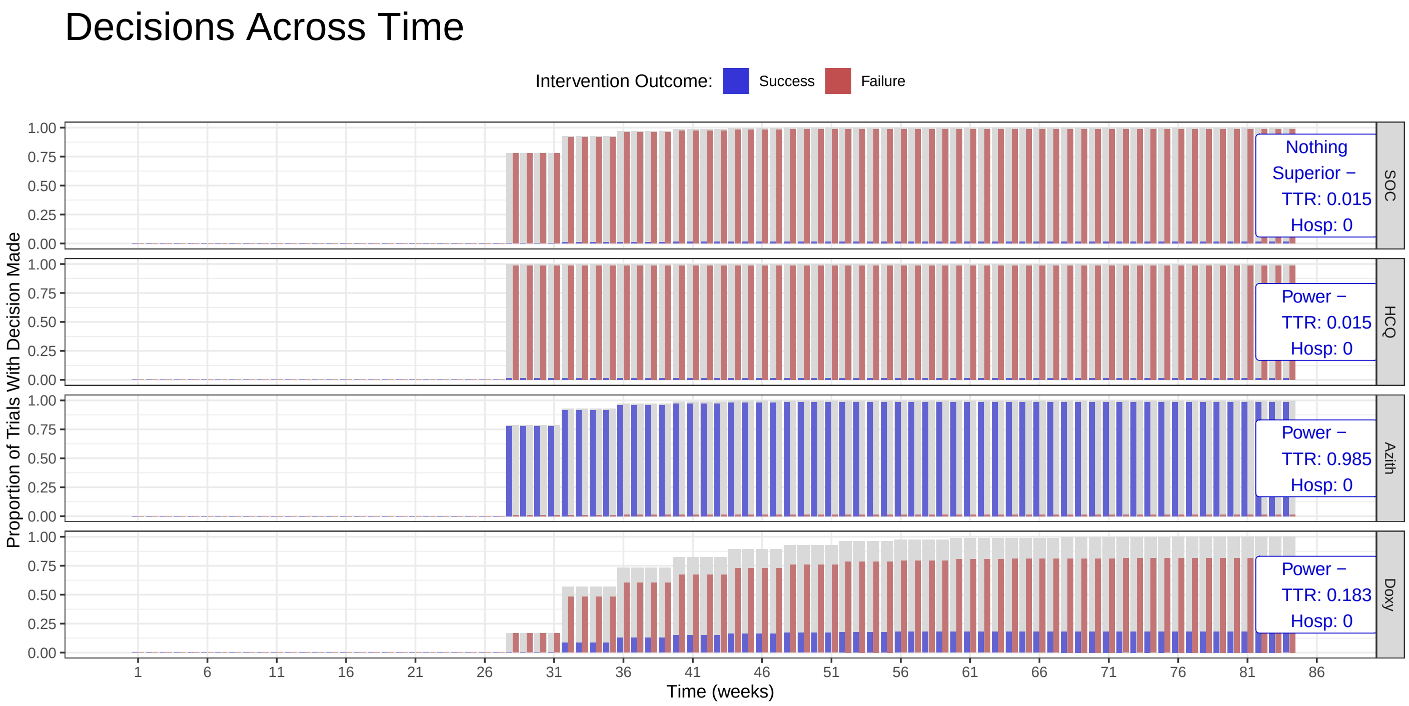

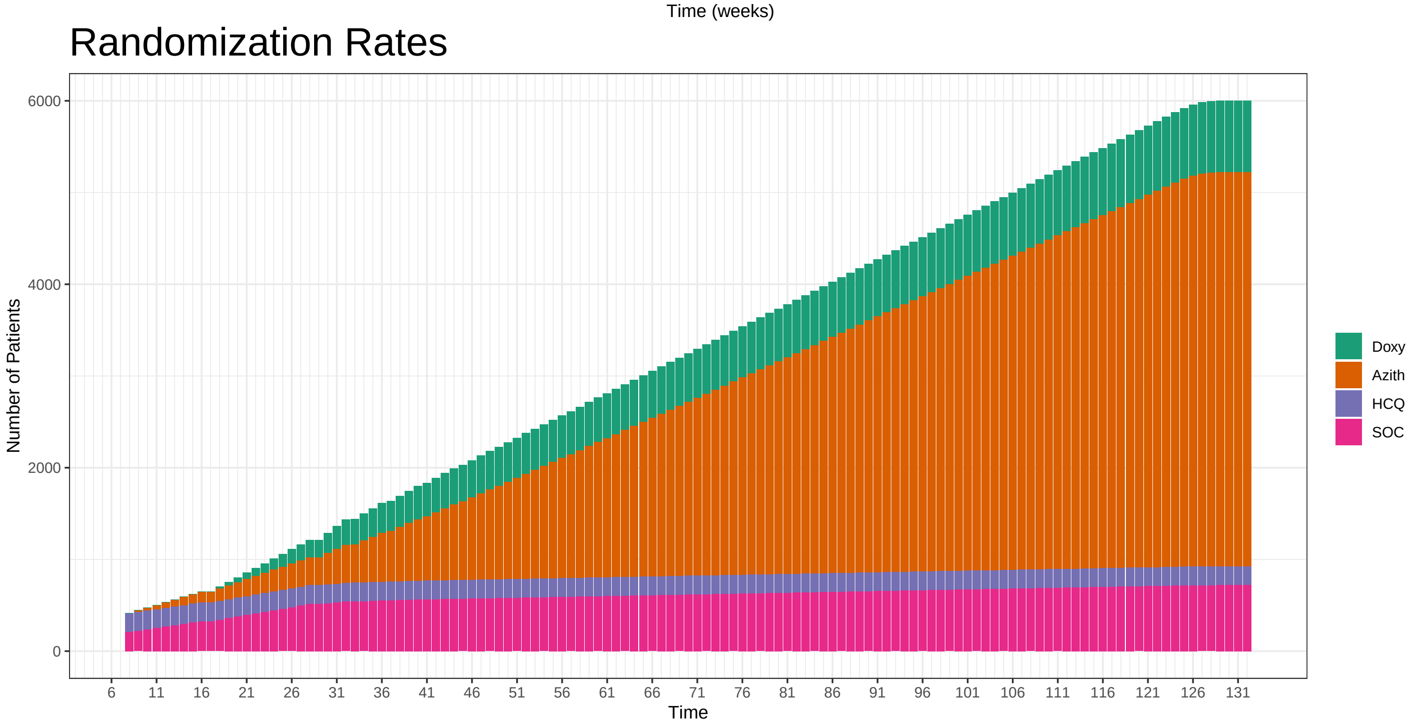

### Hazard Ratios: HCQ: 1 Azith: 1.3 Doxy: 1.2

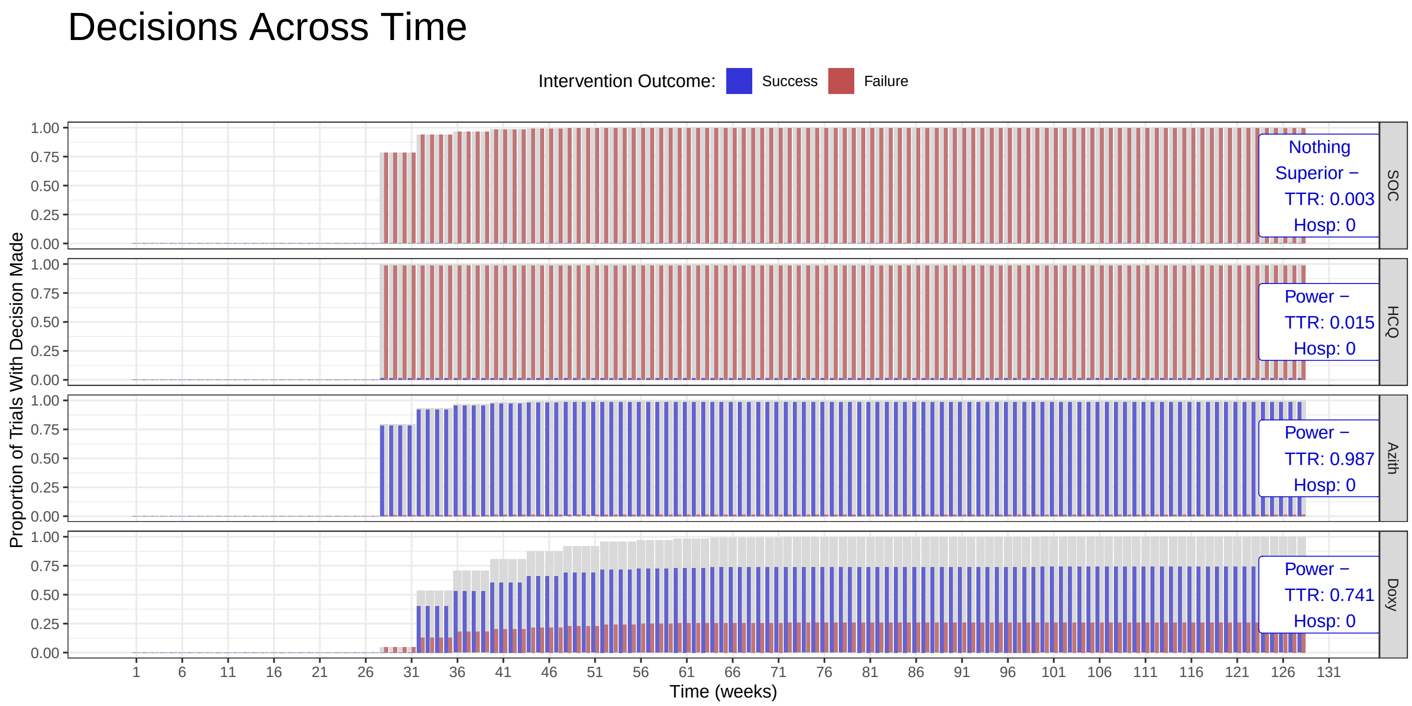

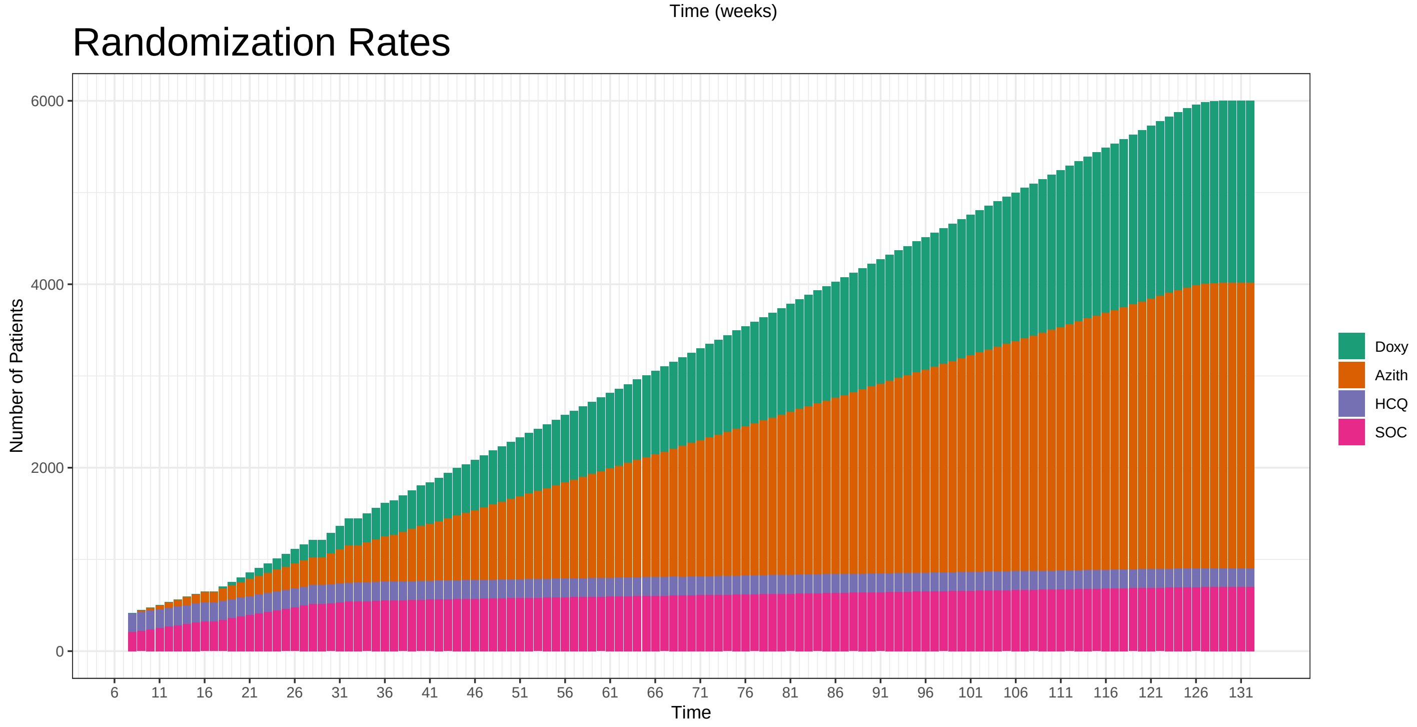

### Hazard Ratios: HCQ: 1 Azith: 1.3 Doxy: 1.3

#### Decisions Across Time

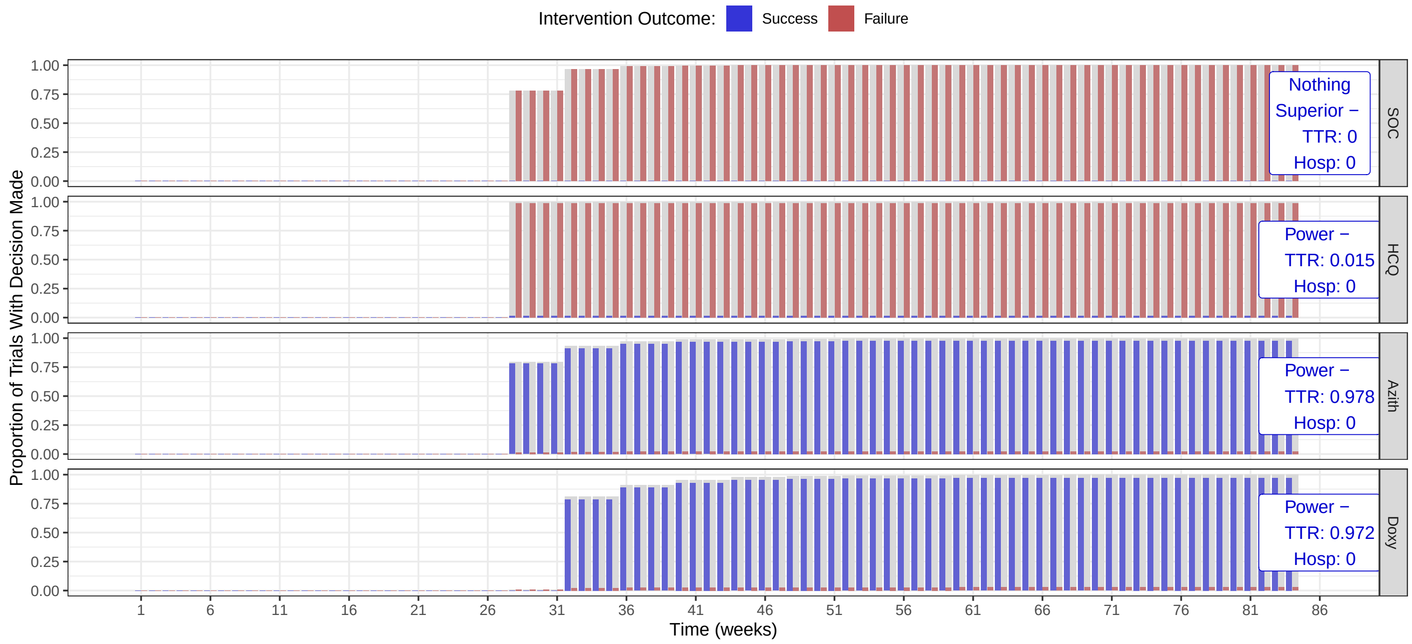

#### Randomization Rates

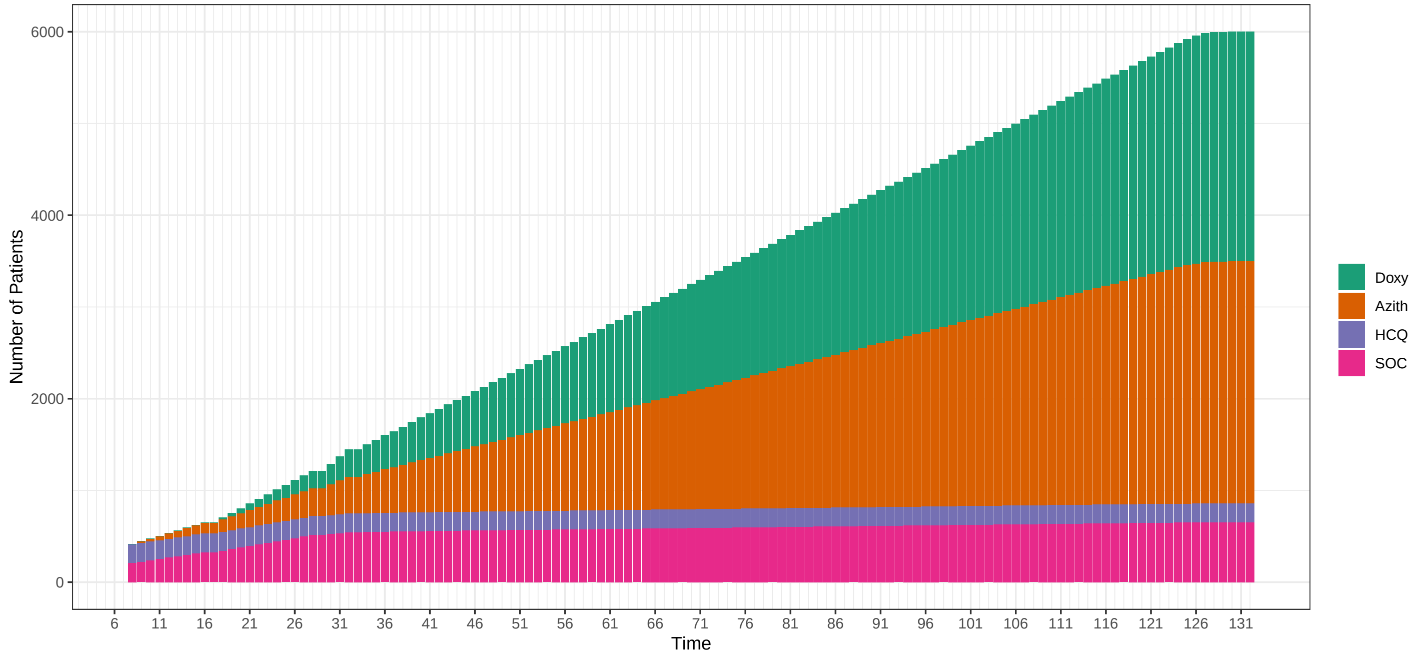

### Hazard Ratios: HCQ: 1.1 Azith: 1.1 Doxy: 1.1

#### Decisions Across Time

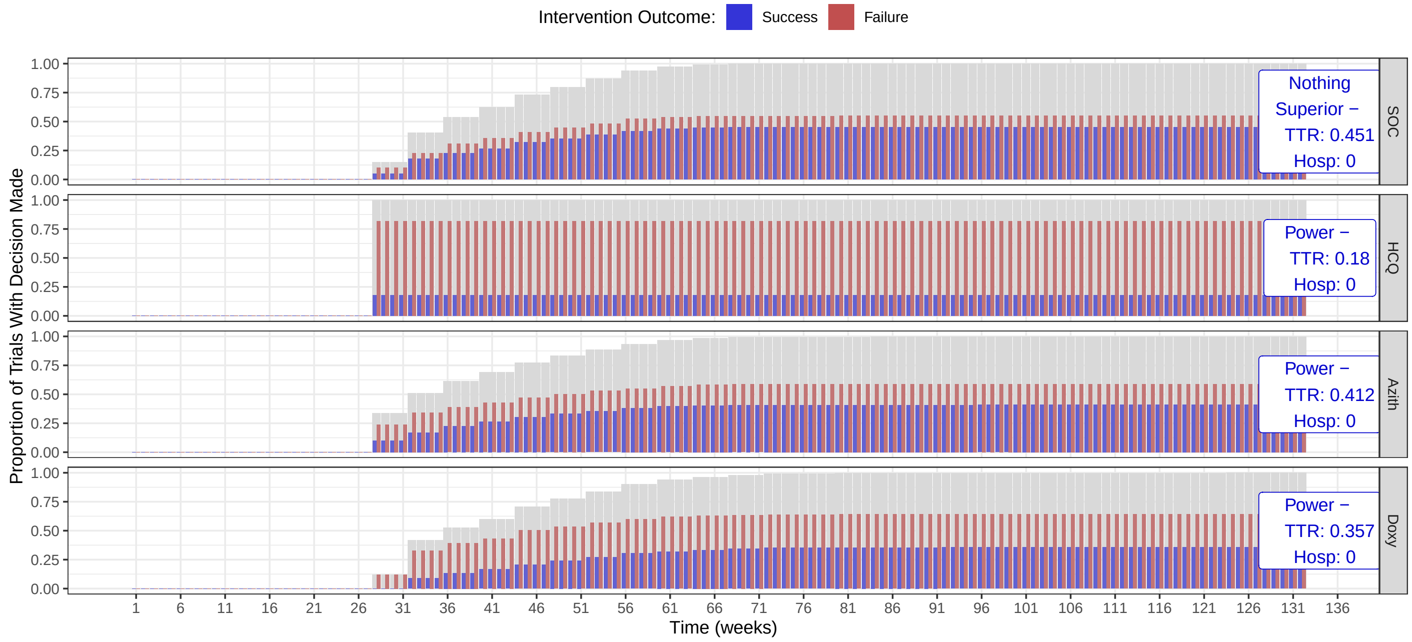

#### Randomization Rates

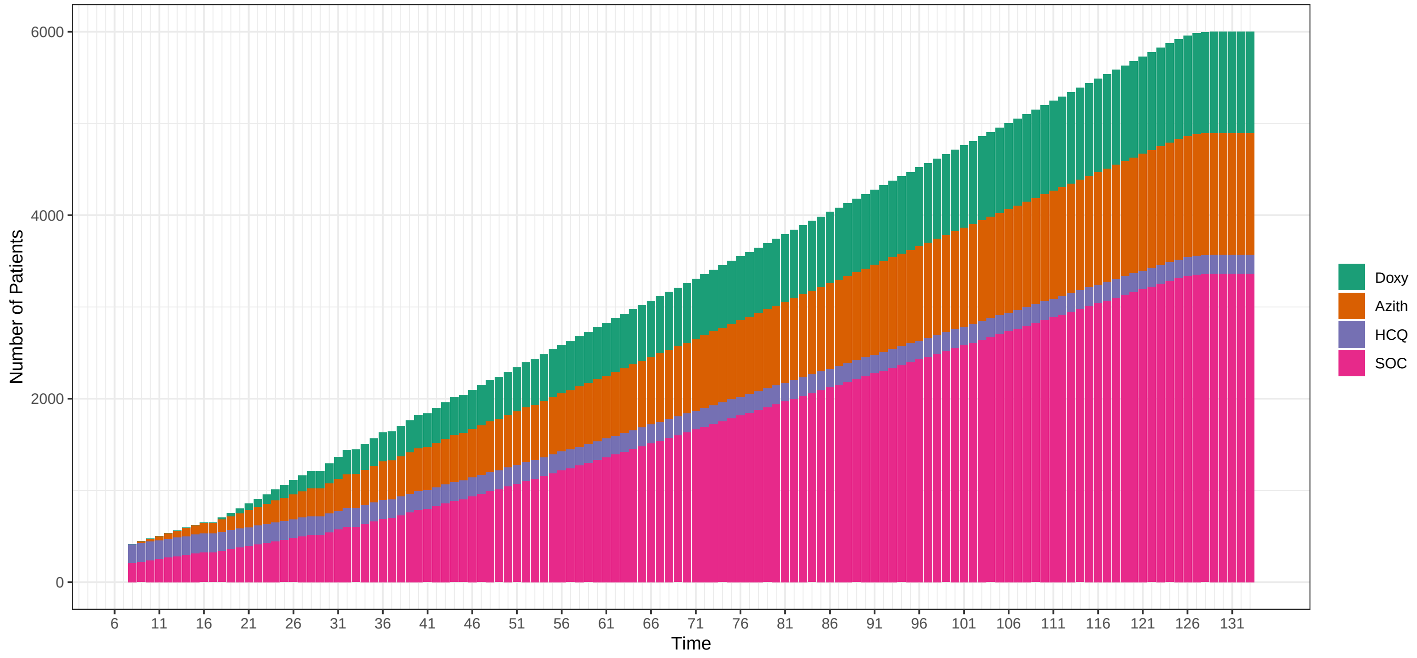

### Hazard Ratios: HCQ: 1.2 Azith: 1.2 Doxy: 1.2

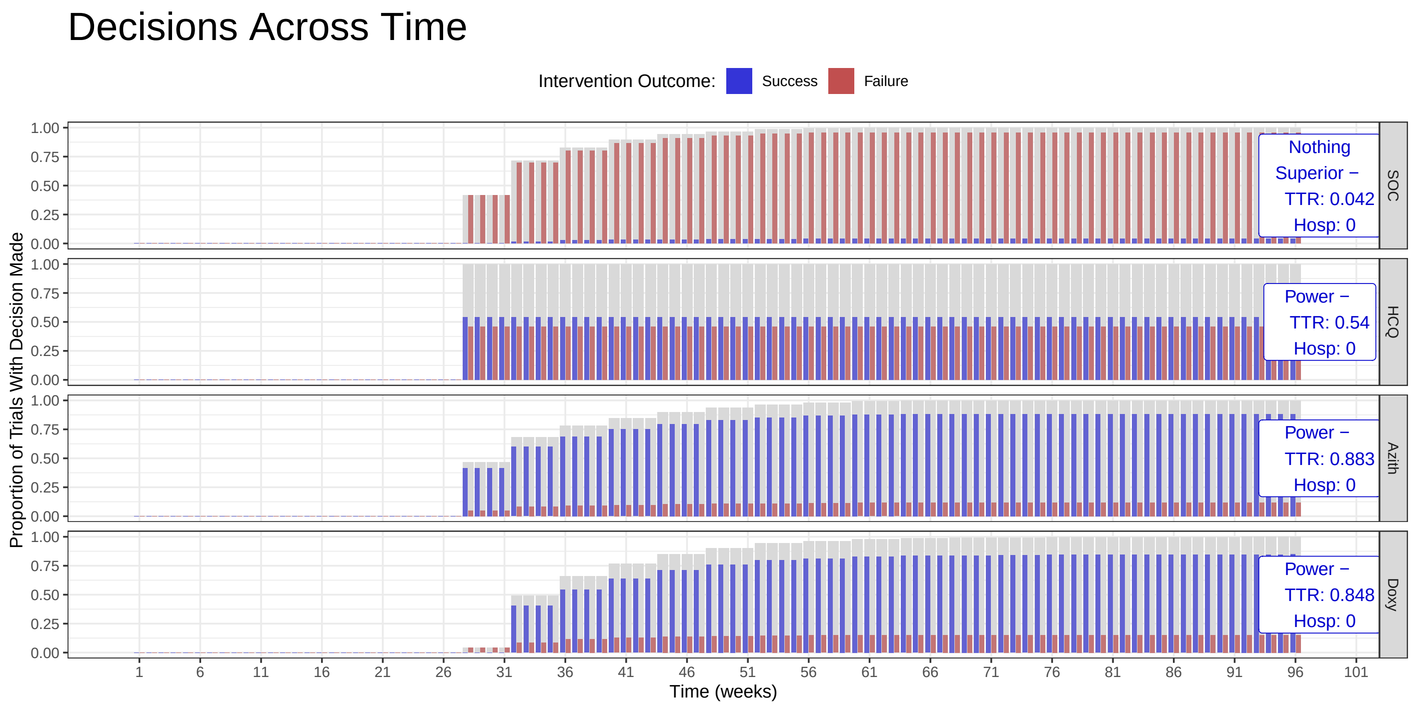

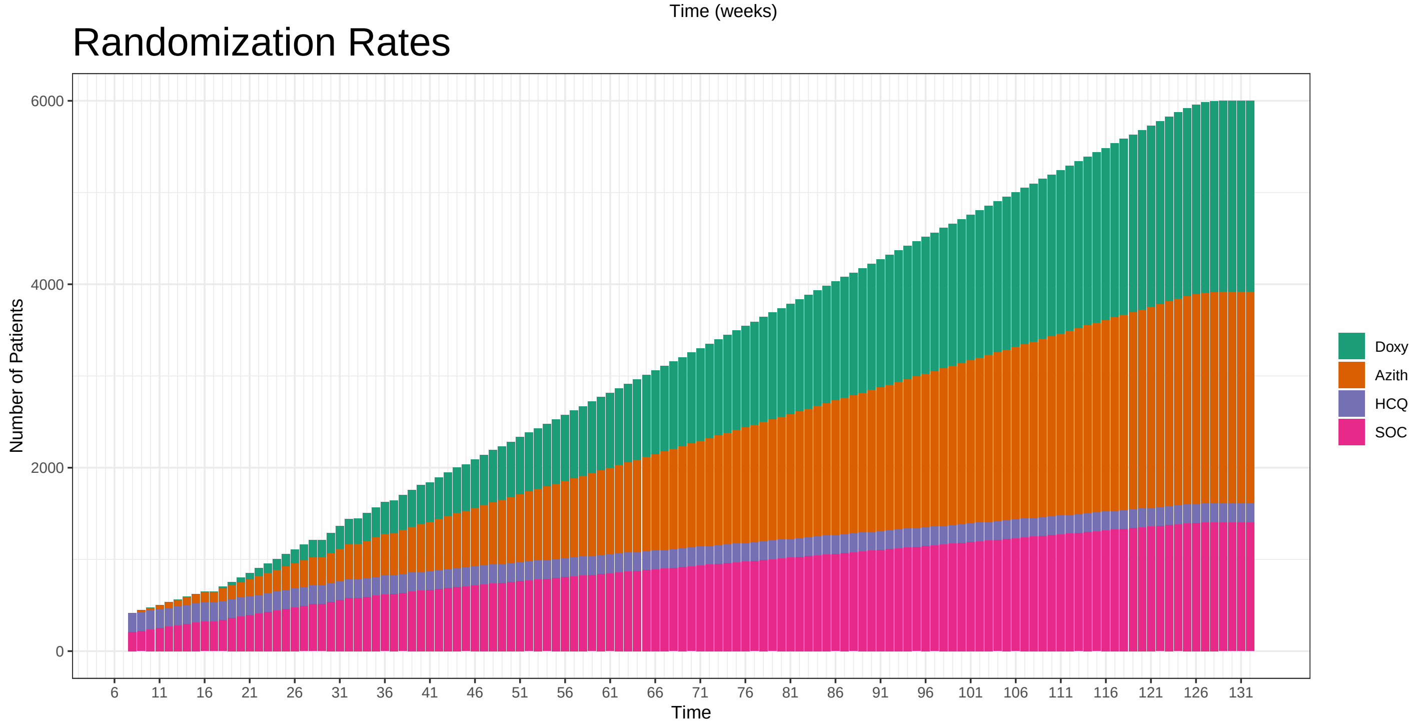

#### Hazard Ratios: HCQ: 1.3 Azith: 1.3 Doxy: 1.3

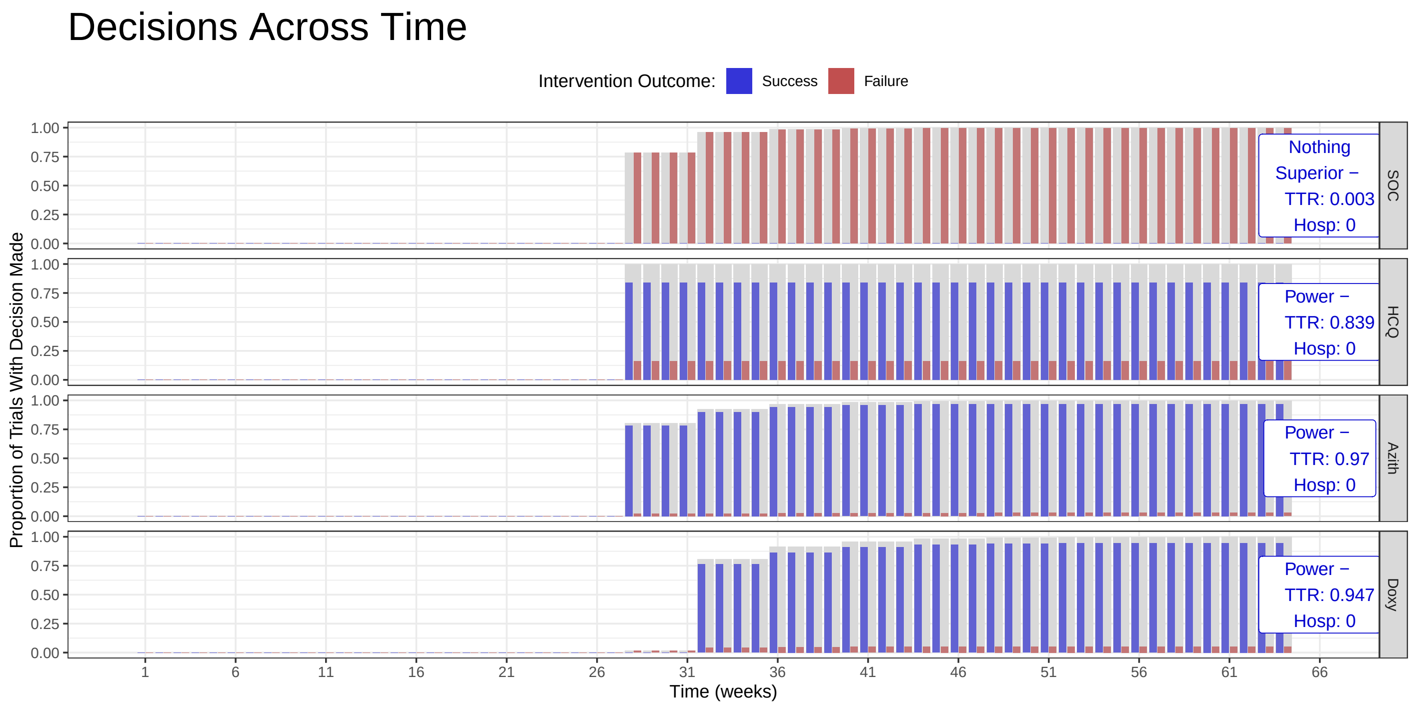

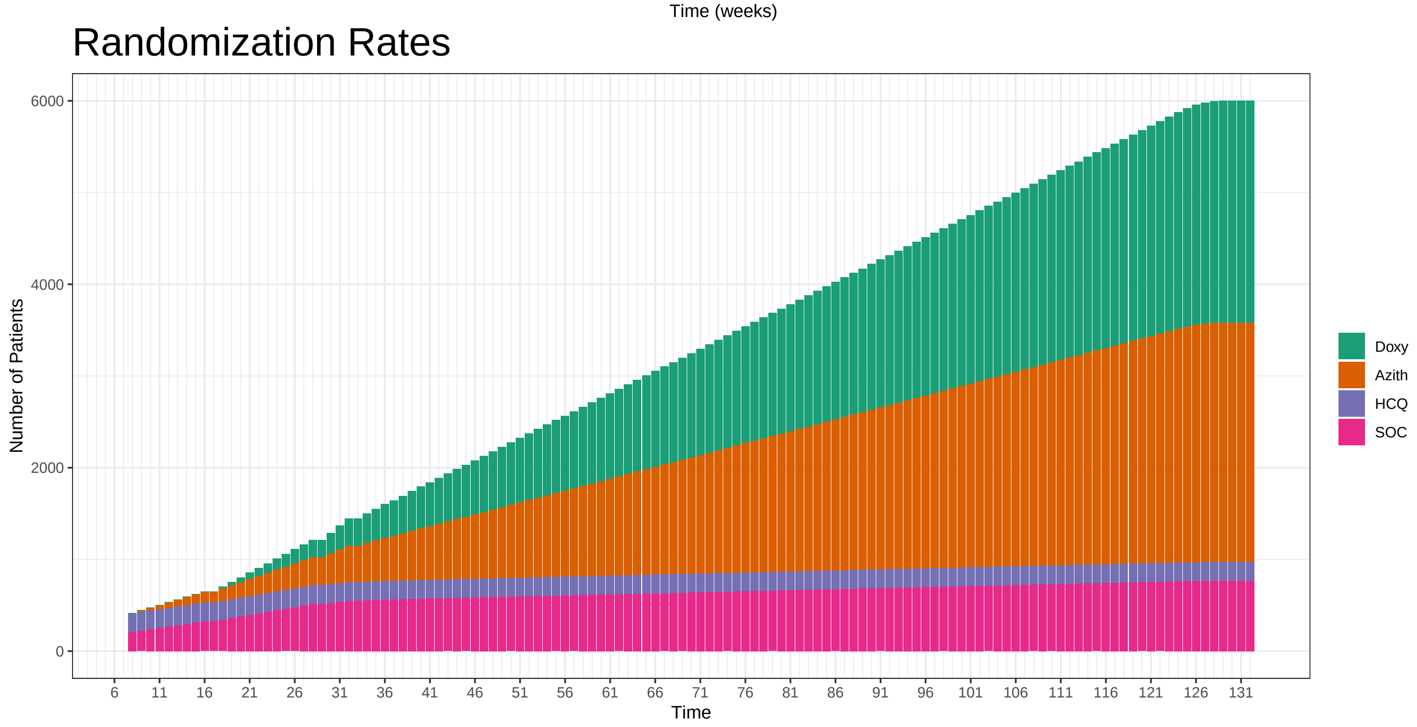

#### 5.3 OCs Additional Scenarios: Null Effect Hospitalization

### Principle OCs

Berry Consultants

10/26/2020

These operating characteristics are based on 1000 simulations per scenario across a variety of assumed treatment effects for experimental arms in the trial. The true number of patients randomized per day from April 1, 2020 to October 19, 2020. After this period is over, the simulations assume a 50 patient per week accrual rate. In absence of a definitive rule, these simulations conduct an interim every 28 days. In addition, the true day of enrollment stopping of HCQ is used as well as the days for opening enrollment to the Azithromycin and Doxycycline arms. As such, up to at least October 19, 2020 (day 199) of the trial, the enrollment rates to each arm in these simulations are expected to align closely with the true enrollment rates to the arms in the trial.

| Median TTR | Hazard Ratio | | | Power for TTR | | | | Hospitalization Rate Benefit (%) | | | Power for TTR & Hosp | | |
|---|---|---|---|---|---|---|---|---|---|---|---|---|---|
| SOC | HCQ | Azith | Doxy | SOC | HCQ | Azith | Doxy | HCQ | Azith | Doxy | HCQ | Azith | Doxy |
| 9 | 1.1 | 1.1 | 1.1 | 0.416 | 0.179 | 0.428 | 0.398 | 0 | 0 | 0 | 0.002 | 0.008 | 0.012 |
| 9 | 1.2 | 1.2 | 1.2 | 0.035 | 0.540 | 0.879 | 0.857 | 0 | 0 | 0 | 0.008 | 0.019 | 0.016 |
| 9 | 1.3 | 1.3 | 1.3 | 0.002 | 0.839 | 0.975 | 0.959 | 0 | 0 | 0 | 0.012 | 0.019 | 0.029 |

Decisions on Time to Recovery Benefit By Time

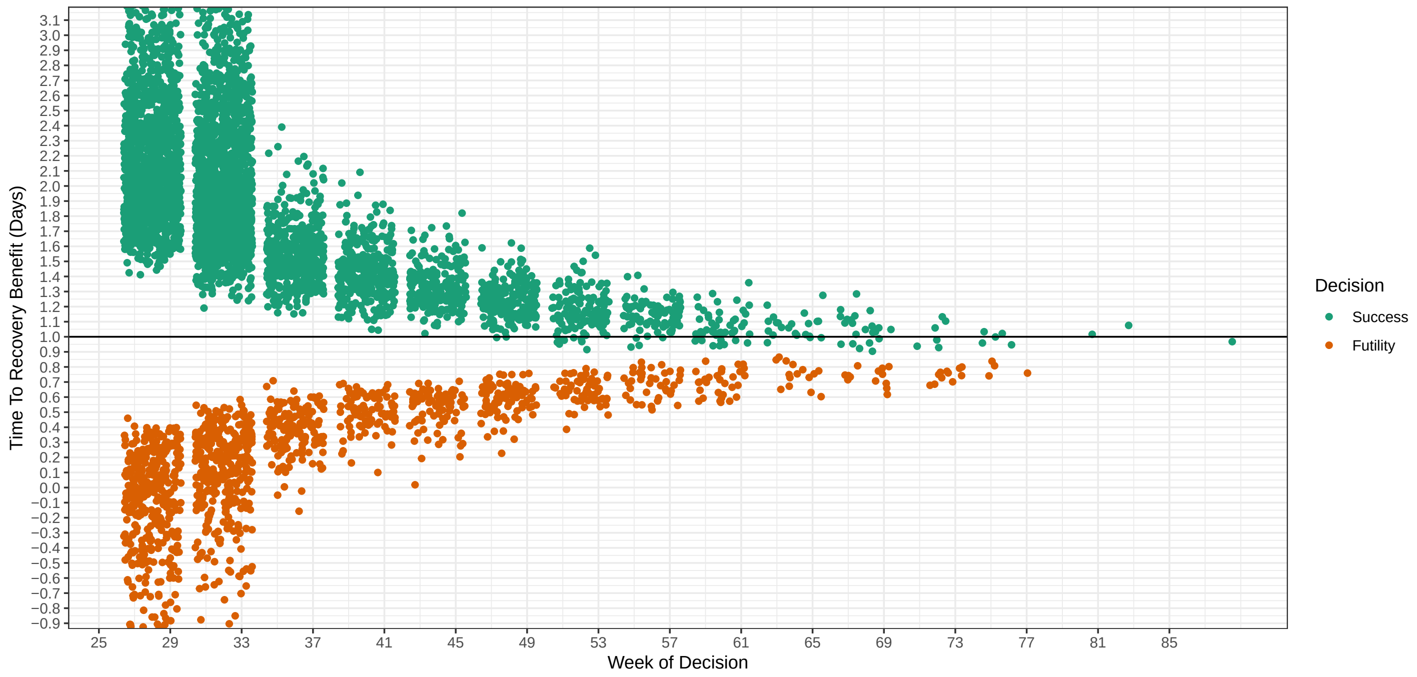

### Hazard Ratios: HCQ: 1.1 Azith: 1.1 Doxy: 1.1

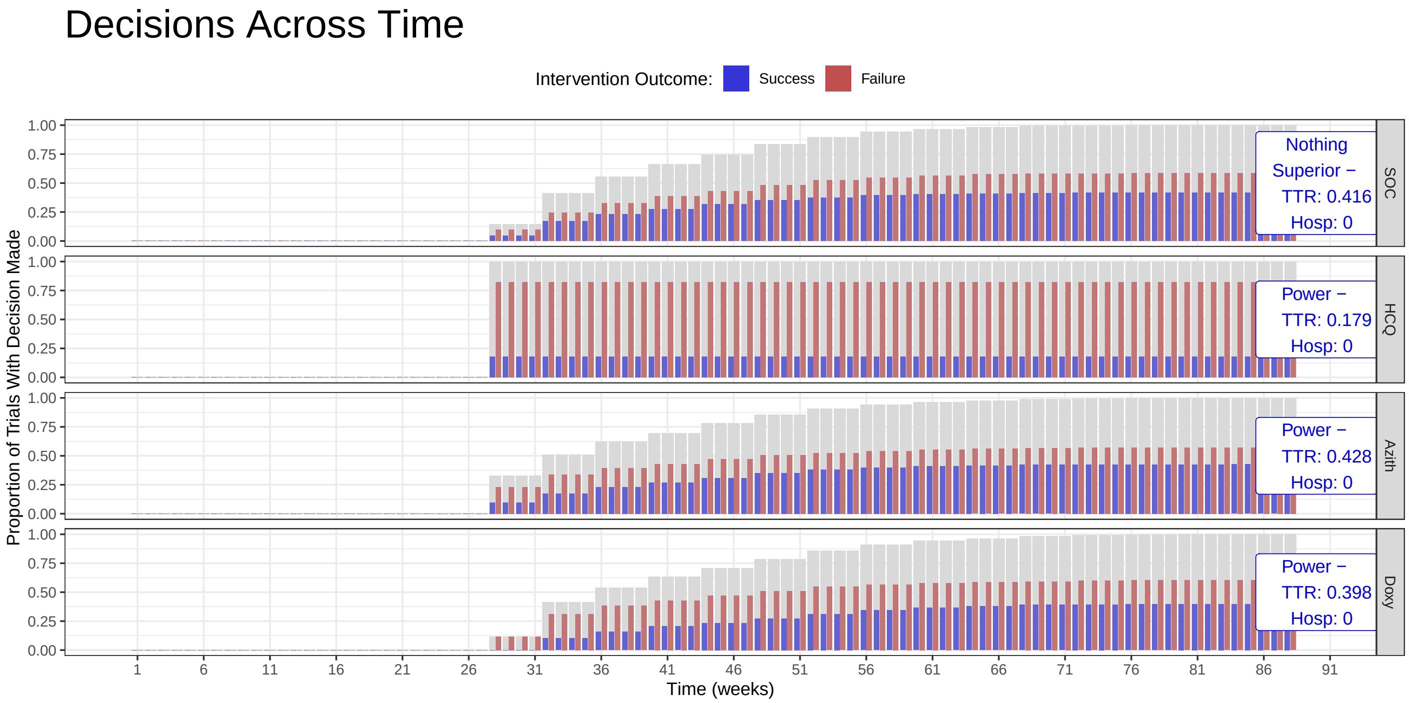

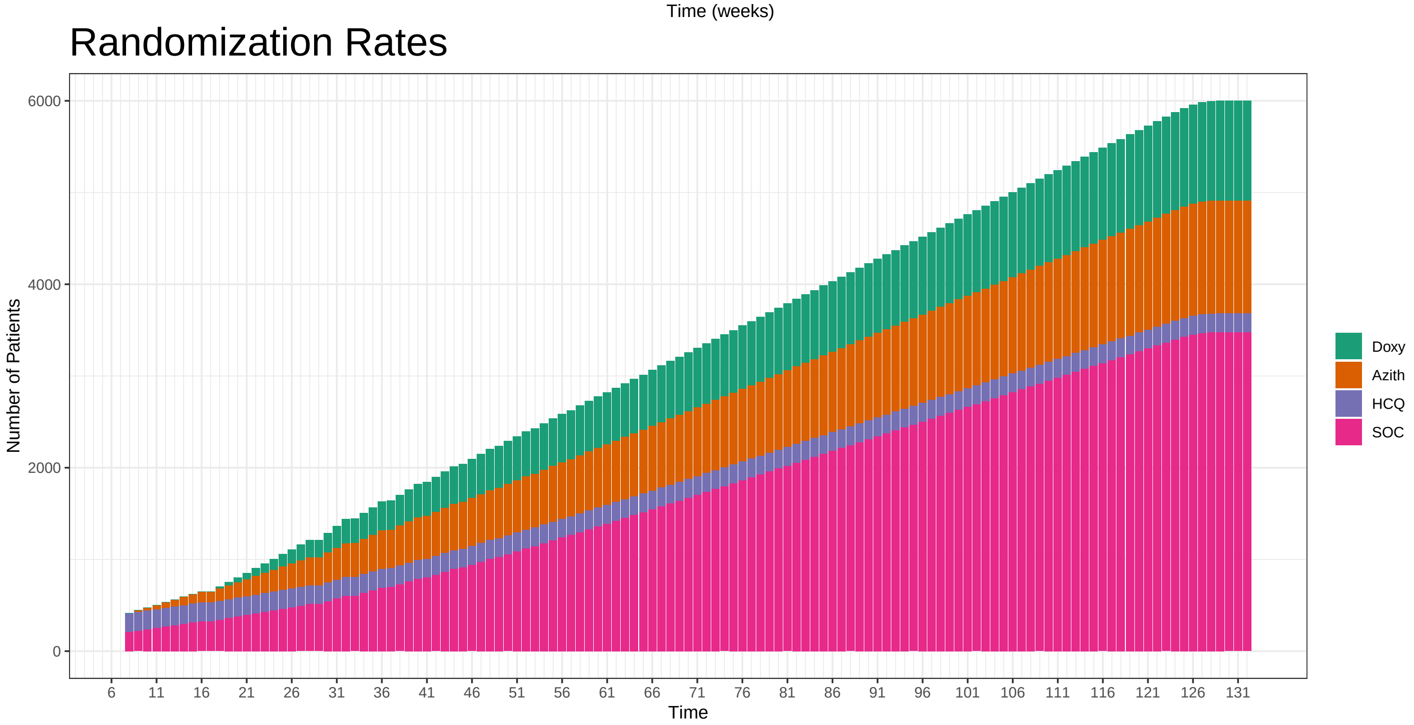

### Hazard Ratios: HCQ: 1.2 Azith: 1.2 Doxy: 1.2

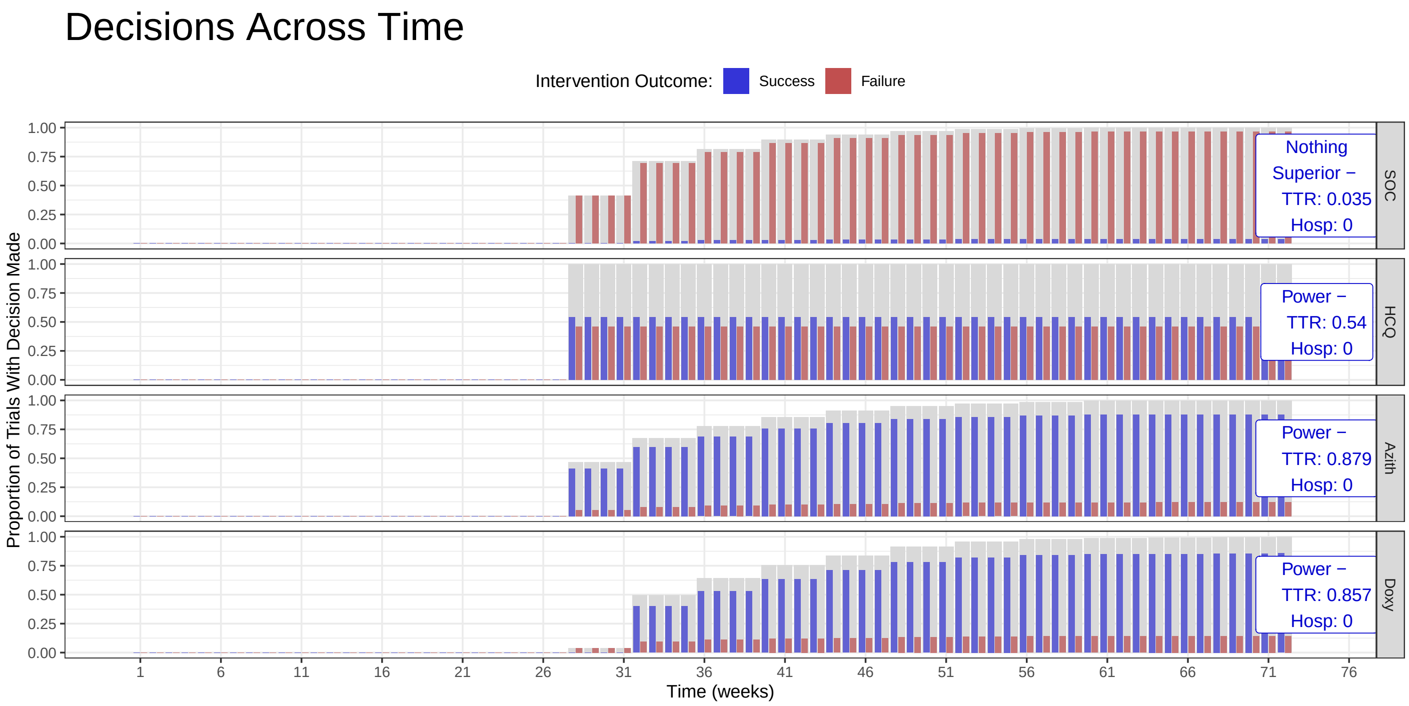

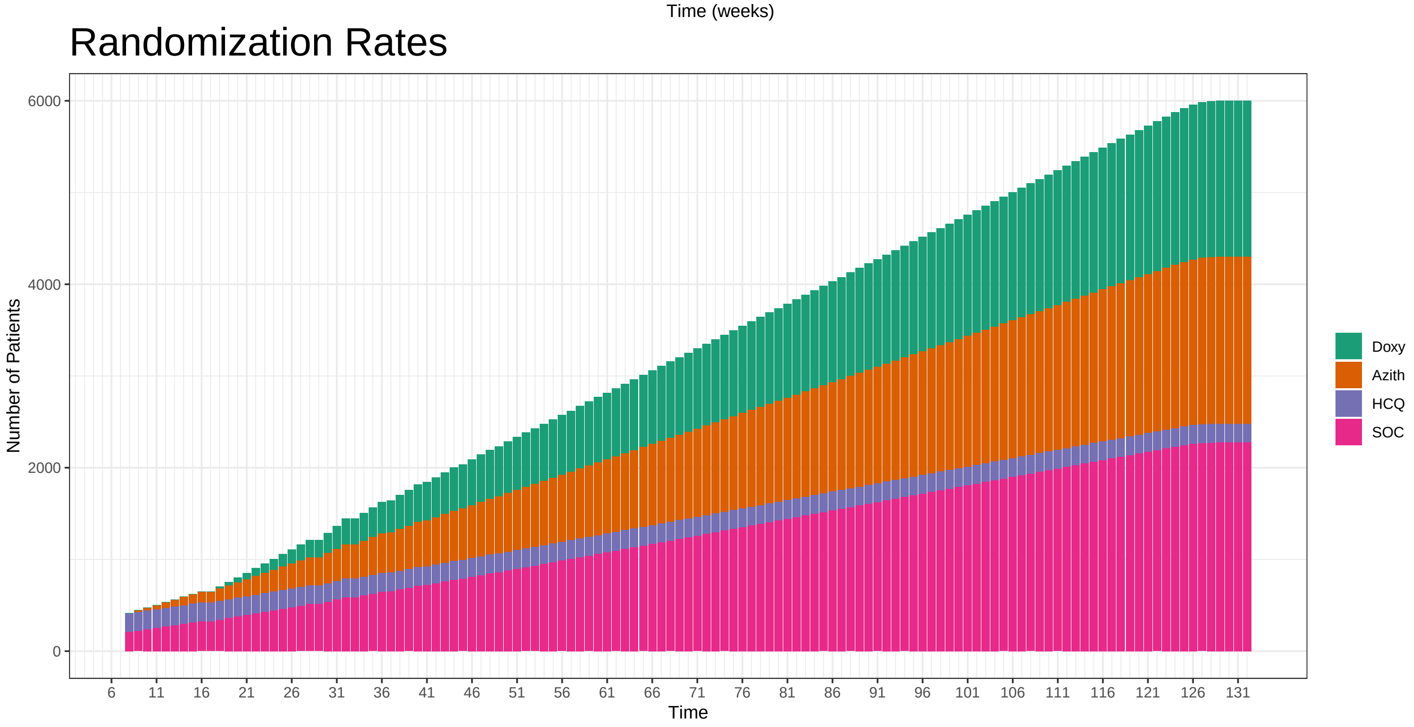

#### Hazard Ratios: HCQ: 1.3 Azith: 1.3 Doxy: 1.3

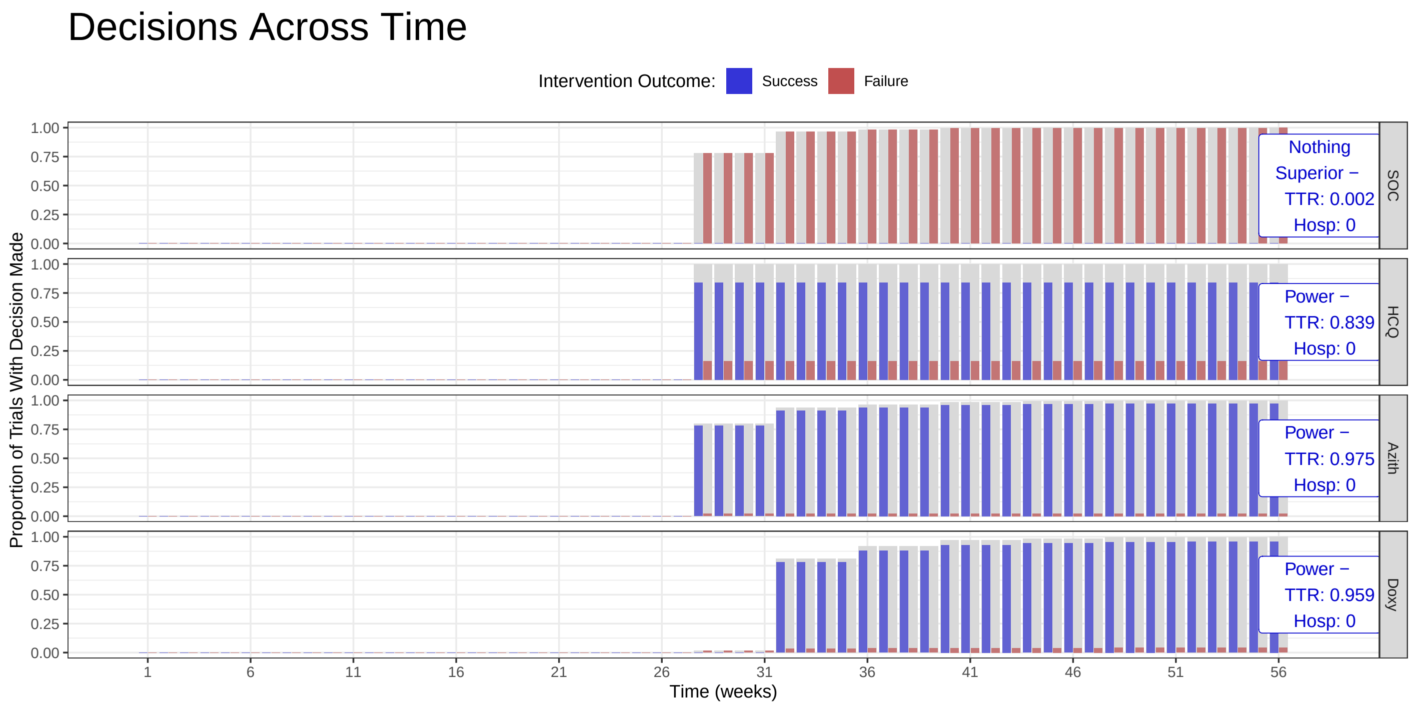

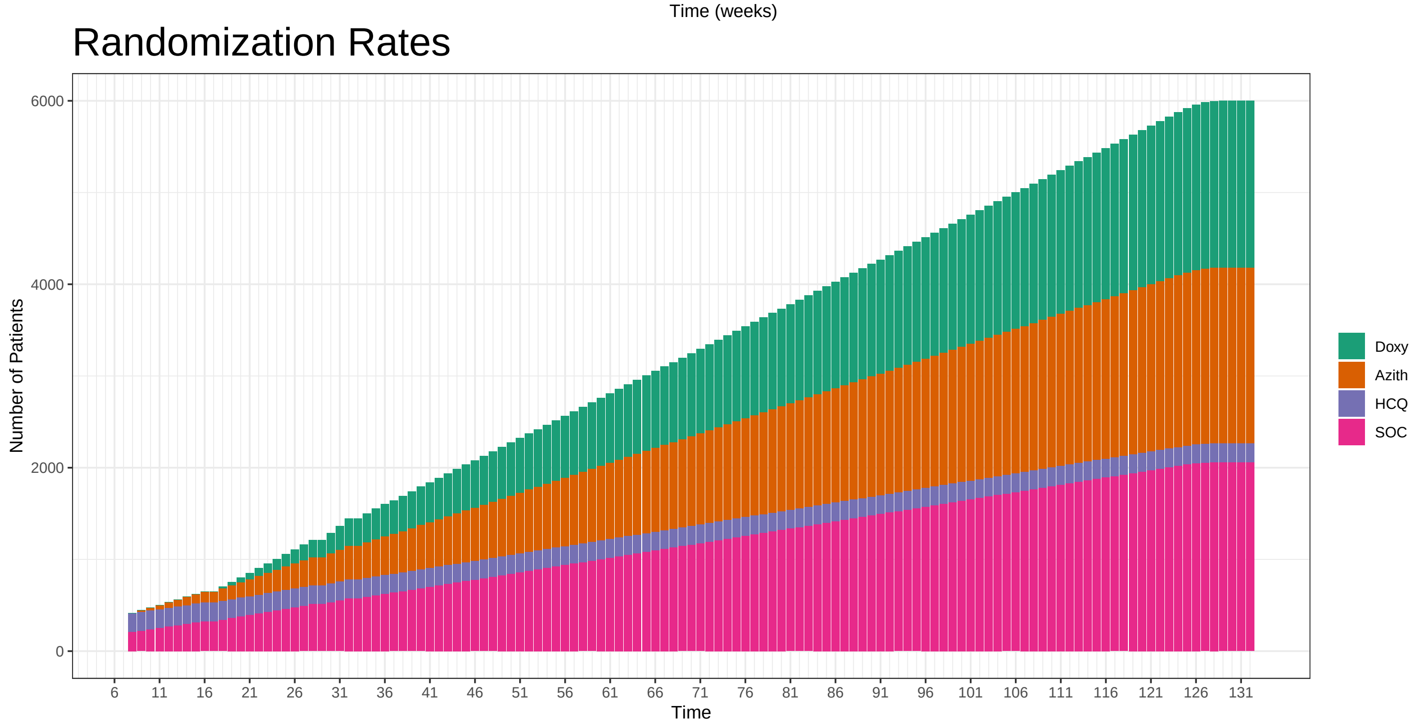

#### 5.4 OCs Additional Scenarios: Modified Time to Recovery Rate for Usual Care Arm

### Principle OCs

Berry Consultants

10/26/2020

These operating characteristics are based on 1000 simulations per scenario across a variety of assumed treatment effects for experimental arms in the trial. The true number of patients randomized per day from April 1, 2020 to October 19, 2020. After this period is over, the simulations assume a 50 patient per week accrual rate. In absence of a definitive rule, these simulations conduct an interim every 28 days. In addition, the true day of enrollment stopping of HCQ is used as well as the days for opening enrollment to the Azithromycin and Doxycycline arms. As such, up to at least October 19, 2020 (day 199) of the trial, the enrollment rates to each arm in these simulations are expected to align closely with the true enrollment rates to the arms in the trial.

| Median TTR | Hazard Ratio | | | Power for TTR | | | | Hospitalization Rate Benefit (%) | | | Power for TTR & Hosp | | |
|---|---|---|---|---|---|---|---|---|---|---|---|---|---|
| SOC | HCQ | Azith | Doxy | SOC | HCQ | Azith | Doxy | HCQ | Azith | Doxy | HCQ | Azith | Doxy |
| 7 | 1 | 1 | 1.0 | 0.979 | 0.009 | 0.012 | 0.010 | 0 | 0 | 0 | 0 | 0 | 0.000 |
| 7 | 1 | 1 | 1.1 | 0.768 | 0.009 | 0.019 | 0.221 | 0 | 0 | 1 | 0 | 0 | 0.033 |
| 7 | 1 | 1 | 1.2 | 0.317 | 0.011 | 0.010 | 0.683 | 0 | 0 | 2 | 0 | 0 | 0.343 |
| 7 | 1 | 1 | 1.3 | 0.059 | 0.007 | 0.007 | 0.941 | 0 | 0 | 3 | 0 | 0 | 0.905 |
| 11 | 1 | 1 | 1.0 | 0.940 | 0.017 | 0.036 | 0.030 | 0 | 0 | 0 | 0 | 0 | 0.000 |
| 11 | 1 | 1 | 1.1 | 0.467 | 0.015 | 0.038 | 0.522 | 0 | 0 | 1 | 0 | 0 | 0.105 |
| 11 | 1 | 1 | 1.2 | 0.057 | 0.015 | 0.014 | 0.943 | 0 | 0 | 2 | 0 | 0 | 0.578 |
| 11 | 1 | 1 | 1.3 | 0.003 | 0.015 | 0.013 | 0.997 | 0 | 0 | 3 | 0 | 0 | 0.963 |

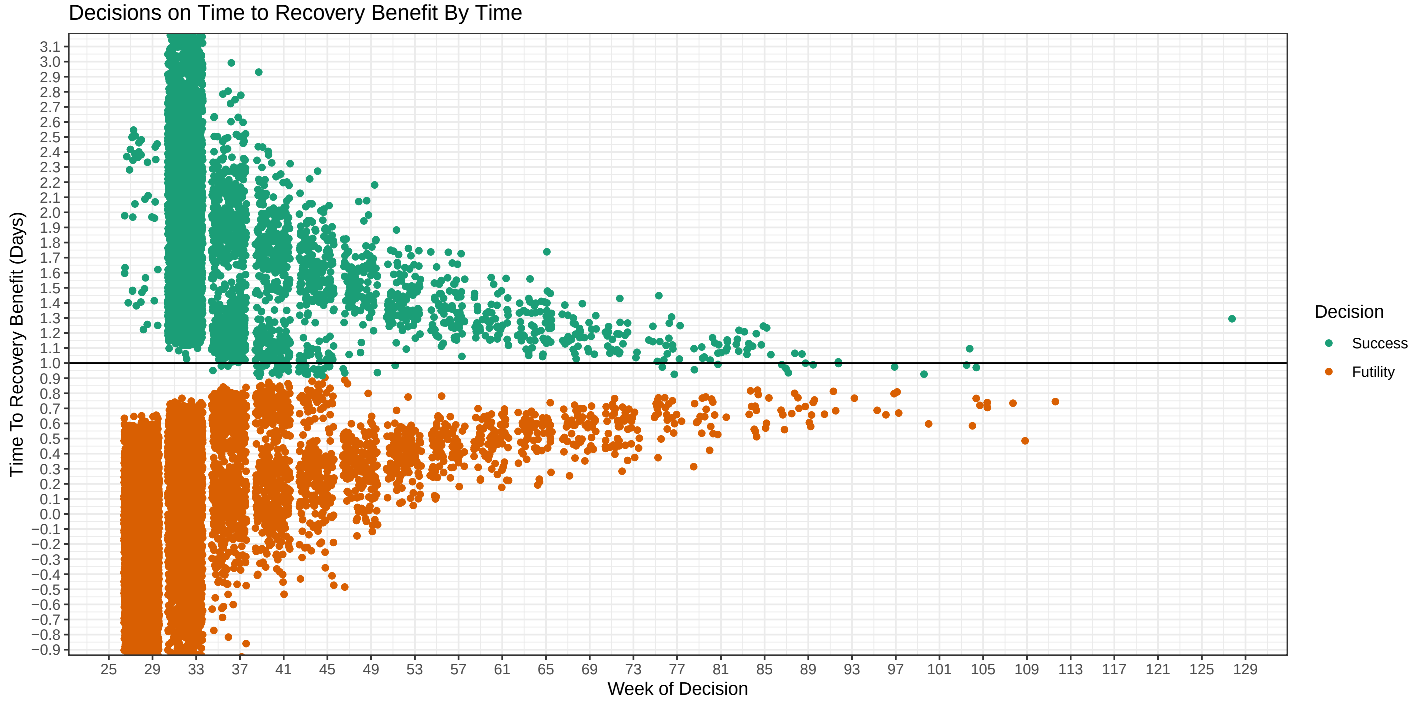

### Hazard Ratios: HCQ: 1 Azith: 1 Doxy: 1

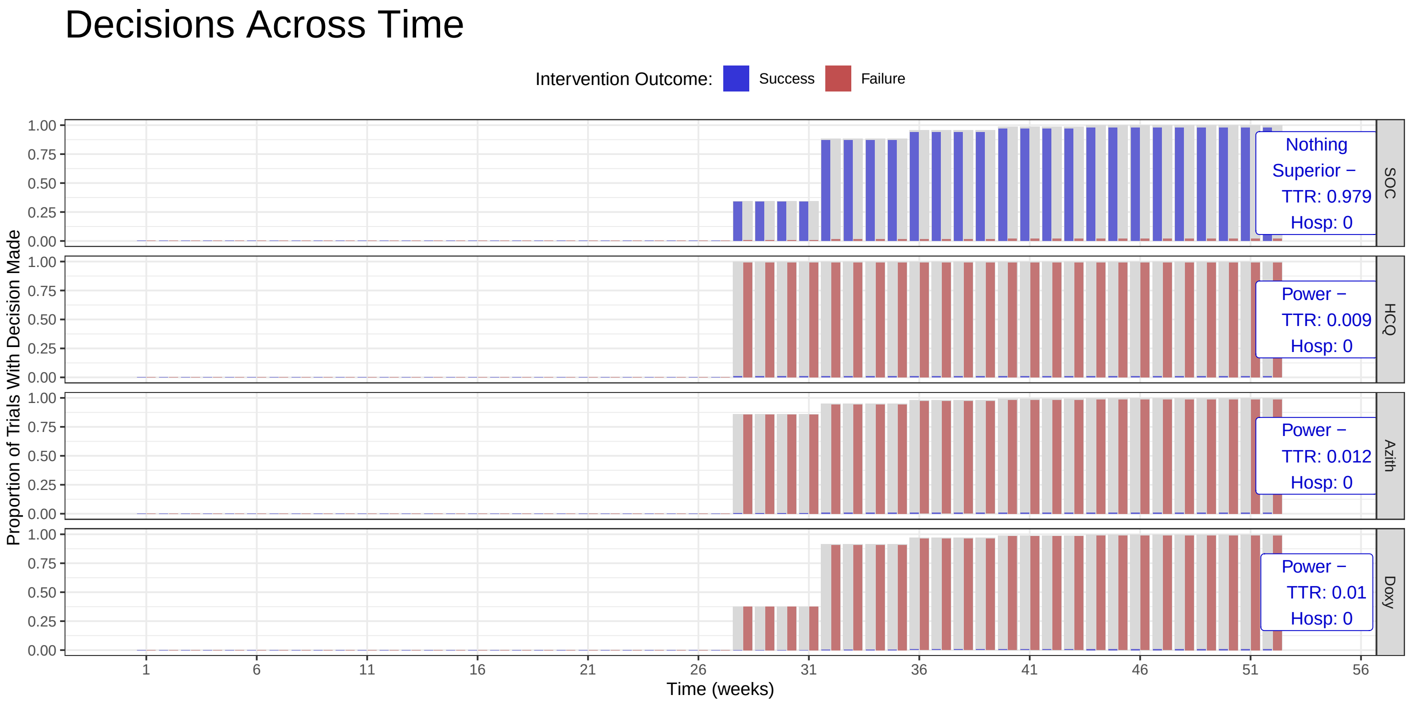

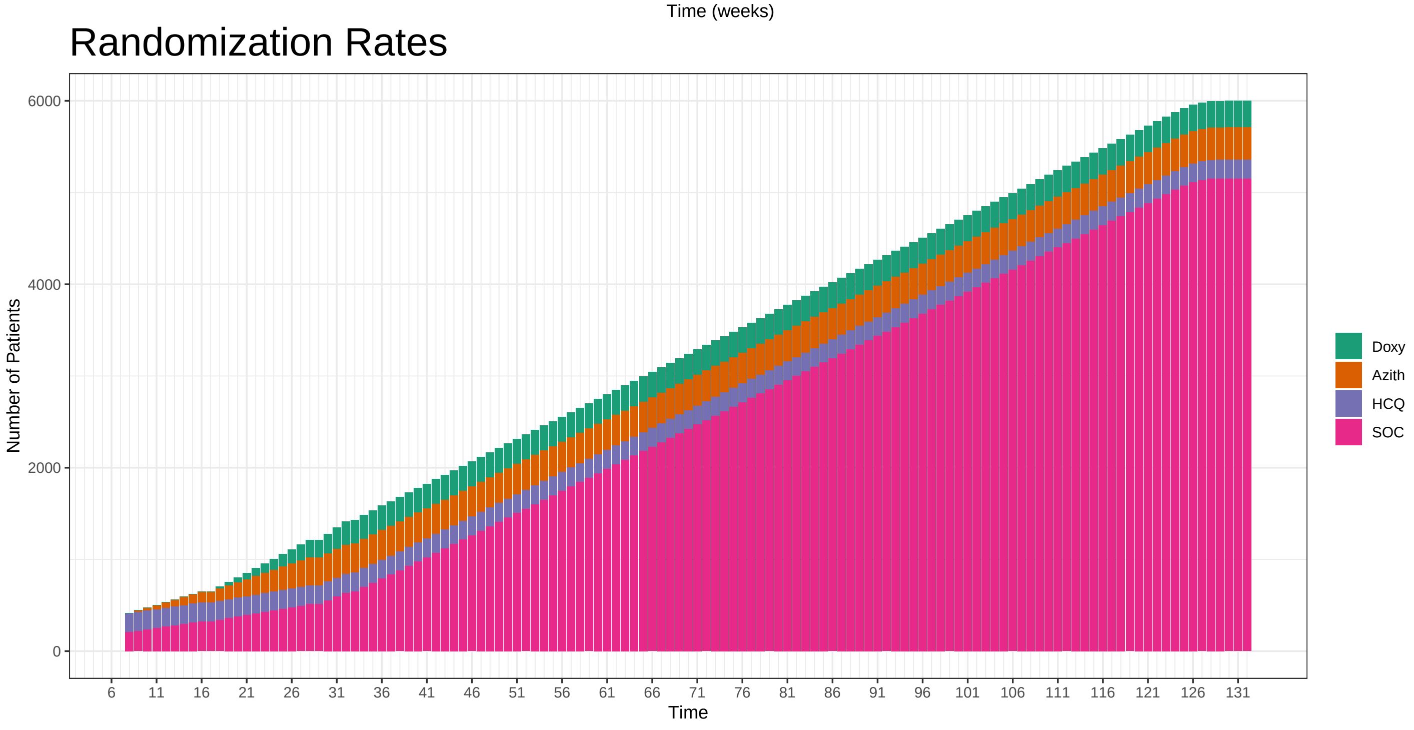

### Hazard Ratios: HCQ: 1 Azith: 1 Doxy: 1.1

#### Decisions Across Time

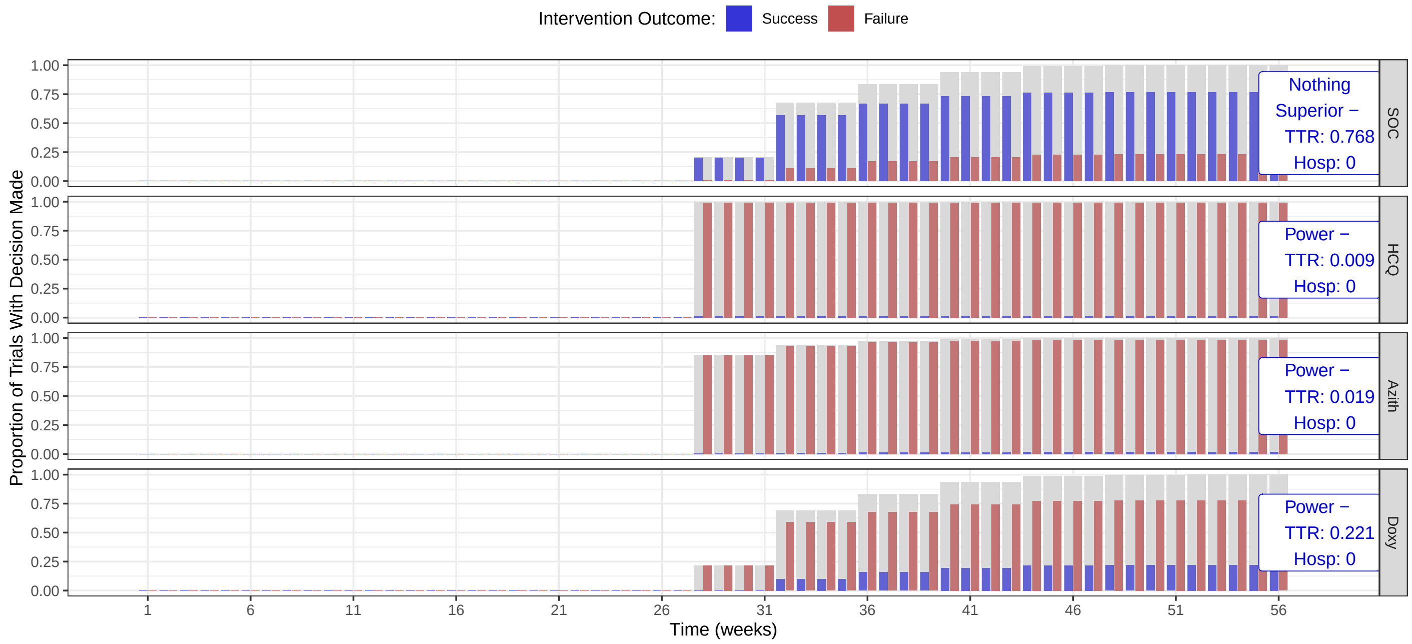

#### Randomization Rates

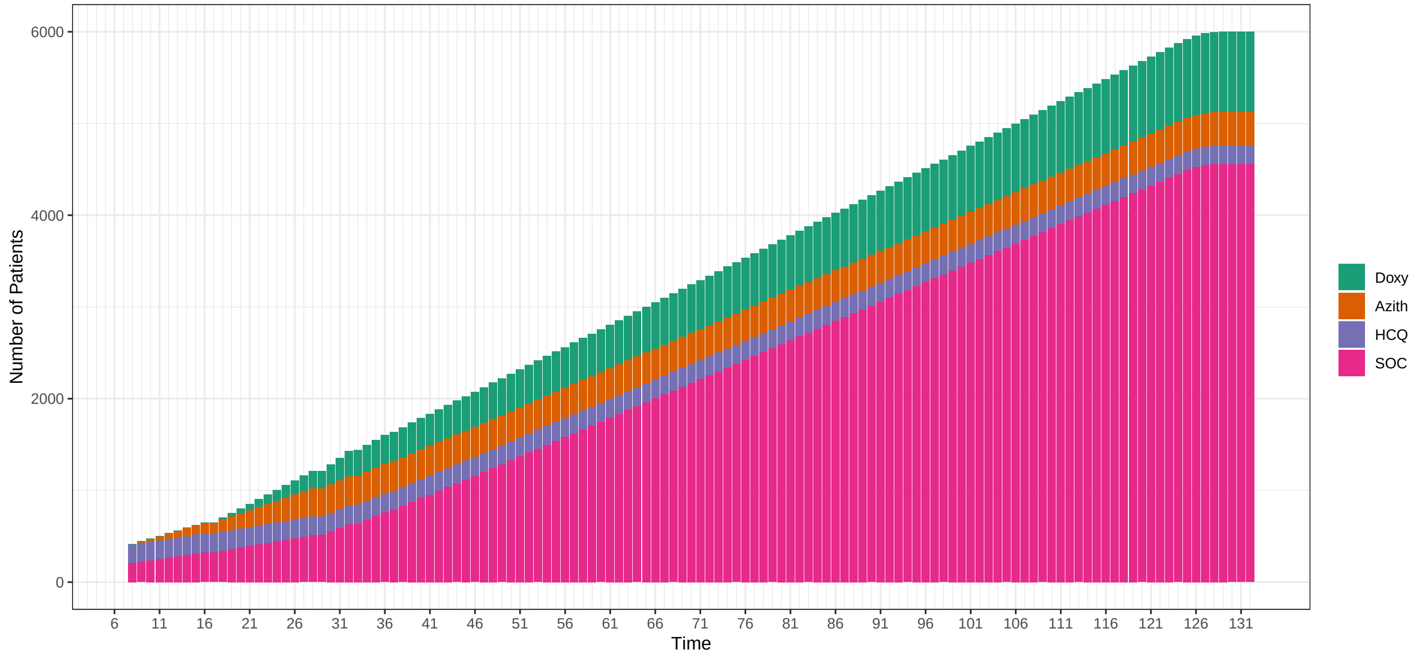

### Hazard Ratios: HCQ: 1 Azith: 1 Doxy: 1.2

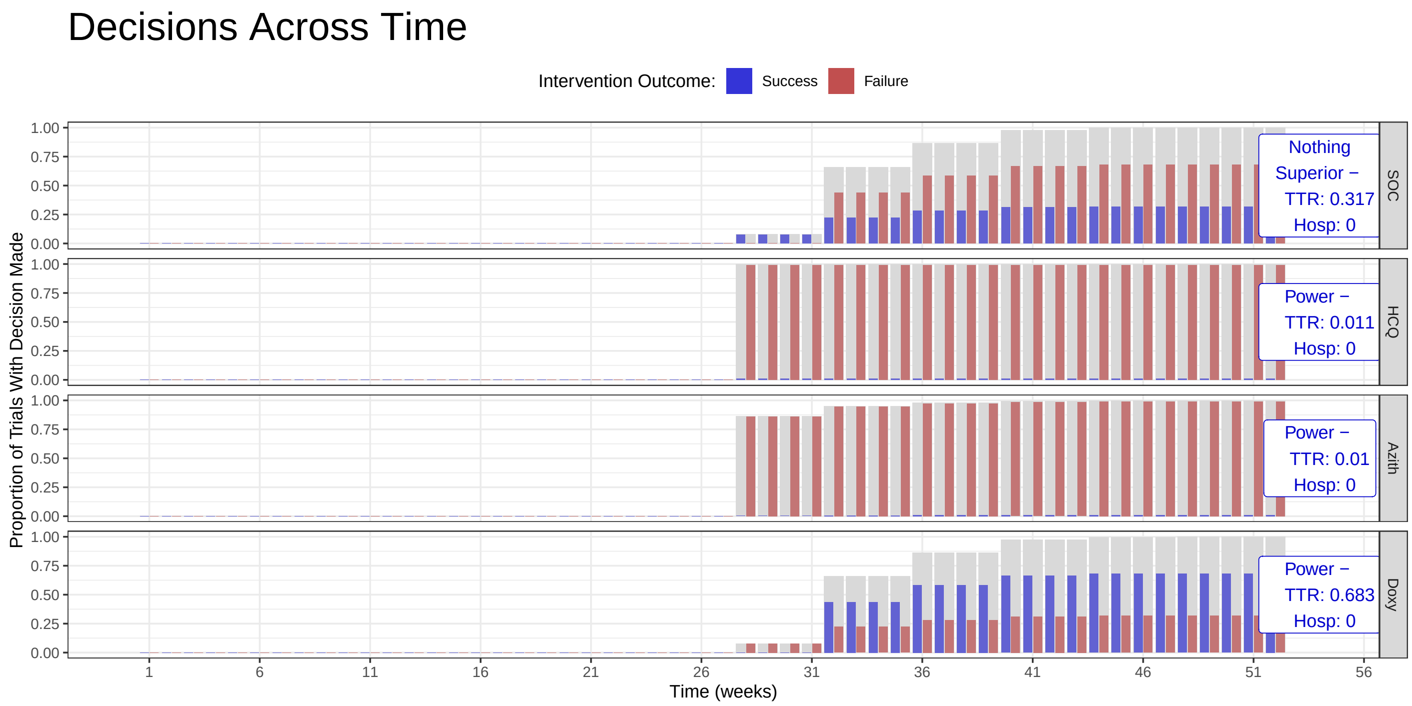

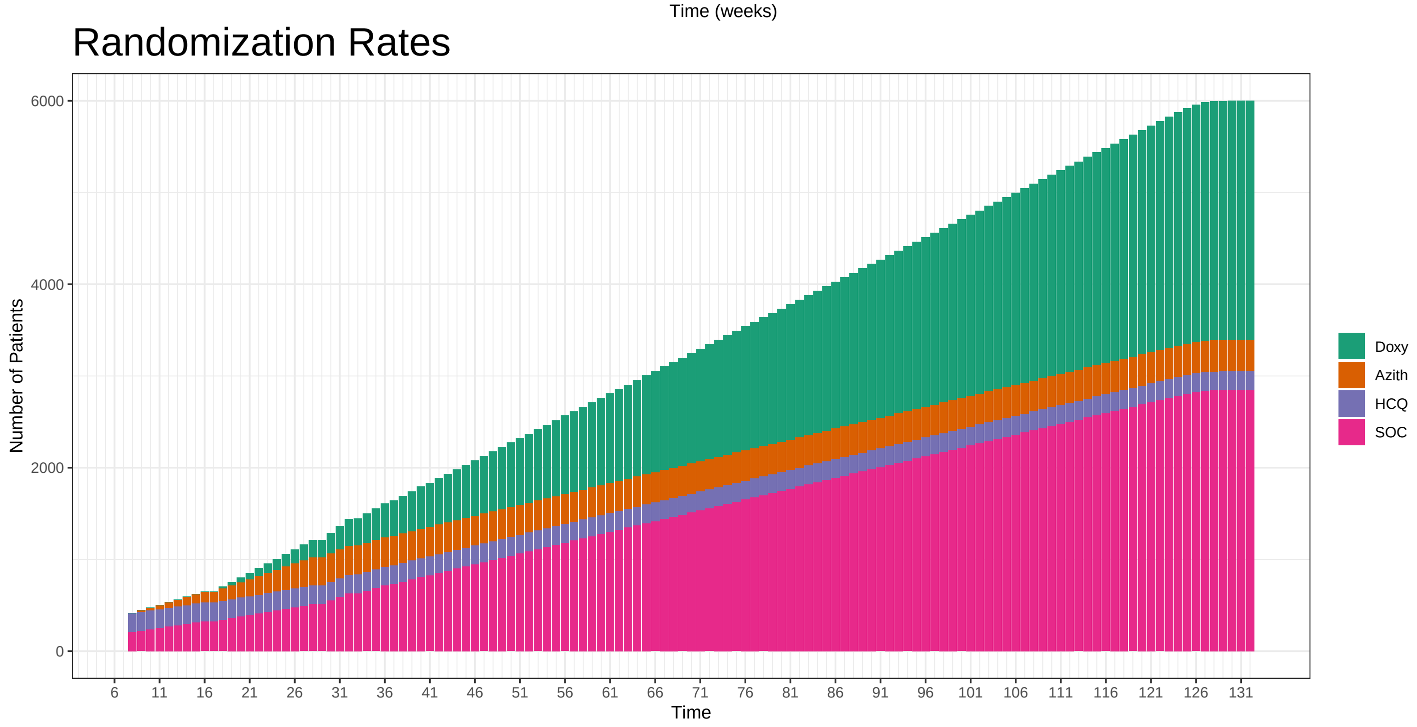

### Hazard Ratios: HCQ: 1 Azith: 1 Doxy: 1.3

#### Decisions Across Time

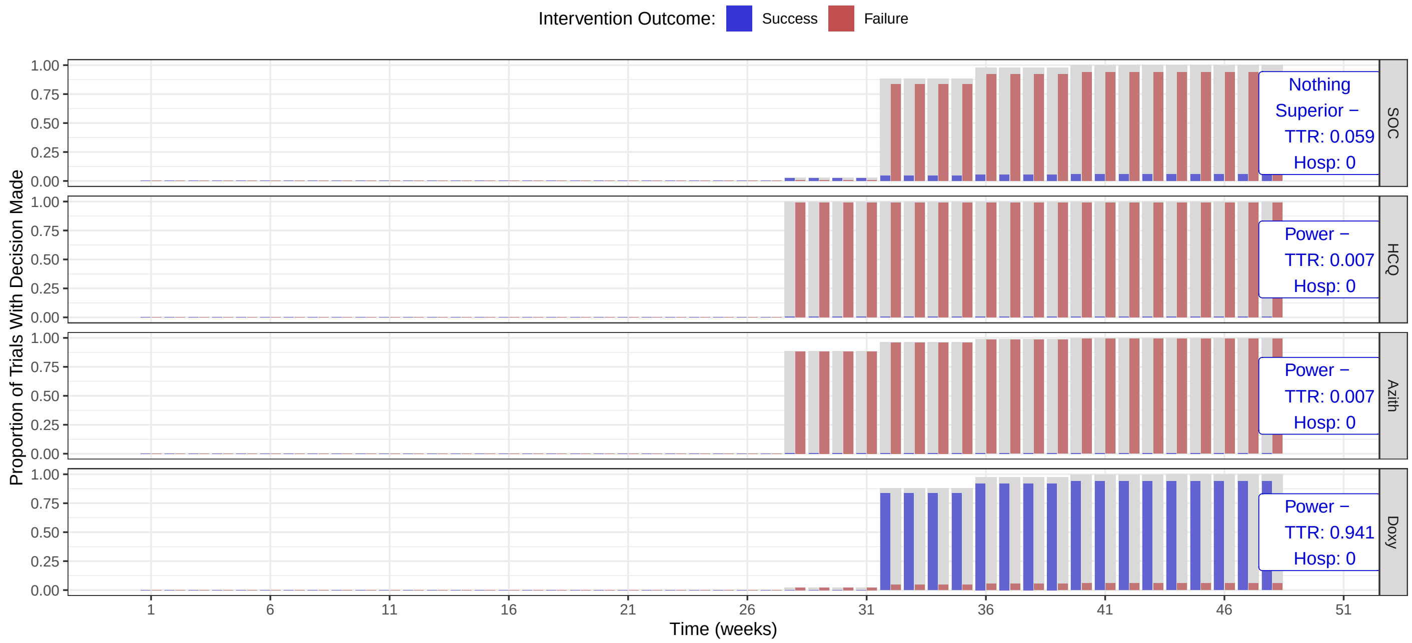

#### Randomization Rates

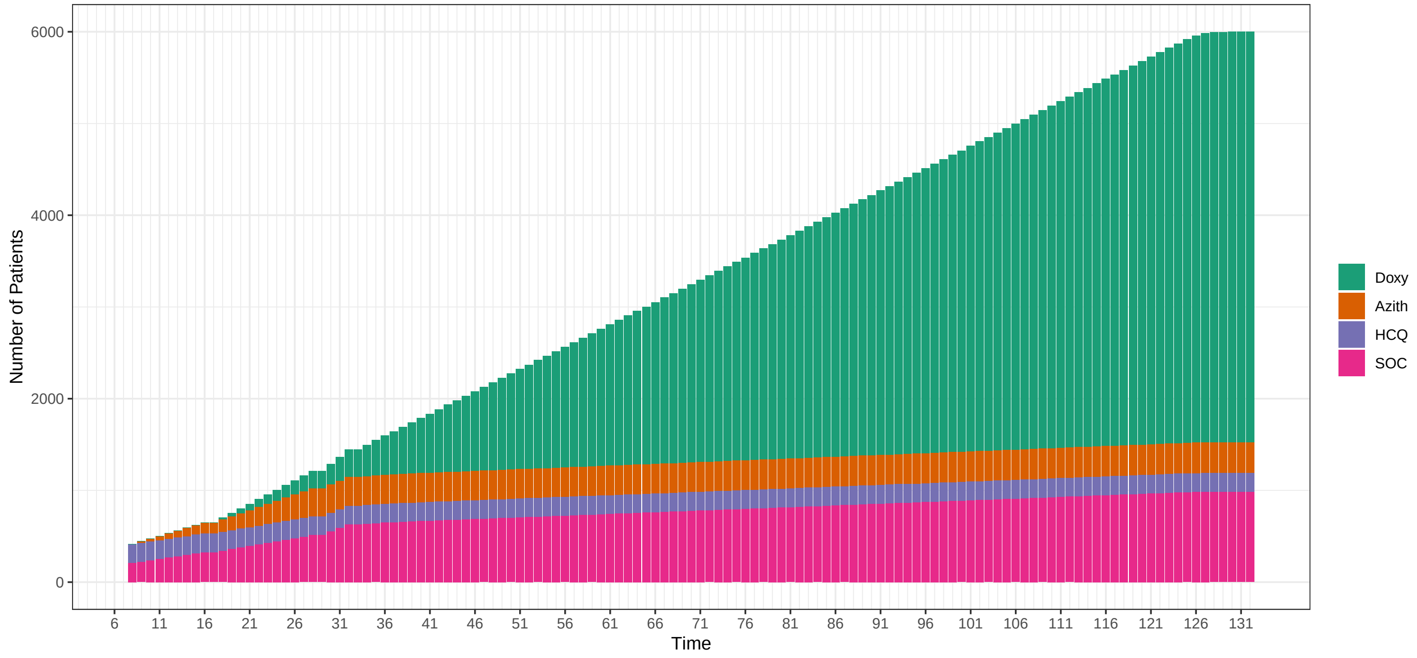

### Hazard Ratios: HCQ: 1 Azith: 1 Doxy: 1

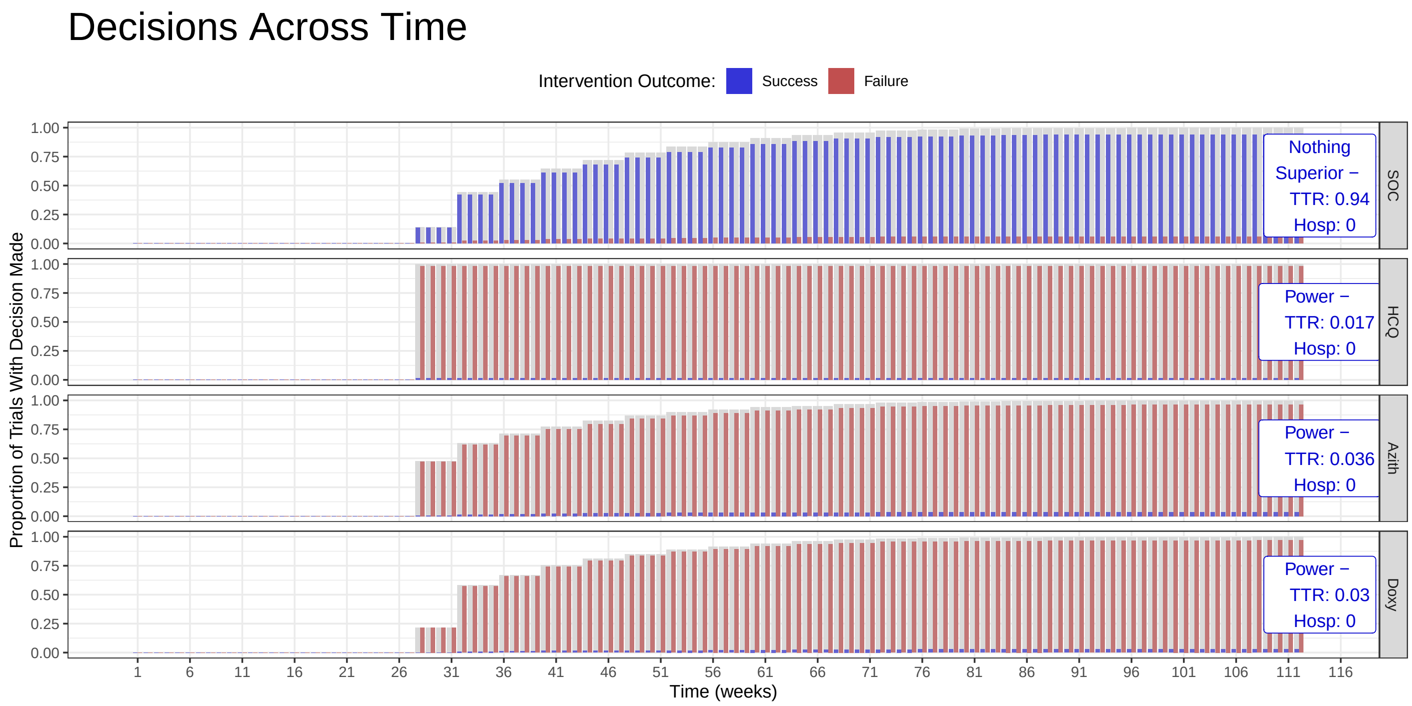

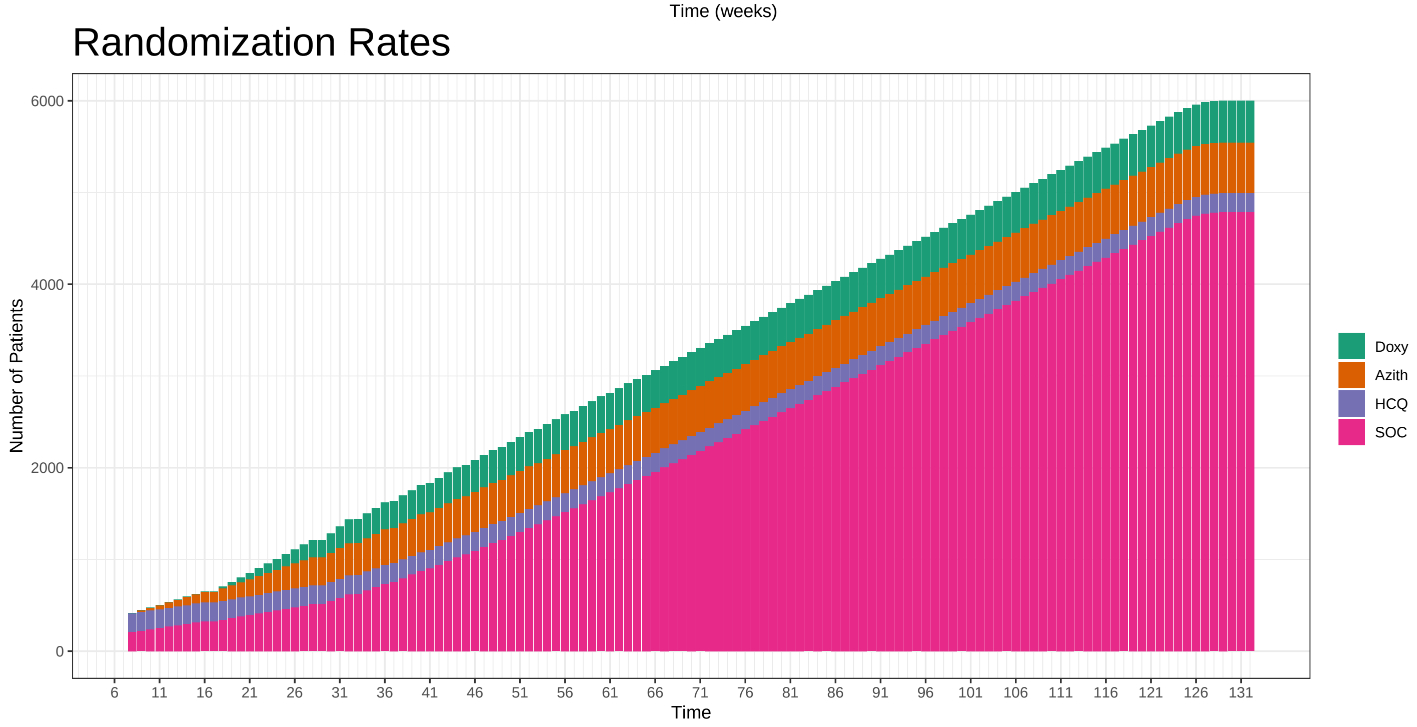

#### Hazard Ratios: HCQ: 1 Azith: 1 Doxy: 1.1

##### Decisions Across Time

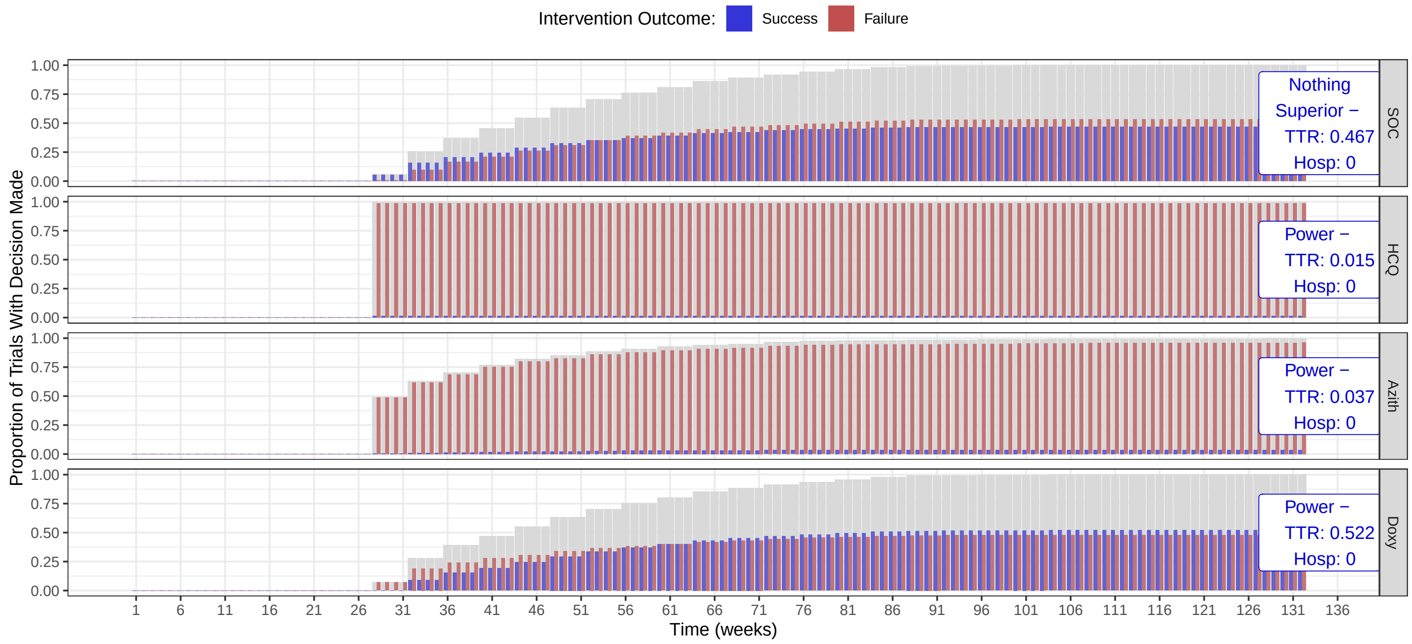

##### Randomization Rates

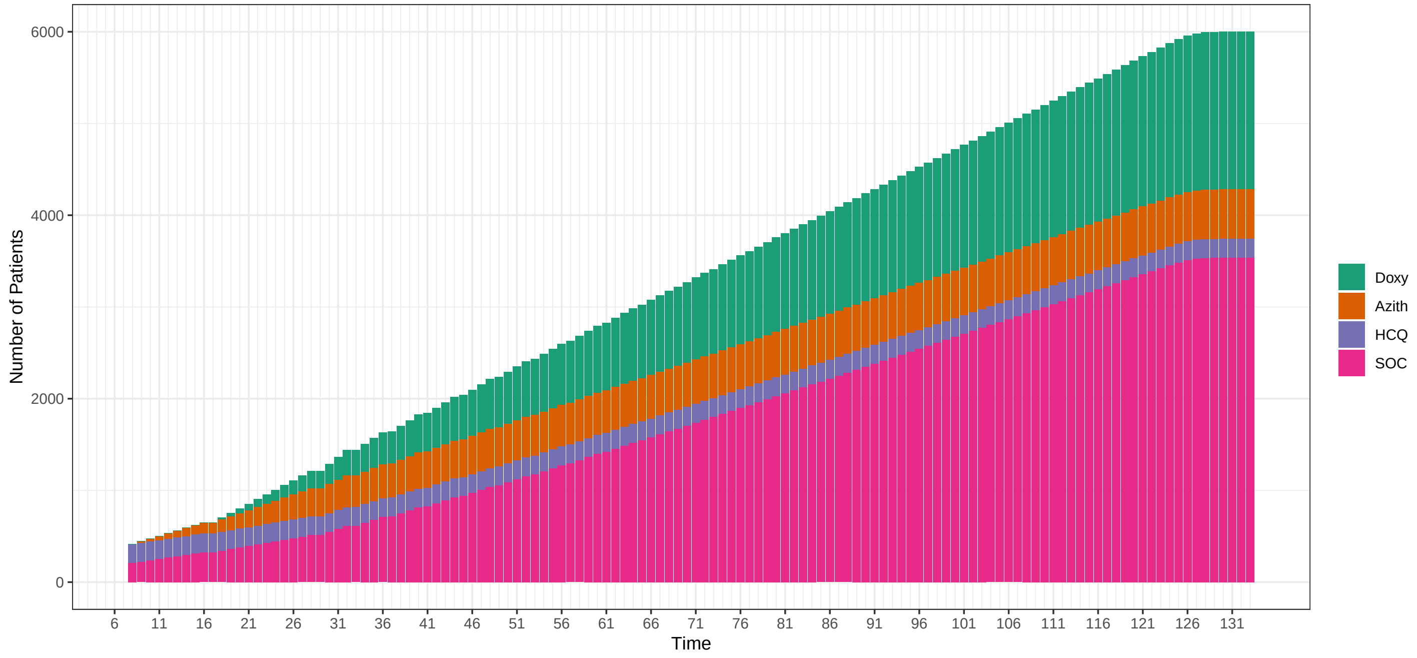

### Hazard Ratios: HCQ: 1 Azith: 1 Doxy: 1.2

#### Decisions Across Time

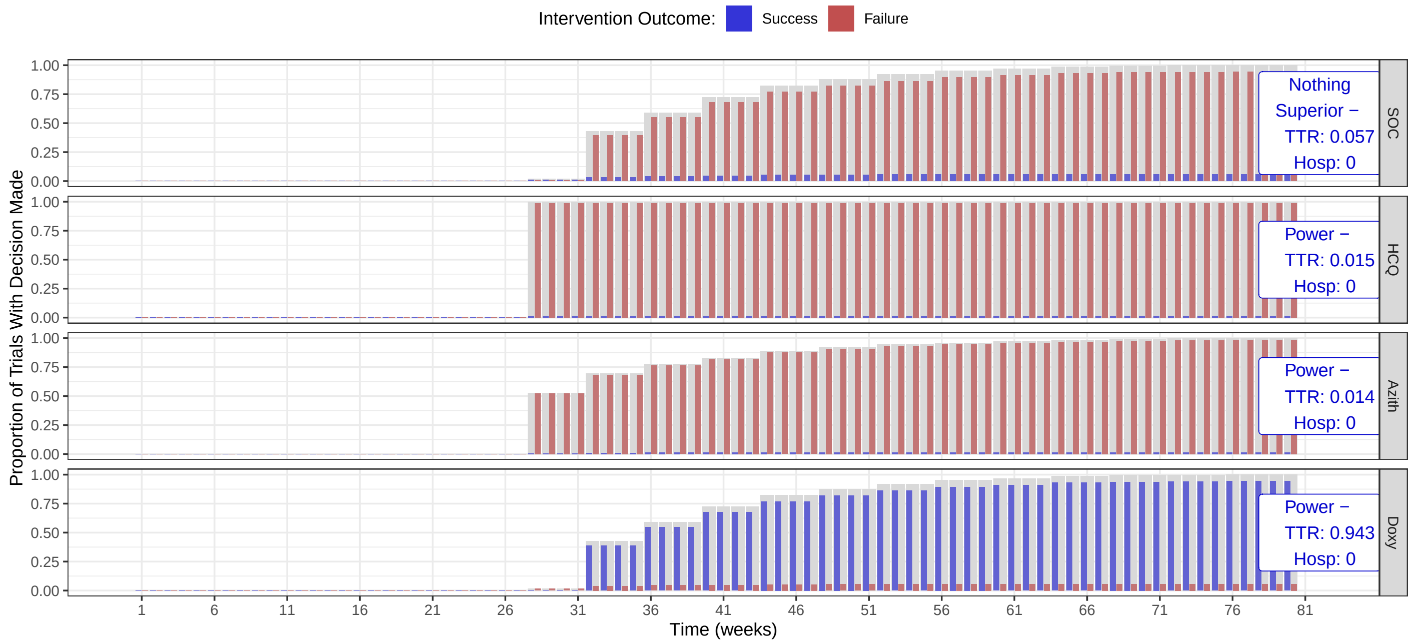

#### Randomization Rates

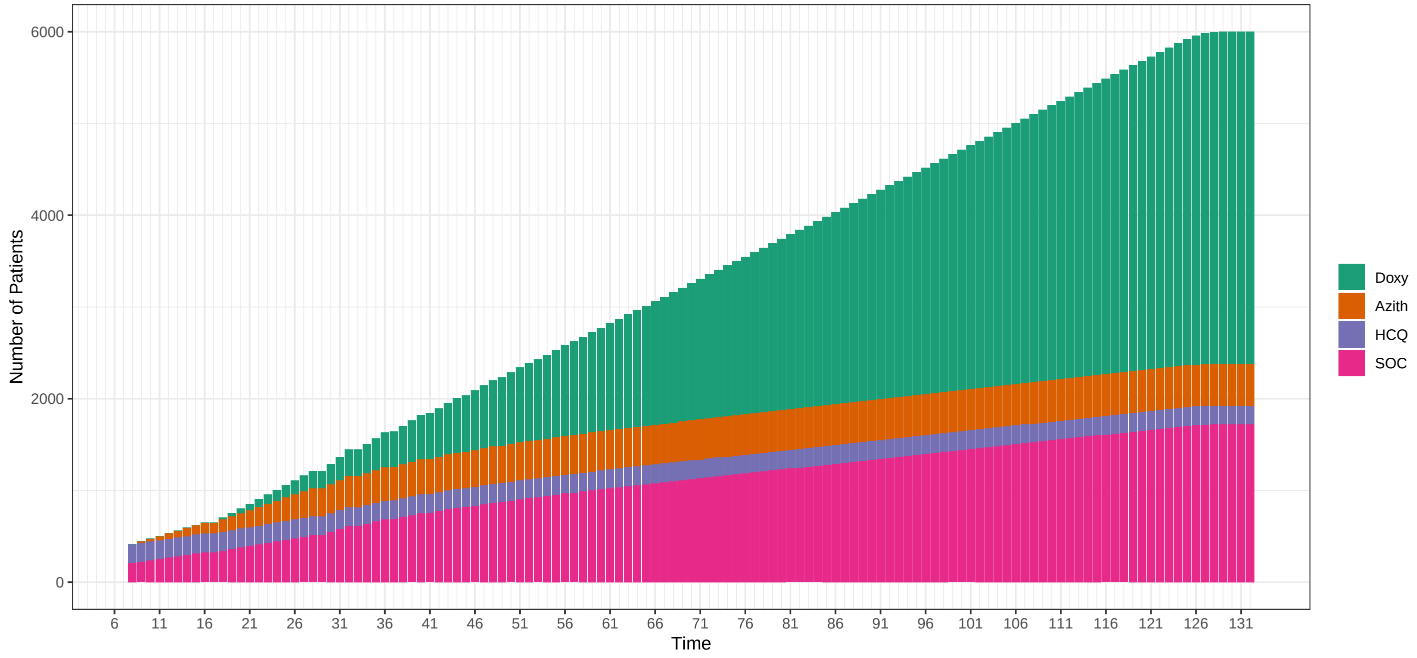

#### Hazard Ratios: HCQ: 1 Azith: 1 Doxy: 1.3

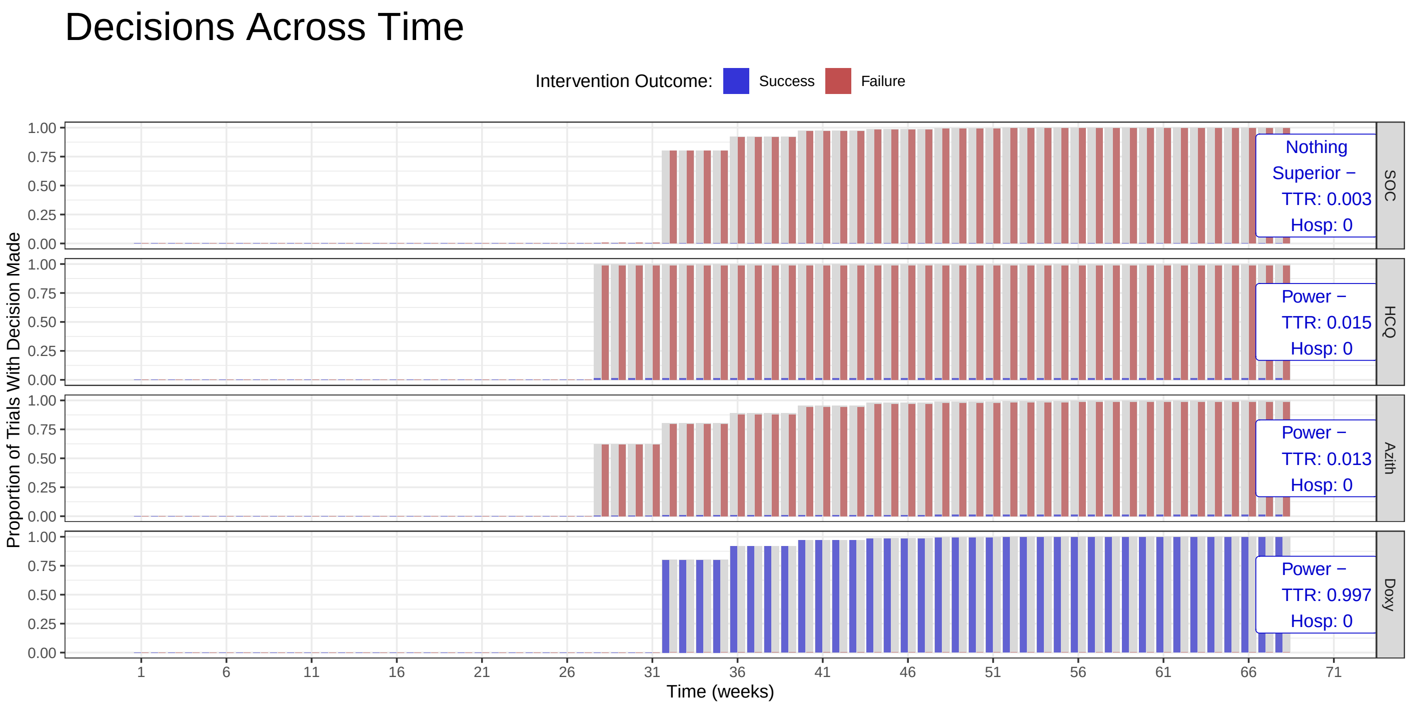

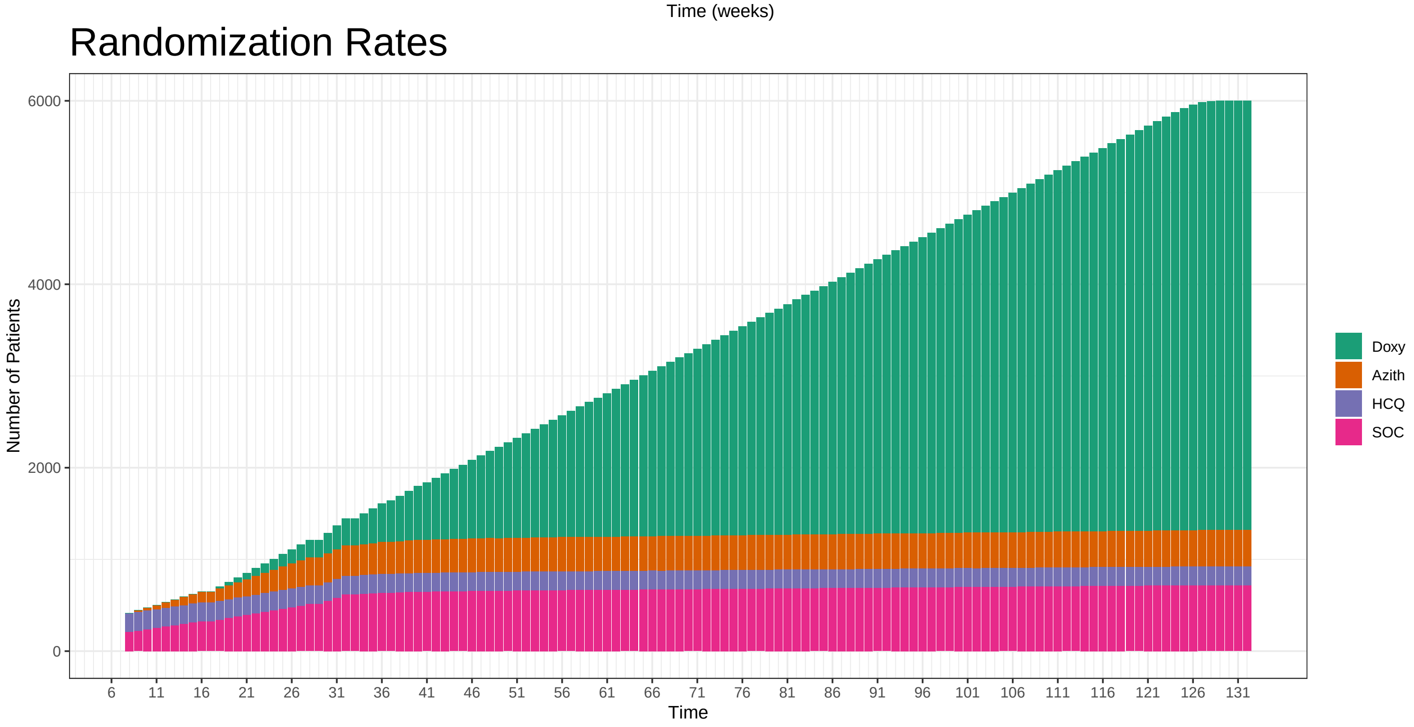

##### 5.5 OCs Additional Scenarios: Fast Accrual

### Principle OCs

Berry Consultants

10/26/2020

These operating characteristics are based on 1000 simulations per scenario across a variety of assumed treatment effects for experimental arms in the trial. The true number of patients randomized per day from April 1, 2020 to October 19, 2020. After this period is over, the simulations assume a faster than expected 100 patient per week accrual rate. In absence of a definitive rule, these simulations conduct an interim every 28 days. In addition, the true day of enrollment stopping of HCQ is used as well as the days for opening enrollment to the Azithromycin and Doxycycline arms. As such, up to at least October 19, 2020 (day 199) of the trial, the enrollment rates to each arm in these simulations are expected to align closely with the true enrollment rates to the arms in the trial.

| Median TTR | Hazard Ratio | | | Power for TTR | | | | Hospitalization Rate Benefit (%) | | | Power for TTR & Hosp | | |
|---|---|---|---|---|---|---|---|---|---|---|---|---|---|
| SOC | HCQ | Azith | Doxy | SOC | HCQ | Azith | Doxy | HCQ | Azith | Doxy | HCQ | Azith | Doxy |
| 9 | 1 | 1 | 1.0 | 0.962 | 0.014 | 0.023 | 0.016 | 0 | 0 | 0 | 0 | 0 | 0.000 |
| 9 | 1 | 1 | 1.1 | 0.613 | 0.015 | 0.020 | 0.378 | 0 | 0 | 1 | 0 | 0 | 0.067 |
| 9 | 1 | 1 | 1.2 | 0.120 | 0.015 | 0.016 | 0.880 | 0 | 0 | 2 | 0 | 0 | 0.588 |
| 9 | 1 | 1 | 1.3 | 0.011 | 0.014 | 0.009 | 0.989 | 0 | 0 | 3 | 0 | 0 | 0.975 |

Decisions on Time to Recovery Benefit By Time

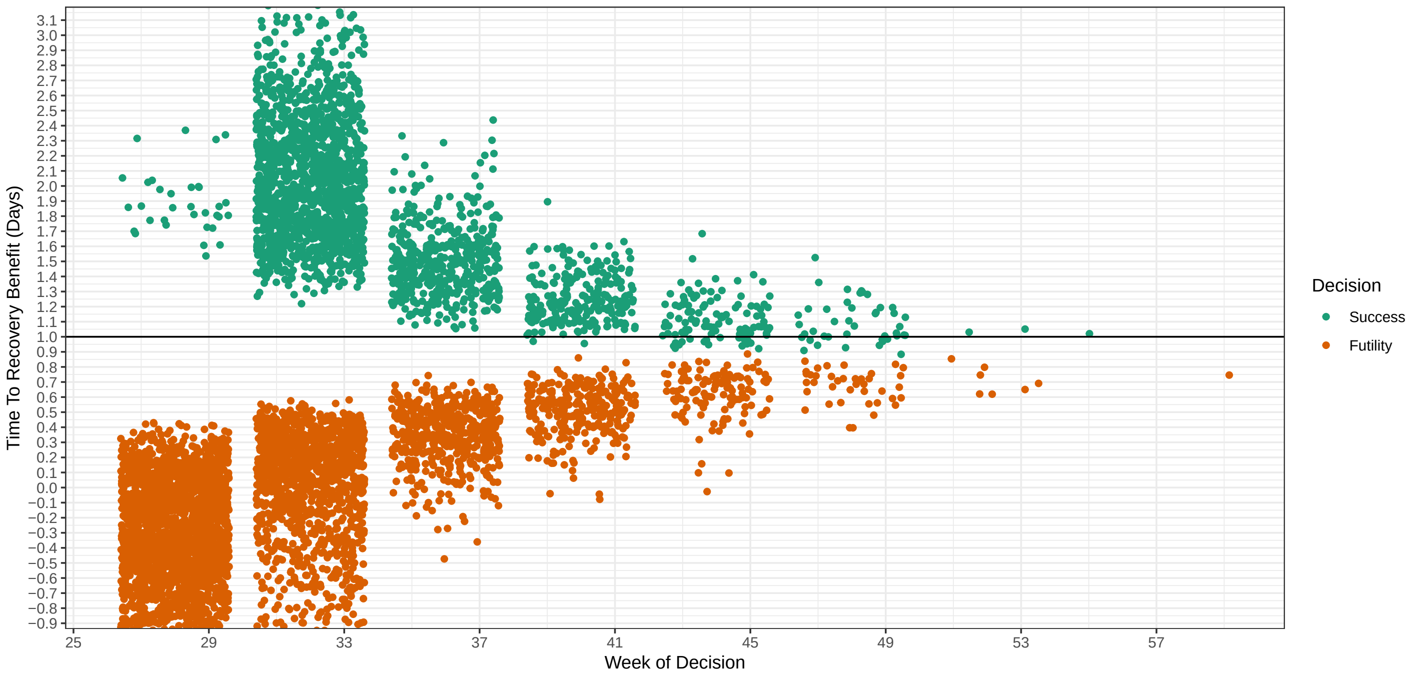

### Hazard Ratios: HCQ: NA Azith: NA Doxy: NA

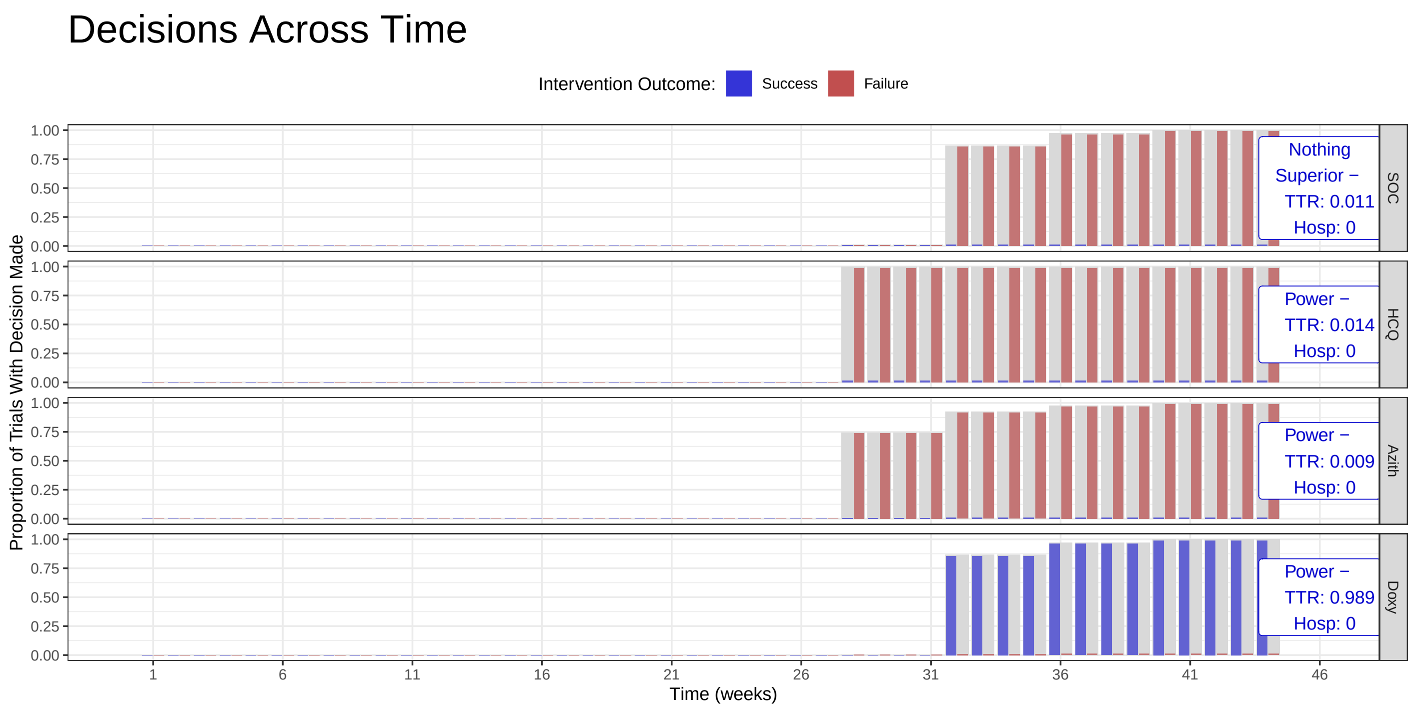

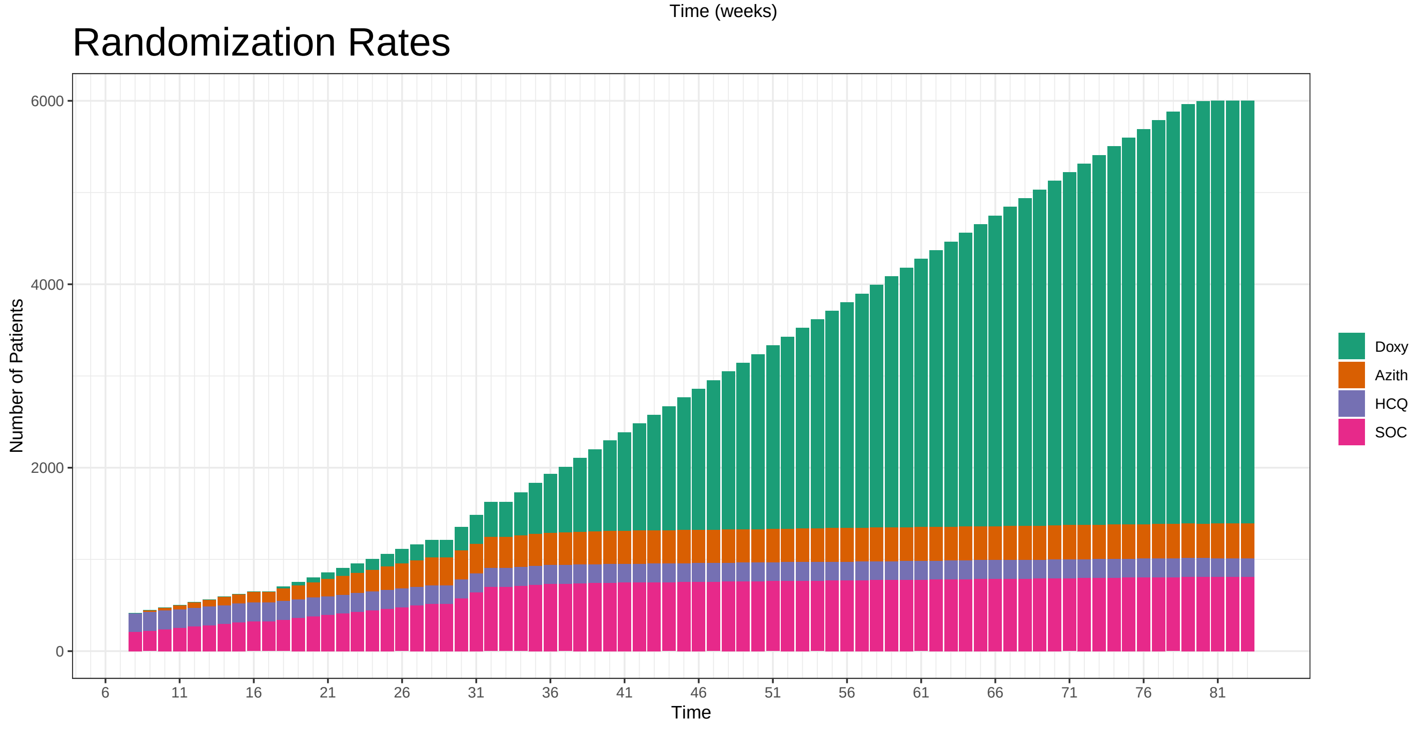

### Hazard Ratios: HCQ: NA Azith: NA Doxy: NA

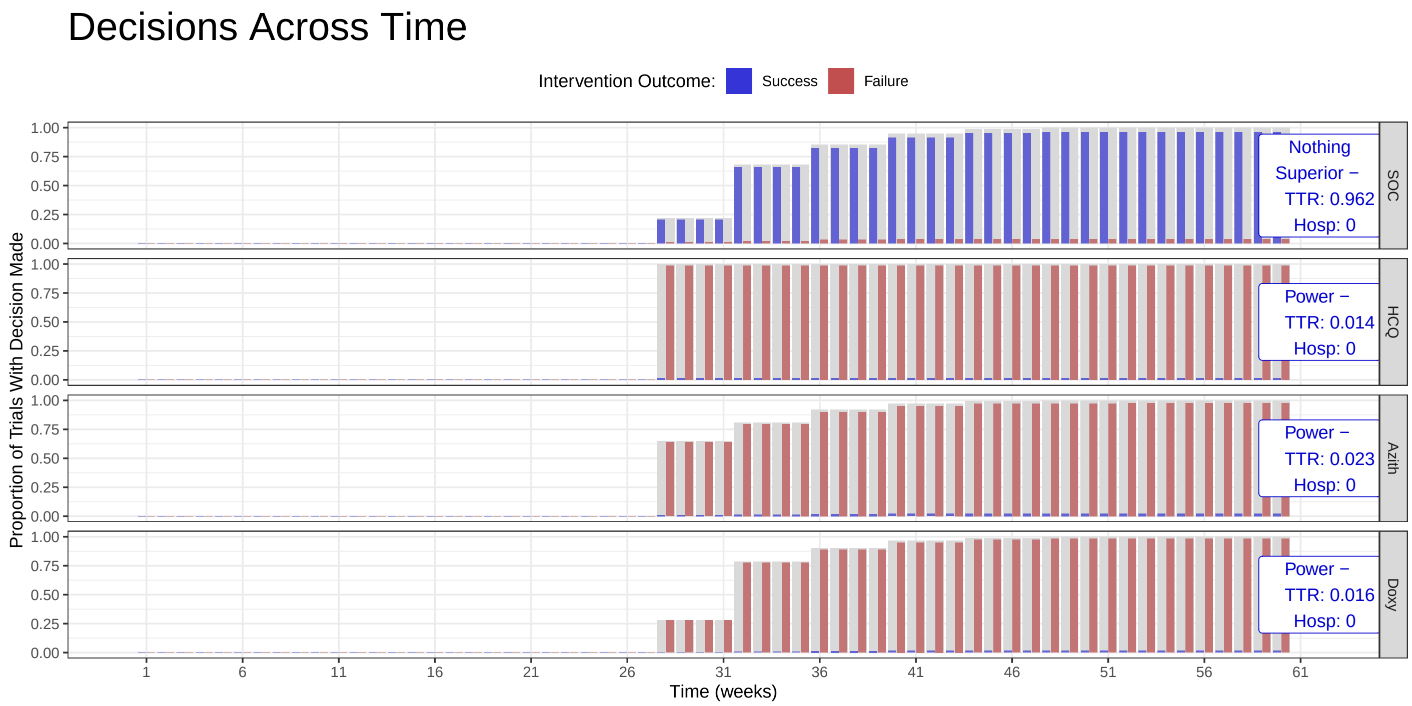

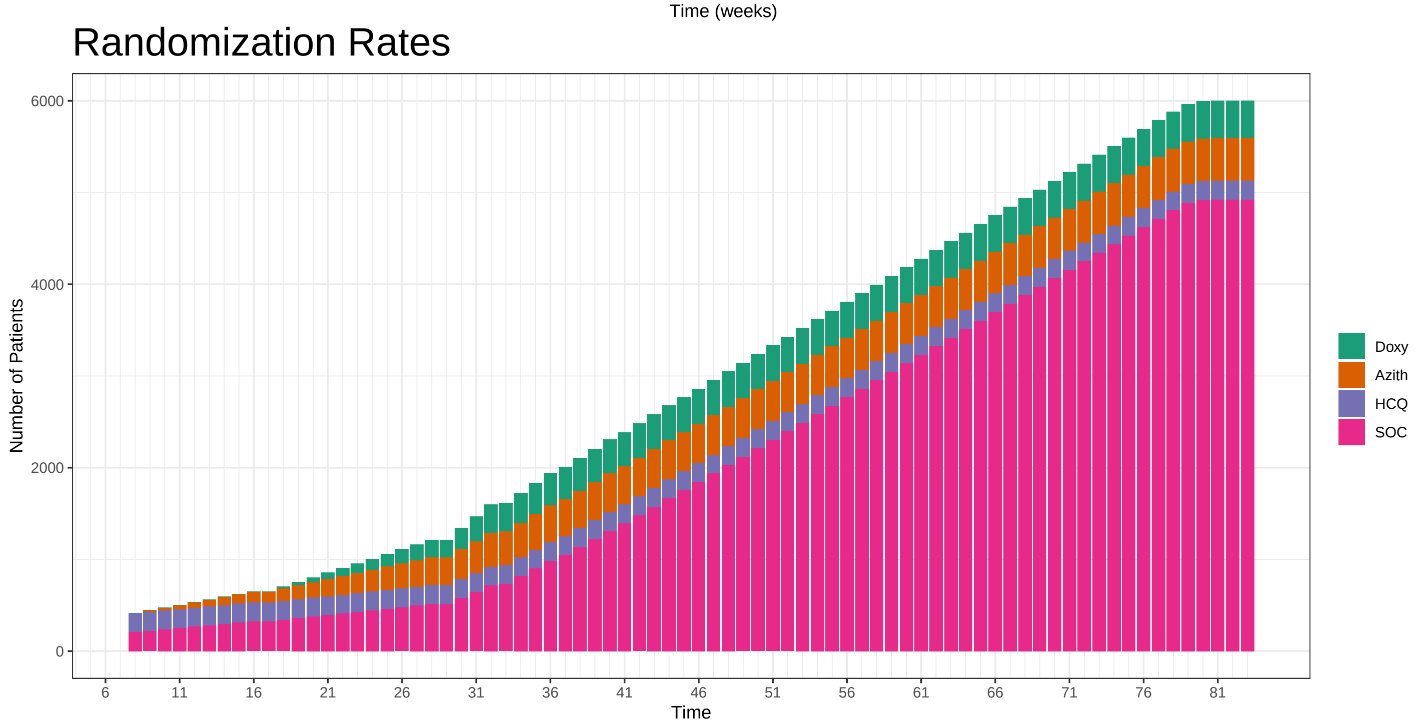

**Hazard Ratios: HCQ: NA Azith: NA Doxy: NA**

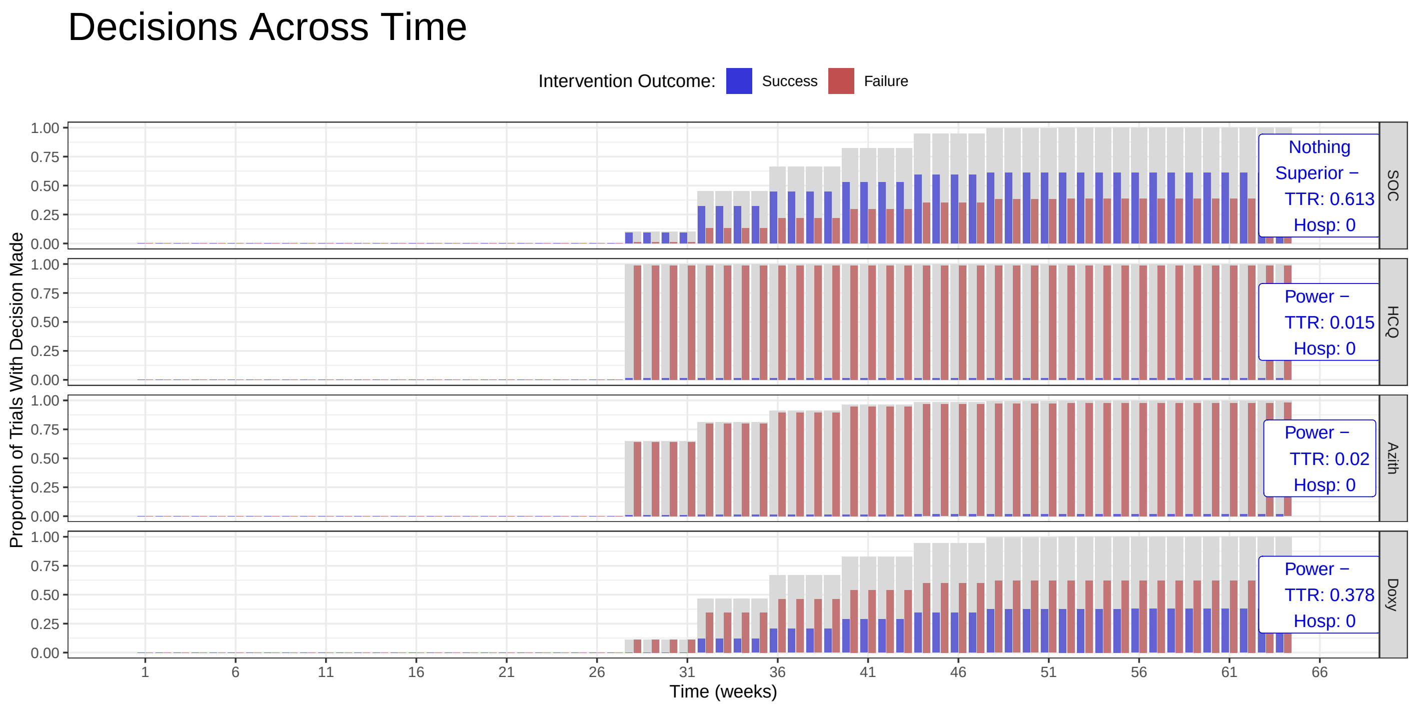

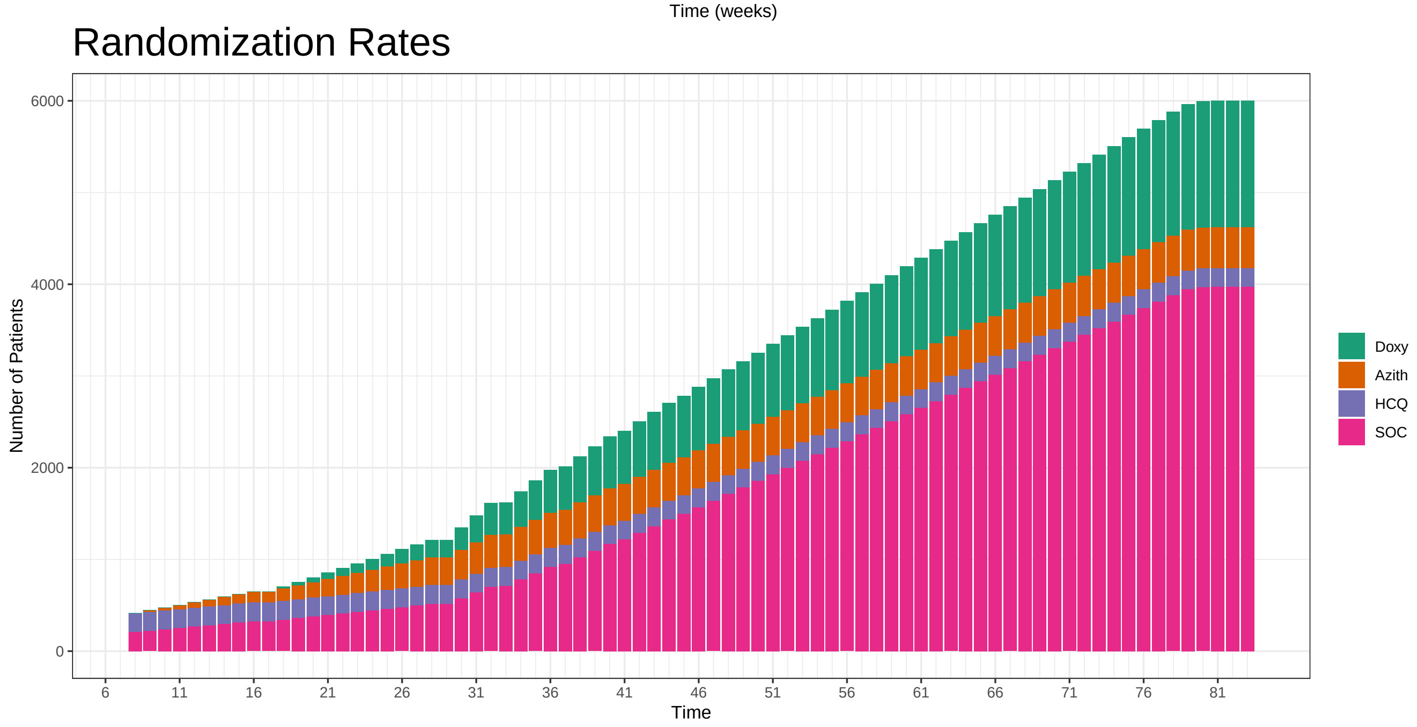

### Hazard Ratios: HCQ: NA Azith: NA Doxy: NA

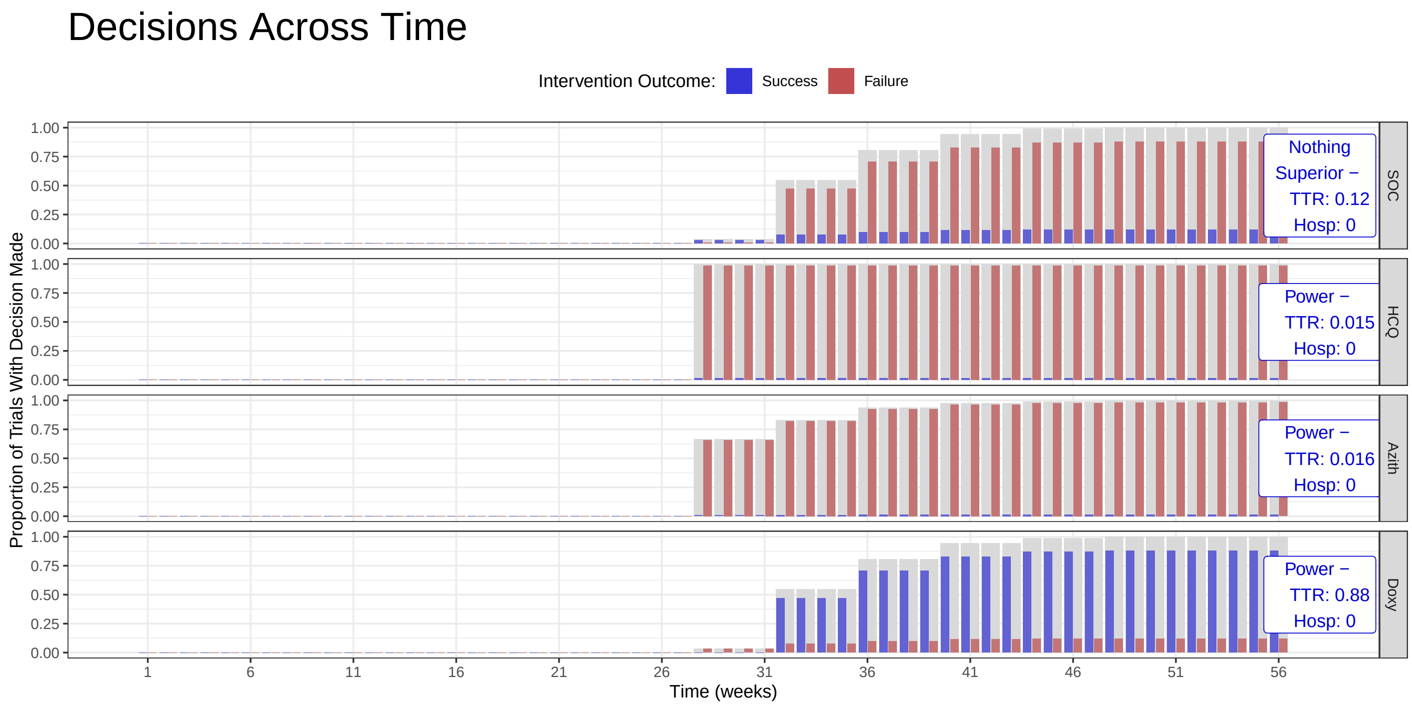

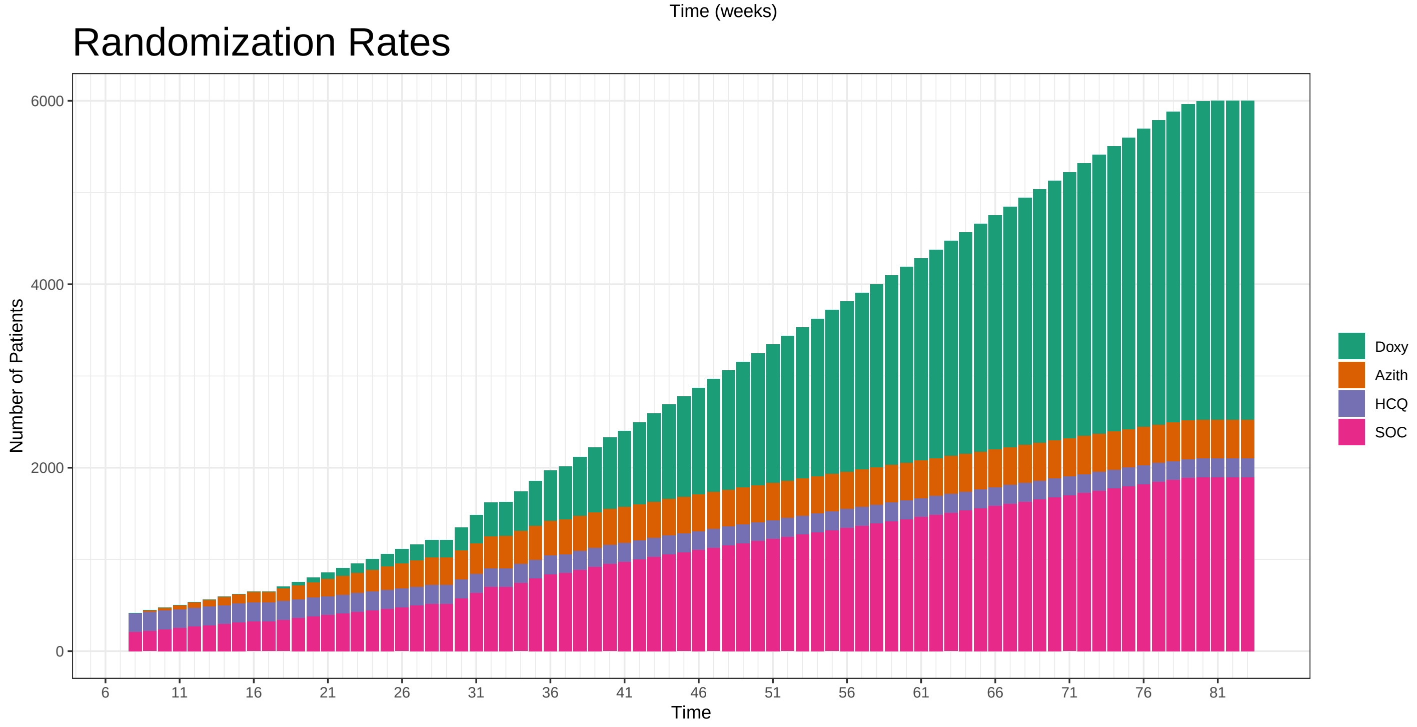

### 6 Simulated Example Trials

#### 6.1 Simulated Example Trials: Three Interventions

This section explores the virtual simulations for the first three interventions versus Usual Care, with staggered entry (and HCQ suspension) as observed in the trial. We simulate virtual example trials and apply the adaptive algorithm exactly as described in the preceding sections. At each interim analysis, we perform an analysis of the data available at the time of the interim. For the purposes of the simulations, each of the trials continues to 4,000 participants in the primary analysis population, but in reality the end of the platform trial will be determined by available resources and the state of the pandemic. The calculations and statistical inferences using these data are summarized in an interim dashboard. The interim number and timing of the interim are provided in the upper left hand corner of the dashboard.

The table provides quantities calculated from the raw data as well as model estimated quantities. The columns denote the following.

**Recovery Data**

- Enrolled: The number of participants randomized to each arm at the time of the interim analysis
- Complete: The number of randomized participants with the opportunity to complete 28-days of follow-up
- Recovered: The number of randomized participants recovered, per primary endpoint definition (i.e. first recovery)
- Exposure Days: The total number of days of observed follow-up prior to a subjects's reported recovery across all participants
- Recoveries Per Day: The number of recoveries divided by the number of exposure days, interpreted as the number of recoveries per day per participant
- Estimated Hazard: The probability of recovery on day $d$ given no recovery up to day $d$, with 95% Bayesian credible interval, averaged across all time segments
- Estimated HR: The estimated hazard ratio comparing the hazard of recovery for intervention $j$ versus the Usual Care, with 95% Bayesian credible interval
- Estimated Median Time to Recovery: The model-based estimated median time to recovery for each treatment arm

**Hospitalization Data:**

- Hospitalizations: Total number of participants with hospitalization or death
- 28 Day Completers: Number of participants with opportunity to complete 28 days of follow-up
- Observed Hosp. Rate: Proportion of participants with hospitalization or death among those with the opportunity to complete 28 days of follow-up
- Est. Hosp. Rate: Model-based estimate for probability of hospitalization with corresponding 95% credible interval

**Recovery Inferences:**

- Pr(Superiority): The model-based estimated probability of superiority with respect to recovery endpoint for intervention $j$ versus Usual Care
- Pr(Meaningful Effect): The model-based estimated probability that the benefit in median time to recovery for intervention $j$ compared to Usual Care is at least 1.5 days
- Pr(Best): The model-based estimated probability that intervention $j$ is superior to all other active interventions with respect to the recovery endpoint
- Randomization Probability: The randomization probabilities to be used for treatment allocation until the next interim analysis

**Hospitalization Inferences:**

- Pr(Superiority): The model-based estimated probability of superiority for intervention $j$ versus Usual Care with respect to the hospitalization/death endpoint
- Pr(Meaningful Effect): The model-based estimated probability that the reduction in proportion of persons with hospitalization/death for intervention $j$ relative to Usual Care 0.02 or greater (i.e., at least 2 percentage points)

The plot and table in the lower left corner of the dashboard provide a more detailed look at the observed data for the recovery endpoint. The plot is a Kaplan-Meier plot showing the proportion of subjects recovered for days 0-28. The table below the plot provides the number not yet recovered by 0, 7, 14, 21, and 28 days for each arm. Both of these summaries are showing observed data that contribute to the statistical modeling.

The two plots on the lower right show summaries of the statistical modeling of the recovery endpoint. The first plot shows the Bayesian posterior distributions of the hazard ratios corresponding to each intervention. Hazard ratios greater than one indicate faster recovery. The second plot (on far right) shows the Bayesian posterior distribution of the median time to recovery in days, with the center of each distribution shown in text above the peak of the distribution.

Below we show four virtual trials to illustrate the adaptive platform trial.

#### 6.2 Example 1

### April 1, 2020

Before Patient Enrollment Begins

#### Recovery Data

| | Enrolled | Complete | Recovered | Exposure Days | Recoveries Per Day | Estimated Hazard | Estimated HR (95% interval) | Estimated Median Survival Time |
|---|---|---|---|---|---|---|---|---|
| SOC | 0 | 0 | 0 | 0 | | | | |
| HCQ | 0 | 0 | 0 | 0 | | | | |
| Azith | 0 | 0 | 0 | 0 | | | | |
| Doxy | 0 | 0 | 0 | 0 | | | | |
| Total | 0 | 0 | 0 | 0 | | | | |

#### Hospitalization Data

| Hospitalizations | 28 Day Completers | Observed Hosp. Rate | Est. Hosp. Rate (95% interval) |
|---|---|---|---|
| 0 | 0 | | |
| 0 | 0 | | |
| 0 | 0 | | |
| 0 | 0 | | |
| 0 | 0 | | |

| Intervention Status |
|---|
| Enrolling |
| Enrolling |
| Unopened |
| Unopened |

#### Observed Patient Recoveries

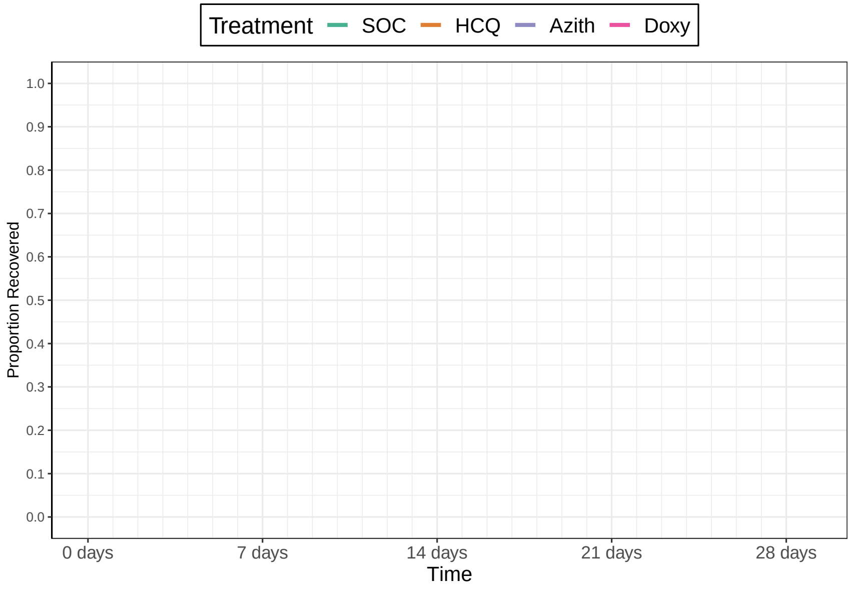

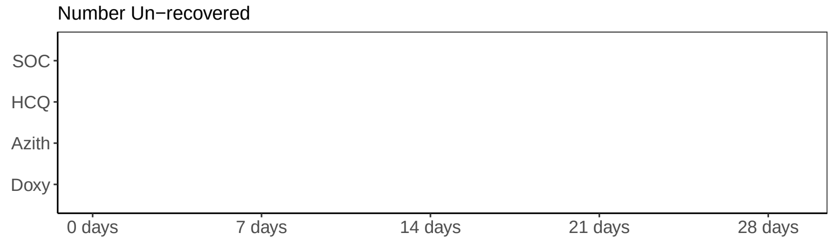

#### Recovery Inferences

| | Pr(Superiority) | Pr(Meaningful Effect) | Pr(Best) | Randomization Probability |
|---|---|---|---|---|
| SOC | | | | 0.5 |
| HCQ | | | | 0.5 |
| Azith | | | | 0 |
| Doxy | | | | 0 |

#### Hospitalization Inferences

| Pr(Superiority) | Pr(Meaningful Effect) |
|---|---|
| 0 | 0 |
| 0 | 0 |
| 0 | 0 |
| 0 | 0 |

#### Hazard Ratio Estimates

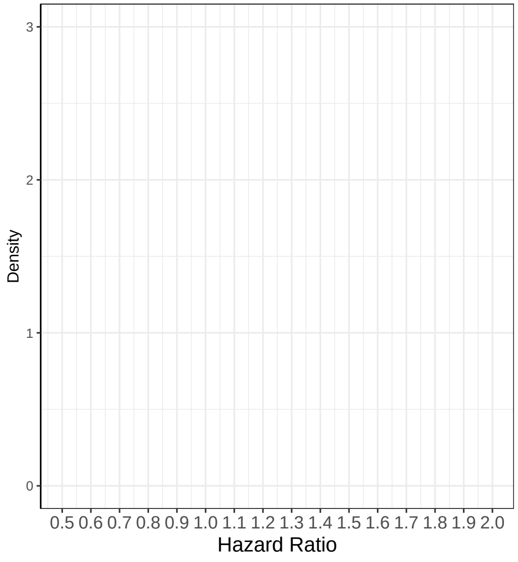

#### Median Time To Recovery Estimates

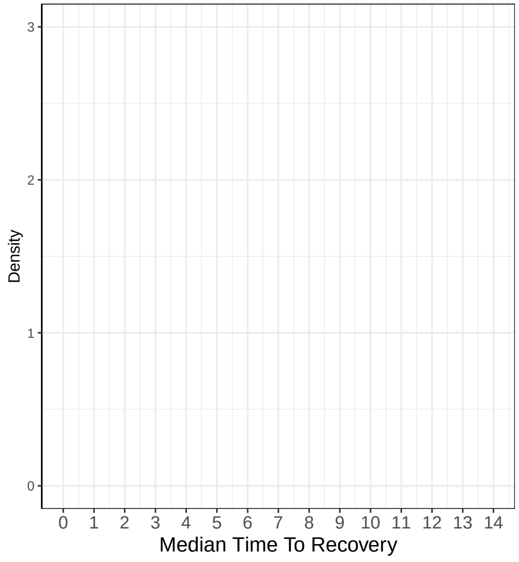

### Remove Drug on May 22, 2020

##### Recovery Data

| | Enrolled | Complete | Recovered | Exposure Days | Recoveries Per Day | Estimated Hazard | Estimated HR (95% interval) | Estimated Median Time to Recovery |
|---|---|---|---|---|---|---|---|---|
| SOC | 207 | 143 | 134 | 1817 | 0.074 | | | |
| HCQ | 206 | 147 | 140 | 1861 | 0.075 | | | |
| Azith | 0 | 0 | 0 | 0 | | | | |
| Doxy | 0 | 0 | 0 | 0 | | | | |
| Total | 413 | 290 | 274 | 3679 | 0.074 | | | |

##### Hospitalization Data

| Hospitalizations | 28 Day Completers | Observed Hosp. Rate | Est. Hosp. Rate (95% interval) |
|---|---|---|---|
| 2 | 79 | 0.0253 | |
| 6 | 78 | 0.0769 | |
| 0 | 0 | NA | |
| 0 | 0 | NA | |
| 8 | 157 | 0.0510 | |

| Intervention Status |
|---|
| Enrolling |
| Removing |
| Unopened |
| Unopened |

##### Observed Patient Recoveries

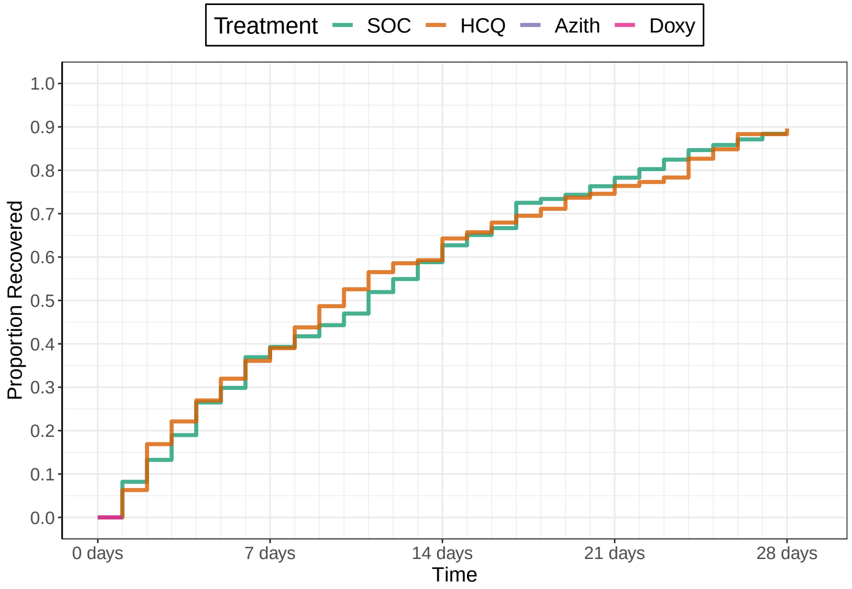

Number Un−recovered

| | 0 days | 7 days | 14 days | 21 days | 28 days |
|---|---|---|---|---|---|
| SOC | 207 | 107 | 53 | 24 | 9 |
| HCQ | 206 | 109 | 57 | 28 | 9 |
| Azith | | | | | |
| Doxy | | | | | |

##### Recovery Inferences

| | Pr(Superiority) | Pr(Meaningful Effect) | Pr(Best) | Randomization Probability |
|---|---|---|---|---|
| SOC | | | | 1 |
| HCQ | | | 0 | 0 |
| Azith | | | 0 | 0 |
| Doxy | | | 0 | 0 |

##### Hospitalization Inferences

| Pr(Superiority) | Pr(Meaningful Effect) |
|---|---|

##### Hazard Ratio Estimates

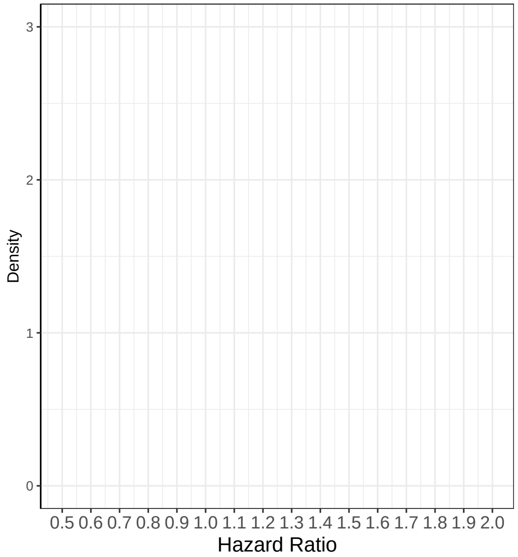

##### Median Time To Recovery Estimates

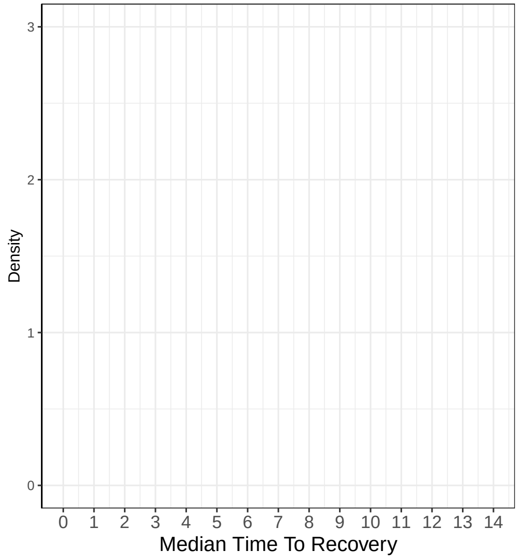

### Add Drug on May 23, 2020

#### Recovery Data

| | Enrolled | Complete | Recovered | Exposure Days | Recoveries Per Day | Estimated Hazard | Estimated HR (95% interval) | Estimated Median Time to Recovery |
|---|---|---|---|---|---|---|---|---|
| SOC | 209 | 147 | 138 | 1881 | 0.073 | | | |
| HCQ | 206 | 151 | 143 | 1918 | 0.075 | | | |
| Azith | 0 | 0 | 0 | 0 | | | | |
| Doxy | 0 | 0 | 0 | 0 | | | | |
| Total | 415 | 298 | 281 | 3799 | 0.074 | | | |

#### Hospitalization Data

| Hospitalizations | 28 Day Completers | Observed Hosp. Rate | Est. Hosp. Rate (95% interval) |
|---|---|---|---|
| 2 | 80 | 0.025 | |
| 6 | 80 | 0.075 | |
| 0 | 0 | NA | |
| 0 | 0 | NA | |
| 8 | 160 | 0.050 | |

| Intervention Status |
|---|
| Enrolling |
| Paused |
| Introducing |
| Unopened |

#### Observed Patient Recoveries

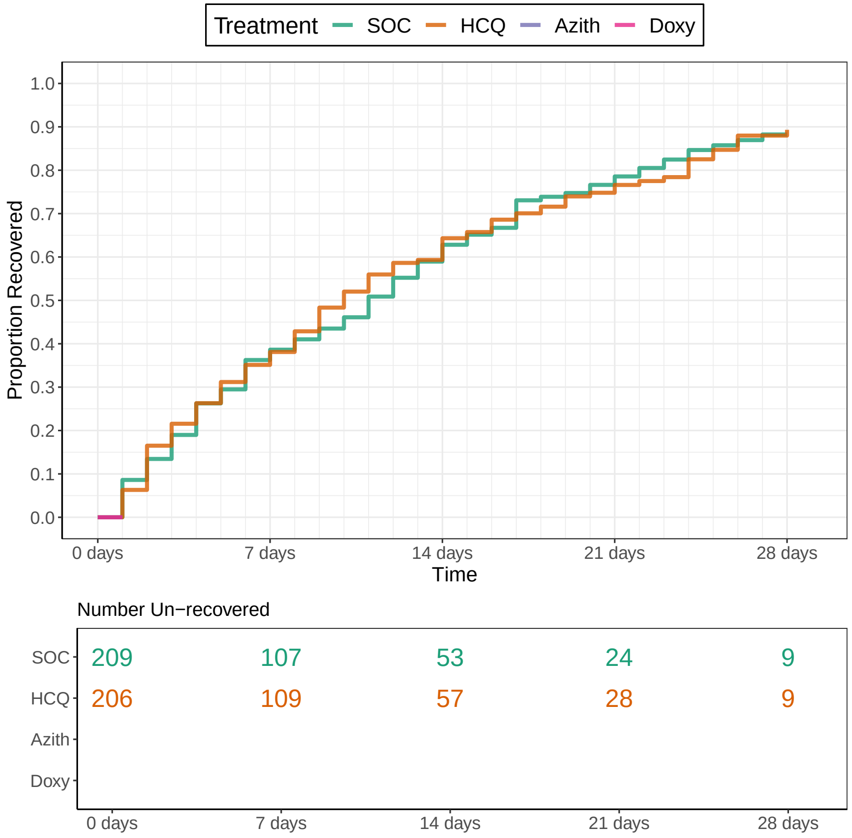

#### Recovery Inferences

| | Pr(Superiority) | Pr(Meaningful Effect) | Pr(Best) | Randomization Probability |
|---|---|---|---|---|
| SOC | | | | 0.5 |
| HCQ | | | 0 | 0 |
| Azith | | | 1 | 0.5 |
| Doxy | | | 0 | 0 |

#### Hospitalization Inferences

| Pr(Superiority) | Pr(Meaningful Effect) |
|---|---|

#### Hazard Ratio Estimates

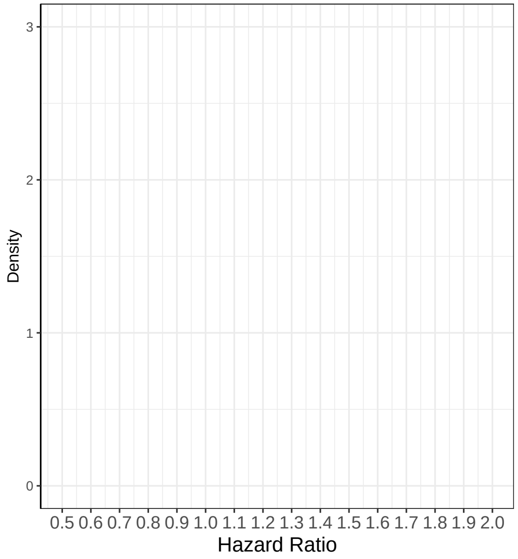

#### Median Time To Recovery Estimates

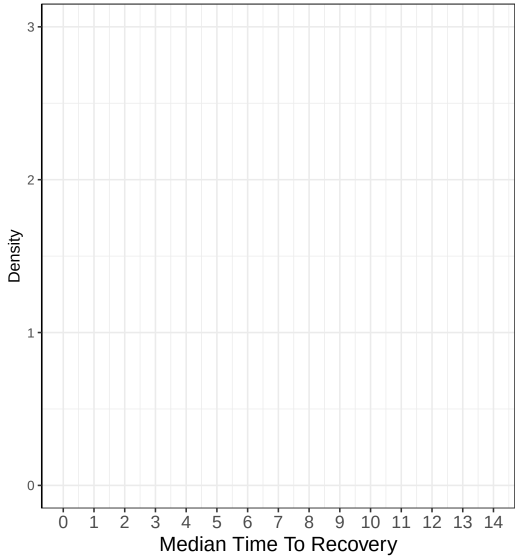

### Add Drug on July 23, 2020

##### Recovery Data

| | Enrolled | Complete | Recovered | Exposure Days | Recoveries Per Day | Estimated Hazard | Estimated HR (95% interval) | Estimated Median Time to Recovery |
|---|---|---|---|---|---|---|---|---|
| SOC | 327 | 308 | 272 | 3615 | 0.075 | | | |
| HCQ | 206 | 206 | 176 | 2546 | 0.069 | | | |
| Azith | 118 | 103 | 94 | 1175 | 0.08 | | | |
| Doxy | 0 | 0 | 0 | 0 | | | | |
| Total | 651 | 617 | 542 | 7336 | 0.074 | | | |

##### Hospitalization Data

| Hospitalizations | 28 Day Completers | Observed Hosp. Rate | Est. Hosp. Rate (95% interval) |
|---|---|---|---|
| 3 | 286 | 0.0105 | |
| 6 | 206 | 0.0291 | |
| 8 | 77 | 0.1039 | |
| 0 | 0 | NA | |
| 17 | 569 | 0.0299 | |

| Intervention Status |
|---|
| Enrolling |
| Paused |
| Introducing |
| Introducing |

##### Observed Patient Recoveries

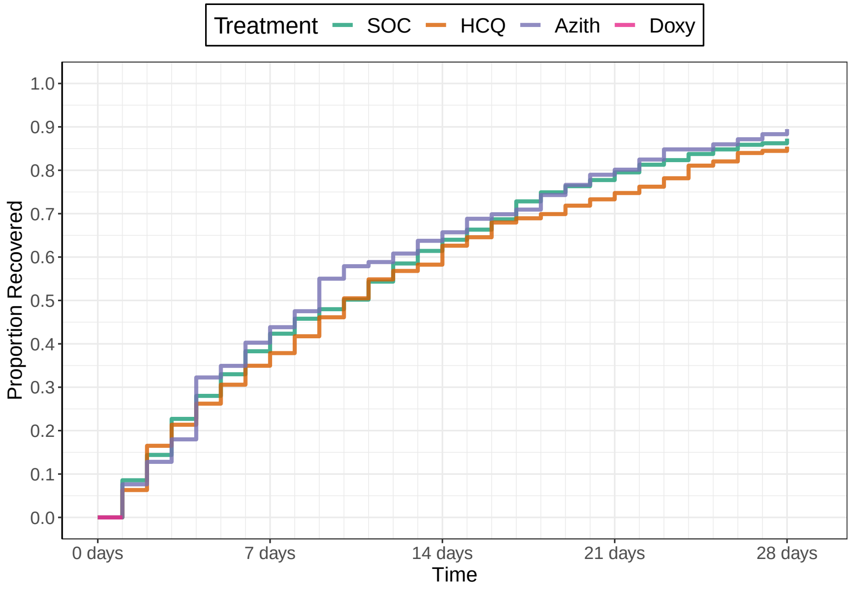

Number Un−recovered

| | 0 days | 7 days | 14 days | 21 days | 28 days |
|---|---|---|---|---|---|
| SOC | 327 | 198 | 120 | 63 | 39 |
| HCQ | 206 | 134 | 86 | 55 | 32 |
| Azith | 118 | 67 | 37 | 18 | 10 |
| Doxy | | | | | |

##### Recovery Inferences

| | Pr(Superiority) | Pr(Meaningful Effect) | Pr(Best) | Randomization Probability |
|---|---|---|---|---|
| SOC | | | | 0.33 |
| HCQ | | | 0 | 0 |
| Azith | | | 1 | 0.33 |
| Doxy | | | 0 | 0.33 |

##### Hospitalization Inferences

| Pr(Superiority) | Pr(Meaningful Effect) |
|---|---|

##### Hazard Ratio Estimates

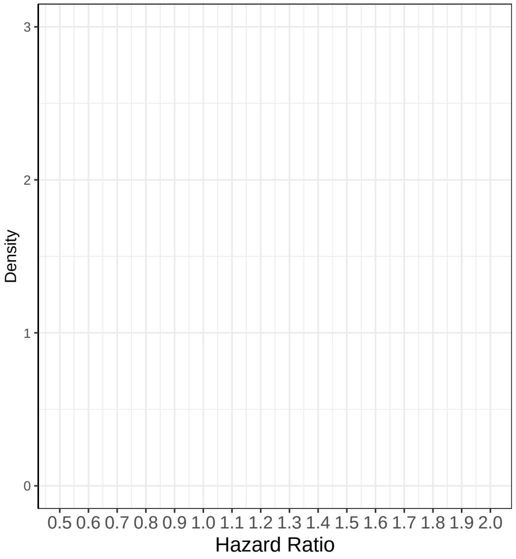

##### Median Time To Recovery Estimates

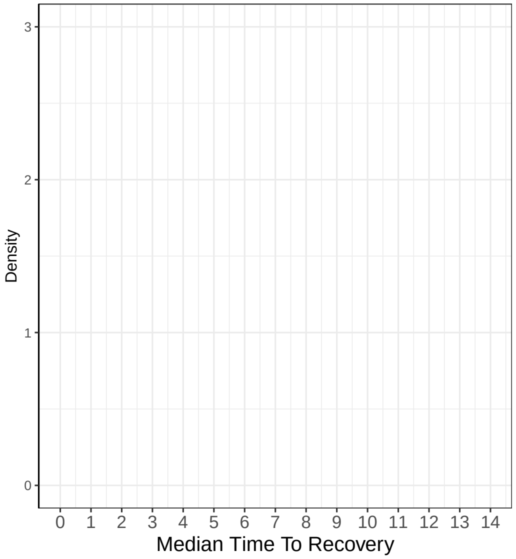

### Interim 1 on October 15, 2020

##### Recovery Data

| | Enrolled | Complete | Recovered | Exposure Days | Recoveries Per Day | Estimated Hazard | Estimated HR (95% interval) | Estimated Median Time to Recovery |
|---|---|---|---|---|---|---|---|---|
| SOC | 496 | 437 | 388 | 5313 | 0.073 | 0.071 | 1 | 9.61 |
| HCQ | 206 | 206 | 176 | 2546 | 0.069 | 0.067 | 0.937 (0.795, 1.113) | 10.27 |
| Azith | 313 | 268 | 252 | 2762 | 0.091 | 0.085 | 1.202 (1.03, 1.4) | 8.01 |
| Doxy | 195 | 151 | 147 | 1353 | 0.109 | 0.097 | 1.368 (1.131, 1.625) | 7.04 |
| Total | 1210 | 1062 | 963 | 11975 | 0.08 | | | |

##### Hospitalization Data

| Hospitalizations | 28 Day Completers | Observed Hosp. Rate | Est. Hosp. Rate (95% interval) |
|---|---|---|---|
| 9 | 395 | 0.0228 | 0.02 (0.0095, 0.0339) |
| 6 | 206 | 0.0291 | 0.0334 (0.0135, 0.061) |
| 12 | 198 | 0.0606 | 0.0413 (0.0219, 0.0666) |
| 7 | 74 | 0.0946 | 0.0407 (0.0179, 0.0713) |
| 34 | 873 | 0.0389 | |

| Intervention Status |
|---|
| LostOnTTR |
| Futility |
| SuccessOnTTRFutilityOnHosp Not Announced |
| Enrolling |

##### Observed Patient Recoveries

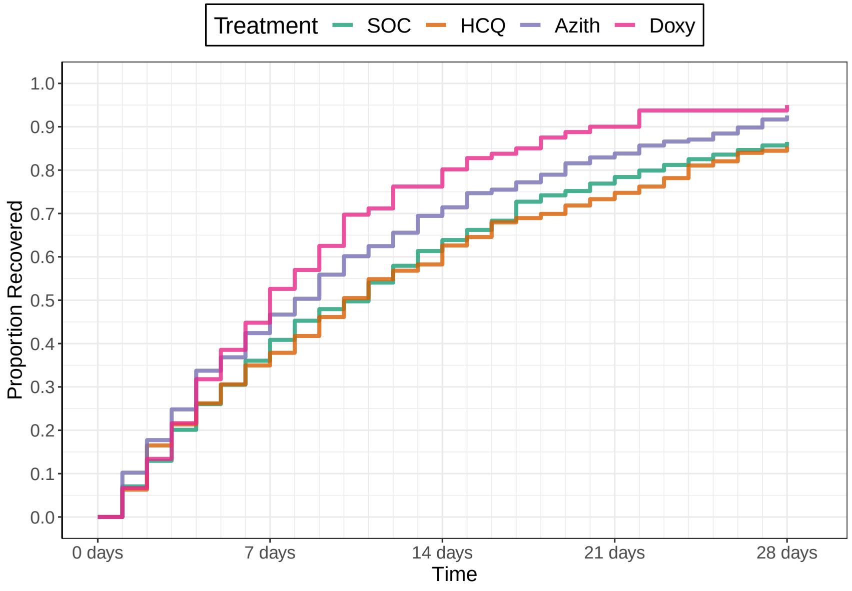

Number Un−recovered

| | 0 days | 7 days | 14 days | 21 days | 28 days |
|---|---|---|---|---|---|
| SOC | 496 | 293 | 168 | 92 | 54 |
| HCQ | 206 | 134 | 86 | 55 | 32 |
| Azith | 313 | 162 | 77 | 38 | 18 |
| Doxy | 195 | 92 | 30 | 8 | 5 |

##### Recovery Inferences

| | Pr(Superiority) | Pr(Meaningful Effect) | Pr(Best) | Randomization Probability |
|---|---|---|---|---|
| SOC | | | | 0.25 |
| HCQ | 0.2183 | 0.009 | 0 | 0 |
| Azith | 0.9903 | 0.5687 | 0.0788 | 0.25 |
| Doxy | 0.9993 | 0.9257 | 0.6712 | 0.5 |

##### Hospitalization Inferences

| Pr(Superiority) | Pr(Meaningful Effect) |
|---|---|
| 0.1652 | 0.0038 |
| 0.0455 | 5e−04 |
| 0.0712 | 0 |

##### Hazard Ratio Estimates

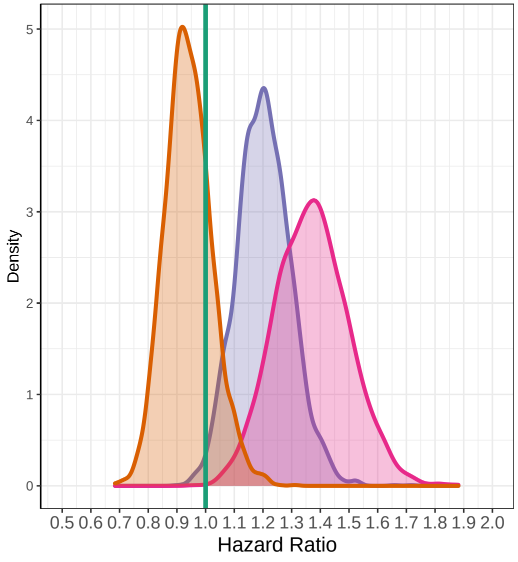

##### Median Time To Recovery Estimates

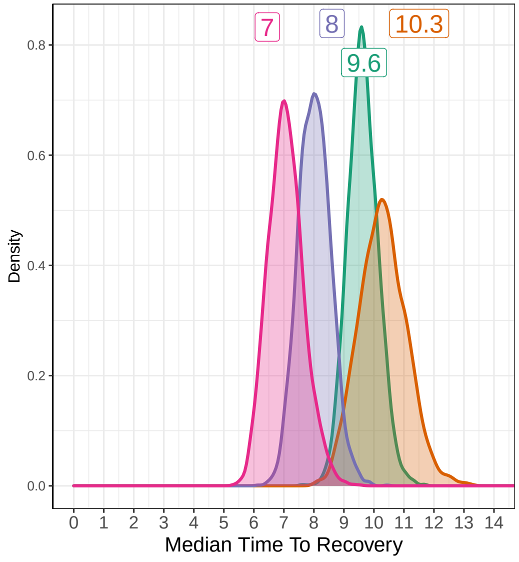

### Interim 2 on November 12, 2020

#### Recovery Data

| | Enrolled | Complete | Recovered | Exposure Days | Recoveries Per Day | Estimated Hazard | Estimated HR (95% interval) | Estimated Median Time to Recovery |
|---|---|---|---|---|---|---|---|---|
| SOC | 550 | 536 | 472 | 6252 | 0.075 | 0.073 | 1 | 9.28 |
| HCQ | 206 | 206 | 176 | 2546 | 0.069 | 0.066 | 0.915 (0.768, 1.081) | 10.17 |
| Azith | 377 | 352 | 327 | 3592 | 0.091 | 0.085 | 1.173 (1.022, 1.343) | 7.89 |
| Doxy | 294 | 275 | 265 | 2468 | 0.107 | 0.098 | 1.348 (1.156, 1.548) | 6.85 |
| Total | 1427 | 1369 | 1240 | 14858 | 0.083 | | | |

#### Hospitalization Data

| Hospitalizations | 28 Day Completers | Observed Hosp. Rate | Est. Hosp. Rate (95% interval) |
|---|---|---|---|
| 14 | 496 | 0.0282 | 0.0272 (0.0153, 0.0422) |
| 6 | 206 | 0.0291 | 0.034 (0.0137, 0.0619) |
| 15 | 313 | 0.0479 | 0.0423 (0.024, 0.0654) |
| 9 | 195 | 0.0462 | 0.0339 (0.0162, 0.057) |
| 44 | 1210 | 0.0364 | |

| Intervention Status |
|---|
| LostOnTTR |
| Dropped |
| SuccessOnTTRFutilityOnHosp Not Announced |
| SuccessOnTTRFutilityOnHosp *Announced* |

#### Observed Patient Recoveries

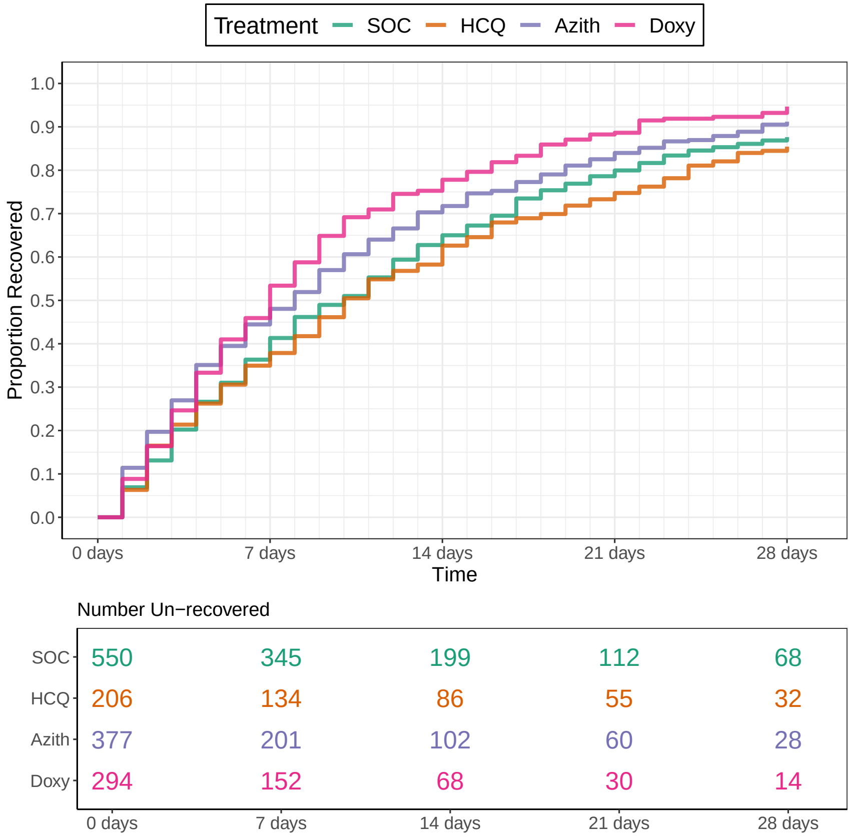

#### Recovery Inferences

| | Pr(Superiority) | Pr(Meaningful Effect) | Pr(Best) | Randomization Probability |
|---|---|---|---|---|
| SOC | | | | 0.33 |
| HCQ | 0.1457 | 0.002 | 0 | 0 |
| Azith | 0.987 | 0.442 | 0.0415 | 0.33 |
| Doxy | 1 | 0.937 | 0.7085 | 0.33 |

#### Hospitalization Inferences

| Pr(Superiority) | Pr(Meaningful Effect) |
|---|---|
| 0.3375 | 0.02 |
| 0.1098 | 0.002 |
| 0.3078 | 0.0132 |

#### Hazard Ratio Estimates

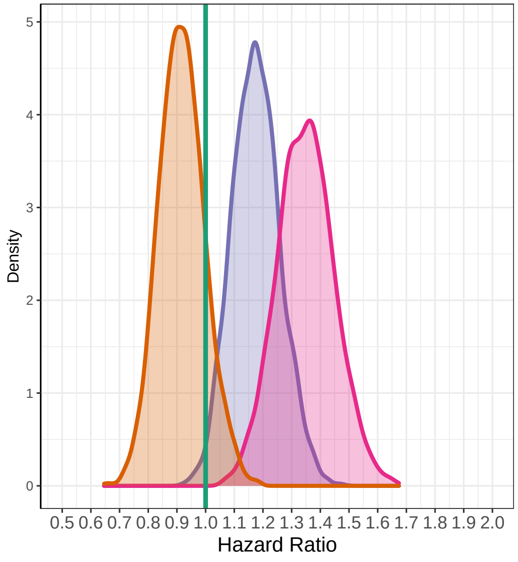

#### Median Time To Recovery Estimates

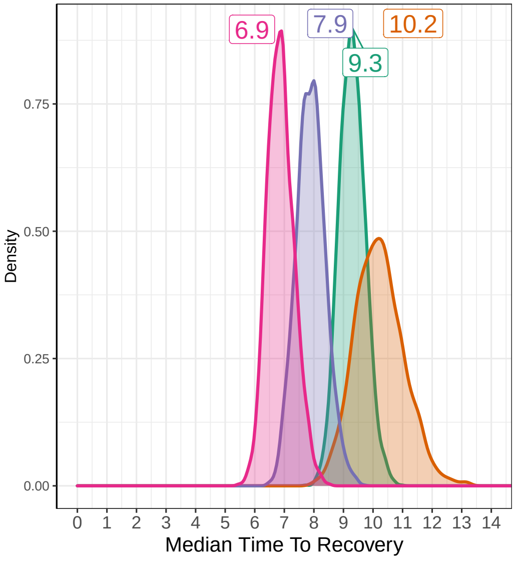

### Interim 3 on September 05, 2022

**Recovery Data**

| | Enrolled | Complete | Recovered | Exposure Days | Recoveries Per Day | Estimated Hazard | Estimated HR (95% interval) | Estimated Median Time to Recovery |
|---|---|---|---|---|---|---|---|---|
| SOC | 2078 | 2078 | 1838 | 24211 | 0.076 | 0.074 | 1 | 9.53 |
| HCQ | 206 | 206 | 176 | 2546 | 0.069 | 0.067 | 0.913 (0.784, 1.056) | 10.41 |
| Azith | 1918 | 1918 | 1769 | 19480 | 0.091 | 0.087 | 1.185 (1.11, 1.266) | 8.08 |
| Doxy | 1798 | 1798 | 1703 | 16353 | 0.104 | 0.099 | 1.347 (1.261, 1.443) | 7.13 |
| Total | 6000 | 6000 | 5486 | 62591 | 0.088 | | | |

**Hospitalization Data**

| Hospitalizations | 28 Day Completers | Observed Hosp. Rate | Est. Hosp. Rate (95% interval) |
|---|---|---|---|
| 51 | 2078 | 0.0245 | 0.0251 (0.019, 0.0322) |
| 6 | 206 | 0.0291 | 0.0334 (0.0137, 0.0612) |
| 60 | 1918 | 0.0313 | 0.0318 (0.0244, 0.0399) |
| 40 | 1798 | 0.0222 | 0.0227 (0.0164, 0.0299) |
| 157 | 6000 | 0.0262 | |

| Intervention Status |
|---|
| Complete |
| Dropped |
| SuccessOnTTRFutilityOnHosp Not Announced |
| SuccessOnTTRFutilityOnHosp *Announced* |

#### Observed Patient Recoveries

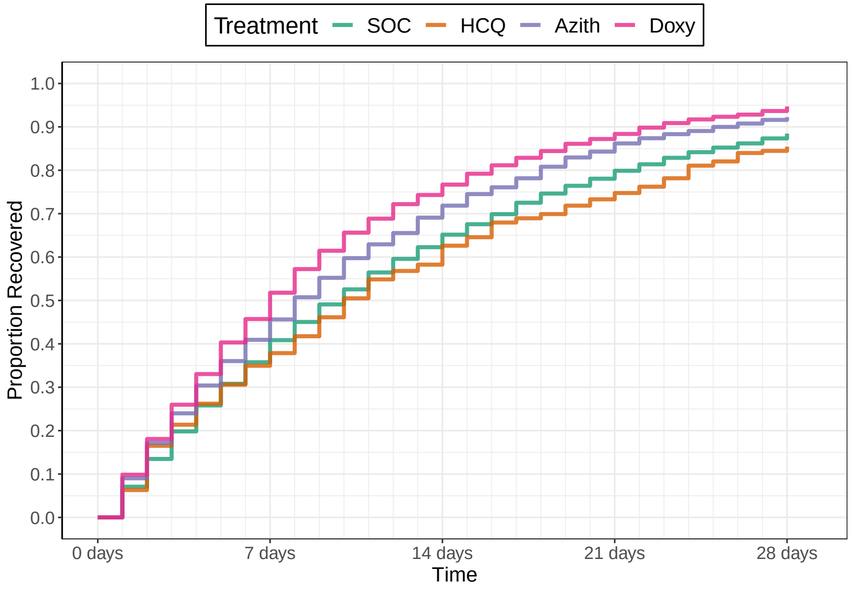

Number Un−recovered

| | 0 days | 7 days | 14 days | 21 days | 28 days |
|---|---|---|---|---|---|
| SOC | 2078 | 1335 | 784 | 456 | 263 |
| HCQ | 206 | 134 | 86 | 55 | 32 |
| Azith | 1918 | 1133 | 593 | 301 | 161 |
| Doxy | 1798 | 976 | 462 | 230 | 114 |

**Recovery Inferences**

| | Pr(Superiority) | Pr(Meaningful Effect) | Pr(Best) | Randomization Probability |
|---|---|---|---|---|
| SOC | | | | |
| HCQ | 0.1137 | 3e−04 | 0 | |
| Azith | 1 | 0.438 | 2e−04 | |
| Doxy | 1 | 0.9997 | 0.7498 | |

**Hospitalization Inferences**

| Pr(Superiority) | Pr(Meaningful Effect) |
|---|---|
| 0.2758 | 0.0012 |
| 0.095 | 0 |
| 0.6968 | 2e−04 |

#### Hazard Ratio Estimates

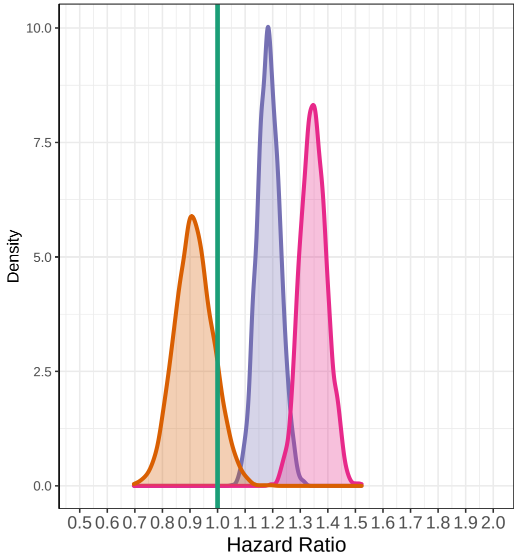

#### Median Time To Recovery Estimates

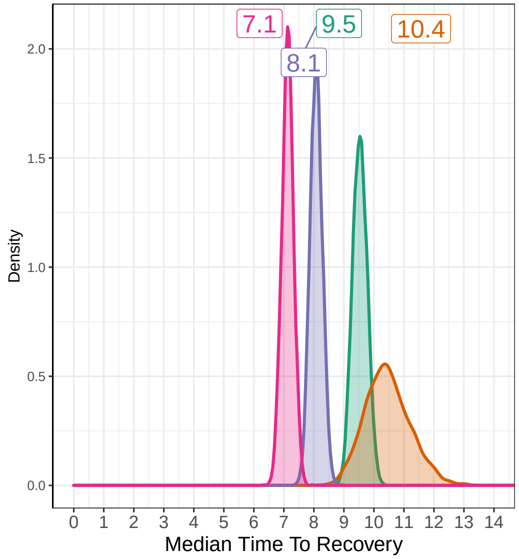

##### 6.3 Example 2

### April 1, 2020

Before Patient Enrollment Begins

#### Recovery Data

| | Enrolled | Complete | Recovered | Exposure Days | Recoveries Per Day | Estimated Hazard | Estimated HR (95% interval) | Estimated Median Survival Time |
|---|---|---|---|---|---|---|---|---|
| SOC | 0 | 0 | 0 | 0 | | | | |
| HCQ | 0 | 0 | 0 | 0 | | | | |
| Azith | 0 | 0 | 0 | 0 | | | | |
| Doxy | 0 | 0 | 0 | 0 | | | | |
| Total | 0 | 0 | 0 | 0 | | | | |

#### Hospitalization Data

| Hospitalizations | 28 Day Completers | Observed Hosp. Rate | Est. Hosp. Rate (95% interval) |
|---|---|---|---|
| 0 | 0 | | |
| 0 | 0 | | |
| 0 | 0 | | |
| 0 | 0 | | |
| 0 | 0 | | |

| Intervention Status |
|---|
| Enrolling |
| Enrolling |
| Unopened |
| Unopened |

#### Observed Patient Recoveries

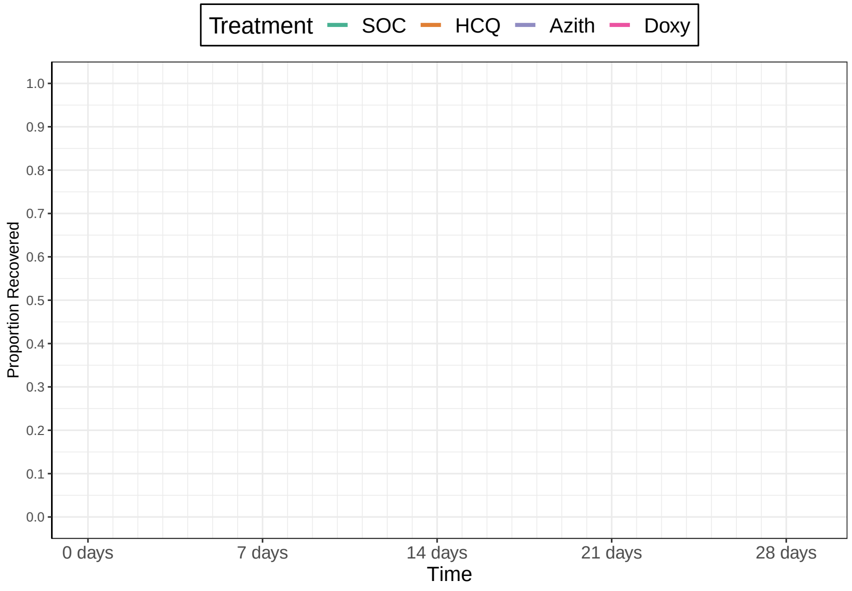

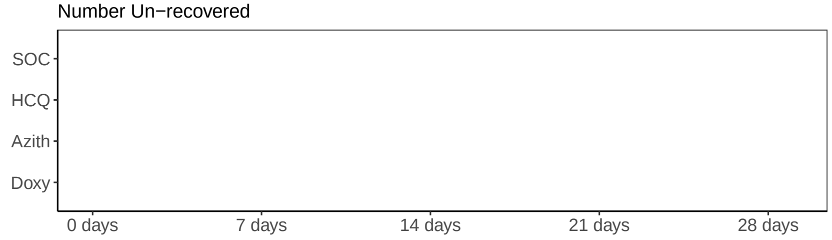

#### Recovery Inferences

| | Pr(Superiority) | Pr(Meaningful Effect) | Pr(Best) | Randomization Probability |
|---|---|---|---|---|
| SOC | | | | 0.5 |
| HCQ | | | | 0.5 |
| Azith | | | | 0 |
| Doxy | | | | 0 |

#### Hospitalization Inferences

| Pr(Superiority) | Pr(Meaningful Effect) |
|---|---|
| 0 | 0 |
| 0 | 0 |
| 0 | 0 |
| 0 | 0 |

#### Hazard Ratio Estimates

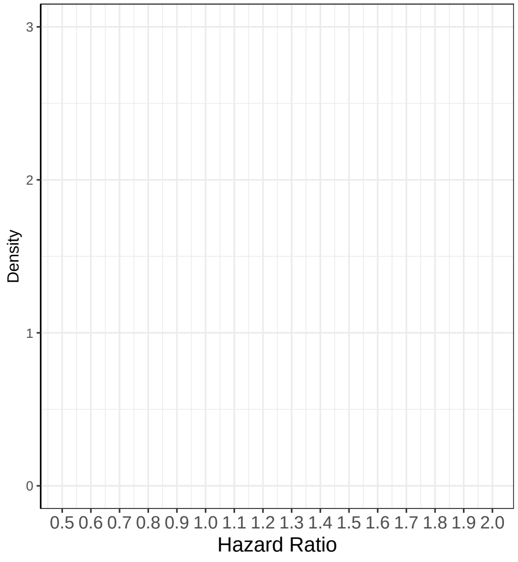

#### Median Time To Recovery Estimates

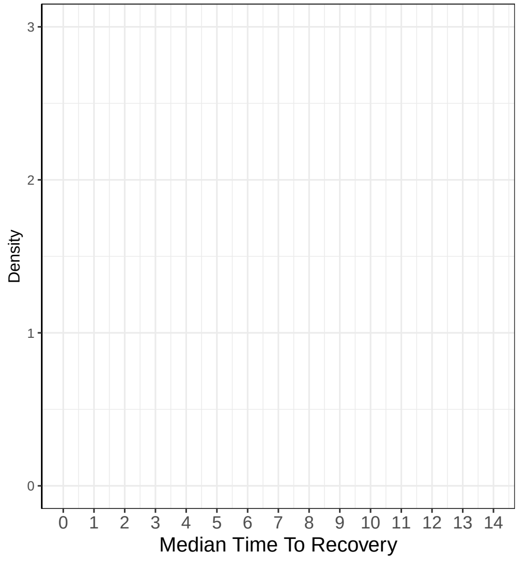

### Remove Drug on May 22, 2020

Recovery Data

| | Enrolled | Complete | Recovered | Exposure Days | Recoveries Per Day | Estimated Hazard | Estimated HR (95% interval) | Estimated Median Time to Recovery |
|---|---|---|---|---|---|---|---|---|
| SOC | 206 | 146 | 136 | 1807 | 0.075 | | | |
| HCQ | 207 | 149 | 143 | 1737 | 0.082 | | | |
| Azith | 0 | 0 | 0 | 0 | | | | |
| Doxy | 0 | 0 | 0 | 0 | | | | |
| Total | 413 | 295 | 279 | 3544 | 0.079 | | | |

Hospitalization Data

| Hospitalizations | 28 Day Completers | Observed Hosp. Rate | Est. Hosp. Rate (95% interval) |
|---|---|---|---|
| 1 | 78 | 0.0128 | |
| 10 | 79 | 0.1266 | |
| 0 | 0 | NA | |
| 0 | 0 | NA | |
| 11 | 157 | 0.0701 | |

| Intervention Status |
|---|
| Enrolling |
| Removing |
| Unopened |
| Unopened |

Observed Patient Recoveries

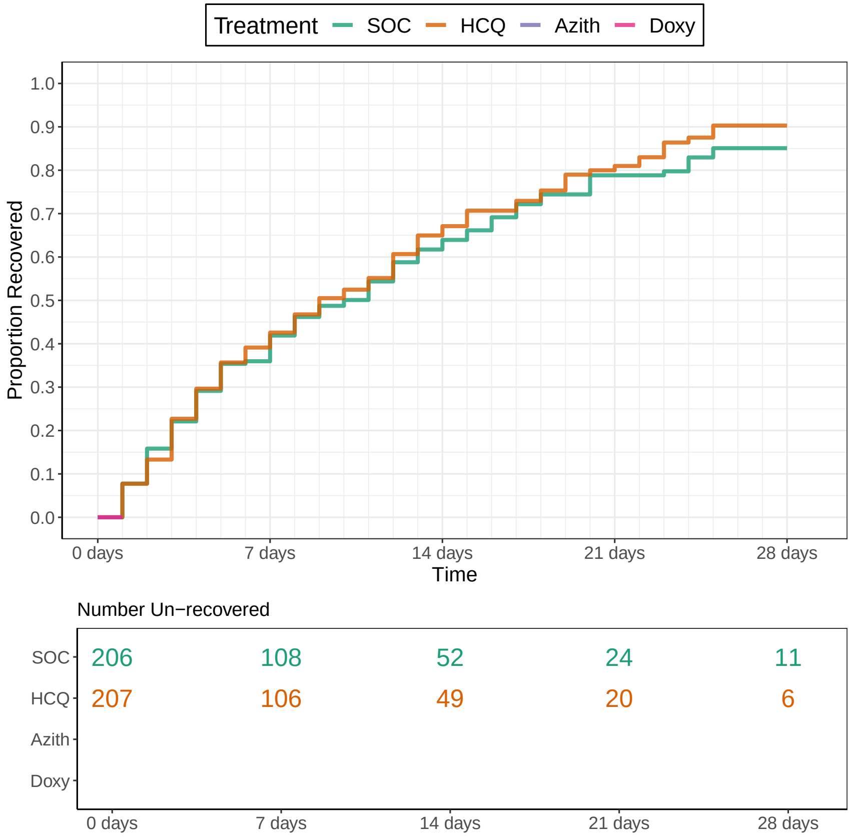

Recovery Inferences

| | Pr(Superiority) | Pr(Meaningful Effect) | Pr(Best) | Randomization Probability |
|---|---|---|---|---|
| SOC | | | | 1 |
| HCQ | | | 0 | 0 |
| Azith | | | 0 | 0 |
| Doxy | | | 0 | 0 |

Hospitalization Inferences

| Pr(Superiority) | Pr(Meaningful Effect) |
|---|---|

Hazard Ratio Estimates

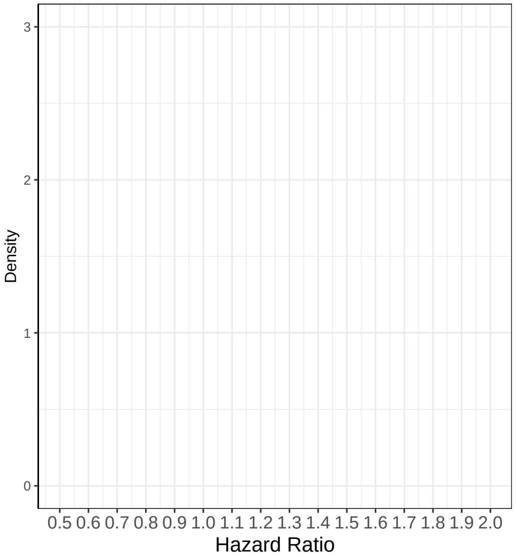

Median Time To Recovery Estimates

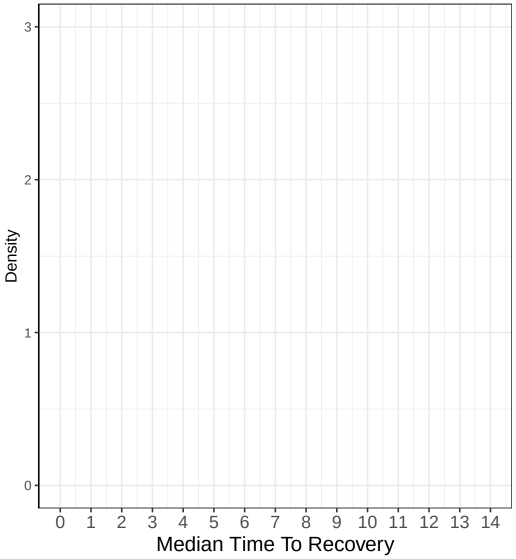

### Add Drug on May 23, 2020

Recovery Data

| | Enrolled | Complete | Recovered | Exposure Days | Recoveries Per Day | Estimated Hazard | Estimated HR (95% interval) | Estimated Median Time to Recovery |
|---|---|---|---|---|---|---|---|---|
| SOC | 208 | 151 | 140 | 1866 | 0.075 | | | |
| HCQ | 207 | 154 | 148 | 1793 | 0.083 | | | |
| Azith | 0 | 0 | 0 | 0 | | | | |
| Doxy | 0 | 0 | 0 | 0 | | | | |
| Total | 415 | 305 | 288 | 3659 | 0.079 | | | |

Hospitalization Data

| Hospitalizations | 28 Day Completers | Observed Hosp. Rate | Est. Hosp. Rate (95% interval) |
|---|---|---|---|
| 1 | 80 | 0.0125 | |
| 10 | 80 | 0.1250 | |
| 0 | 0 | NA | |
| 0 | 0 | NA | |
| 11 | 160 | 0.0688 | |

| Intervention Status |
|---|
| Enrolling |
| Paused |
| Introducing |
| Unopened |

Observed Patient Recoveries

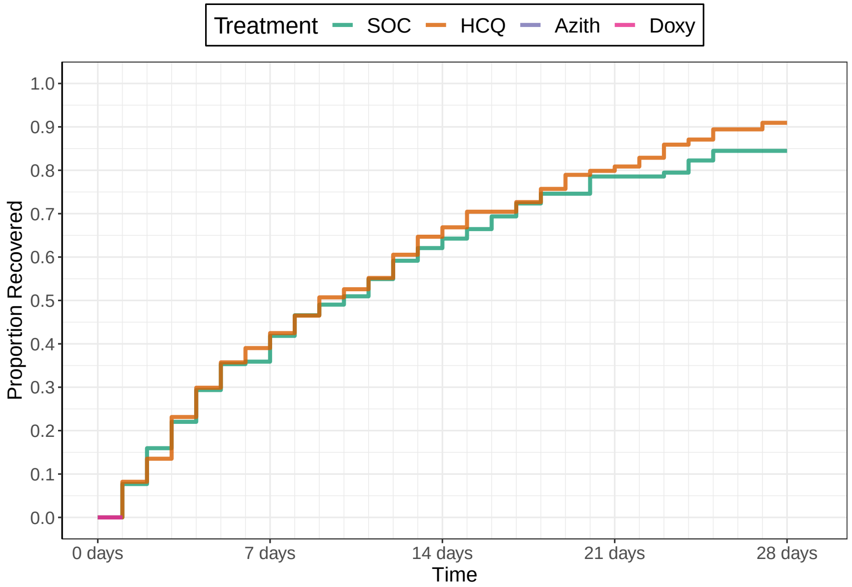

Number Un−recovered

| | 0 days | 7 days | 14 days | 21 days | 28 days |
|---|---|---|---|---|---|
| SOC | 208 | 108 | 52 | 24 | 11 |
| HCQ | 207 | 106 | 49 | 20 | 6 |
| Azith | | | | | |
| Doxy | | | | | |

Recovery Inferences

| | Pr(Superiority) | Pr(Meaningful Effect) | Pr(Best) | Randomization Probability |
|---|---|---|---|---|
| SOC | | | | 0.5 |
| HCQ | | | 0 | 0 |
| Azith | | | 1 | 0.5 |
| Doxy | | | 0 | 0 |

Hospitalization Inferences

| Pr(Superiority) | Pr(Meaningful Effect) |
|---|---|

Hazard Ratio Estimates

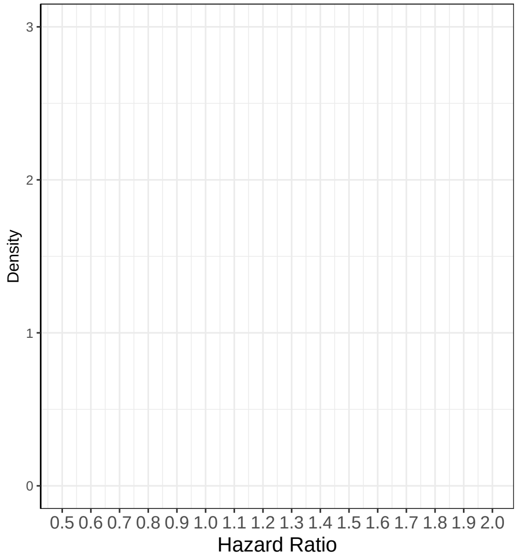

Median Time To Recovery Estimates

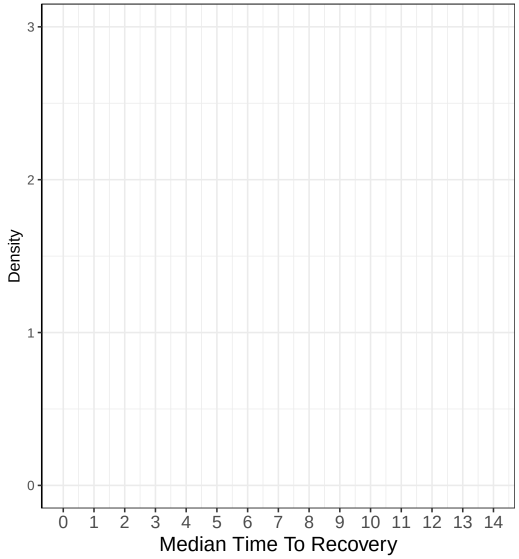

### Add Drug on July 23, 2020

Recovery Data

| | Enrolled | Complete | Recovered | Exposure Days | Recoveries Per Day | Estimated Hazard | Estimated HR (95% interval) | Estimated Median Time to Recovery |
|---|---|---|---|---|---|---|---|---|
| SOC | 326 | 308 | 270 | 3608 | 0.075 | | | |
| HCQ | 207 | 207 | 181 | 2375 | 0.076 | | | |
| Azith | 118 | 106 | 97 | 1148 | 0.084 | | | |
| Doxy | 0 | 0 | 0 | 0 | | | | |
| Total | 651 | 621 | 548 | 7132 | 0.077 | | | |

Hospitalization Data

| Hospitalizations | 28 Day Completers | Observed Hosp. Rate | Est. Hosp. Rate (95% interval) |
|---|---|---|---|
| 7 | 285 | 0.0246 | |
| 10 | 207 | 0.0483 | |
| 4 | 77 | 0.0519 | |
| 0 | 0 | NA | |
| 21 | 569 | 0.0369 | |

| Intervention Status |
|---|
| Enrolling |
| Paused |
| Introducing |
| Introducing |

Observed Patient Recoveries

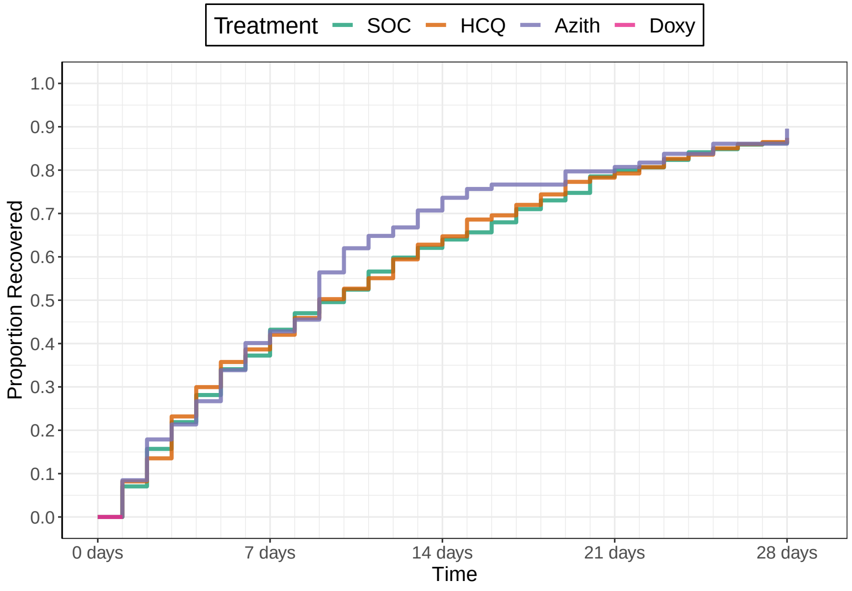

Number Un−recovered

| | 0 days | 7 days | 14 days | 21 days | 28 days |
|---|---|---|---|---|---|
| SOC | 326 | 200 | 118 | 62 | 39 |
| HCQ | 207 | 127 | 77 | 45 | 28 |
| Azith | 118 | 67 | 30 | 20 | 12 |
| Doxy | | | | | |

Recovery Inferences

| | Pr(Superiority) | Pr(Meaningful Effect) | Pr(Best) | Randomization Probability |
|---|---|---|---|---|
| SOC | | | | 0.33 |
| HCQ | | | 0 | 0 |
| Azith | | | 1 | 0.33 |
| Doxy | | | 0 | 0.33 |

Hospitalization Inferences

| Pr(Superiority) | Pr(Meaningful Effect) |
|---|---|

Hazard Ratio Estimates

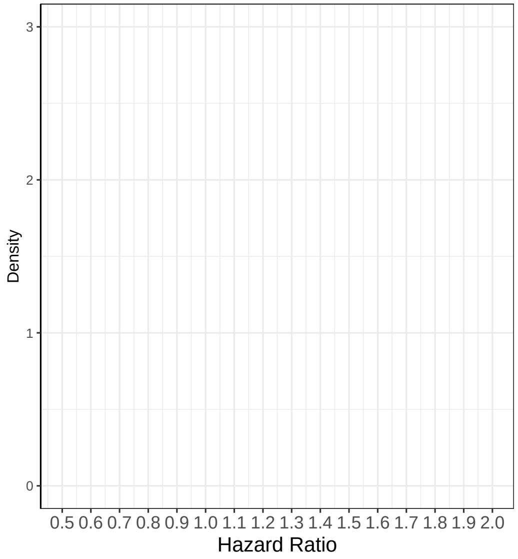

Median Time To Recovery Estimates

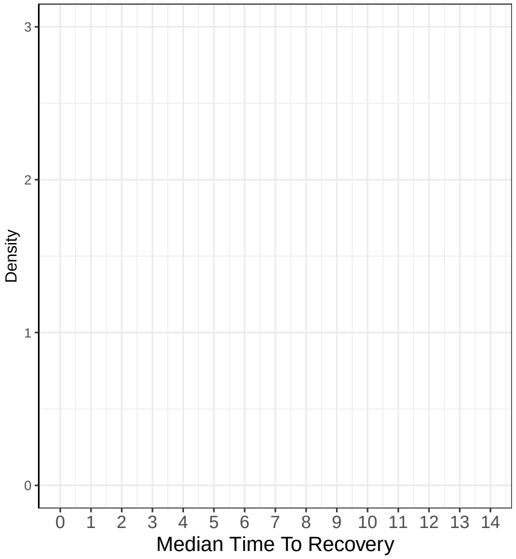

### Interim 1 on October 15, 2020

Recovery Data

| | Enrolled | Complete | Recovered | Exposure Days | Recoveries Per Day | Estimated Hazard | Estimated HR (95% interval) | Estimated Median Time to Recovery |
|---|---|---|---|---|---|---|---|---|
| SOC | 515 | 461 | 415 | 5407 | 0.077 | 0.074 | 1 | 9.22 |
| HCQ | 207 | 207 | 181 | 2375 | 0.076 | 0.073 | 0.985 (0.835, 1.162) | 9.36 |
| Azith | 309 | 256 | 234 | 2846 | 0.082 | 0.077 | 1.045 (0.907, 1.206) | 8.81 |
| Doxy | 179 | 138 | 133 | 1309 | 0.102 | 0.092 | 1.246 (1.029, 1.497) | 7.35 |
| Total | 1210 | 1062 | 963 | 11937 | 0.081 | | | |

Hospitalization Data

| Hospitalizations | 28 Day Completers | Observed Hosp. Rate | Est. Hosp. Rate (95% interval) |
|---|---|---|---|
| 19 | 400 | 0.0475 | 0.0387 (0.0238, 0.0573) |
| 10 | 207 | 0.0483 | 0.0528 (0.0274, 0.0866) |
| 6 | 192 | 0.0312 | 0.0223 (0.0089, 0.0407) |
| 12 | 74 | 0.1622 | 0.0713 (0.0387, 0.1129) |
| 47 | 873 | 0.0538 | |

| Intervention Status |
|---|
| Enrolling |
| Futility |
| Enrolling |
| Enrolling |

Observed Patient Recoveries

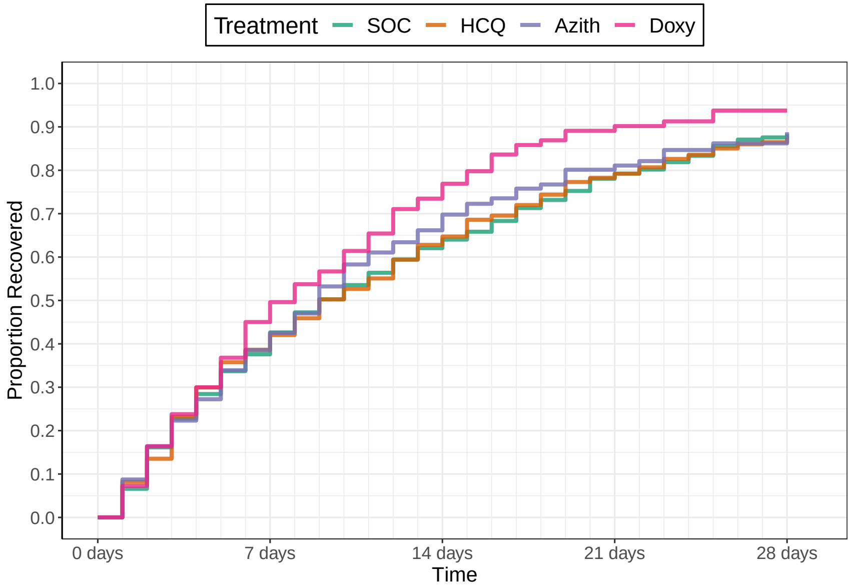

Number Un−recovered

| | 0 days | 7 days | 14 days | 21 days | 28 days |
|---|---|---|---|---|---|
| SOC | 515 | 297 | 173 | 93 | 49 |
| HCQ | 207 | 127 | 77 | 45 | 28 |
| Azith | 309 | 169 | 84 | 41 | 27 |
| Doxy | 179 | 84 | 31 | 10 | 5 |

Recovery Inferences

| | Pr(Superiority) | Pr(Meaningful Effect) | Pr(Best) | Randomization Probability |
|---|---|---|---|---|
| SOC | | | | 0.33 |
| HCQ | 0.4313 | 0.0207 | 0 | 0 |
| Azith | 0.7207 | 0.057 | 0.0362 | 0.28 |
| Doxy | 0.9853 | 0.6817 | 0.7138 | 0.39 |

Hospitalization Inferences

| Pr(Superiority) | Pr(Meaningful Effect) |
|---|---|
| 0.2122 | 0.0148 |
| 0.9192 | 0.3777 |
| 0.0478 | 0.0018 |

Hazard Ratio Estimates

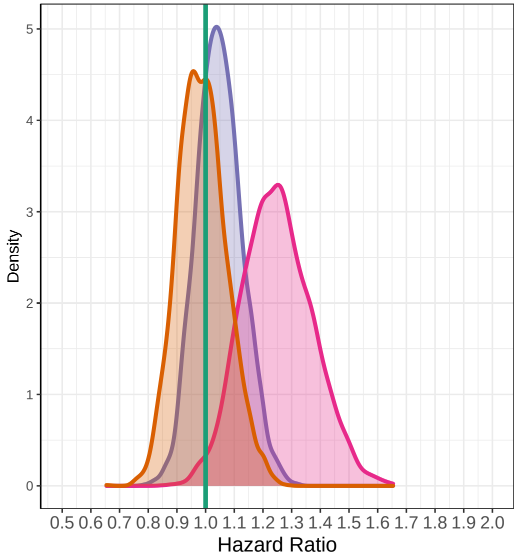

Median Time To Recovery Estimates

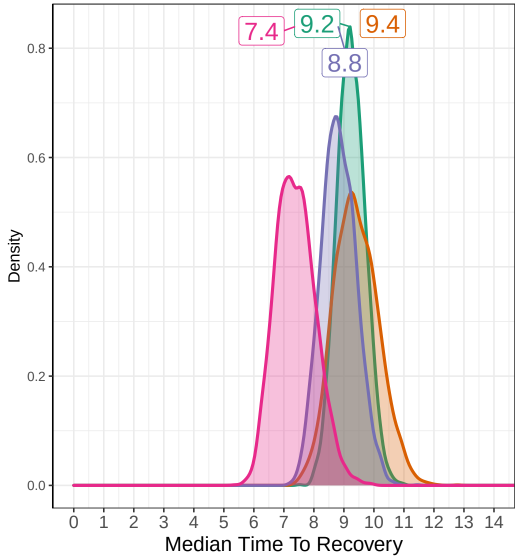

### Interim 2 on November 12, 2020

#### Recovery Data

| | Enrolled | Complete | Recovered | Exposure Days | Recoveries Per Day | Estimated Hazard | Estimated HR (95% interval) | Estimated Median Time to Recovery |
|---|---|---|---|---|---|---|---|---|
| SOC | 593 | 575 | 512 | 6654 | 0.077 | 0.074 | 1 | 9.23 |
| HCQ | 207 | 207 | 181 | 2375 | 0.076 | 0.073 | 0.981 (0.83, 1.152) | 9.43 |
| Azith | 375 | 351 | 321 | 3888 | 0.083 | 0.078 | 1.053 (0.923, 1.206) | 8.77 |
| Doxy | 273 | 243 | 233 | 2182 | 0.107 | 0.098 | 1.323 (1.143, 1.53) | 7.01 |
| Total | 1448 | 1376 | 1247 | 15100 | 0.083 | | | |

#### Hospitalization Data

| Hospitalizations | 28 Day Completers | Observed Hosp. Rate | Est. Hosp. Rate (95% interval) |
|---|---|---|---|
| 22 | 515 | 0.0427 | 0.0387 (0.0251, 0.0548) |
| 10 | 207 | 0.0483 | 0.0526 (0.0266, 0.0875) |
| 7 | 309 | 0.0227 | 0.0212 (0.0092, 0.0381) |
| 14 | 179 | 0.0782 | 0.054 (0.0308, 0.0841) |
| 53 | 1210 | 0.0438 | |

| Intervention Status |
|---|
| LostOnTTR |
| Dropped |
| InferiorToDoxy |
| SuccessOnTTRFutilityOnHosp *Announced* |

#### Observed Patient Recoveries

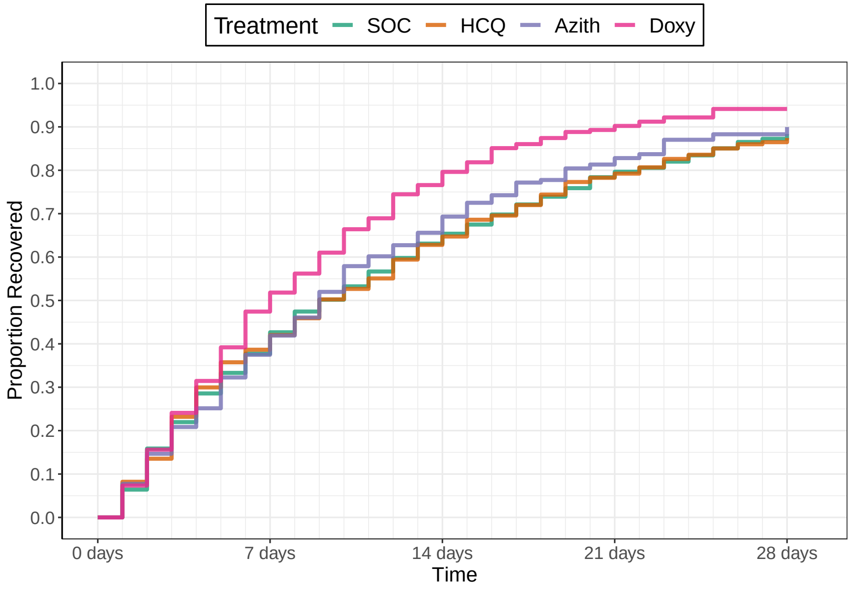

Number Un−recovered

| | 0 days | 7 days | 14 days | 21 days | 28 days |
|---|---|---|---|---|---|
| SOC | 593 | 365 | 210 | 120 | 69 |
| HCQ | 207 | 127 | 77 | 45 | 28 |
| Azith | 375 | 226 | 120 | 63 | 35 |
| Doxy | 273 | 132 | 54 | 23 | 12 |

#### Recovery Inferences

| | Pr(Superiority) | Pr(Meaningful Effect) | Pr(Best) | Randomization Probability |
|---|---|---|---|---|
| SOC | | | | 0.5 |
| HCQ | 0.4057 | 0.0127 | 0 | 0 |
| Azith | 0.772 | 0.0437 | 0 | 0 |
| Doxy | 0.9997 | 0.9037 | 0.75 | 0.5 |

#### Hospitalization Inferences

| Pr(Superiority) | Pr(Meaningful Effect) |
|---|---|
| 0.2158 | 0.0158 |
| 0.9422 | 0.4105 |
| 0.1645 | 0.0045 |

#### Hazard Ratio Estimates

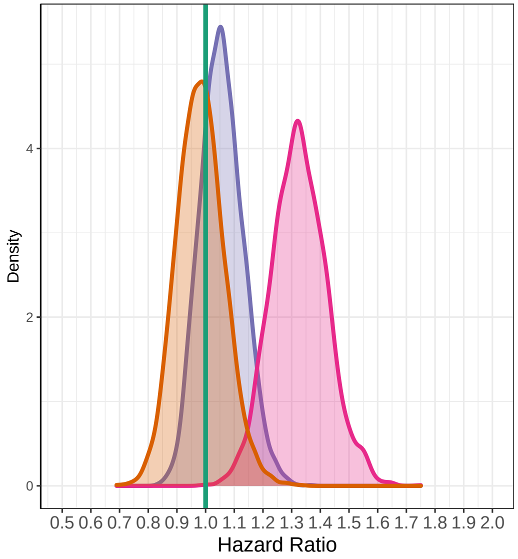

#### Median Time To Recovery Estimates

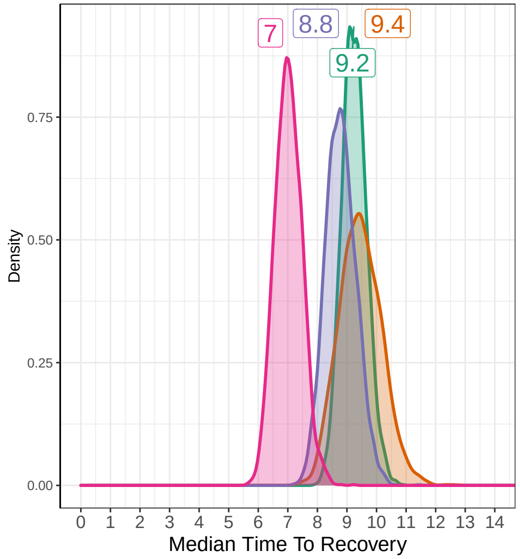

### Interim 3 on August 30, 2022

#### Recovery Data

| | Enrolled | Complete | Recovered | Exposure Days | Recoveries Per Day | Estimated Hazard | Estimated HR (95% interval) | Estimated Median Time to Recovery |
|---|---|---|---|---|---|---|---|---|
| SOC | 2851 | 2851 | 2499 | 33636 | 0.074 | 0.072 | 1 | 9.69 |
| HCQ | 207 | 207 | 181 | 2375 | 0.076 | 0.073 | 1.021 (0.876, 1.173) | 9.5 |
| Azith | 375 | 375 | 339 | 4072 | 0.083 | 0.08 | 1.11 (0.991, 1.238) | 8.75 |
| Doxy | 2567 | 2567 | 2417 | 23633 | 0.102 | 0.097 | 1.354 (1.282, 1.431) | 7.16 |
| Total | 6000 | 6000 | 5436 | 63716 | 0.085 | | | |

#### Hospitalization Data

| Hospitalizations | 28 Day Completers | Observed Hosp. Rate | Est. Hosp. Rate (95% interval) |
|---|---|---|---|
| 83 | 2851 | 0.0291 | 0.0294 (0.0235, 0.0362) |
| 10 | 207 | 0.0483 | 0.053 (0.0272, 0.0879) |
| 7 | 375 | 0.0187 | 0.0212 (0.0093, 0.0381) |
| 62 | 2567 | 0.0242 | 0.0246 (0.0188, 0.0306) |
| 162 | 6000 | 0.0270 | |

| Intervention Status |
|---|
| Complete |
| Dropped |
| Dropped |
| SuccessOnTTRFutilityOnHosp *Announced* |

#### Observed Patient Recoveries

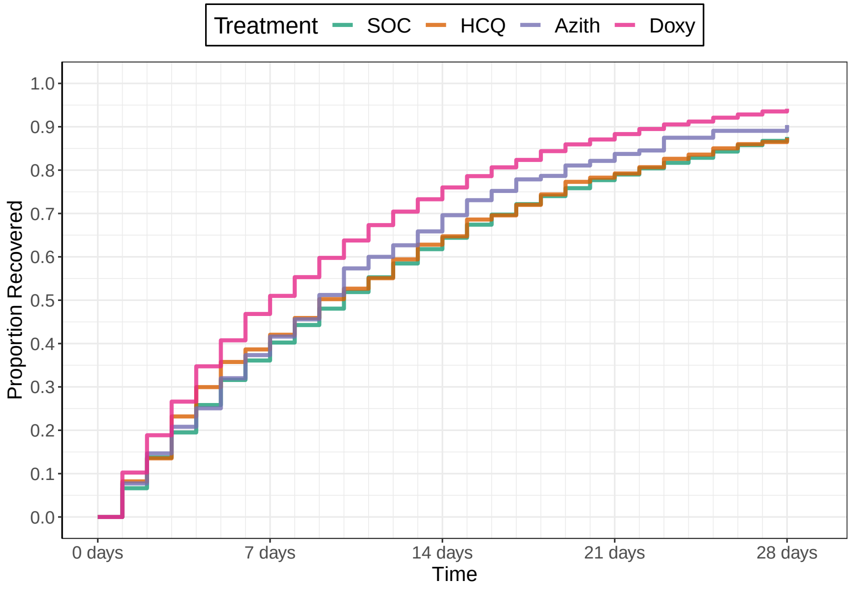

Number Un−recovered

| | 0 days | 7 days | 14 days | 21 days | 28 days |
|---|---|---|---|---|---|
| SOC | 2851 | 1822 | 1090 | 636 | 378 |
| HCQ | 207 | 127 | 77 | 45 | 28 |
| Azith | 375 | 235 | 128 | 67 | 41 |
| Doxy | 2567 | 1365 | 686 | 332 | 166 |

#### Recovery Inferences

| | Pr(Superiority) | Pr(Meaningful Effect) | Pr(Best) | Randomization Probability |
|---|---|---|---|---|
| SOC | | | | |
| HCQ | 0.6043 | 0.021 | 0 | |
| Azith | 0.966 | 0.1283 | 0 | |
| Doxy | 1 | 1 | 0.75 | |

#### Hospitalization Inferences

| Pr(Superiority) | Pr(Meaningful Effect) |
|---|---|
| 0.0495 | 0 |
| 0.8518 | 0.056 |
| 0.866 | 2e−04 |

#### Hazard Ratio Estimates

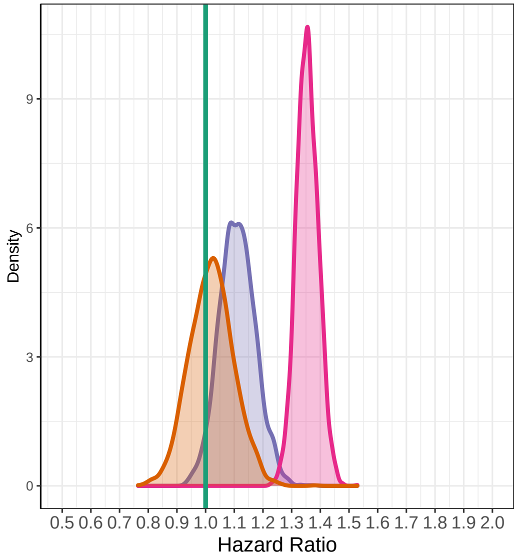

#### Median Time To Recovery Estimates

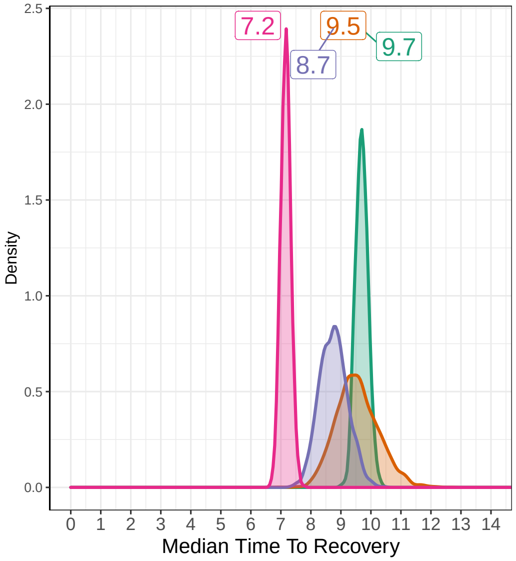

#### 6.4 Example 3

### April 1, 2020

Before Patient Enrollment Begins

#### Recovery Data

| | Enrolled | Complete | Recovered | Exposure Days | Recoveries Per Day | Estimated Hazard | Estimated HR (95% interval) | Estimated Median Survival Time |
|---|---|---|---|---|---|---|---|---|
| SOC | 0 | 0 | 0 | 0 | | | | |
| HCQ | 0 | 0 | 0 | 0 | | | | |
| Azith | 0 | 0 | 0 | 0 | | | | |
| Doxy | 0 | 0 | 0 | 0 | | | | |
| Total | 0 | 0 | 0 | 0 | | | | |

#### Hospitalization Data

| Hospitalizations | 28 Day Completers | Observed Hosp. Rate | Est. Hosp. Rate (95% interval) |
|---|---|---|---|
| 0 | 0 | | |
| 0 | 0 | | |
| 0 | 0 | | |
| 0 | 0 | | |
| 0 | 0 | | |

| Intervention Status |
|---|
| Enrolling |
| Enrolling |
| Unopened |
| Unopened |

#### Observed Patient Recoveries

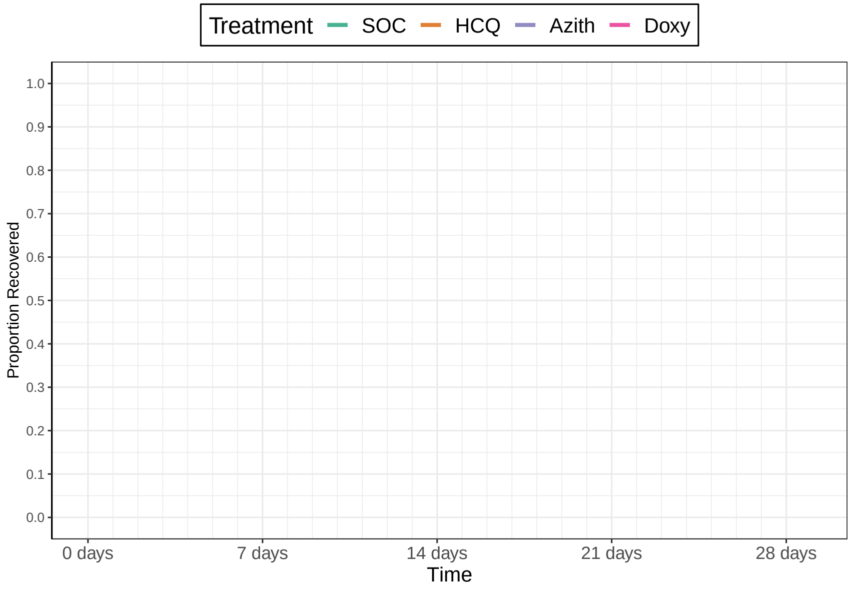

Number Un−recovered

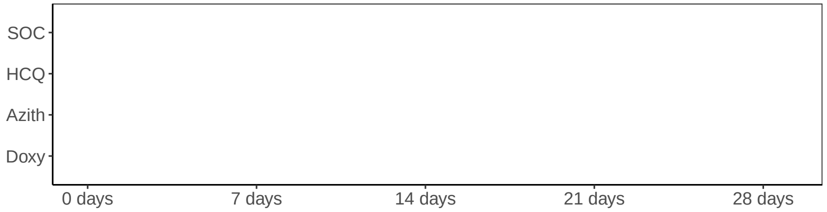

#### Recovery Inferences

| | Pr(Superiority) | Pr(Meaningful Effect) | Pr(Best) | Randomization Probability |
|---|---|---|---|---|
| SOC | | | | 0.5 |
| HCQ | | | | 0.5 |
| Azith | | | | 0 |
| Doxy | | | | 0 |

#### Hospitalization Inferences

| Pr(Superiority) | Pr(Meaningful Effect) |
|---|---|
| 0 | 0 |
| 0 | 0 |
| 0 | 0 |
| 0 | 0 |

#### Hazard Ratio Estimates

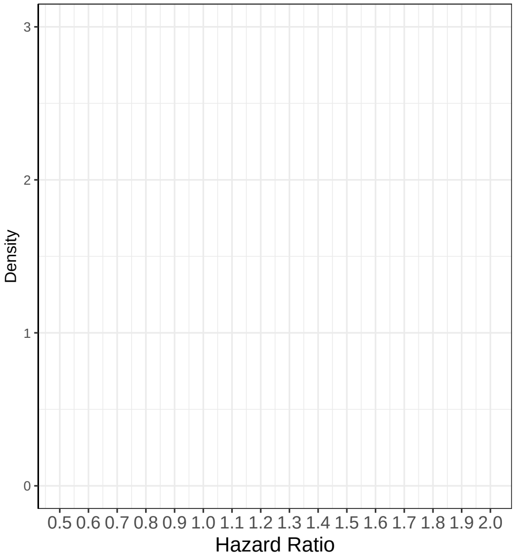

#### Median Time To Recovery Estimates

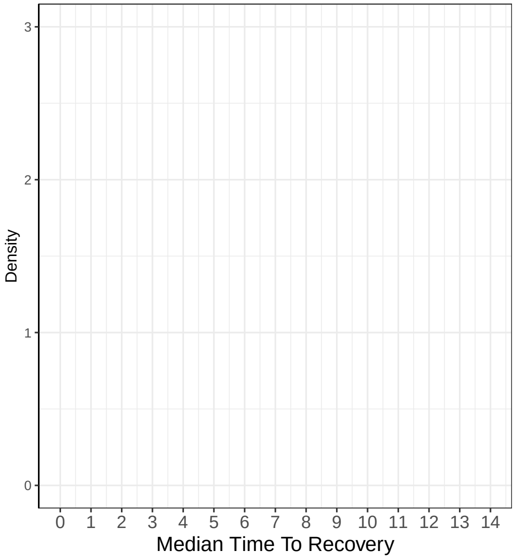

### Remove Drug on May 22, 2020

#### Recovery Data

| | Enrolled | Complete | Recovered | Exposure Days | Recoveries Per Day | Estimated Hazard | Estimated HR (95% interval) | Estimated Median Time to Recovery |
|---|---|---|---|---|---|---|---|---|
| SOC | 206 | 150 | 141 | 1753 | 0.08 | | | |
| HCQ | 207 | 156 | 147 | 1684 | 0.087 | | | |
| Azith | 0 | 0 | 0 | 0 | | | | |
| Doxy | 0 | 0 | 0 | 0 | | | | |
| Total | 413 | 306 | 288 | 3437 | 0.084 | | | |

#### Hospitalization Data

| Hospitalizations | 28 Day Completers | Observed Hosp. Rate | Est. Hosp. Rate (95% interval) |
|---|---|---|---|
| 5 | 78 | 0.0641 | |
| 7 | 79 | 0.0886 | |
| 0 | 0 | NA | |
| 0 | 0 | NA | |
| 12 | 157 | 0.0764 | |

| Intervention Status |
|---|
| Enrolling |
| Removing |
| Unopened |
| Unopened |

#### Observed Patient Recoveries

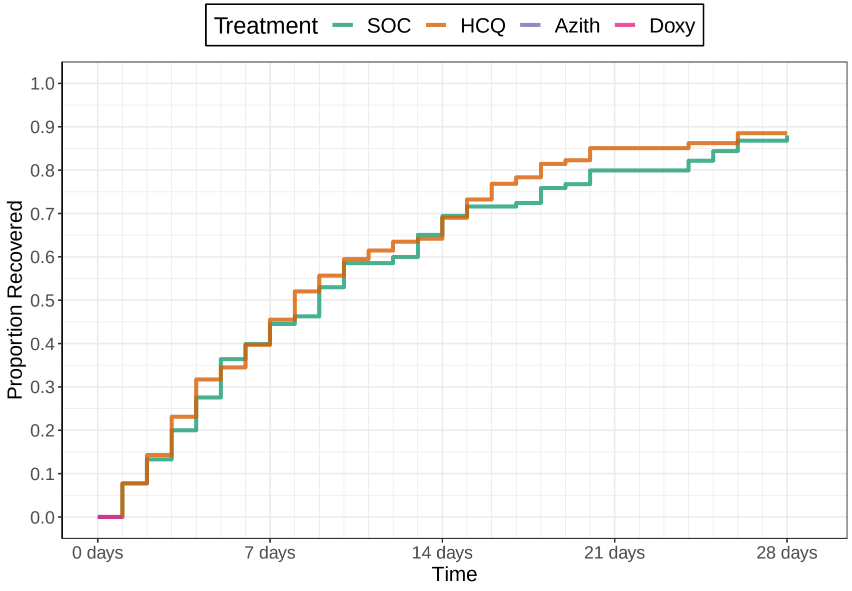

Number Un−recovered

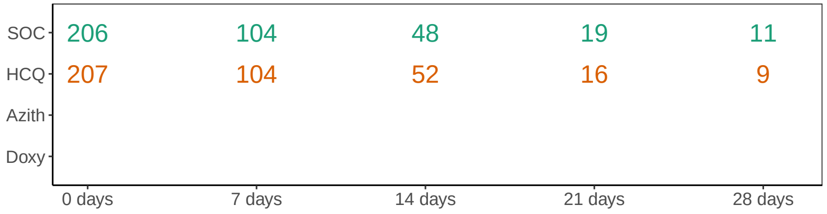

#### Recovery Inferences

| | Pr(Superiority) | Pr(Meaningful Effect) | Pr(Best) | Randomization Probability |
|---|---|---|---|---|
| SOC | | | | 1 |
| HCQ | | | 0 | 0 |
| Azith | | | 0 | 0 |
| Doxy | | | 0 | 0 |

#### Hospitalization Inferences

| Pr(Superiority) | Pr(Meaningful Effect) |
|---|---|

#### Hazard Ratio Estimates

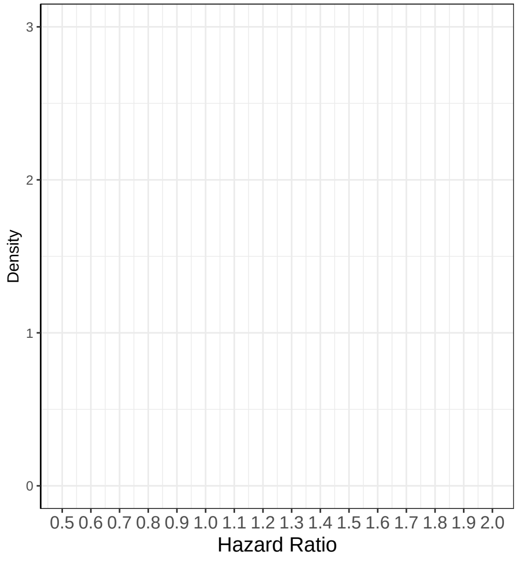

#### Median Time To Recovery Estimates

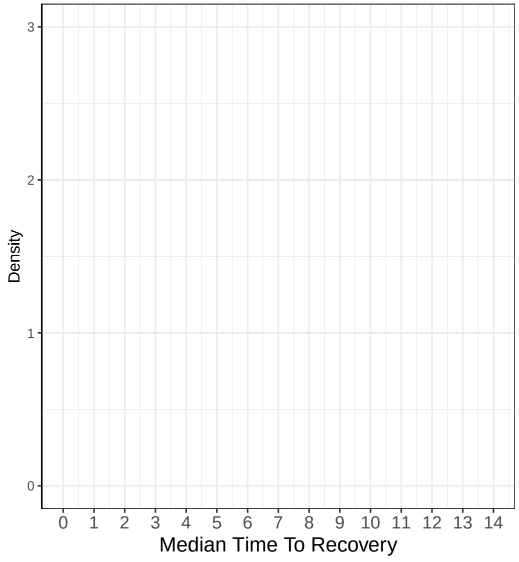

### Add Drug on May 23, 2020

#### Recovery Data

| | Enrolled | Complete | Recovered | Exposure Days | Recoveries Per Day | Estimated Hazard | Estimated HR (95% interval) | Estimated Median Time to Recovery |
|---|---|---|---|---|---|---|---|---|
| SOC | 208 | 155 | 145 | 1808 | 0.08 | | | |
| HCQ | 207 | 166 | 157 | 1728 | 0.091 | | | |
| Azith | 0 | 0 | 0 | 0 | | | | |
| Doxy | 0 | 0 | 0 | 0 | | | | |
| Total | 415 | 321 | 302 | 3536 | 0.085 | | | |

#### Hospitalization Data

| Hospitalizations | 28 Day Completers | Observed Hosp. Rate | Est. Hosp. Rate (95% interval) |
|---|---|---|---|
| 5 | 80 | 0.0625 | |
| 7 | 80 | 0.0875 | |
| 0 | 0 | NA | |
| 0 | 0 | NA | |
| 12 | 160 | 0.0750 | |

| Intervention Status |
|---|
| Enrolling |
| Paused |
| Introducing |
| Unopened |

#### Observed Patient Recoveries

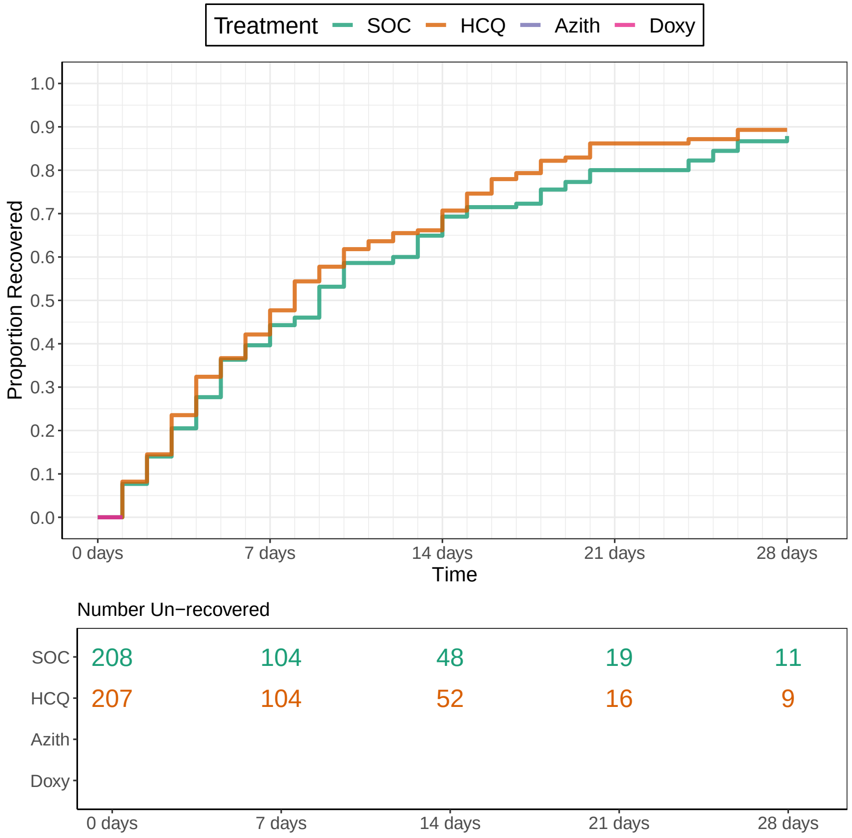

#### Recovery Inferences

| | Pr(Superiority) | Pr(Meaningful Effect) | Pr(Best) | Randomization Probability |
|---|---|---|---|---|
| SOC | | | | 0.5 |
| HCQ | | | 0 | 0 |
| Azith | | | 1 | 0.5 |
| Doxy | | | 0 | 0 |

#### Hospitalization Inferences

| Pr(Superiority) | Pr(Meaningful Effect) |
|---|---|

#### Hazard Ratio Estimates

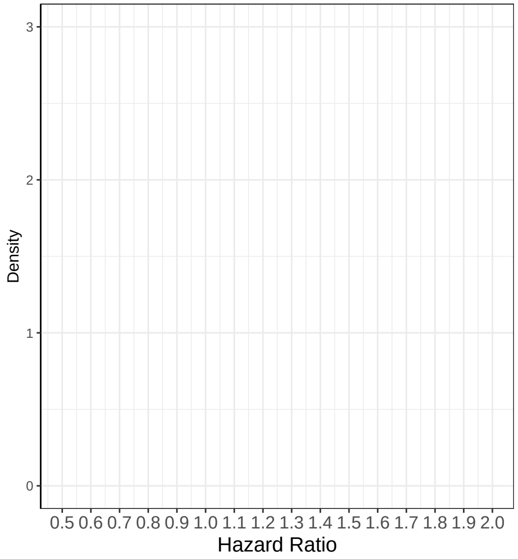

#### Median Time To Recovery Estimates

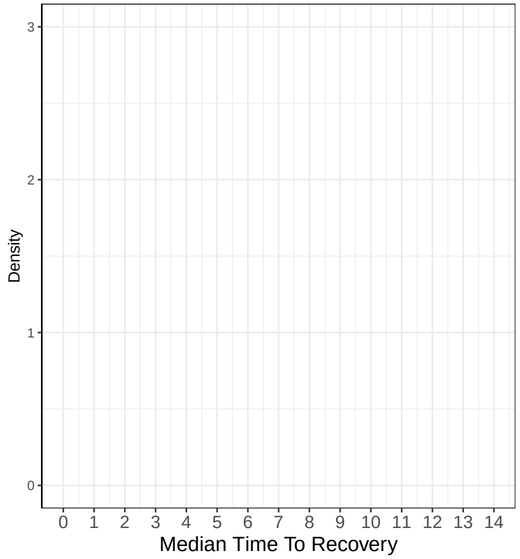

### Add Drug on July 23, 2020

Recovery Data

| | Enrolled | Complete | Recovered | Exposure Days | Recoveries Per Day | Estimated Hazard | Estimated HR (95% interval) | Estimated Median Time to Recovery |
|---|---|---|---|---|---|---|---|---|
| SOC | 326 | 305 | 275 | 3462 | 0.079 | | | |
| HCQ | 207 | 207 | 188 | 2127 | 0.088 | | | |
| Azith | 118 | 105 | 97 | 1041 | 0.093 | | | |
| Doxy | 0 | 0 | 0 | 0 | | | | |
| Total | 651 | 617 | 560 | 6630 | 0.084 | | | |

Hospitalization Data

| Hospitalizations | 28 Day Completers | Observed Hosp. Rate | Est. Hosp. Rate (95% interval) |
|---|---|---|---|
| 6 | 285 | 0.0211 | |
| 7 | 207 | 0.0338 | |
| 3 | 77 | 0.0390 | |
| 0 | 0 | NA | |
| 16 | 569 | 0.0281 | |

| Intervention Status |
|---|
| Enrolling |
| Paused |
| Introducing |
| Introducing |

Observed Patient Recoveries

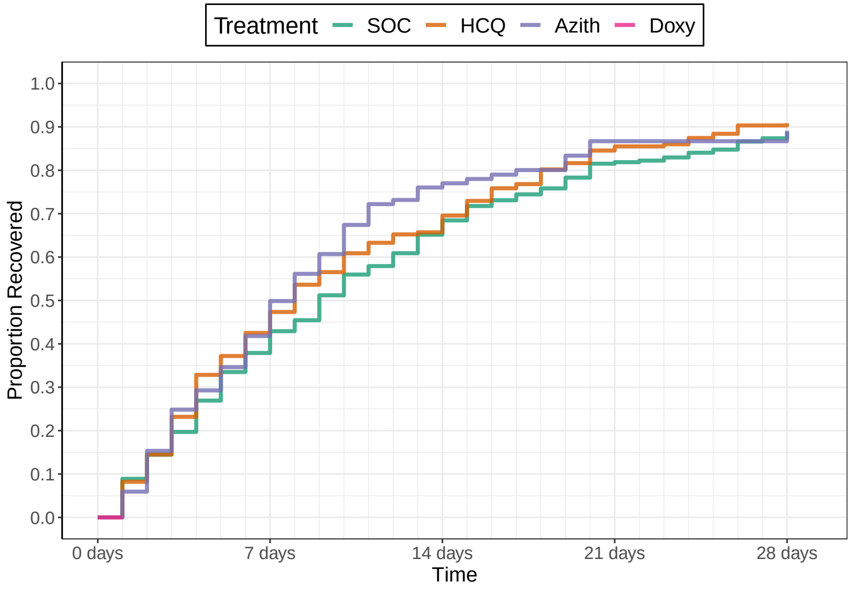

Number Un−recovered

| | 0 days | 7 days | 14 days | 21 days | 28 days |
|---|---|---|---|---|---|
| SOC | 326 | 198 | 106 | 52 | 34 |
| HCQ | 207 | 119 | 71 | 32 | 20 |
| Azith | 118 | 65 | 25 | 12 | 11 |
| Doxy | | | | | |

Recovery Inferences

| | Pr(Superiority) | Pr(Meaningful Effect) | Pr(Best) | Randomization Probability |
|---|---|---|---|---|
| SOC | | | | 0.33 |
| HCQ | | | 0 | 0 |
| Azith | | | 1 | 0.33 |
| Doxy | | | 0 | 0.33 |

Hospitalization Inferences

| Pr(Superiority) | Pr(Meaningful Effect) |
|---|---|

Hazard Ratio Estimates

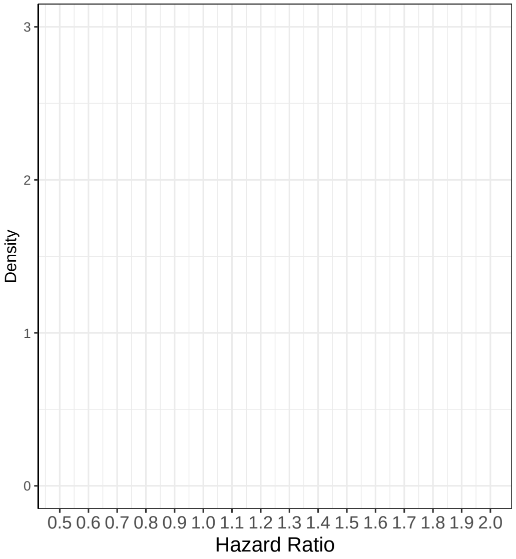

Median Time To Recovery Estimates

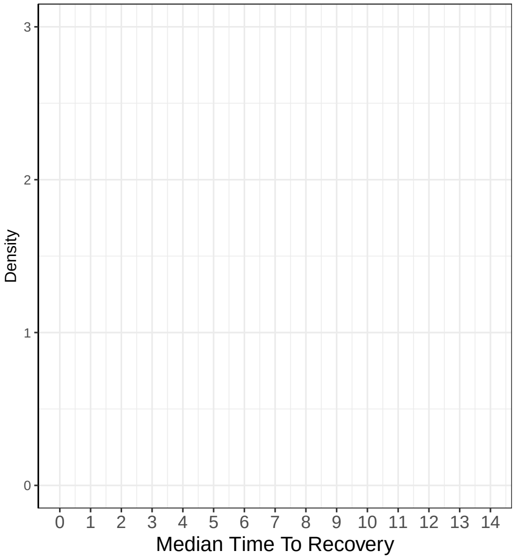

### Interim 1 on October 15, 2020

Recovery Data

| | Enrolled | Complete | Recovered | Exposure Days | Recoveries Per Day | Estimated Hazard | Estimated HR (95% interval) | Estimated Median Time to Recovery |
|---|---|---|---|---|---|---|---|---|
| SOC | 519 | 477 | 433 | 5488 | 0.079 | 0.073 | 1 | 8.96 |
| HCQ | 207 | 207 | 188 | 2127 | 0.088 | 0.08 | 1.098 (0.937, 1.295) | 8.18 |
| Azith | 317 | 262 | 242 | 2746 | 0.088 | 0.079 | 1.083 (0.928, 1.255) | 8.28 |
| Doxy | 167 | 118 | 108 | 1301 | 0.083 | 0.075 | 1.019 (0.831, 1.239) | 8.79 |
| Total | 1210 | 1064 | 971 | 11662 | 0.083 | | | |

Hospitalization Data

| Hospitalizations | 28 Day Completers | Observed Hosp. Rate | Est. Hosp. Rate (95% interval) |
|---|---|---|---|
| 14 | 419 | 0.0334 | 0.0287 (0.0162, 0.0445) |
| 7 | 207 | 0.0338 | 0.038 (0.0165, 0.0671) |
| 9 | 185 | 0.0486 | 0.0314 (0.0155, 0.0526) |
| 6 | 62 | 0.0968 | 0.0409 (0.017, 0.0745) |
| 36 | 873 | 0.0412 | |

| Intervention Status |
|---|
| Enrolling |
| Futility |
| Enrolling |
| Enrolling |

Observed Patient Recoveries

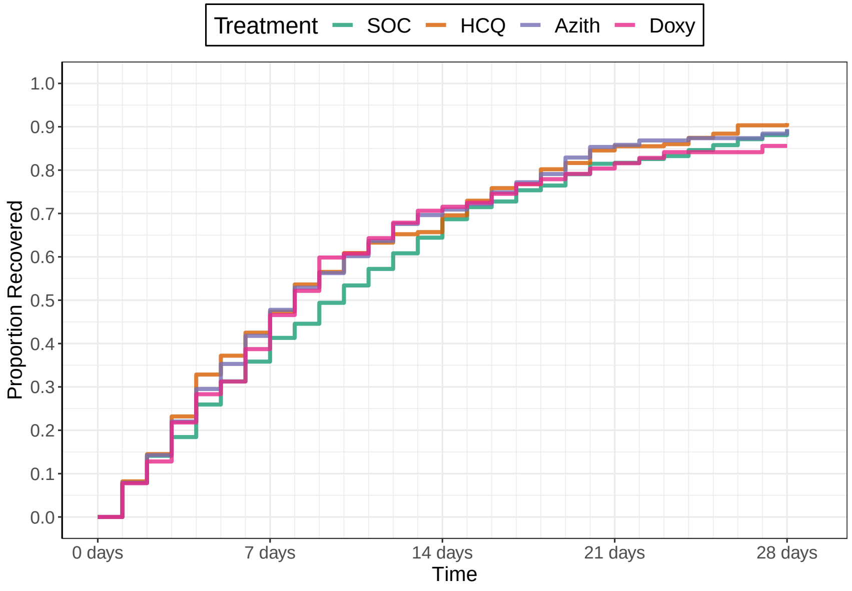

Number Un−recovered

| | 0 days | 7 days | 14 days | 21 days | 28 days |
|---|---|---|---|---|---|
| SOC | 519 | 317 | 167 | 84 | 50 |
| HCQ | 207 | 119 | 71 | 32 | 20 |
| Azith | 317 | 165 | 72 | 30 | 22 |
| Doxy | 167 | 78 | 32 | 16 | 10 |

Recovery Inferences

| | Pr(Superiority) | Pr(Meaningful Effect) | Pr(Best) | Randomization Probability |
|---|---|---|---|---|
| SOC | | | | 0.33 |
| HCQ | 0.8727 | 0.1377 | 0 | 0 |
| Azith | 0.8553 | 0.0917 | 0.525 | 0.4 |
| Doxy | 0.5723 | 0.0587 | 0.225 | 0.27 |

Hospitalization Inferences

| Pr(Superiority) | Pr(Meaningful Effect) |
|---|---|
| 0.2815 | 0.0148 |
| 0.4105 | 0.0232 |
| 0.2388 | 0.0127 |

Hazard Ratio Estimates

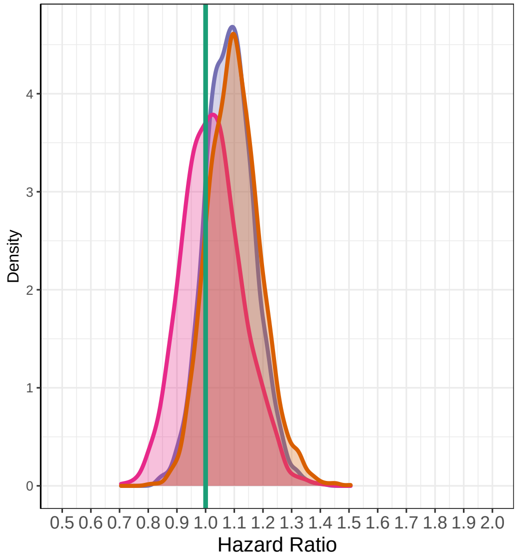

Median Time To Recovery Estimates

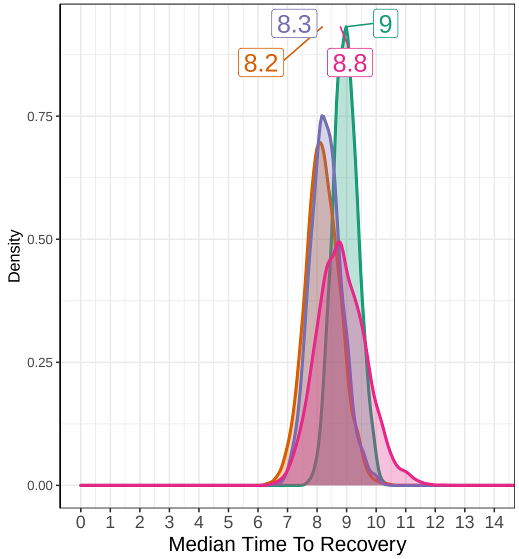

### Interim 2 on November 12, 2020

##### Recovery Data

| | Enrolled | Complete | Recovered | Exposure Days | Recoveries Per Day | Estimated Hazard | Estimated HR (95% interval) | Estimated Median Time to Recovery |
|---|---|---|---|---|---|---|---|---|
| SOC | 605 | 574 | 522 | 6588 | 0.079 | 0.076 | 1 | 9.04 |
| HCQ | 207 | 207 | 188 | 2127 | 0.088 | 0.083 | 1.094 (0.932, 1.284) | 8.3 |
| Azith | 417 | 392 | 362 | 3924 | 0.092 | 0.086 | 1.139 (1.001, 1.305) | 7.98 |
| Doxy | 227 | 204 | 187 | 2176 | 0.086 | 0.08 | 1.065 (0.911, 1.25) | 8.5 |
| Total | 1456 | 1377 | 1259 | 14815 | 0.085 | | | |

##### Hospitalization Data

| Hospitalizations | 28 Day Completers | Observed Hosp. Rate | Est. Hosp. Rate (95% interval) |
|---|---|---|---|
| 18 | 519 | 0.0347 | 0.0311 (0.0189, 0.0462) |
| 7 | 207 | 0.0338 | 0.0382 (0.0162, 0.0686) |
| 12 | 317 | 0.0379 | 0.0309 (0.0164, 0.0497) |
| 9 | 167 | 0.0539 | 0.0436 (0.0213, 0.0734) |
| 46 | 1210 | 0.0380 | |

| Intervention Status |
|---|
| Enrolling |
| Dropped |
| Enrolling |
| Enrolling |

##### Observed Patient Recoveries

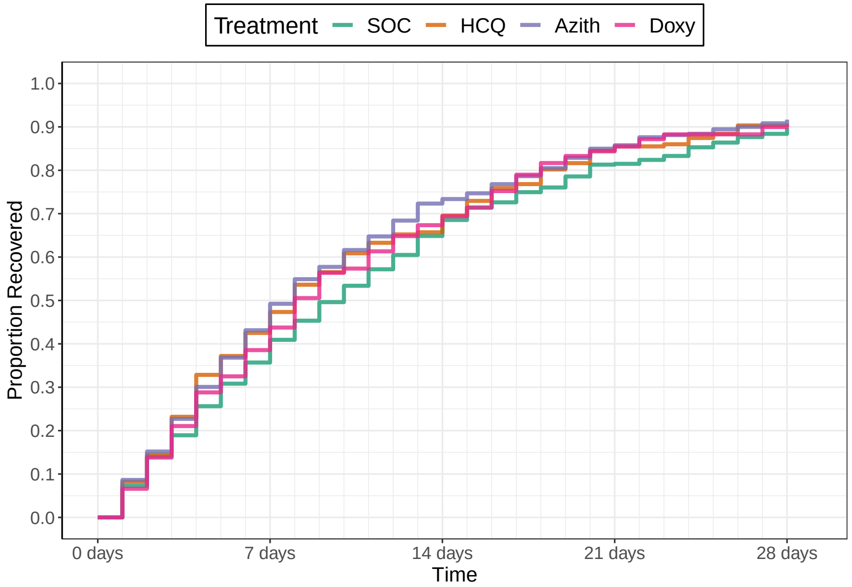

Number Un−recovered

| | 0 days | 7 days | 14 days | 21 days | 28 days |
|---|---|---|---|---|---|
| SOC | 605 | 381 | 200 | 103 | 62 |
| HCQ | 207 | 119 | 71 | 32 | 20 |
| Azith | 417 | 224 | 106 | 57 | 33 |
| Doxy | 227 | 130 | 64 | 28 | 18 |

##### Recovery Inferences

| | Pr(Superiority) | Pr(Meaningful Effect) | Pr(Best) | Randomization Probability |
|---|---|---|---|---|
| SOC | | | | 0.33 |
| HCQ | 0.8647 | 0.133 | 0 | 0 |
| Azith | 0.9757 | 0.22 | 0.5913 | 0.37 |
| Doxy | 0.7733 | 0.077 | 0.1588 | 0.29 |

##### Hospitalization Inferences

| Pr(Superiority) | Pr(Meaningful Effect) |
|---|---|
| 0.3355 | 0.018 |
| 0.5128 | 0.029 |
| 0.205 | 0.0102 |

##### Hazard Ratio Estimates

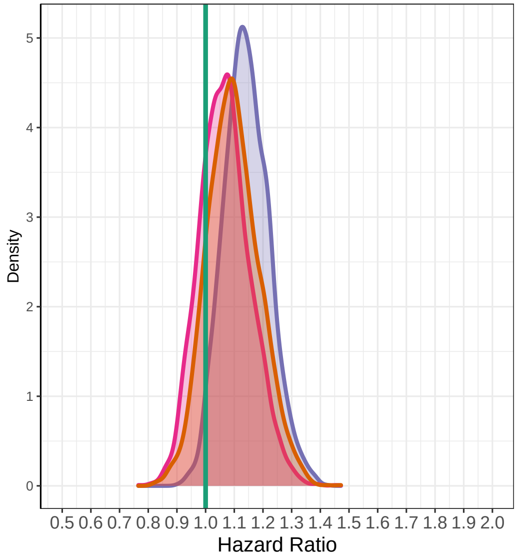

##### Median Time To Recovery Estimates

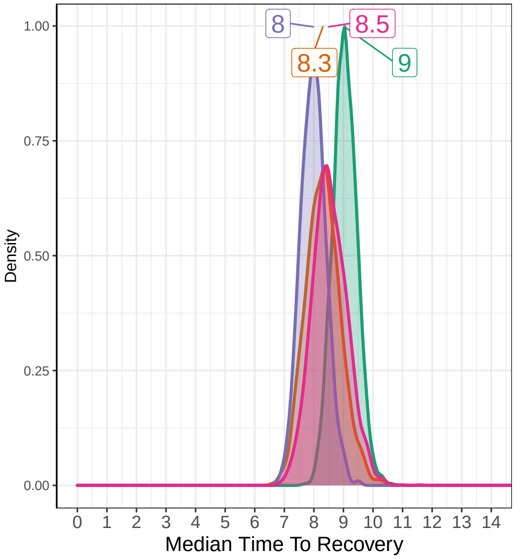

### Interim 3 on December 10, 2020

#### Recovery Data

| | Enrolled | Complete | Recovered | Exposure Days | Recoveries Per Day | Estimated Hazard | Estimated HR (95% interval) | Estimated Median Time to Recovery |
|---|---|---|---|---|---|---|---|---|
| SOC | 668 | 636 | 573 | 7328 | 0.078 | 0.075 | 1 | 9.14 |
| HCQ | 207 | 207 | 188 | 2127 | 0.088 | 0.083 | 1.107 (0.945, 1.296) | 8.29 |
| Azith | 472 | 453 | 419 | 4606 | 0.091 | 0.086 | 1.136 (1.01, 1.286) | 8.07 |
| Doxy | 289 | 264 | 245 | 2946 | 0.083 | 0.079 | 1.042 (0.904, 1.204) | 8.76 |
| Total | 1636 | 1560 | 1425 | 17008 | 0.084 | | | |

#### Hospitalization Data

| Hospitalizations | 28 Day Completers | Observed Hosp. Rate | Est. Hosp. Rate (95% interval) |
|---|---|---|---|
| 22 | 605 | 0.0364 | 0.0344 (0.0222, 0.0488) |
| 7 | 207 | 0.0338 | 0.0388 (0.0169, 0.0687) |
| 15 | 417 | 0.0360 | 0.0337 (0.0194, 0.0518) |
| 9 | 227 | 0.0396 | 0.0341 (0.0168, 0.0574) |
| 53 | 1456 | 0.0364 | |

| Intervention Status |
|---|
| Enrolling |
| Dropped |
| Enrolling |
| Futility |

#### Observed Patient Recoveries

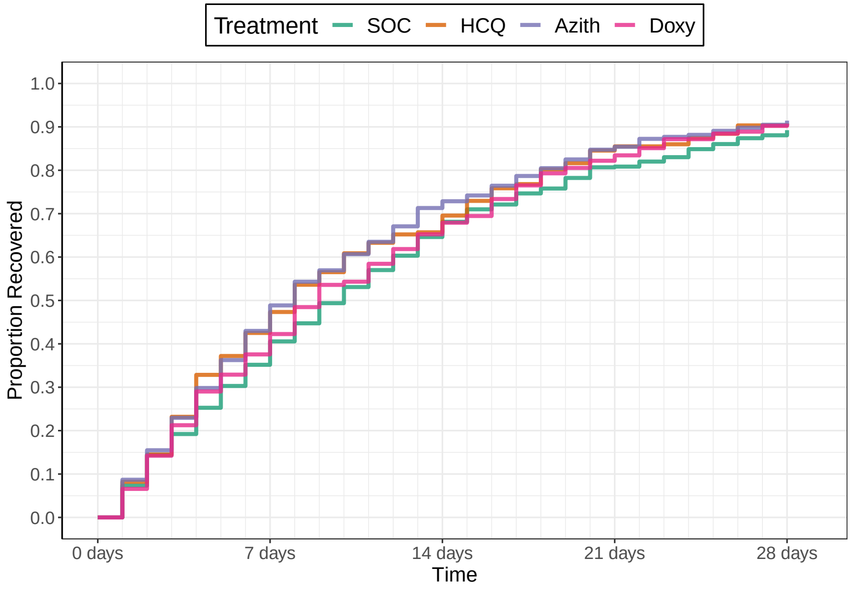

Number Un−recovered

| | 0 days | 7 days | 14 days | 21 days | 28 days |
|---|---|---|---|---|---|
| SOC | 668 | 421 | 222 | 118 | 71 |
| HCQ | 207 | 119 | 71 | 32 | 20 |
| Azith | 472 | 262 | 129 | 68 | 40 |
| Doxy | 289 | 173 | 91 | 43 | 21 |

#### Recovery Inferences

| | Pr(Superiority) | Pr(Meaningful Effect) | Pr(Best) | Randomization Probability |
|---|---|---|---|---|
| SOC | | | | 0.5 |
| HCQ | 0.8873 | 0.153 | 0 | 0 |
| Azith | 0.9803 | 0.2037 | 0.75 | 0.5 |
| Doxy | 0.7243 | 0.034 | 0 | 0 |

#### Hospitalization Inferences

| Pr(Superiority) | Pr(Meaningful Effect) |
|---|---|
| 0.4158 | 0.04 |
| 0.5435 | 0.0325 |
| 0.532 | 0.0452 |

#### Hazard Ratio Estimates

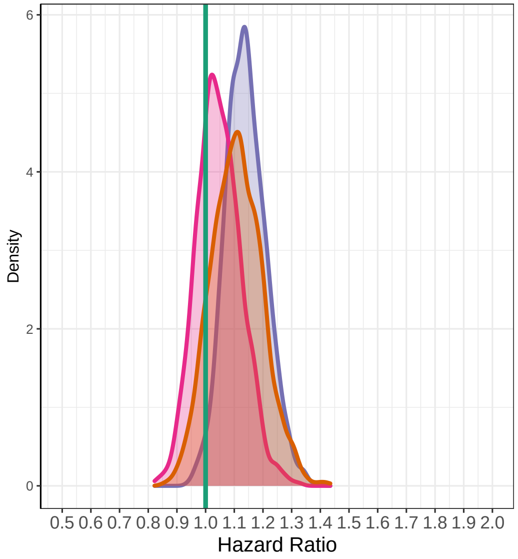

#### Median Time To Recovery Estimates

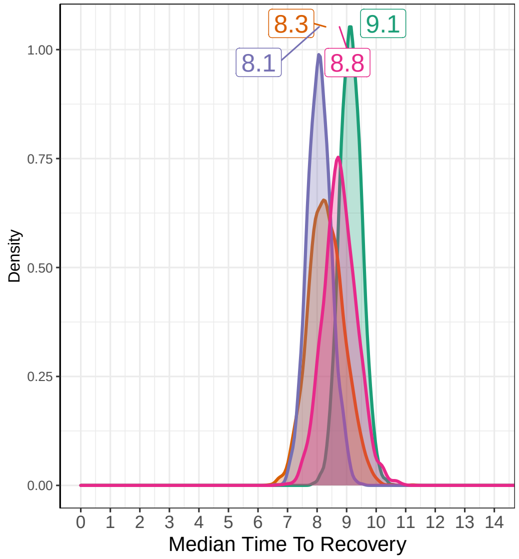

### Interim 4 on January 07, 2021

##### Recovery Data

| | Enrolled | Complete | Recovered | Exposure Days | Recoveries Per Day | Estimated Hazard | Estimated HR (95% interval) | Estimated Median Time to Recovery |
|---|---|---|---|---|---|---|---|---|
| SOC | 770 | 724 | 650 | 8446 | 0.077 | 0.074 | 1 | 9.27 |
| HCQ | 207 | 207 | 188 | 2127 | 0.088 | 0.083 | 1.123 (0.964, 1.31) | 8.27 |
| Azith | 583 | 540 | 497 | 5606 | 0.089 | 0.083 | 1.131 (1.008, 1.266) | 8.23 |
| Doxy | 289 | 289 | 263 | 3158 | 0.083 | 0.079 | 1.069 (0.932, 1.219) | 8.67 |
| Total | 1849 | 1760 | 1598 | 19337 | 0.083 | | | |

##### Hospitalization Data

| Hospitalizations | 28 Day Completers | Observed Hosp. Rate | Est. Hosp. Rate (95% interval) |
|---|---|---|---|
| 25 | 668 | 0.0374 | 0.0337 (0.0221, 0.047) |
| 7 | 207 | 0.0338 | 0.0384 (0.0173, 0.0679) |
| 17 | 472 | 0.0360 | 0.0306 (0.0184, 0.046) |
| 9 | 289 | 0.0311 | 0.0344 (0.0162, 0.0593) |
| 58 | 1636 | 0.0355 | |

| Intervention Status |
|---|
| Enrolling |
| Dropped |
| Enrolling |
| Dropped |

##### Observed Patient Recoveries

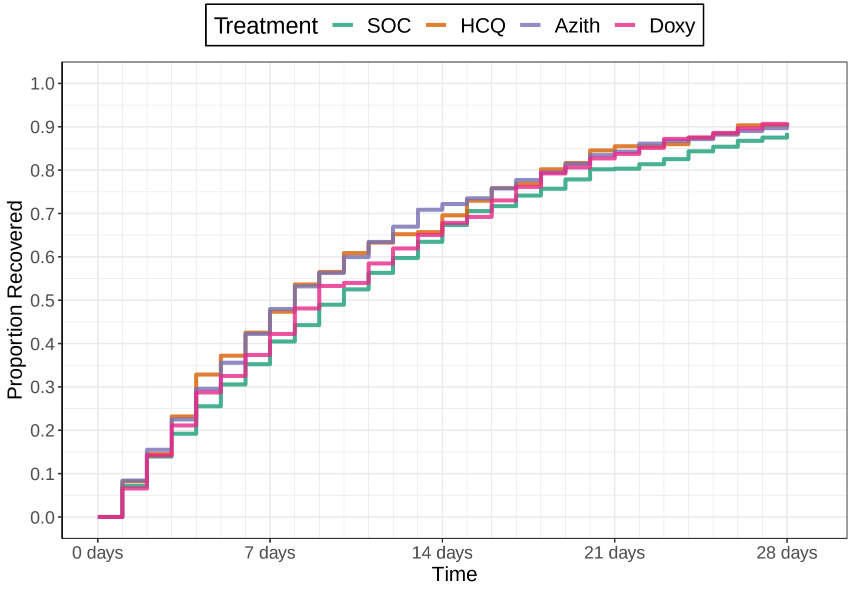

Number Un−recovered

| | 0 days | 7 days | 14 days | 21 days | 28 days |
|---|---|---|---|---|---|
| SOC | 770 | 482 | 264 | 135 | 81 |
| HCQ | 207 | 119 | 71 | 32 | 20 |
| Azith | 583 | 322 | 155 | 82 | 48 |
| Doxy | 289 | 181 | 101 | 50 | 27 |

##### Recovery Inferences

| | Pr(Superiority) | Pr(Meaningful Effect) | Pr(Best) | Randomization Probability |
|---|---|---|---|---|
| SOC | | | | 0.5 |
| HCQ | 0.9317 | 0.218 | 0 | 0 |
| Azith | 0.982 | 0.1753 | 0.75 | 0.5 |
| Doxy | 0.8383 | 0.0533 | 0 | 0 |

##### Hospitalization Inferences

| Pr(Superiority) | Pr(Meaningful Effect) |
|---|---|
| 0.4008 | 0.032 |
| 0.6305 | 0.0352 |
| 0.51 | 0.0415 |

##### Hazard Ratio Estimates

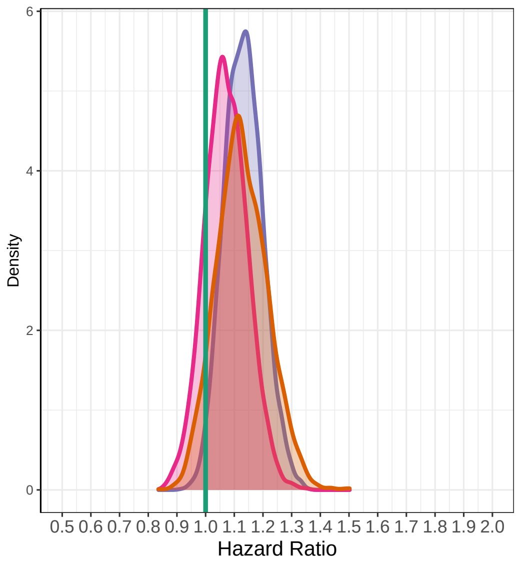

##### Median Time To Recovery Estimates

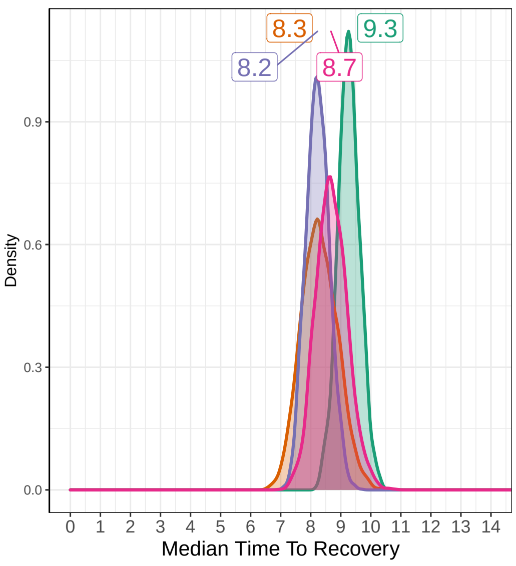

### Interim 5 on February 04, 2021

#### Recovery Data

| | Enrolled | Complete | Recovered | Exposure Days | Recoveries Per Day | Estimated Hazard | Estimated HR (95% interval) | Estimated Median Time to Recovery |
|---|---|---|---|---|---|---|---|---|
| SOC | 881 | 832 | 747 | 9689 | 0.077 | 0.074 | 1 | 9.31 |
| HCQ | 207 | 207 | 188 | 2127 | 0.088 | 0.084 | 1.124 (0.958, 1.325) | 8.31 |
| Azith | 682 | 643 | 590 | 6725 | 0.088 | 0.083 | 1.122 (1.005, 1.246) | 8.34 |
| Doxy | 289 | 289 | 263 | 3158 | 0.083 | 0.079 | 1.069 (0.932, 1.219) | 8.73 |
| Total | 2059 | 1971 | 1788 | 21700 | 0.082 | | | |

#### Hospitalization Data

| Hospitalizations | 28 Day Completers | Observed Hosp. Rate | Est. Hosp. Rate (95% interval) |
|---|---|---|---|
| 28 | 770 | 0.0364 | 0.033 (0.0223, 0.0459) |
| 7 | 207 | 0.0338 | 0.0383 (0.0167, 0.0678) |
| 20 | 583 | 0.0343 | 0.0306 (0.019, 0.0447) |
| 9 | 289 | 0.0311 | 0.0346 (0.017, 0.0578) |
| 64 | 1849 | 0.0346 | |

| Intervention Status |
|---|
| Enrolling |
| Dropped |
| Enrolling |
| Dropped |

#### Observed Patient Recoveries

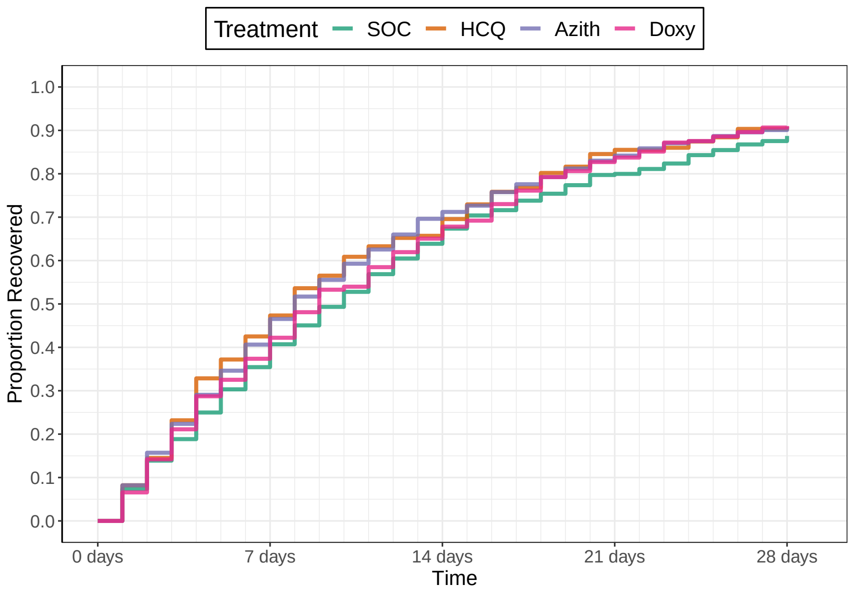

Number Un−recovered

| | 0 days | 7 days | 14 days | 21 days | 28 days |
|---|---|---|---|---|---|
| SOC | 881 | 552 | 299 | 163 | 95 |
| HCQ | 207 | 119 | 71 | 32 | 20 |
| Azith | 682 | 392 | 192 | 102 | 57 |
| Doxy | 289 | 181 | 101 | 50 | 27 |

#### Recovery Inferences

| | Pr(Superiority) | Pr(Meaningful Effect) | Pr(Best) | Randomization Probability |
|---|---|---|---|---|
| SOC | | | | 0.5 |
| HCQ | 0.928 | 0.2117 | 0 | 0 |
| Azith | 0.9803 | 0.125 | 0.75 | 0.5 |
| Doxy | 0.826 | 0.054 | 0 | 0 |

#### Hospitalization Inferences

| Pr(Superiority) | Pr(Meaningful Effect) |
|---|---|
| 0.38 | 0.023 |
| 0.607 | 0.0235 |
| 0.4732 | 0.025 |

#### Hazard Ratio Estimates

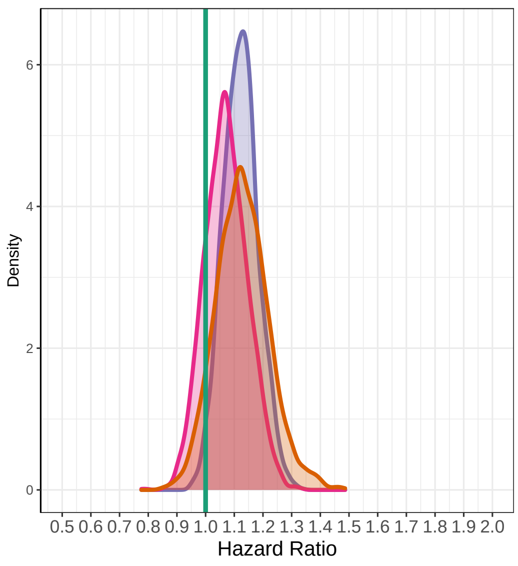

#### Median Time To Recovery Estimates

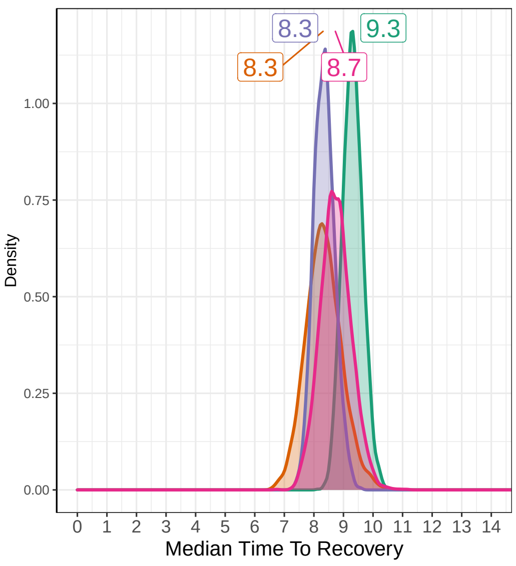

### Interim 6 on March 04, 2021

#### Recovery Data

| | Enrolled | Complete | Recovered | Exposure Days | Recoveries Per Day | Estimated Hazard | Estimated HR (95% interval) | Estimated Median Time to Recovery |
|---|---|---|---|---|---|---|---|---|
| SOC | 1010 | 955 | 861 | 11070 | 0.078 | 0.075 | 1 | 9.3 |
| HCQ | 207 | 207 | 188 | 2127 | 0.088 | 0.084 | 1.113 (0.957, 1.299) | 8.38 |
| Azith | 782 | 737 | 677 | 7695 | 0.088 | 0.084 | 1.115 (1.019, 1.232) | 8.38 |
| Doxy | 289 | 289 | 263 | 3158 | 0.083 | 0.08 | 1.062 (0.926, 1.22) | 8.78 |
| Total | 2288 | 2188 | 1989 | 24050 | 0.083 | | | |

#### Hospitalization Data

| Hospitalizations | 28 Day Completers | Observed Hosp. Rate | Est. Hosp. Rate (95% interval) |
|---|---|---|---|
| 31 | 881 | 0.0352 | 0.0314 (0.0215, 0.0431) |
| 7 | 207 | 0.0338 | 0.0384 (0.0174, 0.0679) |
| 27 | 682 | 0.0396 | 0.0356 (0.0239, 0.0497) |
| 9 | 289 | 0.0311 | 0.0341 (0.0164, 0.0578) |
| 74 | 2059 | 0.0359 | |

| Intervention Status |
|---|
| Enrolling |
| Dropped |
| Enrolling |
| Dropped |

#### Observed Patient Recoveries

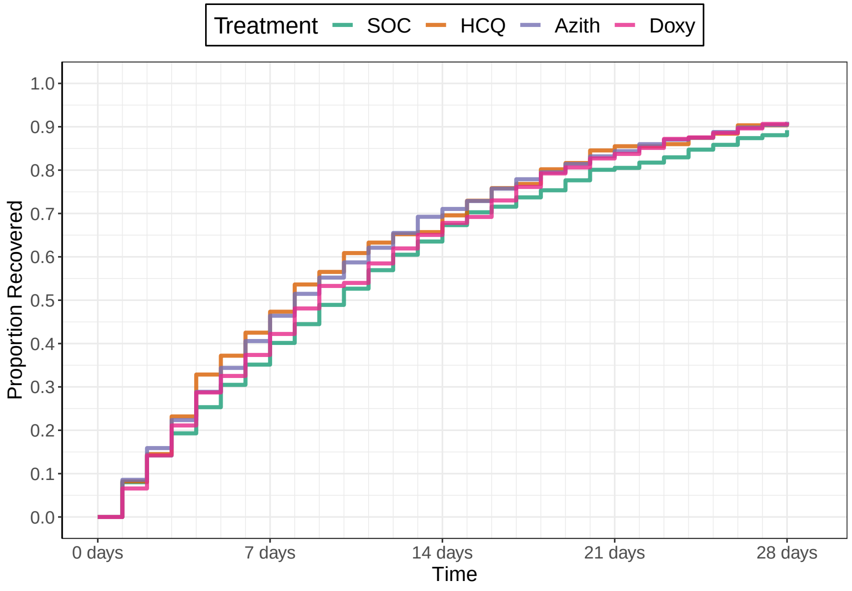

Number Un−recovered

| | 0 days | 7 days | 14 days | 21 days | 28 days |
|---|---|---|---|---|---|
| SOC | 1010 | 635 | 347 | 181 | 104 |
| HCQ | 207 | 119 | 71 | 32 | 20 |
| Azith | 782 | 448 | 221 | 114 | 65 |
| Doxy | 289 | 181 | 101 | 50 | 27 |

#### Recovery Inferences

| | Pr(Superiority) | Pr(Meaningful Effect) | Pr(Best) | Randomization Probability |
|---|---|---|---|---|
| SOC | | | | 0.5 |
| HCQ | 0.9197 | 0.1733 | 0 | 0 |
| Azith | 0.9887 | 0.077 | 0.75 | 0.5 |
| Doxy | 0.8143 | 0.0377 | 0 | 0 |

#### Hospitalization Inferences

| Pr(Superiority) | Pr(Meaningful Effect) |
|---|---|
| 0.331 | 0.0162 |
| 0.3137 | 0.0028 |
| 0.4255 | 0.0168 |

#### Hazard Ratio Estimates

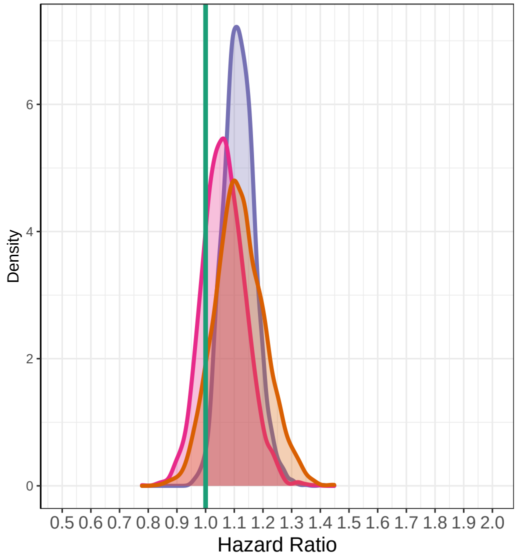

#### Median Time To Recovery Estimates

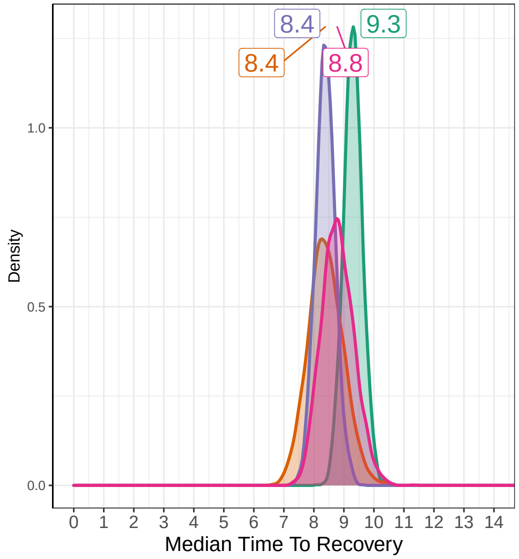

### Interim 7 on April 01, 2021

##### Recovery Data

| | Enrolled | Complete | Recovered | Exposure Days | Recoveries Per Day | Estimated Hazard | Estimated HR (95% interval) | Estimated Median Time to Recovery |
|---|---|---|---|---|---|---|---|---|
| SOC | 1130 | 1085 | 975 | 12506 | 0.078 | 0.076 | 1 | 9.29 |
| HCQ | 207 | 207 | 188 | 2127 | 0.088 | 0.085 | 1.114 (0.96, 1.287) | 8.38 |
| Azith | 884 | 848 | 784 | 8906 | 0.088 | 0.085 | 1.115 (1.015, 1.217) | 8.37 |
| Doxy | 289 | 289 | 263 | 3158 | 0.083 | 0.081 | 1.059 (0.924, 1.206) | 8.8 |
| Total | 2510 | 2429 | 2210 | 26697 | 0.083 | | | |

##### Hospitalization Data

| Hospitalizations | 28 Day Completers | Observed Hosp. Rate | Est. Hosp. Rate (95% interval) |
|---|---|---|---|
| 33 | 1010 | 0.0327 | 0.03 (0.0209, 0.0405) |
| 7 | 207 | 0.0338 | 0.0382 (0.0164, 0.0674) |
| 31 | 782 | 0.0396 | 0.0361 (0.0249, 0.0494) |
| 9 | 289 | 0.0311 | 0.0342 (0.0163, 0.0577) |
| 80 | 2288 | 0.0350 | |

| Intervention Status |
|---|
| Enrolling |
| Dropped |
| Enrolling |
| Dropped |

##### Observed Patient Recoveries

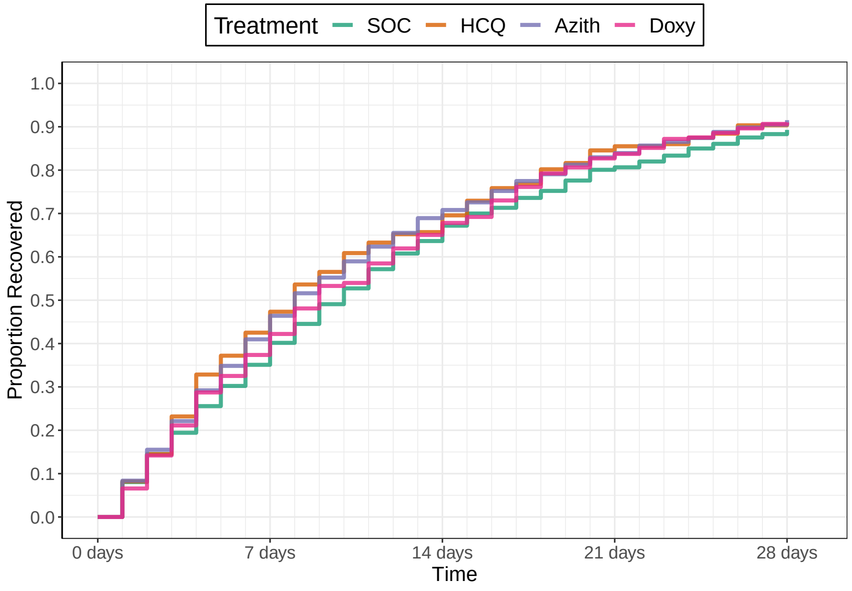

Number Un−recovered

| | 0 days | 7 days | 14 days | 21 days | 28 days |
|---|---|---|---|---|---|
| SOC | 1130 | 717 | 390 | 209 | 120 |
| HCQ | 207 | 119 | 71 | 32 | 20 |
| Azith | 884 | 511 | 265 | 139 | 73 |
| Doxy | 289 | 181 | 101 | 50 | 27 |

##### Recovery Inferences

| | Pr(Superiority) | Pr(Meaningful Effect) | Pr(Best) | Randomization Probability |
|---|---|---|---|---|
| SOC | | | | 0.5 |
| HCQ | 0.927 | 0.1657 | 0 | 0 |
| Azith | 0.9893 | 0.0693 | 0.75 | 0.5 |
| Doxy | 0.803 | 0.036 | 0 | 0 |

##### Hospitalization Inferences

| Pr(Superiority) | Pr(Meaningful Effect) |
|---|---|
| 0.3045 | 0.0082 |
| 0.2268 | 8e−04 |
| 0.3902 | 0.0092 |

##### Hazard Ratio Estimates

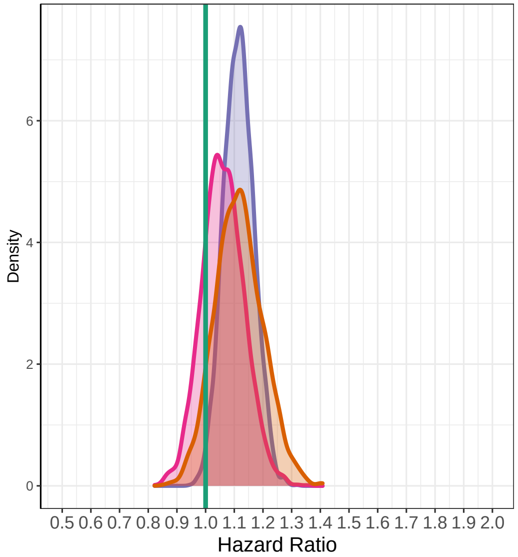

##### Median Time To Recovery Estimates

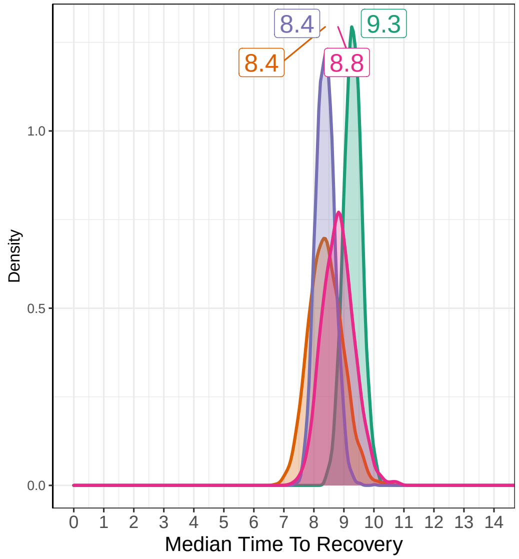

### Interim 8 on April 29, 2021

#### Recovery Data

| | Enrolled | Complete | Recovered | Exposure Days | Recoveries Per Day | Estimated Hazard | Estimated HR (95% interval) | Estimated Median Time to Recovery |
|---|---|---|---|---|---|---|---|---|
| SOC | 1228 | 1194 | 1073 | 13572 | 0.079 | 0.076 | 1 | 9.12 |
| HCQ | 207 | 207 | 188 | 2127 | 0.088 | 0.084 | 1.1 (0.944, 1.283) | 8.33 |
| Azith | 996 | 954 | 876 | 9989 | 0.088 | 0.084 | 1.097 (1.008, 1.198) | 8.35 |
| Doxy | 289 | 289 | 263 | 3158 | 0.083 | 0.08 | 1.049 (0.921, 1.187) | 8.72 |
| Total | 2720 | 2644 | 2400 | 28845 | 0.083 | | | |

#### Hospitalization Data

| Hospitalizations | 28 Day Completers | Observed Hosp. Rate | Est. Hosp. Rate (95% interval) |
|---|---|---|---|
| 34 | 1130 | 0.0301 | 0.0285 (0.0199, 0.0385) |
| 7 | 207 | 0.0338 | 0.0382 (0.0166, 0.0675) |
| 35 | 884 | 0.0396 | 0.0361 (0.0254, 0.0481) |
| 9 | 289 | 0.0311 | 0.0344 (0.0167, 0.0579) |
| 85 | 2510 | 0.0339 | |

| Intervention Status |
|---|
| Enrolling |
| Dropped |
| Futility |
| Dropped |

#### Observed Patient Recoveries

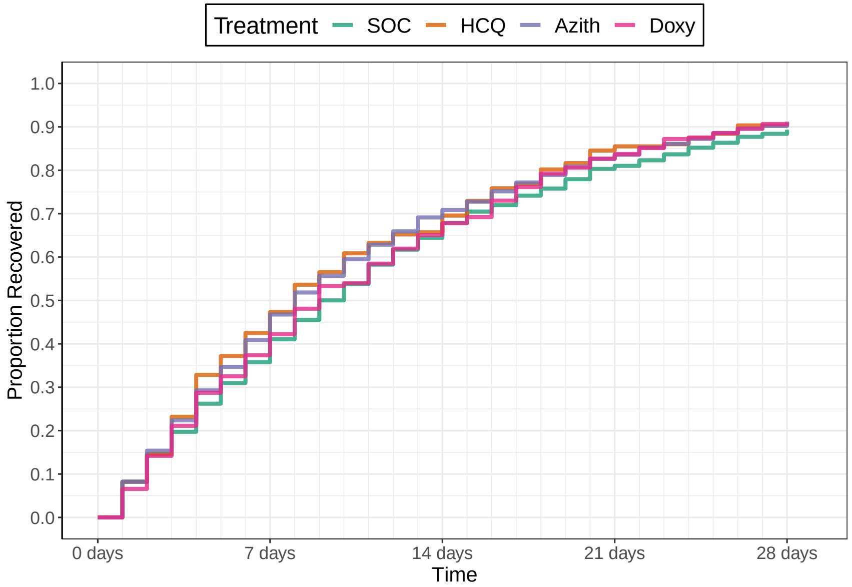

Number Un−recovered

| | 0 days | 7 days | 14 days | 21 days | 28 days |
|---|---|---|---|---|---|
| SOC | 1228 | 777 | 421 | 230 | 133 |
| HCQ | 207 | 119 | 71 | 32 | 20 |
| Azith | 996 | 573 | 288 | 159 | 87 |
| Doxy | 289 | 181 | 101 | 50 | 27 |

#### Recovery Inferences

| | Pr(Superiority) | Pr(Meaningful Effect) | Pr(Best) | Randomization Probability |
|---|---|---|---|---|
| SOC | | | | 1 |
| HCQ | 0.8877 | 0.1253 | 0 | 0 |
| Azith | 0.9853 | 0.027 | 0 | 0 |
| Doxy | 0.759 | 0.019 | 0 | 0 |

#### Hospitalization Inferences

| Pr(Superiority) | Pr(Meaningful Effect) |
|---|---|
| 0.254 | 0.0068 |
| 0.1568 | 0 |
| 0.3295 | 0.006 |

#### Hazard Ratio Estimates

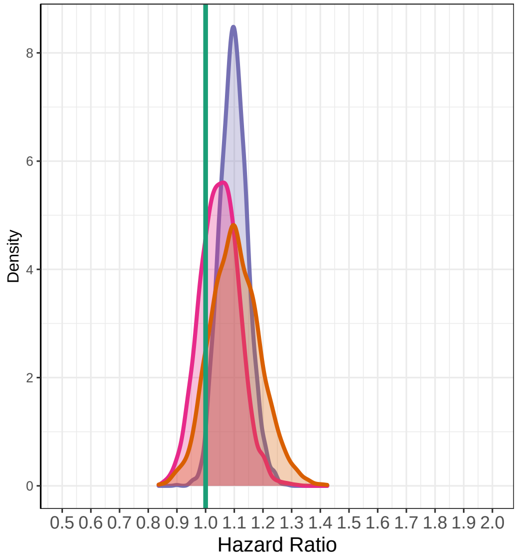

#### Median Time To Recovery Estimates

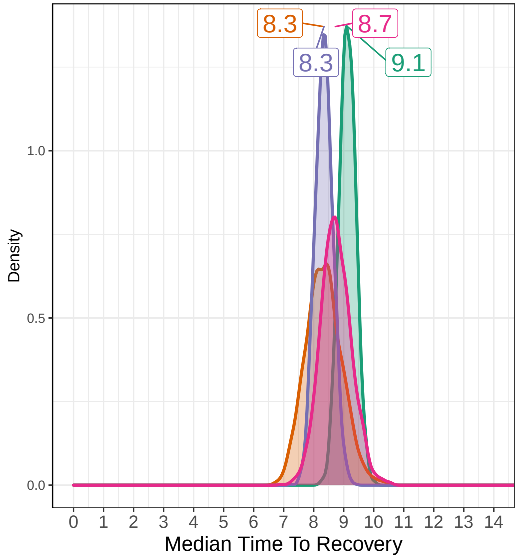

### Interim 9 on August 26, 2022

#### Recovery Data

| | Enrolled | Complete | Recovered | Exposure Days | Recoveries Per Day | Estimated Hazard | Estimated HR (95% interval) | Estimated Median Time to Recovery |
|---|---|---|---|---|---|---|---|---|
| SOC | 4508 | 4508 | 3979 | 52829 | 0.075 | 0.073 | 1 | 9.64 |
| HCQ | 207 | 207 | 188 | 2127 | 0.088 | 0.085 | 1.154 (0.995, 1.322) | 8.39 |
| Azith | 996 | 996 | 908 | 10382 | 0.087 | 0.084 | 1.153 (1.071, 1.235) | 8.4 |
| Doxy | 289 | 289 | 263 | 3158 | 0.083 | 0.081 | 1.098 (0.97, 1.243) | 8.79 |
| Total | 6000 | 6000 | 5338 | 68497 | 0.078 | | | |

#### Hospitalization Data

| Hospitalizations | 28 Day Completers | Observed Hosp. Rate | Est. Hosp. Rate (95% interval) |
|---|---|---|---|
| 138 | 4508 | 0.0306 | 0.0308 (0.026, 0.0361) |
| 7 | 207 | 0.0338 | 0.0387 (0.017, 0.0686) |
| 35 | 996 | 0.0351 | 0.0361 (0.0253, 0.0486) |
| 9 | 289 | 0.0311 | 0.0344 (0.0167, 0.0584) |
| 189 | 6000 | 0.0315 | |

| Intervention Status |
|---|
| Complete |
| Dropped |
| Dropped |
| Dropped |

#### Observed Patient Recoveries

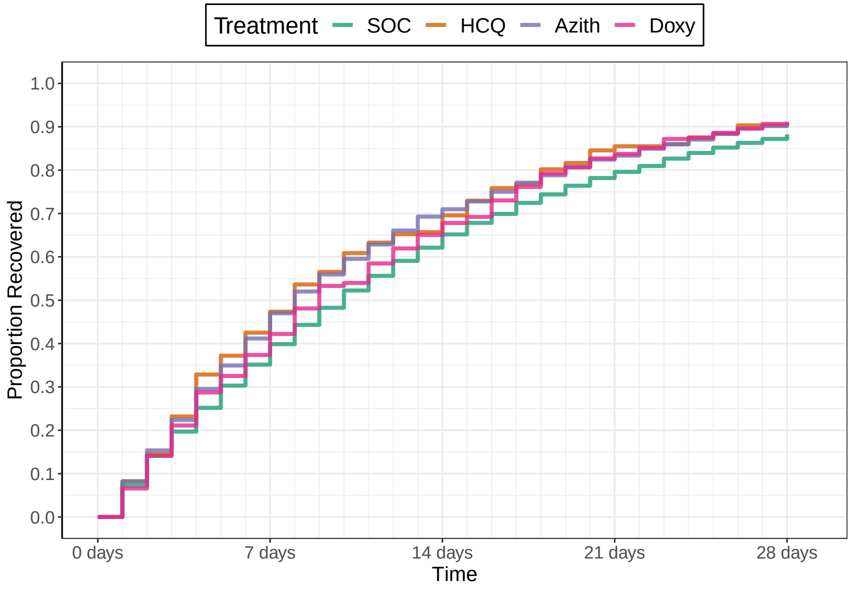

Number Un−recovered

| | 0 days | 7 days | 14 days | 21 days | 28 days |
|---|---|---|---|---|---|
| SOC | 4508 | 2924 | 1708 | 984 | 577 |
| HCQ | 207 | 119 | 71 | 32 | 20 |
| Azith | 996 | 586 | 306 | 175 | 98 |
| Doxy | 289 | 181 | 101 | 50 | 27 |

#### Recovery Inferences

| | Pr(Superiority) | Pr(Meaningful Effect) | Pr(Best) | Randomization Probability |
|---|---|---|---|---|
| SOC | | | | |
| HCQ | 0.9697 | 0.3407 | 0 | |
| Azith | 1 | 0.1867 | 0 | |
| Doxy | 0.928 | 0.1067 | 0 | |

#### Hospitalization Inferences

| Pr(Superiority) | Pr(Meaningful Effect) |
|---|---|
| 0.3088 | 0.005 |
| 0.2218 | 0 |
| 0.4058 | 0.0032 |

#### Hazard Ratio Estimates

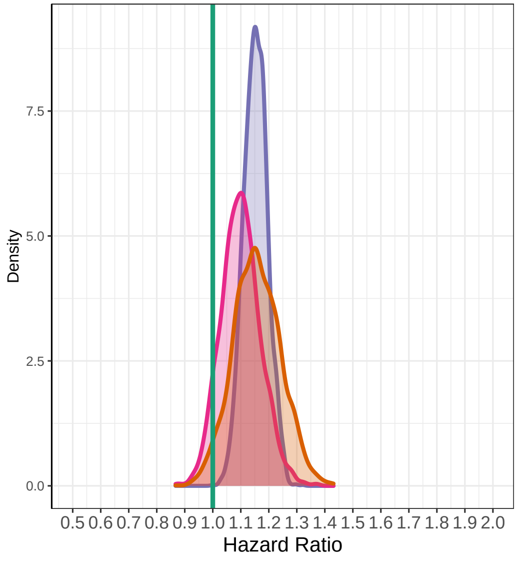

#### Median Time To Recovery Estimates

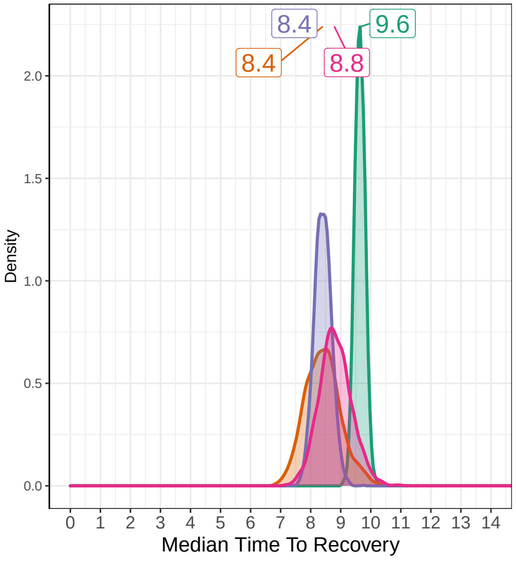

#### 6.5 Example 4

### April 1, 2020

Before Patient Enrollment Begins

#### Recovery Data

| | Enrolled | Complete | Recovered | Exposure Days | Recoveries Per Day | Estimated Hazard | Estimated HR (95% interval) | Estimated Median Survival Time |
|---|---|---|---|---|---|---|---|---|
| SOC | 0 | 0 | 0 | 0 | | | | |
| HCQ | 0 | 0 | 0 | 0 | | | | |
| Azith | 0 | 0 | 0 | 0 | | | | |
| Doxy | 0 | 0 | 0 | 0 | | | | |
| Total | 0 | 0 | 0 | 0 | | | | |

#### Hospitalization Data

| Hospitalizations | 28 Day Completers | Observed Hosp. Rate | Est. Hosp. Rate (95% interval) |
|---|---|---|---|
| 0 | 0 | | |
| 0 | 0 | | |
| 0 | 0 | | |
| 0 | 0 | | |
| 0 | 0 | | |

| Intervention Status |
|---|
| Enrolling |
| Enrolling |
| Unopened |
| Unopened |

#### Observed Patient Recoveries

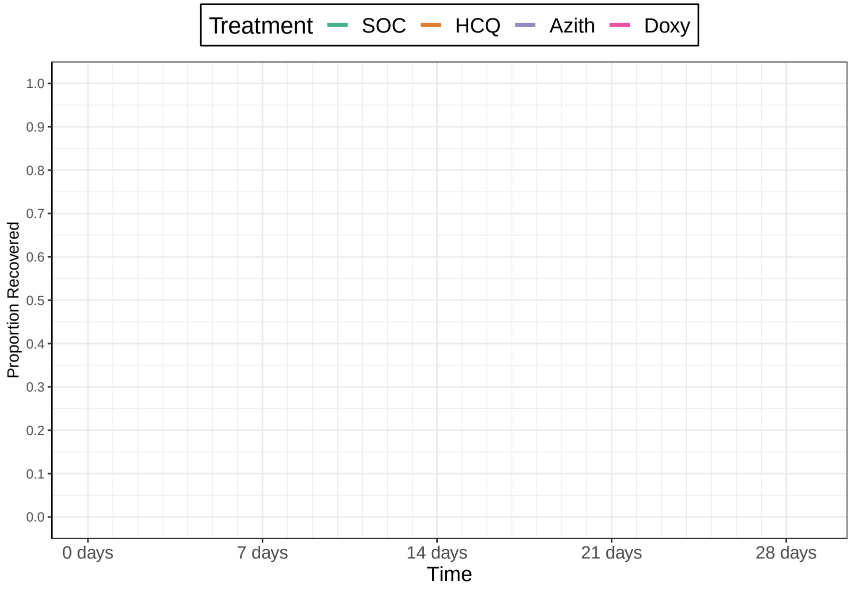

Number Un−recovered

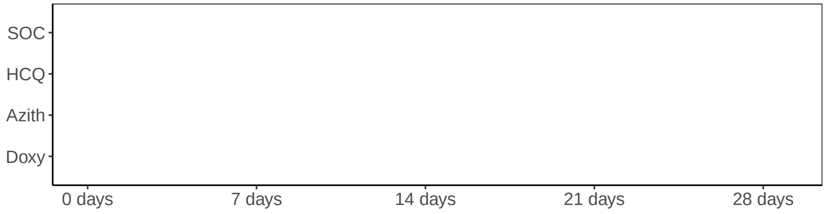

#### Recovery Inferences

| | Pr(Superiority) | Pr(Meaningful Effect) | Pr(Best) | Randomization Probability |
|---|---|---|---|---|
| SOC | | | | 0.5 |
| HCQ | | | | 0.5 |
| Azith | | | | 0 |
| Doxy | | | | 0 |

#### Hospitalization Inferences

| Pr(Superiority) | Pr(Meaningful Effect) |
|---|---|
| 0 | 0 |
| 0 | 0 |
| 0 | 0 |
| 0 | 0 |

#### Hazard Ratio Estimates

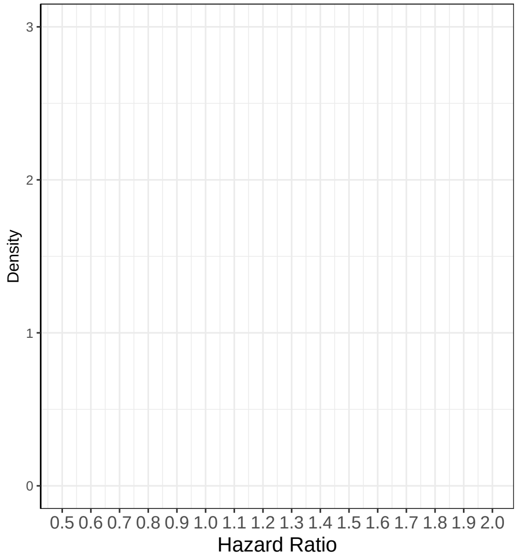

#### Median Time To Recovery Estimates

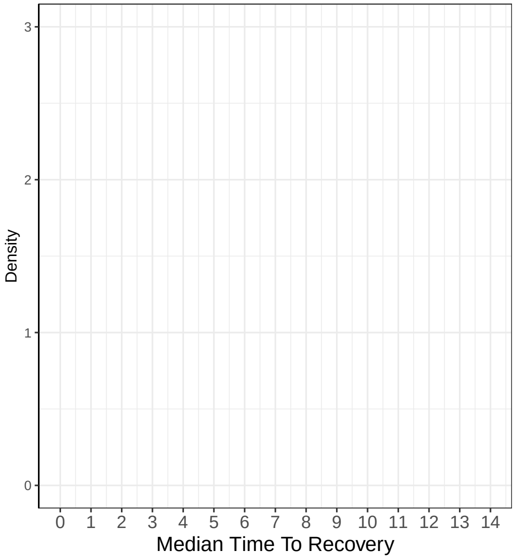

### Remove Drug on May 22, 2020

#### Recovery Data

| | Enrolled | Complete | Recovered | Exposure Days | Recoveries Per Day | Estimated Hazard | Estimated HR (95% interval) | Estimated Median Time to Recovery |
|---|---|---|---|---|---|---|---|---|
| SOC | 206 | 152 | 146 | 1728 | 0.084 | | | |
| HCQ | 207 | 143 | 134 | 1821 | 0.074 | | | |
| Azith | 0 | 0 | 0 | 0 | | | | |
| Doxy | 0 | 0 | 0 | 0 | | | | |
| Total | 413 | 295 | 280 | 3549 | 0.079 | | | |

#### Hospitalization Data

| Hospitalizations | 28 Day Completers | Observed Hosp. Rate | Est. Hosp. Rate (95% interval) |
|---|---|---|---|
| 4 | 79 | 0.0506 | |
| 5 | 78 | 0.0641 | |
| 0 | 0 | NA | |
| 0 | 0 | NA | |
| 9 | 157 | 0.0573 | |

| Intervention Status |
|---|
| Enrolling |
| Removing |
| Unopened |
| Unopened |

#### Observed Patient Recoveries

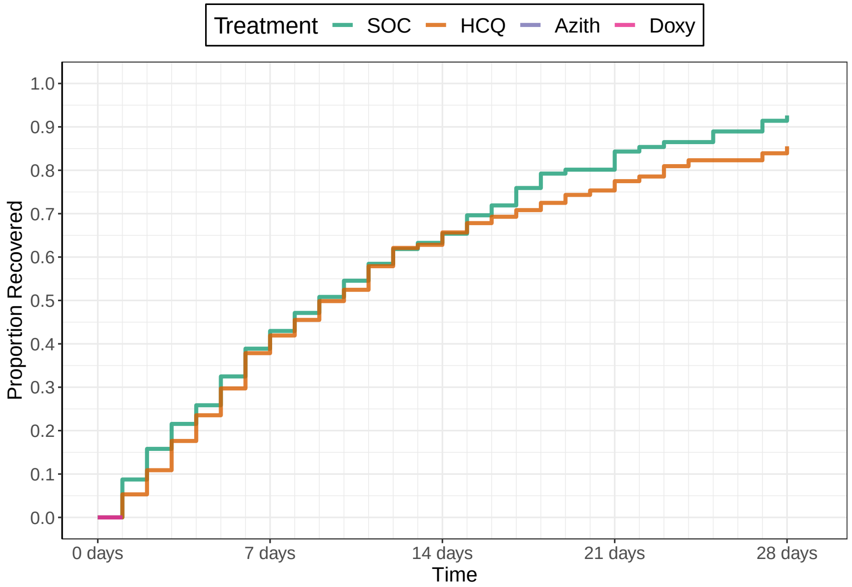

Number Un−recovered

| | 0 days | 7 days | 14 days | 21 days | 28 days |
|---|---|---|---|---|---|
| SOC | 206 | 105 | 52 | 19 | 7 |
| HCQ | 207 | 107 | 52 | 23 | 10 |
| Azith | | | | | |
| Doxy | | | | | |

#### Recovery Inferences

| | Pr(Superiority) | Pr(Meaningful Effect) | Pr(Best) | Randomization Probability |
|---|---|---|---|---|
| SOC | | | | 1 |
| HCQ | | | 0 | 0 |
| Azith | | | 0 | 0 |
| Doxy | | | 0 | 0 |

#### Hospitalization Inferences

| Pr(Superiority) | Pr(Meaningful Effect) |
|---|---|

#### Hazard Ratio Estimates

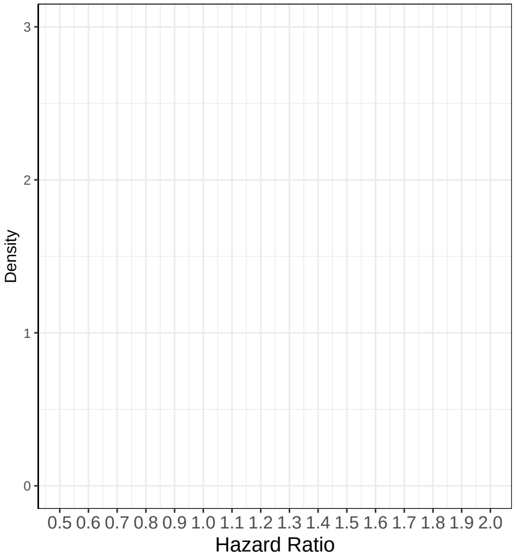

#### Median Time To Recovery Estimates

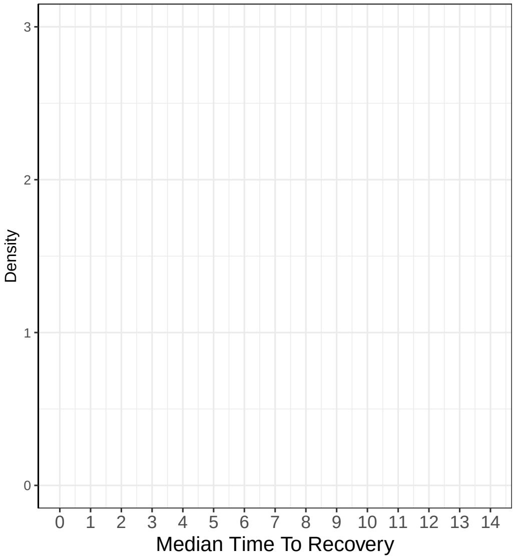

### Add Drug on May 23, 2020

##### Recovery Data

| | Enrolled | Complete | Recovered | Exposure Days | Recoveries Per Day | Estimated Hazard | Estimated HR (95% interval) | Estimated Median Time to Recovery |
|---|---|---|---|---|---|---|---|---|
| SOC | 208 | 155 | 149 | 1782 | 0.084 | | | |
| HCQ | 207 | 147 | 138 | 1884 | 0.073 | | | |
| Azith | 0 | 0 | 0 | 0 | | | | |
| Doxy | 0 | 0 | 0 | 0 | | | | |
| Total | 415 | 302 | 287 | 3666 | 0.078 | | | |

##### Hospitalization Data

| Hospitalizations | 28 Day Completers | Observed Hosp. Rate | Est. Hosp. Rate (95% interval) |
|---|---|---|---|
| 4 | 80 | 0.0500 | |
| 5 | 80 | 0.0625 | |
| 0 | 0 | NA | |
| 0 | 0 | NA | |
| 9 | 160 | 0.0562 | |

| Intervention Status |
|---|
| Enrolling |
| Paused |
| Introducing |
| Unopened |

##### Observed Patient Recoveries

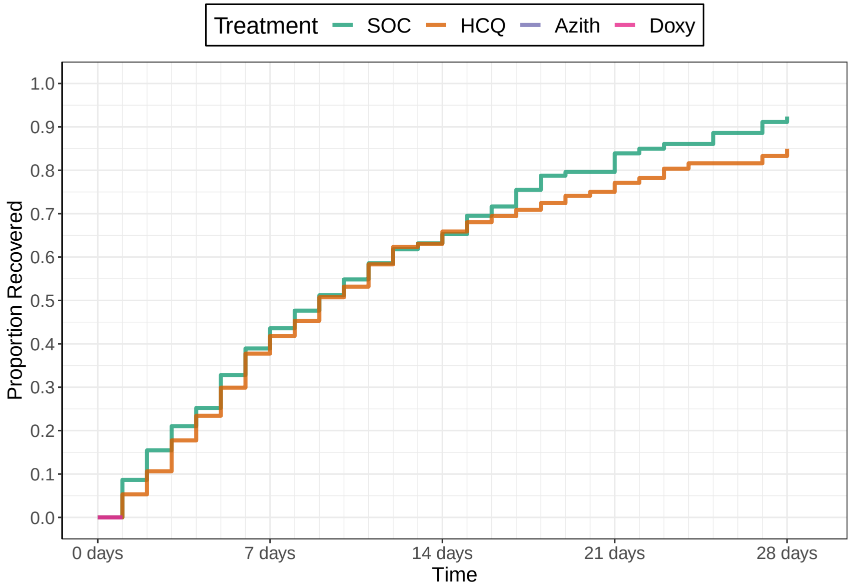

Number Un−recovered

| | 0 days | 7 days | 14 days | 21 days | 28 days |
|---|---|---|---|---|---|
| SOC | 208 | 105 | 52 | 19 | 7 |
| HCQ | 207 | 107 | 52 | 24 | 10 |
| Azith | | | | | |
| Doxy | | | | | |

##### Recovery Inferences

| | Pr(Superiority) | Pr(Meaningful Effect) | Pr(Best) | Randomization Probability |
|---|---|---|---|---|
| SOC | | | | 0.5 |
| HCQ | | | 0 | 0 |
| Azith | | | 1 | 0.5 |
| Doxy | | | 0 | 0 |

##### Hospitalization Inferences

| Pr(Superiority) | Pr(Meaningful Effect) |
|---|---|

##### Hazard Ratio Estimates

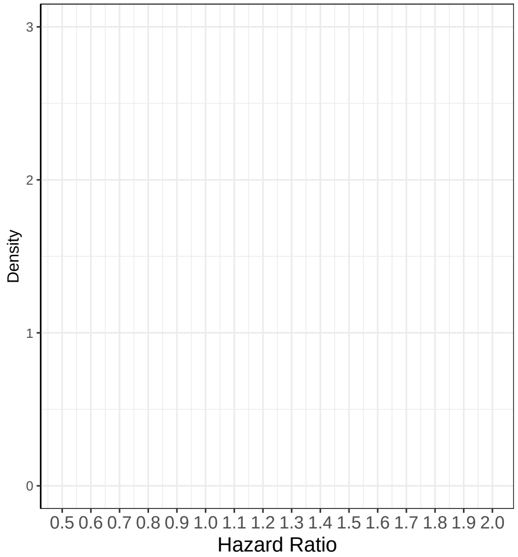

##### Median Time To Recovery Estimates

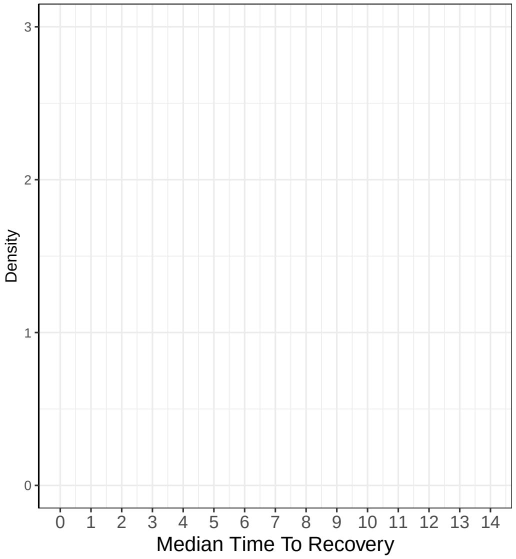

### Add Drug on July 23, 2020

#### Recovery Data

| | Enrolled | Complete | Recovered | Exposure Days | Recoveries Per Day | Estimated Hazard | Estimated HR (95% interval) | Estimated Median Time to Recovery |
|---|---|---|---|---|---|---|---|---|
| SOC | 326 | 310 | 286 | 3499 | 0.082 | | | |
| HCQ | 207 | 207 | 179 | 2453 | 0.073 | | | |
| Azith | 118 | 104 | 100 | 1078 | 0.093 | | | |
| Doxy | 0 | 0 | 0 | 0 | | | | |
| Total | 651 | 621 | 565 | 7030 | 0.08 | | | |

#### Hospitalization Data

| Hospitalizations | 28 Day Completers | Observed Hosp. Rate | Est. Hosp. Rate (95% interval) |
|---|---|---|---|
| 7 | 285 | 0.0246 | |
| 5 | 207 | 0.0242 | |
| 4 | 77 | 0.0519 | |
| 0 | 0 | NA | |
| 16 | 569 | 0.0281 | |

| Intervention Status |
|---|
| Enrolling |
| Paused |
| Introducing |
| Introducing |

#### Observed Patient Recoveries

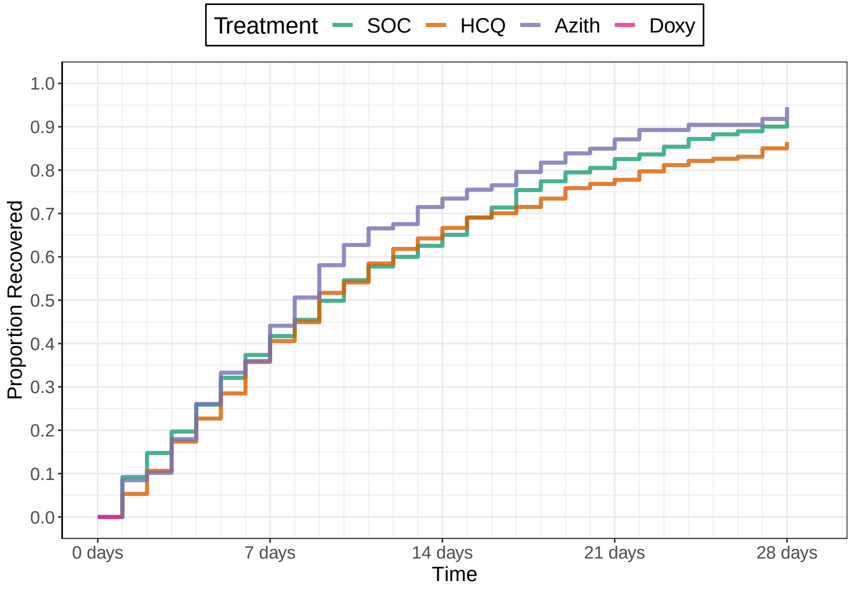

Number Un−recovered

| | 0 days | 7 days | 14 days | 21 days | 28 days |
|---|---|---|---|---|---|
| SOC | 326 | 202 | 118 | 57 | 28 |
| HCQ | 207 | 133 | 74 | 48 | 31 |
| Azith | 118 | 71 | 29 | 14 | 6 |
| Doxy | | | | | |

#### Recovery Inferences

| | Pr(Superiority) | Pr(Meaningful Effect) | Pr(Best) | Randomization Probability |
|---|---|---|---|---|
| SOC | | | | 0.33 |
| HCQ | | | 0 | 0 |
| Azith | | | 1 | 0.33 |
| Doxy | | | 0 | 0.33 |

#### Hospitalization Inferences

| Pr(Superiority) | Pr(Meaningful Effect) |
|---|---|

#### Hazard Ratio Estimates

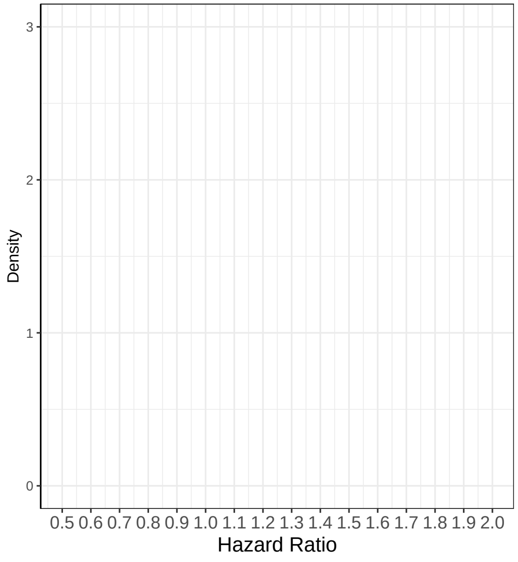

#### Median Time To Recovery Estimates

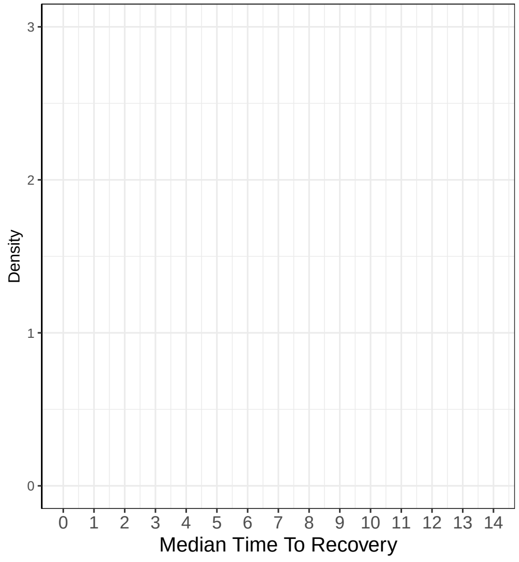

### Interim 1 on October 15, 2020

#### Recovery Data

| | Enrolled | Complete | Recovered | Exposure Days | Recoveries Per Day | Estimated Hazard | Estimated HR (95% interval) | Estimated Median Time to Recovery |
|---|---|---|---|---|---|---|---|---|
| SOC | 482 | 440 | 405 | 4826 | 0.084 | 0.082 | 1 | 8.69 |
| HCQ | 207 | 207 | 179 | 2453 | 0.073 | 0.072 | 0.875 (0.742, 1.03) | 9.88 |
| Azith | 312 | 270 | 260 | 2615 | 0.099 | 0.095 | 1.158 (1.004, 1.349) | 7.53 |
| Doxy | 209 | 151 | 146 | 1571 | 0.093 | 0.088 | 1.08 (0.901, 1.302) | 8.05 |
| Total | 1210 | 1068 | 990 | 11465 | 0.086 | | | |

#### Hospitalization Data

| Hospitalizations | 28 Day Completers | Observed Hosp. Rate | Est. Hosp. Rate (95% interval) |
|---|---|---|---|
| 10 | 387 | 0.0258 | 0.0228 (0.0115, 0.0382) |
| 5 | 207 | 0.0242 | 0.0286 (0.0108, 0.0541) |
| 8 | 200 | 0.0400 | 0.0288 (0.0132, 0.05) |
| 2 | 79 | 0.0253 | 0.0141 (0.003, 0.0346) |
| 25 | 873 | 0.0286 | |

| Intervention Status |
|---|
| Enrolling |
| Futility |
| Enrolling |
| Enrolling |

#### Observed Patient Recoveries

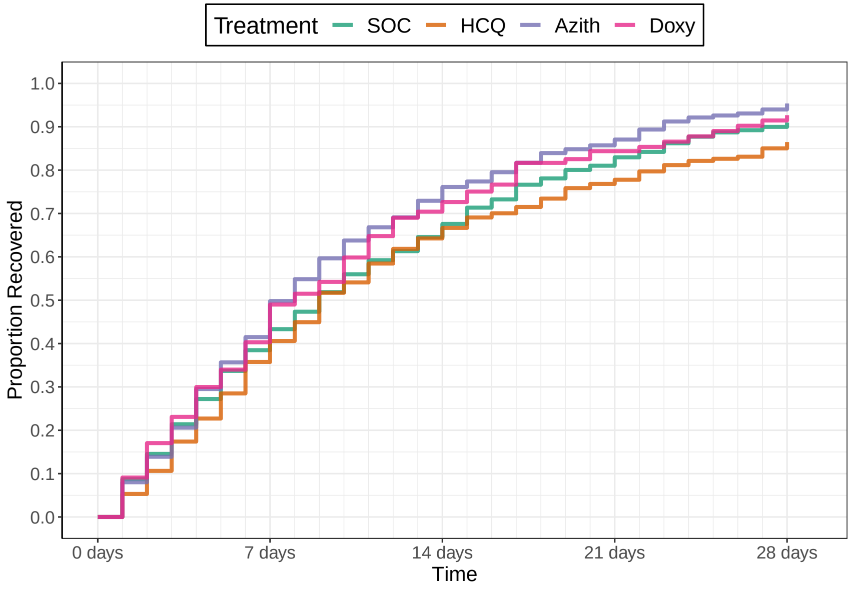

Number Un−recovered

| | 0 days | 7 days | 14 days | 21 days | 28 days |
|---|---|---|---|---|---|
| SOC | 482 | 279 | 152 | 78 | 40 |
| HCQ | 207 | 133 | 74 | 48 | 31 |
| Azith | 312 | 162 | 68 | 32 | 13 |
| Doxy | 209 | 103 | 40 | 16 | 7 |

#### Recovery Inferences

| | Pr(Superiority) | Pr(Meaningful Effect) | Pr(Best) | Randomization Probability |
|---|---|---|---|---|
| SOC | | | | 0.33 |
| HCQ | 0.0513 | 0 | 0 | 0 |
| Azith | 0.9793 | 0.28 | 0.567 | 0.37 |
| Doxy | 0.8027 | 0.116 | 0.183 | 0.3 |

#### Hospitalization Inferences

| Pr(Superiority) | Pr(Meaningful Effect) |
|---|---|
| 0.3352 | 0.0142 |
| 0.3022 | 0.0122 |
| 0.806 | 0.1338 |

#### Hazard Ratio Estimates

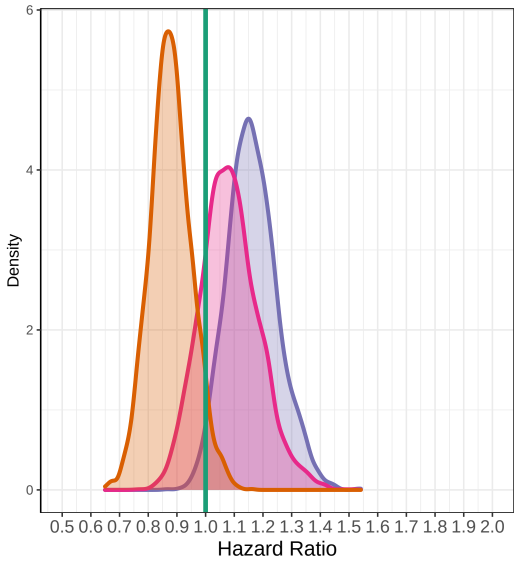

#### Median Time To Recovery Estimates

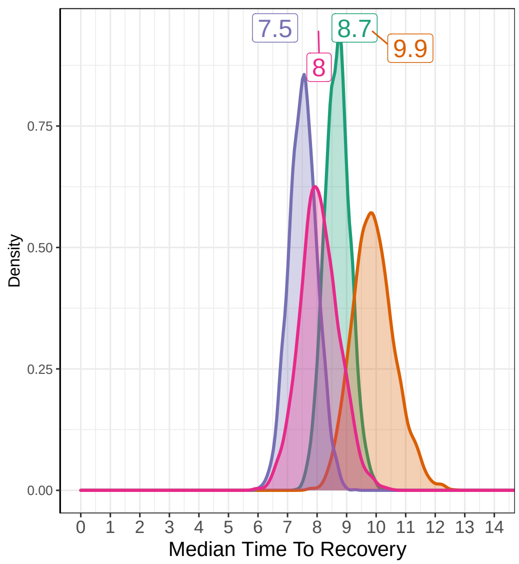

### Interim 2 on November 12, 2020

#### Recovery Data

| | Enrolled | Complete | Recovered | Exposure Days | Recoveries Per Day | Estimated Hazard | Estimated HR (95% interval) | Estimated Median Time to Recovery |
|---|---|---|---|---|---|---|---|---|
| SOC | 573 | 535 | 488 | 5941 | 0.082 | 0.079 | 1 | 8.83 |
| HCQ | 207 | 207 | 179 | 2453 | 0.073 | 0.071 | 0.891 (0.756, 1.049) | 9.85 |
| Azith | 405 | 373 | 354 | 3728 | 0.095 | 0.09 | 1.132 (0.989, 1.296) | 7.85 |
| Doxy | 263 | 246 | 231 | 2421 | 0.095 | 0.09 | 1.134 (0.977, 1.32) | 7.83 |
| Total | 1448 | 1361 | 1252 | 14543 | 0.086 | | | |

#### Hospitalization Data

| Hospitalizations | 28 Day Completers | Observed Hosp. Rate | Est. Hosp. Rate (95% interval) |
|---|---|---|---|
| 15 | 482 | 0.0311 | 0.0279 (0.0162, 0.0424) |
| 5 | 207 | 0.0242 | 0.0285 (0.0109, 0.054) |
| 10 | 312 | 0.0321 | 0.0274 (0.014, 0.0455) |
| 4 | 209 | 0.0191 | 0.019 (0.0062, 0.0387) |
| 34 | 1210 | 0.0281 | |

| Intervention Status |
|---|
| Enrolling |
| Dropped |
| Enrolling |
| Enrolling |

#### Observed Patient Recoveries

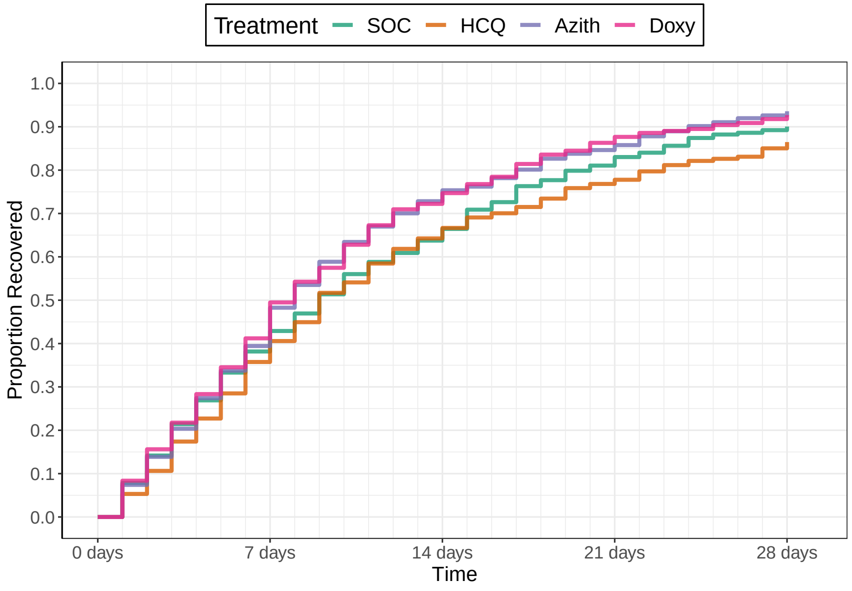

Number Un−recovered

| | 0 days | 7 days | 14 days | 21 days | 28 days |
|---|---|---|---|---|---|
| SOC | 573 | 340 | 188 | 96 | 52 |
| HCQ | 207 | 133 | 74 | 48 | 31 |
| Azith | 405 | 233 | 97 | 54 | 23 |
| Doxy | 263 | 149 | 67 | 30 | 18 |

#### Recovery Inferences

| | Pr(Superiority) | Pr(Meaningful Effect) | Pr(Best) | Randomization Probability |
|---|---|---|---|---|
| SOC | | | | 0.33 |
| HCQ | 0.0867 | 0 | 0 | 0 |
| Azith | 0.966 | 0.1797 | 0.3665 | 0.34 |
| Doxy | 0.944 | 0.1967 | 0.3835 | 0.33 |

#### Hospitalization Inferences

| Pr(Superiority) | Pr(Meaningful Effect) |
|---|---|
| 0.5152 | 0.042 |
| 0.5262 | 0.033 |
| 0.8028 | 0.1468 |

#### Hazard Ratio Estimates

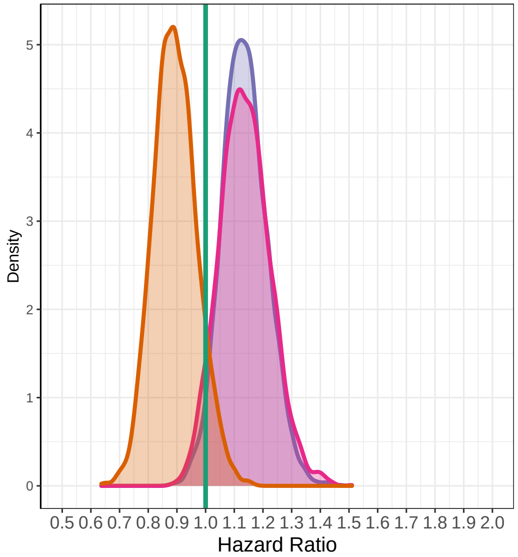

#### Median Time To Recovery Estimates

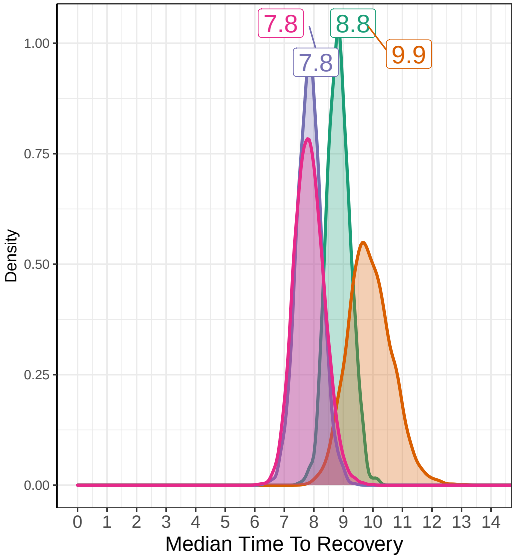

### Interim 3 on December 10, 2020

**Recovery Data**

| | Enrolled | Complete | Recovered | Exposure Days | Recoveries Per Day | Estimated Hazard | Estimated HR (95% interval) | Estimated Median Time to Recovery |
|---|---|---|---|---|---|---|---|---|
| SOC | 638 | 615 | 559 | 6714 | 0.083 | 0.08 | 1 | 8.77 |
| HCQ | 207 | 207 | 179 | 2453 | 0.073 | 0.071 | 0.884 (0.749, 1.036) | 9.85 |
| Azith | 470 | 452 | 422 | 4524 | 0.093 | 0.089 | 1.107 (0.983, 1.254) | 7.99 |
| Doxy | 334 | 307 | 290 | 3140 | 0.092 | 0.088 | 1.093 (0.954, 1.259) | 8.07 |
| Total | 1649 | 1581 | 1450 | 16831 | 0.086 | | | |

**Hospitalization Data**

| Hospitalizations | 28 Day Completers | Observed Hosp. Rate | Est. Hosp. Rate (95% interval) |
|---|---|---|---|
| 18 | 573 | 0.0314 | 0.0295 (0.0181, 0.044) |
| 5 | 207 | 0.0242 | 0.0288 (0.0108, 0.0568) |
| 11 | 405 | 0.0272 | 0.0252 (0.0134, 0.0417) |
| 7 | 263 | 0.0266 | 0.0241 (0.0103, 0.0436) |
| 41 | 1448 | 0.0283 | |

| Intervention Status |
|---|
| Enrolling |
| Dropped |
| Enrolling |
| Enrolling |

#### Observed Patient Recoveries

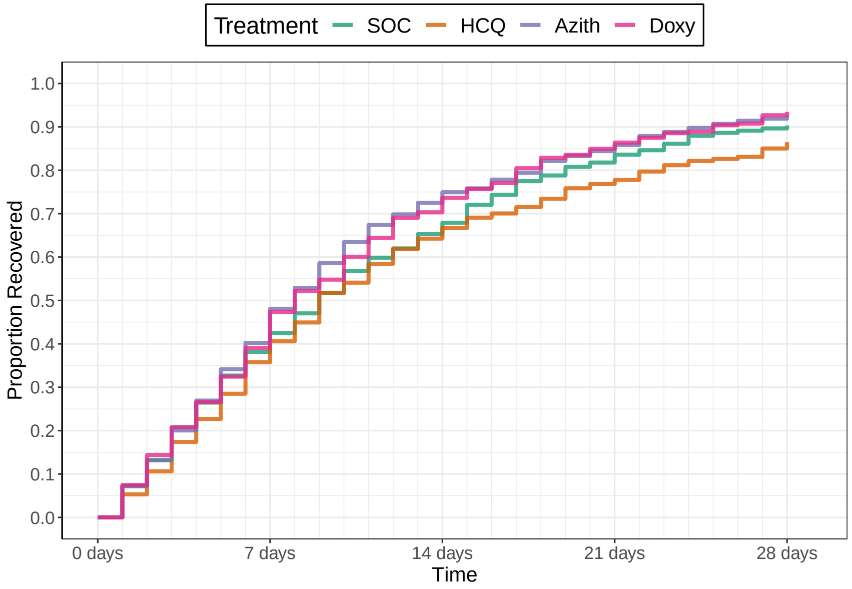

**Number Un−recovered**

| | 0 days | 7 days | 14 days | 21 days | 28 days |
|---|---|---|---|---|---|
| SOC | 638 | 384 | 211 | 110 | 60 |
| HCQ | 207 | 133 | 74 | 48 | 31 |
| Azith | 470 | 274 | 124 | 67 | 33 |
| Doxy | 334 | 191 | 89 | 42 | 19 |

**Recovery Inferences**

| | Pr(Superiority) | Pr(Meaningful Effect) | Pr(Best) | Randomization Probability |
|---|---|---|---|---|
| SOC | | | | 0.33 |
| HCQ | 0.064 | 0 | 0 | 0 |
| Azith | 0.9527 | 0.0717 | 0.4225 | 0.34 |
| Doxy | 0.9047 | 0.0707 | 0.3275 | 0.32 |

**Hospitalization Inferences**

| Pr(Superiority) | Pr(Meaningful Effect) |
|---|---|
| 0.5562 | 0.06 |
| 0.6805 | 0.0505 |
| 0.7072 | 0.0848 |

#### Hazard Ratio Estimates

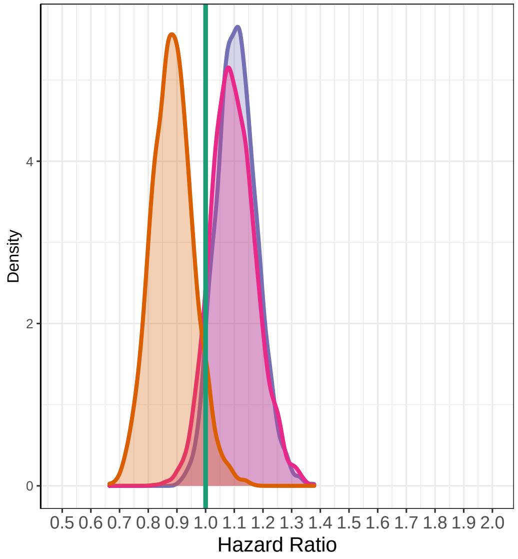

#### Median Time To Recovery Estimates

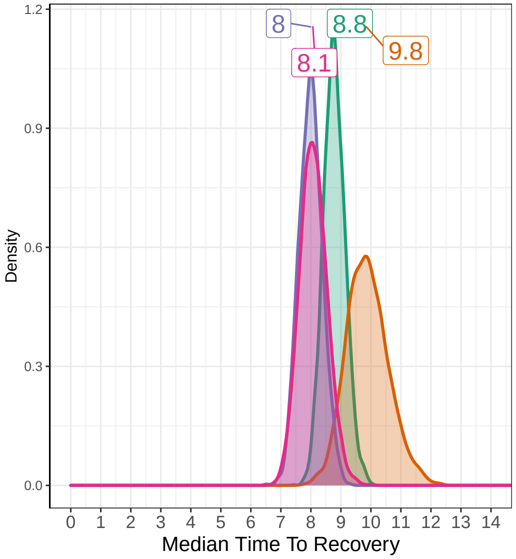

### Interim 4 on January 07, 2021

Recovery Data

| | Enrolled | Complete | Recovered | Exposure Days | Recoveries Per Day | Estimated Hazard | Estimated HR (95% interval) | Estimated Median Time to Recovery |
|---|---|---|---|---|---|---|---|---|
| SOC | 709 | 684 | 620 | 7506 | 0.083 | 0.08 | 1 | 8.89 |
| HCQ | 207 | 207 | 179 | 2453 | 0.073 | 0.071 | 0.886 (0.759, 1.029) | 9.92 |
| Azith | 553 | 516 | 484 | 5173 | 0.094 | 0.09 | 1.118 (1, 1.254) | 8.04 |
| Doxy | 388 | 367 | 347 | 3635 | 0.095 | 0.091 | 1.138 (1.003, 1.285) | 7.92 |
| Total | 1857 | 1774 | 1630 | 18768 | 0.087 | | | |

Hospitalization Data

| Hospitalizations | 28 Day Completers | Observed Hosp. Rate | Est. Hosp. Rate (95% interval) |
|---|---|---|---|
| 18 | 638 | 0.0282 | 0.0267 (0.0163, 0.0397) |
| 5 | 207 | 0.0242 | 0.0289 (0.0106, 0.0564) |
| 12 | 470 | 0.0255 | 0.0235 (0.0125, 0.0373) |
| 10 | 334 | 0.0299 | 0.0283 (0.0143, 0.0466) |
| 45 | 1649 | 0.0273 | |

| Intervention Status |
|---|
| Enrolling |
| Dropped |
| Enrolling |
| Enrolling |

Observed Patient Recoveries

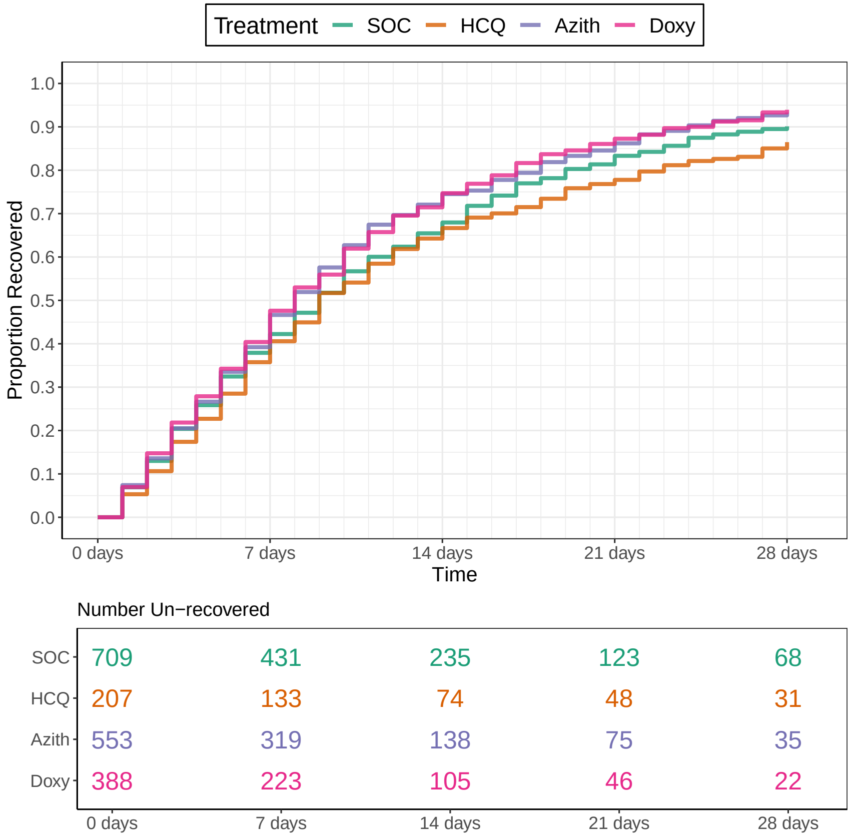

Recovery Inferences

| | Pr(Superiority) | Pr(Meaningful Effect) | Pr(Best) | Randomization Probability |
|---|---|---|---|---|
| SOC | | | | 0.33 |
| HCQ | 0.0633 | 0 | 0 | 0 |
| Azith | 0.9743 | 0.0653 | 0.2918 | 0.33 |
| Doxy | 0.978 | 0.128 | 0.4582 | 0.33 |

Hospitalization Inferences

| Pr(Superiority) | Pr(Meaningful Effect) |
|---|---|
| 0.4572 | 0.0332 |
| 0.643 | 0.0288 |
| 0.461 | 0.0102 |

Hazard Ratio Estimates

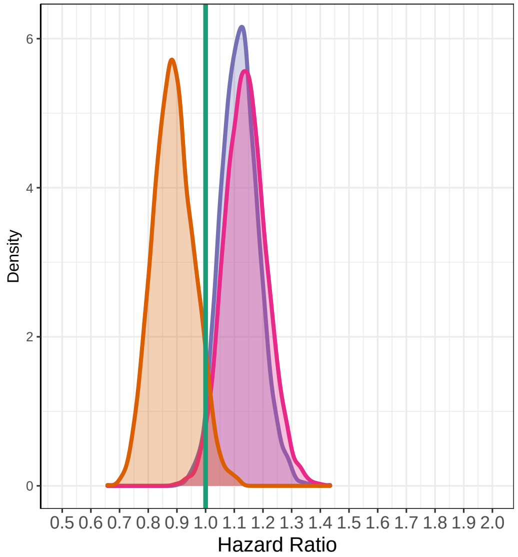

Median Time To Recovery Estimates

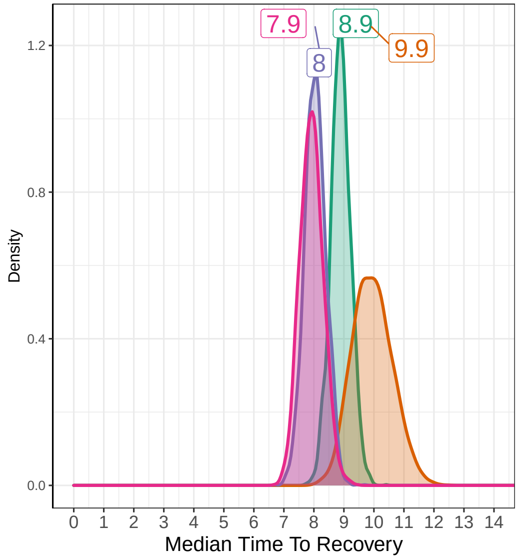

### Interim 5 on February 04, 2021

#### Recovery Data

| | Enrolled | Complete | Recovered | Exposure Days | Recoveries Per Day | Estimated Hazard | Estimated HR (95% interval) | Estimated Median Time to Recovery |
|---|---|---|---|---|---|---|---|---|
| SOC | 770 | 742 | 672 | 8161 | 0.082 | 0.081 | 1 | 8.97 |
| HCQ | 207 | 207 | 179 | 2453 | 0.073 | 0.072 | 0.892 (0.762, 1.037) | 9.94 |
| Azith | 631 | 597 | 561 | 6070 | 0.092 | 0.09 | 1.113 (0.996, 1.241) | 8.14 |
| Doxy | 445 | 426 | 403 | 4196 | 0.096 | 0.093 | 1.154 (1.018, 1.302) | 7.89 |
| Total | 2053 | 1972 | 1815 | 20881 | 0.087 | | | |

#### Hospitalization Data

| Hospitalizations | 28 Day Completers | Observed Hosp. Rate | Est. Hosp. Rate (95% interval) |
|---|---|---|---|
| 18 | 709 | 0.0254 | 0.0245 (0.0146, 0.0368) |
| 5 | 207 | 0.0242 | 0.0289 (0.0106, 0.0552) |
| 14 | 553 | 0.0253 | 0.0238 (0.0135, 0.0369) |
| 11 | 388 | 0.0284 | 0.0269 (0.0138, 0.0433) |
| 48 | 1857 | 0.0258 | |

| Intervention Status |
|---|
| Enrolling |
| Dropped |
| Enrolling |
| Enrolling |

#### Observed Patient Recoveries

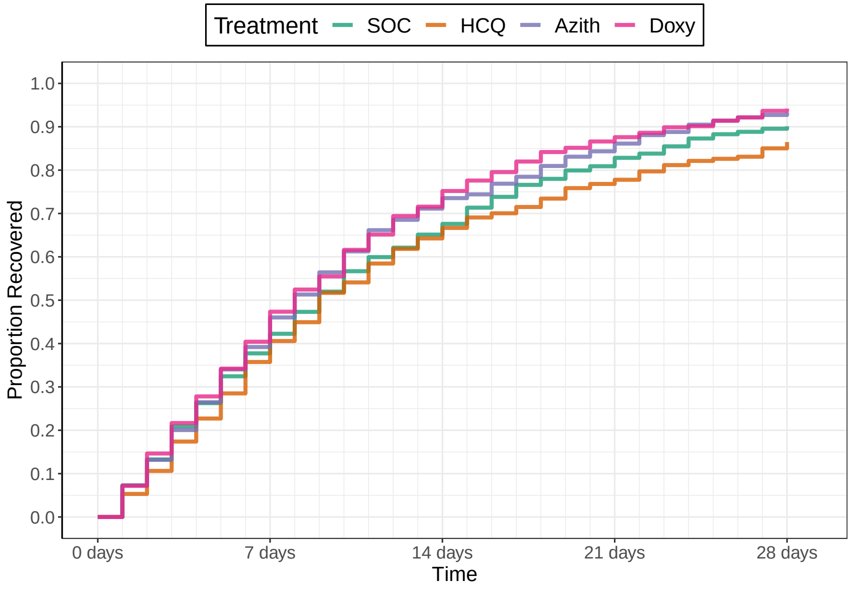

Number Un−recovered

| | 0 days | 7 days | 14 days | 21 days | 28 days |
|---|---|---|---|---|---|
| SOC | 770 | 469 | 253 | 138 | 74 |
| HCQ | 207 | 133 | 74 | 48 | 31 |
| Azith | 631 | 374 | 167 | 88 | 39 |
| Doxy | 445 | 258 | 118 | 54 | 25 |

#### Recovery Inferences

| | Pr(Superiority) | Pr(Meaningful Effect) | Pr(Best) | Randomization Probability |
|---|---|---|---|---|
| SOC | | | | 0.33 |
| HCQ | 0.069 | 0 | 0 | 0 |
| Azith | 0.9693 | 0.0623 | 0.2185 | 0.33 |
| Doxy | 0.9857 | 0.1877 | 0.5315 | 0.34 |

#### Hospitalization Inferences

| Pr(Superiority) | Pr(Meaningful Effect) |
|---|---|
| 0.3985 | 0.0142 |
| 0.5432 | 0.0082 |
| 0.4128 | 0.0068 |

#### Hazard Ratio Estimates

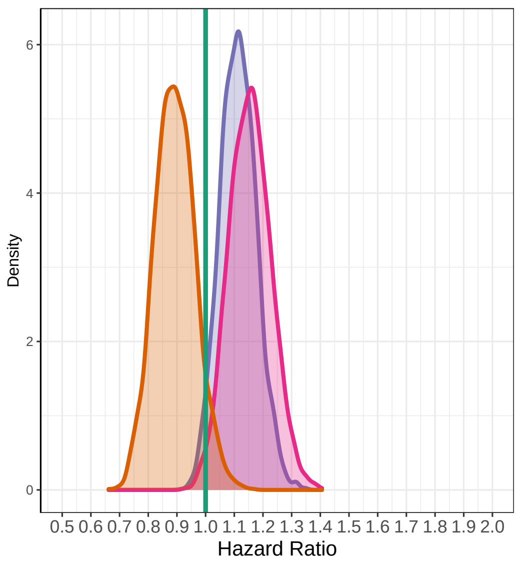

#### Median Time To Recovery Estimates

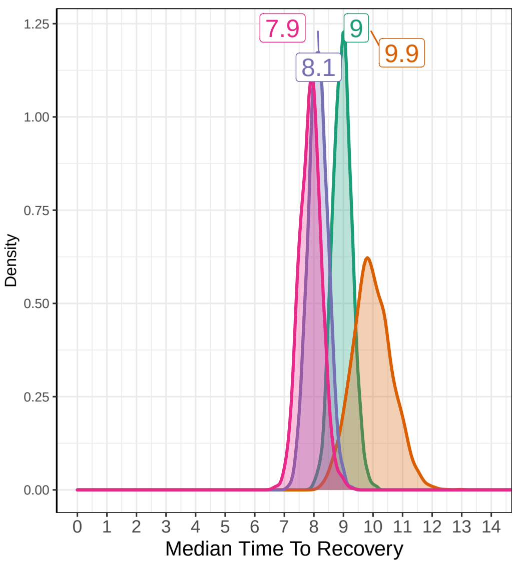

### Interim 6 on March 04, 2021

##### Recovery Data

| | Enrolled | Complete | Recovered | Exposure Days | Recoveries Per Day | Estimated Hazard | Estimated HR (95% interval) | Estimated Median Time to Recovery |
|---|---|---|---|---|---|---|---|---|
| SOC | 847 | 807 | 733 | 8965 | 0.082 | 0.08 | 1 | 9.02 |
| HCQ | 207 | 207 | 179 | 2453 | 0.073 | 0.072 | 0.895 (0.764, 1.043) | 9.97 |
| Azith | 695 | 673 | 629 | 6919 | 0.091 | 0.089 | 1.102 (0.995, 1.216) | 8.26 |
| Doxy | 506 | 482 | 458 | 4790 | 0.096 | 0.093 | 1.156 (1.037, 1.296) | 7.91 |
| Total | 2255 | 2169 | 1999 | 23127 | 0.086 | | | |

##### Hospitalization Data

| Hospitalizations | 28 Day Completers | Observed Hosp. Rate | Est. Hosp. Rate (95% interval) |
|---|---|---|---|
| 19 | 770 | 0.0247 | 0.0236 (0.0145, 0.0346) |
| 5 | 207 | 0.0242 | 0.0287 (0.0106, 0.0557) |
| 16 | 631 | 0.0254 | 0.0245 (0.0145, 0.0368) |
| 14 | 445 | 0.0315 | 0.0296 (0.0168, 0.0463) |
| 54 | 2053 | 0.0263 | |

| Intervention Status |
|---|
| LostOnTTR |
| Dropped |
| Futility |
| SuccessOnTTRFutilityOnHosp Not Announced |

##### Observed Patient Recoveries

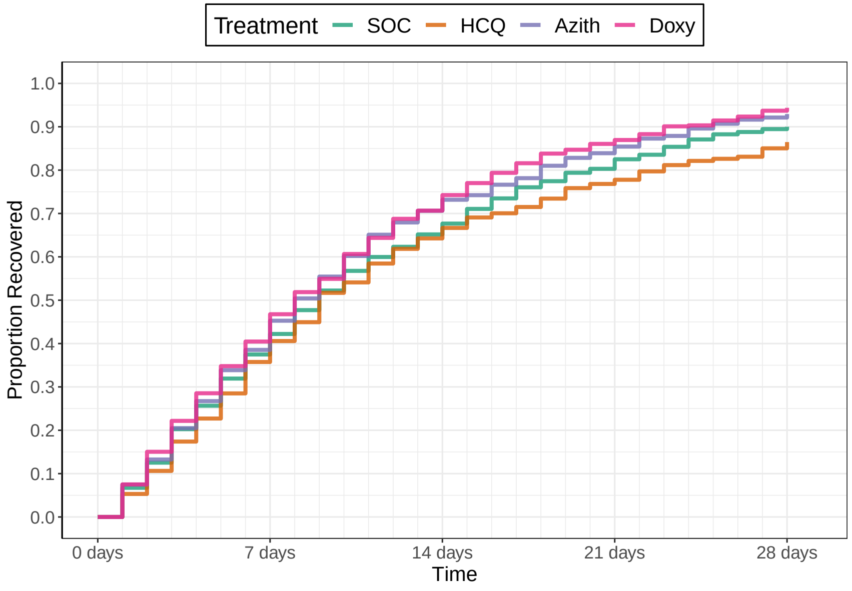

Number Un−recovered

| | 0 days | 7 days | 14 days | 21 days | 28 days |
|---|---|---|---|---|---|
| SOC | 847 | 516 | 279 | 152 | 78 |
| HCQ | 207 | 133 | 74 | 48 | 31 |
| Azith | 695 | 420 | 196 | 105 | 49 |
| Doxy | 506 | 293 | 139 | 62 | 28 |

##### Recovery Inferences

| | Pr(Superiority) | Pr(Meaningful Effect) | Pr(Best) | Randomization Probability |
|---|---|---|---|---|
| SOC | | | | 0.5 |
| HCQ | 0.0757 | 0 | 0 | 0 |
| Azith | 0.9667 | 0.0317 | 0 | 0 |
| Doxy | 0.9933 | 0.1847 | 0.75 | 0.5 |

##### Hospitalization Inferences

| Pr(Superiority) | Pr(Meaningful Effect) |
|---|---|
| 0.369 | 0.0078 |
| 0.464 | 0.0028 |
| 0.2502 | 0.0022 |

##### Hazard Ratio Estimates

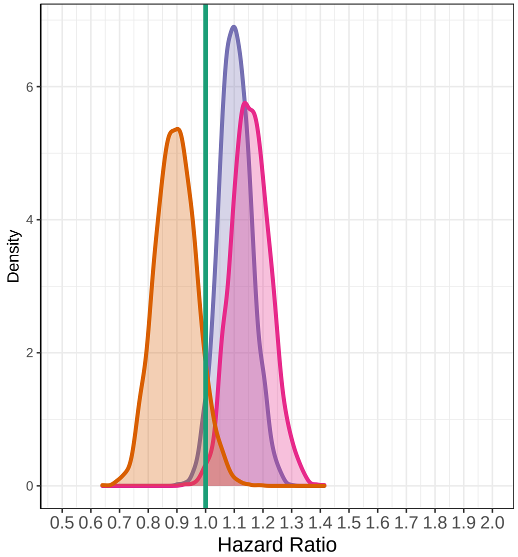

##### Median Time To Recovery Estimates

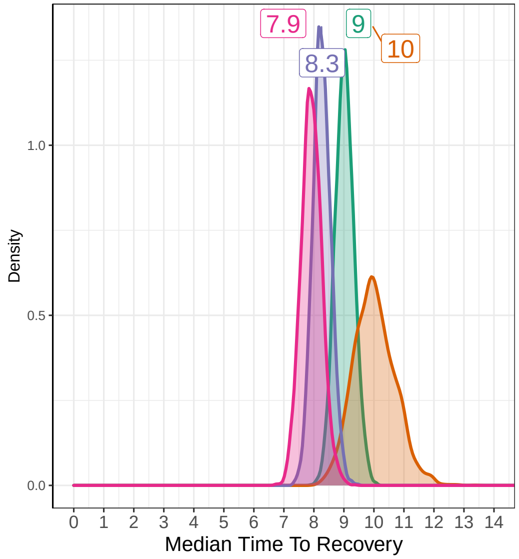

### Interim 7 on August 27, 2022

##### Recovery Data

| | Enrolled | Complete | Recovered | Exposure Days | Recoveries Per Day | Estimated Hazard | Estimated HR (95% interval) | Estimated Median Time to Recovery |
|---|---|---|---|---|---|---|---|---|
| SOC | 2688 | 2688 | 2373 | 30880 | 0.077 | 0.074 | 1 | 9.39 |
| HCQ | 207 | 207 | 179 | 2453 | 0.073 | 0.07 | 0.948 (0.82, 1.088) | 9.86 |
| Azith | 695 | 695 | 646 | 7084 | 0.091 | 0.087 | 1.17 (1.074, 1.275) | 8.1 |
| Doxy | 2410 | 2410 | 2264 | 22450 | 0.101 | 0.096 | 1.293 (1.22, 1.368) | 7.37 |
| Total | 6000 | 6000 | 5462 | 62867 | 0.087 | | | |

##### Hospitalization Data

| Hospitalizations | 28 Day Completers | Observed Hosp. Rate | Est. Hosp. Rate (95% interval) |
|---|---|---|---|
| 88 | 2688 | 0.0327 | 0.0331 (0.0266, 0.0403) |
| 5 | 207 | 0.0242 | 0.0284 (0.0105, 0.0557) |
| 16 | 695 | 0.0230 | 0.0243 (0.0143, 0.037) |
| 68 | 2410 | 0.0282 | 0.0286 (0.0224, 0.0355) |
| 177 | 6000 | 0.0295 | |

| Intervention Status |
|---|
| Complete |
| Dropped |
| Dropped |
| SuccessOnTTRFutilityOnHosp *Announced* |

##### Observed Patient Recoveries

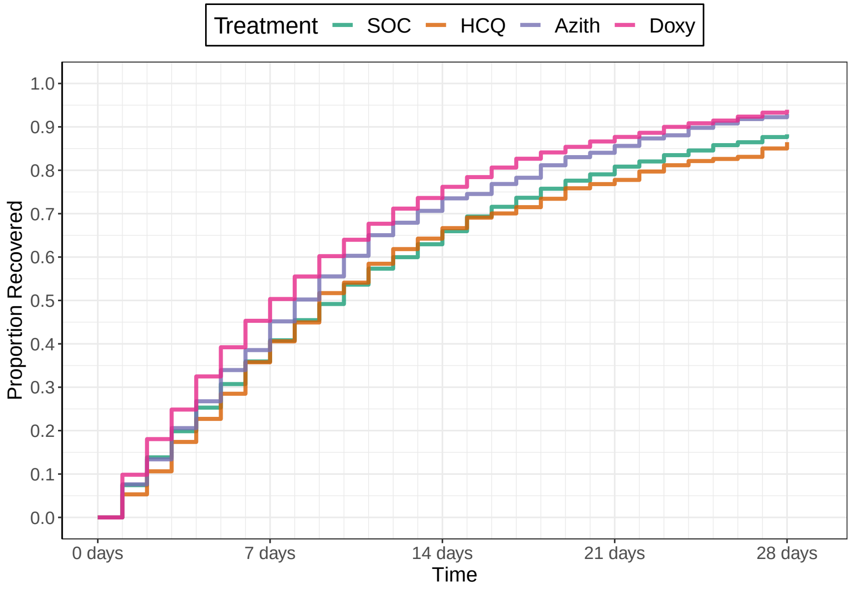

Number Un−recovered

| | 0 days | 7 days | 14 days | 21 days | 28 days |
|---|---|---|---|---|---|
| SOC | 2688 | 1722 | 996 | 563 | 332 |
| HCQ | 207 | 133 | 74 | 48 | 31 |
| Azith | 695 | 427 | 204 | 111 | 54 |
| Doxy | 2410 | 1318 | 636 | 322 | 162 |

##### Recovery Inferences

| | Pr(Superiority) | Pr(Meaningful Effect) | Pr(Best) | Randomization Probability |
|---|---|---|---|---|
| SOC | | | | |
| HCQ | 0.2353 | 3e−04 | 0 | |
| Azith | 1 | 0.271 | 0 | |
| Doxy | 1 | 0.9847 | 0.75 | |

##### Hospitalization Inferences

| Pr(Superiority) | Pr(Meaningful Effect) |
|---|---|
| 0.6968 | 0.0802 |
| 0.9035 | 0.0392 |
| 0.8315 | 8e−04 |

##### Hazard Ratio Estimates

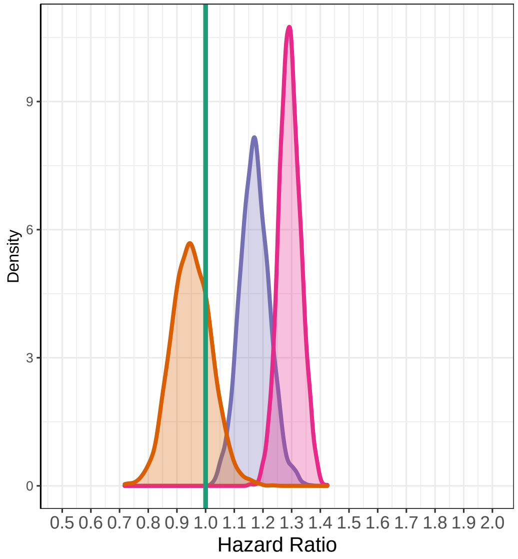

##### Median Time To Recovery Estimates

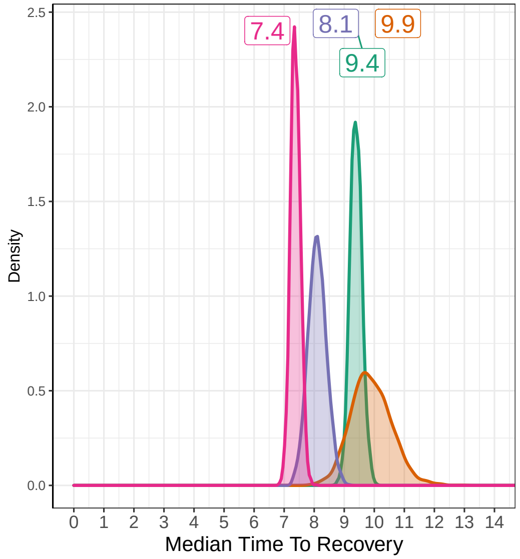

#### 6.6 Computational Details

In the simulations, relevant covariates (age, comorbidity, vaccination status) are regarded as nuisance variables, and hence are not factored into the simulations. For computational efficiency, the primary analysis for the hospitalization endpoint was simulated using beta-binomial posterior distributions; however logistic regression will be used during execution to account for the covariates (as specified in Section 3.2). Operating characteristics are based on a minimum of 1,000 simulations per scenario. Each Bayesian MCMC fit includes at least 4,000 posterior samples with a burn-in of at least 1,000 samples. In trial execution, at least 10,000 MCMC samples will be taken from the Bayesian posterior distribution after an appropriate burn-in. The R software package was used to summarize the simulation output and to create graphics and tables for this report. This document was typeset with LaTeX.

Figure S1 Estimated mean and 95% confidence interval of daily rating of feeling well over 28 days by treatment arm (concurrent randomisation and eligible SARS-CoV-2 positive population)

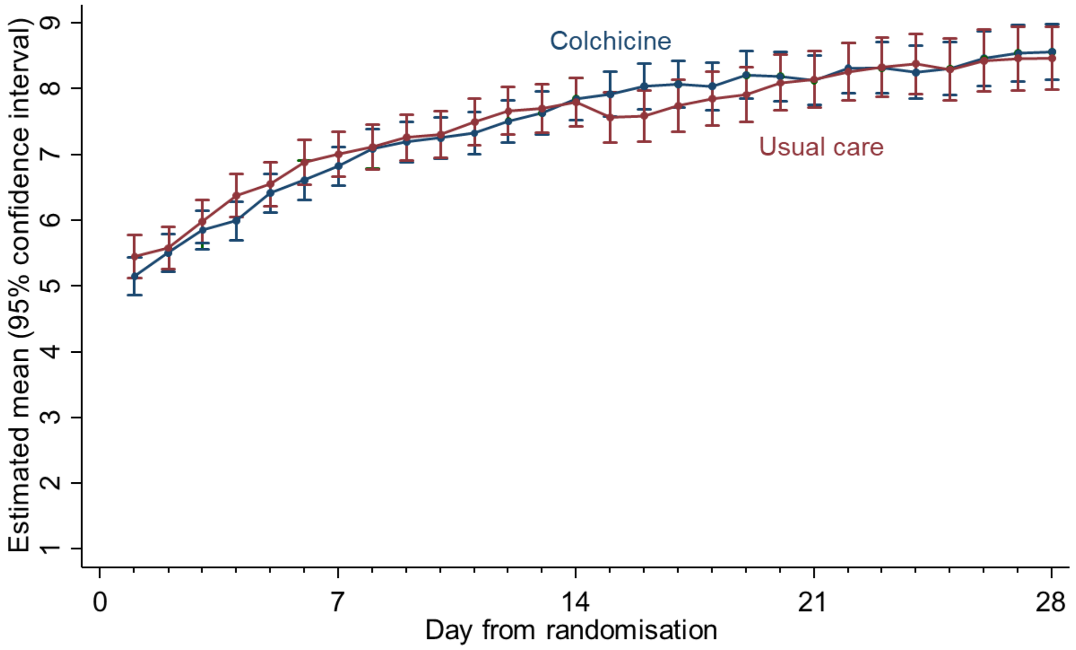

Figure S2 Summary and results of the time to sustained recovery (concurrent randomisation and eligible SARS-CoV-2 positive population population)

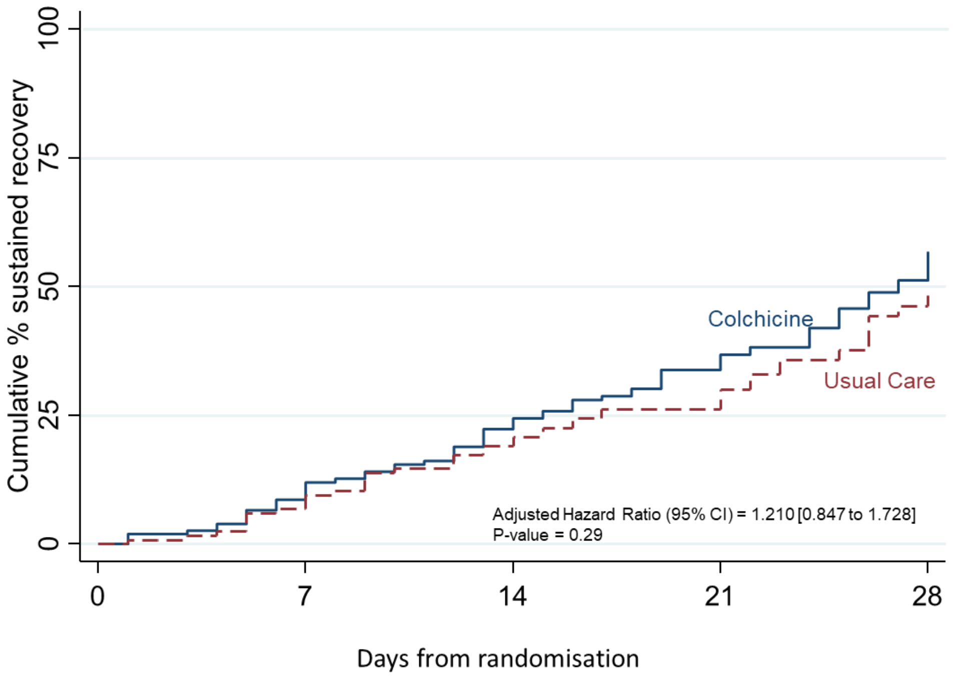

Cumulative number not yet recovered (recovered)

| | | | | | |
|---|---|---|---|---|---|
| Colchicine | 156 (0) | 135 (18) | 112 (36) | 90 (53) | 61 (79) |
| Usual Care | 120 (0) | 107 (8) | 93 (24) | 78 (37) | 54 (53) |

Figure S3 Secondary Time-to-Event Outcomes (the concurrent randomisation and eligible SARS-CoV-2 positive population)

a) Time to alleviation of all symptoms, and each symptom

| Time to Alleviation of Symptom[1] | Colchicine | Usual Care | Adjusted HR (95% CI) | P-value |
|---|---|---|---|---|
| All symptoms | 125/141 | 98/111 | 0.94 ( 0.72, 1.24) | 0.67 |
| Fever | 35/36 | 18/18 | 0.61 ( 0.33, 1.11) | 0.10 |
| Cough | 70/73 | 61/63 | 0.93 ( 0.65, 1.33) | 0.69 |
| Shortness of breath | 64/66 | 35/38 | 1.19 ( 0.78, 1.82) | 0.43 |
| Muscle ache | 62/67 | 58/60 | 1.00 ( 0.69, 1.43) | 0.99 |
| Nausea-vomiting | 18/18 | 13/13 | 0.88 ( 0.39, 1.96) | 0.75 |
| Diarrhoea | 9/10 | 12/12 | 0.61 ( 0.23, 1.64) | 0.33 |
| Generally unwell | 99/105 | 85/87 | 0.83 ( 0.61, 1.13) | 0.24 |
| Loss of smell or taste | 73/78 | 43/48 | 1.11 (0.75, 1.63) | 0.59 |
| Headache | 66/68 | 40/41 | 1.08 ( 0.72, 1.63) | 0.69 |
| Abdominal pain | 17/17 | 8/8 | 1.27 ( 0.54, 3.00) | 0.58 |

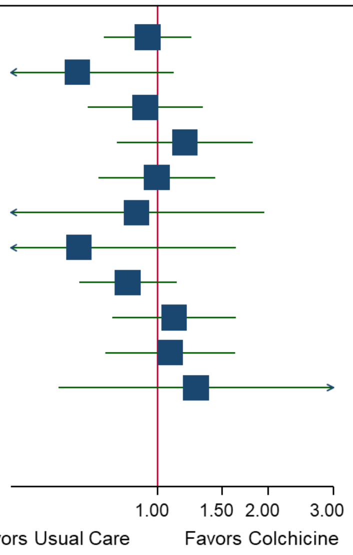

[1]Time to alleviation of symptoms is defined as time from randomisation to all symptoms or each symptom being rated as mild or none, among participants who reported that symptom at baseline.

(b) Time to sustained alleviation of all symptoms, each symptom

| Time to Sustained Alleviation[1] | Colchicine | Usual Care | Adjusted HR (95% CI) | P-value |
|---|---|---|---|---|
| All symptoms | 100/141 | 74/111 | 1.11 (0.81, 1.52) | 0.51 |
| Fever | 27/36 | 13/18 | 0.82 ( 0.41, 1.64) | 0.58 |
| Cough | 59/73 | 49/63 | 1.09 ( 0.73, 1.63) | 0.67 |
| Shortness of breath | 53/66 | 20/38 | 1.96 ( 1.15, 3.35) | 0.014 |
| Muscle ache | 47/67 | 40/60 | 1.25 ( 0.81, 1.91) | 0.31 |
| Nausea-vomiting | 13/18 | 9/13 | 0.90 ( 0.35, 2.35) | 0.83 |
| Diarrhoea | 8/10 | 9/12 | 0.78 ( 0.25, 2.50) | 0.68 |
| Generally unwell | 83/105 | 71/87 | 1.02 ( 0.74, 1.42) | 0.89 |
| Loss of smell or taste | 61/78 | 39/48 | 0.82 ( 0.54, 1.24) | 0.34 |
| Headache | 55/68 | 29/41 | 1.56 ( 0.97, 2.52) | 0.067 |
| Abdominal pain | 13/17 | 6/8 | 1.02 ( 0.38, 2.76) | 0.96 |

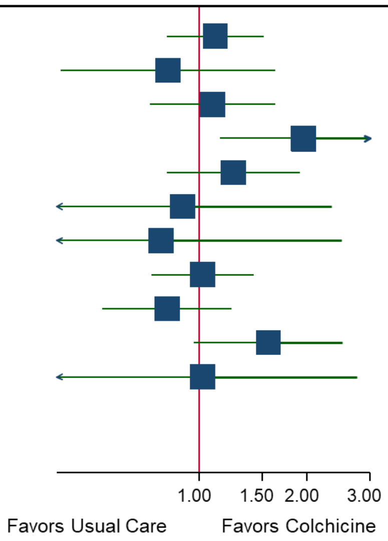

[1]Time to sustained alleviation of symptoms is defined as the time to alleviation of symptoms, with no subsequent symptom being rated as greater than mild.

#### (c) Time to initial reduction of severity of all symptoms, and each symptom

| Time to Initial Reduction of Severity of Symptom[1] | Colchicine | Usual Care | Adjusted HR (95% CI) | P-value |
|---|---|---|---|---|
| All symptoms | 119/156 | 89/120 | 0.96 (0.73, 1.27) | 0.78 |
| Fever | 84/87 | 59/60 | 0.79 (0.55, 1.12) | 0.19 |
| Cough | 129/142 | 106/114 | 0.92 (0.71, 1.20) | 0.54 |
| Shortness of breath | 122/137 | 74/83 | 0.96 (0.72, 1.29) | 0.79 |
| Muscle ache | 110/119 | 90/93 | 0.86 (0.65, 1.15) | 0.32 |
| Nausea-vomiting | 43/44 | 33/33 | 0.73 (0.45, 1.19) | 0.21 |
| Diarrhoea | 27/29 | 30/30 | 0.61 (0.34, 1.09) | 0.093 |
| Generally unwell | 143/152 | 114/118 | 0.88 (0.69, 1.14) | 0.35 |
| Loss of smell or taste | 94/103 | 60/66 | 1.01 (0.72, 1.42) | 0.96 |
| Headache | 104/110 | 79/81 | 0.95 (0.71, 1.29) | 0.76 |
| Abdominal pain | 32/33 | 20/21 | 1.08 (0.60, 1.92) | 0.80 |

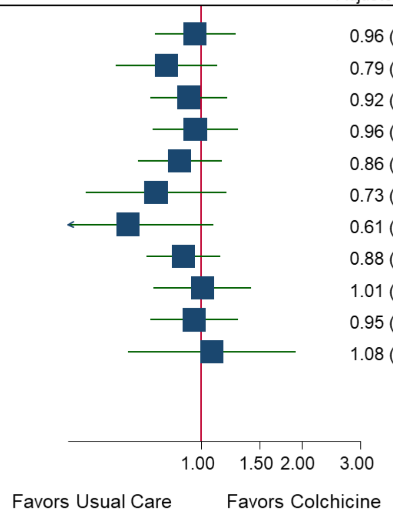

[1]Time to initial reduction of severity of symptoms is defined as time to reduction in severity of each individual symptoms to at least one grade lower among participants who reported that symptom at baseline.

Table S1 Baseline characteristics for participants irrespective of SARS-CoV-2 status by treatment group population

| | Colchicine (N=206) | Usual Care (N=2050) | Overall (N=2256) |
|---|---|---|---|
| **Age, mean(SD)** | 48·5 (13·2) | 61·7 (9·0) | 60·5 (10·2) |
| **Age category, n(%)** | | | |
| *18-49* | 99 (48·1%) | 45 (2·2%) | 144 (6·4%) |
| *50-64* | 75 (36·4%) | 1074 (52·4%) | 1149 (50·9%) |
| *65 and over* | 32 (15·5%) | 931 (45·4%) | 963 (42·7%) |
| **Sex, n(%)** | | | |
| *Female* | 98 (47·6%) | 1118 (54·5%) | 1216 (53·9%) |
| *Male* | 107 (51·9%) | 932 (45·5%) | 1039 (46·1%) |
| *Other* | 1 (0·5%) | 0 (0·0%) | 1 (0·0%) |
| **Ethnicity†, n(%)** | | | |
| *White* | 180 (87·4%) | 1797 (87·7%) | 1977 (87·6%) |
| *Mixed background* | 3 (1·5%) | 29 (1·4%) | 32 (1·4%) |
| *South Asian* | 20 (9·7%) | 105 (5·1%) | 125 (5·5%) |
| *Black* | 1 (0·5%) | 8 (0·4%) | 9 (0·4%) |
| *Other* | 2 (1·0%) | 22 (1·1%) | 24 (1·1%) |
| *Missing, n(%)* | 0 (0·0%) | 89 (4·3%) | 89 (3·9%) |
| **Index of multiple deprivation quintile, n(%)** | | | |
| *(Most Deprived) 1* | 49 (23·8%) | 368 (18·0%) | 417 (18·5%) |
| *2* | 41 (19·9%) | 346 (16·9%) | 387 (17·2%) |
| *3* | 42 (20·4%) | 410 (20·0%) | 452 (20·0%) |
| *4* | 37 (18·0%) | 448 (21·9%) | 485 (21·5%) |
| *(Least Deprived) 5* | 37 (18·0%) | 477 (23·3%) | 514 (22·8%) |
| *Missing, n(%)* | 0 (0·0%) | 1 (0·0%) | 1 (0·0%) |
| **Duration of illness prior to randomisation, median(IQR)** | 6·0 (4·0 to 8·0) | 6·0 (4·0 to 9·0) | 6·0 (4·0 to 9·0) |
| **Smoking status, n(%)** | | | |
| *Current smoker* | 26 (12·6%) | 206 (10·0%) | 232 (10·3%) |
| *Former smoker* | 81 (39·3%) | 787 (38·4%) | 868 (38·5%) |
| *Never smoker* | 98 (47·6%) | 1018 (49·7%) | 1116 (49·5%) |
| *Missing, n(%)* | 1 (0·5%) | 39 (1·9%) | 40 (1·8%) |
| **Swab result, n(%)** | | | |
| *Negative* | 25 (12·1%) | 567 (27·7%) | 592 (26·2%) |
| *Positive* | 174 (84·5%) | 1207 (58·9%) | 1381 (61·2%) |
| *No result* | 0 (0·0%) | 9 (0·4%) | 9 (0·4%) |
| *Missing, n(%)* | 7 (3·4%) | 267 (13·0%) | 274 (12·1%) |
| **Vaccination status, n(%)** | 122 (59·2%) | 169 (8·2%) | 291 (12·9%) |
| **Vaccine doses received, n(%)** | | | |
| *One dose* | 89 (43·2%) | 141 (6·9%) | 230 (10·2%) |
| *Two doses* | 33 (16·0%) | 28 (1·4%) | 61 (2·7%) |
| **Comorbidity, n(%)** | 200 (97·1%) | 1742 (85·0%) | 1942 (86·1%) |
| **Comorbidities** | | | |
| **Asthma, COPD or lung disease, n(%)** | 70 (34·0%) | 492 (24·0%) | 562 (24·9%) |
| **Diabetes, n(%)** | 25 (12·1%) | 423 (20·6%) | 448 (19·9%) |
| **Heart problems‡, n(%)** | 20 (9·7%) | 296 (14·4%) | 316 (14·0%) |
| **High blood pressure required medication, n(%)** | 46 (22·3%) | 869 (42·4%) | 915 (40·6%) |
| **Liver disease, n(%)** | 1 (0·5%) | 57 (2·8%) | 58 (2·6%) |
| **Stroke or other neurological problem, n(%)** | 10 (4·9%) | 116 (5·7%) | 126 (5·6%) |
| **Taking angiotensin-converting enzyme inhibitor§, n(%)** | 21 (10·2%) | 428 (20·9%) | 449 (19·9%) |
| **Baseline symptoms** | | | |
| **Fever, n(%)** | | | |
| *No problem* | 87 (42·2%) | 910 (44·4%) | 997 (44·2%) |
| *Mild problem* | 75 (36·4%) | 710 (34·6%) | 785 (34·8%) |
| *Moderate problem* | 39 (18·9%) | 368 (18·0%) | 407 (18·0%) |
| *Major problem* | 5 (2·4%) | 62 (3·0%) | 67 (3·0%) |
| **Cough, n(%)** | | | |
| *No problem* | 18 (8·7%) | 328 (16·0%) | 346 (15·3%) |
| *Mild problem* | 86 (41·7%) | 860 (42·0%) | 946 (41·9%) |
| *Moderate problem* | 91 (44·2%) | 730 (35·6%) | 821 (36·4%) |
| *Major problem* | 11 (5·3%) | 132 (6·4%) | 143 (6·3%) |
| **Shortness of breath, n(%)** | | | |
| *No problem* | 26 (12·6%) | 813 (39·7%) | 839 (37·2%) |
| *Mild problem* | 93 (45·1%) | 814 (39·7%) | 907 (40·2%) |
| *Moderate problem* | 79 (38·3%) | 371 (18·1%) | 450 (19·9%) |
| *Major problem* | 8 (3·9%) | 52 (2·5%) | 60 (2·7%) |
| **Muscle ache, n(%)** | | | |
| *No problem* | 51 (24·8%) | 561 (27·4%) | 612 (27·1%) |
| *Mild problem* | 64 (31·1%) | 755 (36·8%) | 819 (36·3%) |
| *Moderate problem* | 68 (33·0%) | 545 (26·6%) | 613 (27·2%) |

| | Colchicine (N=206) | Usual Care (N=2050) | Overall (N=2256) |
|---|---|---|---|
| *Major problem* | 23 (11·2%) | 189 (9·2%) | 212 (9·4%) |
| **Nausea, n(%)** | | | |
| *No problem* | 156 (75·7%) | 1470 (71·7%) | 1626 (72·1%) |
| *Mild problem* | 29 (14·1%) | 427 (20·8%) | 456 (20·2%) |
| *Moderate problem* | 17 (8·3%) | 122 (6·0%) | 139 (6·2%) |
| *Major problem* | 4 (1·9%) | 31 (1·5%) | 35 (1·6%) |
| **Feeling generally unwell, n(%)** | | | |
| *No problem* | 4 (1·9%) | 99 (4·8%) | 103 (4·6%) |
| *Mild problem* | 65 (31·6%) | 703 (34·3%) | 768 (34·0%) |
| *Moderate problem* | 112 (54·4%) | 786 (38·3%) | 898 (39·8%) |
| *Major problem* | 25 (12·1%) | 236 (11·5%) | 261 (11·6%) |
| *Missing, n(%)* | 0 (0·0%) | 226 (11·0%) | 226 (10·0%) |
| **Diarrhea, n(%)** | | | |
| *No problem* | 167 (81·1%) | 1365 (66·6%) | 1532 (67·9%) |
| *Mild problem* | 26 (12·6%) | 313 (15·3%) | 339 (15·0%) |
| *Moderate problem* | 12 (5·8%) | 110 (5·4%) | 122 (5·4%) |
| *Major problem* | 1 (0·5%) | 36 (1·8%) | 37 (1·6%) |
| *Missing, n(%)* | 0 (0·0%) | 226 (11·0%) | 226 (10·0%) |
| **Loss of sense of smell/taste, n(%)** | | | |
| *No problem* | 66 (32·0%) | 62 (3·0%) | 128 (5·7%) |
| *Mild problem* | 32 (15·5%) | 27 (1·3%) | 59 (2·6%) |
| *Moderate problem* | 42 (20·4%) | 23 (1·1%) | 65 (2·9%) |
| *Major problem* | 51 (24·8%) | 33 (1·6%) | 84 (3·7%) |
| *Missing, n(%)* | 15 (7·3%) | 1905 (92·9%) | 1920 (85·1%) |
| **Headache, n(%)** | | | |
| *No problem* | 50 (24·3%) | 35 (1·7%) | 85 (3·8%) |
| *Mild problem* | 56 (27·2%) | 55 (2·7%) | 111 (4·9%) |
| *Moderate problem* | 66 (32·0%) | 41 (2·0%) | 107 (4·7%) |
| *Major problem* | 19 (9·2%) | 14 (0·7%) | 33 (1·5%) |
| *Missing, n(%)* | 15 (7·3%) | 1905 (92·9%) | 1920 (85·1%) |
| **Abdominal pain, n(%)** | | | |
| *No problem* | 150 (72·8%) | 116 (5·7%) | 266 (11·8%) |
| *Mild problem* | 21 (10·2%) | 17 (0·8%) | 38 (1·7%) |
| *Moderate problem* | 18 (8·7%) | 11 (0·5%) | 29 (1·3%) |
| *Major problem* | 2 (1·0%) | 1 (0·0%) | 3 (0·1%) |
| *Missing, n(%)* | 15 (7·3%) | 1905 (92·9%) | 1920 (85·1%) |
| **Use of healthcare services** | | | |
| **Have you taken antibiotics since your illness started, n(%)** | 20 (9·7%) | 129 (6·3%) | 149 (6·6%) |
| *Missing, n(%)* | 0 (0·0%) | 2 (0·1%) | 2 (0·1%) |
| **GP, n(%)** | 57 (27·7%) | 524 (25·6%) | 581 (25·8%) |
| **Other primary care services, n(%)** | 6 (2·9%) | 157 (7·7%) | 163 (7·2%) |
| **NHS 111, n(%)** | 36 (17·5%) | 289 (14·1%) | 325 (14·4%) |
| **A&E, n(%)** | 7 (3·4%) | 29 (1·4%) | 36 (1·6%) |
| **Other healthcare services, n(%)** | 5 (2·4%) | 44 (2·1%) | 49 (2·2%) |
| **Rating of how well participant feels (1 worst, 10 best), mean (SD)** | 4·9 (1·4) | 5·1 (1·6) | 5·0 (1·5) |
| *Missing, n(%)* | 15 (0·2%) | 1905 (21·1%) | 1920 (21·3%) |
| **Well-being (WHO5 Questionnaire) ¶, mean(SD)** | 44·8 (23·5) | 47·7 (25·1) | 47·5 (25·0) |
| *Missing, n(%)* | 0 (0·0%) | 24 (0·3%) | 24 (0·3%) |

* Includes participants randomised before the colchicine arm was open· For comparison of concurrently randomised usual care and colchicine participants, see Table S1·

† Data on ethnicity were collected retrospectively via notes review before July 2020

‡ E·g· angina, heart attack, heart failure, atrial fibrillation, valve problems

§ Such as Ramipril, Lisinopril, Perindopril, Captopril or Enalapril

¶ Well-being is measured using the WHO well-being index which includes 5 items relating to well-being measured on a five point scale· A total score is computed by summing the scores to the five individual questions to give a raw score ranging from 0 to 25 which is then multiplied by 4 to give the final score from 0 representing the worst imaginable well-being to 100 representing the best imaginable well-being·

Table S2 Reasons for hospitalisations/deaths unrelated to COVID-19

| Description | Colchicine | Usual Care |
|---|---|---|
| Heart valve surgery | | 1 |
| Back pain, paraesthesia, investigated for spinal cord compression | 1 | |
